# Human germline biallelic complete NFAT1 deficiency causes the triad of progressive joint contractures, osteochondromas, and susceptibility to B cell malignancy

**DOI:** 10.1101/2022.01.30.22269378

**Authors:** Mehul Sharma, Maggie P. Fu, Henry Y. Lu, Ashish A. Sharma, Bhavi P. Modi, Christina Michalski, Susan Lin, Joshua Dalmann, Areesha Salman, Kate L. Del Bel, Meriam Waqas, Jefferson Terry, Audi Setiadi, Pascal M. Lavoie, Wyeth W. Wasserman, Jill Mwenifumbo, Michael S. Kobor, Anna F. Lee, Anna Lehman, the CAUSES Study, Sylvia Cheng, Anthony Cooper, Millan S. Patel, Stuart E. Turvey

**Affiliations:** Department of Pediatrics, BC Children’s Hospital, The University of British Columbia, Vancouver, British Columbia, Canada; Experimental Medicine Program, Faculty of Medicine, The University of British Columbia, British Columbia, Canada; Department of Medical Genetics, BC Children’s Hospital Research Institute, University of British Columbia, Vancouver, British Columbia, Canada; Genome Science and Technology Program, Faculty of Science, The University of British Columbia, Vancouver, British Columbia, Canada; Department of Pathology, Emory University, Atlanta, GA, USA; Department of Pathology and Laboratory Medicine, BC Children’s Hospital, Vancouver, British Columbia, Canada; Centre for Molecular Medicine and Therapeutics, Vancouver, British Columbia, Canada; Department of Orthopaedics, Faculty of Medicine, University of British Columbia

## Abstract

Discovery of humans with monogenic disorders has a rich history of generating new insights into biology. Here we report the first human identified with complete deficiency of nuclear factor of activated T cells 1 (NFAT1). NFAT1, encoded by *NFATC2*, mediates calcium-calcineurin signals that drive cell activation, proliferation, and survival. The patient is homozygous for a damaging germline *NFATC2* variant (c.2023_2026delTACC; p.Tyr675Thrfs*18) and presented with joint contractures, osteochondromas, and B cell lymphoma. Absence of NFAT1 protein in chondrocytes caused enrichment in pro-survival and inflammatory genes. Systematic single-cell-omic analyses revealed an environment that promotes lymphomagenesis with accumulation of naïve B cells (with oncogenic signatures - *MYC*, *JAK1*), exhausted CD4^+^ T cells, impaired T follicular helper cells, and aberrant CD8^+^ T cells. This work highlights the pleiotropic role of human NFAT1, will empower the diagnosis of additional patients with NFAT1 deficiency, and further define detrimental effects a long-term use of calcineurin inhibitors.

## INTRODUCTION

Intracellular calcium signaling orchestrates a diverse range of critical physiological and immunological processes, including lymphocyte activation, differentiation, and effector function^1, 2^. T cell receptor (TCR) or B cell receptor (BCR) engagement activates phospholipase C (PLC), generating inositol-1,4,5-triphosphate (IP3), which, in turn, activates IP3 receptors and creates an influx of Ca^2+^ from the endoplasmic reticulum into the cytosol^2^. Free Ca^2+^ is subsequently bound by calmodulin, resulting in a conformation change that promotes binding of the serine/threonine phosphatase calcineurin^3, 4^. This calmodulin-calcineurin interaction activates an array of transcription factor families, including the nuclear factor of activated T cells (NFAT) family. This family consists of five members: NFAT1 (NFATc2, NFATp), NFAT2 (NFATc1, NFATc), NFAT3 (NFATc4), NFAT4 (NFATc3, NFATx) and NFAT5^5^. These proteins contain an N-terminal transactivation domain (TAD), a regulatory NFAT homology region (NHR), a DNA-binding domain (REL-homology domain, [RHD]) and a carboxy-terminal domain. Activation of NFAT1-4 is dependent on calcineurin-mediated dephosphorylation of the NHR, which exposes a nuclear localization signal and leads to NFAT translocation to be nucleus and transcriptional regulation^5–12^.

Originally discovered in activated T cells as a protein complex that binds the interleukin-2 (IL-2) promoter^13^, our understanding of the NFAT family has grown remarkably over the past few decades. It is now appreciated that NFATs have a role in the differentiation and activation of many T helper subsets, including T_H_1, T_H_2, T_H_17, T follicular helper cells (T_FH_), and T regulatory cells (Tregs)^14, 15^. NFATs are also expressed by and have a function in other immune cells including B cells^16^ and dendritic cells^17^, as well as in non-immune cells, such as cartilage cells^18–20^, adipocytes^21^, cardiac cells^22^ and breast cancer cells^23^. Despite our expanded understanding of this transcription factor family, the specific function of each individual NFAT protein has been difficult to define fully due to the redundancy amongst the family members. This is due to highly homologous RHDs, which all bind to a core DNA sequence of (A/T)GGAAA^11^. On the other hand, their non-redundant roles are attributed to their interaction with other transcription factors via their dissimilar TADs^24^. Furthermore, NFATs can also alter the function of other transcription factors by directly competing for binding sites, as has been reported with nuclear factor kappa B (NF-κB)^25^.

A particularly illustrative example of how critical calcineurin and NFATs are in immunity and clinical medicine is the fact that calcineurin inhibitors (e.g. cyclosporin A (CsA), tacrolimus [FK506], and pimecrolimus), which suppress T cells without significant myelotoxicity, have served as the standard of care over the past 40 years for preventing organ rejection and graft-versus-host disease in transplant recipients^10, 26–28^. Further, they have also been used successfully to treat a variety of immune-mediated disorders including atopic dermatitis, psoriasis, rheumatoid arthritis, and uveitis^28, 29^. However, despite the effectiveness of calcineurin inhibitors, serious adverse side effects have been reported, including nephrotoxicity, lymphomas, and increased susceptibility to infections^30^. Insight into NFAT biology in humans is thus critical to understand the mechanisms underlying the side-effects of these drugs.

One strategy to study human NFAT biology has been to use calcineurin inhibitors^31^. However, since they inhibit all members of the NFAT family, the role of individual members has been challenging to define. Similarly, although targeted genetic disruption of individual NFAT family members in mice has helped uncover their specific functions in murine immunity, this approach is not fully transferable to the human system. A powerful complementary approach is to identify and study rare individuals who carry damaging germline variants in genes encoding members of the NFAT family, allowing us to investigate their roles in an unmodified human system. Here, we report the identification and characterization of the first human with complete NFAT1 deficiency.

## METHODS

### Study participants and consent

All study participants and/or their parents/guardians provided written informed consent to participate. All individuals also provided consent to be published. Research study protocols (H18-02853 and H15-00092) were approved by The University of British Columbia Clinical Research Ethics Board.

### Identification of variant and confirmation by Sanger sequencing

Trio whole exome sequencing (WES) was performed on genomic DNA from the patient and both parents on an Illumina platform at Ambry Genetics (Aliso Viejo, California, USA) after capture of targeted regions using xGen™ Exome Hybridization Panel (IDT, Coraville, Iowa, USA). More than 99% of targeted regions were covered by at least 10 reads that averaged 150 bp in length. Reads were aligned to GRCh37/Hg19 and variants were called using BWA-MEM, GATK 3.5-0, SAMtools 0.1.19, and Picard 1.139, and then annotated and prioritized with VarSeq v.1.5 (Golden Helix, Bozeman, Montana, USA). WES data was analyzed using an updated version of our in-house, open-source, semi-automated bioinformatics pipeline that has been previously described^32, 33^. A predicted null *NFATC2* variant (c.2023_2026delTACC, p.Tyr675Thrfs*18) was selected for further analysis on the basis of: i) the clinical phenotype matching closely to that of *Nfatc2* knockout mice^18, 34^; ii) the variant resulting in a premature stop codon; and iii) the variant segregating with the disease in the extended family. The variant was confirmed by standard Sanger sequencing of genomic DNA from all individuals of the family. Briefly, genomic DNA was extracted from whole blood (QIAmp DNA Blood Mini Kit; Qiagen, Hilden, Germany) and buccal swabs (DNeasy Blood and Tissue Kit; Qiagen) according to manufacturer’s recommendations. The variant region was PCR amplified using GoTaq Green Master Mix (Promega; Madison, Wisconsin, USA) and primers that flank the area: Forward: 5’- CAGGGACAGGAGTCATCCAC-3’; Reverse: 5’-AACACCCATTAGACAGTGGGC-3’ (Integrated DNA Technologies; Coralville, Iowa, USA). The PCR product was purified using a QIAquick PCR Purification Kit (Qiagen) and sequenced using the same primers.

### Cell isolation, culture, and immortalization of lymphoblastoid cell lines (LCLs)

Peripheral blood mononuclear cells (PBMCs) were isolated by standard Ficoll-Paque (GE Healthcare, Chicago, Illinois, USA) density centrifugation from four discrete blood draws and time points from the patient over a span of two years, one time point from four heterozygous family members, and from eight unique age-matched healthy controls. PBMCs were cultured in complete RPMI-1640 (GE Healthcare) supplemented with 10% heat inactivated FBS (Gibco, Life Technologies; Rockville, Maryland, USA), 2mM _L_-glutamine (HyClone, Thermo Fisher Scientific; Waltham, Massachusetts, United States), and 1mM sodium pyruvate (Gibco, Life Technologies; Carlsbad, California, USA) in the presence or absence of various stimuli described in the following sections.

Lymphoblastoid cell lines (LCLs) were derived by EBV transformation and cultured in complete RPMI-1640 as previously described by our group^35, 36^. Briefly, PBMCs from the patient (II-1), one heterozygous control (II-2), and one healthy control individual were cultured in complete RPM1-1640 media (GE Healthcare) supplemented with tacrolimus (AG Scientific, San Diego, California) for 1 h. Cells were then infected with supernatant from a viral replication-permissive marmoset cell line B95-8 (ATCC, Manassas, Virginia, USA) until sufficient B cell blasts were generated. LCLs were cultured in complete RPMI-1640 media without tacrolimus.

Chondrocytes were grown from an osteochondroma that was surgically removed from the patient’s proximal tibia. The small cartilage component was excised from connective tissue and placed in separate wells in a 6-well plate and digested with 200 U/ml collagenase in complete Dulbecco’s Modified Eagle Medium (DMEM; GE Healthcare) supplemented with 10% heat inactivated FBS (Gibco, Life Technologies, Rockville, Md), 2mM L-glutamine (HyClone, Thermo Fisher Scientific), and 1mM sodium pyruvate (Gibco, Life Technologies) and 1x Antibiotic-Antimycotic (Gibco, Thermo Fisher) for 1 day at 37°C. Cells were filtered with a 40μ and allowed to adhere.

### Stable expression of NFAT1 using a Lentivirus vector

Stable expression of NFAT1 was established as previously described^37, 38^. Briefly, the NFATC2 (NM_173091) gene was cloned out of a Myc-DDK-tagged NFATC2 plasmid (Cat#: RC213966, OriGene Technologies; Rockville, Maryland, USA) and cloned into a GFP-tagged Lenti vector (Cat#: PS100071, OriGene Technologies) using EcoRI-HF (Cat#: R3101) and NotI-HF (Cat#: R3189), both from New England BioLabs (Ipswich, Massachusetts, USA). GFP-tagged NFATC2 and GFP-expressing empty vector (EV) were packaged using 3rd generation packaging plasmids, and transfected into HEK 293T cells. Culture media was collected, centrifuged, filtered, concentrated, and stored at −80°C before use.

To establish patient-derived chondrocytes that stably express WT NFAT1, chondrocytes were infected with EV or WT *NFATC2* lentiviral particles and 5ug/ml polybrene (Sigma-Aldrich, St Louis, Mo), cultured, and expanded in high glucose DMEM (HyClone) supplemented with 10% FBS and 2mM _L_-glutamine. Expanded cells were sorted on GFP expression using a BD FACS Aria (BD Biosciences) cell sorter. To establish patient T cells that express WT NFAT1, isolated PBMCs were stimulated with CD3/CD28 T cell activator (Cat#: 10971; Stemcell, Vancouver, Canada) for 3 days, infected with EV or WT *NFATC2* particles for 24 hours at 37°C, and stimulated with CD3/CD28 T cell activator for another 3 days before undergoing downstream flow cytometric analyses.

### Cytometric Bead Array (CBA)

A cytokine bead array was used to measure cytokine production in chondrocytes as previously described^39^. Briefly, patient and WT-*NFATC2*-transduced chondrocytes were seeded in T25 flasks in complete DMEM media (GE Healthcare) without antibiotics and allowed to reach confluency. Chondrocytes were washed and subsequently left untreated in media alone or stimulated with 50ng/mL phorbol 12-myristate 13-acetate (PMA) (Sigma) and 1μM ionomycin (Sigma) (P/I) or 20ng/mL IL-1β (Cat#: 14-8018-62; eBiosciences, San Diego, California, USA) at 37°C for 24h. Supernatants were collected and a human inflammatory cytokine CBA kit was used to measure the concentration of IL-6 (BD Biosciences, Cat#: 552932; Franklin Lakes, New Jersey, USA) according to manufacturer’s recommendations^40^. Samples were acquired using a BD LSRII flow cytometer and analyzed using FlowJo (BD Biosciences).

### Immunoblotting

Changes in NFAT1 expression and phosphorylation status was detected by stimulating LCLs with 10μM ionomycin for 10 min in complete RPMI or empty vector (EV)- or WT-*NFATC2*-transduced chondrocytes with P/I or 20ng/mL IL-1β for 15 min in complete DMEM at 37°C. Whole cell lysates were prepared by lysing untreated and stimulated LCLs or chondrocytes in a modified radio immunoprecipitation assay buffer with the Halt protease and phosphatase inhibitors cocktail (Thermo Fisher Scientific). Lysates were separated by 10% SDS-PAGE and transferred onto polyvinylidene difluoride membranes (Immobilon-FL; MilliporeSigma, Billerica, Massachusetts, USA). Membranes were blocked with 5% BSA in Tris-buffered saline with Tween-20, incubated with primary antibodies for 18h at 4°C, incubated with secondary antibodies for 1h at room temperature, and imaged using a LI-COR Odyssey infrared scanner (LI-COR Biosciences, Lincoln, Nebraska, USA). The primary antibodies used were the following: NFAT1 (Cat#: 4389) and β-actin (Cat#: 3700) both from Cell Signaling Technologies (Danvers, Massachusetts, USA). The secondary antibodies used were the following: goat anti-rabbit IgG DyLight 800 conjugated (611-145-002-0.5; Rockland Immunochemicals, Pottstown, Pennsylvania, USA) and goat anti-mouse IgG IRDye 680RD (926-6870; LI-COR).

### Quantification of *NFATC2* transcript abundance by quantitative PCR (qPCR)

qPCR was conducted as previously described^35^. Briefly, total RNA was extracted from unstimulated or 18h ionomycin-stimulated LCLs using a RNeasy Plus Mini Kit (Qiagen) and converted to cDNA using an iScript cDNA synthesis kit (BioRad Laboratories; Hercules, California, USA). Transcript abundance was measured using a Universal SYBR Green Super Mix (Bio-Rad) and a 7300 Real-Time PCR System (Applied Biosystems, Foster City, California, USA). The primers used for *NFATC2* were the following: Forward: 5’-GTCAGCCTCAGCACTTTACC-3’; Reverse: 5’- ATCAGAGTGGGGTCATATTCATC -3’ (PrimeTime; Integrated DNA Technologies). Relative transcript abundance was quantified relative to *ACTB* using the 2^-ΔΔ^ method^41^.

### RNA-Sequencing

To investigate the global transcriptome, EV- or WT-*NFATC2*-transduced chondrocytes were unstimulated, 4h P/I-stimulated, or 4h 20ng/mL IL-1β-simulated. RNA was extracted in triplicate as previously described^35^ using a RNeasy Mini Plus Kit (Qiagen) according to manufacturer’s recommendations. RNA was prepared following the standard protocol for the NEBNext Ultra II Stranded mRNA (New England Biolabs) and sequenced on the Illumina NextSeq 500 with Paired End 42 bp × 42 bp reads. De-multiplexed read sequences were aligned to a reference sequence using Spliced Transcripts Alignment to a Reference (STAR) aligner. E1 Assembly and expression were estimated using Cufflinks E2 through bioinformatics apps on Illumina BaseSpace.

Expression data were normalized to reads between samples using the edgeR package in R (R Foundation, Vienna, Austria). Normalized counts were filtered to remove low counts using the filterByExpr function in edgeR^42^. Batch correction and differential expression was conducted using Limma^43^. Pathway analysis was done by first performing Gene Set Enrichment Analysis (GSEA) with 1000 permutations using the Molecular Signatures Database Hallmark module. Signal-to-noise ratio was used for gene ranking and the obtained NESs and P-values were further adjusted using the Benjamini-Hochberg method. Pathways with an adjusted P-value < 0.05 were considered significant. Leading edge genes from significant pathways between EV and WT-*NFATC2*-transduced chondrocytes were identified. Expression levels of these genes were then determined in each group.

Sample level enrichment analyses (SLEA) scores were computed as previously described^44^. Briefly, z-scores were computed for gene sets of interest for each sample. The mean expression levels of significant genes were compared to the expression of 1000 random gene sets of the same size. The difference between observed and expected mean expression was then calculated and represented on heatmaps generated using Morpheus (https://software.broadinstitute.org/morpheus). Raw data are deposited on Gene Expression Omnibus at the following accession number GSE193414.

### Single Cell RNA-Sequencing

Targeted single cell RNA-Sequencing of 500 curated immune-related genes was conducted on PBMCs from the patient, three heterozygous controls, and two age-matched healthy controls using the BD Rhapsody Single Cell platform, the Rhapsody Cartridge Reagent Kit (BD, Cat#: 633731), the BD Rhapsody Cartridge Kit (BD, Cat#: 633733), the Rhapsody cDNA Kit (BD, Cat#: 633733), the Rhapsody Targeted mRNA amplification Kit (Cat#: 633774), and the Human Single-Cell Multiplexing Kit (BD, Cat#: 633781) according to manufacturer’s recommendations. Briefly, thawed PBMCs were rested overnight and stimulated with P/I or left untreated for 4h. Cells from each donor were labelled with sample tags, washed with stain buffer and pooled together in cold sample buffer to obtain ∼25,000 cells in 620ul for each of the unstimulated and stimulated samples. Two nanowell cartridges were primed and subsequently loaded with the pooled samples for 15min at room temperature on the Rhapsody Express instrument (BD, Cat#: 633702). Cell capture beads were prepared and loaded onto the cartridge. The cartridge was washed, cells were lysed, and beads were retrieved. The sample then underwent reverse transcription and exonuclease I treatment. cDNA was amplified using a custom immune panel (**Supp. Table 5**) via PCR. mRNA PCR products were separated from sample tag products with double-sided size selection using AMPure beads (Beckman Coulter, Cat#: A63880). Separated products were further amplified by PCR and then purified using AMPure beads. Product quality was checked using Agilent DNA High Sensitivity Kit (Agilent Technologies, Cat#: 5067-4626) on the Agilent 2100 Bioanalyzer (Agilent Technologies, Cat#: G2940CA). PCR products were then PCR amplified using two index primers, one for unstimulated (5’-CGAGGCTG-3’) and one for stimulated (5’-AAGAGGCA-3’) samples. Indexed products were again purified using AMPure beads and sample quality was checked using the Agilent 2100 Bioanalyzer. Libraries were diluted to 650pM and multiplexed for paired-end (2 x 115bp) sequencing on a NextSeq2000 Sequencing System (Illumina) at 200 cycles P3 for 1.1 billion reads.

FASTQ files were processed using the BD Rhapsody Targeted Analysis Pipeline and Seven Bridges (www.sevenbridges.com) according to manufacturer’s recommendations. Briefly, R1 and R2 reads were filtered based on read length, quality, and highest single nucleotide frequency. R1 reads are annotated to identify cell labels, unique molecular identifiers (UMI), and filtered for >6 Poly(T) tail. Bowtie2 version 2.2.9 was used to map the R2 reads to the reference sequences. All reads, R1 and R2, with the same cell label, UMI sequence, and gene were collapsed into a single molecule. Recursive substation error correction (RSEC) and distribution-based error correction (DBEC) were used to adjust for PCR errors in UMI sequences (which can give rise to artifacts). Putative cells were determined by considering all reads associated with DBEC-adjusted molecules. The minimum second derivative along the cumulative reads curve is identified as the inflexion point, after which the cells are labelled as noise. Filtered putative reads that align to a sample tag sequence were used to identify sample labels for a cell. Finally, labelled DBEC-adjusted molecule counts were obtained in a CSV format for downstream analyses.

The R package Seurat^45^ was utilized for all downstream analysis. No gene per cell cutoffs were imposed since we were using a targeted 500-gene panel. Scaling and clustering was performed on each pool of samples independently. Dimensionality reduction using PCA was based on all 500 genes and UMAP was based on the first 20 PCs. Cell identities were first annotated with SingleR using fine labeling from the Novershtern hematopoietic dataset^46, 47^. The annotation was refined manually based on the UMAP clustering patterns, grouping the SingleR labels into 8 main populations: Naïve B cells, Memory B cells, Naïve CD4^+^, Memory CD4^+^, Memory CD8^+^, Monocytes, NK cells, and undefined. For differential gene expression analyses, we utilized the Seurat implementation of negative binomial test, assuming an underlying negative binomial distribution in RNA-Seq data while leveraging the UMI counts to remove technical noise^48, 49^. Raw data are deposited on Gene Expression Omnibus at the following accession number GSE193410. All downstream analysis scripts used in analysis are available on GitHub (https://github.com/maggie-fu/NFATC2_RNAseq/).

### Lactate Dehydrogenase (LDH) Assay

Chondrocyte resistance to cell death was quantified using a lactate dehydrogenase assay (LDH) according to manufacturer’s instructions (Promega, Cat#: G1780). Briefly, EV- or WT-*NFATC2*-transduced chondrocytes were seeded in 96-well plates at a concentration of 5000 cells/well. Cells were rested until they reached confluency and subsequently stimulated with P/I over a time course, including 0, 2, 4, 6, 8, 10, and 12 days at 37°C. 50 μl of cell supernatants were collected, added to 50μl of CytoTox96 reagent, and incubated in the dark for 30 min at room temperature. The reaction was subsequently stopped with 50μl of stop solution. Maximum LDH release was measured at each time point by lysing cells. Absorbance was measured on the Infinite M200 plate reader (Tecan; Männedorf, Switzerland) at 492nm. Cell death was defined as a percentage of maximum LDH release.

### B cell expansion and differentiation

Naïve B cells were isolated from PBMCs of the patient, one heterozygous family member and 6 healthy controls using the EasySep Human Naïve B Cell Isolation Kit (Cat#: 17254; Stemcell) according to manufacturer’s recommendations. Cells were stained with the ‘Plasmablast Differentiation’ panel (**Supp. Table 6**) described in the ‘Flow Cytometry’ section before and after stimulation with ImmunoCult-ACF Human B Cell Expansion Supplement (Cat#: 10974; Stemcell) for 6 days in order to determine the proportion of different subsets by flow cytometry.

### Clinical-Grade Flow Cytometry

Patient clinical immunophenotyping was also carried out at an accredited clinical flow cytometry laboratory. Whole blood was collected and stained using DURAClone tubes (Beckman Coulter; Brea, California, USA) according to the manufacturer’s recommendation. Briefly, fresh blood was incubated with dried reagent tubes and incubated in the dark at room temperature for 15 min. Cells were washed in 3mL of PBS 2 times, before being fixed in 500uL of PBS containing 0.1% formaldehyde and subsequently acquired. Panels focused on the analysis of T cell subsets (DURAClone IM T Cell Subsets, Beckman Coulter; Cat#: B53328) and B cell subset (DURAClone IM B Cell, Beckman Coulter; Cat#: B53318). All flow data were acquired on a Navios flow cytometer and then analyzed using Kaluza software (Beckman Coulter).

### Research-Grade Flow Cytometry

To carry out immunophenotyping and intracellular cytokine detection, PBMCs from the patient, heterozygous family members, and age-matched controls were stimulated with P/I for 4h at 37°C in the presence of GolgiStop (Cat# 554724, BD Biosciences). Following stimulation, cells were stained with a cocktail of antibodies against surface markers for 20 min on ice then fixed with the Foxp3 Fixation/Permeabilization working solution (Cat#: 00-5123-43; Invitrogen, Thermo Fischer Scientific) from the eBioscience Foxp3 Transcription Factor Staining Buffer Set (Cat# 00-5523-00, Invitrogen, Thermo Fisher Scientific) for 20 min. Fixed cells were then stained with antibodies directed against intracellular cytokines or transcription factors for 45 min in 1x Permeabilization Buffer (Cat#: 00-8333-56; Invitrogen, Thermo Fischer Scientific). Samples were then washed and analyzed on the BD FACSymphony flow cytometer (BD Biosciences). Data were analyzed using FlowJo software (BD Biosciences). Antibody panels used for staining are listed in **Supp. Table 6**.

### Histology

Formalin-fixed, paraffin-embedded lymph node tissue was sectioned at 4μM and subjected to routine hematoxylin and eosin (H&E) staining or immunohistochemistry (IHC). The following IHC antibodies were used: CD21 (IF8, Dako), CD20 (L26, Ventana), BCL6 (P6-B6P, Dako), CD30 (Ber-H2, Ventana), and BCL2 (124, Dako). CD21, BCL6, and BCL2 IHC were performed on the Dako Autostainer. CD20 and CD30 IHC were performed on the Dako Omnis Automated Slide Stainer.

## RESULTS

### Clinical features of complete human NFAT1 deficiency

The patient (designated II-1 on the family pedigree) is born to consanguineous parents. After a healthy birth, the patient started developing difficulty bending his knees with no associated swelling, pain, erythema, or morning stiffness in early childhood (ages 0-5 years). Insidiously, many joints developed a similar painless decreased range of motion causing difficulty with ambulation. Medical care was disjointed until the patient was in his teenage years (ages 15-20 years). Assessment then demonstrated a neurodevelopmentally normal young man with marked bilateral fixed flexion contractures of knees, hips, and ankles. Multiple upper limb joints also demonstrated reduced range of motion, including wrists, hands, and shoulders (**Fig. 1A**, **Table 1**). Muscle strength and deep tendon reflexes were all intact. The joint changes significantly impacted his gait, causing him to walk with an increased anterior pelvic tilt, and leading to decreased dynamic hip range of motion with particularly diminished extension through mid to late stance. He also had diminished dynamic knee range of motion with knee flexion throughout stance and swing and mild equinus bilaterally at his ankles (**Supp. Table 1**). He had absent distal digital creases and prominent fetal fat pads on all fingers. Echocardiography showed a mildly hypertrophied left ventricle with multiple cords and an appearance of coarse trabeculae on the septal and free wall surfaces. Dual-energy X-ray absorptiometry (DEXA) scanning revealed low total body bone mineral density (0.888 g/cm^2^, z-score for chronologic age being −1.9 [-0.4 after adjusting for height]). X-rays showed flattening of multiple lower thoracic and lumbar vertebral bodies suggesting mild vertebral compression fractures. Two osteochondromas/exostoses were also identified; one on the medial aspect of the right proximal tibial metaphysis (2cm) and a second on the anterior aspect of the right proximal fibula (3cm) (**Fig. 1A, 1B**). In summary, the main musculoskeletal findings are contractures of the large and small joints of the upper and lower limbs, osteochondromas, and osteopenia (**Table 1**).

**Fig. 1.**
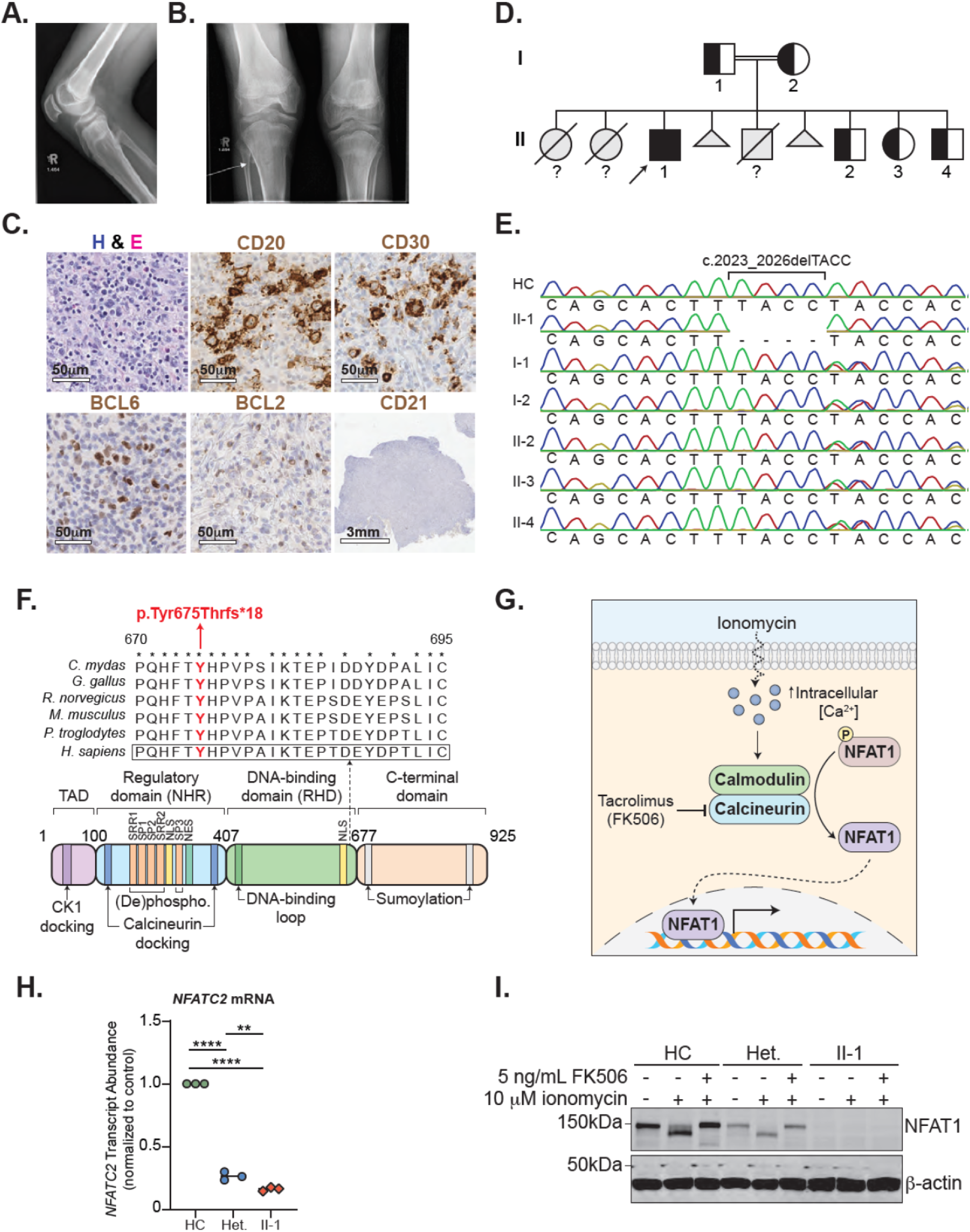
Clinical phenotype of a patient with NFAT1 deficiency, joint contractures, and B cell malignancy. **A)** X-ray demonstrating extent to which knees could be straightened documenting the fixed flexion deformity. **B)** Osteochondroma (indicated by the arrow) on the anterior aspect of the right proximal fibula. **C)** Hematoxylin and eosin (H&E) stain, and CD20, CD21, CD30, BCL2, and BCL6 immunostain of a patient lymph node biopsy. Scale bar for H&E, CD20, CD30, BCL6, and BCL2 is 50μm and for CD21 is 3mm. **D)** Family pedigree. Half-filled symbols=heterozygous unaffected individuals, filled symbols=homozygous affected individual, arrow=proband, question marks=ungenotyped. **E)** Sanger sequencing of DNA extracted from whole blood of the patient, family members, and a healthy control. Site of four base pair deletion is indicated. **F)** Schematic illustrating the protein domains of NFAT1. Location of variant shown in red. Affected region was aligned to other species. Asterisks indicate full conservation. **G)** Schematic illustrating ionomycin-induced activation of calcineurin and NFAT1. **H)** *NFATC2* transcript abundance relative to β-actin (*ACTB*) in the patient (II-1), a heterozygous control (II-2), and a healthy control (HC) determined by qPCR. Red circles=II-1, blue circles=II-2, green circles=HC. ** p<0.01, ****p<0.0001. One-way ANOVA and Tukey’s post-hoc test. **I)** Immunoblot of patient (II-1), heterozygous control (II-2), and control-derived LCLs using an N-terminal NFAT1 antibody before and after 10 min ionomycin stimulation with or without FK506 treatment. N = 3.

**Table 1:** Major Clinical Features. Patient clinical features organized by musculoskeletal or immunological/hematological abnormalities.

| Musculoskeletal | Immunological/Hematological |
| --- | --- |
| Contractures of upper limb joints, hips, knees, ankles | B-cell lymphoma, unclassifiable, with features intermediate between DLBCL and cHL |
| Mobility issues: stiff gait, decreased stride length, toe walking | Widespread lymphadenopathy: abdominal, inguinal, hepatic, splenic, paratracheal, subcarinal, bone. No bone marrow involvement. |
| Decreased arm swing and knee range of motion. Anterior pelvic tilt with adduction of thighs | Lymph node tissue demonstrates malignant, large, mono- or binucleated cells within a benign background inflammatory cell infiltrate composed of small lymphocytes, eosinophils, histiocytes, and neutrophils |
| Ill-defined sclerotic bands present in the metaphyses of both the femoral tibia and fibula | Lymph node: effaced/abnormal architecture, large atypical CD79a<sup>+</sup>CD30<sup>+</sup>CD20<sup>+</sup>CD15<sup>-</sup>PAX5<sup>+</sup> B cells, positive BCL6 rearrangement, negative BCL2 rearrangement |
| Presence of multiple small pedunculated osteochondromas |  |
| Developed severe spinal cord stenosis at level C3 C4, requiring posterior cervical decompression and fusion |  |
| Bone Density Scan (DEXA):<br><u>Lumbar spine L1-4:</u> 0.739 (g/cm<sup>2</sup>); Z-Score for chronologic age: -1.4; Z-score corrected for height: +0.2<br><u>Total left hip:</u> 0.865 (g/cm<sup>2</sup>); Z-Score for chronological age 1.0; Z-score corrected for height: +0.2<br><u>Total body:</u> 0.888 (g/cm<sup>2</sup>); Z-Score for chronologic age: -1.9; Z-score corrected for height: -0.4 |  |
| No evidence of muscle necrosis or regeneration, no increased endomysia fibrous tissue, no inflammation, no atypical cells |  |

In his late teens (ages 15-20 years), the patient presented with abdominal pain, fever, night sweats and weight loss (i.e. B symptoms) and was ultimately diagnosed B-cell lymphoma, unclassifiable, with features intermediate between diffuse large B-cell lymphoma (DLBCL) and classic Hodgkin lymphoma (cHL). Lymphoid tissue biopsy showed effacement of typical node architecture and the absence of clear germinal centers and follicular structure. There was a prominent population of larger atypical lymphocytes (CD79a^+^CD30^+^CD20^+^CD15^-^BCL6^+^) in the background of mature CD4^+^ and CD8^+^ T cells (**Fig. 1C**). He was staged as Ann Arbor stage 4B with widespread disease, including abdominal, inguinal, hepatic, splenic, paratracheal, and subcarinal lymphadenopathy. The patient was treated with 6 cycles of a modified dose-adjusted EPOCH-R regimen without vincristine and went into full remission.

### Whole-exome sequencing (WES) reveals a novel damaging homozygous variant in *NFATC2*

Given the unique phenotype of the patient, consanguinity, and significant family history, we performed whole exome sequencing (WES) to identify a potential genetic etiology. The patient was found to carry a previously unreported homozygous 4-base pair deletion in exon 8 of the gene *nuclear factor of activated T cells 2* (*NFATC2*) encoding the NFAT1 transcription factor. This variant causes a frameshift leading to a premature stop codon (NM_173091:c.2023_2026delTACC; NP_775114:p.Tyr675Thrfs*18). Sanger sequencing of the variant region in whole blood confirmed that it segregated with disease within the patient family (**Fig. 1D, 1E**). The variant localizes to the DNA binding domain of the NFAT1 protein and affects an evolutionarily conserved region (**Fig. 1F**).

This variant led to significantly reduced *NFATC2* transcript abundance in patient-derived lymphoblastoid cell lines (LCLs) when compared to heterozygous control- and unrelated control-derived LCLs (**Fig. 1G**). To investigate the impact on NFAT1 protein function, we measured NFAT1 expression in LCLs pre- and post-stimulation with the NFAT1 activator, ionomycin (**Fig. 1H**), and in the absence or presence of the calcineurin inhibitor tacrolimus (FK506). We confirmed that NFAT1 expression was undetectable in patient LCLs, but reduced in heterozygous control LCLs (**Fig. 1I**). Furthermore, NFAT1 was strongly dephosphorylated in both wild-type and heterozygous control LCLs following ionomycin stimulation and this effect was effectively inhibited by FK506 treatment (**Fig. 1G, 1I**).

### NFAT1-deficient patient chondrocytes are more enriched in cell proliferation and inflammatory genes with elevated IL-6 production and resistance to cell death

Informed by the patient’s joint contractures and osteochondromas, we assessed the impact of NFAT1 deficiency on chondrocytes isolated from the excised tibial osteochondroma. Patient chondrocytes had undetectable NFAT1 expression (**Fig. 2A**). To study the impact of NFAT1 deficiency on chondrocytes, we engineered a *WT NFATC2* lentivirus construct and transduced patient chondrocytes to rescue NFAT1 expression. This restored function to the pathway as demonstrated by NFAT1 dephosphorylation that was sensitive to FK506 inhibition (**Fig. 2A**). Further, treatment with IL-1β, a key cytokine involved in cartilage damage in arthritis^50^, did not change NFAT1 phosphorylation status.

**Fig. 2.**
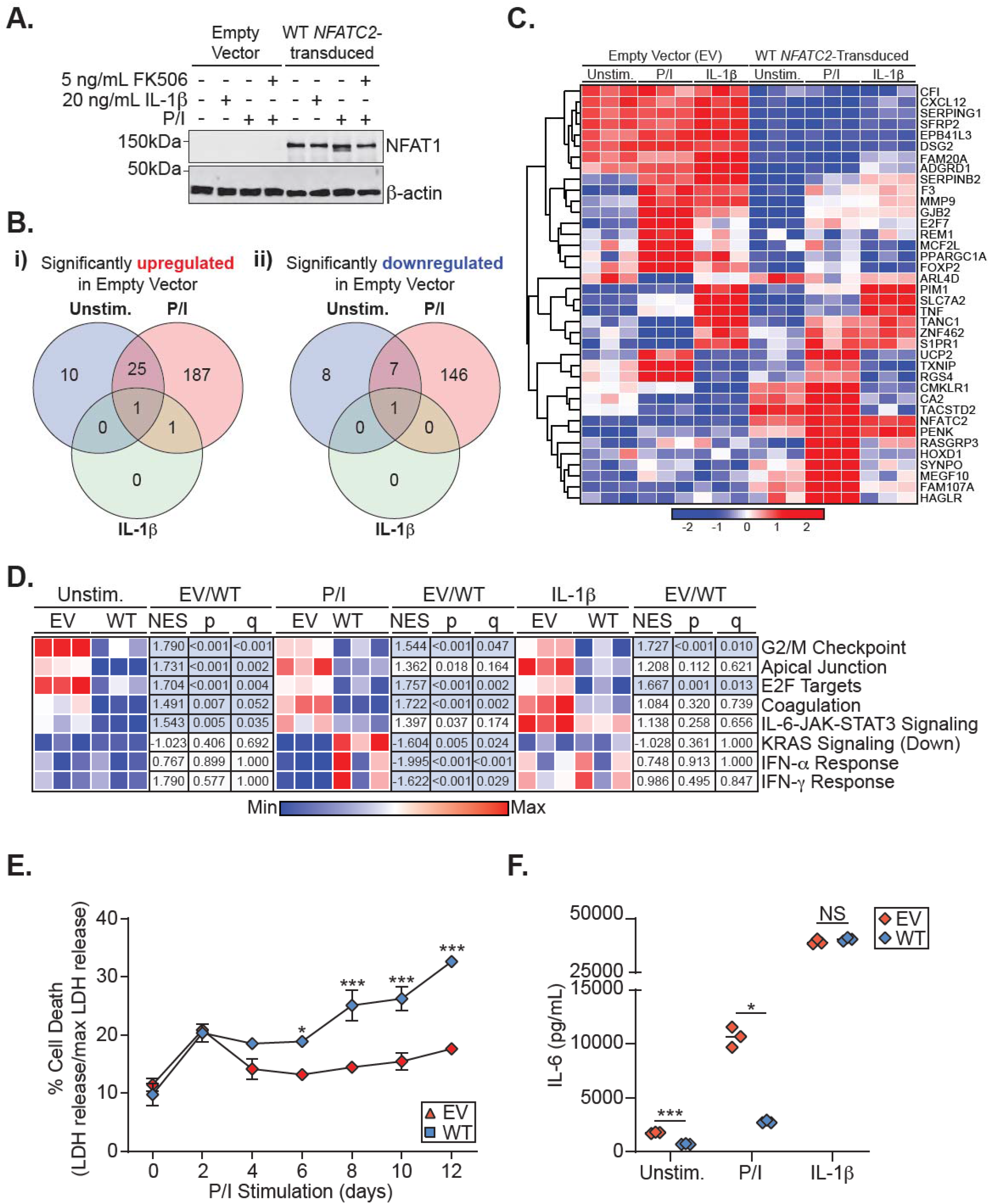
NFAT1 deficiency in chondrocytes leads to an increased pro-survival and proinflammatory phenotype. **A)** Immunoblot of empty vector (EV)- and WT *NFATC2*-transduced patient chondrocytes before and after 15min phorbol 12-myristate 13-acetate/ionomycin (P/I) stimulation, 20ng/mL IL-1β stimulation, or no treatment. n=3. **B-D)** RNA-Seq carried out on EV- or WT *NFATC2*-transduced patient chondrocytes unstimulated or stimulated 24h with P/I or 20ng/mL IL-1β. **B)** Venn diagram of significantly (FDR<0.05) **(i)** upregulated and **(ii)** downregulated genes in EV-transduced patient chondrocytes compared to WT *NFATC2*-transduced chondrocytes. **C)** Heatmap of top genes that meet a FDR<0.025 cutoff between EV and WT *NFATC2*-transduced patient chondrocytes. **D)** Heatmap of sample level enrichment scores (SLEA) scores with normalized enrichment scores (NES), p-val, and q-values for each pathway determined by gene set enrichment analysis (GSEA). Min and max refer to the row-normalized minimum and maximum for each pathway. **E)** Measurement of percent cell death in chondrocytes over 12 days by quantifying lactate dehydrogenase in supernatants. *p<0.05, ***p<0.001 Two-way ANOVA, Dunnett’s post-hoc test. **F)** ELISA detection of IL-6 production from the supernatants of chondrocytes in different conditions. *p<0.05, ***p<0.001 Mann-Whitney U-test.

To define the impact of NFAT1 deficiency on the global transcriptome of chondrocytes, we conducted bulk RNA-Seq on empty vector (EV) or WT *NFATC2*-transduced patient chondrocytes untreated or stimulated with P/I or IL-1β. We identified 187 significantly upregulated and 146 significantly downregulated genes between P/I-stimulated WT *NFATC2*- and EV-transduced chondrocytes, which may represent NFAT1 suppressed and enhanced genes, respectively (**Fig. 2B, 2C**). Gene set enrichment analysis (GSEA) identified significant enrichment in pathways involved with cellular growth and inflammation (**Fig. 2D**). Notable was the enrichment of cell proliferation pathways both at baseline and in response to P/I stimulation in NFAT1-deficient EV-transduced chondrocytes, including G2/M Checkpoint (unstimulated: NES=1.790, FDR<0.001; P/I stimulated: NES=1.544, FDR=0.047) and E2F targets (unstimulated: NES=1.704, FDR=0.004; stimulated: NES=1.667, FDR=0.013) (**Fig. 2D**). To assess the functional consequences of these transcriptomic findings, we stimulated EV- and WT *NFATC2*-transduced chondrocytes over a time course up to 12 days with P/I and monitored cell survival. Here, we discovered that EV-transduced NFAT1-deficient chondrocytes were significantly more resistant to cell death than WT *NFATC2*-transduced chondrocytes (**Fig. 2E**).

The IL-6-JAK-STAT3 signaling pathway was also significantly enriched in unstimulated EV-transduced NFAT1-deficient chondrocytes (NES=1.543, FDR=0.035) (**Fig. 2D**). Confirming this at the protein level, both unstimulated and P/I-stimulated EV-transduced NFAT1-deficient chondrocytes produced significantly more IL-6 than ‘rescued’ WT-transduced chondrocytes (**Fig. 2F**). Taken together, these findings suggest that NFAT1 regulates chondrocyte growth, death, and proinflammatory transcriptional programs, collectively leading to aberrant connective tissue homeostasis.

### NFAT1 deficient B cells are largely naïve *ex vivo* and express proliferation markers

Given that NFAT1 is expressed in most lymphocytes^16^, we conducted targeted single cell RNA-Sequencing (scRNA-Seq) on patient PBMCs alongside 5 healthy controls (2 age-matched sex-matched healthy controls and 3 heterozygous family members) stimulated with P/I to identify cell type-specific defects. Unsupervised clustering analysis revealed markedly reduced memory B cells and elevated memory CD4^+^ T cells in the patient PBMCs (**Fig. 3A**).

**Fig. 3.**
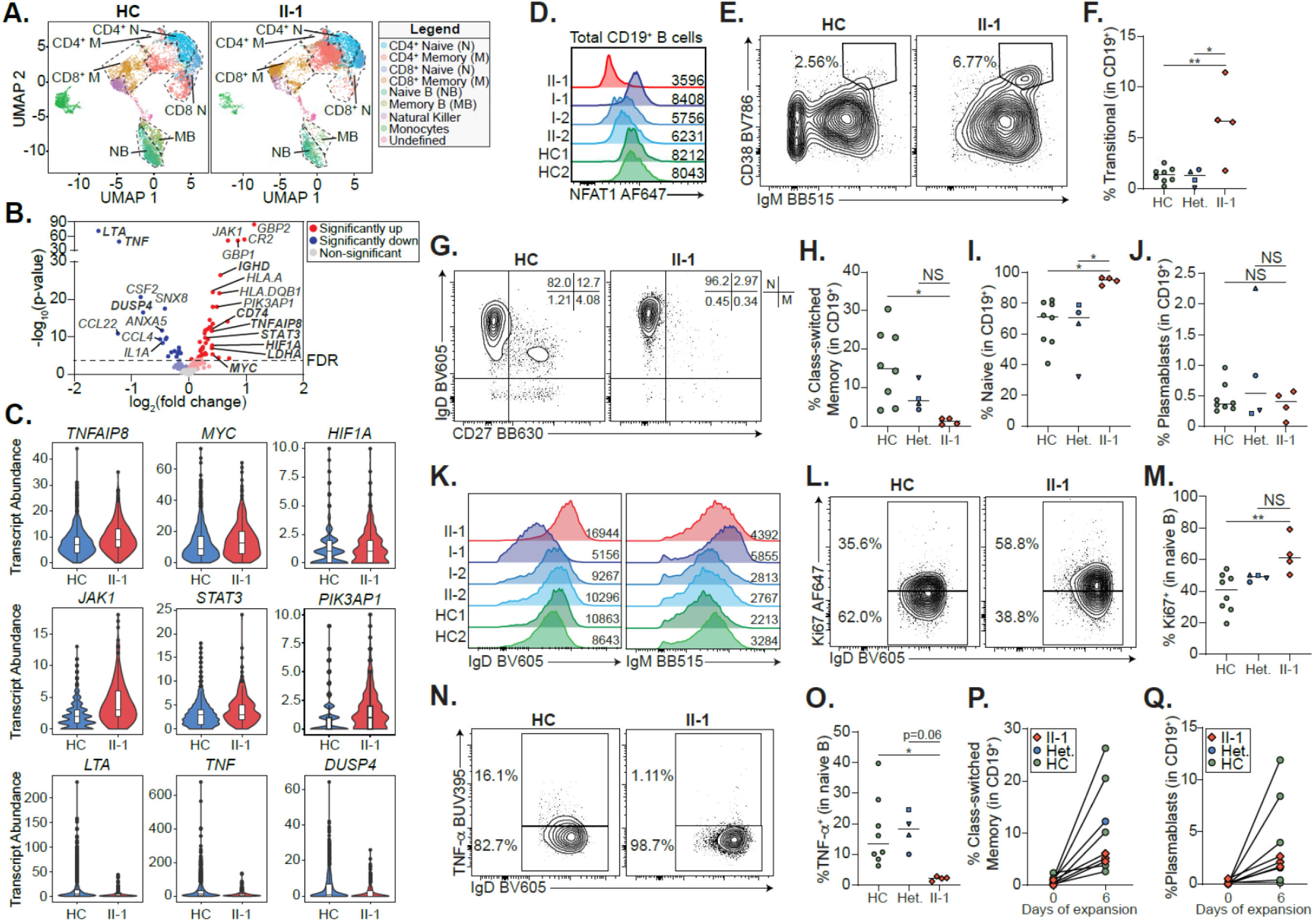
NFAT1 deficiency leads to accumulation of hyperproliferative naïve B cells. **A-C)** Single cell RNA-Sequencing of peripheral blood mononuclear cells from the patient (II-1), heterozygous controls (n=3), and a healthy control (n=2) stimulated with P/I for 4h. **A)** Uniform manifold approximation and projection (UMAP) visualization of stimulated cell subsets. Cell labels indicated on right. **B)** Volcano plot showing differential gene expression in naïve B cells between patient and 5 healthy controls. Dark colors=significance, dim colors=nominal significance but not after adjusted p-value, grey colors=not significant. **C)** Violin plots of significantly differentially expressed genes of interest in naïve B cells comparing patient and healthy controls. **D)** Expression of NFAT1 in patient and control naïve B cells. Mean fluorescence intensities (MFIs) are indicated. **E)** Frequency of IgM^++^CD38^++^ transitional B cells in the patient (II-1) and heterozygous (n=4) and healthy controls (n=8). **F)** Quantification of **E)**. **G)** Frequency of IgD^+^CD27^-^ naïve (N) and IgD^-^CD27^+^ memory (M) B cells in the patient and one representative control. Schematic of quadrants that correspond to each cell population and frequency shown top right. **H,I)** Quantification of **G). J)** Frequency of CD27^+^CD38^+^ plasmablasts in mature CD19^+^ B cells. **K)** IgD and IgM expression in each individual. MFIs are indicated. **l)** Frequency of Ki67^+^ naïve B cells in the patient and a representative control. **M)** Quantification of **l)**. **N)** Frequency of TNF-α^+^ naïve B cells in the patient and a representative control. **O)** Quantification of n). **P-Q)** Isolated naïve B cells from patient, heterozygous (n=1), and healthy controls (n=6) expanded for 6 days. **P)** Frequency of class-switched memory B cells and **Q)** plasmablasts before and after expansion. *p<0.05, **p<0.01. One-way ANOVA and Tukey’s posthoc test. Green circles=healthy control, blue circle=II-2, blue upright triangle=I-2, blue inverted triangle=I-1, blue square=II-3, red diamond=II-1.

Since patient B cells were largely naïve, we focused our differential gene expression (DE) analysis on patient and control naïve B cells (**Fig. 3B**). We discovered significantly increased transcript abundance of pro-survival, proliferation, and anti-apoptotic genes frequently associated with lymphoma in patient cells compared to controls. This includes *MYC*^51^, *HIF1A*^52^, *JAK1*, *STAT3*^53^, *TNFAIP8*^54^, *and PIK3AP1*^55^ (**Fig. 3C**). Consistent with the documented NFAT1 deficiency, classical NFAT1 targets such as *TNF* and *LTA*^56, 57^ were significantly decreased in patient naïve B cells.

To study this B cell phenotype further, we carried out multi-parameter flow cytometric analyses on PBMCs from the patient, 4 heterozygous healthy family member, and 8 healthy controls (see **Supp. Fig. 1A** for gating strategy). Direct detection of NFAT1 protein confirmed the lack of NFAT1 in patient B cells as compared to controls (**Fig. 3D**). Validating our scRNA-Seq data, and consistent with clinical-grade flow cytometry (**Supp. Table 2**), patient B cells showed signs of a developmental arrest, with significantly elevated transitional B cells (**Fig. 3E, 3F)** and naïve B cells (**Fig. 3G, 3I**) with a concurrent reduction in class-switched memory B cells (**Fig. 3G, 3H**). Interestingly, despite the reduced frequency of memory B cells in the patient, plasmablast numbers were comparable to controls (**Fig. 3J**) (**Supp. Fig. 1A**), perhaps explaining the patient’s normal serum immunoglobulin levels.

Expanding our naïve B cell analyses further, we observed markedly elevated IgD expression (**Fig. 3K**) (**Supp. Fig. 1B**) and generally comparable to control IgM expression (**Fig. 3K**) (**Supp. Fig. 1C**) in patient naïve B cells. These IgD^hi^ naïve B cells were characterized by elevated expression of the cell proliferation marker Ki67 (**Fig. 3L, 3M**), but significantly reduced TNF-α expression (**Fig. 3N, 3O**) which was consistent with the scRNA-Seq data.

Given the striking B cell differentiation defect in the patient, we next asked whether this was intrinsic or extrinsic to B cells. We isolated naïve B cells from the patient and controls and quantified the frequency of class-switched memory B cells and plasmablasts before and after treatment with an *in vitro* B cell activation and expansion cocktail. We demonstrated intact differentiation of both memory B cells (**Fig. 3P**) (**Supp. Fig. 1D**) and plasmablasts (**Fig. 3P, 3Q**), suggesting that the differentiation defect is likely to be extrinsic to B cells.

### NFAT1 deficient CD8^+^ T cells show impaired polyfunctionality

Since both NFAT1 deficiency and calcineurin inhibitor treatment are known to suppress CD8^+^ T cell effector function^58, 59^ and suppressed CD8^+^ T cells can create conditions that favour lymphomagenesis^60^, we next analyzed CD8^+^ T cell differentiation and function by scRNA-Seq and flow cytometry. DE analysis of patient memory CD8^+^ T cells revealed a mixed effector function defect, including decreased transcript abundance of *TNF*, *IFNG*, and *FASLG*, but increased *GZMH*, *GNLY*, and *PFN1* (**Fig. 4A, 4B**). Flow cytometric analysis of CD8^+^ T cell subsets revealed no differences in patient naïve and memory populations when compared to controls (**Fig. 4C, 4D**). However, whereas control CD8 populations expressed easily detectable levels of NFAT1 protein, patient CD8 populations were NFAT1-deficient (**Fig. 4E**). In line with the abnormal effector marker profile in scRNA-Seq, patient CD8^+^ memory T cells showed significantly impaired expression of TNF-α and IFN-γ expression in response to stimulation (**Fig. 4F-4H**). In contrast, IL-2 expression was intact in both patient naïve and memory CD8^+^ T cells (**Supp. Fig. 2A-D**). Patient CD8^+^ memory T cells also exhibited abnormal expression of activation markers, including comparable to control expression of CD40L (**Supp. Fig. 2E-F**) and PD-1 (**Fig. 4I**) (**Supp. Fig. 2H**), but elevated CD69 (**Fig. 4I**) (**Supp. Fig. 2G**). These findings indicate a pleiotropic function for NFAT1 in human CD8^+^ T cells.

**Fig. 4.**
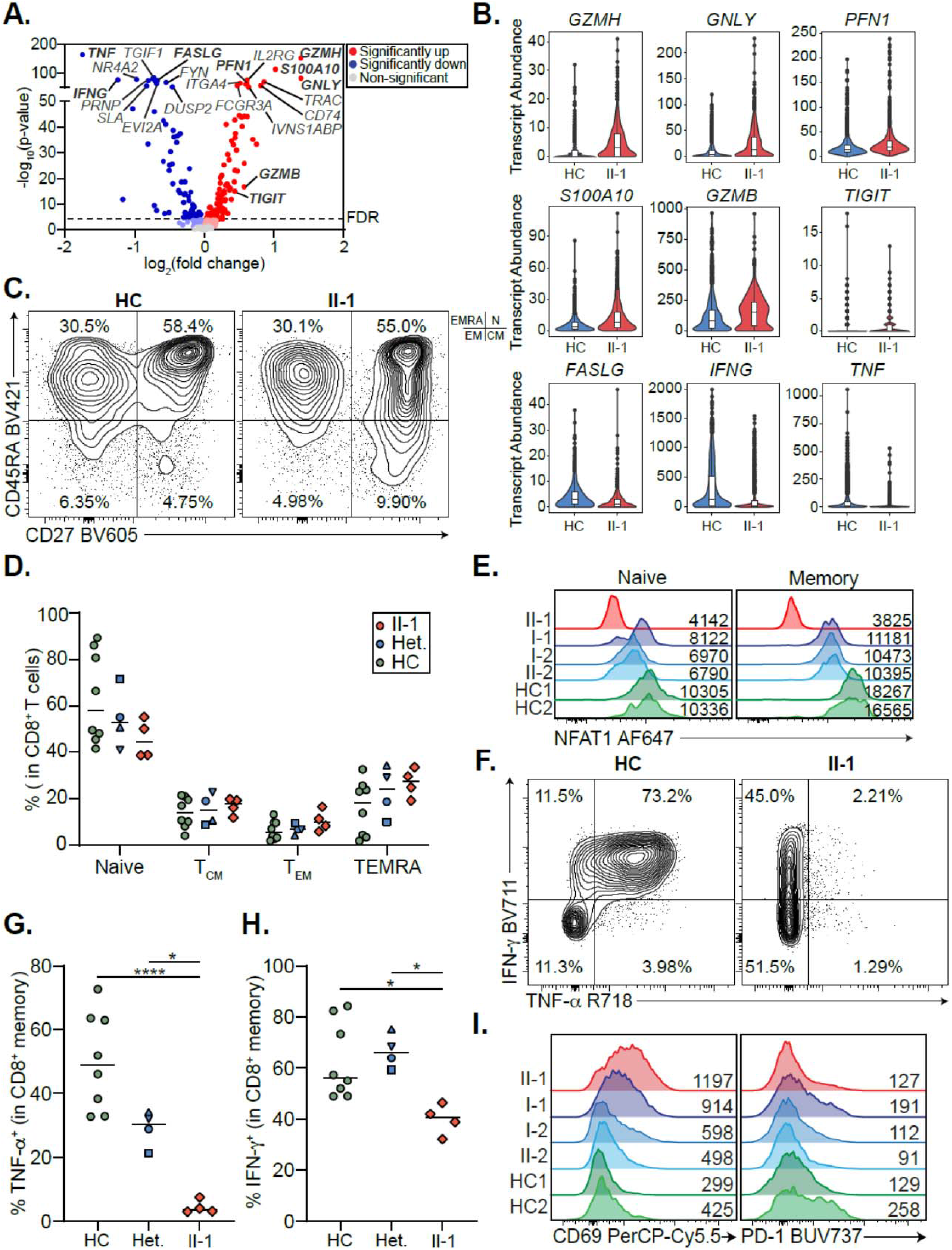
NFAT1 deficiency impairs memory CD8^+^ T cell function. **A)** Volcano plot showing differential gene expression in CD8^+^ memory T cells between patient and 5 healthy controls (3 heterozygous controls, and 2 healthy controls). **B)** Violin plots of significantly differentially expressed genes of interest in CD8^+^ memory T cells in patient and healthy controls. **C)** Frequency of CD8^+^ T cell subsets in the patient and a representative control, including CD45RA^+^CD27^-^ TEMRA, CD45RA^+^CD27^+^ Naïve (N), CD45RA^-^CD27^-^ effector memory (EM), CD45RA^-^CD27^+^ central memory (CM). Quadrants corresponding to each subset shown top right. **D)** Quantification of **C)**. **E)** Expression of NFAT1 in patient and control naïve and memory CD8^+^ T cells. Mean fluorescence intensities (MFIs) are indicated. **F)** TNF-α and IFN-γ expression in memory CD8^+^ T cells in the patient and a control. **G, H)** Quantification of **F)**. **I)** CD69 and PD-1 expression in patient and control memory CD8^+^ T cells after 4h of P/I stimulation. MFIs are indicated. *p<0.05, ****p<0.0001. One-way ANOVA and Tukey’s post-hoc test. Green circles=healthy control, blue circle=II-2, blue upright triangle=I-2, blue inverted triangle=I-1, blue square=II-3, red diamond=II-1.

### NFAT1 deficiency causes an accumulation of non-functional memory CD4^+^ T cells

The role of NFAT1 in CD4 cell functionality and differentiation is well characterized in mouse studies^14^. We sought to see if this held true in human complete NFAT1 deficiency. scRNA-Seq revealed a marked increase in the memory CD4^+^ T cell compartment (**Fig. 3A**). We conducted DE analysis on these cells and found significantly impaired expression of many makers for T helper function (i.e. *TNF* and *CD40LG*) and cell proliferation (i.e. *MYC*) (**Fig. 5A, 5B**). In addition, we observed increased expression of other activation/exhaustion-related genes such as *TIGIT*, *S100A10* and *TNFRSF4* (**Fig. 5A, 5B**). Interestingly, many regulatory components of the NF-ĸB pathway (*NFKBIZ*, *NFKBIA*, *REL*) and AP-1 pathway (*FOS* and *JUN*) showed impaired expression in NFAT1 deficient cells (**Fig. 5A, 5B**), highlighting the broad defects associated with human NFAT1 deficiency.

**Fig. 5.**
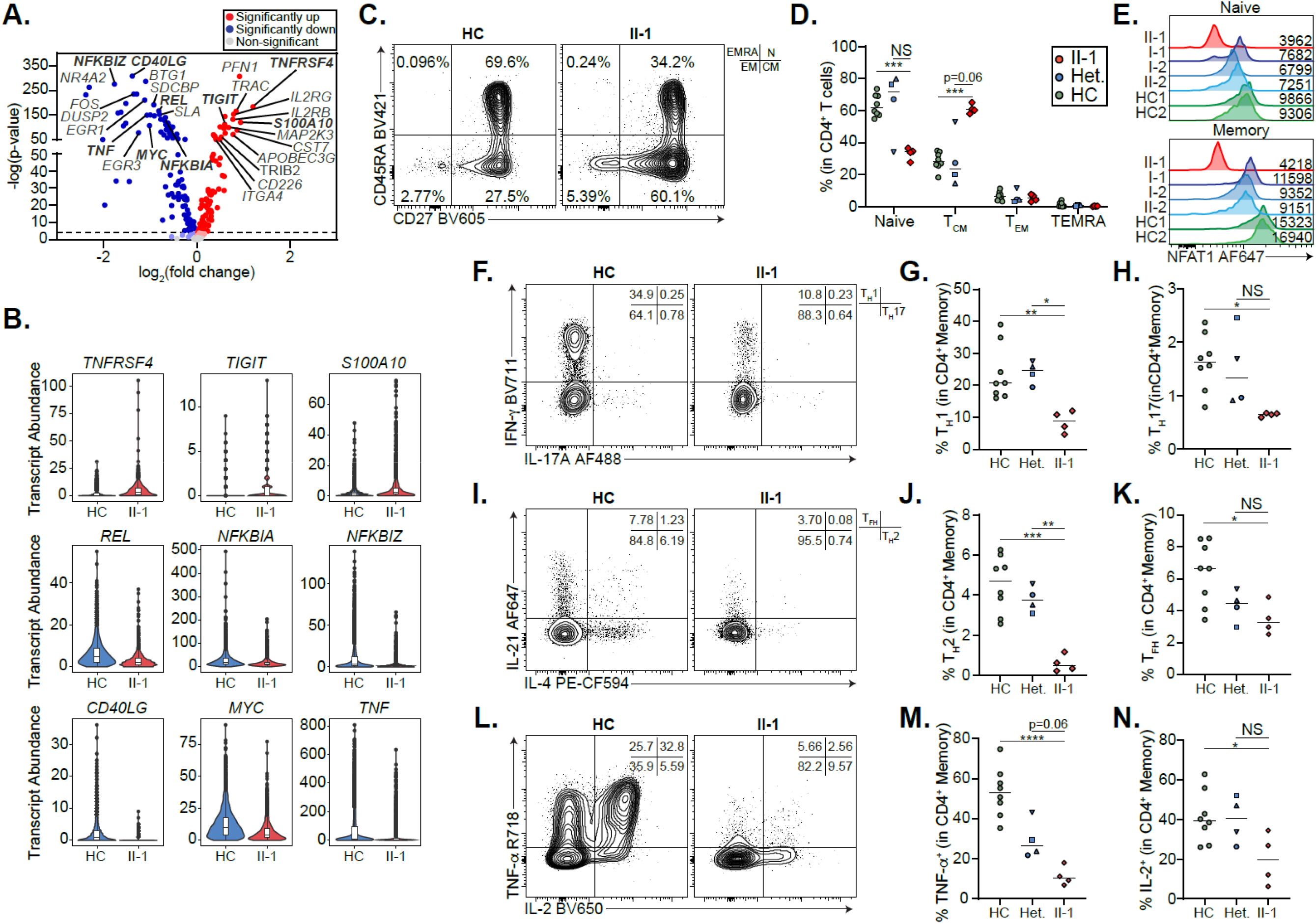
Patient CD4^+^ T cells are exhausted. **A)** Volcano plot showing differential gene expression in CD4^+^ memory T cells between patient and 5 healthy controls. **B)** Violin plots of significantly differentially expressed genes of interest in CD4^+^ memory T cells in patient and healthy controls. **C)** Frequency of CD8^+^ T cell subsets in the patient and a representative control, including CD45RA^+^CD27^-^ TEMRA, CD45RA^+^CD27^+^ Naïve (N), CD45RA^-^CD27^-^ effector memory (EM), CD45RA^-^CD27^+^ central memory (CM). Quadrants corresponding to each subset shown top right. **D)** Quantification of **C)**. **E)** Expression of NFAT1 in patient and control naïve and memory CD4^+^ T cells. Mean fluorescence intensities are indicated. **F)** Frequency of T_H_1 (IFN-γ IL-17A), T_H_17 (IL-17A IFN-γ) cells in memory CD4 T cells of the patient and a representative control. **G-H)** Quantification of **F)**. **I)** Frequency of T_H_2 (IL-4^+^IL-21^-^) and T_FH_ (IL-21^+^IL-4^-^) cells in memory CD4^+^ T cells of the patient and a representative control. **J-K)** Quantification of **I)**. **L)** Frequency of TNF- ^+^ and IL-2^+^ CD4^+^ memory T cells in the patient and a representative control. **M,N)** Quantification of **L)**. *p<0.05, **p<0.01, ***p<0.001, ****p<0.0001. One-way ANOVA and Tukey’s post-hoc test. Green circles=healthy control, blue circle=II-2, blue upright triangle=I-2, blue inverted triangle=I-1, blue square=II-3, red diamond=II-1.

We next examined the impact of NFAT1 deficiency on CD4^+^ T cell differentiation by flow cytometry. Confirming our scRNA-Seq data, the patient possessed decreased naïve CD4^+^ T cells with a concurrent increase in CD4^+^ central memory T cells (**Fig. 5C, 5D, Supp. Table 2**) and both these subsets had undetectable levels of NFAT1 protein, in contrast to controls (**Fig. 5E**). Within the memory CD4^+^ T cell compartment, the patient had significantly decreased T_H_1, T_H_2, T_H_17 and T_FH_ cells as measured by cytokine (i.e. IFN-γ, IL-4, IL-17A and IL-21) production (**Fig. 5F–5K**). Furthermore, classical NFAT1 targets IL-2 and TNF-α ^7^, were significantly reduced in patient memory CD4^+^ T cells (**Fig. 5L, 5M**). In contrast, naïve CD4^+^ T cells did not exhibit any significant differences in the expression of these same cytokines (**Supp. Fig. 3A-3I**).

### Human NFAT1 deficiency does not cause any overt defects in Tregs

Since NFAT1 was shown to be critical for controlling Treg suppressive function in mouse models^61^, we studied whether Treg development and/or function was affected in NFAT1 deficiency. Surprisingly, we found normal frequencies of Tregs and their naïve and activated subsets (**Supp. Fig. 5A-5C**). From a functional standpoint, activated patient Tregs showed significantly reduced TNF-α (**Supp. Fig. 5D-5E**), but normal IFN-γ (**Supp. Fig. 5D-5E**) and IL-2 (**Supp. Fig. 5G-5H**) expression. Similarly, Treg markers of stability and activation were largely normal, including FOXP3, Helios, CD25, CD69, CTLA4, and ICOS (**Supp. Fig. 5I-5N**), except for PD-1, which was significantly elevated (**Supp. Fig. 5O**).

### NFAT1 deficiency impairs T_FH_ function

Given the impaired production of a variety of T cell cytokines (**Fig. 5F-5N**) by CD4^+^ memory T cells, we asked whether the expression of activation markers and critical T cell coreceptors by this T cell population is affected by NFAT1 deficiency. Here we found significantly increased CD69, PD-1, and ICOS, but decreased CD25 and CD40L after P/I treatment (**Fig. 6A**) (**Supp. Fig. 3P-3T**). A similar profile was observed in naïve CD4^+^ T cells (**Supp. Fig. 3K-5O**).

**Fig. 6.**
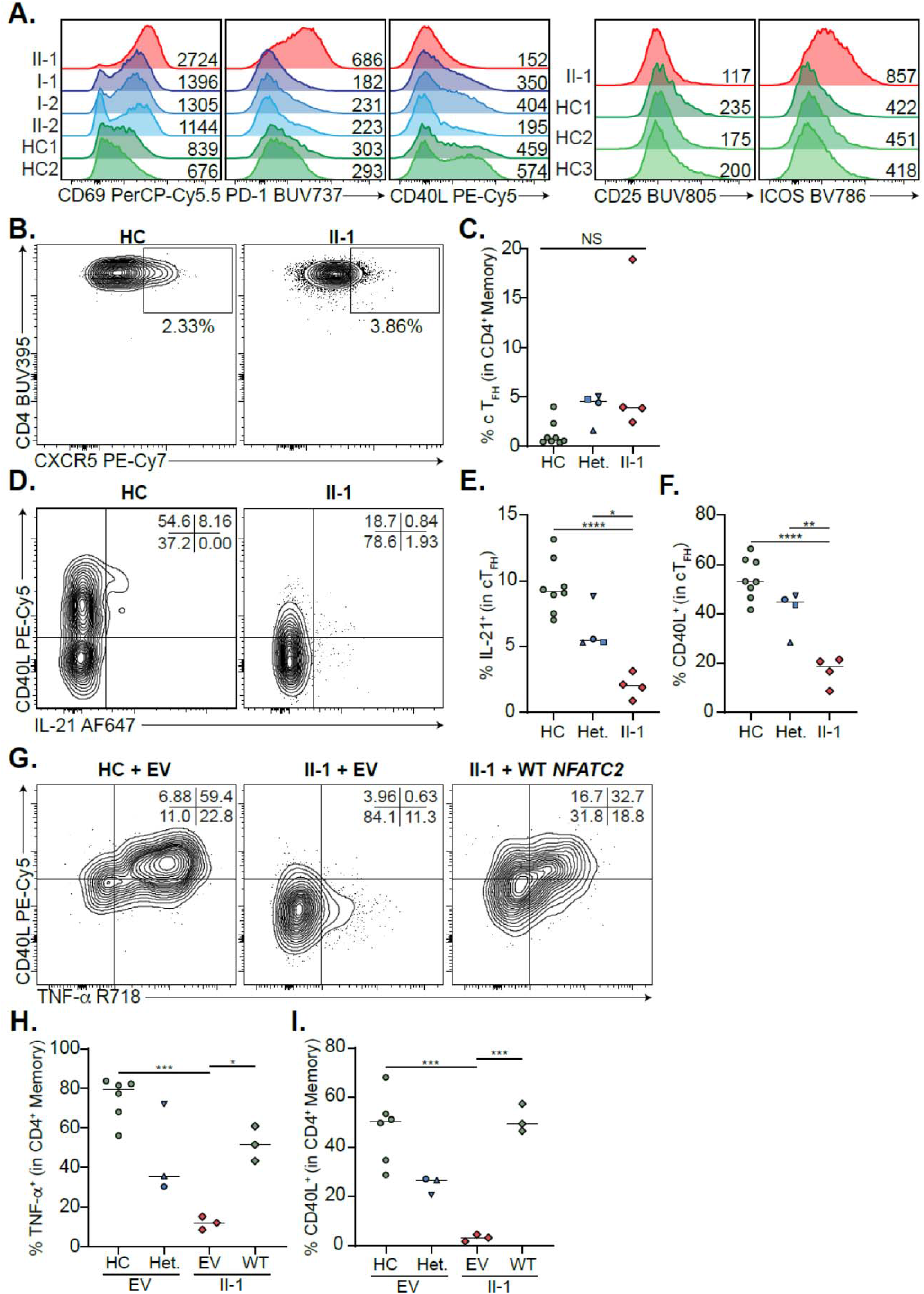
NFAT1 deficiency impairs T_FH_ function. **A)** Expression of activation markers in CD4^+^ memory T cells after P/I treatment, including CD69, PD-1, CD40L, CD25 and ICOS. Mean fluorescence intensities (MFIs) are indicated. **B)** Frequency of circulating T_FH_ (cT_FH_) cells in PD-1^+^ memory CD4^+^ T cells in the patient and a representative control. **C)** Quantification of **B)**. **D)** CD40L and IL-21 expression in patient and control cT_FH_ cells after 4h P/I stimulation. **E,F)** Quantification of **D)**. **G)** Frequency of CD40L^+^ and TNF-α^+^ GFP^+^ CD4^+^ patient or control memory T cells after 4h P/I stimulation of EV or WT *NFATC2* transduction. **H,I)** Quantification of **G)**. *p<0.05, **p<0.01, ***p<0.001, ****p<0.0001. One-way ANOVA and Tukey’s post-hoc test. Green circles=healthy control, blue circle=II-2, blue upright triangle=I-2, blue inverted triangle=I-1, blue square=II-3, red diamond=II-1.

Reduced CD4^+^ memory T cell activation marker expression and cytokine secretion paired with an extrinsic B cell differentiation defect prompted us to assess frequency and function of circulating T_FH_ cells in the patient (**Supp. Fig. 4A**). Confirming cytokine staining (**Fig. 5F-5K**), T_FH_ numbers were comparable to controls (**Fig. 6B, 6C**). Despite this, the expression of CD40L and IL-21, both important for B cell activation and differentiation^62^, was markedly impaired (**Fig. 6D-6F**). Importantly, these defects were reversed when we transduced patient CD4^+^ T cells with WT *NFATC2*, which restored CD40L and TNF-α expression to a level that was comparable to healthy controls (**Fig. 6G-6I**).

## DISCUSSION

Here we present a comprehensive clinical, immunological, biochemical, and transcriptional workup of the first reported case of human complete NFAT1 deficiency in a young man with a clinical triad of: progressive joint contractures, osteochondromas, and B cell malignancy. Based on our study, patients should be worked up for possible NFAT1 deficiency if they present with this triad of features. Although the full phenotype of human NFAT1 deficiency will only be appreciated when more affected individuals are identified, other clinical flags include: i) impaired B cell development (increased naïve, decreased memory); ii) normal CD8^+^ T cell development but impaired TNF-α and IFN-γ production and elevated CD69 expression; and iii) increased CD4^+^ memory proportions with reduced T_H_ subsets and cytokine secretion, and increased CD69, PD-1, ICOS expression. Notably, haploinsufficiency (at least with this particular null variant) was not observed as all heterozygous family members were healthy and unaffected.

Studying *Nfatc2^-/-^* mice has been extremely informative in defining the physiological functions of NFAT1. This includes regulation of chondrogenesis^18^ and bone development^19, 63^, maintenance of steady state hematopoiesis^63^, suppression and regulation of T cells^64, 65^, and B cell lymphomagenesis^34^. Reassuringly, human NFAT1 deficiency recapitulates a variety of the features seen in the knockout mice (**Table 2**). The joint contractures and progressive difficulty in ambulation observed in our NFAT1-deficient patient were accurately predicted by *Nfatc2*^-/-^ mice, as was the with excess differentiation, proliferation, and endochondral ossification affecting cartilage cells^18^. The pro-growth, pro-survival, and proinflammatory phenotype of patient chondrocytes (**Fig. 2**) suggests that human NFAT1 serves as a repressor of cartilage cell growth and differentiation. Future work will focus on studying how osteopenia arises in the context of human NFAT1 deficiency and the contributions of different mesenchymal cells to the patient’s musculoskeletal phenotype. Interestingly, bone fractures and osteopenia have also been observed in *Nfatc2^-/-^* or tacrolimus-treated mice^19^, and organ transplant recipients on immunosuppressive regimens that include glucocorticoids and tacrolimus^66^, further strengthening the link to NFAT1 function. In light of these observations, it is important to consider other findings in *Nfatc2^-/-^* mice, notably the development of chondrosarcomas^18^ and susceptibility to osteoarthiritis^20^, as additional patients with NFAT1 deficiency are identified.

**Table 2:**
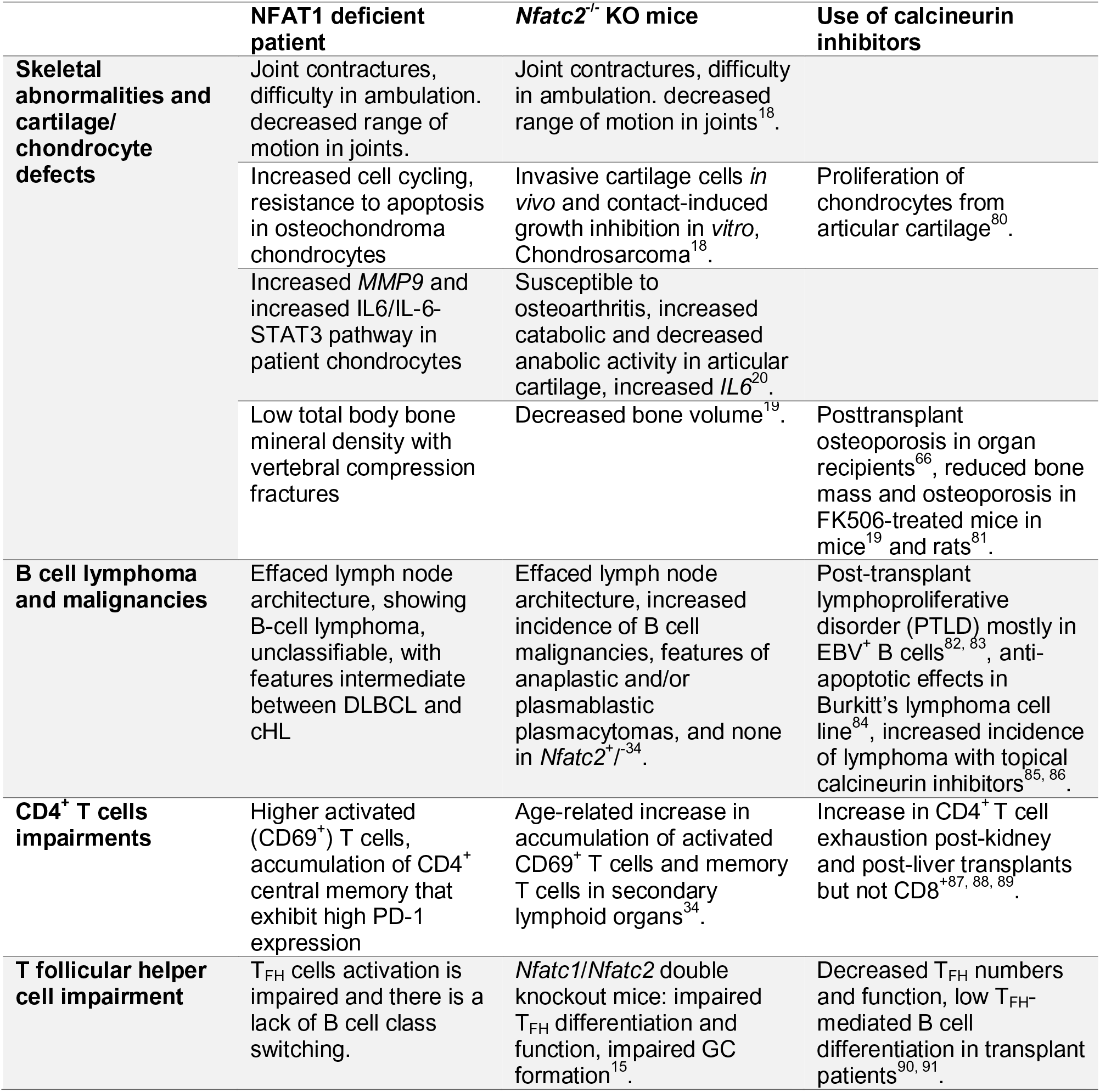
Comparison of human NFAT1 deficiency, Nfatc2-/- mice, and patients on calcineurin inhibitors.

One of the most significant phenotypes in our patient was the development of a rare mature B cell lymphoma characterized by large CD20^+^CD30^+^CD15^-^BCL6^+^ cells (**Fig. 1C**). Intriguingly, an increased incidence of anaplastic and/or plasmablastic plasmacytomas –also rare– was also observed with aging *Nfatc2^-/-^* mice^34^. Abrogation of NFAT1 function in B cells may confer an intrinsic pro-survival transcriptional program. NFAT1 is critical for repressing cell cycle progression through cyclin E (*CCNE2*)^67^ and our findings extend this understanding to suggest that JNK signaling (via *DUSP4*^68^) and AKT signaling (via *PIK3AP1*^55^) (**Fig. 3B, 3C**), might contribute to dysregulated B cell proliferation in the context of NFAT1 deficiency. In parallel, the CD4^+^ T cells of the NFAT1-deficient patient expressed high levels of the inhibitory checkpoint marker, PD-1 (**Fig. 6A**), which has frequently been associated with B cell lymphomas^69, 70^. This could inhibit effective anti-tumour effector functions in the patient. This is likely further exacerbated by the markedly reduced production of effector cytokines (TNF-α, IFN-γ) in patient’s CD4^+^ and CD8^+^ T cells (**Fig. 4-5**). Together these data indicate that ongoing clinical tumor surveillance, and a heightened awareness of tumor risk, will be an important aspect of the long-term care of patients with NFAT1 deficiency.

Studying complete human germline NFAT1 deficiency can provide new insights into the use of calcineurin inhibitors and improves our understanding of the consequences of long-term NFAT1 inhibition by calcineurin inhibitors (**Table 2**). Patients undergoing solid organ or allogeneic hematopoietic stem cell transplantation are frequently treated with an immunosuppressive regimen including a calcineurin inhibitor for extended periods of time. This can lead to a rare complication called post-transplant lymphoproliferative disorder (PTLD). Most PTLD’s are a B cell proliferative disorder believed to be due to impaired CD8^+^ T cell function and EBV infection^71, 72^. Our findings complement this understanding since NFAT1-deficient CD8^+^ T cells express significantly reduced cytotoxic mediators such as *FASLG* and *IFNG* (**Fig. 4B, 4H**). Consistent with NFAT1-dependent regulation of the transcription factor Egr3 being critical for FasL induction^73^, we found that both *EGR3* and *FASLG* were downregulated in CD4^+^ and CD8^+^ patient memory T cells (**Fig. 4-5**). Future work should focus on the role of NFAT1 in intrinsic B cell proliferation and CD4^+^ T cell exhaustion and the role of these factors in the pathophysiology of PTLD.

Mechanistically, human NFAT1 deficiency has revealed the remarkable pleiotropic and cell-type specific functions of NFAT1. Our study supports previous work that ascribed both oncogenic and tumor suppressor roles to NFAT1, depending on the cell type^24^. Informed by this patient, NFAT1 serves as a tumor suppressor in chondrocytes and B cells. This work also clarifies the role of human NFAT1 in B cell class-switching. Specifically, the B cell differentiation block is likely attributable to impaired T cell help. Previous studies demonstrated that both NFAT1 and NFAT2 are important for T_FH_ differentiation^15^. However, this NFAT1-deficient patient demonstrates that human NFAT1 is likely dispensable for T_FH_ differentiation but is critical for T_FH_ activation and function (**Fig. 6B-I**). Similarly, NFAT1 is essential for T_H_ cell differentiation and cytokine production (**Fig. 5F-5N**). One possible explanation for impaired CD4^+^ memory function could be that they are exhausted. This is supported by high PD-1 (**Fig. 6A**), and *TIGIT* (**Fig. 5A-5B**) expression^74^.

Accumulation of memory T cells^34, 75^, along with hyperresponsiveness has been documented in T cells of *Nfatc2^-/-^* mice^64, 65, 75, 76^. We observed suppressed cytokine signaling in the patient memory CD4^+^ and CD8^+^ cells. However, patient naïve CD4^+^ T cells do display signatures of activation, with higher PD-1 and CD69 (**Supp. Fig. 2J–2T**), as well as higher expression of T cell activation markers *CXCR3*^77^, *ITGA4* (an α subunit of integrin receptors), and *IL2RB* (CD122) (**Supp. Table 3**), a cytokine receptor recently found to be involved in homeostatic proliferation of naïve CD4^+^ T cells^78^. Higher activation status in naïve T cells might explain the age-associated accumulation of memory T cells observed in knock-out mice^34^ and the NFAT1-deficient patient.

Given the significant T and B cell dysregulation identified in this NFAT1-deficient patient and the absence of classical features for immunodeficiency (recurrent/severe/unusual infections), we propose to classify human NFAT1 deficiency as a primary immune regulatory disorder^79^. Studying this first reported patient has given us unique insight into the role of NFAT1 in the human musculoskeletal and immune systems. Our findings corroborate decades of *Nfatc2^-/-^* murine studies, while also providing a mechanistic understanding of human NFAT1 deficiency in different cell types. Beyond individual patients, this study also informs the clinical implementation of calcineurin inhibitors and highlights potential adverse consequences of long-term use.

## Data Availability

All data produced in the present work is either contained in the manuscript or deposited on Gene Expression Omnibus (bulk RNA-Seq and single-cell RNA-Seq).

https://www.ncbi.nlm.nih.gov/geo/query/acc.cgi?acc=GSE193415

## Acknowledgements

This work was supported by grants from the Canadian Institutes of Health Research (PJT 178054) (S.E.T.), Genome British Columbia (SIP007) (S.E.T.), and BC Children’s Hospital Foundation. S.E.T. holds a Tier 1 Canada Research Chair in Pediatric Precision Health and the Aubrey J. Tingle Professor of Pediatric Immunology. M.S. is supported by a CIHR Frederick Banting and Charles Best Canada Graduate Scholarships Doctoral Award (CGS-D) and University of British Columbia Four Year Doctoral Fellowship (4YF). H.Y.L. is supported by a CGS-D, 4YF, Killam Doctoral Scholarship, Friedman Award for Scholars in Health, and a BC Children’s Hospital Research Institute Graduate Studentship. Investigators in the CAUSES Study include: Shelin Adam, Nick Dragojlovic, Christèle du Souich, Alison M. Elliott, Anna Lehman, Larry Lynd, Jill Mwenifumbo, Tanya N. Nelson, Clara van Karnebeek, and Jan M. Friedman. We would like to acknowledge Navdeep Sangha at Thomas Jefferson University Hospital for providing his expertise in B cell lymphomas. We would also like to acknowledge the histology labs at BC Children’s Hospital and BC Cancer for their assistance in histological and immunohistochemistry, and the Biomedical Research Centre Sequencing Core (BRC-Seq) for their assistance with RNA-Sequencing and processing.

## Supplementary figures

**Supp Fig. 1.**
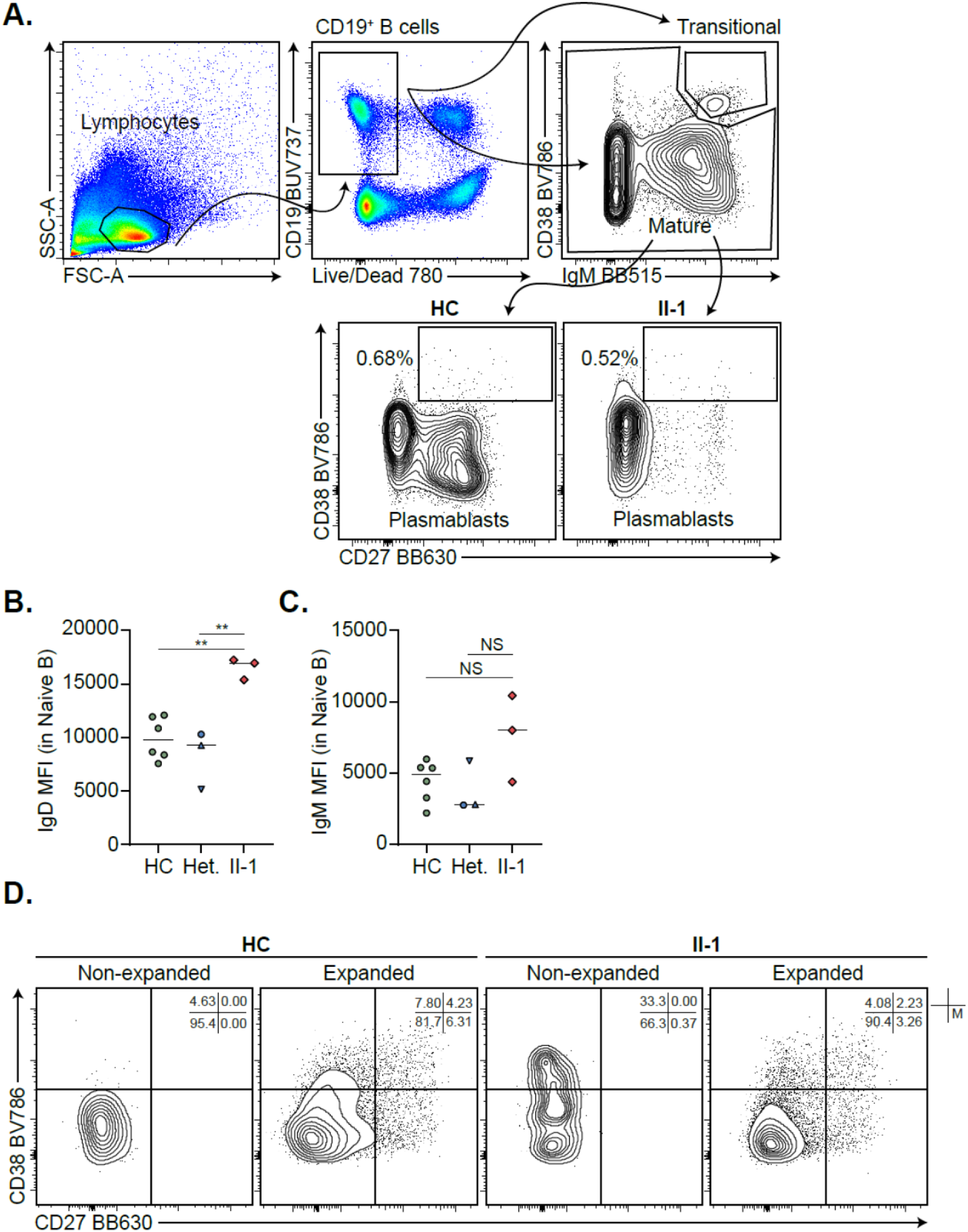
Extended B cell phenotyping. **A)** Gating strategy for transitional B cells and plasmablasts in mature CD19^+^ B cells. **B,C)** Quantification of IgD and IgM MFI in patient and control naïve B cells. **D)** Naïve B cells were isolated from patient and a control and expanded *in vitro*. Frequency of CD27^+^CD38^-^ memory B cells was enumerated at Day 0 (Non-expanded) and Day 6 (Expanded). **p<0.01. One-way ANOVA and Tukey’s post-hoc test. Green circles=healthy control, blue circle=II-2, blue upright triangle=I-2, blue inverted triangle=I-1, blue square=II-3, red diamond=II-1.

**Supp Fig. 2.**
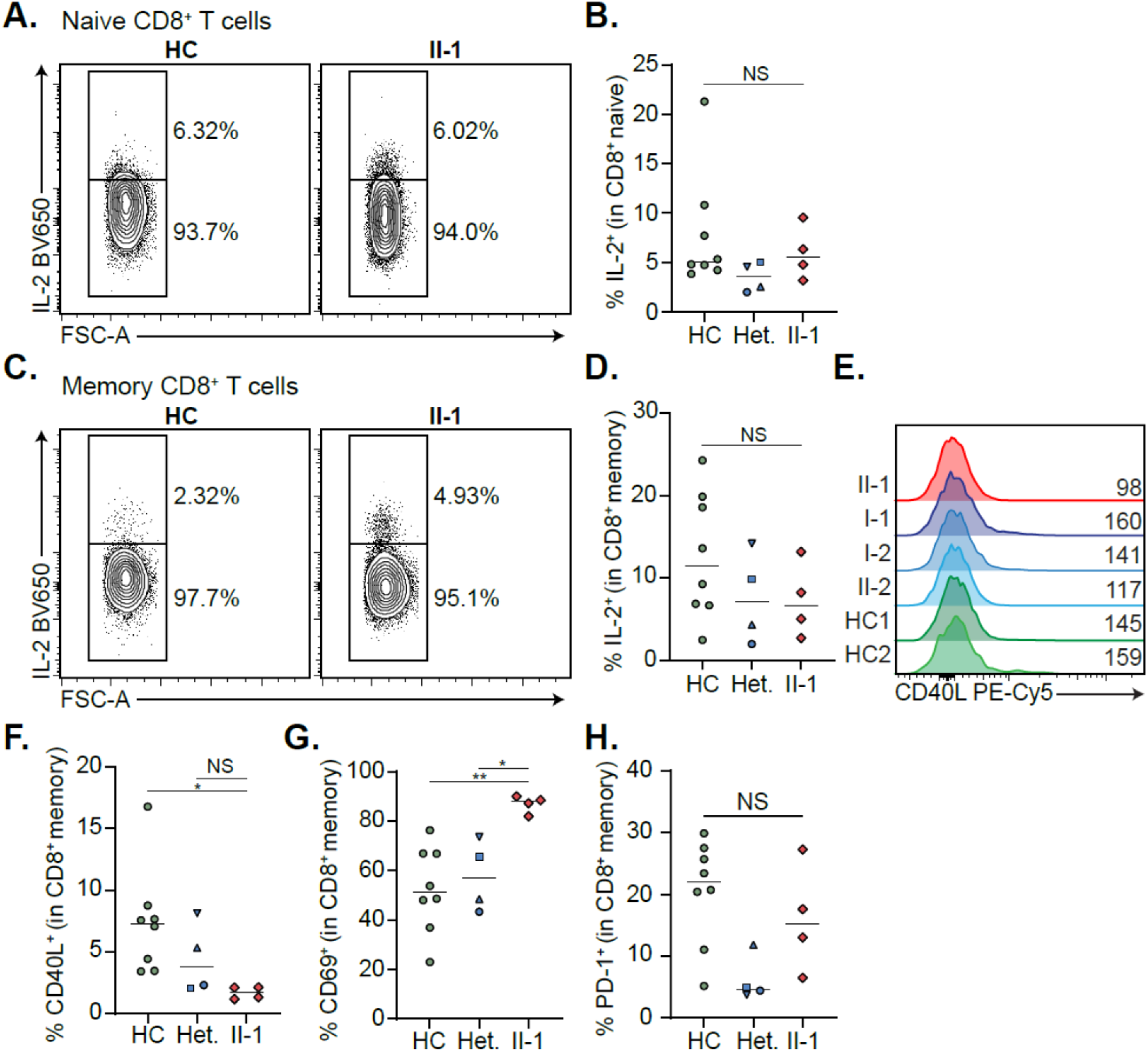
Extended CD8+ T cell phenotyping. **A)** Frequency of IL-2^+^ naïve CD8^+^ T cells in the patient and a representative control. **B)** Quantification of **A)**. **C)** Frequency of IL-2^+^ memory CD8^+^ T cells in the patient and a representative control. **D)** Quantification of **C)**. **E)** CD40L expression in memory CD8^+^ T cells. Mean fluorescence intensities are indicated. **F-H)** Quantification of **F)** CD40L^+^, **G)** CD69^+^, and **H)** PD-1^+^ CD8^+^ memory T cells. *p<0.05, **p<0.01. One-way ANOVA and Tukey’s post-hoc test. Green circles=healthy control, blue circle=II-2, blue upright triangle=I-2, blue inverted triangle=I-1, blue square=II-3, red diamond=II-1.

**Supp Fig. 3.**
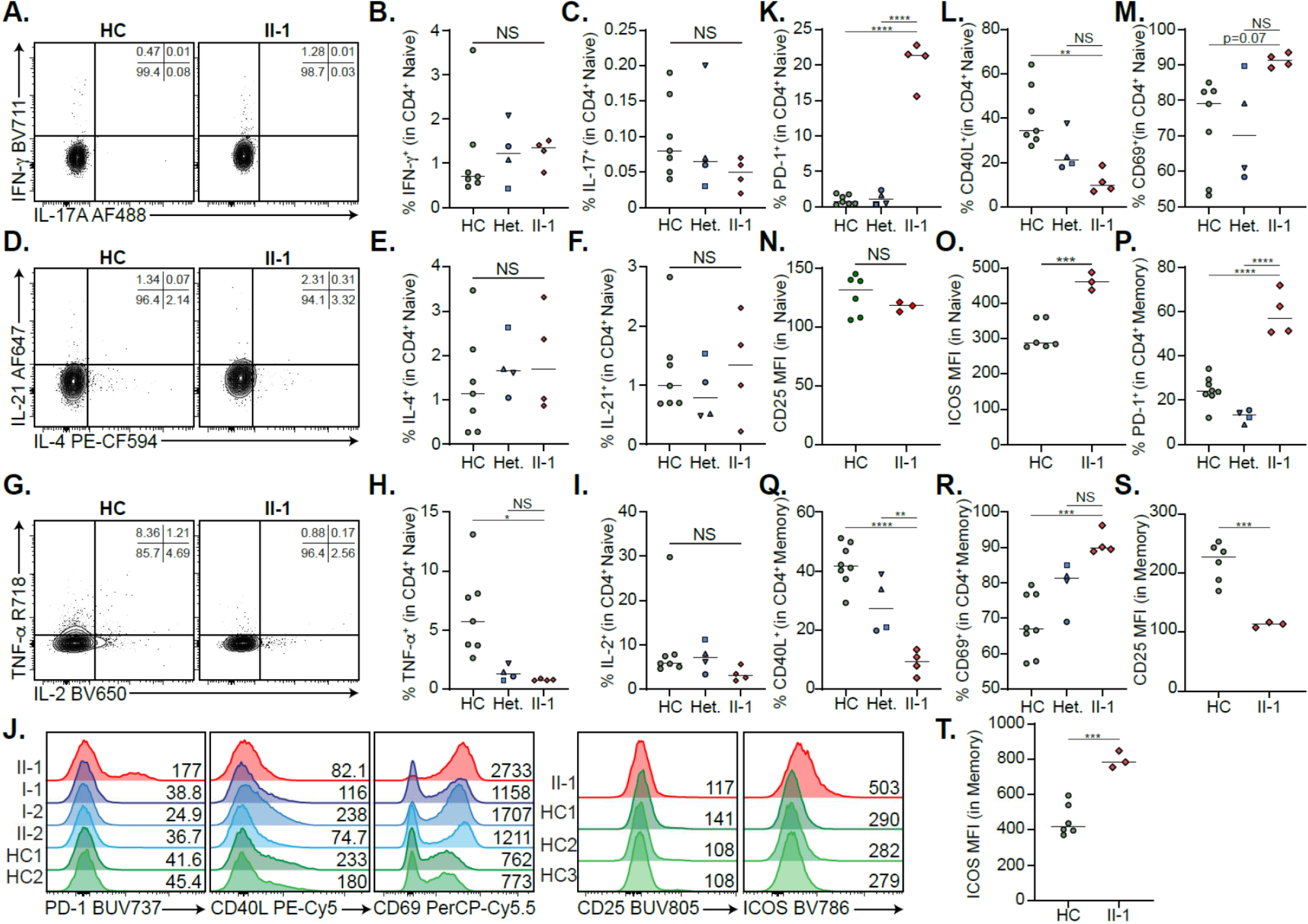
Extended CD4^+^ T cell immunophenotyping. **A)** Frequency of IFN-γ^+^ and IL-17A^+^ naïve CD4^+^ T cells in the patient and a representative control. **B,C)** Quantification of **A)**. **D)** Frequency of IL-21^+^ and IL-4^+^ naïve CD4^+^ T cells in the patient and a representative control. **E,F)** Quantification of **D)**. **G)** Frequency of TNF-α and IL-2 naïve CD4 T cells in the patient and a representative control. **H,I)** Quantification of **G)**. **J)** naïve CD4^+^ T cell expression of activation markers, including PD-1, CD40L, CD69, CD25, and ICOS. Mean fluorescence intensities (MFIs) are indicated. **K-O)** Frequency of same activation markers as in **J)**. **P-T)** Quantification of frequencies and MFIs of CD4^+^ memory T cell activation markers, including PD-1, CD40L, CD69, CD25 and ICOS. *p<0.05, **p<0.01, ***p<0.001, ****p<0.0001. One-way ANOVA and Tukey’s post-hoc test. Green circles=healthy control, blue circle=II-2, blue upright triangle=I-2, blue inverted triangle=I-1, blue square=II-3, red diamond=II-1.

**Supp Fig. 4.**
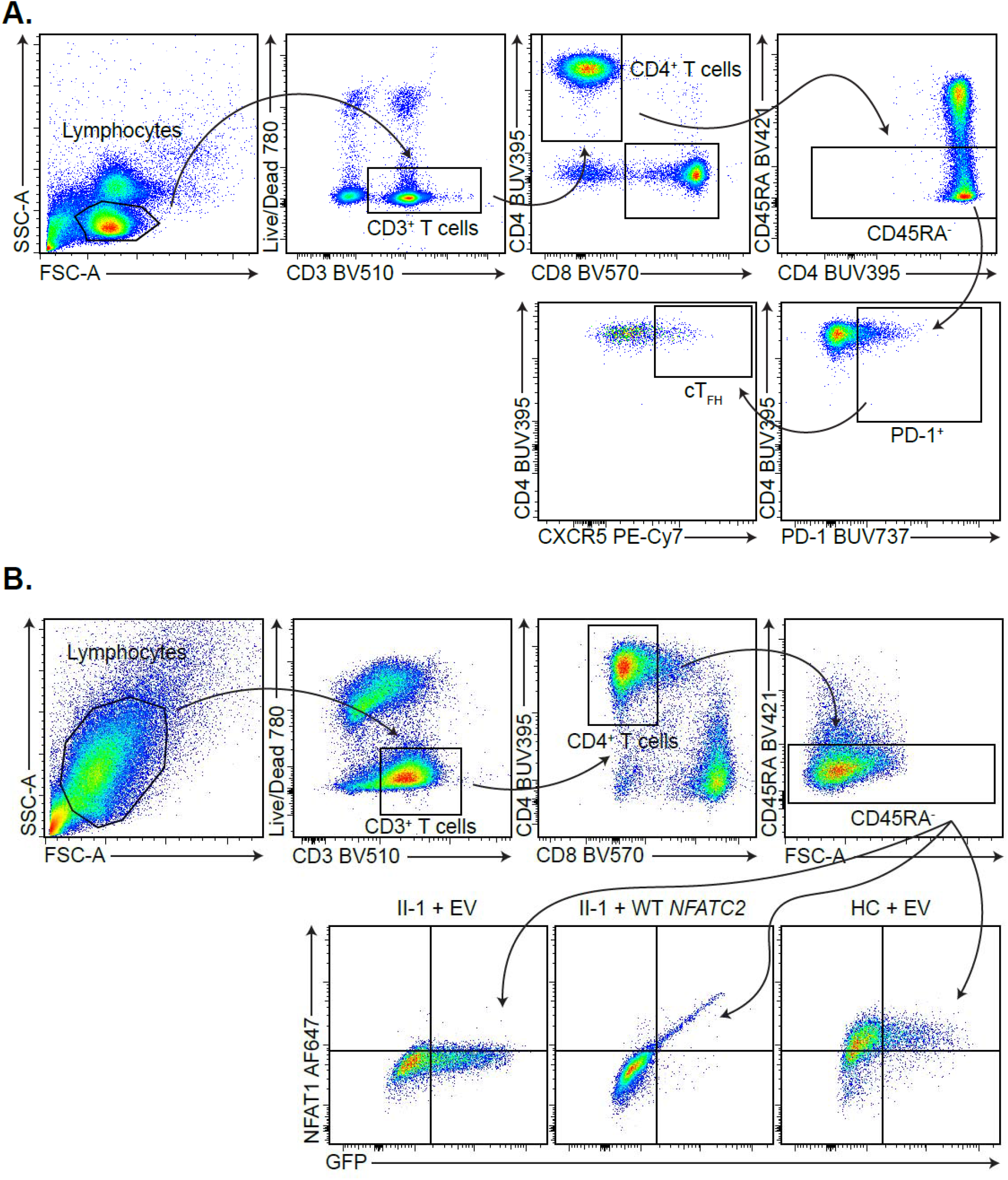
CD4^+^ T cell gating strategies. **A)** Gating strategy for circulating T follicular helper cells (T_FH_) defined as CD3^+^CD4^+^CD8^-^CD45RA^-^CXCR5^+^PD-1^+^. **B)** Gating strategy for identifying WT *NFATC2*-transduced CD4^+^ memory T cells.

**Supp Fig. 5.**
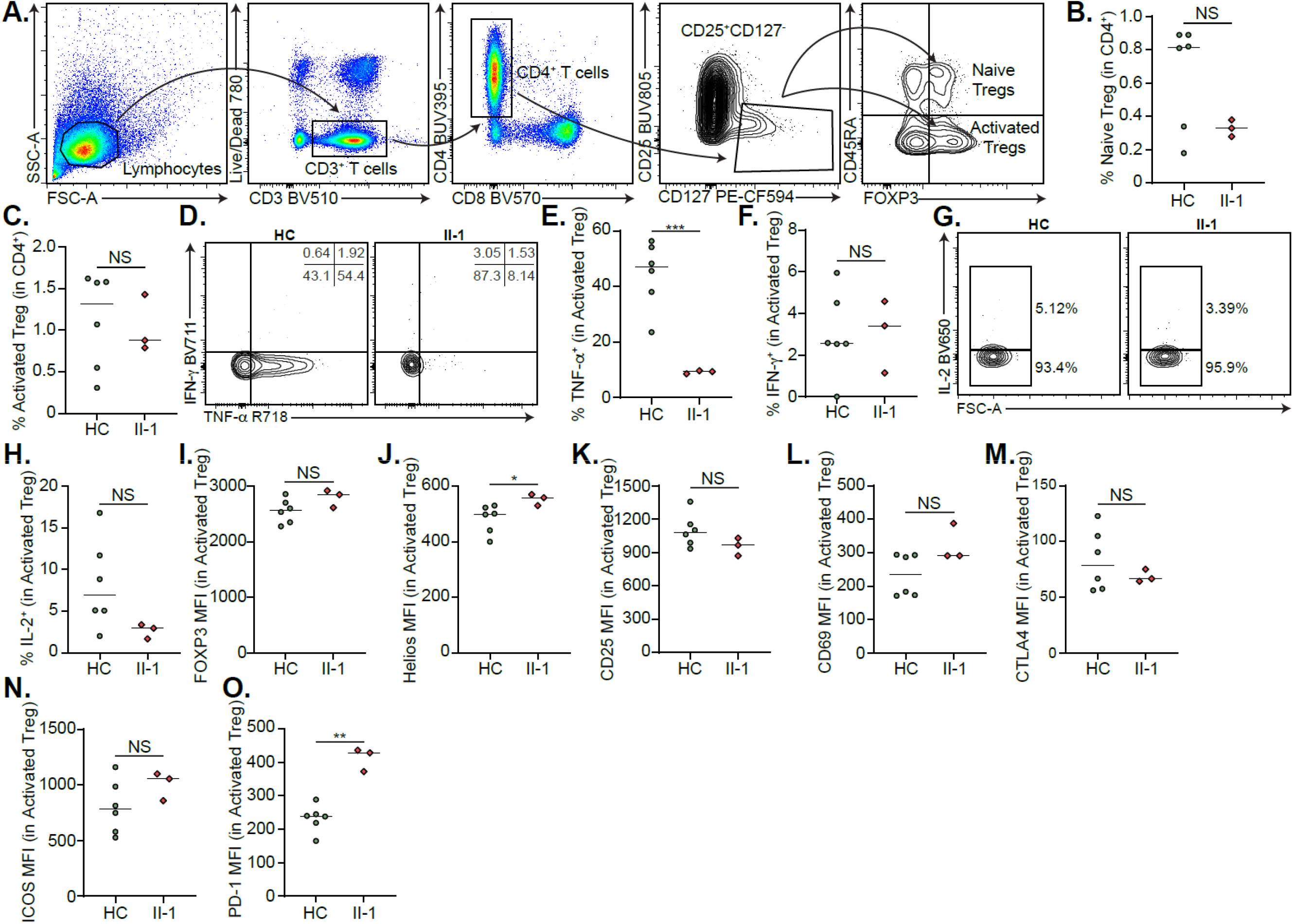
Patient T regulatory cell phenotype. **A)** Gating strategy for naïve and activated T regulatory cells (Treg) defined as CD3^+^CD4^+^CD8^-^CD25^+^CD127^-^FOXP3^+^ and CD45RA^+/-^. **B,C)** Frequency of **B)** naïve and **C)** memory Tregs. **D)** Frequency of TNF-α and IFN-γ activated Tregs in the patient and a representative control. **E,F)** Quantification of **D)**. **G)** Frequency of IL-2^+^ activated Tregs in the patient and a representative control. **H)** Quantification of **G)**. **I-O)** Quantification of Treg stability markers **I)** FOXP3 and **J)** Helios, and activation markers, **K)** CD25, **L)** CD69, **M)** CTLA4, **N)** ICOS, and **O)** PD-1 in activated Tregs. **p<0.01. One-way ANOVA and Tukey’s post-hoc test. Green circles=healthy control, red diamond=II-1.

## Supplementary Tables

**Supplementary Table 1:**
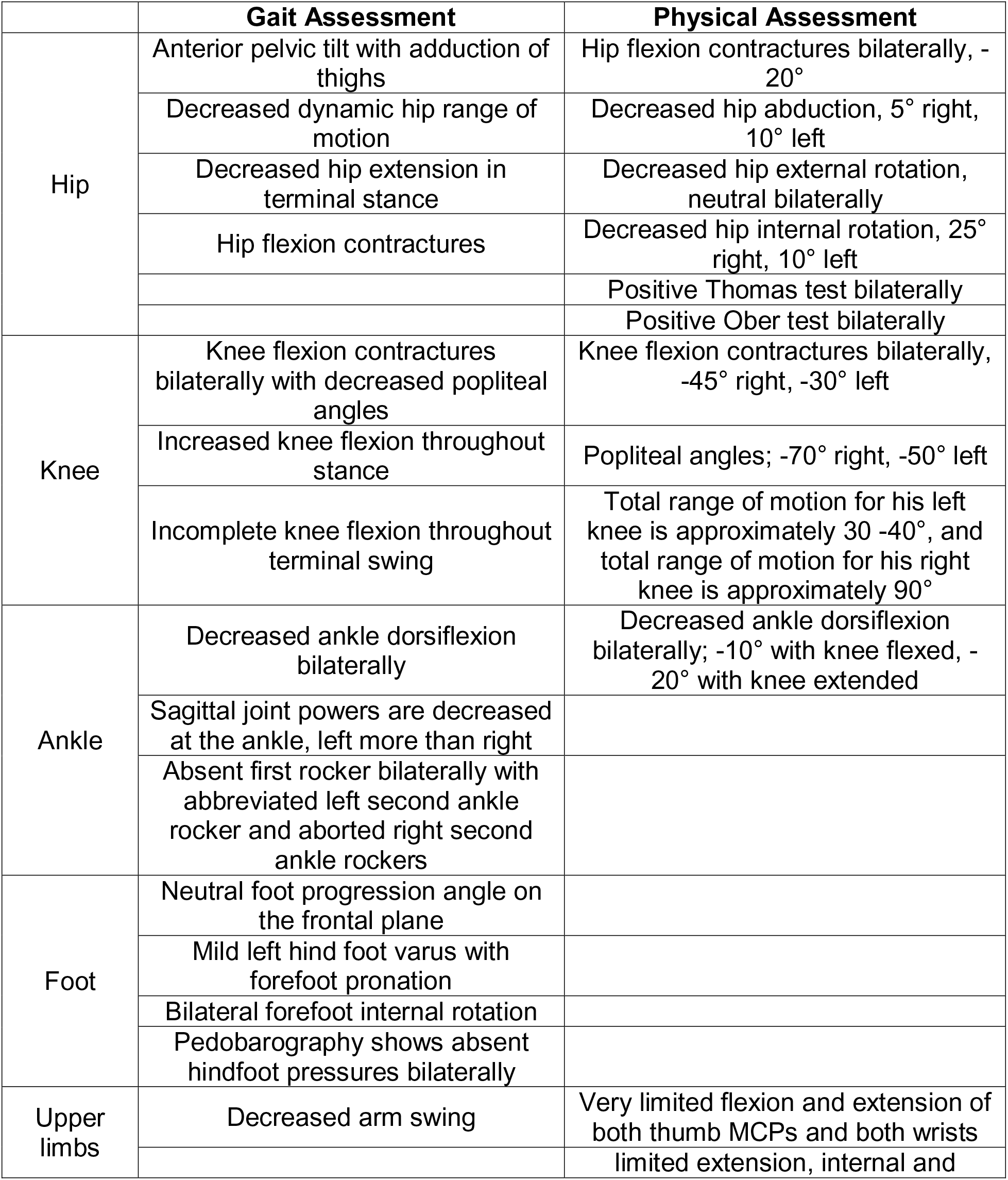
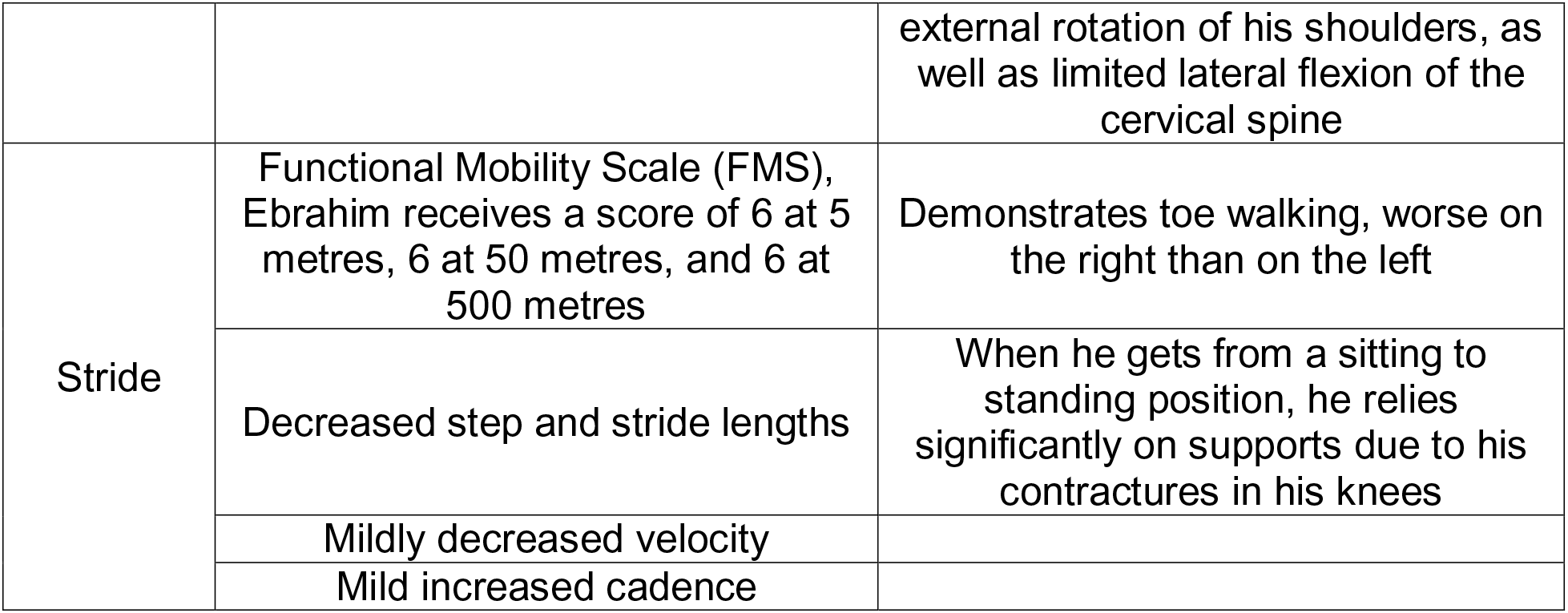
Gait and physical assessment in the patient

**Supplementary Table 2:**
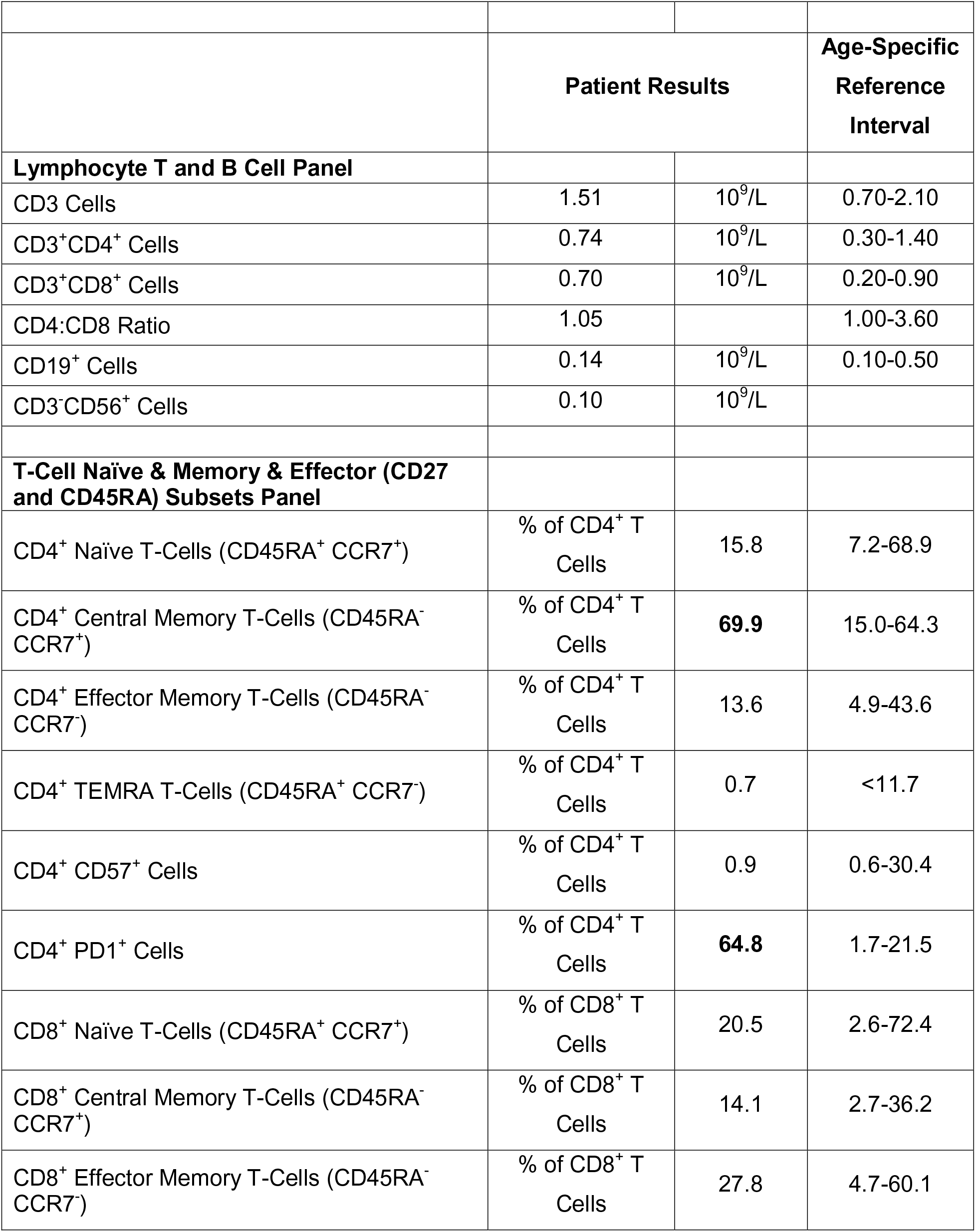
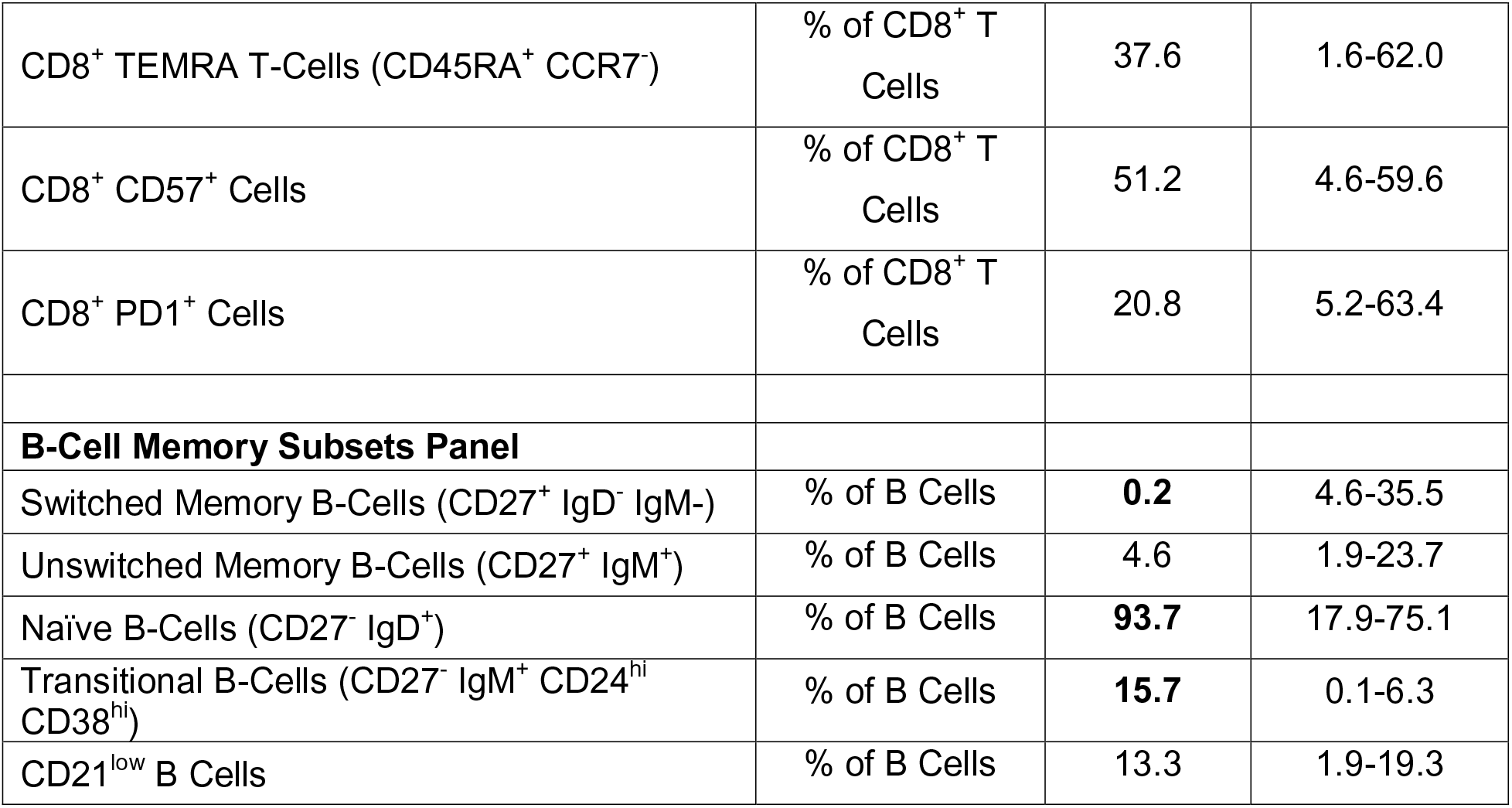
Clinical Flow Cytometry for determination of T cell and B cell subset proportions in the patient along with reference intervals

