## Supplementary Table 3 for "Human germline biallelic complete NFAT1 deficiency causes the triad of progressive joint contractures, osteochondromas, and susceptibility to B cell malignancy"

Supplementary Table 3: Differential expression in different subsets of PBMCs in patient against five healthy controls (3 hetrozygous family members, 2 age-matched healthy controls)

**Unstimulated samples**

CD4+ Naïve

| | p_val | avg_log2FC | Patient | Controls | p_val_adj |
|---|---|---|---|---|---|
| ITGA4 | 1.14E-156 | 1.114652474 | 0.763 | 0.411 | 5.69E-154 |
| CD4 | 1.65E-68 | 0.554509821 | 0.888 | 0.737 | 8.22E-66 |
| CD52 | 7.56E-64 | 0.612750754 | 0.992 | 0.957 | 3.77E-61 |
| TIGIT | 1.05E-61 | 0.4768707 | 0.304 | 0.058 | 5.25E-59 |
| PASK | 3.04E-58 | 1.025426687 | 0.759 | 0.582 | 1.51E-55 |
| ITGB2 | 7.29E-53 | 0.504977744 | 0.897 | 0.727 | 3.63E-50 |
| PFN1 | 6.09E-52 | 0.400448875 | 1 | 1 | 3.03E-49 |
| LAT | 1.07E-48 | 0.494292192 | 0.941 | 0.878 | 5.32E-46 |
| GAPDH | 2.40E-45 | 0.554750474 | 0.959 | 0.892 | 1.20E-42 |
| EVI2B | 6.69E-44 | 0.519573854 | 0.885 | 0.707 | 3.33E-41 |
| DUSP2 | 6.26E-40 | 0.683084059 | 0.433 | 0.242 | 3.12E-37 |
| SELPLG | 5.59E-39 | 0.415622952 | 0.91 | 0.78 | 2.78E-36 |
| STAT1 | 1.56E-37 | -0.678168788 | 0.64 | 0.709 | 7.79E-35 |
| BTG1 | 4.59E-35 | -0.52164304 | 0.946 | 0.975 | 2.28E-32 |
| RPL6 | 2.58E-34 | 0.348239719 | 1 | 1 | 1.29E-31 |
| FYB1 | 6.45E-34 | 0.401657701 | 0.927 | 0.831 | 3.21E-31 |
| TRIB2 | 1.79E-32 | 0.488580897 | 0.688 | 0.451 | 8.93E-30 |
| S100A10 | 4.03E-32 | 0.414296198 | 0.894 | 0.819 | 2.01E-29 |
| GNAI2 | 2.38E-30 | 0.305348892 | 0.997 | 0.989 | 1.19E-27 |
| IFITM2 | 1.46E-27 | 0.412838065 | 0.926 | 0.864 | 7.27E-25 |
| IL32 | 1.30E-25 | 0.426627342 | 0.769 | 0.617 | 6.45E-23 |
| CD3E | 2.46E-25 | 0.282508286 | 0.992 | 0.98 | 1.23E-22 |
| HIF1A | 3.21E-25 | -0.397418078 | 0.412 | 0.548 | 1.60E-22 |
| NFKBIA | 1.41E-22 | -0.460871565 | 0.654 | 0.72 | 7.01E-20 |
| DOCK8 | 2.29E-22 | 0.285887916 | 0.789 | 0.658 | 1.14E-19 |
| CD5 | 1.70E-21 | 0.342644304 | 0.779 | 0.673 | 8.46E-19 |
| CD3D | 1.13E-19 | 0.272896246 | 0.972 | 0.941 | 5.62E-17 |
| IFI6 | 3.28E-18 | -0.315960771 | 0.101 | 0.2 | 1.63E-15 |
| CD226 | 6.87E-18 | 0.183042831 | 0.192 | 0.087 | 3.42E-15 |
| CCR7 | 1.24E-17 | -0.321775909 | 0.959 | 0.962 | 6.18E-15 |
| IL6R | 1.57E-17 | 0.21195398 | 0.304 | 0.172 | 7.84E-15 |
| LCK | 1.02E-16 | 0.243099366 | 0.969 | 0.926 | 5.08E-14 |
| CD27 | 1.19E-16 | 0.309262045 | 0.748 | 0.581 | 5.95E-14 |
| SNX9 | 3.67E-16 | -0.223502753 | 0.116 | 0.24 | 1.83E-13 |
| BIRC3 | 9.83E-15 | -0.363188138 | 0.451 | 0.539 | 4.89E-12 |
| PTPRC | 2.46E-14 | 0.207662081 | 0.988 | 0.981 | 1.22E-11 |
| CD2 | 2.75E-14 | 0.258905714 | 0.896 | 0.853 | 1.37E-11 |
| XBP1 | 6.85E-14 | -0.241356157 | 0.241 | 0.387 | 3.41E-11 |
| POU2AF1 | 9.27E-14 | 0.149360523 | 0.132 | 0.053 | 4.62E-11 |

| | | | | | |
|---|---|---|---|---|---|
| CXCR3 | 1.86E-13 | 0.180087775 | 0.17 | 0.069 | 9.29E-11 |
| CD44 | 1.95E-13 | 0.214599548 | 0.956 | 0.932 | 9.72E-11 |
| IRF1 | 2.24E-13 | -0.359135749 | 0.605 | 0.643 | 1.11E-10 |
| ARPC1B | 2.81E-13 | 0.25050031 | 0.765 | 0.654 | 1.40E-10 |
| CREM | 4.84E-13 | -0.35472934 | 0.198 | 0.257 | 2.41E-10 |
| STAT3 | 7.54E-13 | -0.269563957 | 0.59 | 0.688 | 3.76E-10 |
| PIK3IP1 | 8.05E-13 | -0.254853236 | 0.856 | 0.909 | 4.01E-10 |
| IRF7 | 1.94E-12 | -0.274720008 | 0.328 | 0.418 | 9.68E-10 |
| CST7 | 1.91E-11 | 0.212122197 | 0.282 | 0.202 | 9.49E-09 |
| CD37 | 2.11E-11 | 0.215967758 | 0.895 | 0.828 | 1.05E-08 |
| LAMP3 | 2.83E-11 | -0.175582239 | 0.041 | 0.1 | 1.41E-08 |
| GBP2 | 4.02E-11 | -0.249170449 | 0.374 | 0.455 | 2.00E-08 |
| TRAC | 7.34E-11 | 0.227489726 | 0.992 | 0.982 | 3.66E-08 |
| GBP1 | 2.01E-10 | -0.271623487 | 0.351 | 0.405 | 1.00E-07 |
| ZAP70 | 3.43E-10 | 0.184111166 | 0.409 | 0.298 | 1.71E-07 |
| RGS1 | 3.71E-10 | -0.334811182 | 0.088 | 0.134 | 1.85E-07 |
| PSME2 | 4.34E-10 | -0.235222894 | 0.725 | 0.771 | 2.16E-07 |
| TSPAN32 | 5.19E-10 | 0.117097185 | 0.13 | 0.061 | 2.59E-07 |
| ARHGEF3 | 1.77E-09 | 0.187069933 | 0.458 | 0.346 | 8.80E-07 |
| NFATC1 | 1.81E-09 | 0.164334119 | 0.377 | 0.266 | 9.00E-07 |
| NFKBIZ | 2.46E-09 | -0.225446519 | 0.336 | 0.419 | 1.22E-06 |
| CD247 | 4.24E-09 | 0.186606546 | 0.573 | 0.485 | 2.11E-06 |
| NFKB1 | 1.26E-08 | -0.229553038 | 0.525 | 0.563 | 6.27E-06 |
| TBX21 | 1.28E-08 | 0.122527882 | 0.133 | 0.063 | 6.37E-06 |
| CD274 | 1.56E-08 | 0.105092297 | 0.125 | 0.065 | 7.79E-06 |
| CXCR4 | 2.47E-08 | -0.286212511 | 0.875 | 0.894 | 1.23E-05 |
| CD69 | 3.74E-08 | -0.230280029 | 0.304 | 0.38 | 1.86E-05 |
| VSIR | 3.85E-08 | 0.155542629 | 0.345 | 0.255 | 1.92E-05 |
| CD300A | 4.12E-08 | 0.11573988 | 0.16 | 0.09 | 2.05E-05 |
| IL4R | 4.92E-08 | -0.195259044 | 0.375 | 0.445 | 2.45E-05 |
| IL6ST | 5.95E-08 | -0.197613262 | 0.571 | 0.675 | 2.96E-05 |
| ILF2 | 1.07E-07 | -0.185650332 | 0.492 | 0.555 | 5.35E-05 |
| IL7R | 1.38E-07 | 0.211266583 | 0.647 | 0.586 | 6.87E-05 |
| EIF5A | 1.76E-07 | -0.202156714 | 0.632 | 0.701 | 8.79E-05 |
| TGFBR2 | 2.95E-07 | -0.197400613 | 0.762 | 0.779 | 0.000146979 |
| WAS | 5.76E-07 | 0.1593344 | 0.498 | 0.423 | 0.000286879 |
| CD40LG | 8.49E-07 | 0.081388515 | 0.112 | 0.06 | 0.00042293 |
| CCL22 | 9.48E-07 | -0.22728912 | 0.122 | 0.131 | 0.000472162 |
| SOCS3 | 1.16E-06 | -0.140031802 | 0.134 | 0.198 | 0.000580136 |
| MAP2K3 | 1.21E-06 | 0.16018948 | 0.442 | 0.343 | 0.000602214 |
| CD200 | 1.45E-06 | 0.065344725 | 0.065 | 0.022 | 0.000723518 |
| GIMAP2 | 1.49E-06 | 0.16832648 | 0.771 | 0.727 | 0.000739878 |
| FYN | 1.88E-06 | 0.175978366 | 0.868 | 0.779 | 0.000934101 |
| IL2RG | 2.09E-06 | 0.152235948 | 0.917 | 0.889 | 0.001042754 |
| BIN2 | 2.24E-06 | 0.130524641 | 0.343 | 0.27 | 0.001116453 |
| DPP4 | 2.63E-06 | -0.075180194 | 0.027 | 0.075 | 0.001310015 |
| TIAF1 | 3.57E-06 | 0.086997875 | 0.141 | 0.088 | 0.001780246 |

| | | | | | |
|---|---|---|---|---|---|
| EIF1 | 3.59E-06 | 0.160076897 | 0.97 | 0.946 | 0.001785634 |
| LGALS9 | 3.66E-06 | 0.108411251 | 0.188 | 0.124 | 0.001822486 |
| IGBP1 | 4.64E-06 | 0.130804384 | 0.36 | 0.279 | 0.002308582 |
| ATF4 | 6.88E-06 | -0.189683038 | 0.814 | 0.824 | 0.003424992 |
| AHI1 | 7.46E-06 | 0.061559439 | 0.073 | 0.033 | 0.003713386 |
| TNFAIP8 | 8.03E-06 | 0.147666662 | 0.708 | 0.661 | 0.004000628 |
| BACH2 | 1.93E-05 | -0.09793603 | 0.095 | 0.157 | 0.009632714 |
| NR4A2 | 2.52E-05 | -0.148998035 | 0.036 | 0.075 | 0.012556093 |
| ADA | 2.63E-05 | 0.118805945 | 0.274 | 0.215 | 0.013111383 |
| STAT6 | 3.22E-05 | 0.121985419 | 0.38 | 0.299 | 0.016037559 |
| VPS28 | 3.30E-05 | 0.132760356 | 0.639 | 0.596 | 0.016449357 |
| HLA.DPA1 | 4.78E-05 | 0.163018177 | 0.527 | 0.428 | 0.023780238 |
| IL21 | 4.91E-05 | 0.024545621 | 0.017 | 0.001 | 0.024462985 |
| CD48 | 5.31E-05 | 0.137454945 | 0.958 | 0.916 | 0.026419618 |
| CXCR5 | 7.21E-05 | 0.133628302 | 0.171 | 0.091 | 0.035924649 |
| IRF9 | 8.51E-05 | -0.161347403 | 0.668 | 0.667 | 0.042378189 |
| BCL2 | 9.64E-05 | 0.090776915 | 0.141 | 0.098 | 0.048015034 |
| LRRC8B | 9.87E-05 | 0.051356353 | 0.073 | 0.038 | 0.049140236 |
| PRF1 | 0.000121796 | 0.111624504 | 0.295 | 0.238 | 0.060654324 |
| CASP5 | 0.00015163 | 0.074207879 | 0.157 | 0.11 | 0.075511762 |
| MME | 0.000156326 | 0.025542491 | 0.014 | 0.001 | 0.077850382 |
| HLA.A | 0.000193032 | 0.095332025 | 1 | 1 | 0.096129757 |
| JAK1 | 0.000215002 | 0.108742079 | 0.949 | 0.93 | 0.107070757 |
| PAK1IP1 | 0.000217046 | -0.058312638 | 0.038 | 0.075 | 0.108088992 |
| IER3 | 0.000236437 | 0.128750308 | 0.37 | 0.304 | 0.117745758 |
| CHI3L2 | 0.000259113 | 0.078499723 | 0.13 | 0.084 | 0.12903812 |
| LDHA | 0.000287921 | -0.133571846 | 0.848 | 0.857 | 0.143384532 |
| NANS | 0.000297034 | -0.097087461 | 0.177 | 0.234 | 0.147922966 |
| SLC1A5 | 0.000394011 | 0.078622745 | 0.154 | 0.104 | 0.196217601 |
| APOBEC3G | 0.000397851 | 0.093437147 | 0.248 | 0.197 | 0.198129703 |
| CD3G | 0.000405217 | 0.071194043 | 0.141 | 0.098 | 0.201797866 |
| ARL4C | 0.000407416 | 0.121974951 | 0.706 | 0.642 | 0.202892953 |
| RPN2 | 0.000455452 | 0.108462847 | 0.583 | 0.509 | 0.226814886 |
| IFNG | 0.000467932 | 0.071213831 | 0.045 | 0.017 | 0.23303032 |
| PDCD1 | 0.000481187 | 0.019980071 | 0.013 | 0.002 | 0.239631223 |
| SELL | 0.000550101 | -0.142647496 | 0.85 | 0.907 | 0.27395035 |
| ADGRE1 | 0.000644304 | 0.067032237 | 0.09 | 0.058 | 0.32086322 |
| TNFSF10 | 0.000652779 | 0.118966818 | 0.419 | 0.356 | 0.325083838 |
| FAS | 0.000786848 | 0.055380759 | 0.091 | 0.049 | 0.391850155 |
| KDELR1 | 0.000886951 | 0.09405654 | 0.374 | 0.318 | 0.441701393 |
| IL2RB | 0.00089967 | 0.117745101 | 0.346 | 0.265 | 0.44803583 |
| SKAP1 | 0.001032627 | 0.059221597 | 0.113 | 0.081 | 0.514248319 |
| PDIA6 | 0.001047133 | 0.108887026 | 0.471 | 0.4 | 0.521472161 |
| CD83 | 0.001328981 | -0.086390416 | 0.112 | 0.131 | 0.661832609 |
| GZMK | 0.001567261 | 0.066683132 | 0.043 | 0.038 | 0.780496163 |
| TOP2A | 0.001582172 | 0.024823169 | 0.025 | 0.009 | 0.787921529 |
| EVI2A | 0.001621067 | 0.110728812 | 0.635 | 0.591 | 0.807291607 |

| | | | | | |
|---|---|---|---|---|---|
| TRBC2 | 0.001628431 | 0.147238369 | 0.966 | 0.974 | 0.810958835 |
| PRNP | 0.002144091 | -0.073119696 | 0.127 | 0.176 | 1 |
| ERBIN | 0.002173683 | -0.103771803 | 0.507 | 0.514 | 1 |
| BAX | 0.002594857 | -0.108323577 | 0.432 | 0.449 | 1 |
| KLRK1 | 0.002980066 | 0.066955245 | 0.155 | 0.111 | 1 |
| LAP3 | 0.003792648 | -0.082410587 | 0.235 | 0.269 | 1 |
| PSME3 | 0.004994985 | -0.058063442 | 0.106 | 0.139 | 1 |
| ARID5A | 0.005252199 | -0.081937225 | 0.211 | 0.271 | 1 |
| EIF5 | 0.005402908 | -0.098434105 | 0.527 | 0.527 | 1 |
| IGLC3 | 0.006492521 | -0.140428964 | 0.472 | 0.472 | 1 |
| IFITM3 | 0.00666535 | -0.078592717 | 0.177 | 0.209 | 1 |
| ATF6B | 0.007350059 | 0.074458323 | 0.346 | 0.295 | 1 |
| ITGAX | 0.007424593 | -0.038825655 | 0.042 | 0.065 | 1 |
| CD72 | 0.007509743 | -0.025823986 | 0.012 | 0.027 | 1 |
| UFM1 | 0.008197078 | 0.084684004 | 0.527 | 0.49 | 1 |
| ITK | 0.008421481 | -0.090769523 | 0.836 | 0.845 | 1 |
| IL18R1 | 0.008448369 | 0.033033418 | 0.049 | 0.024 | 1 |
| ABCE1 | 0.009260828 | -0.07246745 | 0.34 | 0.365 | 1 |
| LAG3 | 0.009369845 | 0.06340704 | 0.082 | 0.051 | 1 |
| NAMPT | 0.009680184 | -0.067886375 | 0.162 | 0.192 | 1 |
| RUNX3 | 0.00988298 | -0.092952363 | 0.519 | 0.543 | 1 |
| IRF8 | 0.010329877 | -0.096534072 | 0.198 | 0.171 | 1 |
| LAMP1 | 0.010590043 | 0.080553891 | 0.486 | 0.455 | 1 |
| CD28 | 0.012716002 | 0.063966581 | 0.235 | 0.192 | 1 |
| STAT4 | 0.012735781 | -0.074445084 | 0.252 | 0.303 | 1 |
| KLRB1 | 0.013171125 | 0.012382125 | 0.007 | 0.002 | 1 |
| CCR9 | 0.013228075 | 0.042784582 | 0.037 | 0.015 | 1 |
| VNN2 | 0.013554208 | 0.050037569 | 0.156 | 0.127 | 1 |
| PSMB8 | 0.013774459 | -0.084855073 | 0.824 | 0.842 | 1 |
| CHD7 | 0.014991721 | -0.028656302 | 0.027 | 0.044 | 1 |
| CYCS | 0.016392453 | -0.072405004 | 0.299 | 0.338 | 1 |
| CD40 | 0.016627105 | -0.043957345 | 0.049 | 0.06 | 1 |
| TGFBR1 | 0.016946988 | -0.044160106 | 0.078 | 0.099 | 1 |
| GRB10 | 0.017844929 | -0.025683418 | 0.016 | 0.031 | 1 |
| RNASE6 | 0.018641239 | 0.018476062 | 0.027 | 0.015 | 1 |
| TNFSF8 | 0.019135963 | 0.022502598 | 0.028 | 0.018 | 1 |
| LTA | 0.02006593 | -0.049183861 | 0.083 | 0.105 | 1 |
| CSRNP1 | 0.020109552 | -0.064906008 | 0.172 | 0.203 | 1 |
| TNIK | 0.020137531 | 0.062954749 | 0.221 | 0.18 | 1 |
| TNFRSF25 | 0.020155932 | 0.053737738 | 0.16 | 0.129 | 1 |
| MALT1 | 0.021083244 | 0.050447484 | 0.181 | 0.147 | 1 |
| IKZF1 | 0.02574401 | 0.072646125 | 0.501 | 0.447 | 1 |
| ORAI1 | 0.025868553 | 0.066234446 | 0.404 | 0.361 | 1 |
| NFKB2 | 0.026704087 | -0.037623431 | 0.044 | 0.065 | 1 |
| TCF7 | 0.02844121 | 0.062278245 | 0.322 | 0.275 | 1 |
| CXCL10 | 0.030171772 | -0.034446342 | 0.05 | 0.065 | 1 |
| CD22 | 0.0306793 | -0.036437327 | 0.051 | 0.069 | 1 |

| | | | | | |
|---|---|---|---|---|---|
| MCM2 | 0.031123069 | -0.037812209 | 0.072 | 0.097 | 1 |
| DUSP6 | 0.033812756 | 0.017926818 | 0.029 | 0.017 | 1 |
| SOCS2 | 0.034791952 | 0.029010381 | 0.041 | 0.028 | 1 |
| IL18RAP | 0.035843706 | 0.019546914 | 0.029 | 0.015 | 1 |
| CXCL16 | 0.035991165 | -0.04997283 | 0.155 | 0.178 | 1 |
| SPP1 | 0.036570949 | 0.009818473 | 0.01 | 0.003 | 1 |
| CD24 | 0.039797742 | -0.012881558 | 0.003 | 0.012 | 1 |
| CTLA4 | 0.041060241 | 0.060297287 | 0.174 | 0.116 | 1 |
| NINJ2 | 0.049390264 | 0.018201105 | 0.027 | 0.013 | 1 |
| NFATC2 | 0.049566105 | -0.049861641 | 0.185 | 0.212 | 1 |
| NCAM1 | 0.052745159 | 0.007415872 | 0.006 | 0.002 | 1 |
| PDIA4 | 0.062690643 | 0.066466798 | 0.443 | 0.421 | 1 |
| UGCG | 0.065534961 | 0.049664534 | 0.249 | 0.227 | 1 |
| TYMS | 0.065857078 | 0.00880544 | 0.008 | 0.003 | 1 |
| FOXP1 | 0.076880656 | -0.043562105 | 0.113 | 0.148 | 1 |
| PSMB9 | 0.077487694 | -0.0649819 | 0.529 | 0.519 | 1 |
| KLRC1 | 0.078909205 | 0.015850154 | 0.032 | 0.022 | 1 |
| IL6 | 0.079817741 | 0.018531134 | 0.034 | 0.023 | 1 |
| TUBA1C | 0.091939289 | -0.057338737 | 0.435 | 0.455 | 1 |
| SP140 | 0.092363515 | 0.041571879 | 0.184 | 0.149 | 1 |
| TRAT1 | 0.094656842 | 0.042782859 | 0.207 | 0.181 | 1 |
| TRAF1 | 0.097986327 | 0.04011975 | 0.186 | 0.145 | 1 |
| PCNA | 0.104890958 | -0.046445061 | 0.269 | 0.287 | 1 |
| BATF | 0.104896491 | 0.027739775 | 0.091 | 0.069 | 1 |
| PAXBP1 | 0.105851025 | -0.05032657 | 0.384 | 0.402 | 1 |
| CCL2 | 0.106421163 | 0.076576774 | 0.792 | 0.756 | 1 |
| BCL6 | 0.107654054 | -0.019093449 | 0.031 | 0.041 | 1 |
| MS4A1 | 0.112573082 | -0.051588081 | 0.164 | 0.146 | 1 |
| MMP9 | 0.112633042 | -0.029739079 | 0.084 | 0.094 | 1 |
| AIM2 | 0.114139275 | 0.005756649 | 0.005 | 0.002 | 1 |
| ITGAE | 0.116181562 | -0.031584012 | 0.098 | 0.114 | 1 |
| RORA | 0.119519066 | -0.06379396 | 0.389 | 0.396 | 1 |
| FOXO1 | 0.126179071 | -0.038297044 | 0.168 | 0.179 | 1 |
| JUN | 0.128124881 | -0.02589796 | 0.05 | 0.055 | 1 |
| EBF1 | 0.13110397 | 0.005115308 | 0.006 | 0.002 | 1 |
| S100A12 | 0.132881715 | -0.014939114 | 0.017 | 0.028 | 1 |
| FOSB | 0.134218156 | -0.010340483 | 0.003 | 0.007 | 1 |
| LRRC32 | 0.134433144 | -0.010340483 | 0.003 | 0.007 | 1 |
| GATA3 | 0.139883103 | 0.014233689 | 0.03 | 0.02 | 1 |
| ANXA5 | 0.142378512 | 0.047533291 | 0.444 | 0.386 | 1 |
| EGR1 | 0.151222515 | -0.006022153 | 0.001 | 0.005 | 1 |
| CCL4 | 0.151934698 | -0.062170462 | 0.258 | 0.271 | 1 |
| MYB | 0.154168579 | 0.020294306 | 0.032 | 0.021 | 1 |
| TRDC | 0.156186683 | 0.03870761 | 0.073 | 0.069 | 1 |
| TNFRSF4 | 0.156287477 | 0.007898719 | 0.009 | 0.004 | 1 |
| IL11RA | 0.156319834 | 0.049425142 | 0.422 | 0.404 | 1 |
| CSF2 | 0.158371503 | 0.018117679 | 0.012 | 0.01 | 1 |

| | | | | | |
|---|---|---|---|---|---|
| CXCL3 | 0.158738347 | 0.036796057 | 0.134 | 0.127 | 1 |
| CD80 | 0.173666622 | 0.005115308 | 0.006 | 0.002 | 1 |
| B4GALT5 | 0.179348326 | -0.005382802 | 0.001 | 0.004 | 1 |
| LGALS1 | 0.185344845 | -0.049306578 | 0.418 | 0.419 | 1 |
| LIG4 | 0.188554398 | 0.021991641 | 0.086 | 0.073 | 1 |
| CHD1 | 0.18983672 | -0.007651988 | 0.006 | 0.011 | 1 |
| IGKC | 0.194876741 | -0.059056123 | 0.625 | 0.582 | 1 |
| CD9 | 0.195077646 | -0.0274335 | 0.11 | 0.133 | 1 |
| FCER1G | 0.195207562 | 0.04533939 | 0.444 | 0.407 | 1 |
| SLC25A37 | 0.209203324 | -0.035614185 | 0.2 | 0.224 | 1 |
| IL3RA | 0.211046665 | -0.014550806 | 0.025 | 0.037 | 1 |
| CD79A | 0.214860106 | -0.037425272 | 0.156 | 0.161 | 1 |
| LTB | 0.220804026 | 0.039892161 | 0.31 | 0.299 | 1 |
| B3GAT1 | 0.224976398 | 0.005869336 | 0.007 | 0.004 | 1 |
| ZBTB16 | 0.226897434 | 0.005115308 | 0.003 | 0.002 | 1 |
| VSIG4 | 0.227933668 | 0.010165428 | 0.022 | 0.018 | 1 |
| FURIN | 0.231420948 | -0.039572368 | 0.278 | 0.257 | 1 |
| OSM | 0.232898105 | -0.007016299 | 0.005 | 0.01 | 1 |
| NRP1 | 0.235349751 | -0.011430063 | 0.021 | 0.027 | 1 |
| IGHG2 | 0.235989588 | -0.016544195 | 0.031 | 0.04 | 1 |
| CBLB | 0.243827774 | 0.009431711 | 0.021 | 0.016 | 1 |
| GAB2 | 0.24417857 | 0.017127721 | 0.059 | 0.049 | 1 |
| REL | 0.244594194 | -0.032278299 | 0.178 | 0.18 | 1 |
| IL15 | 0.248915058 | -0.006768686 | 0.003 | 0.008 | 1 |
| CXCL9 | 0.254732988 | -0.007263301 | 0.007 | 0.011 | 1 |
| LYN | 0.255609727 | 0.02276951 | 0.126 | 0.109 | 1 |
| IGHM | 0.258359597 | -0.04330562 | 0.225 | 0.228 | 1 |
| IGHD | 0.264574906 | -0.022644228 | 0.067 | 0.069 | 1 |
| TGFBI | 0.267055591 | 0.033229282 | 0.303 | 0.286 | 1 |
| SRGN | 0.268009398 | -0.018784274 | 0.062 | 0.072 | 1 |
| SDCBP | 0.271314956 | -0.036225602 | 0.672 | 0.69 | 1 |
| FUT4 | 0.273770946 | 0.014514944 | 0.051 | 0.04 | 1 |
| ICAM1 | 0.275427339 | 0.022570693 | 0.119 | 0.097 | 1 |
| KLRG1 | 0.275623479 | 0.013891285 | 0.041 | 0.033 | 1 |
| ZNF683 | 0.279489015 | 0.014252279 | 0.053 | 0.041 | 1 |
| CR2 | 0.282192221 | 0.013800052 | 0.036 | 0.03 | 1 |
| SNCA | 0.285553644 | 0.007212775 | 0.015 | 0.011 | 1 |
| IL15RA | 0.292168479 | -0.01642728 | 0.057 | 0.064 | 1 |
| HNRNPC | 0.293665 | -0.036295156 | 0.579 | 0.609 | 1 |
| CTSW | 0.295511218 | -0.013408391 | 0.039 | 0.049 | 1 |
| GEM | 0.296018607 | -0.006520459 | 0.003 | 0.005 | 1 |
| PCLAF | 0.302045275 | 0.007150024 | 0.003 | 0.002 | 1 |
| IL1R1 | 0.308995221 | -0.005992764 | 0.007 | 0.011 | 1 |
| THEMIS | 0.319753051 | 0.017146245 | 0.087 | 0.083 | 1 |
| FCER2 | 0.320498078 | -0.010749735 | 0.025 | 0.031 | 1 |
| DUSP4 | 0.321472557 | 0.021081065 | 0.047 | 0.041 | 1 |
| MAST4 | 0.323087051 | 0.019701941 | 0.118 | 0.104 | 1 |

| | | | | | |
|---|---|---|---|---|---|
| F5 | 0.334291973 | 0.017291898 | 0.05 | 0.047 | 1 |
| POU2F2 | 0.33647221 | 0.022956188 | 0.172 | 0.158 | 1 |
| IFRD1 | 0.339378759 | -0.023350087 | 0.17 | 0.167 | 1 |
| GZMB | 0.347804439 | 0.024209781 | 0.185 | 0.176 | 1 |
| FCGR3A | 0.360919514 | -0.021181952 | 0.148 | 0.162 | 1 |
| PIK3AP1 | 0.367625003 | -0.011488483 | 0.037 | 0.042 | 1 |
| IL1RN | 0.380661012 | 0.016649419 | 0.08 | 0.067 | 1 |
| CD244 | 0.381708333 | 0.004704794 | 0.01 | 0.007 | 1 |
| IL12A | 0.38640114 | -0.002822562 | 0.001 | 0.003 | 1 |
| PRDM1 | 0.387110054 | 0.014006101 | 0.066 | 0.052 | 1 |
| PTPN7 | 0.38840444 | 0.023194201 | 0.237 | 0.22 | 1 |
| KLRC4 | 0.389844589 | 0.0035721 | 0.007 | 0.004 | 1 |
| VPREB3 | 0.389889199 | -0.013191191 | 0.042 | 0.05 | 1 |
| GZMA | 0.394672426 | -0.011197399 | 0.038 | 0.048 | 1 |
| FOXP3 | 0.394856157 | 0.024590363 | 0.023 | 0.018 | 1 |
| CCL20 | 0.398636827 | -0.003968145 | 0.003 | 0.005 | 1 |
| TNFSF13B | 0.400173578 | -0.018621426 | 0.079 | 0.075 | 1 |
| CD33 | 0.402177885 | -0.007191142 | 0.019 | 0.023 | 1 |
| SNX8 | 0.406008023 | -0.014557446 | 0.065 | 0.074 | 1 |
| PTGDR2 | 0.411744363 | -0.006768686 | 0.003 | 0.004 | 1 |
| RAPH1 | 0.416242553 | -0.006210401 | 0.014 | 0.017 | 1 |
| BLNK | 0.418312251 | 0.007381882 | 0.025 | 0.02 | 1 |
| EIF3J | 0.420774627 | 0.011916047 | 0.059 | 0.052 | 1 |
| IL12RB2 | 0.421815395 | -0.01239799 | 0.029 | 0.028 | 1 |
| CXCL2 | 0.423913839 | 0.016581196 | 0.123 | 0.114 | 1 |
| TLR7 | 0.424501622 | -0.003076276 | 0.002 | 0.004 | 1 |
| HLA.DMA | 0.427456563 | 0.023323723 | 0.253 | 0.236 | 1 |
| CTSD | 0.428365234 | 0.023164172 | 0.314 | 0.312 | 1 |
| CSF1 | 0.434751509 | -0.005219987 | 0.009 | 0.012 | 1 |
| CD8A | 0.436525478 | -0.011544854 | 0.052 | 0.059 | 1 |
| TCF4 | 0.446778304 | -0.003968145 | 0.003 | 0.004 | 1 |
| FTH1 | 0.447455888 | -0.035452908 | 0.987 | 0.972 | 1 |
| LGALS3 | 0.464815678 | -0.015352482 | 0.086 | 0.092 | 1 |
| CXCL1 | 0.469682477 | 0.029530517 | 0.287 | 0.283 | 1 |
| YBX3 | 0.4702158 | 0.004704794 | 0.008 | 0.006 | 1 |
| MYC | 0.470442351 | -0.035021066 | 0.575 | 0.551 | 1 |
| TRIB1 | 0.473492342 | -0.006912722 | 0.023 | 0.026 | 1 |
| NT5E | 0.47505492 | -0.006456488 | 0.015 | 0.015 | 1 |
| NKG7 | 0.475308378 | 0.019971443 | 0.291 | 0.28 | 1 |
| TCL1A | 0.476119895 | 0.010376504 | 0.046 | 0.034 | 1 |
| CX3CR1 | 0.481580258 | -0.008361628 | 0.028 | 0.03 | 1 |
| DUSP1 | 0.482646379 | 0.021684241 | 0.234 | 0.219 | 1 |
| CXCR6 | 0.489911741 | -0.005716412 | 0.01 | 0.012 | 1 |
| IGHG1 | 0.490388754 | -0.024961647 | 0.222 | 0.213 | 1 |
| IGHG3 | 0.493109133 | -0.011172032 | 0.053 | 0.053 | 1 |
| CD6 | 0.494440982 | -0.023868948 | 0.832 | 0.829 | 1 |
| MCM4 | 0.495826403 | -0.007741248 | 0.027 | 0.032 | 1 |

| | | | | | |
|---|---|---|---|---|---|
| SOCS1 | 0.501954906 | 0.002813118 | 0.002 | 0.003 | 1 |
| CD209 | 0.510449942 | 0.002552794 | 0.006 | 0.004 | 1 |
| IL12RB1 | 0.513918036 | -0.006531264 | 0.023 | 0.026 | 1 |
| CCR4 | 0.515030299 | 0.029369779 | 0.176 | 0.12 | 1 |
| IL2RA | 0.515256147 | 0.006949614 | 0.013 | 0.007 | 1 |
| CCL13 | 0.518177134 | -0.005826077 | 0.013 | 0.016 | 1 |
| CD163 | 0.518892467 | -0.010899768 | 0.082 | 0.087 | 1 |
| CCR8 | 0.528244011 | -0.002181791 | 0.001 | 0.002 | 1 |
| CCR5 | 0.530782368 | -0.006043146 | 0.021 | 0.024 | 1 |
| IL2 | 0.53654124 | 0.015130034 | 0.024 | 0.021 | 1 |
| CXCL13 | 0.547274426 | 0.002932466 | 0.005 | 0.005 | 1 |
| CD70 | 0.549366505 | -0.002690293 | 0.003 | 0.004 | 1 |
| TNFRSF17 | 0.550167362 | -0.001540736 | 0.001 | 0.002 | 1 |
| LILRB4 | 0.571840856 | 0.009364741 | 0.084 | 0.078 | 1 |
| MITF | 0.572333256 | -0.005171073 | 0.02 | 0.023 | 1 |
| PGM3 | 0.574860018 | -0.010847038 | 0.075 | 0.084 | 1 |
| AURKB | 0.574918669 | 0.002932466 | 0.007 | 0.004 | 1 |
| CLEC4E | 0.575494929 | -0.004677735 | 0.017 | 0.021 | 1 |
| CLEC10A | 0.579612175 | 0.005237343 | 0.027 | 0.022 | 1 |
| STAT5A | 0.580640328 | -0.017217039 | 0.389 | 0.396 | 1 |
| BCL2A1 | 0.58463685 | 0.014422167 | 0.153 | 0.136 | 1 |
| CCND2 | 0.591465088 | 0.016135824 | 0.265 | 0.246 | 1 |
| TGFB3 | 0.596152177 | -0.003063789 | 0.008 | 0.01 | 1 |
| IKZF2 | 0.597084101 | -0.001540736 | 0.001 | 0.002 | 1 |
| QPCT | 0.59713998 | -0.00255862 | 0.006 | 0.008 | 1 |
| GIMAP5 | 0.604581703 | 0.018066481 | 0.865 | 0.833 | 1 |
| AQP9 | 0.608447533 | 0.005898494 | 0.041 | 0.034 | 1 |
| CD8B | 0.610560424 | -0.00255862 | 0.006 | 0.007 | 1 |
| GZMH | 0.615753789 | 0.005424146 | 0.037 | 0.033 | 1 |
| ICOS | 0.616057184 | -0.014764265 | 0.25 | 0.269 | 1 |
| SLA | 0.616300248 | 0.016067754 | 0.373 | 0.355 | 1 |
| CXCL8 | 0.619155467 | 0.021554077 | 0.433 | 0.431 | 1 |
| BCL10 | 0.619187066 | -0.008686924 | 0.075 | 0.08 | 1 |
| IRF4 | 0.631723935 | -0.011457587 | 0.081 | 0.072 | 1 |
| JAK3 | 0.638140331 | 0.015880559 | 0.75 | 0.74 | 1 |
| BCL2L1 | 0.644355862 | 0.004010518 | 0.022 | 0.017 | 1 |
| GNLY | 0.647219027 | 0.015451133 | 0.385 | 0.377 | 1 |
| STIM1 | 0.647603947 | 0.010495359 | 0.193 | 0.186 | 1 |
| CXCL11 | 0.652533779 | -0.001540736 | 0.001 | 0.001 | 1 |
| TREM1 | 0.656404694 | 0.001912876 | 0.005 | 0.004 | 1 |
| HLA.DQB1 | 0.658809288 | -0.014826354 | 0.226 | 0.212 | 1 |
| BCL11B | 0.659285618 | 0.016108912 | 0.719 | 0.707 | 1 |
| TMEM97 | 0.663020016 | -0.004768384 | 0.025 | 0.027 | 1 |
| FOSL1 | 0.664372995 | -0.002050942 | 0.003 | 0.004 | 1 |
| TGFB1 | 0.667880309 | 0.003637385 | 0.02 | 0.016 | 1 |
| CD109 | 0.66863926 | -0.003301675 | 0.015 | 0.016 | 1 |
| FBXO11 | 0.669939221 | -0.011992364 | 0.278 | 0.274 | 1 |

| | | | | | |
|---|---|---|---|---|---|
| LIPA | 0.671584727 | -0.013344627 | 0.462 | 0.482 | 1 |
| MGST1 | 0.671704852 | -0.003419388 | 0.019 | 0.021 | 1 |
| BTLA | 0.677799939 | 0.006250773 | 0.056 | 0.049 | 1 |
| PLCG2 | 0.680775113 | -0.00474675 | 0.032 | 0.033 | 1 |
| TNFSF14 | 0.684390898 | -0.001920682 | 0.006 | 0.007 | 1 |
| CD7 | 0.697968734 | -0.011720247 | 0.26 | 0.269 | 1 |
| BLK | 0.699604655 | -0.000899396 | 0.001 | 0.002 | 1 |
| APOE | 0.699604655 | -0.000899396 | 0.001 | 0.002 | 1 |
| EGR3 | 0.699604655 | -0.000899396 | 0.001 | 0.002 | 1 |
| KIT | 0.699604655 | -0.000899396 | 0.001 | 0.002 | 1 |
| IFNGR1 | 0.706428714 | -0.009874984 | 0.216 | 0.223 | 1 |
| VMO1 | 0.708388452 | -0.008133843 | 0.091 | 0.095 | 1 |
| FCN1 | 0.713635392 | 0.007271927 | 0.109 | 0.093 | 1 |
| FASLG | 0.71457193 | -0.002297048 | 0.01 | 0.012 | 1 |
| TIMM9 | 0.716389696 | 0.011665242 | 0.405 | 0.393 | 1 |
| CCL3 | 0.726726897 | 0.014038272 | 0.288 | 0.272 | 1 |
| CD86 | 0.729792978 | 0.004629355 | 0.046 | 0.036 | 1 |
| LAT2 | 0.740599522 | -0.003536291 | 0.019 | 0.019 | 1 |
| CD38 | 0.757031661 | -0.003751676 | 0.03 | 0.032 | 1 |
| JAK2 | 0.75765398 | 0.001273242 | 0.006 | 0.005 | 1 |
| CMKLR1 | 0.7645836 | -0.001914842 | 0.012 | 0.013 | 1 |
| CD79B | 0.766336129 | 0.006017254 | 0.095 | 0.093 | 1 |
| IRF3 | 0.770785057 | 0.005402631 | 0.086 | 0.086 | 1 |
| ENTPD1 | 0.802485358 | 0.001639443 | 0.013 | 0.011 | 1 |
| IL1A | 0.807676684 | 0.001517925 | 0.009 | 0.008 | 1 |
| TGIF1 | 0.810091102 | -0.009099277 | 0.432 | 0.418 | 1 |
| LIF | 0.810675773 | 0.00089166 | 0.005 | 0.004 | 1 |
| JUNB | 0.815794379 | -0.007105379 | 0.245 | 0.229 | 1 |
| FOS | 0.817637082 | 0.005352457 | 0.148 | 0.137 | 1 |
| CD63 | 0.818820137 | 0.007859494 | 0.512 | 0.513 | 1 |
| FN1 | 0.823595896 | -0.006873297 | 0.282 | 0.297 | 1 |
| TLR2 | 0.828269051 | -0.001903359 | 0.023 | 0.024 | 1 |
| DCLRE1C | 0.828724962 | 0.004568817 | 0.105 | 0.103 | 1 |
| NDFIP2 | 0.829313844 | -0.000771389 | 0.003 | 0.004 | 1 |
| IL1B | 0.829434892 | 0.007472642 | 0.206 | 0.19 | 1 |
| CCR1 | 0.829760077 | -0.003724057 | 0.079 | 0.083 | 1 |
| CCR6 | 0.829916809 | -0.002159942 | 0.016 | 0.015 | 1 |
| ITGAM | 0.837150686 | -0.001903359 | 0.022 | 0.023 | 1 |
| KCNE3 | 0.837278448 | 0.001504148 | 0.015 | 0.015 | 1 |
| MZB1 | 0.8381697 | -0.001657578 | 0.016 | 0.015 | 1 |
| HLA.DRA | 0.847782046 | 0.00842321 | 0.648 | 0.62 | 1 |
| THBS1 | 0.849900501 | -0.006843078 | 0.442 | 0.435 | 1 |
| RORC | 0.851157045 | 0.001015263 | 0.006 | 0.005 | 1 |
| CYBB | 0.859396806 | 0.00466502 | 0.259 | 0.253 | 1 |
| CXCL5 | 0.860015499 | -0.007291557 | 0.493 | 0.506 | 1 |
| TNF | 0.862535417 | 0.003238638 | 0.072 | 0.069 | 1 |
| CCL5 | 0.869057345 | -0.004419857 | 0.153 | 0.155 | 1 |

| | | | | | |
|---|---|---|---|---|---|
| HAVCR2 | 0.873148286 | 0.002460175 | 0.062 | 0.064 | 1 |
| KLRC3 | 0.875063805 | -0.002118403 | 0.049 | 0.051 | 1 |
| NFKBID | 0.876104633 | 0.000509299 | 0.003 | 0.003 | 1 |
| IL18 | 0.876104633 | 0.000509299 | 0.003 | 0.003 | 1 |
| BCAT1 | 0.877343928 | 0.000881395 | 0.01 | 0.01 | 1 |
| SLC7A1 | 0.885930076 | 0.00260939 | 0.087 | 0.084 | 1 |
| KLRF1 | 0.894122165 | 0.00111519 | 0.019 | 0.02 | 1 |
| SMAD2 | 0.894167253 | -0.003325283 | 0.233 | 0.224 | 1 |
| IL23A | 0.900284996 | 0.003058031 | 0.164 | 0.162 | 1 |
| ALAS2 | 0.900946214 | -0.00025777 | 0.001 | 0.001 | 1 |
| SPAG9 | 0.901010535 | 0.002727539 | 0.148 | 0.143 | 1 |
| CNOT2 | 0.907636772 | 0.003216713 | 0.341 | 0.341 | 1 |
| NIBAN3 | 0.92262331 | -0.000387892 | 0.005 | 0.005 | 1 |
| NGDN | 0.927572271 | -0.000899396 | 0.023 | 0.021 | 1 |
| CARD11 | 0.929839728 | -0.000517107 | 0.008 | 0.007 | 1 |
| ZNF341 | 0.937229122 | 0.000851296 | 0.022 | 0.024 | 1 |
| RBCK1 | 0.946761726 | 0.001894565 | 0.247 | 0.256 | 1 |
| CD74 | 0.948781569 | -0.002710998 | 0.904 | 0.869 | 1 |
| EEF1E1 | 0.949622829 | -0.000899396 | 0.046 | 0.047 | 1 |
| SLC7A7 | 0.950746813 | 0.00091114 | 0.061 | 0.057 | 1 |
| DUSP5 | 0.953340668 | 0.000822198 | 0.036 | 0.037 | 1 |
| TNFRSF13C | 0.955721493 | 0.001321844 | 0.133 | 0.118 | 1 |
| S100A9 | 0.960764087 | -0.001821808 | 0.341 | 0.33 | 1 |
| IGHG4 | 0.963205441 | 0.000126156 | 0.002 | 0.002 | 1 |
| IGHA1 | 0.964047051 | -0.000393719 | 0.015 | 0.013 | 1 |
| NFATC3 | 0.96472417 | -0.001318114 | 0.394 | 0.391 | 1 |
| CD14 | 0.971408683 | -0.001225281 | 0.378 | 0.379 | 1 |
| JCHAIN | 0.975504749 | 0.000462163 | 0.031 | 0.025 | 1 |
| LEF1 | 0.977620149 | 0.000949411 | 0.904 | 0.899 | 1 |
| LY86 | 0.979855477 | 0.000411008 | 0.067 | 0.071 | 1 |
| PSEN1 | 0.981812697 | -0.000440742 | 0.102 | 0.106 | 1 |
| CD1C | 0.982333809 | -0.000135594 | 0.009 | 0.008 | 1 |
| CD36 | 0.982891512 | -0.000450028 | 0.123 | 0.123 | 1 |
| IVNS1ABP | 0.993771965 | 0.000249094 | 0.457 | 0.451 | 1 |
| IL17RB | 0.996188075 | -0.005131869 | 0 | 0.002 | 1 |
| CXCR2 | 0.99632963 | -0.002568216 | 0 | 0.002 | 1 |

CD4+ Memory

| | p_val | avg_log2FC | Patient | Controls | p_val_adj |
|---|---|---|---|---|---|
| ITGA4 | 3.55E-49 | 1.053507483 | 0.745 | 0.438 | 1.77E-46 |
| CXCR4 | 5.23E-45 | -1.073017843 | 0.833 | 0.963 | 2.60E-42 |
| CD52 | 2.88E-41 | 0.812140134 | 0.993 | 0.953 | 1.43E-38 |
| LAT | 1.05E-36 | 0.761500074 | 0.911 | 0.731 | 5.21E-34 |
| CD4 | 4.50E-33 | 0.617619791 | 0.904 | 0.755 | 2.24E-30 |
| BTG1 | 1.54E-31 | -0.79167569 | 0.94 | 0.971 | 7.68E-29 |
| STAT1 | 4.34E-30 | -1.015821292 | 0.603 | 0.682 | 2.16E-27 |

| | | | | | |
|---|---|---|---|---|---|
| ITGB2 | 6.04E-30 | 0.614149537 | 0.883 | 0.706 | 3.01E-27 |
| NFKBIA | 3.74E-27 | -0.980750097 | 0.706 | 0.763 | 1.86E-24 |
| PASK | 1.17E-24 | 0.924598537 | 0.817 | 0.593 | 5.83E-22 |
| BIRC3 | 2.71E-24 | -0.833265746 | 0.509 | 0.638 | 1.35E-21 |
| RGS1 | 8.93E-24 | -1.184736702 | 0.106 | 0.388 | 4.45E-21 |
| GBP1 | 8.70E-22 | -0.82507885 | 0.367 | 0.463 | 4.33E-19 |
| TRIB2 | 3.83E-21 | 0.583057155 | 0.661 | 0.386 | 1.91E-18 |
| FTH1 | 1.64E-20 | -0.664258574 | 0.993 | 0.99 | 8.17E-18 |
| IRF7 | 2.05E-20 | -0.625683766 | 0.319 | 0.495 | 1.02E-17 |
| IFI6 | 4.71E-20 | -0.742946604 | 0.117 | 0.297 | 2.34E-17 |
| IRF1 | 3.56E-18 | -0.711827491 | 0.619 | 0.714 | 1.77E-15 |
| RPL6 | 6.82E-18 | 0.364487599 | 1 | 1 | 3.40E-15 |
| LAMP3 | 1.25E-17 | -0.878702636 | 0.041 | 0.156 | 6.24E-15 |
| LCK | 4.52E-17 | 0.418082208 | 0.938 | 0.852 | 2.25E-14 |
| CD3E | 9.67E-17 | 0.372515489 | 0.993 | 0.961 | 4.82E-14 |
| FYB1 | 1.32E-16 | 0.455612074 | 0.876 | 0.726 | 6.56E-14 |
| CD69 | 1.19E-15 | -0.625650927 | 0.33 | 0.531 | 5.94E-13 |
| CREM | 1.24E-15 | -0.662055489 | 0.19 | 0.393 | 6.18E-13 |
| SELPLG | 2.27E-15 | 0.423765312 | 0.933 | 0.787 | 1.13E-12 |
| SNX9 | 4.13E-15 | -0.371281348 | 0.101 | 0.294 | 2.06E-12 |
| NFKB1 | 8.57E-15 | -0.594147907 | 0.475 | 0.542 | 4.27E-12 |
| CCR7 | 1.99E-14 | -0.776323164 | 0.702 | 0.718 | 9.92E-12 |
| PSME2 | 2.63E-13 | -0.531402922 | 0.766 | 0.779 | 1.31E-10 |
| CD74 | 5.20E-13 | -0.656079073 | 0.936 | 0.932 | 2.59E-10 |
| CCL22 | 4.01E-12 | -0.862566508 | 0.156 | 0.144 | 2.00E-09 |
| IFITM2 | 1.36E-11 | 0.428557355 | 0.931 | 0.849 | 6.77E-09 |
| CD83 | 7.12E-11 | -0.600001183 | 0.154 | 0.187 | 3.55E-08 |
| WAS | 9.64E-11 | 0.323134776 | 0.592 | 0.401 | 4.80E-08 |
| GNAI2 | 2.99E-10 | 0.289176922 | 0.998 | 0.985 | 1.49E-07 |
| NFKBIZ | 3.32E-10 | -0.441301348 | 0.278 | 0.411 | 1.65E-07 |
| CD27 | 3.45E-10 | 0.38584491 | 0.608 | 0.438 | 1.72E-07 |
| STAT3 | 1.29E-09 | -0.38497467 | 0.553 | 0.675 | 6.41E-07 |
| HLA.DRA | 1.88E-09 | -0.61524207 | 0.706 | 0.653 | 9.36E-07 |
| FOS | 2.01E-09 | -0.488317957 | 0.213 | 0.252 | 9.99E-07 |
| CSRNP1 | 1.08E-08 | -0.285759456 | 0.144 | 0.274 | 5.38E-06 |
| ZAP70 | 1.37E-08 | 0.261431739 | 0.358 | 0.222 | 6.83E-06 |
| HIF1A | 2.05E-08 | -0.339905643 | 0.397 | 0.453 | 1.02E-05 |
| TIGIT | 2.76E-08 | 0.307347152 | 0.278 | 0.131 | 1.38E-05 |
| DUSP5 | 3.40E-08 | -0.389389017 | 0.128 | 0.175 | 1.70E-05 |
| JUN | 4.41E-08 | -0.218956036 | 0.055 | 0.157 | 2.20E-05 |
| PFN1 | 4.48E-08 | 0.292484277 | 1 | 1 | 2.23E-05 |
| TUBA1C | 4.60E-08 | -0.40828838 | 0.495 | 0.526 | 2.29E-05 |
| TRAC | 4.69E-08 | 0.299434014 | 0.998 | 0.974 | 2.34E-05 |
| CXCL16 | 4.95E-08 | -0.342635885 | 0.17 | 0.237 | 2.46E-05 |
| EIF5A | 9.20E-08 | -0.332211679 | 0.658 | 0.711 | 4.58E-05 |
| CD3D | 1.23E-07 | 0.260285673 | 0.982 | 0.914 | 6.13E-05 |
| BIN2 | 1.96E-07 | 0.233428899 | 0.388 | 0.253 | 9.78E-05 |

| | | | | | |
|---|---|---|---|---|---|
| ATF4 | 1.97E-07 | -0.353050352 | 0.862 | 0.836 | 9.80E-05 |
| POU2AF1 | 3.58E-07 | 0.138594082 | 0.124 | 0.044 | 0.000178433 |
| NAMPT | 3.98E-07 | -0.291342023 | 0.165 | 0.244 | 0.000198364 |
| IL2RB | 4.89E-07 | -0.369466948 | 0.516 | 0.508 | 0.000243601 |
| GAPDH | 6.26E-07 | 0.296228808 | 0.982 | 0.946 | 0.000311745 |
| IRF4 | 7.25E-07 | -0.300450031 | 0.069 | 0.122 | 0.000360857 |
| GBP2 | 2.52E-06 | -0.287930593 | 0.351 | 0.419 | 0.001256042 |
| CD37 | 2.53E-06 | 0.255580294 | 0.878 | 0.768 | 0.001259949 |
| LGALS1 | 2.91E-06 | -0.397336159 | 0.583 | 0.664 | 0.001449485 |
| PRNP | 3.10E-06 | -0.215871018 | 0.2 | 0.317 | 0.001541953 |
| IRF9 | 3.38E-06 | -0.298255842 | 0.638 | 0.685 | 0.001683068 |
| RUNX3 | 3.40E-06 | -0.259741687 | 0.615 | 0.682 | 0.001691421 |
| TSPAN32 | 3.71E-06 | 0.132867636 | 0.133 | 0.057 | 0.001847038 |
| JUNB | 3.75E-06 | -0.241479351 | 0.222 | 0.305 | 0.001866312 |
| FURIN | 4.77E-06 | -0.341058831 | 0.333 | 0.391 | 0.002375324 |
| CD5 | 5.16E-06 | 0.281336482 | 0.745 | 0.664 | 0.002567581 |
| LAP3 | 6.89E-06 | -0.29860393 | 0.284 | 0.302 | 0.003429314 |
| PSMB9 | 9.51E-06 | -0.266837494 | 0.624 | 0.636 | 0.004734106 |
| CD86 | 9.97E-06 | -0.346554739 | 0.032 | 0.054 | 0.004962733 |
| AHI1 | 1.98E-05 | 0.132823294 | 0.096 | 0.039 | 0.009858942 |
| PIK3IP1 | 2.11E-05 | -0.256988183 | 0.743 | 0.792 | 0.010501083 |
| CXCL10 | 2.38E-05 | -0.297306179 | 0.069 | 0.083 | 0.011842174 |
| CD226 | 2.48E-05 | 0.176674121 | 0.25 | 0.133 | 0.012332946 |
| EVI2B | 2.58E-05 | 0.236641044 | 0.894 | 0.807 | 0.012867051 |
| IGKC | 3.38E-05 | 0.282165399 | 0.601 | 0.537 | 0.016853678 |
| CD247 | 3.75E-05 | 0.207622709 | 0.514 | 0.404 | 0.018654306 |
| IL6R | 3.87E-05 | 0.152494309 | 0.259 | 0.143 | 0.019287376 |
| TNFSF10 | 4.27E-05 | 0.243259373 | 0.557 | 0.442 | 0.021272149 |
| IL4R | 5.08E-05 | -0.24555888 | 0.346 | 0.455 | 0.02529608 |
| TNFAIP8 | 5.94E-05 | 0.221443991 | 0.764 | 0.635 | 0.029562241 |
| DOCK8 | 6.25E-05 | 0.202415891 | 0.766 | 0.615 | 0.031137115 |
| VSIR | 7.74E-05 | 0.184405406 | 0.415 | 0.338 | 0.038556745 |
| NFKB2 | 8.27E-05 | -0.121932523 | 0.037 | 0.096 | 0.041191323 |
| ICAM1 | 0.000188088 | -0.176105455 | 0.096 | 0.133 | 0.093667592 |
| CXCL1 | 0.00020457 | 0.303710299 | 0.296 | 0.284 | 0.101875661 |
| XBP1 | 0.00024671 | -0.207399477 | 0.394 | 0.476 | 0.122861795 |
| DUSP4 | 0.000346249 | -0.183826094 | 0.099 | 0.157 | 0.172432167 |
| ARID5A | 0.000374758 | -0.162674447 | 0.179 | 0.276 | 0.186629448 |
| TGIF1 | 0.000432601 | -0.213372699 | 0.443 | 0.508 | 0.215435345 |
| IL32 | 0.000432844 | 0.253512779 | 0.89 | 0.773 | 0.215556161 |
| SOCS3 | 0.000435849 | -0.150789897 | 0.099 | 0.159 | 0.217052647 |
| HLA.A | 0.000480219 | -0.144106298 | 1 | 1 | 0.239149019 |
| ARHGEF3 | 0.000501461 | 0.170482287 | 0.447 | 0.372 | 0.249727779 |
| MYC | 0.00055236 | 0.272678612 | 0.507 | 0.377 | 0.275075397 |
| ARPC1B | 0.000574663 | 0.19088589 | 0.828 | 0.75 | 0.286181951 |
| STAT5A | 0.000756296 | -0.181205436 | 0.31 | 0.388 | 0.37663538 |
| BCL2A1 | 0.000770076 | -0.255299098 | 0.195 | 0.151 | 0.383497756 |

| | | | | | |
|---|---|---|---|---|---|
| IL15RA | 0.000939347 | -0.132601316 | 0.092 | 0.136 | 0.467794916 |
| CXCL9 | 0.000969646 | -0.383118828 | 0.023 | 0.028 | 0.482883837 |
| NFATC2 | 0.001013034 | -0.134792191 | 0.197 | 0.281 | 0.50449084 |
| SDCBP | 0.001145371 | -0.180981156 | 0.651 | 0.687 | 0.570394853 |
| HLA.DPA1 | 0.00128063 | -0.319790978 | 0.693 | 0.518 | 0.637753693 |
| REL | 0.001403365 | -0.147988942 | 0.177 | 0.224 | 0.698875873 |
| CCL3 | 0.001516207 | 0.231444462 | 0.305 | 0.297 | 0.755070956 |
| STAT6 | 0.001611955 | 0.152900152 | 0.339 | 0.242 | 0.802753561 |
| IGBP1 | 0.001671601 | 0.144389909 | 0.353 | 0.276 | 0.832457075 |
| DUSP2 | 0.001710636 | 0.287098111 | 0.491 | 0.438 | 0.851896746 |
| CCL20 | 0.001747029 | -0.318974743 | 0.009 | 0.019 | 0.870020197 |
| VPS28 | 0.002167838 | 0.150469051 | 0.693 | 0.602 | 1 |
| THBS1 | 0.002219277 | -0.196428516 | 0.454 | 0.472 | 1 |
| PTPRC | 0.002241603 | 0.134686146 | 0.998 | 0.974 | 1 |
| CCR4 | 0.002523796 | -0.246932657 | 0.287 | 0.357 | 1 |
| PSMB8 | 0.002537825 | -0.164466666 | 0.862 | 0.857 | 1 |
| CD3G | 0.002578017 | 0.079199524 | 0.131 | 0.068 | 1 |
| IL1B | 0.002848075 | 0.176674121 | 0.248 | 0.209 | 1 |
| SPAG9 | 0.002932872 | -0.108895199 | 0.106 | 0.154 | 1 |
| JAK3 | 0.002996585 | 0.159800302 | 0.663 | 0.594 | 1 |
| STAT4 | 0.003167584 | -0.146361377 | 0.241 | 0.299 | 1 |
| CD300A | 0.003376237 | 0.118571166 | 0.266 | 0.192 | 1 |
| RAPH1 | 0.003675991 | -0.050791271 | 0.005 | 0.036 | 1 |
| IFNGR1 | 0.003972543 | -0.133413508 | 0.216 | 0.274 | 1 |
| NR4A2 | 0.004044322 | -0.311551339 | 0.037 | 0.123 | 1 |
| CD48 | 0.004061039 | 0.155973212 | 0.975 | 0.885 | 1 |
| GIMAP2 | 0.004111355 | 0.161349644 | 0.7 | 0.615 | 1 |
| TGFBR2 | 0.004496049 | -0.161401396 | 0.635 | 0.648 | 1 |
| CXCL2 | 0.005467605 | 0.128938354 | 0.126 | 0.11 | 1 |
| RORC | 0.006092576 | 0.088758276 | 0.071 | 0.041 | 1 |
| CCL5 | 0.006199233 | -0.206928856 | 0.234 | 0.295 | 1 |
| RBCK1 | 0.006390536 | -0.118974649 | 0.234 | 0.276 | 1 |
| PRF1 | 0.007161387 | 0.15126041 | 0.376 | 0.312 | 1 |
| CHI3L2 | 0.007292199 | 0.061573144 | 0.069 | 0.034 | 1 |
| BAX | 0.007610249 | -0.154825952 | 0.385 | 0.406 | 1 |
| LDHA | 0.008151562 | -0.168727309 | 0.892 | 0.877 | 1 |
| CXCL8 | 0.008531949 | 0.212930006 | 0.463 | 0.45 | 1 |
| IL11RA | 0.008656759 | 0.136701135 | 0.42 | 0.336 | 1 |
| IL23A | 0.009882746 | -0.117447963 | 0.138 | 0.156 | 1 |
| FOXO1 | 0.010822335 | -0.099501384 | 0.144 | 0.195 | 1 |
| CD70 | 0.011090733 | -0.061058846 | 0.011 | 0.028 | 1 |
| DUSP1 | 0.011718164 | -0.171134039 | 0.369 | 0.419 | 1 |
| CYCS | 0.012532603 | -0.12450886 | 0.323 | 0.381 | 1 |
| NINJ2 | 0.013486286 | 0.041744541 | 0.05 | 0.023 | 1 |
| TNFSF13B | 0.014847928 | -0.14222534 | 0.119 | 0.148 | 1 |
| CXCL5 | 0.015001194 | 0.160962874 | 0.509 | 0.464 | 1 |
| CTSW | 0.015062728 | -0.064188904 | 0.05 | 0.083 | 1 |

| | | | | | |
|---|---|---|---|---|---|
| ARL4C | 0.015181067 | -0.141544134 | 0.693 | 0.756 | 1 |
| EIF5 | 0.015231798 | -0.145107127 | 0.486 | 0.516 | 1 |
| CD8A | 0.015267734 | 0.06759108 | 0.112 | 0.078 | 1 |
| IL1R1 | 0.015302363 | -0.053345228 | 0.014 | 0.031 | 1 |
| TNFRSF13C | 0.015340133 | -0.075227542 | 0.085 | 0.13 | 1 |
| IL12RB2 | 0.015546462 | -0.133114054 | 0.117 | 0.13 | 1 |
| LGALS3 | 0.018269186 | -0.112291046 | 0.174 | 0.213 | 1 |
| CD2 | 0.019256951 | 0.131559353 | 0.947 | 0.875 | 1 |
| LTA | 0.020528555 | -0.108055356 | 0.076 | 0.11 | 1 |
| IGHG1 | 0.020584217 | 0.114709819 | 0.225 | 0.208 | 1 |
| IFITM3 | 0.021851895 | -0.120561769 | 0.177 | 0.174 | 1 |
| FOSB | 0.022605138 | -0.13134774 | 0.009 | 0.016 | 1 |
| NT5E | 0.023408787 | -0.073421229 | 0.028 | 0.034 | 1 |
| IL6ST | 0.023466362 | -0.14085824 | 0.378 | 0.42 | 1 |
| TRAF1 | 0.023563489 | -0.102087158 | 0.163 | 0.177 | 1 |
| IER3 | 0.023849481 | -0.162289345 | 0.28 | 0.258 | 1 |
| IL2RA | 0.026186179 | -0.113439819 | 0.06 | 0.078 | 1 |
| CXCR3 | 0.026290017 | 0.121853201 | 0.275 | 0.209 | 1 |
| HLA.DQB1 | 0.026633771 | -0.198972801 | 0.333 | 0.281 | 1 |
| PDIA6 | 0.027698076 | 0.123228934 | 0.571 | 0.5 | 1 |
| SLC1A5 | 0.028730721 | 0.098361719 | 0.255 | 0.203 | 1 |
| SRGN | 0.029234262 | -0.087343382 | 0.099 | 0.13 | 1 |
| KDELR1 | 0.029684774 | 0.098064286 | 0.399 | 0.347 | 1 |
| LAG3 | 0.03261484 | -0.111451266 | 0.089 | 0.112 | 1 |
| VMO1 | 0.035049151 | -0.071284964 | 0.096 | 0.122 | 1 |
| PTPN7 | 0.035388019 | 0.100204982 | 0.351 | 0.295 | 1 |
| CASP5 | 0.035595418 | 0.071390003 | 0.183 | 0.144 | 1 |
| IL3RA | 0.036092804 | 0.052570338 | 0.06 | 0.041 | 1 |
| CD7 | 0.03619435 | -0.09804209 | 0.163 | 0.209 | 1 |
| IL15 | 0.036833403 | -0.040943485 | 0.011 | 0.024 | 1 |
| FOXP1 | 0.036890543 | -0.06790338 | 0.064 | 0.106 | 1 |
| LEF1 | 0.037157054 | 0.131031455 | 0.768 | 0.653 | 1 |
| CSF1 | 0.037456888 | -0.047011617 | 0.021 | 0.039 | 1 |
| TNFRSF25 | 0.038527147 | 0.07805824 | 0.151 | 0.107 | 1 |
| LRRC8B | 0.03898907 | 0.037171811 | 0.046 | 0.026 | 1 |
| SLA | 0.042341805 | 0.106889824 | 0.509 | 0.432 | 1 |
| IFRD1 | 0.045182898 | -0.092685923 | 0.167 | 0.206 | 1 |
| DPP4 | 0.045620284 | -0.086360285 | 0.071 | 0.093 | 1 |
| ENTPD1 | 0.050886409 | -0.031912501 | 0.009 | 0.028 | 1 |
| LIPA | 0.0538387 | -0.105830149 | 0.413 | 0.505 | 1 |
| NANS | 0.063234541 | -0.086360285 | 0.183 | 0.216 | 1 |
| IL12RB1 | 0.063669294 | -0.038770826 | 0.034 | 0.06 | 1 |
| PTGDR2 | 0.064007638 | -0.053193421 | 0.005 | 0.016 | 1 |
| CD40 | 0.064319221 | -0.078186353 | 0.078 | 0.065 | 1 |
| CXCL11 | 0.065069665 | -0.026824521 | 0.002 | 0.011 | 1 |
| GRB10 | 0.065149672 | -0.021231473 | 0.005 | 0.018 | 1 |
| BACH2 | 0.06542599 | -0.041171511 | 0.037 | 0.065 | 1 |

| | | | | | |
|---|---|---|---|---|---|
| CHD7 | 0.0659863 | -0.029907016 | 0.016 | 0.032 | 1 |
| TIAF1 | 0.067814594 | 0.067967862 | 0.17 | 0.128 | 1 |
| IRF8 | 0.068022705 | -0.07967892 | 0.135 | 0.144 | 1 |
| EEF1E1 | 0.068538863 | 0.039728808 | 0.071 | 0.047 | 1 |
| BCL6 | 0.073380868 | -0.032657118 | 0.021 | 0.047 | 1 |
| IL21 | 0.076934622 | 0.015041747 | 0.014 | 0.003 | 1 |
| BATF | 0.077516234 | -0.068438377 | 0.096 | 0.112 | 1 |
| RORA | 0.077603176 | -0.113268158 | 0.55 | 0.547 | 1 |
| JAK2 | 0.078990086 | -0.017620694 | 0.002 | 0.015 | 1 |
| ORAI1 | 0.079539419 | 0.086598777 | 0.445 | 0.367 | 1 |
| ERBIN | 0.081035103 | -0.088902479 | 0.498 | 0.506 | 1 |
| IL7R | 0.082095654 | 0.118723271 | 0.69 | 0.657 | 1 |
| GIMAP5 | 0.082985808 | 0.104688008 | 0.766 | 0.696 | 1 |
| BCL11B | 0.085379636 | 0.103819423 | 0.656 | 0.584 | 1 |
| PAXBP1 | 0.08667934 | -0.081743657 | 0.323 | 0.367 | 1 |
| CXCL3 | 0.087052428 | 0.070692499 | 0.14 | 0.13 | 1 |
| ICOS | 0.089227275 | -0.084653963 | 0.232 | 0.237 | 1 |
| ILF2 | 0.092235741 | -0.094587489 | 0.55 | 0.534 | 1 |
| BTLA | 0.093034239 | 0.03691109 | 0.055 | 0.031 | 1 |
| PCNA | 0.095355991 | -0.077412049 | 0.284 | 0.308 | 1 |
| TRDC | 0.09706233 | -0.06699496 | 0.071 | 0.08 | 1 |
| CD28 | 0.098392409 | 0.067804291 | 0.25 | 0.19 | 1 |
| CX3CR1 | 0.106521218 | 0.023264206 | 0.034 | 0.016 | 1 |
| AURKB | 0.109956245 | 0.016893677 | 0.016 | 0.006 | 1 |
| MCM4 | 0.110565571 | -0.020543687 | 0.011 | 0.026 | 1 |
| S100A10 | 0.111733532 | 0.096375516 | 0.968 | 0.893 | 1 |
| SNX8 | 0.112468469 | 0.042173085 | 0.085 | 0.067 | 1 |
| PIK3AP1 | 0.115149891 | 0.030679136 | 0.057 | 0.034 | 1 |
| KLRB1 | 0.120200601 | -0.028910013 | 0.018 | 0.028 | 1 |
| FASLG | 0.1203892 | -0.019246089 | 0.007 | 0.023 | 1 |
| TRBC2 | 0.120733083 | 0.116384894 | 0.972 | 0.968 | 1 |
| KLRG1 | 0.123749293 | 0.035570435 | 0.076 | 0.057 | 1 |
| VNN2 | 0.124651667 | 0.032725605 | 0.064 | 0.045 | 1 |
| TGFBI | 0.132948738 | -0.07975755 | 0.344 | 0.393 | 1 |
| BLNK | 0.13608536 | -0.025664353 | 0.021 | 0.032 | 1 |
| STIM1 | 0.137313167 | 0.050774782 | 0.181 | 0.127 | 1 |
| FCN1 | 0.144113863 | -0.038714913 | 0.076 | 0.106 | 1 |
| MS4A1 | 0.144754576 | -0.053809713 | 0.135 | 0.172 | 1 |
| GATA3 | 0.146704511 | 0.032725605 | 0.06 | 0.044 | 1 |
| AQP9 | 0.153505524 | 0.031871482 | 0.06 | 0.041 | 1 |
| MYB | 0.153726425 | -0.019246089 | 0.009 | 0.018 | 1 |
| QPCT | 0.157164368 | -0.014323104 | 0.005 | 0.013 | 1 |
| FYN | 0.160363658 | -0.079775631 | 0.833 | 0.843 | 1 |
| LTB | 0.162303531 | 0.073695657 | 0.394 | 0.323 | 1 |
| CSF2 | 0.162413316 | -0.044793792 | 0.018 | 0.019 | 1 |
| ATF6B | 0.168455579 | 0.063178769 | 0.358 | 0.321 | 1 |
| LIF | 0.171318486 | 0.011774034 | 0.009 | 0.003 | 1 |

| | | | | | |
|---|---|---|---|---|---|
| ITK | 0.177375204 | -0.077032081 | 0.819 | 0.782 | 1 |
| TLR2 | 0.178818006 | -0.018573926 | 0.016 | 0.029 | 1 |
| CTSD | 0.179408306 | 0.066808488 | 0.319 | 0.292 | 1 |
| CD24 | 0.180616988 | -0.010679085 | 0.002 | 0.01 | 1 |
| IL2 | 0.18148538 | -0.066315205 | 0.032 | 0.032 | 1 |
| VPREB3 | 0.185332033 | -0.027732555 | 0.032 | 0.05 | 1 |
| SLC7A1 | 0.192502586 | -0.037974254 | 0.085 | 0.099 | 1 |
| IL23R | 0.197277879 | -0.015310533 | 0.002 | 0.006 | 1 |
| CD22 | 0.200644602 | 0.031151764 | 0.073 | 0.057 | 1 |
| CD200 | 0.202636919 | -0.054511419 | 0.048 | 0.024 | 1 |
| CD72 | 0.204874673 | 0.020473261 | 0.034 | 0.024 | 1 |
| CCL4 | 0.207659898 | 0.10609555 | 0.28 | 0.258 | 1 |
| CCL2 | 0.209986645 | 0.096429598 | 0.78 | 0.758 | 1 |
| CCR8 | 0.214883428 | -0.014323104 | 0.005 | 0.01 | 1 |
| LAT2 | 0.215409116 | 0.017762658 | 0.032 | 0.018 | 1 |
| TNF | 0.229646246 | -0.07038157 | 0.197 | 0.224 | 1 |
| S100A9 | 0.234066946 | -0.067887625 | 0.337 | 0.349 | 1 |
| LYN | 0.234305187 | -0.048735213 | 0.135 | 0.136 | 1 |
| FBXO11 | 0.236923241 | -0.049288698 | 0.234 | 0.274 | 1 |
| CD109 | 0.23800252 | -0.018573926 | 0.016 | 0.023 | 1 |
| CD1C | 0.24092154 | 0.009441462 | 0.011 | 0.005 | 1 |
| FOSL1 | 0.248624376 | -0.008357773 | 0.002 | 0.008 | 1 |
| CD9 | 0.249866087 | 0.039685019 | 0.142 | 0.112 | 1 |
| CD40LG | 0.25286709 | -0.040109654 | 0.106 | 0.104 | 1 |
| IGHA1 | 0.254025011 | -0.012359703 | 0.009 | 0.018 | 1 |
| CTLA4 | 0.254295265 | -0.073815031 | 0.257 | 0.242 | 1 |
| CD6 | 0.255462762 | -0.067758726 | 0.759 | 0.76 | 1 |
| CNOT2 | 0.256820558 | -0.047817609 | 0.239 | 0.281 | 1 |
| ADA | 0.257838311 | 0.057267973 | 0.278 | 0.205 | 1 |
| GZMK | 0.260331678 | -0.069308014 | 0.128 | 0.154 | 1 |
| IL1A | 0.260365147 | 0.016413763 | 0.014 | 0.011 | 1 |
| IGLC3 | 0.263951321 | -0.06997619 | 0.445 | 0.505 | 1 |
| SNCA | 0.264101804 | -0.013679145 | 0.011 | 0.019 | 1 |
| DUSP6 | 0.268844219 | 0.019089014 | 0.039 | 0.024 | 1 |
| GZMA | 0.270065004 | 0.042030251 | 0.117 | 0.091 | 1 |
| BCL2 | 0.272411498 | 0.050789681 | 0.151 | 0.153 | 1 |
| GEM | 0.275786316 | -0.052617859 | 0.007 | 0.018 | 1 |
| APOBEC3G | 0.277028261 | 0.058464827 | 0.456 | 0.317 | 1 |
| SKAP1 | 0.277160727 | 0.034655116 | 0.108 | 0.084 | 1 |
| TNFSF8 | 0.280932258 | 0.02304461 | 0.053 | 0.032 | 1 |
| CYBB | 0.28639247 | -0.048370699 | 0.264 | 0.307 | 1 |
| IKZF1 | 0.287552919 | 0.056984118 | 0.445 | 0.365 | 1 |
| CD79B | 0.290237095 | 0.035574774 | 0.149 | 0.117 | 1 |
| NFKBID | 0.291102549 | -0.008357773 | 0.002 | 0.006 | 1 |
| JCHAIN | 0.299834008 | 0.016811673 | 0.041 | 0.029 | 1 |
| ABCE1 | 0.303873254 | -0.046609636 | 0.3 | 0.313 | 1 |
| NFATC3 | 0.306241399 | 0.047515641 | 0.369 | 0.347 | 1 |

| | | | | | |
|---|---|---|---|---|---|
| NFATC1 | 0.306538262 | 0.046179656 | 0.314 | 0.282 | 1 |
| THEMIS | 0.30660828 | 0.02778726 | 0.096 | 0.073 | 1 |
| IL18RAP | 0.310128816 | 0.034281022 | 0.073 | 0.057 | 1 |
| SMAD2 | 0.311632791 | -0.041798244 | 0.181 | 0.211 | 1 |
| FN1 | 0.322292075 | -0.054917237 | 0.261 | 0.278 | 1 |
| LGALS9 | 0.332518062 | 0.04540754 | 0.225 | 0.182 | 1 |
| CCR6 | 0.333930123 | -0.056646061 | 0.239 | 0.256 | 1 |
| IL6 | 0.336572181 | 0.025430541 | 0.016 | 0.024 | 1 |
| MMP9 | 0.341771351 | -0.029855914 | 0.103 | 0.115 | 1 |
| IL2RG | 0.350166853 | 0.048601044 | 0.954 | 0.914 | 1 |
| SPP1 | 0.351954802 | -0.00603272 | 0.002 | 0.006 | 1 |
| ZNF341 | 0.354572669 | 0.009929729 | 0.018 | 0.011 | 1 |
| MCM2 | 0.356336279 | -0.021568832 | 0.062 | 0.07 | 1 |
| CST7 | 0.359944068 | 0.05547241 | 0.505 | 0.417 | 1 |
| LRRC32 | 0.361724023 | -0.014015644 | 0.014 | 0.021 | 1 |
| ITGAX | 0.363700663 | 0.024312639 | 0.092 | 0.086 | 1 |
| HNRNPC | 0.373977884 | -0.05086719 | 0.622 | 0.586 | 1 |
| TRIB1 | 0.380686686 | 0.019839119 | 0.057 | 0.047 | 1 |
| IVNS1ABP | 0.39241349 | -0.044307903 | 0.436 | 0.476 | 1 |
| KLRC3 | 0.395178488 | 0.019367222 | 0.064 | 0.052 | 1 |
| LILRB4 | 0.400814648 | -0.019097058 | 0.05 | 0.068 | 1 |
| IL18R1 | 0.405672327 | 0.029276602 | 0.131 | 0.093 | 1 |
| TLR7 | 0.410658505 | 0.005216318 | 0.007 | 0.003 | 1 |
| KLRC4 | 0.410658505 | 0.005216318 | 0.007 | 0.003 | 1 |
| IRF3 | 0.415496511 | -0.022969175 | 0.078 | 0.084 | 1 |
| CD274 | 0.420079949 | -0.033465336 | 0.133 | 0.102 | 1 |
| IGHG2 | 0.42016616 | 0.012719489 | 0.037 | 0.026 | 1 |
| FUT4 | 0.424009192 | -0.014374172 | 0.034 | 0.047 | 1 |
| IGHD | 0.432463175 | 0.019301492 | 0.078 | 0.058 | 1 |
| IFNG | 0.434977714 | 0.027097765 | 0.069 | 0.041 | 1 |
| CXCR6 | 0.438288711 | -0.05227929 | 0.083 | 0.094 | 1 |
| LIG4 | 0.447032198 | 0.019630411 | 0.094 | 0.08 | 1 |
| SP140 | 0.447643626 | 0.031424268 | 0.28 | 0.263 | 1 |
| IL17RB | 0.450177789 | 0.020554919 | 0.009 | 0.006 | 1 |
| PSME3 | 0.451923671 | -0.028179214 | 0.128 | 0.144 | 1 |
| ZNF683 | 0.45304419 | 0.01541734 | 0.05 | 0.039 | 1 |
| F13A1 | 0.455746303 | 0.01038037 | 0.005 | 0.006 | 1 |
| CCND2 | 0.463761068 | -0.041966165 | 0.317 | 0.294 | 1 |
| VSIG4 | 0.468212661 | -0.009442914 | 0.014 | 0.023 | 1 |
| CCL13 | 0.48114907 | 0.008994334 | 0.011 | 0.008 | 1 |
| TNIK | 0.483217906 | -0.028701516 | 0.144 | 0.161 | 1 |
| BCL2L1 | 0.485080348 | -0.013089088 | 0.032 | 0.039 | 1 |
| FCER2 | 0.49178863 | -0.009797948 | 0.021 | 0.026 | 1 |
| SOCS1 | 0.495184428 | -0.005060183 | 0.005 | 0.008 | 1 |
| EBF1 | 0.495184428 | -0.005060183 | 0.005 | 0.008 | 1 |
| UFM1 | 0.498775078 | -0.03607853 | 0.532 | 0.511 | 1 |
| S100A12 | 0.500050681 | 0.011356674 | 0.037 | 0.029 | 1 |

| | | | | | |
|---|---|---|---|---|---|
| TGFBR1 | 0.502512268 | -0.021050348 | 0.085 | 0.102 | 1 |
| MALT1 | 0.504705759 | 0.023557407 | 0.161 | 0.141 | 1 |
| CD44 | 0.506801424 | -0.033971603 | 0.961 | 0.935 | 1 |
| BCL10 | 0.51818716 | -0.01719826 | 0.073 | 0.076 | 1 |
| SELL | 0.529888539 | -0.060328995 | 0.567 | 0.584 | 1 |
| CR2 | 0.548126094 | -0.006781785 | 0.011 | 0.015 | 1 |
| CCR1 | 0.554323488 | -0.016571955 | 0.096 | 0.101 | 1 |
| MZB1 | 0.559729767 | -0.007151102 | 0.016 | 0.021 | 1 |
| CCR2 | 0.565421575 | -0.008478113 | 0.016 | 0.021 | 1 |
| TMEM97 | 0.566758076 | 0.007180483 | 0.023 | 0.018 | 1 |
| PDCD1 | 0.577859677 | 0.009965832 | 0.025 | 0.016 | 1 |
| MITF | 0.581352309 | -0.007517006 | 0.021 | 0.026 | 1 |
| TCL1A | 0.581579059 | -0.011185894 | 0.032 | 0.042 | 1 |
| CBLB | 0.583555579 | -0.008478113 | 0.016 | 0.019 | 1 |
| CD38 | 0.583555579 | -0.008478113 | 0.016 | 0.019 | 1 |
| ZBTB16 | 0.58476645 | 0.004787603 | 0.011 | 0.008 | 1 |
| TRAT1 | 0.596035688 | -0.021153155 | 0.163 | 0.183 | 1 |
| FOXP3 | 0.604238435 | -0.072071898 | 0.067 | 0.081 | 1 |
| CD63 | 0.606485698 | 0.029351536 | 0.674 | 0.654 | 1 |
| YBX3 | 0.60697606 | 0.006676751 | 0.014 | 0.008 | 1 |
| RNASE6 | 0.610259178 | 0.008602659 | 0.028 | 0.023 | 1 |
| CCR9 | 0.6304266 | 0.024913518 | 0.039 | 0.029 | 1 |
| FCER1G | 0.634837634 | -0.027574465 | 0.427 | 0.433 | 1 |
| TOP2A | 0.649347859 | 0.00674912 | 0.028 | 0.019 | 1 |
| FCGR3A | 0.651288872 | 0.015564617 | 0.156 | 0.157 | 1 |
| ANXA5 | 0.663803632 | 0.023693261 | 0.647 | 0.596 | 1 |
| PDIA4 | 0.673367593 | 0.023557407 | 0.484 | 0.43 | 1 |
| GZMH | 0.675980837 | -0.007648309 | 0.037 | 0.037 | 1 |
| CD79A | 0.676522141 | -0.014259236 | 0.14 | 0.148 | 1 |
| TNFRSF4 | 0.679796675 | -0.007517006 | 0.018 | 0.016 | 1 |
| BCAT1 | 0.683398507 | -0.004475318 | 0.011 | 0.013 | 1 |
| FAS | 0.684654441 | 0.0169971 | 0.197 | 0.166 | 1 |
| KLRK1 | 0.687853414 | -0.014305348 | 0.161 | 0.183 | 1 |
| MAP2K3 | 0.688109624 | 0.021498652 | 0.47 | 0.414 | 1 |
| CD244 | 0.688200189 | -0.00273513 | 0.005 | 0.006 | 1 |
| MAST4 | 0.690854147 | -0.012153913 | 0.119 | 0.12 | 1 |
| RPN2 | 0.694583146 | 0.019285923 | 0.589 | 0.567 | 1 |
| HLA.DMA | 0.695772975 | -0.023641794 | 0.296 | 0.237 | 1 |
| KLRF1 | 0.696337992 | 0.005305703 | 0.014 | 0.013 | 1 |
| CXCR5 | 0.725762389 | -0.017625108 | 0.128 | 0.128 | 1 |
| CD33 | 0.727346835 | 0.005393102 | 0.03 | 0.023 | 1 |
| NDFIP2 | 0.732097167 | -0.00273513 | 0.005 | 0.005 | 1 |
| CLEC10A | 0.734356911 | 0.005898223 | 0.034 | 0.029 | 1 |
| OSM | 0.735565699 | -0.008947422 | 0.03 | 0.026 | 1 |
| PRDM1 | 0.748300069 | -0.015199256 | 0.202 | 0.209 | 1 |
| KLRC1 | 0.751861349 | 0.004460943 | 0.028 | 0.024 | 1 |
| LY86 | 0.75937994 | -0.006489904 | 0.048 | 0.049 | 1 |

| | | | | | |
|---|---|---|---|---|---|
| ITGAE | 0.76107281 | -0.012012943 | 0.119 | 0.146 | 1 |
| KCNE3 | 0.764242673 | -0.003898125 | 0.016 | 0.021 | 1 |
| PSEN1 | 0.767697134 | 0.008932165 | 0.089 | 0.093 | 1 |
| PAK1IP1 | 0.780428145 | -0.005921669 | 0.055 | 0.058 | 1 |
| NKG7 | 0.781248139 | 0.013175389 | 0.328 | 0.33 | 1 |
| EVI2A | 0.78334991 | 0.015620447 | 0.599 | 0.571 | 1 |
| LAMP1 | 0.789399625 | -0.013581069 | 0.44 | 0.446 | 1 |
| EIF3J | 0.789667248 | 0.006612701 | 0.067 | 0.068 | 1 |
| POU2F2 | 0.792347751 | -0.009806782 | 0.147 | 0.157 | 1 |
| ADGRE1 | 0.795803416 | 0.007023162 | 0.055 | 0.057 | 1 |
| TCF7 | 0.796770918 | 0.010073651 | 0.186 | 0.161 | 1 |
| JAK1 | 0.800474917 | -0.011247026 | 0.938 | 0.945 | 1 |
| CD36 | 0.804406748 | -0.008782626 | 0.124 | 0.135 | 1 |
| DCLRE1C | 0.808053375 | 0.007213831 | 0.089 | 0.088 | 1 |
| TBX21 | 0.811689618 | -0.012087522 | 0.216 | 0.172 | 1 |
| PGM3 | 0.814104302 | -0.006072496 | 0.064 | 0.067 | 1 |
| GNLY | 0.816306185 | -0.015970957 | 0.413 | 0.406 | 1 |
| TIMM9 | 0.820696271 | 0.010971118 | 0.356 | 0.347 | 1 |
| CD209 | 0.821020261 | -0.001770113 | 0.007 | 0.008 | 1 |
| TGFB1 | 0.825518643 | 0.003525219 | 0.023 | 0.018 | 1 |
| CD163 | 0.828050614 | 0.00587928 | 0.094 | 0.081 | 1 |
| PLCG2 | 0.852863184 | -0.003146917 | 0.037 | 0.039 | 1 |
| CARD11 | 0.85515112 | 0.001512471 | 0.009 | 0.008 | 1 |
| HAVCR2 | 0.860956646 | 0.005006329 | 0.08 | 0.08 | 1 |
| F5 | 0.865807031 | -0.005164202 | 0.06 | 0.055 | 1 |
| NRP1 | 0.866976829 | 0.002702884 | 0.032 | 0.032 | 1 |
| IL1RN | 0.867516067 | -0.004259205 | 0.071 | 0.071 | 1 |
| B4GALT5 | 0.885896407 | 0.001512471 | 0.007 | 0.006 | 1 |
| TNFSF14 | 0.904415342 | 0.001642993 | 0.018 | 0.018 | 1 |
| CLEC4E | 0.910094329 | -0.001602667 | 0.018 | 0.015 | 1 |
| CMKLR1 | 0.914875133 | -0.001207604 | 0.011 | 0.015 | 1 |
| SLC7A7 | 0.918292353 | -0.002254652 | 0.064 | 0.067 | 1 |
| SLC25A37 | 0.919575973 | -0.003731387 | 0.133 | 0.135 | 1 |
| UGCG | 0.92815238 | -0.004043682 | 0.259 | 0.256 | 1 |
| CCR5 | 0.932178577 | -0.002823491 | 0.067 | 0.06 | 1 |
| SOCS2 | 0.936749178 | -0.003898125 | 0.083 | 0.062 | 1 |
| TGFB3 | 0.939548968 | 0.00055494 | 0.007 | 0.006 | 1 |
| GAB2 | 0.941726914 | -0.00229602 | 0.078 | 0.07 | 1 |
| GZMB | 0.943319428 | -0.003113582 | 0.193 | 0.174 | 1 |
| TREM1 | 0.947718379 | -0.000406324 | 0.005 | 0.005 | 1 |
| IGHG3 | 0.955184321 | 0.001102556 | 0.053 | 0.05 | 1 |
| IGHM | 0.961178497 | -0.002091368 | 0.218 | 0.235 | 1 |
| NGDN | 0.962672549 | -0.000652465 | 0.016 | 0.021 | 1 |
| MGST1 | 0.97499839 | -0.000499548 | 0.03 | 0.032 | 1 |
| CD14 | 0.980959022 | 0.001272615 | 0.401 | 0.404 | 1 |
| EGR1 | 0.99051429 | 0.000148708 | 0.009 | 0.008 | 1 |
| EIF1 | 0.993572069 | -0.000459116 | 0.972 | 0.955 | 1 |

| | | | | | |
|---|---|---|---|---|---|
| EGR3 | 0.99553413 | -0.013984229 | 0 | 0.006 | 1 |
| PCLAF | 0.995577738 | -0.011662918 | 0 | 0.008 | 1 |
| CCR3 | 0.995631111 | -0.009337865 | 0 | 0.005 | 1 |
| IGHG4 | 0.99569992 | -0.007009059 | 0 | 0.005 | 1 |
| NCAM1 | 0.99569992 | -0.007009059 | 0 | 0.005 | 1 |
| CD80 | 0.996119928 | -0.000104551 | 0.018 | 0.018 | 1 |
| TNFRSF8 | 0.996618246 | 0.013175389 | 0.007 | 0 | 1 |
| ITGAM | 0.998181497 | 4.14E-05 | 0.034 | 0.037 | 1 |
| IL22 | 0.999991026 | -0.493101643 | 0 | 0.006 | 1 |

CD8+ Naïve

| | p_val | avg_log2FC | Patient | Controls | p_val_adj |
|---|---|---|---|---|---|
| ITGA4 | 7.48E-64 | 0.699557073 | 0.733 | 0.498 | 3.72E-61 |
| STAT1 | 3.22E-42 | -0.81978999 | 0.634 | 0.709 | 1.60E-39 |
| PASK | 1.93E-36 | 0.699916108 | 0.878 | 0.705 | 9.62E-34 |
| TIGIT | 6.44E-34 | 0.350042499 | 0.234 | 0.066 | 3.21E-31 |
| SELPLG | 2.12E-33 | 0.4027842 | 0.918 | 0.803 | 1.06E-30 |
| CD52 | 3.66E-29 | 0.401587138 | 0.999 | 0.972 | 1.82E-26 |
| CD27 | 1.28E-28 | 0.422019885 | 0.807 | 0.626 | 6.36E-26 |
| BTG1 | 4.67E-24 | -0.431369936 | 0.965 | 0.98 | 2.33E-21 |
| IRF1 | 1.57E-23 | -0.517972981 | 0.551 | 0.646 | 7.81E-21 |
| S100A10 | 1.69E-22 | 0.352650202 | 0.94 | 0.865 | 8.40E-20 |
| HIF1A | 6.93E-22 | -0.377494495 | 0.398 | 0.532 | 3.45E-19 |
| ITGB2 | 9.06E-22 | 0.350267651 | 0.837 | 0.694 | 4.51E-19 |
| TRIB2 | 1.68E-21 | 0.42410371 | 0.553 | 0.371 | 8.36E-19 |
| IRF7 | 2.19E-21 | -0.391570559 | 0.333 | 0.471 | 1.09E-18 |
| CCL5 | 5.20E-20 | 0.629640964 | 0.352 | 0.217 | 2.59E-17 |
| ATF4 | 2.42E-17 | -0.363977315 | 0.815 | 0.86 | 1.20E-14 |
| PFN1 | 4.68E-17 | 0.237907256 | 1 | 0.999 | 2.33E-14 |
| KLRK1 | 3.51E-16 | 0.459952643 | 0.89 | 0.799 | 1.75E-13 |
| CCR7 | 5.91E-15 | -0.311448174 | 0.958 | 0.966 | 2.94E-12 |
| EVI2B | 1.25E-14 | 0.29748109 | 0.878 | 0.771 | 6.24E-12 |
| CD3E | 2.69E-14 | 0.22957578 | 0.995 | 0.97 | 1.34E-11 |
| PSME2 | 3.67E-13 | -0.292583057 | 0.743 | 0.792 | 1.83E-10 |
| IL6ST | 4.34E-13 | -0.282281512 | 0.454 | 0.605 | 2.16E-10 |
| CD226 | 1.12E-12 | 0.139679556 | 0.138 | 0.057 | 5.58E-10 |
| RPL6 | 2.40E-12 | 0.210050741 | 1 | 0.999 | 1.19E-09 |
| IFI6 | 2.64E-12 | -0.264590185 | 0.118 | 0.199 | 1.31E-09 |
| STAT3 | 5.14E-12 | -0.26817791 | 0.601 | 0.684 | 2.56E-09 |
| LCK | 6.88E-12 | 0.21536905 | 0.978 | 0.928 | 3.42E-09 |
| FYB1 | 7.47E-12 | 0.242447052 | 0.891 | 0.828 | 3.72E-09 |
| DOCK8 | 7.68E-12 | 0.220821969 | 0.754 | 0.667 | 3.82E-09 |
| TSPAN32 | 1.04E-11 | 0.171601505 | 0.232 | 0.128 | 5.19E-09 |
| GZMK | 1.12E-11 | 0.385473557 | 0.137 | 0.062 | 5.56E-09 |
| LAT | 3.13E-11 | 0.238891617 | 0.923 | 0.886 | 1.56E-08 |
| CXCR3 | 7.63E-11 | 0.224487453 | 0.221 | 0.107 | 3.80E-08 |

| | | | | | |
|---|---|---|---|---|---|
| GIMAP2 | 2.47E-10 | 0.235478184 | 0.795 | 0.703 | 1.23E-07 |
| CD300A | 2.63E-10 | 0.153875862 | 0.195 | 0.106 | 1.31E-07 |
| DUSP2 | 4.82E-10 | 0.323438588 | 0.357 | 0.275 | 2.40E-07 |
| PSMB8 | 5.35E-10 | -0.229963756 | 0.854 | 0.879 | 2.67E-07 |
| LAMP3 | 9.78E-10 | -0.20521274 | 0.067 | 0.119 | 4.87E-07 |
| IER3 | 1.24E-09 | 0.226972406 | 0.411 | 0.285 | 6.19E-07 |
| CTLA4 | 1.42E-09 | 0.149601822 | 0.139 | 0.056 | 7.08E-07 |
| GBP1 | 2.06E-09 | -0.276492384 | 0.422 | 0.441 | 1.02E-06 |
| CD48 | 2.11E-09 | 0.20670421 | 0.99 | 0.941 | 1.05E-06 |
| IRF9 | 2.80E-09 | -0.259745595 | 0.632 | 0.689 | 1.39E-06 |
| NFKBIA | 3.21E-09 | -0.293268017 | 0.669 | 0.734 | 1.60E-06 |
| IL32 | 5.65E-09 | 0.243195886 | 0.778 | 0.668 | 2.81E-06 |
| BIRC3 | 1.79E-08 | -0.274428477 | 0.414 | 0.476 | 8.91E-06 |
| ZAP70 | 2.89E-08 | 0.169087325 | 0.407 | 0.321 | 1.44E-05 |
| IL2RB | 5.51E-08 | 0.209672571 | 0.454 | 0.326 | 2.75E-05 |
| EIF5A | 6.44E-08 | -0.203021224 | 0.684 | 0.746 | 3.21E-05 |
| NFKB1 | 1.21E-07 | -0.226938468 | 0.593 | 0.627 | 6.05E-05 |
| ARPC1B | 1.27E-07 | 0.183686329 | 0.792 | 0.735 | 6.33E-05 |
| WAS | 1.67E-07 | 0.17614029 | 0.535 | 0.45 | 8.33E-05 |
| TNFAIP8 | 1.72E-07 | 0.182509168 | 0.744 | 0.656 | 8.59E-05 |
| RGS1 | 1.74E-07 | -0.211886636 | 0.063 | 0.109 | 8.64E-05 |
| LAP3 | 1.77E-07 | -0.165949119 | 0.195 | 0.266 | 8.81E-05 |
| CYCS | 1.80E-07 | -0.167896556 | 0.308 | 0.402 | 8.96E-05 |
| ILF2 | 2.94E-07 | -0.174966549 | 0.496 | 0.591 | 0.000146206 |
| CD69 | 3.29E-07 | -0.227397248 | 0.278 | 0.35 | 0.000163616 |
| CHI3L2 | 3.29E-07 | 0.1304251 | 0.139 | 0.086 | 0.000163766 |
| IFITM2 | 3.70E-07 | 0.201761145 | 0.904 | 0.866 | 0.000184316 |
| GAPDH | 3.96E-07 | 0.208265022 | 0.928 | 0.913 | 0.000197417 |
| LDHA | 5.02E-07 | -0.191849904 | 0.842 | 0.857 | 0.000250242 |
| NFKBIZ | 5.75E-07 | -0.201768437 | 0.349 | 0.429 | 0.000286464 |
| CXCR4 | 8.02E-07 | -0.246336034 | 0.928 | 0.923 | 0.000399471 |
| CREM | 8.06E-07 | -0.213766166 | 0.15 | 0.194 | 0.000401504 |
| SNX9 | 1.17E-06 | -0.146121611 | 0.169 | 0.255 | 0.000582148 |
| CD5 | 1.34E-06 | 0.186200388 | 0.659 | 0.598 | 0.000664962 |
| ARHGEF3 | 1.40E-06 | 0.168045647 | 0.465 | 0.389 | 0.00069862 |
| BIN2 | 1.84E-06 | 0.142884768 | 0.364 | 0.28 | 0.000914265 |
| VSIR | 1.94E-06 | 0.145424246 | 0.376 | 0.285 | 0.000966021 |
| CD37 | 2.43E-06 | 0.164788321 | 0.9 | 0.853 | 0.001209432 |
| CSRNP1 | 2.83E-06 | -0.134224277 | 0.128 | 0.193 | 0.001408677 |
| BCL2 | 3.07E-06 | 0.113481098 | 0.174 | 0.105 | 0.001528281 |
| CD4 | 7.08E-06 | 0.201191952 | 0.324 | 0.296 | 0.003525633 |
| CCL22 | 7.69E-06 | -0.228255269 | 0.132 | 0.129 | 0.003829214 |
| GNAI2 | 7.82E-06 | 0.131968558 | 0.99 | 0.985 | 0.003896411 |
| CR2 | 8.48E-06 | 0.095520147 | 0.114 | 0.052 | 0.004221193 |
| CD6 | 1.11E-05 | -0.174490946 | 0.734 | 0.775 | 0.00552272 |
| LAG3 | 1.12E-05 | 0.104958654 | 0.11 | 0.054 | 0.005581371 |
| NFKB2 | 1.21E-05 | -0.085962737 | 0.035 | 0.076 | 0.006049856 |

| | | | | | |
|---|---|---|---|---|---|
| CD3G | 1.44E-05 | 0.0839757 | 0.131 | 0.082 | 0.007182603 |
| CD8A | 1.65E-05 | 0.244809431 | 0.717 | 0.608 | 0.008227193 |
| SOCS3 | 2.55E-05 | -0.106719571 | 0.079 | 0.136 | 0.012682925 |
| GBP2 | 2.87E-05 | -0.182599005 | 0.441 | 0.479 | 0.014284889 |
| PCNA | 4.91E-05 | -0.124581644 | 0.236 | 0.3 | 0.024429259 |
| BAX | 6.47E-05 | -0.156025595 | 0.455 | 0.485 | 0.03222374 |
| NR4A2 | 7.76E-05 | -0.126308948 | 0.035 | 0.071 | 0.038663856 |
| IFITM3 | 7.98E-05 | -0.143641353 | 0.266 | 0.308 | 0.039763424 |
| PIK3IP1 | 8.49E-05 | -0.1401584 | 0.911 | 0.914 | 0.042301448 |
| CD44 | 9.27E-05 | 0.126311263 | 0.939 | 0.928 | 0.04615239 |
| ARID5A | 0.000118897 | -0.117147505 | 0.193 | 0.259 | 0.059210862 |
| TIAF1 | 0.000140742 | 0.079026484 | 0.124 | 0.08 | 0.070089629 |
| JUN | 0.000160827 | -0.072633963 | 0.037 | 0.071 | 0.08009203 |
| FTH1 | 0.000189728 | -0.168941015 | 0.98 | 0.979 | 0.094484319 |
| IL7R | 0.000191793 | 0.164264213 | 0.63 | 0.574 | 0.095512708 |
| CD2 | 0.00024219 | 0.130932859 | 0.897 | 0.845 | 0.120610427 |
| LGALS1 | 0.000275533 | -0.155786974 | 0.433 | 0.482 | 0.137215669 |
| CTSW | 0.000324893 | 0.089092865 | 0.199 | 0.155 | 0.161796848 |
| CD83 | 0.000330696 | -0.106594812 | 0.127 | 0.146 | 0.164686835 |
| TBX21 | 0.000332803 | 0.068484928 | 0.108 | 0.068 | 0.165736073 |
| MAP2K3 | 0.000344226 | 0.128095088 | 0.444 | 0.36 | 0.171424426 |
| CD7 | 0.000378182 | -0.116321874 | 0.221 | 0.284 | 0.188334536 |
| MS4A1 | 0.000390508 | -0.122652601 | 0.158 | 0.194 | 0.19447282 |
| PSMB9 | 0.000396119 | -0.13576837 | 0.586 | 0.59 | 0.197267485 |
| CD8B | 0.000401722 | 0.11288185 | 0.265 | 0.184 | 0.200057681 |
| HAVCR2 | 0.000438547 | 0.071373495 | 0.103 | 0.069 | 0.218396541 |
| FOS | 0.000457983 | -0.107288324 | 0.137 | 0.192 | 0.228075574 |
| FYN | 0.000464693 | 0.13805946 | 0.821 | 0.743 | 0.231417155 |
| ADA | 0.000488601 | 0.100384901 | 0.272 | 0.202 | 0.243323184 |
| CCL2 | 0.000725486 | -0.16855427 | 0.785 | 0.783 | 0.361292173 |
| TCF7 | 0.000779151 | 0.096155665 | 0.332 | 0.262 | 0.38801715 |
| NT5E | 0.000887493 | 0.106397701 | 0.185 | 0.137 | 0.441971512 |
| BCL6 | 0.000932754 | -0.042369919 | 0.016 | 0.043 | 0.464511615 |
| CCL3 | 0.001050949 | -0.138426165 | 0.285 | 0.314 | 0.523372835 |
| CD247 | 0.00125406 | 0.110191154 | 0.487 | 0.444 | 0.624521925 |
| NINJ2 | 0.001407005 | 0.027898398 | 0.027 | 0.01 | 0.700688418 |
| JAK3 | 0.00159708 | -0.117282619 | 0.686 | 0.716 | 0.795345775 |
| IGHG3 | 0.001808038 | -0.065191629 | 0.032 | 0.052 | 0.90040288 |
| ITK | 0.00205314 | -0.110495572 | 0.83 | 0.844 | 1 |
| AHI1 | 0.002213999 | 0.052996685 | 0.066 | 0.039 | 1 |
| IGBP1 | 0.003224329 | 0.089591552 | 0.343 | 0.279 | 1 |
| POU2AF1 | 0.003260003 | 0.067391381 | 0.09 | 0.056 | 1 |
| NFATC1 | 0.003366231 | 0.083673878 | 0.288 | 0.238 | 1 |
| JUNB | 0.003413263 | -0.086292248 | 0.172 | 0.206 | 1 |
| CD274 | 0.003547747 | 0.048756956 | 0.091 | 0.057 | 1 |
| PDIA4 | 0.003658755 | 0.110996051 | 0.469 | 0.41 | 1 |
| APOBEC3G | 0.003829264 | 0.089428028 | 0.309 | 0.261 | 1 |

| | | | | | |
|---|---|---|---|---|---|
| CCL4 | 0.003891158 | -0.132222057 | 0.284 | 0.307 | 1 |
| EIF5 | 0.004044611 | -0.108305825 | 0.511 | 0.525 | 1 |
| XBP1 | 0.004145498 | -0.097353116 | 0.295 | 0.347 | 1 |
| FAS | 0.004647875 | 0.04682231 | 0.08 | 0.045 | 1 |
| DUSP5 | 0.005513964 | -0.042860656 | 0.02 | 0.035 | 1 |
| IGHG2 | 0.005659542 | -0.058286744 | 0.025 | 0.036 | 1 |
| MYB | 0.007375174 | 0.04793755 | 0.058 | 0.038 | 1 |
| GZMA | 0.008664506 | 0.046663395 | 0.075 | 0.055 | 1 |
| HLA.DRA | 0.008980663 | -0.138531996 | 0.655 | 0.658 | 1 |
| CCR1 | 0.009711707 | 0.047671267 | 0.114 | 0.085 | 1 |
| TNFSF13B | 0.010727862 | -0.055059181 | 0.054 | 0.077 | 1 |
| NAMPT | 0.011275944 | -0.074252062 | 0.189 | 0.215 | 1 |
| TNFRSF13C | 0.011461318 | 0.075756152 | 0.195 | 0.145 | 1 |
| IL6R | 0.012658256 | 0.056934065 | 0.15 | 0.118 | 1 |
| SOCS2 | 0.013082466 | 0.03894112 | 0.047 | 0.025 | 1 |
| CD3D | 0.01393041 | 0.080115152 | 0.965 | 0.95 | 1 |
| TOP2A | 0.015017184 | 0.015124371 | 0.015 | 0.006 | 1 |
| PRF1 | 0.015216727 | 0.084463426 | 0.352 | 0.328 | 1 |
| IFRD1 | 0.015286972 | -0.064718459 | 0.166 | 0.203 | 1 |
| SLC25A37 | 0.015649313 | 0.071376895 | 0.252 | 0.202 | 1 |
| CASP5 | 0.01760912 | 0.050029118 | 0.139 | 0.12 | 1 |
| IGHM | 0.018732377 | 0.098740365 | 0.271 | 0.259 | 1 |
| HLA.DQB1 | 0.02038654 | -0.100342368 | 0.252 | 0.251 | 1 |
| CD74 | 0.020853812 | -0.113024946 | 0.92 | 0.907 | 1 |
| SELL | 0.021336481 | 0.095357567 | 0.916 | 0.895 | 1 |
| IL2 | 0.02167719 | -0.055693909 | 0.02 | 0.024 | 1 |
| PRNP | 0.022217921 | -0.06066681 | 0.167 | 0.197 | 1 |
| FURIN | 0.022917004 | -0.075444865 | 0.27 | 0.275 | 1 |
| ATF6B | 0.023609632 | 0.069653386 | 0.364 | 0.298 | 1 |
| CXCL13 | 0.023650492 | -0.151941303 | 0.009 | 0.005 | 1 |
| POU2F2 | 0.024781633 | 0.059849769 | 0.204 | 0.166 | 1 |
| HNRNPC | 0.025617145 | -0.079353166 | 0.634 | 0.646 | 1 |
| GNLY | 0.027409457 | -0.089545062 | 0.42 | 0.474 | 1 |
| NFATC3 | 0.027690893 | 0.067788018 | 0.474 | 0.426 | 1 |
| GEM | 0.028856386 | -0.017766589 | 0.003 | 0.01 | 1 |
| IL6 | 0.028957096 | -0.036352894 | 0.022 | 0.03 | 1 |
| LIPA | 0.029591654 | -0.075015428 | 0.465 | 0.507 | 1 |
| PAK1IP1 | 0.030694789 | -0.036663223 | 0.053 | 0.081 | 1 |
| TNIK | 0.032553534 | 0.054592897 | 0.171 | 0.132 | 1 |
| IGHG1 | 0.033369711 | -0.080762789 | 0.208 | 0.232 | 1 |
| VPS28 | 0.034570106 | 0.069311563 | 0.654 | 0.626 | 1 |
| SKAP1 | 0.036988756 | 0.040360648 | 0.094 | 0.072 | 1 |
| GAB2 | 0.040139626 | -0.032695218 | 0.034 | 0.056 | 1 |
| CD28 | 0.040857631 | 0.049090253 | 0.166 | 0.136 | 1 |
| RBCK1 | 0.041614934 | -0.061100767 | 0.24 | 0.277 | 1 |
| BACH2 | 0.041870412 | -0.051996198 | 0.143 | 0.161 | 1 |
| CD40 | 0.042893195 | -0.039666557 | 0.063 | 0.073 | 1 |

| | | | | | |
|---|---|---|---|---|---|
| FCER1G | 0.04664545 | -0.074455702 | 0.53 | 0.546 | 1 |
| IGKC | 0.048809849 | -0.110071208 | 0.629 | 0.614 | 1 |
| PDIA6 | 0.051532146 | 0.065265771 | 0.445 | 0.409 | 1 |
| IVNS1ABP | 0.053536899 | -0.06564169 | 0.482 | 0.49 | 1 |
| REL | 0.053896184 | -0.055186633 | 0.186 | 0.198 | 1 |
| TRAC | 0.054321395 | 0.071366582 | 0.994 | 0.988 | 1 |
| VNN2 | 0.058939544 | 0.048247352 | 0.222 | 0.19 | 1 |
| RPN2 | 0.058966114 | 0.061791517 | 0.592 | 0.538 | 1 |
| SPP1 | 0.059644644 | -0.019428843 | 0.005 | 0.01 | 1 |
| ITGAE | 0.060324676 | -0.04629852 | 0.114 | 0.142 | 1 |
| IFNG | 0.061886311 | -0.036450734 | 0.023 | 0.025 | 1 |
| IRF8 | 0.062191395 | -0.050570197 | 0.165 | 0.175 | 1 |
| BCL2A1 | 0.062753392 | 0.055126848 | 0.189 | 0.139 | 1 |
| CD9 | 0.063538937 | -0.048566482 | 0.139 | 0.161 | 1 |
| CARD11 | 0.065286821 | 0.01048401 | 0.011 | 0.005 | 1 |
| AURKB | 0.065505654 | 0.008619399 | 0.005 | 0.002 | 1 |
| MKI67 | 0.065723964 | 0.006525101 | 0.004 | 0.001 | 1 |
| CHD1 | 0.066733862 | -0.011380766 | 0.004 | 0.012 | 1 |
| ABCE1 | 0.066929377 | -0.054980763 | 0.347 | 0.374 | 1 |
| KLRC4 | 0.067043509 | 0.034266506 | 0.094 | 0.071 | 1 |
| TRDC | 0.069169168 | 0.09313839 | 0.113 | 0.097 | 1 |
| NRP1 | 0.069899396 | -0.01967631 | 0.023 | 0.036 | 1 |
| SLA | 0.071689716 | 0.063098814 | 0.371 | 0.325 | 1 |
| EVI2A | 0.075567964 | 0.063475496 | 0.665 | 0.629 | 1 |
| JAK1 | 0.075661626 | 0.054812848 | 0.953 | 0.927 | 1 |
| CD33 | 0.080454162 | -0.016551176 | 0.014 | 0.024 | 1 |
| CXCR5 | 0.082827946 | 0.047071464 | 0.1 | 0.072 | 1 |
| IL1A | 0.085320399 | -0.010153405 | 0.003 | 0.009 | 1 |
| CXCL1 | 0.088437781 | -0.072425621 | 0.302 | 0.299 | 1 |
| FCGR3A | 0.090672202 | 0.042736561 | 0.196 | 0.175 | 1 |
| CD86 | 0.090779392 | -0.039630202 | 0.048 | 0.044 | 1 |
| CST7 | 0.10024599 | 0.064404028 | 0.295 | 0.276 | 1 |
| CD40LG | 0.101056617 | -0.01729772 | 0.02 | 0.03 | 1 |
| F5 | 0.102537216 | 0.017262368 | 0.024 | 0.016 | 1 |
| ERBIN | 0.105647708 | -0.056740266 | 0.53 | 0.567 | 1 |
| ICOS | 0.107533077 | 0.044339492 | 0.196 | 0.162 | 1 |
| CD14 | 0.108141329 | 0.055558636 | 0.428 | 0.403 | 1 |
| CCND2 | 0.109745911 | 0.051527991 | 0.27 | 0.243 | 1 |
| NKG7 | 0.111006197 | -0.06197249 | 0.376 | 0.4 | 1 |
| KLRC3 | 0.115158047 | -0.051909351 | 0.113 | 0.137 | 1 |
| IL2RG | 0.116281668 | 0.053865238 | 0.926 | 0.901 | 1 |
| RNASE6 | 0.12000112 | 0.01370893 | 0.028 | 0.018 | 1 |
| LEF1 | 0.127139741 | -0.054680306 | 0.906 | 0.905 | 1 |
| IL18 | 0.132259307 | 0.007358499 | 0.008 | 0.004 | 1 |
| CLEC10A | 0.133313418 | 0.016561068 | 0.034 | 0.025 | 1 |
| IL3RA | 0.139655406 | -0.01934098 | 0.034 | 0.048 | 1 |
| CSF2 | 0.144666305 | -0.019715478 | 0.01 | 0.01 | 1 |

| | | | | | |
|---|---|---|---|---|---|
| STAT6 | 0.146868075 | 0.044849368 | 0.345 | 0.32 | 1 |
| CD79B | 0.155974073 | 0.032495101 | 0.125 | 0.105 | 1 |
| ZNF683 | 0.157509059 | 0.025302241 | 0.063 | 0.049 | 1 |
| TIMM9 | 0.157565919 | -0.047613544 | 0.441 | 0.461 | 1 |
| CD63 | 0.157774816 | -0.052701302 | 0.572 | 0.58 | 1 |
| IL18R1 | 0.157778296 | 0.016561068 | 0.037 | 0.021 | 1 |
| DUSP4 | 0.162899443 | -0.026777918 | 0.032 | 0.035 | 1 |
| UGCG | 0.16930036 | 0.036167125 | 0.213 | 0.188 | 1 |
| LTB | 0.170714787 | 0.045728457 | 0.299 | 0.289 | 1 |
| IL23A | 0.172591035 | 0.036833148 | 0.223 | 0.174 | 1 |
| CHD7 | 0.173377479 | 0.015320599 | 0.035 | 0.027 | 1 |
| CTSD | 0.179907303 | 0.045588752 | 0.332 | 0.33 | 1 |
| BCL2L1 | 0.18345972 | -0.011281739 | 0.014 | 0.022 | 1 |
| TRBC2 | 0.184458008 | 0.06534983 | 0.966 | 0.975 | 1 |
| TGIF1 | 0.185830528 | -0.053247429 | 0.411 | 0.413 | 1 |
| TUBA1C | 0.188826015 | -0.047870939 | 0.446 | 0.455 | 1 |
| NFATC2 | 0.192738416 | -0.034244005 | 0.176 | 0.201 | 1 |
| LTA | 0.197676548 | -0.029186812 | 0.091 | 0.115 | 1 |
| EBF1 | 0.198085107 | -0.007861594 | 0.003 | 0.005 | 1 |
| CD79A | 0.198808204 | -0.05057994 | 0.19 | 0.213 | 1 |
| CXCL9 | 0.19940052 | 0.017546821 | 0.023 | 0.018 | 1 |
| F13A1 | 0.202198259 | -0.00585884 | 0.001 | 0.004 | 1 |
| TNFRSF25 | 0.20808691 | 0.026411 | 0.104 | 0.09 | 1 |
| TCF4 | 0.217449008 | -0.006331694 | 0.003 | 0.006 | 1 |
| MALT1 | 0.218513043 | 0.026327892 | 0.152 | 0.133 | 1 |
| CCR9 | 0.218751151 | 0.023591865 | 0.025 | 0.018 | 1 |
| SLC7A1 | 0.219325672 | -0.022613225 | 0.075 | 0.081 | 1 |
| TGFBI | 0.220907605 | -0.040460874 | 0.312 | 0.334 | 1 |
| TCL1A | 0.221048232 | 0.019900865 | 0.038 | 0.037 | 1 |
| LILRB4 | 0.221523244 | -0.020838535 | 0.06 | 0.07 | 1 |
| SDCBP | 0.223349006 | -0.042878459 | 0.672 | 0.694 | 1 |
| TGFBR2 | 0.229725269 | -0.046164072 | 0.834 | 0.842 | 1 |
| CD200 | 0.239816539 | 0.016820282 | 0.034 | 0.018 | 1 |
| CD38 | 0.243633583 | 0.016872962 | 0.037 | 0.028 | 1 |
| IL11RA | 0.246566331 | 0.040498565 | 0.37 | 0.356 | 1 |
| FBXO11 | 0.24854161 | 0.034470633 | 0.292 | 0.274 | 1 |
| YBX3 | 0.251031841 | 0.014084213 | 0.035 | 0.027 | 1 |
| SPAG9 | 0.252407158 | -0.02680828 | 0.129 | 0.151 | 1 |
| TRAF1 | 0.258715305 | -0.029151213 | 0.156 | 0.161 | 1 |
| MCM4 | 0.269631328 | -0.013286152 | 0.025 | 0.033 | 1 |
| MCM2 | 0.270344486 | 0.022224751 | 0.103 | 0.087 | 1 |
| ITGAM | 0.270589202 | -0.012543069 | 0.028 | 0.034 | 1 |
| FOXO1 | 0.273226338 | -0.028624009 | 0.175 | 0.186 | 1 |
| EGR1 | 0.277902054 | -0.005092061 | 0.001 | 0.003 | 1 |
| CD72 | 0.280412897 | -0.012543069 | 0.027 | 0.034 | 1 |
| GRB10 | 0.281691722 | -0.008847117 | 0.011 | 0.015 | 1 |
| KLRC1 | 0.283178213 | -0.021875046 | 0.044 | 0.049 | 1 |

| | | | | | |
|---|---|---|---|---|---|
| TYMS | 0.289990183 | 0.003164969 | 0.004 | 0.002 | 1 |
| MME | 0.289990183 | 0.003164969 | 0.004 | 0.002 | 1 |
| TGFB1 | 0.290180438 | -0.011151189 | 0.016 | 0.021 | 1 |
| CD244 | 0.292627688 | -0.008554206 | 0.013 | 0.018 | 1 |
| TNF | 0.293644852 | -0.019700229 | 0.053 | 0.06 | 1 |
| CCL20 | 0.295806121 | -0.004800171 | 0.003 | 0.005 | 1 |
| NANS | 0.2992263 | -0.02986163 | 0.176 | 0.195 | 1 |
| SP140 | 0.300679192 | 0.027324827 | 0.171 | 0.149 | 1 |
| CXCL2 | 0.302891452 | -0.023133009 | 0.117 | 0.127 | 1 |
| CD1C | 0.303519958 | 0.007757274 | 0.016 | 0.012 | 1 |
| SOCS1 | 0.311526984 | -0.003557279 | 0.001 | 0.004 | 1 |
| DPP4 | 0.324466113 | 0.014997904 | 0.049 | 0.042 | 1 |
| CX3CR1 | 0.331109397 | -0.013191484 | 0.042 | 0.051 | 1 |
| CXCL8 | 0.332171827 | -0.042156671 | 0.485 | 0.463 | 1 |
| TNFSF14 | 0.333228193 | 0.005056937 | 0.009 | 0.005 | 1 |
| GZMB | 0.333496066 | -0.026535265 | 0.188 | 0.203 | 1 |
| CD36 | 0.338368458 | -0.021975594 | 0.133 | 0.142 | 1 |
| SNCA | 0.33888489 | -0.006396113 | 0.009 | 0.013 | 1 |
| IL4R | 0.34524628 | -0.035027885 | 0.359 | 0.354 | 1 |
| CD209 | 0.3470189 | -0.004508959 | 0.004 | 0.007 | 1 |
| CD80 | 0.347345861 | -0.007273363 | 0.005 | 0.006 | 1 |
| TRIB1 | 0.350256819 | -0.01060191 | 0.023 | 0.032 | 1 |
| RORA | 0.350904023 | 0.039802433 | 0.454 | 0.427 | 1 |
| IKZF1 | 0.352939497 | -0.032880617 | 0.465 | 0.488 | 1 |
| TNFSF8 | 0.355774213 | 0.010515951 | 0.033 | 0.022 | 1 |
| IL12A | 0.361560186 | -0.004033799 | 0.003 | 0.005 | 1 |
| CYBB | 0.364534012 | 0.02564126 | 0.288 | 0.26 | 1 |
| BCL11B | 0.367643949 | 0.036594489 | 0.674 | 0.638 | 1 |
| RUNX3 | 0.38027268 | -0.035343584 | 0.659 | 0.666 | 1 |
| DUSP1 | 0.381712129 | -0.029061 | 0.245 | 0.239 | 1 |
| IGHA1 | 0.391023544 | -0.010399589 | 0.013 | 0.018 | 1 |
| VSIG4 | 0.392729568 | -0.008142439 | 0.014 | 0.022 | 1 |
| LAT2 | 0.39297917 | -0.010927781 | 0.03 | 0.033 | 1 |
| FN1 | 0.393708443 | -0.029883674 | 0.295 | 0.297 | 1 |
| IRF3 | 0.396150276 | 0.016347611 | 0.096 | 0.086 | 1 |
| PDCD1 | 0.398052623 | 0.00344735 | 0.005 | 0.002 | 1 |
| HLA.A | 0.399014883 | -0.02267407 | 1 | 1 | 1 |
| B3GAT1 | 0.410755746 | 0.003729075 | 0.005 | 0.004 | 1 |
| STIM1 | 0.413054185 | -0.020049832 | 0.196 | 0.204 | 1 |
| GIMAP5 | 0.41733772 | 0.031075056 | 0.83 | 0.807 | 1 |
| TRAT1 | 0.42102428 | 0.016426146 | 0.106 | 0.093 | 1 |
| IL15 | 0.422308128 | 0.004570335 | 0.009 | 0.007 | 1 |
| CD22 | 0.427301161 | 0.015746684 | 0.082 | 0.061 | 1 |
| CD109 | 0.429359988 | -0.006749709 | 0.014 | 0.017 | 1 |
| TMEM97 | 0.43893252 | -0.009732714 | 0.027 | 0.032 | 1 |
| IRF4 | 0.442350323 | -0.016123774 | 0.054 | 0.057 | 1 |
| CD70 | 0.447349622 | 0.004570335 | 0.01 | 0.006 | 1 |

| | | | | | |
|---|---|---|---|---|---|
| TNFRSF4 | 0.450518402 | 0.002395737 | 0.004 | 0.002 | 1 |
| THBS1 | 0.453121383 | -0.029306138 | 0.479 | 0.494 | 1 |
| PSME3 | 0.454393354 | -0.016551176 | 0.128 | 0.142 | 1 |
| CCR6 | 0.458975423 | 0.008149264 | 0.019 | 0.018 | 1 |
| LY86 | 0.462487507 | -0.012580714 | 0.07 | 0.078 | 1 |
| IGLC3 | 0.474662994 | 0.042287916 | 0.539 | 0.498 | 1 |
| FOXP1 | 0.47765753 | -0.017080703 | 0.117 | 0.137 | 1 |
| IL12RB1 | 0.482814433 | -0.008076517 | 0.028 | 0.029 | 1 |
| BTLA | 0.491425679 | 0.00923285 | 0.046 | 0.038 | 1 |
| FASLG | 0.492235735 | 0.003805183 | 0.01 | 0.007 | 1 |
| IGHD | 0.496951074 | 0.0168569 | 0.085 | 0.073 | 1 |
| KLRG1 | 0.497104381 | 0.007845478 | 0.037 | 0.032 | 1 |
| FOSL1 | 0.503953451 | -0.002977435 | 0.004 | 0.006 | 1 |
| IL12RB2 | 0.504776582 | -0.007734638 | 0.019 | 0.023 | 1 |
| CXCR6 | 0.510302315 | -0.006282841 | 0.01 | 0.013 | 1 |
| VMO1 | 0.51711519 | -0.01546276 | 0.098 | 0.1 | 1 |
| CBLB | 0.518863404 | -0.007502316 | 0.029 | 0.033 | 1 |
| BCAT1 | 0.519562031 | 0.005751218 | 0.019 | 0.012 | 1 |
| LGALS9 | 0.521659073 | -0.015756522 | 0.122 | 0.13 | 1 |
| RORC | 0.524951212 | 0.00296148 | 0.006 | 0.004 | 1 |
| ZBTB16 | 0.524962143 | 0.002678938 | 0.004 | 0.003 | 1 |
| DUSP6 | 0.529651504 | -0.006058802 | 0.019 | 0.023 | 1 |
| PSEN1 | 0.535467485 | 0.012750081 | 0.108 | 0.098 | 1 |
| TNFSF10 | 0.536949644 | 0.019951078 | 0.226 | 0.218 | 1 |
| CD163 | 0.539716014 | 0.011237686 | 0.1 | 0.099 | 1 |
| NCAM1 | 0.544793795 | -0.002020862 | 0.001 | 0.002 | 1 |
| CCL13 | 0.546125335 | -0.004766608 | 0.013 | 0.014 | 1 |
| TGFBR1 | 0.549534405 | 0.012179 | 0.123 | 0.113 | 1 |
| FOSB | 0.550053184 | -0.010106883 | 0.01 | 0.012 | 1 |
| LIG4 | 0.551282517 | 0.011977037 | 0.101 | 0.093 | 1 |
| IL2RA | 0.566602416 | -0.00392856 | 0.005 | 0.007 | 1 |
| LAMP1 | 0.567198024 | 0.019223561 | 0.492 | 0.485 | 1 |
| BATF | 0.567604709 | -0.008562409 | 0.048 | 0.051 | 1 |
| STAT5A | 0.568197858 | -0.018545226 | 0.37 | 0.389 | 1 |
| IL15RA | 0.570905965 | -0.010760899 | 0.075 | 0.072 | 1 |
| CNOT2 | 0.579053761 | -0.017073797 | 0.364 | 0.348 | 1 |
| CLEC4E | 0.58169225 | 0.005952968 | 0.03 | 0.022 | 1 |
| MMP9 | 0.583386134 | 0.011536623 | 0.119 | 0.116 | 1 |
| FCN1 | 0.584438077 | -0.011132988 | 0.087 | 0.088 | 1 |
| OSM | 0.600153766 | -0.002688523 | 0.005 | 0.006 | 1 |
| ZNF341 | 0.600954535 | 0.005132651 | 0.025 | 0.021 | 1 |
| UFM1 | 0.606609912 | -0.017631629 | 0.534 | 0.534 | 1 |
| EEF1E1 | 0.611617614 | -0.008379682 | 0.052 | 0.067 | 1 |
| IGHG4 | 0.613729555 | -0.002499833 | 0.001 | 0.003 | 1 |
| THBD | 0.615469195 | 0.001342234 | 0.003 | 0.002 | 1 |
| TGFB3 | 0.619205043 | -0.003824105 | 0.009 | 0.011 | 1 |
| FOXP3 | 0.620772125 | 0.009550926 | 0.016 | 0.017 | 1 |

| | | | | | |
|---|---|---|---|---|---|
| FCER2 | 0.622398551 | -0.006188007 | 0.029 | 0.033 | 1 |
| LIF | 0.640393371 | -0.00125204 | 0.001 | 0.002 | 1 |
| EIF3J | 0.643670246 | -0.007383901 | 0.063 | 0.066 | 1 |
| MYC | 0.648868405 | -0.023183693 | 0.584 | 0.544 | 1 |
| S100A9 | 0.656007054 | -0.016346698 | 0.345 | 0.322 | 1 |
| EIF1 | 0.656447281 | 0.016888926 | 0.957 | 0.939 | 1 |
| PAXBP1 | 0.660277271 | 0.014614681 | 0.422 | 0.413 | 1 |
| KLRF1 | 0.663981957 | -0.004628009 | 0.024 | 0.028 | 1 |
| CXCL16 | 0.667401431 | -0.011476271 | 0.196 | 0.196 | 1 |
| KIT | 0.667917531 | -0.002977435 | 0.004 | 0.003 | 1 |
| IFNGR1 | 0.674513615 | -0.011418512 | 0.233 | 0.235 | 1 |
| PRDM1 | 0.679583125 | -0.006825269 | 0.047 | 0.045 | 1 |
| MZB1 | 0.680102375 | 0.003475074 | 0.016 | 0.016 | 1 |
| MGST1 | 0.689837285 | -0.004163608 | 0.024 | 0.026 | 1 |
| PIK3AP1 | 0.694637606 | 0.005205795 | 0.043 | 0.035 | 1 |
| SMAD2 | 0.70469195 | 0.010548017 | 0.251 | 0.238 | 1 |
| ICAM1 | 0.711752781 | -0.007643508 | 0.105 | 0.102 | 1 |
| PTPN7 | 0.71289122 | -0.010235777 | 0.181 | 0.19 | 1 |
| THEMIS | 0.71632997 | -0.008513857 | 0.117 | 0.119 | 1 |
| STAT4 | 0.720527071 | -0.011919728 | 0.349 | 0.341 | 1 |
| PTPRC | 0.725078557 | 0.010536983 | 0.985 | 0.983 | 1 |
| SNX8 | 0.730980428 | -0.00705789 | 0.081 | 0.081 | 1 |
| ANXA5 | 0.73381073 | 0.012240117 | 0.413 | 0.408 | 1 |
| SRGN | 0.73382246 | -0.00623759 | 0.074 | 0.074 | 1 |
| MAST4 | 0.736039618 | 0.006505443 | 0.081 | 0.072 | 1 |
| KCNE3 | 0.737387394 | 0.002717157 | 0.019 | 0.017 | 1 |
| LRRC8B | 0.737696659 | 0.004122392 | 0.037 | 0.033 | 1 |
| ADGRE1 | 0.738921614 | -0.006052527 | 0.046 | 0.055 | 1 |
| ARL4C | 0.74016195 | -0.012515663 | 0.659 | 0.646 | 1 |
| ITGAX | 0.743355445 | 0.006116015 | 0.085 | 0.083 | 1 |
| KDELR1 | 0.749137488 | 0.009824239 | 0.341 | 0.335 | 1 |
| PGM3 | 0.751590663 | -0.005567041 | 0.062 | 0.065 | 1 |
| HLA.DMA | 0.752860629 | -0.011265543 | 0.259 | 0.241 | 1 |
| GATA3 | 0.754315522 | 0.002717157 | 0.016 | 0.016 | 1 |
| TREM1 | 0.761463044 | 0.00171103 | 0.006 | 0.006 | 1 |
| LGALS3 | 0.761745296 | 0.007427687 | 0.122 | 0.116 | 1 |
| MITF | 0.763257124 | 0.003150738 | 0.024 | 0.024 | 1 |
| VPREB3 | 0.767587063 | 0.004882147 | 0.056 | 0.056 | 1 |
| QPCT | 0.773330418 | -0.001635534 | 0.005 | 0.007 | 1 |
| TLR7 | 0.773853068 | -0.000964234 | 0.003 | 0.003 | 1 |
| S100A12 | 0.778294903 | -0.002947091 | 0.024 | 0.022 | 1 |
| PCLAF | 0.780253622 | 0.001143339 | 0.005 | 0.004 | 1 |
| APOE | 0.798189798 | -0.000964234 | 0.003 | 0.003 | 1 |
| IKZF2 | 0.809081478 | 0.000858502 | 0.004 | 0.003 | 1 |
| NDFIP2 | 0.830318758 | 0.000858502 | 0.004 | 0.003 | 1 |
| ENTPD1 | 0.831372711 | -0.001918184 | 0.014 | 0.013 | 1 |
| CCR5 | 0.83483676 | 0.002122331 | 0.027 | 0.022 | 1 |

| | | | | | |
|---|---|---|---|---|---|
| RAPH1 | 0.84299802 | 0.001314307 | 0.013 | 0.012 | 1 |
| BCL10 | 0.84850461 | 0.003696534 | 0.093 | 0.092 | 1 |
| CXCL3 | 0.859217294 | 0.004760866 | 0.152 | 0.131 | 1 |
| SLC1A5 | 0.86463483 | 0.00407336 | 0.122 | 0.122 | 1 |
| LRRC32 | 0.869751042 | 0.000573002 | 0.001 | 0.002 | 1 |
| CD24 | 0.888628049 | -0.001159869 | 0.015 | 0.014 | 1 |
| SLC7A7 | 0.888952097 | -0.002252793 | 0.066 | 0.066 | 1 |
| CSF1 | 0.891236661 | -0.000969041 | 0.011 | 0.013 | 1 |
| IL1B | 0.893033671 | -0.004779661 | 0.242 | 0.214 | 1 |
| CXCL10 | 0.895215319 | -0.002862863 | 0.07 | 0.061 | 1 |
| IL18RAP | 0.89584489 | 0.001286935 | 0.02 | 0.02 | 1 |
| GZMH | 0.898795774 | -0.001444284 | 0.033 | 0.034 | 1 |
| FUT4 | 0.906561125 | -0.001628125 | 0.044 | 0.042 | 1 |
| HLA.DPA1 | 0.914172862 | 0.005265492 | 0.531 | 0.48 | 1 |
| PTGDR2 | 0.922714727 | 0.000748624 | 0.008 | 0.007 | 1 |
| BLNK | 0.923100183 | -0.001070189 | 0.025 | 0.028 | 1 |
| CCR4 | 0.923425254 | -0.003236823 | 0.066 | 0.069 | 1 |
| CMKLR1 | 0.928166924 | -0.00078386 | 0.02 | 0.02 | 1 |
| JAK2 | 0.929565404 | -0.000390627 | 0.005 | 0.005 | 1 |
| DCLRE1C | 0.930911297 | -0.0019538 | 0.11 | 0.111 | 1 |
| IL1RN | 0.938729421 | 0.001737743 | 0.072 | 0.06 | 1 |
| NIBAN3 | 0.939061066 | -0.000390627 | 0.004 | 0.005 | 1 |
| TLR2 | 0.942942301 | -0.000692293 | 0.023 | 0.024 | 1 |
| PLCG2 | 0.944864142 | -0.000987344 | 0.048 | 0.05 | 1 |
| NFKBID | 0.950162809 | -0.000195821 | 0.003 | 0.003 | 1 |
| KLRB1 | 0.956940451 | -0.000195821 | 0.003 | 0.002 | 1 |
| JCHAIN | 0.958411584 | -0.000699667 | 0.03 | 0.027 | 1 |
| ORAI1 | 0.958523538 | 0.001616598 | 0.374 | 0.38 | 1 |
| NGDN | 0.966778779 | -0.000411474 | 0.025 | 0.024 | 1 |
| IL1R1 | 0.968614494 | 0.00026947 | 0.01 | 0.011 | 1 |
| LYN | 0.98105102 | -0.000631086 | 0.148 | 0.143 | 1 |
| B4GALT5 | 0.994055235 | -0.011513988 | 0 | 0.007 | 1 |
| TNFRSF8 | 0.994350054 | -0.004616628 | 0 | 0.003 | 1 |
| AQP9 | 0.996396476 | -6.97E-05 | 0.046 | 0.041 | 1 |
| ZBED2 | 0.996413542 | -0.003848215 | 0 | 0.002 | 1 |
| CMTM2 | 0.996457089 | -0.003079393 | 0 | 0.002 | 1 |
| PIGF | 0.996513232 | -0.002310161 | 0 | 0.002 | 1 |
| CXCL5 | 0.998188808 | 9.20E-05 | 0.531 | 0.54 | 1 |
| IL17RB | 0.99903478 | -0.008452565 | 0 | 0.002 | 1 |

CD8+ Memory

| | p_val | avg_log2FC | Patient | Controls | p_val_adj |
|---|---|---|---|---|---|
| CD52 | 2.13E-57 | 0.589463254 | 0.998 | 0.969 | 1.06E-54 |
| CXCR4 | 3.84E-56 | -0.703710338 | 0.914 | 0.962 | 1.91E-53 |
| ZNF683 | 3.65E-37 | 1.173910334 | 0.58 | 0.295 | 1.82E-34 |
| CCL4 | 3.81E-37 | -0.865575332 | 0.617 | 0.652 | 1.90E-34 |

| | | | | | |
|---|---|---|---|---|---|
| GZMH | 9.72E-33 | 0.603117175 | 0.672 | 0.464 | 4.84E-30 |
| IRF7 | 9.76E-33 | -0.481771928 | 0.312 | 0.497 | 4.86E-30 |
| FCGR3A | 6.75E-32 | 0.616841764 | 0.67 | 0.507 | 3.36E-29 |
| NFKBIA | 5.20E-31 | -0.510128718 | 0.638 | 0.769 | 2.59E-28 |
| NKG7 | 3.94E-30 | 0.588288454 | 0.934 | 0.93 | 1.96E-27 |
| STAT1 | 8.35E-28 | -0.528224284 | 0.629 | 0.685 | 4.16E-25 |
| SELPLG | 7.92E-25 | 0.332668013 | 0.927 | 0.852 | 3.94E-22 |
| CCL22 | 5.22E-24 | -0.841667123 | 0.148 | 0.151 | 2.60E-21 |
| IFI6 | 1.98E-23 | -0.433197049 | 0.172 | 0.325 | 9.87E-21 |
| DUSP2 | 2.53E-23 | -0.52714846 | 0.46 | 0.683 | 1.26E-20 |
| GBP1 | 7.78E-23 | -0.445112168 | 0.313 | 0.429 | 3.87E-20 |
| ITGB2 | 9.03E-21 | 0.343550636 | 0.94 | 0.848 | 4.50E-18 |
| IRF9 | 5.57E-20 | -0.378098173 | 0.625 | 0.71 | 2.78E-17 |
| IGLC3 | 5.51E-18 | -0.574102602 | 0.473 | 0.51 | 2.74E-15 |
| PSME2 | 1.18E-17 | -0.346748216 | 0.708 | 0.776 | 5.88E-15 |
| CX3CR1 | 7.35E-17 | 0.461160424 | 0.593 | 0.435 | 3.66E-14 |
| FTH1 | 2.34E-16 | -0.370704951 | 0.978 | 0.98 | 1.16E-13 |
| ITGA4 | 2.78E-16 | 0.300525814 | 0.633 | 0.547 | 1.38E-13 |
| FYN | 2.25E-15 | -0.264709343 | 0.861 | 0.899 | 1.12E-12 |
| TRBC2 | 2.29E-14 | 0.385803607 | 0.972 | 0.952 | 1.14E-11 |
| TSPAN32 | 2.43E-14 | 0.196682859 | 0.267 | 0.147 | 1.21E-11 |
| KLRF1 | 2.96E-14 | -0.289075611 | 0.112 | 0.239 | 1.47E-11 |
| CD69 | 3.45E-14 | -0.333459347 | 0.357 | 0.466 | 1.72E-11 |
| PASK | 1.06E-13 | 0.426278917 | 0.407 | 0.375 | 5.27E-11 |
| CD8A | 1.26E-13 | 0.544859642 | 0.743 | 0.573 | 6.29E-11 |
| KLRK1 | 2.74E-13 | 0.288156775 | 0.93 | 0.891 | 1.37E-10 |
| CD226 | 4.81E-13 | 0.183706355 | 0.251 | 0.144 | 2.40E-10 |
| TRDC | 1.56E-12 | -0.550592246 | 0.298 | 0.453 | 7.77E-10 |
| LAG3 | 2.80E-12 | -0.291800437 | 0.324 | 0.4 | 1.40E-09 |
| PLCG2 | 4.22E-12 | -0.207460283 | 0.116 | 0.215 | 2.10E-09 |
| IFITM2 | 9.39E-12 | 0.27409366 | 0.941 | 0.91 | 4.68E-09 |
| IL32 | 3.11E-11 | 0.256125638 | 0.896 | 0.812 | 1.55E-08 |
| HIF1A | 5.93E-11 | -0.220997238 | 0.316 | 0.435 | 2.95E-08 |
| IL12RB2 | 4.08E-10 | -0.145193358 | 0.038 | 0.102 | 2.03E-07 |
| CD3E | 6.10E-10 | 0.201992961 | 0.974 | 0.933 | 3.04E-07 |
| CD83 | 7.29E-10 | -0.261338524 | 0.102 | 0.159 | 3.63E-07 |
| LAMP3 | 7.48E-10 | -0.266497128 | 0.029 | 0.062 | 3.73E-07 |
| FYB1 | 1.01E-09 | 0.214449782 | 0.73 | 0.617 | 5.05E-07 |
| KLRC4 | 1.39E-09 | 0.156961481 | 0.217 | 0.131 | 6.93E-07 |
| LCK | 1.50E-09 | 0.192646021 | 0.923 | 0.886 | 7.48E-07 |
| S100A10 | 2.57E-09 | 0.205382951 | 0.926 | 0.897 | 1.28E-06 |
| NFKB1 | 4.42E-09 | -0.24099781 | 0.406 | 0.478 | 2.20E-06 |
| BIRC3 | 5.54E-09 | -0.253210837 | 0.309 | 0.371 | 2.76E-06 |
| IL18RAP | 6.78E-09 | -0.220971391 | 0.103 | 0.214 | 3.37E-06 |
| TRIB2 | 1.32E-08 | 0.205734572 | 0.37 | 0.274 | 6.56E-06 |
| CCL3 | 1.40E-08 | -0.311420464 | 0.308 | 0.353 | 6.98E-06 |
| LAT | 2.88E-08 | 0.19848727 | 0.798 | 0.757 | 1.43E-05 |

| | | | | | |
|---|---|---|---|---|---|
| GZMB | 4.18E-08 | 0.343368341 | 0.791 | 0.746 | 2.08E-05 |
| TRAC | 4.32E-08 | 0.24728431 | 0.918 | 0.909 | 2.15E-05 |
| IRF1 | 5.56E-08 | -0.223156946 | 0.791 | 0.812 | 2.77E-05 |
| GNAI2 | 8.61E-08 | 0.148738843 | 0.997 | 0.994 | 4.29E-05 |
| BTG1 | 1.04E-07 | -0.185745808 | 0.973 | 0.983 | 5.17E-05 |
| NFKBIZ | 1.99E-07 | -0.162025724 | 0.2 | 0.267 | 9.92E-05 |
| CREM | 2.57E-07 | -0.182989957 | 0.118 | 0.21 | 0.000127884 |
| PFN1 | 4.35E-07 | 0.149658219 | 1 | 1 | 0.000216505 |
| GBP2 | 4.89E-07 | -0.179705581 | 0.371 | 0.432 | 0.000243443 |
| RGS1 | 5.89E-07 | -0.201608472 | 0.099 | 0.151 | 0.000293545 |
| LAP3 | 1.05E-06 | -0.166878224 | 0.246 | 0.294 | 0.000522929 |
| ITGAM | 1.11E-06 | 0.139622616 | 0.213 | 0.132 | 0.000550664 |
| MS4A1 | 1.59E-06 | -0.201787552 | 0.176 | 0.205 | 0.000790516 |
| CXCL10 | 1.65E-06 | -0.166365764 | 0.044 | 0.067 | 0.000823663 |
| IGHG1 | 2.85E-06 | -0.20683286 | 0.235 | 0.234 | 0.001421398 |
| FCER1G | 3.27E-06 | -0.19161096 | 0.46 | 0.511 | 0.001630869 |
| CD40 | 4.08E-06 | -0.121541438 | 0.064 | 0.093 | 0.002029556 |
| NR4A2 | 4.30E-06 | -0.119495997 | 0.03 | 0.067 | 0.002140372 |
| CCL5 | 4.55E-06 | 0.20386647 | 0.924 | 0.906 | 0.002265371 |
| IRF8 | 4.70E-06 | -0.146510039 | 0.147 | 0.18 | 0.002341079 |
| SNX9 | 5.92E-06 | -0.093975462 | 0.066 | 0.133 | 0.002946493 |
| LDHA | 8.65E-06 | -0.167439643 | 0.864 | 0.868 | 0.00430855 |
| EIF5A | 1.08E-05 | -0.171671007 | 0.609 | 0.666 | 0.005383031 |
| CD2 | 1.42E-05 | 0.164414752 | 0.911 | 0.87 | 0.007060946 |
| CD74 | 1.78E-05 | -0.207897522 | 0.961 | 0.973 | 0.008863574 |
| CMKLR1 | 2.21E-05 | 0.075983901 | 0.091 | 0.048 | 0.011012849 |
| CD5 | 3.07E-05 | 0.167507957 | 0.458 | 0.396 | 0.015312022 |
| PRF1 | 3.65E-05 | 0.20094865 | 0.918 | 0.912 | 0.018158208 |
| SLA | 3.93E-05 | -0.15372001 | 0.479 | 0.529 | 0.019569123 |
| ZBTB16 | 5.15E-05 | -0.061059372 | 0.018 | 0.05 | 0.025626415 |
| NFATC2 | 5.75E-05 | -0.119700559 | 0.301 | 0.365 | 0.028641377 |
| BIN2 | 6.92E-05 | 0.117929611 | 0.416 | 0.363 | 0.034483928 |
| LGALS9 | 7.34E-05 | -0.107864968 | 0.12 | 0.157 | 0.036567683 |
| CCR7 | 7.43E-05 | -0.255232587 | 0.468 | 0.484 | 0.036987645 |
| IFNGR1 | 8.19E-05 | -0.116474614 | 0.247 | 0.306 | 0.040786432 |
| IL12RB1 | 9.06E-05 | -0.063481411 | 0.045 | 0.083 | 0.045137768 |
| CD7 | 0.000102374 | -0.116223005 | 0.187 | 0.245 | 0.050982411 |
| CD3D | 0.00010318 | 0.140910016 | 0.924 | 0.889 | 0.05138375 |
| FOS | 0.000122446 | -0.122972219 | 0.151 | 0.158 | 0.060978326 |
| SDCBP | 0.000129855 | -0.141886197 | 0.632 | 0.654 | 0.06466776 |
| KLRC3 | 0.000139643 | 0.253832877 | 0.589 | 0.544 | 0.069542201 |
| PSMB9 | 0.000140656 | -0.138592013 | 0.714 | 0.745 | 0.070046512 |
| ZAP70 | 0.000146212 | 0.109251932 | 0.398 | 0.334 | 0.072813404 |
| STAT4 | 0.000169524 | -0.121635335 | 0.453 | 0.516 | 0.084423067 |
| IGHG2 | 0.000177725 | -0.100764022 | 0.036 | 0.041 | 0.08850728 |
| IGKC | 0.000179722 | -0.217992945 | 0.618 | 0.608 | 0.089501612 |
| ARID5A | 0.000205532 | -0.094038702 | 0.16 | 0.21 | 0.102355108 |

| | | | | | |
|---|---|---|---|---|---|
| CD3G | 0.000218327 | 0.081053149 | 0.15 | 0.102 | 0.108726707 |
| DOCK8 | 0.000223562 | 0.115697458 | 0.743 | 0.691 | 0.111333794 |
| CD8B | 0.000335221 | 0.09135271 | 0.168 | 0.109 | 0.166940095 |
| CD79A | 0.000355243 | -0.143341775 | 0.16 | 0.188 | 0.17691078 |
| VNN2 | 0.000369453 | 0.093045613 | 0.231 | 0.174 | 0.183987411 |
| IL7R | 0.000405875 | -0.18554453 | 0.32 | 0.433 | 0.202125968 |
| VSIR | 0.000453521 | 0.104419227 | 0.386 | 0.323 | 0.225853295 |
| IGHG3 | 0.000506503 | -0.100120413 | 0.048 | 0.052 | 0.25223856 |
| TUBA1C | 0.000543404 | -0.1351137 | 0.461 | 0.475 | 0.270615204 |
| DUSP5 | 0.000652345 | -0.104051424 | 0.067 | 0.085 | 0.324867567 |
| SPAG9 | 0.00067027 | -0.073776159 | 0.12 | 0.169 | 0.333794387 |
| ITK | 0.00067048 | 0.118464424 | 0.729 | 0.67 | 0.333899032 |
| BCL6 | 0.000816163 | -0.035885377 | 0.016 | 0.04 | 0.406449235 |
| CXCR6 | 0.001161455 | -0.153247401 | 0.087 | 0.159 | 0.578404483 |
| CD86 | 0.001287062 | -0.071660023 | 0.04 | 0.05 | 0.640956665 |
| HLA.DPA1 | 0.001469903 | 0.147148655 | 0.837 | 0.735 | 0.732011934 |
| TGFBR2 | 0.001476909 | -0.106500594 | 0.678 | 0.727 | 0.735500643 |
| NFATC3 | 0.001485847 | 0.098796324 | 0.528 | 0.483 | 0.739951845 |
| IL6ST | 0.001560054 | -0.098974164 | 0.192 | 0.249 | 0.776906787 |
| LGALS1 | 0.001646088 | 0.151711277 | 0.691 | 0.629 | 0.819751908 |
| DUSP4 | 0.001830627 | -0.11207156 | 0.096 | 0.144 | 0.911652336 |
| TBX21 | 0.001947752 | 0.122110912 | 0.633 | 0.586 | 0.969980416 |
| CXCL9 | 0.0020367 | -0.095585484 | 0.016 | 0.018 | 1 |
| REL | 0.002143779 | -0.096786927 | 0.243 | 0.274 | 1 |
| IL2RB | 0.00216499 | -0.12463164 | 0.847 | 0.811 | 1 |
| IL15 | 0.002189153 | -0.035358119 | 0.013 | 0.028 | 1 |
| IKZF1 | 0.002350087 | 0.094001261 | 0.44 | 0.369 | 1 |
| LTA | 0.002694645 | -0.066459466 | 0.052 | 0.074 | 1 |
| HLA.DRA | 0.002702078 | -0.173545494 | 0.728 | 0.709 | 1 |
| IL15RA | 0.00275764 | -0.065896315 | 0.101 | 0.135 | 1 |
| MZB1 | 0.002946649 | -0.038977972 | 0.013 | 0.027 | 1 |
| BCL2A1 | 0.003051818 | -0.108335373 | 0.219 | 0.199 | 1 |
| STAT5A | 0.003545158 | -0.083110506 | 0.254 | 0.293 | 1 |
| CTSW | 0.003736973 | 0.11135798 | 0.607 | 0.557 | 1 |
| ADA | 0.003905931 | 0.091586905 | 0.344 | 0.276 | 1 |
| WAS | 0.004717365 | 0.088733829 | 0.553 | 0.497 | 1 |
| CCR9 | 0.004883603 | 0.02174761 | 0.011 | 0.002 | 1 |
| SKAP1 | 0.005031124 | 0.055324414 | 0.116 | 0.092 | 1 |
| ICAM1 | 0.005099032 | -0.073776159 | 0.118 | 0.129 | 1 |
| SP140 | 0.005147897 | -0.081636435 | 0.235 | 0.286 | 1 |
| TGIF1 | 0.005454276 | -0.103156167 | 0.423 | 0.48 | 1 |
| IGBP1 | 0.005495168 | 0.075656716 | 0.287 | 0.244 | 1 |
| APOBEC3G | 0.005891066 | 0.112141052 | 0.813 | 0.767 | 1 |
| CCR6 | 0.006319035 | -0.041243915 | 0.03 | 0.054 | 1 |
| IL3RA | 0.006508973 | -0.045162755 | 0.034 | 0.043 | 1 |
| NAMPT | 0.006528425 | -0.073540322 | 0.148 | 0.168 | 1 |
| CTSD | 0.006686283 | -0.092358701 | 0.362 | 0.395 | 1 |

| | | | | | |
|---|---|---|---|---|---|
| PIK3IP1 | 0.00716149 | -0.102695463 | 0.659 | 0.685 | 1 |
| RUNX3 | 0.007401546 | 0.085180917 | 0.881 | 0.868 | 1 |
| CD37 | 0.007526389 | 0.093998687 | 0.84 | 0.803 | 1 |
| CSF1 | 0.007527731 | -0.031323439 | 0.009 | 0.021 | 1 |
| NANS | 0.00781145 | -0.064005294 | 0.113 | 0.145 | 1 |
| CD44 | 0.00783195 | -0.088730382 | 0.927 | 0.937 | 1 |
| NGDN | 0.007832123 | 0.023478192 | 0.029 | 0.013 | 1 |
| CXCL5 | 0.008019055 | -0.111401818 | 0.482 | 0.527 | 1 |
| DUSP1 | 0.00804677 | -0.104377731 | 0.284 | 0.319 | 1 |
| CXCL8 | 0.008854262 | -0.121190429 | 0.417 | 0.461 | 1 |
| CD9 | 0.009472002 | -0.069636429 | 0.114 | 0.131 | 1 |
| IVNS1ABP | 0.009503785 | 0.082791485 | 0.428 | 0.382 | 1 |
| TRAF1 | 0.009768554 | -0.064016514 | 0.133 | 0.149 | 1 |
| ARHGEF3 | 0.010417887 | 0.083624097 | 0.517 | 0.452 | 1 |
| FURIN | 0.011552199 | -0.097535817 | 0.322 | 0.359 | 1 |
| ANXA5 | 0.011630567 | 0.091012866 | 0.653 | 0.585 | 1 |
| EVI2A | 0.01229552 | -0.087234009 | 0.502 | 0.536 | 1 |
| NINJ2 | 0.012820868 | 0.028360337 | 0.041 | 0.025 | 1 |
| SOCS3 | 0.013804665 | -0.035547909 | 0.028 | 0.043 | 1 |
| CXCL16 | 0.013899561 | -0.081927183 | 0.175 | 0.167 | 1 |
| ITGAX | 0.015470387 | 0.062553051 | 0.144 | 0.113 | 1 |
| JAK3 | 0.015955239 | -0.086634121 | 0.516 | 0.553 | 1 |
| NFKB2 | 0.016433485 | -0.033034203 | 0.036 | 0.052 | 1 |
| KLRC1 | 0.016575367 | -0.14087105 | 0.317 | 0.355 | 1 |
| GAB2 | 0.016668881 | -0.042908236 | 0.057 | 0.073 | 1 |
| MAP2K3 | 0.017230629 | -0.082302089 | 0.556 | 0.569 | 1 |
| GZMK | 0.019170925 | -0.18636104 | 0.235 | 0.318 | 1 |
| PRNP | 0.022096428 | -0.060629497 | 0.188 | 0.223 | 1 |
| RORA | 0.023034833 | -0.090780123 | 0.554 | 0.592 | 1 |
| CD24 | 0.024040447 | -0.021364718 | 0.009 | 0.017 | 1 |
| BACH2 | 0.024360892 | -0.032926217 | 0.035 | 0.054 | 1 |
| TCF4 | 0.02486914 | -0.019705423 | 0.004 | 0.011 | 1 |
| GNLY | 0.026181653 | 0.173101619 | 0.818 | 0.811 | 1 |
| TNF | 0.02773105 | -0.063526955 | 0.083 | 0.092 | 1 |
| ITGAE | 0.02844294 | -0.054723346 | 0.107 | 0.159 | 1 |
| ADGRG3 | 0.028543323 | -0.018384879 | 0.004 | 0.011 | 1 |
| OSM | 0.028699149 | 0.014190029 | 0.015 | 0.007 | 1 |
| CYBB | 0.029320132 | -0.06840197 | 0.249 | 0.264 | 1 |
| BCL2 | 0.033599221 | 0.040655826 | 0.094 | 0.079 | 1 |
| MMP9 | 0.036419325 | -0.046074709 | 0.111 | 0.123 | 1 |
| TNFSF8 | 0.036525652 | -0.014839589 | 0.005 | 0.014 | 1 |
| CD27 | 0.036961413 | 0.093858228 | 0.294 | 0.301 | 1 |
| IL6 | 0.037175046 | 0.027052956 | 0.029 | 0.026 | 1 |
| SLC25A37 | 0.038072812 | -0.048172128 | 0.124 | 0.146 | 1 |
| AURKB | 0.038524523 | 0.010461869 | 0.011 | 0.005 | 1 |
| ENTPD1 | 0.039236675 | -0.025370597 | 0.011 | 0.02 | 1 |
| CHD7 | 0.039303548 | -0.019326423 | 0.011 | 0.021 | 1 |

| | | | | | |
|---|---|---|---|---|---|
| SOCS2 | 0.039505125 | 0.030155226 | 0.052 | 0.031 | 1 |
| CSRNP1 | 0.043304417 | -0.049434549 | 0.137 | 0.164 | 1 |
| XBP1 | 0.043503836 | -0.068326182 | 0.369 | 0.388 | 1 |
| NCAM1 | 0.04651006 | -0.028351088 | 0.029 | 0.05 | 1 |
| HAVCR2 | 0.046729749 | -0.053417184 | 0.104 | 0.127 | 1 |
| JUNB | 0.049118044 | -0.051104959 | 0.164 | 0.171 | 1 |
| FASLG | 0.049509864 | -0.055696774 | 0.175 | 0.199 | 1 |
| BCAT1 | 0.049825235 | -0.016104201 | 0.007 | 0.017 | 1 |
| EEF1E1 | 0.05075973 | -0.024702925 | 0.028 | 0.043 | 1 |
| IL18 | 0.051098559 | -0.011335646 | 0.003 | 0.01 | 1 |
| CNOT2 | 0.052542342 | 0.052039183 | 0.313 | 0.295 | 1 |
| FUT4 | 0.054243411 | 0.026094292 | 0.052 | 0.036 | 1 |
| STAT3 | 0.057861315 | -0.070968545 | 0.579 | 0.593 | 1 |
| KDELR1 | 0.059811448 | 0.056308252 | 0.375 | 0.347 | 1 |
| ILF2 | 0.060494754 | -0.063632082 | 0.443 | 0.453 | 1 |
| EVI2B | 0.060539368 | 0.060960865 | 0.893 | 0.865 | 1 |
| CD70 | 0.062406067 | -0.021459085 | 0.013 | 0.019 | 1 |
| CD1C | 0.067733917 | -0.017598943 | 0.011 | 0.016 | 1 |
| NT5E | 0.067777724 | 0.022486429 | 0.037 | 0.028 | 1 |
| CXCR5 | 0.067903467 | -0.047837764 | 0.063 | 0.069 | 1 |
| CCL20 | 0.068576285 | -0.025136909 | 0.005 | 0.008 | 1 |
| TRIB1 | 0.069526086 | -0.01747533 | 0.017 | 0.027 | 1 |
| RBCK1 | 0.071374779 | -0.051137432 | 0.236 | 0.274 | 1 |
| PSMB8 | 0.072216298 | -0.059332241 | 0.865 | 0.885 | 1 |
| RORC | 0.075488857 | -0.023498959 | 0.014 | 0.024 | 1 |
| MGST1 | 0.07590575 | -0.024143786 | 0.026 | 0.033 | 1 |
| TNFSF13B | 0.076785693 | -0.033183135 | 0.057 | 0.064 | 1 |
| CD72 | 0.078883786 | -0.028847589 | 0.038 | 0.056 | 1 |
| TGFBR1 | 0.079399473 | 0.042932417 | 0.236 | 0.211 | 1 |
| ICOS | 0.082071214 | 0.034589798 | 0.102 | 0.082 | 1 |
| CXCL13 | 0.083128489 | -0.018803039 | 0.005 | 0.007 | 1 |
| FOSB | 0.083171069 | -0.017242525 | 0.007 | 0.009 | 1 |
| CD4 | 0.083606605 | 0.074057179 | 0.257 | 0.233 | 1 |
| MYB | 0.084976856 | 0.01197938 | 0.014 | 0.007 | 1 |
| IL2RA | 0.088241223 | -0.016582706 | 0.007 | 0.009 | 1 |
| YBX3 | 0.097811551 | 0.015804112 | 0.026 | 0.015 | 1 |
| PRDM1 | 0.098831126 | -0.048006453 | 0.248 | 0.253 | 1 |
| CCL2 | 0.102440188 | -0.078587495 | 0.793 | 0.776 | 1 |
| UFM1 | 0.103100514 | -0.052221005 | 0.493 | 0.492 | 1 |
| LGALS3 | 0.103154633 | -0.042707367 | 0.121 | 0.128 | 1 |
| NFKBID | 0.103242285 | -0.007807881 | 0.001 | 0.005 | 1 |
| TOP2A | 0.105784331 | 0.00890975 | 0.01 | 0.004 | 1 |
| MME | 0.107327489 | 0.006703171 | 0.004 | 0.002 | 1 |
| SRGN | 0.111645775 | -0.034610306 | 0.121 | 0.144 | 1 |
| B3GAT1 | 0.112758308 | 0.039606425 | 0.176 | 0.142 | 1 |
| CYCS | 0.118869314 | -0.048499151 | 0.346 | 0.355 | 1 |
| JAK2 | 0.122479802 | -0.008442075 | 0.004 | 0.01 | 1 |

| | | | | | |
|---|---|---|---|---|---|
| NIBAN3 | 0.122488822 | -0.009537993 | 0.005 | 0.011 | 1 |
| CXCL3 | 0.123058465 | -0.042862044 | 0.132 | 0.142 | 1 |
| BATF | 0.124831585 | 0.032735125 | 0.124 | 0.101 | 1 |
| IFNG | 0.125833883 | 0.036696962 | 0.083 | 0.041 | 1 |
| KLRB1 | 0.128656257 | -0.019807865 | 0.02 | 0.038 | 1 |
| IL1R1 | 0.129271664 | -0.013042607 | 0.008 | 0.011 | 1 |
| LRRC32 | 0.129801498 | -0.010670962 | 0.003 | 0.006 | 1 |
| RNASE6 | 0.139114363 | 0.013373957 | 0.028 | 0.02 | 1 |
| CD38 | 0.146779549 | -0.026638518 | 0.041 | 0.051 | 1 |
| NRP1 | 0.148294094 | 0.013954888 | 0.037 | 0.026 | 1 |
| VSIG4 | 0.14887862 | 0.013337239 | 0.032 | 0.024 | 1 |
| GRB10 | 0.15840361 | -0.006240586 | 0.002 | 0.006 | 1 |
| IL6R | 0.158886154 | 0.020106426 | 0.052 | 0.037 | 1 |
| CTLA4 | 0.159095636 | 0.03232312 | 0.085 | 0.077 | 1 |
| EIF5 | 0.159745727 | -0.046133555 | 0.458 | 0.457 | 1 |
| IGHG4 | 0.163368208 | -0.009107065 | 0.004 | 0.007 | 1 |
| FOSL1 | 0.164205514 | -0.007342945 | 0.003 | 0.007 | 1 |
| CD200 | 0.16605922 | -0.014024734 | 0.015 | 0.018 | 1 |
| PIK3AP1 | 0.167932656 | 0.035173744 | 0.196 | 0.165 | 1 |
| LEF1 | 0.169937632 | 0.065076499 | 0.404 | 0.339 | 1 |
| CD80 | 0.173032959 | -0.010005973 | 0.003 | 0.005 | 1 |
| TNFRSF25 | 0.181713575 | -0.020764389 | 0.049 | 0.059 | 1 |
| SLC1A5 | 0.183727803 | 0.040193007 | 0.234 | 0.185 | 1 |
| TIAF1 | 0.185424523 | 0.031459553 | 0.174 | 0.152 | 1 |
| TNFRSF13C | 0.193800314 | -0.028974396 | 0.081 | 0.091 | 1 |
| CD48 | 0.194280655 | -0.04106477 | 0.971 | 0.961 | 1 |
| CD6 | 0.201022001 | 0.051358866 | 0.652 | 0.617 | 1 |
| IL4R | 0.203410041 | -0.03967915 | 0.313 | 0.318 | 1 |
| GIMAP2 | 0.206322018 | -0.044783226 | 0.597 | 0.653 | 1 |
| PCNA | 0.209146863 | -0.036109397 | 0.221 | 0.237 | 1 |
| IL2RG | 0.216183356 | 0.039466325 | 0.952 | 0.934 | 1 |
| PDIA4 | 0.216453683 | -0.045040815 | 0.425 | 0.43 | 1 |
| GIMAP5 | 0.216532378 | -0.049131482 | 0.591 | 0.639 | 1 |
| CST7 | 0.219749531 | 0.045723943 | 0.958 | 0.943 | 1 |
| MITF | 0.22121735 | -0.009660514 | 0.013 | 0.02 | 1 |
| IGHA1 | 0.222961733 | -0.011867315 | 0.013 | 0.014 | 1 |
| CCR2 | 0.225633858 | 0.006018892 | 0.01 | 0.005 | 1 |
| FBXO11 | 0.226093355 | 0.032647585 | 0.248 | 0.237 | 1 |
| TIGIT | 0.227773215 | 0.044247988 | 0.333 | 0.285 | 1 |
| CXCR3 | 0.228621729 | -0.055034409 | 0.371 | 0.389 | 1 |
| CD274 | 0.234077223 | 0.022613242 | 0.087 | 0.073 | 1 |
| JAK1 | 0.24630241 | -0.033571283 | 0.939 | 0.94 | 1 |
| RPL6 | 0.25207986 | 0.033671355 | 0.999 | 1 | 1 |
| CCR1 | 0.258564087 | -0.023225457 | 0.103 | 0.115 | 1 |
| TMEM97 | 0.262518967 | -0.013039795 | 0.018 | 0.025 | 1 |
| FCN1 | 0.26288735 | -0.028132967 | 0.107 | 0.099 | 1 |
| BLNK | 0.264643622 | -0.013593 | 0.027 | 0.031 | 1 |

| | | | | | |
|---|---|---|---|---|---|
| CCL13 | 0.274918242 | -0.009233457 | 0.012 | 0.016 | 1 |
| IL18R1 | 0.276209848 | -0.023221408 | 0.081 | 0.103 | 1 |
| IFITM3 | 0.278682789 | -0.044588203 | 0.274 | 0.265 | 1 |
| PCLAF | 0.280107782 | 0.005131804 | 0.009 | 0.005 | 1 |
| EBF1 | 0.283287353 | -0.005342443 | 0.003 | 0.006 | 1 |
| POU2F2 | 0.283475396 | -0.020659486 | 0.085 | 0.097 | 1 |
| EIF3J | 0.284132271 | -0.014576069 | 0.046 | 0.054 | 1 |
| IL11RA | 0.284477068 | 0.031102702 | 0.243 | 0.235 | 1 |
| PDIA6 | 0.284940211 | -0.037547888 | 0.534 | 0.494 | 1 |
| IL1RN | 0.289226708 | -0.018341331 | 0.063 | 0.072 | 1 |
| TLR2 | 0.289982461 | 0.008818773 | 0.023 | 0.016 | 1 |
| BTLA | 0.290811095 | 0.009454705 | 0.025 | 0.019 | 1 |
| BCL11B | 0.296118855 | -0.038781928 | 0.53 | 0.521 | 1 |
| AIM2 | 0.298704882 | 0.002687421 | 0.003 | 0.001 | 1 |
| HNRNPC | 0.300748367 | -0.03612948 | 0.57 | 0.551 | 1 |
| TGFB1 | 0.301224503 | -0.011044576 | 0.024 | 0.03 | 1 |
| SLC7A7 | 0.302431588 | -0.018280679 | 0.064 | 0.074 | 1 |
| FOXP3 | 0.306996349 | -0.020253299 | 0.012 | 0.011 | 1 |
| EOMES | 0.309804234 | -0.006418651 | 0.007 | 0.011 | 1 |
| BCL2L1 | 0.310841581 | -0.011796062 | 0.032 | 0.041 | 1 |
| LIPA | 0.316884852 | -0.034183836 | 0.508 | 0.51 | 1 |
| TCF7 | 0.330489783 | 0.020815216 | 0.116 | 0.097 | 1 |
| CD63 | 0.339792282 | -0.034783617 | 0.802 | 0.805 | 1 |
| MALT1 | 0.340760066 | -0.017262119 | 0.084 | 0.101 | 1 |
| GZMA | 0.341536996 | 0.044064656 | 0.55 | 0.522 | 1 |
| SELL | 0.346216307 | 0.051515893 | 0.481 | 0.467 | 1 |
| IL1A | 0.353896687 | -0.005319358 | 0.007 | 0.009 | 1 |
| CD36 | 0.354270002 | -0.020863032 | 0.124 | 0.136 | 1 |
| TGFBI | 0.357350868 | -0.031912934 | 0.338 | 0.35 | 1 |
| LTB | 0.359254407 | -0.022862307 | 0.094 | 0.1 | 1 |
| MCM2 | 0.366314262 | -0.016488705 | 0.075 | 0.08 | 1 |
| JCHAIN | 0.373177156 | -0.015369371 | 0.033 | 0.029 | 1 |
| TNIK | 0.374953546 | -0.020736026 | 0.121 | 0.137 | 1 |
| CCR5 | 0.382035139 | -0.025216412 | 0.111 | 0.133 | 1 |
| KCNE3 | 0.38428403 | -0.007225932 | 0.016 | 0.02 | 1 |
| ARPC1B | 0.386159386 | 0.030515668 | 0.77 | 0.752 | 1 |
| TNFRSF17 | 0.389222918 | -0.003796678 | 0.001 | 0.002 | 1 |
| THBS1 | 0.398799736 | -0.032870474 | 0.485 | 0.505 | 1 |
| ARL4C | 0.401789197 | 0.028004039 | 0.788 | 0.767 | 1 |
| SPP1 | 0.402170383 | -0.007544861 | 0.005 | 0.004 | 1 |
| ZNF341 | 0.40555086 | -0.00909367 | 0.023 | 0.028 | 1 |
| PTPRC | 0.415374657 | 0.022032388 | 0.99 | 0.99 | 1 |
| CR2 | 0.418493385 | 0.006855482 | 0.016 | 0.014 | 1 |
| CCND2 | 0.430725722 | -0.025259917 | 0.21 | 0.22 | 1 |
| IL23R | 0.431219619 | -0.002898692 | 0.002 | 0.004 | 1 |
| PTPN7 | 0.435606135 | -0.024614599 | 0.357 | 0.381 | 1 |
| EIF1 | 0.435705937 | 0.028070291 | 0.94 | 0.927 | 1 |

| | | | | | |
|---|---|---|---|---|---|
| HLA.DQB1 | 0.4387077 | -0.037067976 | 0.366 | 0.297 | 1 |
| B4GALT5 | 0.439522759 | 0.005761905 | 0.018 | 0.013 | 1 |
| CD14 | 0.444249565 | -0.028495619 | 0.384 | 0.409 | 1 |
| RPN2 | 0.446385999 | -0.024597019 | 0.519 | 0.544 | 1 |
| FAS | 0.451681375 | 0.014232744 | 0.096 | 0.073 | 1 |
| PTGDR2 | 0.452633311 | -0.00356769 | 0.002 | 0.003 | 1 |
| IRF4 | 0.459643548 | -0.021362905 | 0.131 | 0.129 | 1 |
| ERBIN | 0.465172996 | 0.024286963 | 0.526 | 0.516 | 1 |
| CLEC4E | 0.468153869 | -0.006137544 | 0.014 | 0.017 | 1 |
| IER3 | 0.468687535 | -0.025290876 | 0.263 | 0.257 | 1 |
| CCR4 | 0.470211468 | -0.017329585 | 0.049 | 0.045 | 1 |
| IL23A | 0.471640527 | 0.013919153 | 0.07 | 0.068 | 1 |
| PAXBP1 | 0.48434157 | -0.021557484 | 0.294 | 0.291 | 1 |
| KIT | 0.486154726 | -0.004861506 | 0.004 | 0.011 | 1 |
| TCL1A | 0.493268286 | 0.014458321 | 0.055 | 0.051 | 1 |
| TLR7 | 0.494173127 | -0.003550318 | 0.005 | 0.007 | 1 |
| STIM1 | 0.497014911 | -0.015791108 | 0.195 | 0.212 | 1 |
| TYMS | 0.497328378 | 0.003574658 | 0.007 | 0.004 | 1 |
| ORAI1 | 0.498684268 | 0.020612891 | 0.421 | 0.392 | 1 |
| SNX8 | 0.503048149 | -0.014114737 | 0.069 | 0.078 | 1 |
| POU2AF1 | 0.504873724 | 0.012417186 | 0.065 | 0.052 | 1 |
| PAK1IP1 | 0.507032096 | -0.00923618 | 0.048 | 0.054 | 1 |
| BCL10 | 0.510840996 | 0.011747567 | 0.096 | 0.086 | 1 |
| AHI1 | 0.518149246 | 0.007619466 | 0.038 | 0.031 | 1 |
| DPP4 | 0.521147683 | -0.011784912 | 0.049 | 0.059 | 1 |
| CCR3 | 0.532136171 | 0.002016556 | 0.002 | 0.002 | 1 |
| IL1B | 0.53404041 | 0.023739293 | 0.222 | 0.215 | 1 |
| RAG1 | 0.537301819 | -0.003111095 | 0.004 | 0.005 | 1 |
| LILRB4 | 0.537655603 | -0.01038868 | 0.071 | 0.079 | 1 |
| CASP5 | 0.544033055 | 0.013946287 | 0.208 | 0.198 | 1 |
| CSF2 | 0.547286481 | -0.01089492 | 0.011 | 0.011 | 1 |
| CD40LG | 0.5531112 | 0.005089087 | 0.02 | 0.016 | 1 |
| TRAT1 | 0.560663839 | -0.009473219 | 0.06 | 0.067 | 1 |
| CXCR2 | 0.563099835 | -0.005634577 | 0.021 | 0.026 | 1 |
| MYC | 0.57019114 | 0.025416608 | 0.21 | 0.185 | 1 |
| STAT6 | 0.570265253 | 0.015530637 | 0.254 | 0.244 | 1 |
| IKZF2 | 0.57849219 | 0.004008444 | 0.011 | 0.008 | 1 |
| ATF6B | 0.585029871 | 0.016031448 | 0.393 | 0.381 | 1 |
| CD160 | 0.593638607 | -0.005909605 | 0.012 | 0.013 | 1 |
| PSME3 | 0.596414661 | -0.011103991 | 0.126 | 0.127 | 1 |
| CXCL2 | 0.602403469 | -0.011779016 | 0.131 | 0.128 | 1 |
| EGR3 | 0.609527258 | -0.003550318 | 0.004 | 0.005 | 1 |
| LAT2 | 0.609716641 | -0.012247012 | 0.143 | 0.147 | 1 |
| RAPH1 | 0.610649027 | -0.003306039 | 0.009 | 0.011 | 1 |
| ATF4 | 0.615434837 | -0.019394271 | 0.785 | 0.77 | 1 |
| GEM | 0.62065105 | 0.003568901 | 0.007 | 0.007 | 1 |
| VPREB3 | 0.622101026 | -0.009657844 | 0.055 | 0.056 | 1 |

| | | | | | |
|---|---|---|---|---|---|
| FCER1A | 0.627426889 | 0.001121826 | 0.002 | 0.001 | 1 |
| TLR8 | 0.630535943 | -0.001116334 | 0.001 | 0.002 | 1 |
| HLA.DMA | 0.63534326 | 0.019358612 | 0.387 | 0.342 | 1 |
| CARD11 | 0.641280638 | -0.003306039 | 0.009 | 0.01 | 1 |
| TNFRSF8 | 0.646176077 | -0.001559764 | 0.002 | 0.003 | 1 |
| TNFSF14 | 0.658342757 | -0.004326603 | 0.023 | 0.024 | 1 |
| CXCL1 | 0.663931871 | -0.017345296 | 0.292 | 0.307 | 1 |
| CD300A | 0.668005035 | -0.017173085 | 0.546 | 0.521 | 1 |
| VMO1 | 0.677769821 | 0.010764774 | 0.113 | 0.099 | 1 |
| IFRD1 | 0.682082917 | -0.009013752 | 0.164 | 0.155 | 1 |
| PGM3 | 0.688465037 | -0.005843689 | 0.047 | 0.05 | 1 |
| SLC7A1 | 0.695585473 | 0.006767819 | 0.069 | 0.066 | 1 |
| MCM4 | 0.700177028 | -0.004083563 | 0.026 | 0.027 | 1 |
| F5 | 0.701813971 | -0.00242963 | 0.007 | 0.007 | 1 |
| CD247 | 0.701889746 | -0.013220283 | 0.56 | 0.536 | 1 |
| AQP9 | 0.702113308 | -0.004853891 | 0.04 | 0.042 | 1 |
| HLA.A | 0.703684042 | -0.008941217 | 1 | 1 | 1 |
| CD163 | 0.726878312 | -0.00597113 | 0.089 | 0.089 | 1 |
| TNFSF10 | 0.729277658 | 0.010919289 | 0.24 | 0.24 | 1 |
| IRF3 | 0.737912184 | 0.00605503 | 0.086 | 0.092 | 1 |
| CD79B | 0.743437694 | -0.00800662 | 0.102 | 0.107 | 1 |
| GATA3 | 0.748664289 | 0.003314031 | 0.03 | 0.028 | 1 |
| VPS28 | 0.751467484 | 0.010022528 | 0.639 | 0.641 | 1 |
| PSEN1 | 0.755238184 | 0.006017589 | 0.094 | 0.096 | 1 |
| LRRC8B | 0.755968123 | 0.002453729 | 0.017 | 0.015 | 1 |
| THBD | 0.759401002 | -0.000889834 | 0.002 | 0.003 | 1 |
| LIF | 0.759401002 | -0.000889834 | 0.002 | 0.003 | 1 |
| MAST4 | 0.772188168 | -0.0031934 | 0.032 | 0.031 | 1 |
| CD244 | 0.773989134 | -0.007860754 | 0.196 | 0.189 | 1 |
| QPCT | 0.788736304 | 0.001793003 | 0.01 | 0.01 | 1 |
| THEMIS | 0.79420595 | -0.004594716 | 0.074 | 0.075 | 1 |
| UGCG | 0.795013767 | -0.006973875 | 0.186 | 0.169 | 1 |
| JUN | 0.800550919 | 0.007097035 | 0.125 | 0.122 | 1 |
| CD22 | 0.82613009 | 0.004635753 | 0.074 | 0.08 | 1 |
| BLK | 0.828548285 | -0.000445469 | 0.001 | 0.001 | 1 |
| TNIP3 | 0.828548285 | -0.000445469 | 0.001 | 0.001 | 1 |
| CLEC4D | 0.828548285 | -0.000445469 | 0.001 | 0.001 | 1 |
| CD209 | 0.838923066 | -0.000881131 | 0.005 | 0.006 | 1 |
| TREM1 | 0.838923066 | -0.000881131 | 0.005 | 0.006 | 1 |
| PDCD1 | 0.845080708 | -0.000663789 | 0.003 | 0.004 | 1 |
| CBLB | 0.848471005 | -0.002102665 | 0.033 | 0.034 | 1 |
| CD109 | 0.852526859 | -0.00130291 | 0.012 | 0.013 | 1 |
| ADGRE1 | 0.856249764 | 0.001572593 | 0.015 | 0.013 | 1 |
| ABCE1 | 0.857196932 | -0.004584893 | 0.252 | 0.246 | 1 |
| FOXO1 | 0.861265838 | -0.003589078 | 0.104 | 0.102 | 1 |
| IGHD | 0.873169552 | -0.004706965 | 0.082 | 0.082 | 1 |
| IGHM | 0.877794619 | -0.006622805 | 0.254 | 0.254 | 1 |

| | | | | | |
|---|---|---|---|---|---|
| TIMM9 | 0.878005529 | 0.004435605 | 0.271 | 0.264 | 1 |
| FCER2 | 0.880740147 | -0.001882655 | 0.025 | 0.033 | 1 |
| CMTM2 | 0.881361755 | 0.000450961 | 0.002 | 0.001 | 1 |
| SMAD2 | 0.882467082 | -0.003690182 | 0.218 | 0.222 | 1 |
| LY86 | 0.88333924 | -0.002545932 | 0.076 | 0.071 | 1 |
| LIG4 | 0.887670956 | 0.002408356 | 0.084 | 0.081 | 1 |
| DUSP6 | 0.890087815 | -0.001464381 | 0.028 | 0.03 | 1 |
| LYN | 0.904456246 | -0.003696833 | 0.275 | 0.282 | 1 |
| CHD1 | 0.908828847 | -0.000438199 | 0.004 | 0.005 | 1 |
| S100A9 | 0.909647581 | 0.00442528 | 0.372 | 0.36 | 1 |
| BAX | 0.910921519 | 0.003696295 | 0.437 | 0.421 | 1 |
| CD28 | 0.939665606 | -0.001307568 | 0.071 | 0.072 | 1 |
| SNCA | 0.945697518 | 0.000463943 | 0.011 | 0.01 | 1 |
| TNFAIP8 | 0.951878277 | 0.002153362 | 0.508 | 0.49 | 1 |
| FN1 | 0.955297524 | 0.001946932 | 0.309 | 0.291 | 1 |
| IL2 | 0.95888222 | 0.00092273 | 0.018 | 0.014 | 1 |
| CHI3L2 | 0.961418214 | 0.000974569 | 0.07 | 0.074 | 1 |
| FOXP1 | 0.965938766 | 0.000762875 | 0.078 | 0.071 | 1 |
| TGFB3 | 0.973227784 | -0.000206574 | 0.008 | 0.007 | 1 |
| GAPDH | 0.973690065 | 0.001249666 | 0.977 | 0.966 | 1 |
| IL12A | 0.974680449 | -0.000200129 | 0.012 | 0.012 | 1 |
| CD33 | 0.976827063 | 0.000260666 | 0.021 | 0.023 | 1 |
| S100A12 | 0.980372819 | 0.00027037 | 0.025 | 0.027 | 1 |
| LAMP1 | 0.98677642 | 0.000557184 | 0.501 | 0.5 | 1 |
| EGR1 | 0.989828517 | -0.015372851 | 0 | 0.008 | 1 |
| DCLRE1C | 0.992168491 | -0.000193086 | 0.108 | 0.113 | 1 |
| F13A1 | 0.993772556 | -0.006703985 | 0 | 0.003 | 1 |
| SOCS1 | 0.993949939 | -0.004026128 | 0 | 0.003 | 1 |
| TNFRSF4 | 0.993949939 | -0.004026128 | 0 | 0.002 | 1 |
| CXCL11 | 0.99592682 | -0.010044332 | 0 | 0.002 | 1 |
| CLEC10A | 0.995952274 | 6.21E-05 | 0.032 | 0.029 | 1 |
| KLRG1 | 0.996139049 | -0.000150021 | 0.269 | 0.269 | 1 |
| CD34 | 0.996205209 | -0.002685334 | 0 | 0.002 | 1 |
| CCR8 | 0.996205209 | -0.002685334 | 0 | 0.002 | 1 |
| NDFIP2 | 0.996205209 | -0.002685334 | 0 | 0.002 | 1 |
| NFATC1 | 0.999501759 | -1.53E-05 | 0.198 | 0.196 | 1 |

Naïve B-cells

| | p_val | avg_log2FC | Patient | Controls | p_val_adj |
|---|---|---|---|---|---|
| IGHD | 5.28E-34 | 0.799974926 | 0.996 | 0.928 | 2.63E-31 |
| TCL1A | 4.90E-29 | 0.929785582 | 0.933 | 0.822 | 2.44E-26 |
| STAT1 | 2.22E-22 | -1.002926911 | 0.531 | 0.706 | 1.10E-19 |
| CD52 | 1.12E-18 | 0.534301533 | 0.992 | 0.988 | 5.57E-16 |
| IRF1 | 5.00E-18 | -0.824056472 | 0.681 | 0.778 | 2.49E-15 |
| CD37 | 7.79E-17 | 0.431112928 | 0.988 | 0.994 | 3.88E-14 |
| IGHM | 1.12E-15 | 0.539112397 | 0.996 | 0.986 | 5.57E-13 |

| | | | | | |
|---|---|---|---|---|---|
| CD79A | 4.00E-15 | 0.421646298 | 1 | 0.997 | 1.99E-12 |
| ITGB2 | 6.99E-13 | 0.5142768 | 0.677 | 0.534 | 3.48E-10 |
| CD79B | 1.35E-11 | 0.401916196 | 0.953 | 0.898 | 6.74E-09 |
| GBP1 | 4.23E-10 | -0.633830021 | 0.307 | 0.385 | 2.11E-07 |
| CD27 | 5.35E-10 | -0.46553621 | 0.114 | 0.252 | 2.66E-07 |
| CD74 | 5.50E-10 | 0.363607186 | 1 | 1 | 2.74E-07 |
| SELL | 8.12E-10 | 0.408274168 | 0.811 | 0.773 | 4.05E-07 |
| IRF7 | 5.89E-09 | -0.42765207 | 0.472 | 0.577 | 2.93E-06 |
| CD22 | 6.36E-09 | 0.48410464 | 0.866 | 0.783 | 3.17E-06 |
| HLA.DMA | 1.41E-08 | 0.371989043 | 0.984 | 0.93 | 7.02E-06 |
| TRIB2 | 2.85E-08 | 0.30866251 | 0.343 | 0.171 | 1.42E-05 |
| IFI6 | 1.04E-07 | -0.336156157 | 0.094 | 0.214 | 5.19E-05 |
| IGHA1 | 1.15E-07 | -0.368883475 | 0.024 | 0.114 | 5.71E-05 |
| HLA.DQB1 | 1.27E-07 | 0.41476816 | 0.996 | 0.951 | 6.32E-05 |
| IGHG2 | 1.33E-07 | -0.614319502 | 0.209 | 0.378 | 6.60E-05 |
| MS4A1 | 1.78E-07 | 0.361417761 | 0.992 | 0.991 | 8.87E-05 |
| IL4R | 2.36E-07 | 0.373402621 | 0.78 | 0.67 | 0.000117358 |
| CXCR4 | 2.88E-07 | -0.374754612 | 0.961 | 0.969 | 0.000143186 |
| IGHG3 | 4.39E-07 | -0.620872085 | 0.224 | 0.393 | 0.000218822 |
| CR2 | 2.50E-06 | 0.259640514 | 0.425 | 0.235 | 0.001247081 |
| CD72 | 3.11E-06 | 0.3805002 | 0.567 | 0.431 | 0.001546655 |
| IRF9 | 4.36E-06 | -0.326694441 | 0.689 | 0.789 | 0.002172279 |
| GBP2 | 6.15E-06 | -0.394221256 | 0.327 | 0.452 | 0.003062283 |
| JCHAIN | 1.43E-05 | -0.261800068 | 0.071 | 0.204 | 0.007117499 |
| GAPDH | 2.05E-05 | 0.521430167 | 0.965 | 0.93 | 0.010196705 |
| LGALS1 | 2.14E-05 | -0.407272466 | 0.437 | 0.472 | 0.010668916 |
| CHI3L2 | 2.52E-05 | -0.188957622 | 0.012 | 0.103 | 0.012538252 |
| BLNK | 3.89E-05 | 0.283789007 | 0.622 | 0.567 | 0.019361123 |
| UGCG | 4.06E-05 | 0.346140905 | 0.437 | 0.401 | 0.020236331 |
| HLA.A | 5.36E-05 | 0.259795925 | 1 | 1 | 0.026699613 |
| RNASE6 | 8.09E-05 | -0.157975432 | 0.063 | 0.15 | 0.04027758 |
| TNFAIP8 | 8.25E-05 | 0.279932677 | 0.724 | 0.616 | 0.041081492 |
| HLA.DRA | 0.000113746 | 0.272304009 | 0.996 | 0.996 | 0.056645534 |
| PCNA | 0.000123735 | 0.409570059 | 0.299 | 0.278 | 0.061620064 |
| LAT2 | 0.000149027 | 0.226042491 | 0.602 | 0.532 | 0.074215635 |
| CXCL10 | 0.000257787 | -0.353219881 | 0.051 | 0.082 | 0.128378022 |
| CD44 | 0.000287527 | 0.245315185 | 0.937 | 0.905 | 0.143188414 |
| NFATC3 | 0.0003536 | 0.191894047 | 0.504 | 0.43 | 0.176092782 |
| BCL2 | 0.000370529 | -0.232250408 | 0.331 | 0.416 | 0.18452357 |
| RBCK1 | 0.000404398 | -0.201318548 | 0.236 | 0.328 | 0.20138996 |
| MAP2K3 | 0.000527829 | 0.36220315 | 0.646 | 0.573 | 0.262858825 |
| GNAI2 | 0.000588536 | 0.214978732 | 0.996 | 0.983 | 0.293090772 |
| TNFRSF17 | 0.000676705 | 0.09967801 | 0.094 | 0.045 | 0.336999073 |
| IL3RA | 0.000699463 | 0.175012433 | 0.197 | 0.097 | 0.348332796 |
| IGLC3 | 0.000746572 | 0.637834071 | 0.76 | 0.743 | 0.371792753 |
| CYCS | 0.000827287 | 0.376907983 | 0.433 | 0.356 | 0.411989031 |
| LY86 | 0.000903406 | 0.201982412 | 0.587 | 0.495 | 0.44989613 |

| | | | | | |
|---|---|---|---|---|---|
| PDIA4 | 0.001036565 | 0.315883717 | 0.433 | 0.392 | 0.516209218 |
| DOCK8 | 0.001217524 | 0.181995766 | 0.772 | 0.663 | 0.60632694 |
| LTA | 0.0013395 | 0.229271994 | 0.075 | 0.098 | 0.667070914 |
| LDHA | 0.001898134 | 0.453812158 | 0.736 | 0.732 | 0.945270582 |
| LGALS9 | 0.00191194 | 0.214372541 | 0.575 | 0.468 | 0.952145875 |
| CCL22 | 0.012301574 | -0.77712935 | 0.201 | 0.198 | 1 |
| CCL4 | 0.002622 | -0.761920949 | 0.346 | 0.318 | 1 |
| TBX21 | 0.010125827 | -0.320480378 | 0.11 | 0.136 | 1 |
| CXCL9 | 0.00384342 | -0.239625048 | 0.012 | 0.025 | 1 |
| NFKBIA | 0.015262936 | -0.208464155 | 0.764 | 0.797 | 1 |
| RUNX3 | 0.026101888 | -0.191013505 | 0.555 | 0.608 | 1 |
| PSMB9 | 0.037999192 | -0.171507779 | 0.752 | 0.713 | 1 |
| CD86 | 0.018677239 | -0.14567531 | 0.185 | 0.222 | 1 |
| CD200 | 0.03213046 | -0.133602478 | 0.185 | 0.216 | 1 |
| NFATC2 | 0.003348865 | -0.121962028 | 0.118 | 0.186 | 1 |
| RGS1 | 0.013711545 | -0.107986889 | 0.063 | 0.112 | 1 |
| NFKB2 | 0.005904287 | -0.10521964 | 0.063 | 0.111 | 1 |
| ITGAE | 0.047942257 | -0.070006199 | 0.087 | 0.123 | 1 |
| SOCS3 | 0.032128967 | -0.052570032 | 0.02 | 0.047 | 1 |
| CX3CR1 | 0.031653038 | -0.041646028 | 0.008 | 0.037 | 1 |
| F13A1 | 0.018880083 | 0.028160842 | 0.016 | 0.004 | 1 |
| NRP1 | 0.048193361 | 0.039180174 | 0.059 | 0.028 | 1 |
| PAX5 | 0.033015567 | 0.047150789 | 0.067 | 0.033 | 1 |
| MMP9 | 0.018725504 | 0.076379218 | 0.134 | 0.088 | 1 |
| ZBED2 | 0.002679842 | 0.076526378 | 0.051 | 0.019 | 1 |
| IFNG | 0.002363446 | 0.080588403 | 0.039 | 0.018 | 1 |
| ITGAX | 0.024834666 | 0.090125058 | 0.098 | 0.072 | 1 |
| TMEM97 | 0.036674121 | 0.09374009 | 0.083 | 0.068 | 1 |
| IL6 | 0.018907185 | 0.094221226 | 0.091 | 0.037 | 1 |
| CXCL3 | 0.015238398 | 0.095171931 | 0.165 | 0.113 | 1 |
| LTB | 0.019886901 | 0.09844552 | 0.193 | 0.13 | 1 |
| IKZF1 | 0.044848181 | 0.104490089 | 0.295 | 0.278 | 1 |
| TIAF1 | 0.010110278 | 0.111839337 | 0.213 | 0.151 | 1 |
| SPAG9 | 0.01293235 | 0.11644859 | 0.209 | 0.156 | 1 |
| TGFBR1 | 0.004432271 | 0.11684663 | 0.197 | 0.132 | 1 |
| ATF6B | 0.021333292 | 0.123100298 | 0.488 | 0.419 | 1 |
| BACH2 | 0.011546531 | 0.127881857 | 0.335 | 0.268 | 1 |
| HIF1A | 0.024861097 | 0.128222236 | 0.657 | 0.605 | 1 |
| PTPRC | 0.022483627 | 0.128844906 | 0.917 | 0.902 | 1 |
| PIK3AP1 | 0.038064707 | 0.13004109 | 0.354 | 0.347 | 1 |
| NT5E | 0.013490913 | 0.130392315 | 0.264 | 0.171 | 1 |
| TRIB1 | 0.008662446 | 0.13412877 | 0.232 | 0.135 | 1 |
| PAK1IP1 | 0.023034286 | 0.13601915 | 0.146 | 0.096 | 1 |
| FTH1 | 0.020847289 | 0.139111138 | 0.98 | 0.973 | 1 |
| ORAI1 | 0.032830482 | 0.140298902 | 0.516 | 0.432 | 1 |
| BIRC3 | 0.041408033 | 0.140353666 | 0.815 | 0.735 | 1 |
| IFNGR1 | 0.014641215 | 0.143439104 | 0.472 | 0.433 | 1 |

| | | | | | |
|---|---|---|---|---|---|
| TIMM9 | 0.038661377 | 0.147278361 | 0.39 | 0.377 | 1 |
| IL2RG | 0.020731356 | 0.14766599 | 0.87 | 0.862 | 1 |
| SLA | 0.010536273 | 0.152903362 | 0.461 | 0.359 | 1 |
| EVI2A | 0.007832256 | 0.154447943 | 0.406 | 0.341 | 1 |
| RPL6 | 0.026784929 | 0.161268887 | 1 | 1 | 1 |
| PSME3 | 0.037967479 | 0.165768704 | 0.24 | 0.18 | 1 |
| ERBIN | 0.005964894 | 0.166514478 | 0.535 | 0.469 | 1 |
| EIF5 | 0.006810476 | 0.184537817 | 0.539 | 0.497 | 1 |
| NANS | 0.011831935 | 0.185110686 | 0.264 | 0.199 | 1 |
| RPN2 | 0.007172255 | 0.190557649 | 0.579 | 0.497 | 1 |
| PLCG2 | 0.004981424 | 0.198241844 | 0.713 | 0.623 | 1 |
| ATF4 | 0.020029375 | 0.204060484 | 0.791 | 0.744 | 1 |
| ARPC1B | 0.025984214 | 0.204609919 | 0.72 | 0.733 | 1 |
| SP140 | 0.002341112 | 0.205715929 | 0.685 | 0.621 | 1 |
| EIF1 | 0.009129859 | 0.213131738 | 0.941 | 0.893 | 1 |
| HLA.DPA1 | 0.002751078 | 0.223034924 | 1 | 0.986 | 1 |
| MCM4 | 0.018913085 | 0.238316401 | 0.087 | 0.059 | 1 |
| PFN1 | 0.008513837 | 0.246409414 | 0.996 | 0.999 | 1 |
| ABCE1 | 0.006934599 | 0.252631361 | 0.358 | 0.303 | 1 |
| VPREB3 | 0.006612902 | 0.256321351 | 0.744 | 0.617 | 1 |
| ILF2 | 0.023839439 | 0.256937009 | 0.547 | 0.523 | 1 |
| EIF5A | 0.031846135 | 0.312540555 | 0.661 | 0.636 | 1 |
| MCM2 | 0.002837235 | 0.339332226 | 0.28 | 0.258 | 1 |
| IGKC | 0.013961072 | 0.362729073 | 0.866 | 0.857 | 1 |

Memory B-cells

| | p_val | avg_log2FC | Patient | Controls | p_val_adj |
|---|---|---|---|---|---|
| CD37 | 4.56E-09 | 1.155719718 | 1 | 0.98 | 2.27E-06 |
| CD52 | 4.93E-08 | 1.273860231 | 1 | 0.992 | 2.45E-05 |
| CD79B | 3.51E-06 | 0.986859682 | 0.95 | 0.867 | 0.001749349 |
| ARL4C | 2.60E-05 | 0.77583313 | 0.5 | 0.235 | 0.012966201 |
| IGHM | 3.02E-05 | 1.891273355 | 0.95 | 0.763 | 0.015063105 |
| CTSD | 3.44E-05 | 0.735400132 | 0.5 | 0.31 | 0.017134902 |
| ATF6B | 3.67E-05 | 0.664222603 | 0.65 | 0.487 | 0.018267837 |
| FTH1 | 4.40E-05 | 0.847570618 | 1 | 0.984 | 0.021902485 |
| IGHD | 4.80E-05 | 1.666644558 | 0.85 | 0.515 | 0.023887327 |
| PTPRC | 7.34E-05 | 0.812137711 | 1 | 0.917 | 0.036556265 |
| TRIB2 | 0.000101503 | 0.991071934 | 0.4 | 0.157 | 0.050548613 |
| ITGA4 | 0.000107127 | 0.737603109 | 0.8 | 0.507 | 0.053349112 |
| TSPAN32 | 0.000170256 | 0.382234435 | 0.2 | 0.034 | 0.084787558 |
| LYN | 0.000256489 | 0.966183259 | 0.95 | 0.781 | 0.127731273 |
| ITGB2 | 0.000284665 | 1.035597353 | 0.9 | 0.606 | 0.141763174 |
| TREM1 | 0.000343656 | 0.184526605 | 0.15 | 0.012 | 0.171140674 |
| SP140 | 0.000691863 | 0.869039112 | 0.8 | 0.722 | 0.344547724 |
| CD22 | 0.000774925 | 1.069647166 | 0.8 | 0.73 | 0.385912565 |
| RNASE6 | 0.000853005 | 0.482561493 | 0.35 | 0.089 | 0.424796645 |

| | | | | | |
|---|---|---|---|---|---|
| VPREB3 | 0.000935019 | -1.172244996 | 0.25 | 0.507 | 0.465639496 |
| DOCK8 | 0.001045595 | 0.616862585 | 0.85 | 0.73 | 0.520706236 |
| GNAI2 | 0.001291689 | 0.74351108 | 1 | 0.998 | 0.643261086 |
| CD72 | 0.001419339 | 1.185471705 | 0.7 | 0.233 | 0.706830998 |
| EVI2A | 0.001424532 | 0.660417769 | 0.75 | 0.473 | 0.709416912 |
| IGHA1 | 0.001528675 | -1.406407194 | 0.1 | 0.362 | 0.761279995 |
| CYBB | 0.001662895 | 0.742998788 | 0.8 | 0.561 | 0.828121467 |
| VSIR | 0.002125119 | 0.582537617 | 0.45 | 0.221 | 1 |
| ERBIN | 0.002411604 | 0.625963052 | 0.8 | 0.581 | 1 |
| FCER1G | 0.00265052 | 0.610395161 | 0.8 | 0.507 | 1 |
| CD27 | 0.002772053 | -0.848765299 | 0.35 | 0.652 | 1 |
| EVI2B | 0.002916106 | 0.625050237 | 1 | 0.889 | 1 |
| ITGAX | 0.002963567 | 1.015963987 | 0.7 | 0.334 | 1 |
| MS4A1 | 0.003556163 | 0.754068946 | 1 | 0.988 | 1 |
| JAK1 | 0.003799238 | 0.537009277 | 1 | 0.899 | 1 |
| LAT2 | 0.003842246 | 0.645671673 | 0.75 | 0.563 | 1 |
| HLA.DMA | 0.004244347 | 0.735487446 | 1 | 0.94 | 1 |
| ARPC1B | 0.004525694 | 0.953574786 | 0.95 | 0.839 | 1 |
| CD48 | 0.004808356 | 0.5829662 | 0.95 | 0.974 | 1 |
| LIPA | 0.004959948 | 0.541800697 | 0.8 | 0.555 | 1 |
| PLCG2 | 0.005239788 | 0.713593958 | 0.9 | 0.742 | 1 |
| FAS | 0.006117926 | 0.319911156 | 0.2 | 0.074 | 1 |
| CD74 | 0.006405571 | 0.662042663 | 1 | 1 | 1 |
| PIK3AP1 | 0.006860341 | 0.650877242 | 0.55 | 0.435 | 1 |
| STAT1 | 0.006988883 | -0.972998715 | 0.65 | 0.72 | 1 |
| CD5 | 0.007159222 | 0.545192482 | 0.4 | 0.117 | 1 |
| CCL4 | 0.007903011 | -1.282791907 | 0.25 | 0.352 | 1 |
| IFNGR1 | 0.008949151 | 0.537009277 | 0.6 | 0.41 | 1 |
| LCK | 0.009426418 | -0.671008895 | 0.15 | 0.404 | 1 |
| POU2F2 | 0.009825854 | 0.549099706 | 0.7 | 0.469 | 1 |
| HLA.DQB1 | 0.010008304 | 0.784507859 | 1 | 0.974 | 1 |
| ITGAM | 0.011223435 | 0.555624956 | 0.35 | 0.151 | 1 |
| HLA.DPA1 | 0.013491879 | 0.700584853 | 1 | 0.996 | 1 |
| BCL11B | 0.01378097 | 0.367084276 | 0.2 | 0.07 | 1 |
| IL2RG | 0.013861973 | 0.557633814 | 0.9 | 0.851 | 1 |
| IKZF1 | 0.014877308 | 0.416288274 | 0.5 | 0.338 | 1 |
| CD79A | 0.015092962 | 0.498478547 | 1 | 0.994 | 1 |
| NFATC1 | 0.017641883 | 0.547807518 | 0.6 | 0.382 | 1 |
| HLA.DRA | 0.018875912 | 0.656072105 | 1 | 0.996 | 1 |
| CD63 | 0.020953142 | 0.623737556 | 0.85 | 0.805 | 1 |
| CD109 | 0.021312951 | 0.390565709 | 0.1 | 0.024 | 1 |
| TNFAIP8 | 0.028608562 | 0.520115296 | 0.9 | 0.684 | 1 |
| BTLA | 0.031511635 | 0.302494482 | 0.3 | 0.109 | 1 |
| BCL6 | 0.032005522 | 0.145381315 | 0.15 | 0.04 | 1 |
| CXCR3 | 0.037705201 | 0.954182521 | 0.35 | 0.135 | 1 |
| CD86 | 0.038376952 | 0.859317479 | 0.4 | 0.352 | 1 |
| FOXO1 | 0.038510168 | 0.342240205 | 0.25 | 0.175 | 1 |

| | | | | | |
|---|---|---|---|---|---|
| CD4 | 0.038653676 | 0.22622174 | 0.15 | 0.105 | 1 |
| PIK3IP1 | 0.038828627 | 0.377692376 | 0.5 | 0.421 | 1 |
| MAP2K3 | 0.03917561 | 0.683408385 | 0.85 | 0.686 | 1 |
| IRF8 | 0.039756537 | -0.597640249 | 0.8 | 0.879 | 1 |
| SKAP1 | 0.040233084 | 0.212309433 | 0.05 | 0.034 | 1 |
| CCR9 | 0.040487662 | 0.592662335 | 0.2 | 0.014 | 1 |
| IFI6 | 0.042604329 | -0.549859984 | 0.1 | 0.235 | 1 |
| CD7 | 0.042623328 | 0.159240163 | 0.1 | 0.028 | 1 |
| HAVCR2 | 0.043538271 | 0.365809247 | 0.3 | 0.089 | 1 |
| TRIB1 | 0.046322425 | 0.460467723 | 0.35 | 0.195 | 1 |
| NFKB2 | 0.052231156 | 0.257626878 | 0.25 | 0.107 | 1 |
| UGCG | 0.054043918 | 0.594494772 | 0.6 | 0.465 | 1 |
| FYN | 0.05511903 | 0.451922221 | 0.35 | 0.356 | 1 |
| LGALS3 | 0.0573022 | 0.652486495 | 0.35 | 0.195 | 1 |
| FYB1 | 0.059536652 | 0.323594452 | 0.3 | 0.183 | 1 |
| CNOT2 | 0.060114158 | 0.357382188 | 0.35 | 0.342 | 1 |
| LY86 | 0.061806943 | 0.444891076 | 0.7 | 0.592 | 1 |
| FCGR3A | 0.068337712 | 0.22622174 | 0.25 | 0.177 | 1 |
| CD36 | 0.069001022 | 0.235839743 | 0.4 | 0.159 | 1 |
| DUSP5 | 0.069663073 | 0.97602564 | 0.35 | 0.137 | 1 |
| CLEC10A | 0.071131688 | 0.111918114 | 0.1 | 0.016 | 1 |
| EGR3 | 0.075657199 | 0.455185055 | 0.2 | 0.032 | 1 |
| IL1B | 0.080035609 | -0.401380377 | 0.1 | 0.276 | 1 |
| CCL22 | 0.084891469 | -1.673984995 | 0.4 | 0.388 | 1 |
| JAK3 | 0.088375721 | 0.357030611 | 0.75 | 0.588 | 1 |
| CXCL10 | 0.09275599 | -1.072839184 | 0.05 | 0.08 | 1 |
| BIN2 | 0.09390311 | 0.218499893 | 0.25 | 0.095 | 1 |
| TCF4 | 0.096369761 | 0.507585517 | 0.4 | 0.235 | 1 |
| TLR7 | 0.099180405 | 0.217549438 | 0.3 | 0.135 | 1 |
| ENTPD1 | 0.102481784 | -0.449590152 | 0.2 | 0.32 | 1 |
| LGALS1 | 0.109842424 | -0.669829369 | 0.75 | 0.797 | 1 |
| ARHGEF3 | 0.112155111 | 0.276920436 | 0.45 | 0.26 | 1 |
| REL | 0.116911421 | -0.527219527 | 0.65 | 0.646 | 1 |
| S100A10 | 0.121854971 | 0.396329096 | 0.85 | 0.791 | 1 |
| CD1C | 0.126056781 | 1.082260537 | 0.5 | 0.187 | 1 |
| CD44 | 0.127152538 | 0.3305584 | 1 | 0.928 | 1 |
| POU2AF1 | 0.127869199 | 0.424309145 | 0.85 | 0.807 | 1 |
| STAT4 | 0.128871048 | -0.270345645 | 0.05 | 0.165 | 1 |
| ITK | 0.13157484 | 0.250724226 | 0.2 | 0.171 | 1 |
| BIRC3 | 0.138844106 | 0.401603647 | 0.75 | 0.666 | 1 |
| FASLG | 0.140334984 | 0.056119231 | 0.05 | 0.01 | 1 |
| MYB | 0.140334984 | 0.056119231 | 0.05 | 0.01 | 1 |
| BTG1 | 0.142051384 | 0.279547852 | 0.95 | 0.982 | 1 |
| CXCL8 | 0.148518136 | -0.407718139 | 0.45 | 0.507 | 1 |
| IRF1 | 0.150395404 | -0.489109783 | 0.65 | 0.724 | 1 |
| HLA.A | 0.150705031 | 0.315603253 | 1 | 1 | 1 |
| DUSP4 | 0.154072833 | -0.677434391 | 0.15 | 0.215 | 1 |

| | | | | | |
|---|---|---|---|---|---|
| SELPLG | 0.159425573 | 0.255881714 | 0.3 | 0.247 | 1 |
| LAT | 0.162414526 | 0.276920436 | 0.4 | 0.245 | 1 |
| TGFBR2 | 0.167333457 | 0.315878505 | 0.95 | 0.799 | 1 |
| TLR2 | 0.168006444 | 0.086778551 | 0.1 | 0.036 | 1 |
| CASP5 | 0.175358465 | 0.199415041 | 0.5 | 0.278 | 1 |
| MITF | 0.179252805 | 0.100690858 | 0.05 | 0.024 | 1 |
| MGST1 | 0.179252835 | 0.100690858 | 0.1 | 0.022 | 1 |
| CLEC4E | 0.179252835 | 0.100690858 | 0.1 | 0.022 | 1 |
| CMKLR1 | 0.184611091 | 0.053282072 | 0.05 | 0.012 | 1 |
| JAK2 | 0.184611091 | 0.053282072 | 0.05 | 0.012 | 1 |
| NFKBID | 0.184810565 | 0.137129462 | 0.1 | 0.038 | 1 |
| PFN1 | 0.187931903 | 0.437077686 | 1 | 0.998 | 1 |
| CD24 | 0.190936897 | 0.406141752 | 0.3 | 0.362 | 1 |
| PAXBP1 | 0.193479851 | -0.254404101 | 0.2 | 0.364 | 1 |
| LTB | 0.201580928 | 0.200359677 | 0.25 | 0.131 | 1 |
| CD274 | 0.202698041 | 0.279932327 | 0.2 | 0.093 | 1 |
| CST7 | 0.204966009 | -0.20981395 | 0.15 | 0.252 | 1 |
| ANXA5 | 0.211931612 | 0.361546093 | 0.7 | 0.684 | 1 |
| IRF3 | 0.215563036 | 0.144057842 | 0.25 | 0.113 | 1 |
| NFATC3 | 0.216053255 | 0.236855904 | 0.5 | 0.394 | 1 |
| TNFRSF13C | 0.216281686 | 0.259527161 | 0.85 | 0.877 | 1 |
| RGS1 | 0.226692228 | 0.500263495 | 0.2 | 0.175 | 1 |
| CD2 | 0.230860399 | -0.196816609 | 0.15 | 0.245 | 1 |
| NGDN | 0.231593492 | 0.050450481 | 0.05 | 0.014 | 1 |
| DUSP1 | 0.23590091 | -0.235038776 | 0.2 | 0.302 | 1 |
| CXCL16 | 0.236933161 | -0.21169965 | 0.15 | 0.274 | 1 |
| CXCL5 | 0.237962122 | 0.29493449 | 0.85 | 0.634 | 1 |
| SNX9 | 0.242214668 | 0.260169072 | 0.35 | 0.328 | 1 |
| GBP1 | 0.242294389 | -0.40186587 | 0.35 | 0.414 | 1 |
| PSME2 | 0.244544954 | -0.456573981 | 0.9 | 0.845 | 1 |
| S100A9 | 0.24753561 | 0.265463372 | 0.45 | 0.37 | 1 |
| FBXO11 | 0.248738855 | 0.193054876 | 0.35 | 0.334 | 1 |
| TGIF1 | 0.255420758 | 0.282322214 | 0.5 | 0.505 | 1 |
| DUSP6 | 0.257577508 | 0.078495112 | 0.1 | 0.04 | 1 |
| IGKC | 0.257742056 | 0.571550136 | 0.9 | 0.879 | 1 |
| IGBP1 | 0.259964144 | 0.231529024 | 0.55 | 0.374 | 1 |
| LRRC8B | 0.262933725 | 0.084012117 | 0.1 | 0.034 | 1 |
| GEM | 0.276294326 | 0.190325084 | 0.1 | 0.032 | 1 |
| AIM2 | 0.279752027 | 0.075744501 | 0.1 | 0.042 | 1 |
| RPL6 | 0.281271188 | 0.271203122 | 1 | 1 | 1 |
| KLRK1 | 0.282657066 | -0.193505276 | 0.2 | 0.31 | 1 |
| GZMB | 0.283273706 | -0.250391038 | 0.15 | 0.221 | 1 |
| CD3E | 0.284178097 | 0.213813436 | 0.45 | 0.334 | 1 |
| CD40 | 0.285475448 | -0.314579616 | 0.6 | 0.698 | 1 |
| LGALS9 | 0.287321223 | 0.256977268 | 0.6 | 0.417 | 1 |
| CCL3 | 0.288956935 | -0.397537661 | 0.3 | 0.322 | 1 |
| VMO1 | 0.290218405 | 0.14913776 | 0.1 | 0.103 | 1 |

| | | | | | |
|---|---|---|---|---|---|
| EIF1 | 0.290364838 | 0.305278992 | 0.95 | 0.944 | 1 |
| IL4R | 0.294859953 | 0.299740808 | 0.55 | 0.364 | 1 |
| BCL10 | 0.296846495 | 0.131339092 | 0.3 | 0.159 | 1 |
| IL2RB | 0.300212213 | -0.221982623 | 0.1 | 0.219 | 1 |
| PAX5 | 0.304912318 | 0.078495112 | 0.1 | 0.038 | 1 |
| IGHG1 | 0.316279071 | 0.353088266 | 0.85 | 0.865 | 1 |
| LAMP1 | 0.317557028 | 0.193054876 | 0.55 | 0.555 | 1 |
| WAS | 0.319116547 | 0.220863535 | 0.75 | 0.565 | 1 |
| PRF1 | 0.320392578 | -0.173299133 | 0.2 | 0.308 | 1 |
| SRGN | 0.321464487 | -0.135105172 | 0.05 | 0.107 | 1 |
| ICOS | 0.329637985 | 0.044803918 | 0.05 | 0.018 | 1 |
| ADA | 0.330102357 | -0.181415582 | 0.15 | 0.284 | 1 |
| SOCS3 | 0.333201339 | 0.091240063 | 0.1 | 0.074 | 1 |
| BCL2 | 0.336644558 | -0.214792245 | 0.3 | 0.473 | 1 |
| SLC1A5 | 0.338999836 | 0.399420258 | 0.6 | 0.37 | 1 |
| BAX | 0.340235329 | 0.243774634 | 0.7 | 0.511 | 1 |
| NR4A2 | 0.344853636 | -0.169515203 | 0.05 | 0.082 | 1 |
| IL12A | 0.350688787 | 0.067523994 | 0.1 | 0.048 | 1 |
| IER3 | 0.354140983 | 0.193054876 | 0.4 | 0.256 | 1 |
| SNCA | 0.362167881 | 0.050450481 | 0.05 | 0.012 | 1 |
| CCR4 | 0.362167881 | 0.050450481 | 0.05 | 0.012 | 1 |
| PSME3 | 0.366260653 | -0.231902102 | 0.35 | 0.306 | 1 |
| LEF1 | 0.3695228 | -0.163430441 | 0.1 | 0.237 | 1 |
| RUNX3 | 0.380882659 | -0.249333111 | 0.7 | 0.724 | 1 |
| IRF7 | 0.381319112 | 0.232225474 | 0.6 | 0.573 | 1 |
| CXCR4 | 0.382863806 | -0.262139749 | 0.85 | 0.917 | 1 |
| PRNP | 0.390450415 | -0.125447744 | 0.1 | 0.207 | 1 |
| FOS | 0.403613975 | 0.131249592 | 0.3 | 0.181 | 1 |
| FOXP1 | 0.406886996 | 0.174439198 | 0.2 | 0.147 | 1 |
| JUNB | 0.409199636 | -0.130013238 | 0.1 | 0.193 | 1 |
| THBS1 | 0.411280302 | 0.162772309 | 0.6 | 0.567 | 1 |
| GZMA | 0.414445857 | 0.067523994 | 0.05 | 0.046 | 1 |
| UFM1 | 0.416375477 | 0.18651203 | 0.55 | 0.584 | 1 |
| MYC | 0.420009818 | 0.423954936 | 0.7 | 0.33 | 1 |
| MZB1 | 0.420331872 | -0.283581472 | 0.25 | 0.298 | 1 |
| IVNS1ABP | 0.42056896 | 0.156866569 | 0.4 | 0.346 | 1 |
| CHD1 | 0.420975618 | 0.044803918 | 0.05 | 0.016 | 1 |
| GIMAP5 | 0.424748417 | 0.121705032 | 0.15 | 0.153 | 1 |
| RPN2 | 0.430060573 | 0.205765928 | 0.7 | 0.604 | 1 |
| SPAG9 | 0.436373515 | 0.13341193 | 0.35 | 0.199 | 1 |
| IRF4 | 0.437294746 | 0.310594048 | 0.65 | 0.421 | 1 |
| NT5E | 0.447720734 | -0.125447744 | 0.15 | 0.173 | 1 |
| SDCBP | 0.450737111 | 0.178430942 | 0.7 | 0.65 | 1 |
| NIBAN3 | 0.455671783 | 0.121705032 | 0.2 | 0.143 | 1 |
| PCNA | 0.464168794 | -0.258641094 | 0.25 | 0.354 | 1 |
| BACH2 | 0.464639482 | 0.119661618 | 0.2 | 0.183 | 1 |
| KDELR1 | 0.468174309 | 0.155170116 | 0.6 | 0.479 | 1 |

| | | | | | |
|---|---|---|---|---|---|
| NANS | 0.468287414 | 0.220212291 | 0.5 | 0.296 | 1 |
| PGM3 | 0.475665854 | 0.095341938 | 0.15 | 0.095 | 1 |
| IGLC3 | 0.484312949 | 0.480473794 | 0.7 | 0.73 | 1 |
| LILRB4 | 0.486241325 | -0.143110456 | 0.1 | 0.097 | 1 |
| MCM2 | 0.490342506 | 0.284202764 | 0.5 | 0.332 | 1 |
| CCL13 | 0.494545834 | 0.047624437 | 0.05 | 0.012 | 1 |
| CHD7 | 0.495613828 | -0.0767296 | 0.05 | 0.083 | 1 |
| CCL2 | 0.502568856 | 0.171359805 | 0.9 | 0.801 | 1 |
| FURIN | 0.504241601 | 0.127183205 | 0.5 | 0.421 | 1 |
| CD3D | 0.5148451 | 0.128924539 | 0.3 | 0.344 | 1 |
| IFITM2 | 0.519455962 | 0.17289868 | 0.95 | 0.944 | 1 |
| BCL2L1 | 0.526023208 | -0.081900555 | 0.05 | 0.07 | 1 |
| IFNG | 0.527498608 | 0.036375296 | 0.05 | 0.022 | 1 |
| SNX8 | 0.529310853 | 0.225065271 | 0.55 | 0.475 | 1 |
| NKG7 | 0.529407202 | -0.127968424 | 0.3 | 0.37 | 1 |
| IL11RA | 0.531596145 | -0.093078228 | 0.15 | 0.171 | 1 |
| EIF5 | 0.532970163 | 0.144368344 | 0.6 | 0.584 | 1 |
| TRDC | 0.536064219 | -0.0767296 | 0.05 | 0.072 | 1 |
| JCHAIN | 0.538578995 | 0.435740637 | 0.2 | 0.235 | 1 |
| GAPDH | 0.545781745 | 0.249895514 | 1 | 0.982 | 1 |
| NINJ2 | 0.565423685 | 0.033576662 | 0.05 | 0.024 | 1 |
| IFITM3 | 0.566749527 | 0.096761215 | 0.25 | 0.193 | 1 |
| TGFBR1 | 0.569982489 | 0.086976357 | 0.35 | 0.254 | 1 |
| CBLB | 0.571451497 | 0.030783447 | 0.05 | 0.028 | 1 |
| KCNE3 | 0.571451497 | 0.030783447 | 0.05 | 0.028 | 1 |
| TIAF1 | 0.577586087 | 0.093519203 | 0.35 | 0.225 | 1 |
| IL7R | 0.59440351 | 0.062306285 | 0.15 | 0.082 | 1 |
| BATF | 0.59979875 | 0.119661618 | 0.25 | 0.133 | 1 |
| IL23A | 0.601756921 | 0.043139836 | 0.1 | 0.064 | 1 |
| ICAM1 | 0.602981444 | -0.154868427 | 0.25 | 0.201 | 1 |
| LAP3 | 0.611100674 | -0.16385721 | 0.6 | 0.437 | 1 |
| TGFBI | 0.612344008 | 0.081161996 | 0.45 | 0.356 | 1 |
| APOBEC3G | 0.616163154 | 0.140294731 | 0.75 | 0.73 | 1 |
| FCER2 | 0.616685807 | -0.152885341 | 0.25 | 0.167 | 1 |
| NRP1 | 0.616808512 | 0.02799563 | 0.05 | 0.03 | 1 |
| IL12RB2 | 0.623176474 | 0.115352519 | 0.1 | 0.024 | 1 |
| TNFSF10 | 0.624932495 | 0.091334955 | 0.35 | 0.211 | 1 |
| JUN | 0.626303809 | 0.065021487 | 0.1 | 0.119 | 1 |
| CXCL3 | 0.626653583 | 0.097897643 | 0.2 | 0.131 | 1 |
| CCL5 | 0.633210226 | -0.07543396 | 0.2 | 0.199 | 1 |
| NAMPT | 0.636007964 | -0.160744993 | 0.3 | 0.35 | 1 |
| EIF5A | 0.644482564 | -0.207036253 | 1 | 0.755 | 1 |
| ORAI1 | 0.654615427 | 0.111113263 | 0.7 | 0.459 | 1 |
| PTPN7 | 0.655091347 | 0.089961383 | 0.25 | 0.167 | 1 |
| RBCK1 | 0.661995187 | 0.081737853 | 0.45 | 0.398 | 1 |
| MMP9 | 0.662027306 | -0.052988813 | 0.1 | 0.115 | 1 |
| CXCL2 | 0.662118537 | -0.074249967 | 0.15 | 0.135 | 1 |

| | | | | | |
|---|---|---|---|---|---|
| IL15RA | 0.665009502 | -0.079963618 | 0.15 | 0.169 | 1 |
| STAT3 | 0.667475839 | 0.118183612 | 0.65 | 0.664 | 1 |
| PDIA4 | 0.669265211 | 0.152640608 | 0.6 | 0.487 | 1 |
| TRAC | 0.67010731 | 0.111113263 | 0.6 | 0.654 | 1 |
| RORA | 0.670804117 | -0.055500032 | 0.05 | 0.109 | 1 |
| CD247 | 0.671953636 | 0.043139836 | 0.05 | 0.056 | 1 |
| PRDM1 | 0.675149148 | 0.067523994 | 0.05 | 0.062 | 1 |
| FCN1 | 0.676428799 | 0.046765434 | 0.15 | 0.097 | 1 |
| CX3CR1 | 0.680715102 | 0.025213189 | 0.05 | 0.03 | 1 |
| PASK | 0.681226008 | 0.084319237 | 0.45 | 0.431 | 1 |
| CREM | 0.681476108 | 0.097897643 | 0.2 | 0.229 | 1 |
| IGHG3 | 0.682609579 | 0.147420882 | 0.8 | 0.789 | 1 |
| IL12RB1 | 0.683397853 | 0.051927139 | 0.1 | 0.085 | 1 |
| CSF1 | 0.686024569 | -0.061104591 | 0.05 | 0.044 | 1 |
| GBP2 | 0.686384966 | -0.114042414 | 0.25 | 0.324 | 1 |
| STAT6 | 0.693097069 | 0.083742505 | 0.5 | 0.525 | 1 |
| SOCS2 | 0.701326365 | -0.058483891 | 0.05 | 0.042 | 1 |
| LAMP3 | 0.709790542 | -0.047953223 | 0.05 | 0.048 | 1 |
| ZNF341 | 0.712135382 | -0.032011679 | 0.05 | 0.068 | 1 |
| TNFRSF17 | 0.714449952 | 0.067523994 | 0.15 | 0.085 | 1 |
| DUSP2 | 0.715396131 | -0.180403519 | 0.4 | 0.378 | 1 |
| BLK | 0.72351062 | -0.029337545 | 0.05 | 0.07 | 1 |
| NFKBIZ | 0.728456261 | 0.101312308 | 0.5 | 0.465 | 1 |
| FN1 | 0.731065413 | 0.059531203 | 0.25 | 0.326 | 1 |
| ABCE1 | 0.732989862 | 0.105224703 | 0.5 | 0.394 | 1 |
| LTA | 0.734146935 | 0.082023564 | 0.1 | 0.089 | 1 |
| CXCL1 | 0.734766738 | 0.092102017 | 0.4 | 0.376 | 1 |
| CCND2 | 0.737042138 | -0.159278094 | 0.4 | 0.227 | 1 |
| IL6ST | 0.747968734 | 0.060860264 | 0.25 | 0.195 | 1 |
| TNF | 0.74889011 | -0.119474521 | 0.35 | 0.157 | 1 |
| BLNK | 0.748972496 | -0.084069184 | 0.55 | 0.571 | 1 |
| TBX21 | 0.753690447 | -0.129622476 | 0.4 | 0.264 | 1 |
| VSIG4 | 0.759102344 | 0.022436105 | 0.05 | 0.028 | 1 |
| TIGIT | 0.759399681 | 0.035102517 | 0.1 | 0.054 | 1 |
| SLC7A7 | 0.759894257 | -0.052057622 | 0.3 | 0.288 | 1 |
| TGFB1 | 0.759907704 | 0.022436105 | 0.05 | 0.028 | 1 |
| IFRD1 | 0.764546565 | 0.035325172 | 0.2 | 0.155 | 1 |
| CD83 | 0.767809613 | -0.152154042 | 0.55 | 0.6 | 1 |
| CD69 | 0.769199459 | -0.074555606 | 0.25 | 0.318 | 1 |
| PSEN1 | 0.769996032 | 0.035325172 | 0.2 | 0.149 | 1 |
| IRF9 | 0.770124874 | 0.069752095 | 0.9 | 0.807 | 1 |
| STIM1 | 0.770545491 | -0.035839844 | 0.15 | 0.159 | 1 |
| DCLRE1C | 0.771225025 | -0.043507318 | 0.2 | 0.229 | 1 |
| BCAT1 | 0.773222736 | 0.094725722 | 0.1 | 0.145 | 1 |
| MALT1 | 0.773324888 | -0.041000463 | 0.2 | 0.189 | 1 |
| NFKBIA | 0.773910368 | 0.084579158 | 0.8 | 0.753 | 1 |
| HIF1A | 0.774808933 | -0.057171753 | 0.65 | 0.636 | 1 |

| | | | | | |
|---|---|---|---|---|---|
| ATF4 | 0.776187996 | 0.084477127 | 0.9 | 0.853 | 1 |
| IL32 | 0.781094613 | -0.041000463 | 0.15 | 0.189 | 1 |
| SMAD2 | 0.781230907 | 0.05158245 | 0.25 | 0.306 | 1 |
| TCL1A | 0.782893638 | 0.123233427 | 0.15 | 0.151 | 1 |
| TUBA1C | 0.7847708 | -0.131896681 | 0.65 | 0.634 | 1 |
| EEF1E1 | 0.788725321 | -0.050263384 | 0.15 | 0.147 | 1 |
| CD80 | 0.791696524 | -0.047953223 | 0.15 | 0.151 | 1 |
| MCM4 | 0.792140519 | 0.085445902 | 0.1 | 0.105 | 1 |
| LRRC32 | 0.793408076 | -0.042658924 | 0.05 | 0.032 | 1 |
| EIF3J | 0.79718824 | 0.036497099 | 0.1 | 0.085 | 1 |
| NFKB1 | 0.799684773 | -0.108460214 | 0.65 | 0.646 | 1 |
| CXCR5 | 0.80094179 | -0.074753283 | 0.7 | 0.799 | 1 |
| VPS28 | 0.807101806 | 0.054113075 | 0.7 | 0.736 | 1 |
| CR2 | 0.808063879 | 0.048988043 | 0.25 | 0.252 | 1 |
| CSRNP1 | 0.811724178 | 0.043677252 | 0.3 | 0.316 | 1 |
| TYMS | 0.812975159 | 0.121105034 | 0.05 | 0.018 | 1 |
| CD300A | 0.819978332 | 0.031390208 | 0.1 | 0.091 | 1 |
| CCR1 | 0.824123002 | -0.027633239 | 0.1 | 0.091 | 1 |
| HNRNPC | 0.825520843 | -0.077906445 | 0.8 | 0.696 | 1 |
| ZNF683 | 0.827636922 | -0.02397436 | 0.05 | 0.044 | 1 |
| TRBC2 | 0.828841635 | 0.052904205 | 0.95 | 0.847 | 1 |
| PSMB8 | 0.83160015 | -0.061285153 | 0.95 | 0.897 | 1 |
| STAT5A | 0.840837883 | 0.056492957 | 0.55 | 0.376 | 1 |
| IL15 | 0.840916792 | -0.018591164 | 0.05 | 0.05 | 1 |
| CCR6 | 0.853650437 | -0.03642697 | 0.4 | 0.316 | 1 |
| SELL | 0.854522945 | -0.052899046 | 0.65 | 0.63 | 1 |
| IGHG2 | 0.861516516 | 0.062306285 | 0.75 | 0.783 | 1 |
| XBP1 | 0.865374926 | 0.049951365 | 0.3 | 0.288 | 1 |
| PAK1IP1 | 0.866133451 | -0.037242743 | 0.25 | 0.181 | 1 |
| IGHG4 | 0.866663936 | 0.050227171 | 0.35 | 0.368 | 1 |
| IL1A | 0.866872779 | 0.014136781 | 0.05 | 0.03 | 1 |
| GAB2 | 0.877035244 | -0.02558541 | 0.2 | 0.195 | 1 |
| PCLAF | 0.879319683 | 0.054514934 | 0.05 | 0.026 | 1 |
| CYCS | 0.879948172 | -0.062922956 | 0.65 | 0.499 | 1 |
| BCL2A1 | 0.883691839 | -0.079317394 | 0.5 | 0.404 | 1 |
| SLC7A1 | 0.885957351 | 0.032334566 | 0.2 | 0.177 | 1 |
| ILF2 | 0.894647756 | 0.046864524 | 0.7 | 0.674 | 1 |
| PDIA6 | 0.89502278 | -0.053166219 | 0.85 | 0.664 | 1 |
| GZMH | 0.895818515 | -0.010478518 | 0.05 | 0.052 | 1 |
| GNLY | 0.896181358 | 0.025213189 | 0.5 | 0.461 | 1 |
| ZBED2 | 0.897678179 | 0.024455272 | 0.05 | 0.036 | 1 |
| CD6 | 0.90564804 | -0.021069929 | 0.1 | 0.183 | 1 |
| VNN2 | 0.908880519 | 0.017535858 | 0.2 | 0.225 | 1 |
| PSMB9 | 0.911641591 | -0.029111789 | 0.85 | 0.843 | 1 |
| ARID5A | 0.915793909 | 0.015861874 | 0.15 | 0.139 | 1 |
| CCR7 | 0.926144709 | -0.031146167 | 0.9 | 0.895 | 1 |
| ITGAE | 0.926592229 | -0.012849422 | 0.2 | 0.169 | 1 |

| | | | | | |
|---|---|---|---|---|---|
| CD8A | 0.928668538 | -0.011303622 | 0.15 | 0.133 | 1 |
| F5 | 0.928818389 | 0.011380916 | 0.05 | 0.02 | 1 |
| CTSW | 0.930289993 | 0.005884928 | 0.05 | 0.046 | 1 |
| IL3RA | 0.932584231 | -0.030778079 | 0.3 | 0.097 | 1 |
| CD14 | 0.945228126 | -0.013925819 | 0.35 | 0.421 | 1 |
| CD9 | 0.946040307 | 0.008630305 | 0.15 | 0.113 | 1 |
| KLRC3 | 0.946746193 | -0.007022959 | 0.1 | 0.091 | 1 |
| LDHA | 0.94697129 | 0.029898922 | 0.95 | 0.843 | 1 |
| IL1RN | 0.949807706 | -0.007022959 | 0.1 | 0.087 | 1 |
| TIMM9 | 0.951042434 | 0.015861874 | 0.45 | 0.459 | 1 |
| GIMAP2 | 0.971650551 | 0.006641752 | 0.35 | 0.302 | 1 |
| CD163 | 0.971979764 | 0.003393657 | 0.1 | 0.089 | 1 |
| LIG4 | 0.975362087 | 0.003620942 | 0.15 | 0.127 | 1 |
| EBF1 | 0.980822435 | 0.004361974 | 0.25 | 0.237 | 1 |
| SLA | 0.989105192 | 0.00305646 | 0.35 | 0.314 | 1 |
| NFATC2 | 0.990698145 | -0.002016939 | 0.15 | 0.254 | 1 |
| CD70 | 0.992912553 | -0.642134254 | 0 | 0.235 | 1 |
| TRAF1 | 0.993180327 | -0.352014897 | 0 | 0.141 | 1 |
| TMEM97 | 0.993340671 | -0.239904531 | 0 | 0.111 | 1 |
| SLC25A37 | 0.993432374 | -0.00163803 | 0.3 | 0.252 | 1 |
| MAST4 | 0.9934739 | -0.172790335 | 0 | 0.103 | 1 |
| TNFSF13B | 0.993492072 | -0.165136763 | 0 | 0.099 | 1 |
| IL6 | 0.993574988 | -0.134109867 | 0 | 0.056 | 1 |
| CHI3L2 | 0.993651804 | -0.110393798 | 0 | 0.062 | 1 |
| CD38 | 0.993661388 | -0.107734452 | 0 | 0.06 | 1 |
| AHI1 | 0.993681314 | -0.102401007 | 0 | 0.072 | 1 |
| CD200 | 0.993691685 | -0.099726873 | 0 | 0.054 | 1 |
| IL6R | 0.993736268 | -0.088980491 | 0 | 0.06 | 1 |
| GZMK | 0.993773529 | -0.080867846 | 0 | 0.054 | 1 |
| IL18R1 | 0.99377353 | -0.080867846 | 0 | 0.04 | 1 |
| IL2RA | 0.993786812 | -0.078153461 | 0 | 0.038 | 1 |
| TCF7 | 0.993814863 | -0.072709322 | 0 | 0.046 | 1 |
| FUT4 | 0.993829709 | -0.06997953 | 0 | 0.044 | 1 |
| KLRC1 | 0.99386127 | -0.064504399 | 0 | 0.038 | 1 |
| YBX3 | 0.99386127 | -0.064504399 | 0 | 0.036 | 1 |
| TOP2A | 0.993914173 | -0.056252546 | 0 | 0.028 | 1 |
| AQP9 | 0.993933588 | -0.053491407 | 0 | 0.038 | 1 |
| TNIK | 0.993933588 | -0.053491407 | 0 | 0.034 | 1 |
| CD226 | 0.993933588 | -0.053491407 | 0 | 0.032 | 1 |
| EGR1 | 0.993933589 | -0.053491407 | 0 | 0.026 | 1 |
| LAG3 | 0.993954054 | -0.050724972 | 0 | 0.03 | 1 |
| CARD11 | 0.993975689 | -0.047953223 | 0 | 0.03 | 1 |
| S100A12 | 0.99397569 | -0.047953223 | 0 | 0.024 | 1 |
| CCR5 | 0.993998636 | -0.045176139 | 0 | 0.03 | 1 |
| TNFRSF8 | 0.993998638 | -0.045176139 | 0 | 0.018 | 1 |
| KLRF1 | 0.994023066 | -0.042393698 | 0 | 0.024 | 1 |
| CTLA4 | 0.994023066 | -0.042393698 | 0 | 0.022 | 1 |

| | | | | | |
|---|---|---|---|---|---|
| FOSL1 | 0.994049181 | -0.039605881 | 0 | 0.02 | 1 |
| B4GALT5 | 0.994077232 | -0.036812666 | 0 | 0.026 | 1 |
| KLRG1 | 0.994077232 | -0.036812666 | 0 | 0.024 | 1 |
| CD33 | 0.994107529 | -0.034014032 | 0 | 0.024 | 1 |
| PDCD1 | 0.994140466 | -0.031209959 | 0 | 0.016 | 1 |
| TNFRSF25 | 0.994176541 | -0.028400426 | 0 | 0.02 | 1 |
| ZAP70 | 0.994216424 | -0.02558541 | 0 | 0.014 | 1 |
| CXCL9 | 0.995705099 | -0.266376742 | 0 | 0.036 | 1 |
| IL2 | 0.9958469 | -0.149706722 | 0 | 0.024 | 1 |
| CD28 | 0.996262476 | -0.02558541 | 0 | 0.018 | 1 |
| TRAT1 | 0.996262476 | -0.02558541 | 0 | 0.016 | 1 |
| CXCR6 | 0.996289518 | -0.022764891 | 0 | 0.01 | 1 |
| QPCT | 0.996320175 | -0.019938847 | 0 | 0.014 | 1 |
| IL18 | 0.996320175 | -0.019938847 | 0 | 0.012 | 1 |
| SPP1 | 0.996320175 | -0.019938847 | 0 | 0.012 | 1 |
| FOSB | 0.996320175 | -0.019938847 | 0 | 0.01 | 1 |
| IL18RAP | 0.996355566 | -0.017107256 | 0 | 0.012 | 1 |
| CD209 | 0.996355566 | -0.017107256 | 0 | 0.01 | 1 |
| NFATC4 | 0.996355566 | -0.017107256 | 0 | 0.01 | 1 |
| CCL20 | 0.996355567 | -0.017107256 | 0 | 0.008 | 1 |
| ZBTB16 | 0.996397425 | -0.014270097 | 0 | 0.01 | 1 |
| THEMIS | 0.996397425 | -0.014270097 | 0 | 0.01 | 1 |
| SOCS1 | 0.996397425 | -0.014270097 | 0 | 0.01 | 1 |
| GRB10 | 0.996397425 | -0.014270097 | 0 | 0.01 | 1 |
| OSM | 0.996397425 | -0.014270097 | 0 | 0.01 | 1 |
| CD8B | 0.996448655 | -0.011427347 | 0 | 0.008 | 1 |
| IL1R1 | 0.996448655 | -0.011427347 | 0 | 0.008 | 1 |
| B3GAT1 | 0.996448656 | -0.011427347 | 0 | 0.006 | 1 |
| TGFB3 | 0.996448656 | -0.011427347 | 0 | 0.006 | 1 |
| TNFSF14 | 0.996514704 | -0.008578985 | 0 | 0.006 | 1 |
| RAPH1 | 0.996514704 | -0.008578985 | 0 | 0.006 | 1 |
| CSF2 | 0.997363647 | -0.128873219 | 0 | 0.014 | 1 |
| AURKB | 0.997628816 | -0.019938847 | 0 | 0.006 | 1 |
| CXCL11 | 0.998334461 | -0.105070194 | 0 | 0.008 | 1 |

Monocytes

| | p_val | avg_log2FC | Patient | Controls | p_val_adj |
|---|---|---|---|---|---|
| FCGR3A | 1.60E-51 | 1.645438528 | 0.954 | 0.851 | 7.99E-49 |
| CXCL5 | 1.28E-47 | -3.006364285 | 0.861 | 0.85 | 6.38E-45 |
| IL1B | 1.65E-47 | -3.890495786 | 0.384 | 0.443 | 8.20E-45 |
| CD44 | 2.69E-46 | -1.056559739 | 0.966 | 0.979 | 1.34E-43 |
| FN1 | 1.55E-45 | -3.220263278 | 0.439 | 0.651 | 7.73E-43 |
| CCL3 | 2.55E-35 | -2.433612776 | 0.608 | 0.78 | 1.27E-32 |
| CXCL1 | 2.99E-31 | -3.036943303 | 0.511 | 0.578 | 1.49E-28 |
| S100A10 | 3.16E-30 | -0.917655545 | 0.962 | 0.978 | 1.57E-27 |
| CXCL8 | 1.10E-29 | -2.202139726 | 0.781 | 0.849 | 5.49E-27 |

| | | | | | |
|---|---|---|---|---|---|
| CLEC4E | 1.66E-29 | -1.178952789 | 0.165 | 0.49 | 8.24E-27 |
| S100A9 | 2.75E-24 | -1.125035356 | 0.932 | 0.951 | 1.37E-21 |
| CCL4 | 3.57E-24 | -2.222614217 | 0.46 | 0.617 | 1.78E-21 |
| DUSP1 | 3.99E-23 | -1.066137142 | 0.814 | 0.875 | 1.99E-20 |
| IL1RN | 2.41E-22 | -2.327538552 | 0.194 | 0.345 | 1.20E-19 |
| AQP9 | 3.84E-22 | -1.118329594 | 0.363 | 0.665 | 1.91E-19 |
| CXCR4 | 4.91E-22 | -0.961453916 | 0.831 | 0.91 | 2.45E-19 |
| MMP9 | 5.10E-22 | -1.371136483 | 0.667 | 0.811 | 2.54E-19 |
| CCL22 | 1.27E-18 | -2.337368759 | 0.203 | 0.216 | 6.31E-16 |
| ICAM1 | 4.21E-18 | -0.846366876 | 0.751 | 0.86 | 2.10E-15 |
| CXCL3 | 5.49E-17 | -1.912402175 | 0.498 | 0.566 | 2.73E-14 |
| ITGAM | 7.35E-17 | -0.801199309 | 0.308 | 0.56 | 3.66E-14 |
| IER3 | 1.54E-14 | -0.979308539 | 0.831 | 0.879 | 7.65E-12 |
| CXCL2 | 2.09E-14 | -1.404462907 | 0.578 | 0.62 | 1.04E-11 |
| LIPA | 1.79E-13 | 0.932648563 | 0.983 | 0.89 | 8.92E-11 |
| MGST1 | 4.43E-13 | -0.745143253 | 0.321 | 0.555 | 2.20E-10 |
| BCL2A1 | 6.72E-13 | -1.0641407 | 0.232 | 0.424 | 3.35E-10 |
| IL6 | 6.96E-13 | -1.604324691 | 0.03 | 0.099 | 3.47E-10 |
| S100A12 | 1.24E-12 | -0.891974776 | 0.27 | 0.456 | 6.19E-10 |
| TGFBR2 | 1.73E-12 | 0.508626054 | 0.895 | 0.751 | 8.63E-10 |
| SNX8 | 3.80E-12 | 0.508599829 | 0.916 | 0.784 | 1.89E-09 |
| NFKBIA | 3.96E-12 | -0.639759967 | 0.987 | 0.966 | 1.97E-09 |
| CD9 | 1.32E-11 | -0.867396805 | 0.696 | 0.81 | 6.56E-09 |
| VMO1 | 1.91E-11 | 1.429288479 | 0.84 | 0.483 | 9.49E-09 |
| TREM1 | 3.79E-11 | -0.47438606 | 0.072 | 0.227 | 1.89E-08 |
| CXCL10 | 5.85E-11 | -2.18567499 | 0.093 | 0.167 | 2.91E-08 |
| WAS | 5.93E-11 | 0.371148773 | 0.916 | 0.846 | 2.95E-08 |
| SOCS3 | 1.18E-10 | -0.493765436 | 0.097 | 0.234 | 5.86E-08 |
| ARL4C | 1.30E-10 | 0.734150249 | 0.726 | 0.599 | 6.49E-08 |
| BTG1 | 5.50E-10 | -0.587964267 | 0.84 | 0.873 | 2.74E-07 |
| CD274 | 7.22E-10 | -0.71443231 | 0.186 | 0.286 | 3.59E-07 |
| STAT6 | 9.96E-10 | 0.342382209 | 0.688 | 0.538 | 4.96E-07 |
| IL1A | 1.72E-09 | -0.823381872 | 0.017 | 0.119 | 8.56E-07 |
| CMKLR1 | 2.00E-09 | 0.454823283 | 0.713 | 0.545 | 9.97E-07 |
| ITGA4 | 2.12E-09 | 0.436062355 | 0.565 | 0.438 | 1.06E-06 |
| PCNA | 3.04E-09 | 0.405132871 | 0.688 | 0.538 | 1.51E-06 |
| CD36 | 3.26E-09 | 0.698586689 | 0.928 | 0.817 | 1.63E-06 |
| ARPC1B | 5.63E-09 | 0.362723363 | 0.996 | 0.971 | 2.80E-06 |
| VNN2 | 6.60E-09 | -0.42574369 | 0.063 | 0.177 | 3.29E-06 |
| IL3RA | 9.51E-09 | -0.686084075 | 0.527 | 0.637 | 4.74E-06 |
| IVNS1ABP | 1.44E-08 | 0.377590466 | 0.831 | 0.676 | 7.17E-06 |
| APOE | 1.72E-08 | -0.332917341 | 0.03 | 0.17 | 8.59E-06 |
| GBP1 | 2.66E-08 | -0.961498738 | 0.616 | 0.571 | 1.32E-05 |
| IGKC | 2.70E-08 | -0.654854272 | 0.722 | 0.707 | 1.34E-05 |
| NAMPT | 2.95E-08 | -0.724247442 | 0.785 | 0.677 | 1.47E-05 |
| IL2RG | 3.50E-08 | -0.536806855 | 0.608 | 0.719 | 1.74E-05 |
| CX3CR1 | 9.07E-08 | 0.407259549 | 0.131 | 0.057 | 4.52E-05 |

| | | | | | |
|---|---|---|---|---|---|
| PFN1 | 1.15E-07 | 0.307712725 | 1 | 1 | 5.70E-05 |
| QPCT | 1.82E-07 | -0.343306358 | 0.101 | 0.241 | 9.05E-05 |
| GNAI2 | 2.03E-07 | 0.303181112 | 1 | 0.993 | 0.000101329 |
| NFKB1 | 2.12E-07 | -0.419271475 | 0.848 | 0.86 | 0.000105802 |
| THBS1 | 3.26E-07 | 0.973541479 | 0.97 | 0.848 | 0.000162271 |
| LILRB4 | 4.32E-07 | -0.420400414 | 0.949 | 0.916 | 0.000215036 |
| BIRC3 | 4.82E-07 | -0.598383947 | 0.409 | 0.51 | 0.000240089 |
| RAPH1 | 5.11E-07 | 0.284810861 | 0.451 | 0.291 | 0.000254419 |
| FOS | 5.76E-07 | -0.699559367 | 0.861 | 0.817 | 0.00028704 |
| SLC7A7 | 6.51E-07 | -0.375098275 | 0.92 | 0.885 | 0.000324206 |
| CSRNP1 | 9.12E-07 | -0.338291338 | 0.207 | 0.322 | 0.000454162 |
| IFNGR1 | 1.74E-06 | 0.32610089 | 0.97 | 0.909 | 0.00086806 |
| TRAF1 | 1.88E-06 | -0.328204645 | 0.046 | 0.138 | 0.000938692 |
| IL15RA | 2.69E-06 | -0.420490283 | 0.207 | 0.259 | 0.001339314 |
| CSF1 | 3.77E-06 | -0.482846216 | 0.055 | 0.134 | 0.00187615 |
| SRGN | 4.49E-06 | 0.301101106 | 0.696 | 0.599 | 0.002236289 |
| CD33 | 5.58E-06 | 0.31190873 | 0.7 | 0.649 | 0.002780839 |
| LAT | 5.98E-06 | -0.426208738 | 0.418 | 0.519 | 0.002977725 |
| SLA | 6.57E-06 | 0.354614114 | 0.865 | 0.734 | 0.003272557 |
| GIMAP2 | 7.34E-06 | 0.303920641 | 0.662 | 0.532 | 0.003656836 |
| EIF5A | 1.08E-05 | 0.289970934 | 0.966 | 0.912 | 0.005384028 |
| CD83 | 1.48E-05 | -0.612211485 | 0.527 | 0.628 | 0.007390735 |
| FCN1 | 1.51E-05 | -0.752110274 | 0.519 | 0.643 | 0.007502124 |
| TGFBI | 1.87E-05 | 0.474678762 | 0.992 | 0.944 | 0.009298107 |
| CCR7 | 2.00E-05 | -0.732895604 | 0.481 | 0.455 | 0.009952269 |
| NFATC3 | 3.27E-05 | 0.247079942 | 0.633 | 0.476 | 0.016301165 |
| TGFBR1 | 4.80E-05 | 0.282737182 | 0.629 | 0.49 | 0.023883073 |
| CLEC4D | 5.20E-05 | -0.167569119 | 0.013 | 0.105 | 0.025877699 |
| CD300A | 5.62E-05 | 0.35947443 | 0.633 | 0.524 | 0.028001335 |
| CD48 | 6.25E-05 | -0.314997536 | 0.975 | 0.956 | 0.031121443 |
| SLC25A37 | 6.33E-05 | -0.373514471 | 0.287 | 0.337 | 0.031542383 |
| SNX9 | 7.43E-05 | 0.271906757 | 0.755 | 0.637 | 0.037003225 |
| JUN | 7.70E-05 | -0.364172512 | 0.414 | 0.504 | 0.038331091 |
| LGALS9 | 7.75E-05 | 0.314166017 | 0.949 | 0.873 | 0.038577264 |
| TNFSF10 | 8.00E-05 | -0.526372023 | 0.219 | 0.256 | 0.039831584 |
| ORAI1 | 0.000118324 | 0.247281296 | 0.785 | 0.675 | 0.058925485 |
| FOSB | 0.00012791 | -0.235928506 | 0.021 | 0.082 | 0.063699201 |
| BIN2 | 0.000128034 | 0.211062946 | 0.544 | 0.409 | 0.063760956 |
| TLR2 | 0.000128817 | -0.374858574 | 0.553 | 0.588 | 0.064150761 |
| CD38 | 0.000150075 | -0.183689736 | 0.038 | 0.119 | 0.074737234 |
| FBXO11 | 0.00015096 | 0.222073944 | 0.667 | 0.545 | 0.075177912 |
| THBD | 0.000154195 | 0.213320655 | 0.127 | 0.043 | 0.076789327 |
| LAMP3 | 0.000190536 | -1.056543441 | 0.059 | 0.109 | 0.094886711 |
| FTH1 | 0.000203882 | -0.295634026 | 1 | 0.999 | 0.101533304 |
| DUSP4 | 0.000265568 | -0.263353501 | 0.042 | 0.096 | 0.132252836 |
| NFKB2 | 0.00028711 | -0.152457965 | 0.072 | 0.154 | 0.142981027 |
| OSM | 0.000309761 | -0.209504384 | 0.051 | 0.101 | 0.154261202 |

| | | | | | |
|---|---|---|---|---|---|
| SLC1A5 | 0.000354607 | 0.292233283 | 0.848 | 0.758 | 0.176594323 |
| IGLC3 | 0.000416768 | 0.373563581 | 0.684 | 0.611 | 0.207550301 |
| IL7R | 0.000431817 | -0.250394941 | 0.156 | 0.221 | 0.215044697 |
| DUSP2 | 0.000434403 | -0.324325418 | 0.354 | 0.412 | 0.216332915 |
| JUNB | 0.00044706 | -0.258113184 | 0.447 | 0.553 | 0.222636026 |
| GNLY | 0.000599677 | -0.322703344 | 0.544 | 0.524 | 0.298639335 |
| DUSP5 | 0.000600263 | -0.464661261 | 0.253 | 0.238 | 0.298930859 |
| PAK1IP1 | 0.00060266 | 0.174035618 | 0.405 | 0.282 | 0.300124574 |
| RNASE6 | 0.000713638 | 0.293398853 | 0.671 | 0.511 | 0.355391705 |
| LY86 | 0.000827877 | 0.303745278 | 0.814 | 0.725 | 0.412282767 |
| SPP1 | 0.000882004 | -0.564536249 | 0.034 | 0.046 | 0.439237932 |
| VSIG4 | 0.000996185 | 0.558773983 | 0.451 | 0.339 | 0.496100313 |
| KCNE3 | 0.001196056 | 0.214178014 | 0.781 | 0.659 | 0.595636032 |
| CCL20 | 0.001364683 | -0.254210988 | 0.017 | 0.036 | 0.67961227 |
| ABCE1 | 0.001627086 | 0.204359821 | 0.624 | 0.554 | 0.810288694 |
| EVI2B | 0.001668949 | 0.212676336 | 0.966 | 0.926 | 0.831136442 |
| TRIB1 | 0.001762153 | 0.275643029 | 0.743 | 0.604 | 0.877552332 |
| LGALS1 | 0.001772388 | -0.262255817 | 0.996 | 0.99 | 0.882649249 |
| TNIK | 0.001912139 | -0.199659897 | 0.249 | 0.314 | 0.952245415 |
| HNRNPC | 0.001942194 | 0.195013361 | 0.924 | 0.842 | 0.967212616 |
| CD209 | 0.001959825 | 0.248565042 | 0.397 | 0.228 | 0.975992775 |
| RGS1 | 0.002006895 | -0.282929805 | 0.076 | 0.152 | 0.999433721 |
| CD226 | 0.002233499 | -0.111070981 | 0.025 | 0.077 | 1 |
| GAB2 | 0.002685013 | 0.196104117 | 0.688 | 0.57 | 1 |
| ATF6B | 0.002702774 | 0.174540047 | 0.827 | 0.73 | 1 |
| EIF1 | 0.002870444 | 0.182523576 | 0.996 | 0.973 | 1 |
| ATF4 | 0.003006854 | 0.194205307 | 0.962 | 0.921 | 1 |
| KDELR1 | 0.003245441 | 0.174394785 | 0.907 | 0.875 | 1 |
| SDCBP | 0.003398998 | 0.234875965 | 0.992 | 0.977 | 1 |
| ITGAX | 0.003781392 | 0.221111524 | 0.932 | 0.917 | 1 |
| BCL6 | 0.003851924 | -0.109506707 | 0.059 | 0.123 | 1 |
| DUSP6 | 0.004124731 | 0.345101417 | 0.709 | 0.596 | 1 |
| LYN | 0.004175295 | -0.259801358 | 0.962 | 0.923 | 1 |
| IL15 | 0.00429751 | -0.124415387 | 0.063 | 0.101 | 1 |
| TIMM9 | 0.004448344 | 0.167013309 | 0.633 | 0.518 | 1 |
| FURIN | 0.004617345 | 0.19292756 | 0.865 | 0.762 | 1 |
| CD109 | 0.004753429 | 0.276799549 | 0.401 | 0.318 | 1 |
| JCHAIN | 0.004863235 | -0.106285919 | 0.025 | 0.067 | 1 |
| CD163 | 0.005285782 | 0.327119786 | 0.857 | 0.775 | 1 |
| VPS28 | 0.005650631 | 0.157041621 | 0.97 | 0.895 | 1 |
| SMAD2 | 0.005729868 | 0.162762825 | 0.658 | 0.566 | 1 |
| FCER1G | 0.006669523 | -0.211184199 | 0.992 | 0.986 | 1 |
| IL2RA | 0.006688634 | -0.104327109 | 0.004 | 0.038 | 1 |
| BATF | 0.006822955 | 0.129234785 | 0.291 | 0.168 | 1 |
| PRDM1 | 0.006843393 | 0.216471788 | 0.473 | 0.4 | 1 |
| CD40 | 0.007583627 | -0.299042211 | 0.565 | 0.494 | 1 |
| ICOS | 0.007600376 | 0.068821844 | 0.059 | 0.029 | 1 |

| | | | | | |
|---|---|---|---|---|---|
| B4GALT5 | 0.007995186 | -0.0819928 | 0.03 | 0.086 | 1 |
| BCL10 | 0.008307725 | 0.135638876 | 0.447 | 0.335 | 1 |
| CD3G | 0.008580592 | 0.043314084 | 0.034 | 0.015 | 1 |
| DOCK8 | 0.008699425 | 0.171531083 | 0.831 | 0.726 | 1 |
| ADA | 0.009102301 | 0.282718918 | 0.865 | 0.75 | 1 |
| IKZF1 | 0.009510353 | 0.144180526 | 0.435 | 0.365 | 1 |
| IRF7 | 0.009604929 | -0.235080021 | 0.608 | 0.651 | 1 |
| IL12RB1 | 0.009707292 | -0.089429628 | 0.042 | 0.094 | 1 |
| BCL11B | 0.010011831 | 0.124842842 | 0.19 | 0.124 | 1 |
| LDHA | 0.010402066 | -0.197665436 | 0.949 | 0.942 | 1 |
| CD7 | 0.010504211 | 0.07418925 | 0.076 | 0.043 | 1 |
| TBX21 | 0.011010878 | -0.137900348 | 0.068 | 0.095 | 1 |
| PRNP | 0.011169192 | 0.172233379 | 0.481 | 0.408 | 1 |
| CD28 | 0.011781425 | 0.079102857 | 0.076 | 0.046 | 1 |
| CD86 | 0.012094801 | -0.23631136 | 0.776 | 0.789 | 1 |
| CNOT2 | 0.01300519 | 0.138543752 | 0.578 | 0.477 | 1 |
| DCLRE1C | 0.013307021 | 0.13093152 | 0.287 | 0.211 | 1 |
| PASK | 0.013697187 | 0.191492111 | 0.278 | 0.268 | 1 |
| IRF9 | 0.014561593 | 0.178026983 | 0.945 | 0.888 | 1 |
| LAT2 | 0.016575876 | 0.193684825 | 0.759 | 0.664 | 1 |
| PTPRC | 0.017077027 | 0.150642529 | 0.979 | 0.951 | 1 |
| POU2F2 | 0.017459167 | 0.129761303 | 0.257 | 0.19 | 1 |
| BLNK | 0.018467309 | 0.153183421 | 0.46 | 0.351 | 1 |
| PSME3 | 0.019596398 | 0.131310698 | 0.536 | 0.462 | 1 |
| STAT3 | 0.020854425 | 0.140058903 | 0.962 | 0.88 | 1 |
| MS4A1 | 0.020986389 | -0.193890684 | 0.245 | 0.25 | 1 |
| IGHG2 | 0.022186687 | -0.108583973 | 0.063 | 0.084 | 1 |
| IRF1 | 0.023005643 | -0.247296907 | 0.848 | 0.831 | 1 |
| LGALS3 | 0.02539502 | 0.238323705 | 0.911 | 0.91 | 1 |
| KLRK1 | 0.028733203 | 0.168582044 | 0.439 | 0.397 | 1 |
| LIF | 0.028856204 | -0.056342108 | 0.008 | 0.031 | 1 |
| CD8A | 0.030231115 | 0.13035158 | 0.219 | 0.195 | 1 |
| NRP1 | 0.030274692 | 0.212341169 | 0.692 | 0.689 | 1 |
| TGFB1 | 0.031143254 | 0.066944318 | 0.114 | 0.069 | 1 |
| UGCG | 0.031685235 | -0.210720681 | 0.511 | 0.518 | 1 |
| IL1R1 | 0.032186458 | 0.186989093 | 0.35 | 0.317 | 1 |
| CD14 | 0.032907778 | 0.232368016 | 0.983 | 0.938 | 1 |
| RBCK1 | 0.035375545 | 0.121173699 | 0.506 | 0.446 | 1 |
| EIF5 | 0.040233231 | 0.121030164 | 0.899 | 0.848 | 1 |
| NR4A2 | 0.042260623 | -0.102014408 | 0.034 | 0.048 | 1 |
| CCL2 | 0.042905736 | -0.340662283 | 0.97 | 0.957 | 1 |
| IRF4 | 0.043390615 | -0.146090235 | 0.122 | 0.124 | 1 |
| CCR1 | 0.046182316 | 0.157005711 | 0.945 | 0.909 | 1 |
| CD74 | 0.04863581 | -0.21177408 | 1 | 0.993 | 1 |
| EVI2A | 0.05310404 | 0.134609534 | 0.878 | 0.771 | 1 |
| IFNG | 0.05493389 | -0.066437985 | 0.017 | 0.029 | 1 |
| TNF | 0.055467033 | -0.266224523 | 0.376 | 0.344 | 1 |

| | | | | | |
|---|---|---|---|---|---|
| NFATC1 | 0.055752206 | 0.101554404 | 0.439 | 0.339 | 1 |
| STAT5A | 0.056343342 | 0.118276962 | 0.65 | 0.568 | 1 |
| CLEC10A | 0.056400239 | -0.297763531 | 0.418 | 0.44 | 1 |
| TCF4 | 0.057079816 | -0.055645929 | 0.03 | 0.064 | 1 |
| FOXO1 | 0.057367807 | 0.064765362 | 0.143 | 0.11 | 1 |
| PLCG2 | 0.057674497 | 0.110641612 | 0.667 | 0.58 | 1 |
| PSMB8 | 0.058132029 | 0.128482181 | 0.979 | 0.942 | 1 |
| CTSD | 0.059777425 | 0.222158429 | 0.958 | 0.946 | 1 |
| CXCL16 | 0.061442965 | -0.181382524 | 0.958 | 0.95 | 1 |
| NANS | 0.065158845 | 0.124303136 | 0.911 | 0.858 | 1 |
| GAPDH | 0.065394731 | 0.125280734 | 1 | 0.987 | 1 |
| HLA.DRA | 0.06586296 | -0.200285055 | 0.987 | 0.976 | 1 |
| CD69 | 0.066856321 | -0.110108457 | 0.135 | 0.168 | 1 |
| MAP2K3 | 0.067069457 | -0.146431026 | 0.835 | 0.814 | 1 |
| PIK3IP1 | 0.06857251 | 0.124799356 | 0.443 | 0.41 | 1 |
| NINJ2 | 0.069610726 | 0.079857814 | 0.211 | 0.155 | 1 |
| CD80 | 0.073063529 | -0.124482781 | 0.072 | 0.114 | 1 |
| PDIA6 | 0.074893352 | 0.155170225 | 0.966 | 0.918 | 1 |
| ARHGEF3 | 0.075210151 | -0.088805798 | 0.177 | 0.214 | 1 |
| FCER2 | 0.075686118 | 0.326843097 | 0.451 | 0.245 | 1 |
| KLRG1 | 0.076482318 | -0.035040785 | 0.013 | 0.034 | 1 |
| RPL6 | 0.076585474 | 0.103132937 | 1 | 0.995 | 1 |
| CTSW | 0.080550416 | 0.07071198 | 0.097 | 0.069 | 1 |
| CD8B | 0.084963332 | 0.030586469 | 0.025 | 0.015 | 1 |
| CCR5 | 0.086113345 | -0.16496619 | 0.557 | 0.539 | 1 |
| SPAG9 | 0.086795463 | 0.11119891 | 0.633 | 0.579 | 1 |
| CASP5 | 0.089226738 | -0.117389173 | 0.477 | 0.465 | 1 |
| DPP4 | 0.090649622 | -0.031672567 | 0.004 | 0.019 | 1 |
| BAX | 0.090871494 | 0.105122125 | 0.857 | 0.84 | 1 |
| CREM | 0.0916271 | 0.114176334 | 0.363 | 0.292 | 1 |
| CD4 | 0.093271864 | 0.171789187 | 0.831 | 0.713 | 1 |
| GRB10 | 0.103613446 | 0.060057588 | 0.093 | 0.085 | 1 |
| HAVCR2 | 0.103629177 | 0.150148814 | 0.84 | 0.78 | 1 |
| IFITM2 | 0.10748468 | 0.131537909 | 0.84 | 0.827 | 1 |
| TRBC2 | 0.108235109 | 0.142719033 | 0.722 | 0.756 | 1 |
| RPN2 | 0.113518181 | 0.103273968 | 0.962 | 0.921 | 1 |
| SNCA | 0.114384269 | -0.106376731 | 0.333 | 0.412 | 1 |
| CYCS | 0.114965327 | 0.108980678 | 0.852 | 0.79 | 1 |
| EIF3J | 0.115865288 | 0.062715485 | 0.181 | 0.145 | 1 |
| EGR1 | 0.116120212 | -0.071090344 | 0.059 | 0.091 | 1 |
| IGHG1 | 0.118308247 | -0.121436006 | 0.291 | 0.334 | 1 |
| ILF2 | 0.120222017 | 0.090906956 | 0.873 | 0.855 | 1 |
| GEM | 0.122022877 | -0.041276988 | 0.017 | 0.027 | 1 |
| AHI1 | 0.130629266 | 0.039172203 | 0.059 | 0.041 | 1 |
| APOBEC3G | 0.13100825 | -0.115508592 | 0.511 | 0.499 | 1 |
| POU2AF1 | 0.131645794 | -0.068300729 | 0.059 | 0.091 | 1 |
| CCND2 | 0.134503295 | -0.074705601 | 0.089 | 0.091 | 1 |

| | | | | | |
|---|---|---|---|---|---|
| ADGRE1 | 0.134769697 | 0.085747485 | 0.156 | 0.099 | 1 |
| NFATC2 | 0.137599077 | -0.043964124 | 0.038 | 0.064 | 1 |
| LCK | 0.14098667 | 0.102141846 | 0.35 | 0.344 | 1 |
| CST7 | 0.14162772 | -0.152559868 | 0.325 | 0.327 | 1 |
| CD72 | 0.147512475 | 0.086092505 | 0.131 | 0.138 | 1 |
| IGHG3 | 0.153044298 | -0.072751759 | 0.114 | 0.108 | 1 |
| IL18R1 | 0.15968268 | -0.022515381 | 0.008 | 0.022 | 1 |
| CCR6 | 0.165513359 | 0.037225574 | 0.021 | 0.023 | 1 |
| IL1R2 | 0.166898428 | -0.027691903 | 0.008 | 0.018 | 1 |
| CD52 | 0.168350347 | -0.147938148 | 0.949 | 0.925 | 1 |
| XBP1 | 0.168990697 | 0.091159478 | 0.848 | 0.767 | 1 |
| BACH2 | 0.170133999 | -0.025967027 | 0.017 | 0.032 | 1 |
| MCM2 | 0.172559868 | -0.071927998 | 0.236 | 0.256 | 1 |
| GZMB | 0.172906161 | -0.125166313 | 0.262 | 0.261 | 1 |
| HLA.A | 0.17382732 | 0.081579705 | 1 | 1 | 1 |
| CXCL13 | 0.187647634 | -0.096063154 | 0.013 | 0.011 | 1 |
| NT5E | 0.18807942 | -0.033546104 | 0.03 | 0.041 | 1 |
| ZBED2 | 0.192961509 | 0.010729875 | 0.013 | 0.005 | 1 |
| CCR4 | 0.196320235 | -0.029198585 | 0.021 | 0.032 | 1 |
| IL18RAP | 0.196691021 | -0.032396412 | 0.025 | 0.035 | 1 |
| PSME2 | 0.204631194 | -0.107815784 | 0.979 | 0.962 | 1 |
| PRF1 | 0.205949551 | -0.109987597 | 0.354 | 0.381 | 1 |
| PTPN7 | 0.207379526 | -0.074731526 | 0.228 | 0.272 | 1 |
| IGHA1 | 0.20938692 | -0.034016508 | 0.013 | 0.017 | 1 |
| CD24 | 0.211010765 | -0.021477844 | 0.008 | 0.018 | 1 |
| CCR3 | 0.211509052 | 0.014616324 | 0.008 | 0.007 | 1 |
| NFKBIZ | 0.221503902 | -0.123688202 | 0.443 | 0.484 | 1 |
| JAK3 | 0.223641609 | -0.083857843 | 0.278 | 0.303 | 1 |
| IL6ST | 0.224624973 | -0.079080765 | 0.713 | 0.698 | 1 |
| MME | 0.230816478 | -0.028884183 | 0.004 | 0.019 | 1 |
| CYBB | 0.23590861 | 0.094026687 | 0.987 | 0.957 | 1 |
| CD200 | 0.24271614 | -0.04766935 | 0.021 | 0.03 | 1 |
| IL18 | 0.243206795 | -0.073515817 | 0.16 | 0.214 | 1 |
| PDIA4 | 0.251011595 | 0.096148515 | 0.937 | 0.881 | 1 |
| SKAP1 | 0.251426494 | 0.021876901 | 0.03 | 0.025 | 1 |
| PIK3AP1 | 0.253356957 | 0.093928581 | 0.831 | 0.713 | 1 |
| TNFSF13B | 0.253491149 | -0.119758727 | 0.582 | 0.56 | 1 |
| HLA.DPA1 | 0.258500355 | -0.140502365 | 0.92 | 0.912 | 1 |
| TRAC | 0.259089492 | 0.10030276 | 0.557 | 0.563 | 1 |
| LEF1 | 0.260946453 | 0.075195427 | 0.266 | 0.229 | 1 |
| B3GAT1 | 0.264241035 | -0.015236904 | 0.008 | 0.019 | 1 |
| IL32 | 0.26574496 | -0.07088389 | 0.27 | 0.264 | 1 |
| TNFSF8 | 0.269034531 | -0.019599735 | 0.013 | 0.021 | 1 |
| CD6 | 0.269200465 | 0.06239615 | 0.198 | 0.2 | 1 |
| FAS | 0.273827833 | -0.038204305 | 0.076 | 0.096 | 1 |
| ITGAE | 0.278647314 | -0.102963861 | 0.308 | 0.332 | 1 |
| CD70 | 0.283433118 | -0.015017894 | 0.004 | 0.01 | 1 |

| | | | | | |
|---|---|---|---|---|---|
| TNIP3 | 0.28470716 | -0.012922476 | 0.004 | 0.012 | 1 |
| CXCR2 | 0.286913432 | -0.034344356 | 0.042 | 0.06 | 1 |
| RUNX3 | 0.295886503 | 0.089242774 | 0.595 | 0.556 | 1 |
| IRF8 | 0.296506456 | 0.091688445 | 0.911 | 0.847 | 1 |
| CHI3L2 | 0.297265919 | -0.024268531 | 0.025 | 0.032 | 1 |
| TCL1A | 0.297615075 | 0.0638527 | 0.072 | 0.06 | 1 |
| STAT4 | 0.297760906 | -0.062286342 | 0.156 | 0.165 | 1 |
| HIF1A | 0.305661076 | 0.081627582 | 0.928 | 0.886 | 1 |
| LAMP1 | 0.307147439 | 0.097995031 | 0.945 | 0.922 | 1 |
| F13A1 | 0.307670336 | 0.087933094 | 0.046 | 0.024 | 1 |
| IL23A | 0.313752451 | -0.036064336 | 0.046 | 0.042 | 1 |
| TNFRSF25 | 0.31381962 | 0.022172032 | 0.034 | 0.032 | 1 |
| CHD1 | 0.321536715 | 0.006821477 | 0.008 | 0.004 | 1 |
| CBLB | 0.32170279 | 0.022462519 | 0.051 | 0.039 | 1 |
| SELL | 0.325453436 | 0.079984076 | 0.388 | 0.36 | 1 |
| STIM1 | 0.326269523 | 0.044941006 | 0.338 | 0.289 | 1 |
| AURKB | 0.329313589 | 0.010729875 | 0.008 | 0.004 | 1 |
| CTLA4 | 0.334030995 | -0.041993228 | 0.025 | 0.037 | 1 |
| IFRD1 | 0.347599214 | 0.054479675 | 0.376 | 0.337 | 1 |
| KLRC4 | 0.353261999 | 0.013930277 | 0.025 | 0.013 | 1 |
| IRF3 | 0.353341166 | 0.031190622 | 0.11 | 0.104 | 1 |
| LRRC32 | 0.358269129 | -0.018360749 | 0.004 | 0.012 | 1 |
| MCM4 | 0.358615917 | 0.026547641 | 0.084 | 0.063 | 1 |
| MALT1 | 0.359411344 | 0.043473222 | 0.245 | 0.206 | 1 |
| LRRC8B | 0.364169541 | 0.023393425 | 0.063 | 0.048 | 1 |
| CD1C | 0.369627527 | 0.045144079 | 0.038 | 0.041 | 1 |
| CCL13 | 0.370599347 | 0.267873834 | 0.186 | 0.108 | 1 |
| PGM3 | 0.373252652 | -0.047706288 | 0.278 | 0.295 | 1 |
| TNFSF14 | 0.37416495 | -0.016702454 | 0.017 | 0.023 | 1 |
| IL12RB2 | 0.375827548 | -0.016491594 | 0.013 | 0.017 | 1 |
| BCAT1 | 0.376468727 | -0.067553328 | 0.456 | 0.433 | 1 |
| TRAT1 | 0.395168423 | -0.013376742 | 0.013 | 0.019 | 1 |
| TNFRSF8 | 0.403406769 | 0.030775131 | 0.097 | 0.076 | 1 |
| ARID5A | 0.404689382 | -0.045243763 | 0.257 | 0.281 | 1 |
| JAK1 | 0.406619935 | -0.049893839 | 0.932 | 0.913 | 1 |
| CD63 | 0.413740136 | 0.064138647 | 1 | 0.991 | 1 |
| TIGIT | 0.414679116 | 0.022748471 | 0.055 | 0.041 | 1 |
| NGDN | 0.42166554 | 0.016024175 | 0.034 | 0.022 | 1 |
| CD247 | 0.422413789 | 0.033657849 | 0.122 | 0.105 | 1 |
| SOCS2 | 0.431720701 | 0.051085318 | 0.181 | 0.1 | 1 |
| TUBA1C | 0.432541914 | -0.053900453 | 0.996 | 0.976 | 1 |
| TSPAN32 | 0.432584482 | -0.022848717 | 0.046 | 0.071 | 1 |
| CD79B | 0.434778386 | 0.04140232 | 0.093 | 0.096 | 1 |
| VSIR | 0.436370443 | 0.067022925 | 0.886 | 0.797 | 1 |
| IGHM | 0.44783149 | -0.069447991 | 0.346 | 0.345 | 1 |
| LIG4 | 0.456506059 | -0.036162402 | 0.262 | 0.274 | 1 |
| LAG3 | 0.461576968 | -0.025466204 | 0.055 | 0.046 | 1 |

| | | | | | |
|---|---|---|---|---|---|
| PSMB9 | 0.462609019 | -0.071089344 | 0.823 | 0.728 | 1 |
| CXCL11 | 0.469042055 | -0.044772533 | 0.004 | 0.02 | 1 |
| GIMAP5 | 0.469465405 | -0.058615987 | 0.565 | 0.54 | 1 |
| LAP3 | 0.47035685 | -0.069436907 | 0.924 | 0.875 | 1 |
| MITF | 0.471792346 | -0.06042257 | 0.692 | 0.667 | 1 |
| UFM1 | 0.474283471 | 0.044387976 | 0.857 | 0.817 | 1 |
| NFKBID | 0.479137008 | 0.012218959 | 0.034 | 0.023 | 1 |
| GBP2 | 0.481926014 | 0.059950368 | 0.426 | 0.392 | 1 |
| CSF2 | 0.487521275 | -0.025458094 | 0.013 | 0.022 | 1 |
| ITK | 0.488045739 | 0.041498837 | 0.278 | 0.215 | 1 |
| KLRF1 | 0.491525624 | 0.019058235 | 0.025 | 0.043 | 1 |
| IFI6 | 0.4966299 | 0.086749961 | 0.671 | 0.638 | 1 |
| SP140 | 0.498628007 | 0.030135274 | 0.148 | 0.107 | 1 |
| CD5 | 0.499565089 | 0.038206449 | 0.186 | 0.125 | 1 |
| GZMH | 0.50655773 | 0.025448278 | 0.072 | 0.063 | 1 |
| HLA.DQB1 | 0.506616462 | 0.082926543 | 0.781 | 0.785 | 1 |
| FYN | 0.52553862 | 0.048892624 | 0.346 | 0.358 | 1 |
| FYB1 | 0.528975418 | 0.043996286 | 0.895 | 0.833 | 1 |
| TIAF1 | 0.529045538 | -0.033201656 | 0.274 | 0.282 | 1 |
| IL2 | 0.531035501 | 0.038319756 | 0.021 | 0.023 | 1 |
| ZBTB16 | 0.532356717 | -0.005564453 | 0.004 | 0.008 | 1 |
| GZMA | 0.533292083 | -0.025322132 | 0.089 | 0.084 | 1 |
| TNFAIP8 | 0.538333123 | -0.048589984 | 0.895 | 0.809 | 1 |
| NIBAN3 | 0.543116042 | -0.008968849 | 0.008 | 0.01 | 1 |
| KLRC3 | 0.550727317 | -0.033667651 | 0.139 | 0.133 | 1 |
| PAXBP1 | 0.553478892 | 0.035441629 | 0.426 | 0.404 | 1 |
| MZB1 | 0.559986666 | -0.011760893 | 0.021 | 0.022 | 1 |
| SLC7A1 | 0.560607575 | -0.028635953 | 0.169 | 0.182 | 1 |
| STAT1 | 0.563717605 | 0.058830837 | 0.966 | 0.926 | 1 |
| GZMK | 0.583103504 | 0.020273648 | 0.068 | 0.06 | 1 |
| ENTPD1 | 0.587103889 | -0.03226087 | 0.186 | 0.178 | 1 |
| VPREB3 | 0.59291451 | -0.02453683 | 0.08 | 0.077 | 1 |
| MYC | 0.597251056 | 0.040962762 | 0.371 | 0.288 | 1 |
| EBF1 | 0.609043583 | 0.006197948 | 0.017 | 0.012 | 1 |
| CXCR5 | 0.609221247 | -0.026884417 | 0.089 | 0.075 | 1 |
| CD40LG | 0.616900135 | -0.004510236 | 0.004 | 0.007 | 1 |
| CD79A | 0.618677923 | -0.042614408 | 0.241 | 0.241 | 1 |
| CD22 | 0.631512007 | 0.045752161 | 0.489 | 0.458 | 1 |
| PCLAF | 0.636372958 | 0.005454933 | 0.013 | 0.007 | 1 |
| VEGFA | 0.638012177 | 0.006935201 | 0.021 | 0.014 | 1 |
| IL6R | 0.64745 | 0.028253716 | 0.536 | 0.488 | 1 |
| CXCR3 | 0.653076082 | -0.013970565 | 0.051 | 0.057 | 1 |
| TGIF1 | 0.661561533 | 0.032950692 | 0.726 | 0.683 | 1 |
| TLR7 | 0.673674401 | -0.024830665 | 0.177 | 0.175 | 1 |
| TLR8 | 0.68270673 | -0.012642857 | 0.063 | 0.071 | 1 |
| ANXA5 | 0.683084754 | 0.029259018 | 0.996 | 0.985 | 1 |
| CD244 | 0.687000034 | -0.01383141 | 0.068 | 0.068 | 1 |

| | | | | | |
|---|---|---|---|---|---|
| JAK2 | 0.688388233 | 0.00888463 | 0.046 | 0.041 | 1 |
| RNASE2 | 0.688580654 | 0.00879778 | 0.03 | 0.026 | 1 |
| IFITM3 | 0.691150243 | 0.04456456 | 0.937 | 0.905 | 1 |
| BCL2 | 0.697005901 | 0.017232715 | 0.156 | 0.127 | 1 |
| CARD11 | 0.69832903 | 0.004706088 | 0.008 | 0.003 | 1 |
| CR2 | 0.699454043 | -0.00659179 | 0.021 | 0.023 | 1 |
| ITGB2 | 0.703810642 | -0.033779234 | 0.987 | 0.957 | 1 |
| FOXP1 | 0.709528425 | 0.014758097 | 0.105 | 0.089 | 1 |
| IGHD | 0.719145111 | 0.022544172 | 0.135 | 0.105 | 1 |
| FUT4 | 0.721189189 | 0.018405515 | 0.295 | 0.246 | 1 |
| CD2 | 0.730058557 | 0.02540977 | 0.346 | 0.319 | 1 |
| BCL2L1 | 0.743597618 | 0.016302446 | 0.207 | 0.204 | 1 |
| MAST4 | 0.752967568 | -0.003991631 | 0.013 | 0.015 | 1 |
| CD37 | 0.753201485 | -0.021697733 | 0.861 | 0.85 | 1 |
| CXCL9 | 0.759484336 | 0.152315859 | 0.042 | 0.055 | 1 |
| CHD7 | 0.762801535 | -0.006065367 | 0.021 | 0.03 | 1 |
| CD27 | 0.768735537 | 0.014850059 | 0.169 | 0.142 | 1 |
| CD3E | 0.768832059 | 0.023991608 | 0.549 | 0.498 | 1 |
| CCR2 | 0.771278325 | -0.007585022 | 0.021 | 0.033 | 1 |
| TCF7 | 0.771450587 | -0.006814003 | 0.042 | 0.04 | 1 |
| TRDC | 0.776142402 | -0.016820053 | 0.152 | 0.109 | 1 |
| ZNF341 | 0.782120977 | 0.007260113 | 0.055 | 0.044 | 1 |
| LTA | 0.791477538 | -0.006814003 | 0.034 | 0.037 | 1 |
| TYMS | 0.795663937 | -0.00239949 | 0.004 | 0.004 | 1 |
| SELPLG | 0.812006635 | 0.020564128 | 0.789 | 0.759 | 1 |
| ZAP70 | 0.823548348 | -0.00528933 | 0.038 | 0.052 | 1 |
| FOXP3 | 0.826784613 | -0.003991631 | 0.013 | 0.009 | 1 |
| NCAM1 | 0.84977705 | 0.004684606 | 0.034 | 0.024 | 1 |
| EEF1E1 | 0.853840363 | -0.006656757 | 0.131 | 0.13 | 1 |
| NKG7 | 0.854265355 | -0.016064463 | 0.485 | 0.471 | 1 |
| TMEM97 | 0.855910116 | 0.004096783 | 0.042 | 0.038 | 1 |
| ZNF683 | 0.856765375 | -0.006731739 | 0.063 | 0.074 | 1 |
| TRIB2 | 0.8667371 | 0.00737786 | 0.089 | 0.115 | 1 |
| F5 | 0.867087117 | -0.001342957 | 0.004 | 0.004 | 1 |
| MYB | 0.87217458 | 0.002759525 | 0.021 | 0.015 | 1 |
| IL2RB | 0.878290453 | 0.009834756 | 0.16 | 0.154 | 1 |
| KLRC1 | 0.88420872 | 0.006302473 | 0.046 | 0.065 | 1 |
| BTLA | 0.887540286 | 0.00200711 | 0.017 | 0.013 | 1 |
| THEMIS | 0.897573725 | 0.00200711 | 0.017 | 0.012 | 1 |
| RORA | 0.897962292 | 0.006127257 | 0.118 | 0.136 | 1 |
| RORC | 0.901003357 | 0.000772432 | 0.004 | 0.004 | 1 |
| IL12A | 0.901003357 | 0.000772432 | 0.004 | 0.004 | 1 |
| FCER1A | 0.902164822 | 0.003350345 | 0.008 | 0.003 | 1 |
| REL | 0.910854481 | -0.008223371 | 0.57 | 0.499 | 1 |
| CD3D | 0.913713728 | -0.007993178 | 0.422 | 0.421 | 1 |
| IGBP1 | 0.925834382 | -0.005037441 | 0.354 | 0.347 | 1 |
| FOSL1 | 0.927670459 | 0.003817766 | 0.114 | 0.099 | 1 |

| | | | | | |
|---|---|---|---|---|---|
| YBX3 | 0.938559826 | 0.002883075 | 0.131 | 0.111 | 1 |
| CCL5 | 0.943692457 | -0.006065367 | 0.274 | 0.266 | 1 |
| IL4R | 0.956443655 | -0.003396616 | 0.722 | 0.703 | 1 |
| KIT | 0.965993087 | -0.00028565 | 0.004 | 0.004 | 1 |
| TOP2A | 0.965993087 | -0.00028565 | 0.004 | 0.004 | 1 |
| CXCR6 | 0.968153504 | -0.001463252 | 0.008 | 0.021 | 1 |
| ERBIN | 0.96839098 | 0.002778896 | 0.857 | 0.781 | 1 |
| KLRB1 | 0.970557654 | -0.00028565 | 0.004 | 0.004 | 1 |
| PSEN1 | 0.980711043 | 0.001330642 | 0.468 | 0.42 | 1 |
| LTB | 0.985124623 | -0.000514094 | 0.042 | 0.044 | 1 |
| TNFRSF13C | 0.985153689 | 0.000840968 | 0.131 | 0.129 | 1 |
| HLA.DMA | 0.985705457 | -0.001732296 | 0.924 | 0.904 | 1 |
| FASLG | 0.991528707 | -0.037747082 | 0 | 0.023 | 1 |
| IL11RA | 0.992895715 | 0.000335731 | 0.105 | 0.112 | 1 |
| EGR3 | 0.994701358 | -0.02422984 | 0 | 0.007 | 1 |
| TNFRSF4 | 0.994867239 | -0.013745092 | 0 | 0.006 | 1 |
| IGHG4 | 0.994915809 | -0.011638967 | 0 | 0.007 | 1 |
| TNFRSF17 | 0.99494352 | -0.010584751 | 0 | 0.007 | 1 |
| SOCS1 | 0.995008397 | -0.008474004 | 0 | 0.005 | 1 |
| PDCD1 | 0.995008397 | -0.008474004 | 0 | 0.004 | 1 |
| TPSAB1 | 0.99504722 | -0.007417472 | 0 | 0.004 | 1 |
| NDFIP2 | 0.995145047 | -0.005302083 | 0 | 0.004 | 1 |
| TGFB3 | 0.995145047 | -0.005302083 | 0 | 0.004 | 1 |
| CCR9 | 0.995145047 | -0.005302083 | 0 | 0.003 | 1 |
| CTSG | 0.995209924 | -0.004243224 | 0 | 0.003 | 1 |
| CD34 | 0.995209924 | -0.004243224 | 0 | 0.003 | 1 |
| IL22 | 0.995209924 | -0.004243224 | 0 | 0.003 | 1 |
| CD160 | 0.996724579 | -0.012692414 | 0 | 0.003 | 1 |
| GATA3 | 0.996969046 | -0.003183587 | 0 | 0.002 | 1 |
| AIM2 | 0.996969046 | -0.003183587 | 0 | 0.002 | 1 |
| PAX5 | 0.996969046 | -0.003183587 | 0 | 0.002 | 1 |

NK cells

| | p_val | avg_log2FC | Patient | Controls | p_val_adj |
|---|---|---|---|---|---|
| FCGR3A | 2.37E-16 | 0.867451942 | 0.907 | 0.757 | 1.18E-13 |
| CD52 | 1.71E-10 | 0.866997284 | 0.963 | 0.855 | 8.50E-08 |
| CD3E | 4.32E-10 | 0.847105598 | 0.806 | 0.596 | 2.15E-07 |
| ZNF683 | 1.69E-09 | 1.507683197 | 0.565 | 0.213 | 8.41E-07 |
| IFI6 | 1.87E-08 | -0.727908896 | 0.148 | 0.351 | 9.33E-06 |
| NKG7 | 1.91E-08 | 0.46641519 | 1 | 0.995 | 9.51E-06 |
| GZMH | 2.66E-08 | 0.752446356 | 0.593 | 0.365 | 1.32E-05 |
| SELPLG | 1.88E-07 | 0.418593064 | 0.926 | 0.844 | 9.38E-05 |
| IRF7 | 8.93E-07 | -0.591508045 | 0.343 | 0.518 | 0.000444603 |
| CXCR4 | 1.02E-05 | -0.5368812 | 0.843 | 0.941 | 0.005094291 |
| EVI2B | 1.69E-05 | 0.35718156 | 0.898 | 0.8 | 0.008430526 |
| S100A10 | 2.08E-05 | 0.388012403 | 0.88 | 0.801 | 0.0103357 |

| | | | | | |
|---|---|---|---|---|---|
| DUSP2 | 3.54E-05 | -0.650609948 | 0.296 | 0.456 | 0.01765359 |
| FYN | 4.18E-05 | -0.362014235 | 0.815 | 0.9 | 0.020812134 |
| IGKC | 7.90E-05 | 0.714648075 | 0.685 | 0.622 | 0.039363511 |
| CD2 | 9.17E-05 | 0.473530384 | 0.815 | 0.696 | 0.045689291 |
| NFKBIA | 9.95E-05 | -0.448436744 | 0.824 | 0.806 | 0.049548915 |
| STAT1 | 0.000119515 | -0.510927833 | 0.685 | 0.683 | 0.059518581 |
| ITGB2 | 0.00012841 | 0.320271771 | 0.954 | 0.943 | 0.06394795 |
| IGLC3 | 0.000156078 | -0.549216882 | 0.38 | 0.502 | 0.077726665 |
| KLRC3 | 0.000182819 | 0.458501662 | 0.87 | 0.829 | 0.091044046 |
| GBP1 | 0.000458706 | -0.415231435 | 0.259 | 0.381 | 0.228435361 |
| CD38 | 0.000476528 | -0.351894716 | 0.157 | 0.284 | 0.237310945 |
| CD8A | 0.000493098 | 0.573284609 | 0.333 | 0.324 | 0.245562671 |
| CD3D | 0.000535886 | 0.508463054 | 0.444 | 0.32 | 0.266871291 |
| IFNG | 0.000630327 | -1.399651586 | 0.083 | 0.063 | 0.313902889 |
| FURIN | 0.000771098 | -0.37328447 | 0.454 | 0.573 | 0.384007006 |
| B3GAT1 | 0.000808922 | 0.227681613 | 0.241 | 0.105 | 0.40284301 |
| HAVCR2 | 0.00085654 | -0.39918955 | 0.157 | 0.262 | 0.426556673 |
| CD69 | 0.000857048 | -0.415936679 | 0.333 | 0.485 | 0.426809895 |
| TNFSF10 | 0.000882495 | -0.360706404 | 0.296 | 0.381 | 0.439482668 |
| LAP3 | 0.001125242 | -0.282699539 | 0.148 | 0.293 | 0.560370396 |
| IL32 | 0.0022372 | 0.399312383 | 0.796 | 0.627 | 1 |
| SKAP1 | 0.002442473 | 0.132848016 | 0.167 | 0.086 | 1 |
| TSPAN32 | 0.003169849 | 0.20797333 | 0.361 | 0.21 | 1 |
| SRGN | 0.00344337 | 0.184554687 | 0.296 | 0.154 | 1 |
| TRAC | 0.00381605 | 0.375739015 | 0.463 | 0.498 | 1 |
| RGS1 | 0.004053537 | -0.248080502 | 0.065 | 0.153 | 1 |
| IRF1 | 0.004320303 | -0.322022254 | 0.796 | 0.851 | 1 |
| RBCK1 | 0.005471627 | -0.223195449 | 0.213 | 0.336 | 1 |
| GNAI2 | 0.007060172 | 0.163715626 | 1 | 1 | 1 |
| JCHAIN | 0.007587338 | 0.206542955 | 0.028 | 0.037 | 1 |
| PFN1 | 0.00784019 | 0.184316977 | 1 | 0.998 | 1 |
| SOCS2 | 0.008471893 | 0.151705564 | 0.148 | 0.039 | 1 |
| HLA.DPA1 | 0.008866094 | 0.286077542 | 0.815 | 0.675 | 1 |
| FYB1 | 0.009132326 | 0.222392421 | 0.648 | 0.59 | 1 |
| KLRC4 | 0.009307711 | 0.179933473 | 0.315 | 0.18 | 1 |
| BIRC3 | 0.010862188 | -0.242732362 | 0.222 | 0.351 | 1 |
| IL18RAP | 0.012129919 | -0.300905834 | 0.306 | 0.561 | 1 |
| IL15RA | 0.012252566 | -0.164499934 | 0.074 | 0.16 | 1 |
| SP140 | 0.013321813 | -0.184860887 | 0.185 | 0.276 | 1 |
| CCL2 | 0.013669723 | -0.278957087 | 0.852 | 0.835 | 1 |
| IRF8 | 0.0142258 | -0.255073371 | 0.204 | 0.297 | 1 |
| LEF1 | 0.014246165 | 0.198058021 | 0.213 | 0.16 | 1 |
| IRF9 | 0.015905473 | -0.260809741 | 0.667 | 0.746 | 1 |
| FCN1 | 0.017081528 | 0.104671934 | 0.12 | 0.062 | 1 |
| FCER1G | 0.017653259 | -0.250691815 | 0.648 | 0.804 | 1 |
| CD74 | 0.017662109 | -0.265773625 | 0.972 | 0.978 | 1 |
| PSMB9 | 0.018237705 | -0.227615863 | 0.722 | 0.784 | 1 |

| | | | | | |
|---|---|---|---|---|---|
| TRIB2 | 0.018329216 | 0.236280004 | 0.435 | 0.376 | 1 |
| ZBTB16 | 0.018541378 | -0.137914607 | 0.065 | 0.154 | 1 |
| KLRF1 | 0.019320643 | -0.232147025 | 0.481 | 0.744 | 1 |
| UGCG | 0.019506117 | -0.140969971 | 0.065 | 0.133 | 1 |
| LCK | 0.019532203 | 0.18695134 | 0.898 | 0.837 | 1 |
| IL7R | 0.020434329 | 0.159233779 | 0.185 | 0.119 | 1 |
| CCR1 | 0.022416749 | 0.153837788 | 0.176 | 0.109 | 1 |
| IFITM2 | 0.022471437 | 0.226184257 | 0.981 | 0.966 | 1 |
| CXCL10 | 0.022608813 | 0.135976054 | 0.065 | 0.071 | 1 |
| SELL | 0.025185418 | 0.349176784 | 0.593 | 0.465 | 1 |
| CD226 | 0.026302525 | 0.166229695 | 0.278 | 0.191 | 1 |
| STAT4 | 0.027556158 | -0.187365796 | 0.519 | 0.626 | 1 |
| CD1C | 0.028076112 | 0.032874878 | 0.028 | 0.005 | 1 |
| KCNE3 | 0.030679892 | 0.045422355 | 0.046 | 0.014 | 1 |
| IGBP1 | 0.031266544 | 0.148341319 | 0.315 | 0.231 | 1 |
| CD8B | 0.032064763 | 0.064170755 | 0.056 | 0.015 | 1 |
| NFATC2 | 0.032566001 | -0.177536546 | 0.361 | 0.441 | 1 |
| ADGRG3 | 0.033302362 | -0.131792683 | 0.046 | 0.092 | 1 |
| ITGAM | 0.033314085 | 0.229099408 | 0.454 | 0.307 | 1 |
| GBP2 | 0.035526794 | -0.188205992 | 0.352 | 0.455 | 1 |
| CSF2 | 0.035860976 | -0.63707767 | 0.009 | 0.023 | 1 |
| KLRG1 | 0.036338435 | 0.128713194 | 0.241 | 0.126 | 1 |
| CD244 | 0.038159942 | -0.172609137 | 0.343 | 0.473 | 1 |
| TCF7 | 0.038313763 | 0.095073444 | 0.102 | 0.059 | 1 |
| VSIR | 0.039305045 | 0.156137349 | 0.37 | 0.299 | 1 |
| VMO1 | 0.041161567 | 0.138481954 | 0.148 | 0.106 | 1 |
| IFITM3 | 0.042031446 | -0.343903125 | 0.509 | 0.55 | 1 |
| JAK1 | 0.043965518 | 0.150706955 | 0.954 | 0.963 | 1 |
| HIF1A | 0.044428285 | -0.202792656 | 0.435 | 0.505 | 1 |
| FASLG | 0.045471677 | -0.187555079 | 0.278 | 0.33 | 1 |
| PDIA6 | 0.045942743 | -0.195714712 | 0.444 | 0.538 | 1 |
| IGHD | 0.049996607 | -0.138787644 | 0.037 | 0.065 | 1 |
| LAT2 | 0.05431514 | -0.167372488 | 0.361 | 0.455 | 1 |
| CX3CR1 | 0.054513121 | 0.270743121 | 0.685 | 0.578 | 1 |
| TRBC2 | 0.055082625 | 0.231348011 | 0.898 | 0.883 | 1 |
| MS4A1 | 0.05516718 | -0.226594025 | 0.148 | 0.153 | 1 |
| EIF5A | 0.058600976 | -0.19636641 | 0.574 | 0.65 | 1 |
| SLC1A5 | 0.060375022 | 0.186088858 | 0.278 | 0.16 | 1 |
| GZMK | 0.060676827 | -0.215252266 | 0.083 | 0.146 | 1 |
| KLRB1 | 0.061321149 | -0.083963373 | 0.019 | 0.076 | 1 |
| SPAG9 | 0.062360334 | -0.110452108 | 0.083 | 0.16 | 1 |
| CLEC4E | 0.06246897 | 0.041043914 | 0.046 | 0.017 | 1 |
| TRIB1 | 0.06246897 | 0.041043914 | 0.046 | 0.017 | 1 |
| LTA | 0.066257314 | -0.21463665 | 0.028 | 0.043 | 1 |
| WAS | 0.067984927 | -0.154744681 | 0.417 | 0.512 | 1 |
| EIF1 | 0.068707786 | 0.168874535 | 0.926 | 0.898 | 1 |
| SLA | 0.072148862 | -0.194045823 | 0.537 | 0.562 | 1 |

| | | | | | |
|---|---|---|---|---|---|
| KDELR1 | 0.073403023 | -0.134348388 | 0.269 | 0.325 | 1 |
| TIGIT | 0.073756003 | -0.166329605 | 0.296 | 0.374 | 1 |
| ITGAX | 0.074216548 | 0.255807596 | 0.407 | 0.285 | 1 |
| PSME2 | 0.074517576 | -0.185856301 | 0.769 | 0.809 | 1 |
| ICAM1 | 0.078836838 | -0.136336165 | 0.12 | 0.176 | 1 |
| CD4 | 0.079498942 | 0.109709092 | 0.139 | 0.089 | 1 |
| BCL2 | 0.079999059 | 0.107956605 | 0.093 | 0.054 | 1 |
| IGHM | 0.080645474 | -0.245356088 | 0.231 | 0.228 | 1 |
| APOBEC3G | 0.084500918 | 0.161782951 | 0.88 | 0.864 | 1 |
| DOCK8 | 0.093271289 | 0.139041355 | 0.806 | 0.687 | 1 |
| TRDC | 0.095731016 | -0.199703654 | 0.759 | 0.806 | 1 |
| FOXP1 | 0.09623463 | -0.075099729 | 0.037 | 0.077 | 1 |
| CNOT2 | 0.096421018 | -0.118830502 | 0.259 | 0.314 | 1 |
| BIN2 | 0.100736433 | 0.120645158 | 0.472 | 0.425 | 1 |
| TNFRSF13C | 0.102491796 | -0.103484776 | 0.037 | 0.082 | 1 |
| BCL2L1 | 0.103162364 | 0.074981008 | 0.111 | 0.062 | 1 |
| ATF4 | 0.105551142 | 0.151983067 | 0.815 | 0.77 | 1 |
| TRAF1 | 0.106768871 | -0.084262393 | 0.065 | 0.12 | 1 |
| DUSP5 | 0.11344979 | -0.067545613 | 0.028 | 0.062 | 1 |
| LAT | 0.11639899 | 0.147219732 | 0.611 | 0.582 | 1 |
| PTPRC | 0.117443536 | 0.101818103 | 0.981 | 0.997 | 1 |
| IER3 | 0.117799621 | 0.145504894 | 0.38 | 0.277 | 1 |
| LAG3 | 0.124827985 | -0.151946265 | 0.185 | 0.19 | 1 |
| GAPDH | 0.124969187 | 0.152946573 | 0.972 | 0.949 | 1 |
| AHI1 | 0.125226722 | -0.072414136 | 0.046 | 0.082 | 1 |
| IKZF1 | 0.125577645 | 0.118154777 | 0.444 | 0.37 | 1 |
| POU2F2 | 0.130006093 | -0.064889531 | 0.028 | 0.074 | 1 |
| IVNS1ABP | 0.130481753 | 0.131618187 | 0.472 | 0.478 | 1 |
| PASK | 0.133486382 | 0.125186188 | 0.231 | 0.219 | 1 |
| TUBA1C | 0.146648862 | -0.134511672 | 0.435 | 0.453 | 1 |
| STAT5A | 0.148060283 | -0.108274876 | 0.194 | 0.251 | 1 |
| PSEN1 | 0.150030795 | -0.075777847 | 0.074 | 0.122 | 1 |
| DCLRE1C | 0.150331212 | 0.088639417 | 0.176 | 0.089 | 1 |
| CD72 | 0.151745822 | -0.062275689 | 0.046 | 0.083 | 1 |
| NFKB1 | 0.155868432 | -0.139436484 | 0.5 | 0.516 | 1 |
| DUSP4 | 0.156418442 | -0.116947036 | 0.083 | 0.108 | 1 |
| NFKB2 | 0.156980821 | -0.056669024 | 0.037 | 0.072 | 1 |
| LILRB4 | 0.158645338 | -0.06517462 | 0.056 | 0.106 | 1 |
| ARPC1B | 0.158918235 | 0.12453781 | 0.759 | 0.695 | 1 |
| TNIK | 0.160663 | 0.076225247 | 0.157 | 0.126 | 1 |
| CD14 | 0.161002366 | 0.126360965 | 0.454 | 0.385 | 1 |
| CCL4 | 0.163370359 | -0.316885665 | 0.648 | 0.59 | 1 |
| ADA | 0.169954726 | 0.105263187 | 0.37 | 0.288 | 1 |
| CCL5 | 0.170675307 | 0.180555965 | 0.889 | 0.854 | 1 |
| PRF1 | 0.17094062 | 0.128544552 | 1 | 0.995 | 1 |
| CMKLR1 | 0.173973455 | 0.060543624 | 0.12 | 0.074 | 1 |
| MCM2 | 0.176899166 | -0.082308845 | 0.074 | 0.092 | 1 |

| | | | | | |
|---|---|---|---|---|---|
| IRF4 | 0.186228897 | -0.085479935 | 0.074 | 0.097 | 1 |
| LYN | 0.187946854 | 0.101760339 | 0.546 | 0.451 | 1 |
| CXCL2 | 0.191023587 | 0.094855643 | 0.139 | 0.122 | 1 |
| LAMP3 | 0.192726971 | -0.059266135 | 0.028 | 0.046 | 1 |
| RORA | 0.197896498 | 0.125624535 | 0.583 | 0.556 | 1 |
| CD3G | 0.201145688 | 0.032326697 | 0.046 | 0.022 | 1 |
| ARID5A | 0.20148912 | -0.09393233 | 0.194 | 0.234 | 1 |
| UFM1 | 0.201764658 | 0.109865388 | 0.546 | 0.464 | 1 |
| PDIA4 | 0.209730696 | -0.127133432 | 0.417 | 0.456 | 1 |
| ENTPD1 | 0.209898448 | -0.065480194 | 0.019 | 0.042 | 1 |
| ORAI1 | 0.211572716 | 0.092021335 | 0.426 | 0.385 | 1 |
| CD5 | 0.212793797 | 0.073191186 | 0.083 | 0.082 | 1 |
| CST7 | 0.214748841 | 0.099447025 | 1 | 0.98 | 1 |
| IL2 | 0.215503564 | -0.122064426 | 0.037 | 0.037 | 1 |
| LAMP1 | 0.215610556 | 0.103841316 | 0.63 | 0.549 | 1 |
| LGALS9 | 0.216006083 | -0.094771661 | 0.222 | 0.276 | 1 |
| CCR4 | 0.216850398 | 0.028219874 | 0.028 | 0.017 | 1 |
| NFATC3 | 0.219262722 | 0.099673005 | 0.648 | 0.569 | 1 |
| B4GALT5 | 0.219508694 | 0.038859667 | 0.028 | 0.015 | 1 |
| GAB2 | 0.219935251 | 0.054088447 | 0.074 | 0.042 | 1 |
| REL | 0.222377841 | -0.105251702 | 0.324 | 0.351 | 1 |
| NR4A2 | 0.224096422 | -0.063411814 | 0.019 | 0.04 | 1 |
| TREM1 | 0.231360565 | 0.024051028 | 0.028 | 0.009 | 1 |
| IL12RB1 | 0.233742512 | -0.058657577 | 0.074 | 0.102 | 1 |
| CASP5 | 0.234557513 | -0.077502527 | 0.204 | 0.243 | 1 |
| TNFRSF25 | 0.235664634 | 0.034501068 | 0.037 | 0.018 | 1 |
| PAK1IP1 | 0.240749896 | -0.042563257 | 0.028 | 0.051 | 1 |
| GATA3 | 0.242081376 | 0.038214522 | 0.065 | 0.032 | 1 |
| CCND2 | 0.243176231 | 0.114015491 | 0.241 | 0.182 | 1 |
| CD9 | 0.243520213 | 0.063139045 | 0.157 | 0.128 | 1 |
| POU2AF1 | 0.247737557 | -0.073778269 | 0.037 | 0.062 | 1 |
| CD83 | 0.247999742 | -0.116771322 | 0.12 | 0.126 | 1 |
| IL6ST | 0.249732309 | 0.07732612 | 0.204 | 0.168 | 1 |
| CCL22 | 0.25007562 | -0.105240032 | 0.12 | 0.129 | 1 |
| PRDM1 | 0.250344363 | 0.089787697 | 0.269 | 0.222 | 1 |
| CD36 | 0.25243207 | -0.057646791 | 0.093 | 0.122 | 1 |
| IFRD1 | 0.256880752 | 0.058202464 | 0.157 | 0.116 | 1 |
| IL12RB2 | 0.265307565 | -0.157668368 | 0.222 | 0.228 | 1 |
| CXCR6 | 0.266066789 | 0.07769023 | 0.056 | 0.037 | 1 |
| CD48 | 0.267740573 | -0.091402856 | 0.935 | 0.946 | 1 |
| CHI3L2 | 0.26815039 | -0.051598034 | 0.046 | 0.077 | 1 |
| GIMAP5 | 0.269759183 | -0.101656479 | 0.574 | 0.633 | 1 |
| PSMB8 | 0.270342597 | -0.096493386 | 0.861 | 0.858 | 1 |
| KLRK1 | 0.271084007 | 0.084449793 | 1 | 0.994 | 1 |
| NANS | 0.275380409 | 0.068458364 | 0.167 | 0.114 | 1 |
| IL1RN | 0.283370098 | 0.060641351 | 0.102 | 0.102 | 1 |
| RPL6 | 0.283638309 | 0.075357429 | 1 | 1 | 1 |

| | | | | | |
|---|---|---|---|---|---|
| TRAT1 | 0.28743812 | 0.021853472 | 0.028 | 0.011 | 1 |
| IL1B | 0.287543566 | 0.114534882 | 0.259 | 0.2 | 1 |
| CD22 | 0.294395328 | -0.072501227 | 0.074 | 0.069 | 1 |
| CLEC10A | 0.296334005 | 0.034214779 | 0.046 | 0.026 | 1 |
| CD40 | 0.300925169 | -0.055690594 | 0.093 | 0.109 | 1 |
| SNCA | 0.304028638 | -0.036249483 | 0.019 | 0.048 | 1 |
| CXCL3 | 0.307189239 | 0.080227177 | 0.12 | 0.116 | 1 |
| CXCR3 | 0.30796116 | -0.110784297 | 0.38 | 0.404 | 1 |
| IL15 | 0.309430894 | -0.034138736 | 0.028 | 0.048 | 1 |
| CD163 | 0.31403949 | -0.044679799 | 0.065 | 0.105 | 1 |
| IGHG1 | 0.317186702 | 0.091686292 | 0.306 | 0.245 | 1 |
| IL2RG | 0.322242431 | 0.076897362 | 0.972 | 0.968 | 1 |
| NFKBIZ | 0.322407059 | -0.078506607 | 0.25 | 0.25 | 1 |
| KLRC1 | 0.322704429 | 0.131444363 | 0.713 | 0.598 | 1 |
| DUSP6 | 0.329972802 | -0.042169454 | 0.028 | 0.051 | 1 |
| TNFSF13B | 0.330347179 | 0.039699371 | 0.065 | 0.046 | 1 |
| ITGAE | 0.339410515 | -0.078094499 | 0.148 | 0.185 | 1 |
| GZMA | 0.341089331 | -0.115640838 | 0.648 | 0.69 | 1 |
| ARL4C | 0.346805276 | 0.084686825 | 0.824 | 0.786 | 1 |
| CD44 | 0.347157673 | 0.097381898 | 0.917 | 0.901 | 1 |
| FTH1 | 0.347230957 | -0.089235248 | 0.981 | 0.969 | 1 |
| SMAD2 | 0.350428451 | 0.061883876 | 0.25 | 0.23 | 1 |
| BATF | 0.35173402 | 0.040807127 | 0.111 | 0.071 | 1 |
| THBS1 | 0.352667312 | -0.093727058 | 0.546 | 0.538 | 1 |
| IFNGR1 | 0.354172033 | -0.068164663 | 0.213 | 0.277 | 1 |
| LIG4 | 0.356645673 | -0.039354439 | 0.056 | 0.074 | 1 |
| PGM3 | 0.359484946 | 0.040807127 | 0.093 | 0.071 | 1 |
| TGFBR1 | 0.362038669 | 0.061118354 | 0.296 | 0.277 | 1 |
| SLC7A7 | 0.365961972 | 0.038750541 | 0.102 | 0.074 | 1 |
| SNX9 | 0.366032841 | -0.037303699 | 0.056 | 0.076 | 1 |
| CYBB | 0.366968313 | -0.064176561 | 0.25 | 0.263 | 1 |
| STIM1 | 0.367134832 | 0.054313176 | 0.241 | 0.237 | 1 |
| IL6R | 0.375783948 | -0.019667941 | 0.009 | 0.023 | 1 |
| NRP1 | 0.378644995 | -0.023770543 | 0.019 | 0.035 | 1 |
| LIF | 0.384062523 | -0.092932568 | 0.019 | 0.011 | 1 |
| IGHA1 | 0.38489025 | -0.059813293 | 0.009 | 0.017 | 1 |
| CREM | 0.385369563 | -0.061738394 | 0.093 | 0.149 | 1 |
| CD247 | 0.389414252 | -0.076932234 | 0.667 | 0.721 | 1 |
| CXCL16 | 0.393132414 | -0.047727157 | 0.12 | 0.157 | 1 |
| MGST1 | 0.398652232 | -0.023770543 | 0.019 | 0.032 | 1 |
| PSME3 | 0.399508331 | -0.047018854 | 0.111 | 0.119 | 1 |
| CD274 | 0.402636128 | -0.039354439 | 0.056 | 0.065 | 1 |
| VPREB3 | 0.403465918 | -0.052683676 | 0.037 | 0.072 | 1 |
| TCF4 | 0.40684203 | 0.019659258 | 0.028 | 0.011 | 1 |
| DPP4 | 0.408013511 | 0.019659258 | 0.028 | 0.011 | 1 |
| CTSD | 0.409048742 | -0.073326368 | 0.509 | 0.501 | 1 |
| GNLY | 0.409238076 | 0.144300139 | 0.954 | 0.894 | 1 |

| | | | | | |
|---|---|---|---|---|---|
| NT5E | 0.415248021 | 0.017468377 | 0.028 | 0.014 | 1 |
| IL3RA | 0.416357296 | -0.029624201 | 0.028 | 0.052 | 1 |
| CTSW | 0.423202041 | -0.092119395 | 0.722 | 0.8 | 1 |
| TNFAIP8 | 0.423614852 | 0.076497427 | 0.417 | 0.388 | 1 |
| HLA.A | 0.430765974 | 0.046741026 | 1 | 1 | 1 |
| CD27 | 0.430776795 | 0.051519961 | 0.13 | 0.119 | 1 |
| ERBIN | 0.431805839 | 0.066808371 | 0.593 | 0.549 | 1 |
| AQP9 | 0.446651095 | -0.023538431 | 0.028 | 0.042 | 1 |
| VNN2 | 0.449378659 | 0.053283266 | 0.204 | 0.185 | 1 |
| IL18 | 0.450868899 | 0.039377664 | 0.065 | 0.045 | 1 |
| STAT6 | 0.465698117 | 0.056312181 | 0.287 | 0.279 | 1 |
| ICOS | 0.466580269 | 0.017331558 | 0.037 | 0.025 | 1 |
| TMEM97 | 0.47746366 | 0.021503728 | 0.037 | 0.028 | 1 |
| CD37 | 0.482533867 | 0.069744226 | 0.796 | 0.769 | 1 |
| ABCE1 | 0.485436714 | -0.04472575 | 0.176 | 0.214 | 1 |
| RNASE6 | 0.492260507 | -0.023538431 | 0.028 | 0.035 | 1 |
| TYMS | 0.494544002 | -0.079256737 | 0.009 | 0.005 | 1 |
| LGALS3 | 0.496903203 | -0.033995523 | 0.083 | 0.102 | 1 |
| BAX | 0.498473317 | 0.053680448 | 0.398 | 0.419 | 1 |
| TGFB3 | 0.500217529 | 0.010994875 | 0.019 | 0.011 | 1 |
| ANXA5 | 0.501399933 | 0.057934098 | 0.444 | 0.371 | 1 |
| LIPA | 0.503546312 | 0.054152457 | 0.546 | 0.507 | 1 |
| S100A9 | 0.507036316 | -0.062172 | 0.287 | 0.327 | 1 |
| IL4R | 0.510481028 | -0.055919377 | 0.296 | 0.334 | 1 |
| PLCG2 | 0.524506046 | -0.061147129 | 0.463 | 0.488 | 1 |
| IL23A | 0.528615335 | 0.021333488 | 0.046 | 0.035 | 1 |
| MITF | 0.529269309 | 0.017197172 | 0.046 | 0.034 | 1 |
| CCL3 | 0.531093967 | -0.135118122 | 0.398 | 0.418 | 1 |
| HLA.DQB1 | 0.537196917 | -0.079015183 | 0.352 | 0.273 | 1 |
| IGHG2 | 0.537632358 | 0.041135762 | 0.056 | 0.042 | 1 |
| XBP1 | 0.538286876 | 0.066750695 | 0.519 | 0.481 | 1 |
| SDCBP | 0.548434594 | -0.054113319 | 0.574 | 0.638 | 1 |
| TCL1A | 0.549891157 | -0.03525004 | 0.037 | 0.046 | 1 |
| CXCR5 | 0.557838923 | -0.030847156 | 0.056 | 0.06 | 1 |
| MYC | 0.56132587 | -0.043945516 | 0.111 | 0.106 | 1 |
| PCLAF | 0.562939581 | -0.034797466 | 0.009 | 0.006 | 1 |
| IL18R1 | 0.564485397 | 0.054454624 | 0.194 | 0.216 | 1 |
| CYCS | 0.575362381 | 0.046198806 | 0.38 | 0.313 | 1 |
| DUSP1 | 0.580388745 | 0.043143319 | 0.278 | 0.25 | 1 |
| BCL2A1 | 0.582887052 | -0.041012429 | 0.148 | 0.131 | 1 |
| ILF2 | 0.588942275 | -0.048305551 | 0.417 | 0.43 | 1 |
| S100A12 | 0.597597346 | -0.015001749 | 0.019 | 0.039 | 1 |
| CCR7 | 0.599057899 | 0.061013996 | 0.407 | 0.37 | 1 |
| CD300A | 0.599431294 | -0.050730069 | 0.778 | 0.829 | 1 |
| GIMAP2 | 0.609258476 | -0.046478526 | 0.62 | 0.632 | 1 |
| TGFBR2 | 0.610975211 | -0.041102767 | 0.741 | 0.69 | 1 |
| VPS28 | 0.62016947 | 0.040640268 | 0.583 | 0.592 | 1 |

| | | | | | |
|---|---|---|---|---|---|
| IL11RA | 0.622885645 | -0.037303699 | 0.167 | 0.177 | 1 |
| EVI2A | 0.623314298 | 0.041999124 | 0.407 | 0.471 | 1 |
| FCER2 | 0.627680715 | 0.021333488 | 0.056 | 0.025 | 1 |
| BACH2 | 0.633000687 | 0.018903406 | 0.046 | 0.055 | 1 |
| LGALS1 | 0.635678103 | 0.058214614 | 0.759 | 0.669 | 1 |
| FUT4 | 0.640316656 | -0.012859662 | 0.028 | 0.037 | 1 |
| IGHG3 | 0.640955229 | 0.030793817 | 0.046 | 0.049 | 1 |
| BCL6 | 0.641053628 | 0.014935717 | 0.056 | 0.042 | 1 |
| MMP9 | 0.646951788 | 0.021907031 | 0.12 | 0.114 | 1 |
| CARD11 | 0.647174585 | -0.008763164 | 0.009 | 0.014 | 1 |
| CTLA4 | 0.653137994 | -0.010831488 | 0.019 | 0.025 | 1 |
| NINJ2 | 0.653244838 | 0.012999137 | 0.028 | 0.025 | 1 |
| TGFBI | 0.65677914 | -0.036567818 | 0.296 | 0.364 | 1 |
| IRF3 | 0.658028554 | 0.020372012 | 0.111 | 0.083 | 1 |
| CSF1 | 0.670871224 | -0.008763164 | 0.009 | 0.012 | 1 |
| CD6 | 0.677525102 | 0.037603953 | 0.407 | 0.348 | 1 |
| CXCL8 | 0.683614778 | 0.045772381 | 0.481 | 0.487 | 1 |
| MALT1 | 0.704299834 | -0.016153864 | 0.074 | 0.091 | 1 |
| CBLB | 0.707437166 | 0.010761347 | 0.046 | 0.039 | 1 |
| SNX8 | 0.708822506 | 0.01834433 | 0.102 | 0.08 | 1 |
| NFATC1 | 0.71041172 | 0.021283001 | 0.148 | 0.133 | 1 |
| PTPN7 | 0.714483543 | 0.030713222 | 0.398 | 0.428 | 1 |
| GEM | 0.715837822 | 0.004432309 | 0.009 | 0.006 | 1 |
| CD209 | 0.715837822 | 0.004432309 | 0.009 | 0.006 | 1 |
| ADGRE1 | 0.715837822 | 0.004432309 | 0.009 | 0.006 | 1 |
| RAG1 | 0.715837822 | 0.004432309 | 0.009 | 0.006 | 1 |
| FOXO1 | 0.719448048 | -0.014406009 | 0.065 | 0.068 | 1 |
| CCR6 | 0.721531148 | -0.006572283 | 0.009 | 0.012 | 1 |
| HNRNPC | 0.72925284 | 0.031224537 | 0.556 | 0.499 | 1 |
| JUNB | 0.731683323 | 0.019641718 | 0.148 | 0.112 | 1 |
| BLNK | 0.739901941 | -0.008663651 | 0.019 | 0.02 | 1 |
| ATF6B | 0.741038231 | 0.024386315 | 0.333 | 0.388 | 1 |
| MAP2K3 | 0.74145587 | 0.029426393 | 0.639 | 0.598 | 1 |
| CCR5 | 0.748159016 | 0.012809416 | 0.028 | 0.039 | 1 |
| CD79A | 0.749670843 | -0.045393355 | 0.222 | 0.16 | 1 |
| CD63 | 0.761379784 | 0.026527913 | 0.889 | 0.903 | 1 |
| FOSB | 0.761971572 | -0.017315521 | 0.019 | 0.008 | 1 |
| ZAP70 | 0.762693164 | 0.022524227 | 0.417 | 0.394 | 1 |
| CXCR2 | 0.762865965 | 0.012717039 | 0.046 | 0.045 | 1 |
| CD79B | 0.763154929 | 0.024420411 | 0.157 | 0.12 | 1 |
| FAS | 0.767949329 | -0.012212718 | 0.074 | 0.076 | 1 |
| RPN2 | 0.773148423 | 0.025484571 | 0.546 | 0.482 | 1 |
| CD33 | 0.777679859 | -0.006492552 | 0.019 | 0.022 | 1 |
| TGFB1 | 0.777679859 | -0.006492552 | 0.019 | 0.022 | 1 |
| RUNX3 | 0.783781711 | 0.022494494 | 0.917 | 0.909 | 1 |
| NAMPT | 0.785355008 | -0.018134423 | 0.167 | 0.162 | 1 |
| EOMES | 0.787336943 | -0.00437807 | 0.009 | 0.012 | 1 |

| | | | | | |
|---|---|---|---|---|---|
| CHD7 | 0.787336943 | -0.00437807 | 0.009 | 0.012 | 1 |
| GZMB | 0.789059619 | -0.035616583 | 0.944 | 0.935 | 1 |
| CD160 | 0.800729018 | -0.01259405 | 0.046 | 0.031 | 1 |
| IL2RB | 0.803751724 | 0.025322313 | 0.926 | 0.977 | 1 |
| NCAM1 | 0.803758734 | -0.024247546 | 0.148 | 0.214 | 1 |
| PAXBP1 | 0.808347588 | 0.019616268 | 0.296 | 0.288 | 1 |
| ARHGEF3 | 0.811853569 | 0.020556488 | 0.565 | 0.487 | 1 |
| JUN | 0.812976811 | 0.017213839 | 0.139 | 0.125 | 1 |
| IL2RA | 0.813546009 | -0.008763164 | 0.009 | 0.005 | 1 |
| KIT | 0.816160941 | 0.027248689 | 0.056 | 0.057 | 1 |
| MZB1 | 0.817235784 | 0.008622436 | 0.037 | 0.031 | 1 |
| FN1 | 0.819066463 | 0.01939052 | 0.25 | 0.296 | 1 |
| TOP2A | 0.81950551 | -0.006572283 | 0.009 | 0.006 | 1 |
| CXCL5 | 0.822405759 | 0.024357684 | 0.481 | 0.493 | 1 |
| CXCL1 | 0.822512598 | -0.025534374 | 0.25 | 0.291 | 1 |
| IL1A | 0.825003706 | -0.00437807 | 0.009 | 0.009 | 1 |
| PIK3IP1 | 0.829547288 | -0.019293897 | 0.648 | 0.653 | 1 |
| SLC7A1 | 0.830661116 | -0.008282993 | 0.046 | 0.054 | 1 |
| STAT3 | 0.832366712 | 0.020372012 | 0.593 | 0.598 | 1 |
| IL12A | 0.844158731 | 0.004412224 | 0.009 | 0.014 | 1 |
| PCNA | 0.848564339 | -0.01714066 | 0.259 | 0.262 | 1 |
| PIK3AP1 | 0.849255745 | 0.013489491 | 0.306 | 0.297 | 1 |
| NGDN | 0.849579377 | 0.004392502 | 0.028 | 0.025 | 1 |
| TLR2 | 0.849579377 | 0.004392502 | 0.028 | 0.025 | 1 |
| TNFSF14 | 0.850875962 | -0.006337348 | 0.028 | 0.035 | 1 |
| SLC25A37 | 0.851093247 | 0.010065418 | 0.12 | 0.111 | 1 |
| MAST4 | 0.8560739 | 0.004392502 | 0.028 | 0.023 | 1 |
| JAK3 | 0.85791396 | 0.015583048 | 0.574 | 0.565 | 1 |
| CSRNP1 | 0.861522174 | -0.010967935 | 0.148 | 0.149 | 1 |
| EIF3J | 0.863250694 | 0.006412462 | 0.065 | 0.054 | 1 |
| LDHA | 0.864364638 | -0.01714066 | 0.796 | 0.801 | 1 |
| CD28 | 0.865311386 | -0.004318181 | 0.019 | 0.017 | 1 |
| HLA.DRA | 0.87030992 | 0.022906428 | 0.722 | 0.716 | 1 |
| BTG1 | 0.874914033 | 0.012851271 | 0.991 | 0.988 | 1 |
| EIF5 | 0.88516616 | -0.011450272 | 0.435 | 0.448 | 1 |
| VSIG4 | 0.888611939 | 0.004392502 | 0.028 | 0.017 | 1 |
| CXCL13 | 0.889877446 | 0.002224665 | 0.009 | 0.006 | 1 |
| BCL10 | 0.890350192 | -0.005903216 | 0.083 | 0.092 | 1 |
| HLA.DMA | 0.891444366 | -0.014649153 | 0.38 | 0.317 | 1 |
| MCM4 | 0.892065206 | -0.006337348 | 0.037 | 0.022 | 1 |
| EEF1E1 | 0.896834513 | 0.004354105 | 0.046 | 0.037 | 1 |
| BCL11B | 0.902444993 | 0.011269805 | 0.296 | 0.297 | 1 |
| CD24 | 0.902489968 | -0.004259368 | 0.019 | 0.022 | 1 |
| ITGA4 | 0.905130946 | 0.011457945 | 0.481 | 0.516 | 1 |
| CD7 | 0.913179979 | 0.00948535 | 0.361 | 0.342 | 1 |
| LTB | 0.919436509 | 0.004335412 | 0.046 | 0.039 | 1 |
| SOCS3 | 0.922117785 | -0.002140527 | 0.019 | 0.018 | 1 |

| | | | | | |
|---|---|---|---|---|---|
| FOS | 0.922902865 | 0.007461313 | 0.213 | 0.199 | 1 |
| ZNF341 | 0.927274802 | -0.002140527 | 0.019 | 0.017 | 1 |
| CR2 | 0.927274802 | -0.002140527 | 0.019 | 0.017 | 1 |
| TBX21 | 0.929961279 | 0.00840324 | 0.75 | 0.75 | 1 |
| EGR1 | 0.93883092 | 0.004392502 | 0.019 | 0.008 | 1 |
| CD86 | 0.947550119 | 0.002224665 | 0.028 | 0.042 | 1 |
| ITK | 0.951685232 | -0.005203719 | 0.611 | 0.627 | 1 |
| TIAF1 | 0.953354023 | -0.003444085 | 0.139 | 0.151 | 1 |
| FBXO11 | 0.954211394 | 0.003883889 | 0.287 | 0.276 | 1 |
| TGIF1 | 0.958862992 | 0.004884012 | 0.435 | 0.391 | 1 |
| LY86 | 0.966594129 | -0.001915065 | 0.065 | 0.06 | 1 |
| TIMM9 | 0.968874482 | -0.002846324 | 0.25 | 0.243 | 1 |
| TNF | 0.993470249 | -0.406118182 | 0 | 0.068 | 1 |
| OSM | 0.99426675 | -0.039468283 | 0 | 0.014 | 1 |
| CD109 | 0.994304863 | -0.035135862 | 0 | 0.023 | 1 |
| IL6 | 0.994397953 | -0.026431794 | 0 | 0.015 | 1 |
| BCAT1 | 0.994426109 | -0.024247546 | 0 | 0.012 | 1 |
| CD200 | 0.994491043 | -0.019869106 | 0 | 0.014 | 1 |
| CCL13 | 0.994491043 | -0.019869106 | 0 | 0.014 | 1 |
| THEMIS | 0.994529156 | -0.017674892 | 0 | 0.011 | 1 |
| SPP1 | 0.994572365 | -0.015477337 | 0 | 0.011 | 1 |
| BTLA | 0.994572365 | -0.015477337 | 0 | 0.011 | 1 |
| TLR7 | 0.994572365 | -0.015477337 | 0 | 0.011 | 1 |
| JAK2 | 0.994622247 | -0.013276428 | 0 | 0.008 | 1 |
| PRNP | 0.995326429 | 0.000404229 | 0.176 | 0.166 | 1 |
| FOXP3 | 0.996511689 | -0.015477337 | 0 | 0.008 | 1 |
| IGHG4 | 0.996511689 | -0.015477337 | 0 | 0.008 | 1 |
| TNFRSF4 | 0.996511689 | -0.015477337 | 0 | 0.006 | 1 |
| NFKBID | 0.996541944 | -0.013276428 | 0 | 0.009 | 1 |
| NIBAN3 | 0.996541944 | -0.013276428 | 0 | 0.009 | 1 |
| IL23R | 0.996541944 | -0.013276428 | 0 | 0.005 | 1 |
| RAPH1 | 0.996577727 | -0.011072157 | 0 | 0.008 | 1 |
| EBF1 | 0.996577728 | -0.011072157 | 0 | 0.006 | 1 |
| CD80 | 0.996621524 | -0.008864514 | 0 | 0.006 | 1 |
| YBX3 | 0.996621524 | -0.008864514 | 0 | 0.006 | 1 |
| BLK | 0.996621524 | -0.008864514 | 0 | 0.005 | 1 |
| LRRC32 | 0.996621524 | -0.008864514 | 0 | 0.005 | 1 |
| CD70 | 0.997604485 | -0.058804768 | 0 | 0.009 | 1 |
| LRRC8B | 0.99886094 | 2.04E-05 | 0.009 | 0.009 | 1 |
| CXCL9 | 0.99886094 | 2.04E-05 | 0.009 | 0.009 | 1 |
| IKZF2 | 0.99886094 | 2.04E-05 | 0.009 | 0.009 | 1 |
| QPCT | 0.99886094 | 2.04E-05 | 0.009 | 0.009 | 1 |
| IL1R1 | 0.999015968 | 2.04E-05 | 0.009 | 0.008 | 1 |
| CCL20 | 0.999170338 | 2.04E-05 | 0.009 | 0.006 | 1 |

**Stimulated samples (4 hours stimulation with 50ng/mL PMA + 1uM Ionomycin)**

CD4+ Naïve

| | p_val | avg_log2FC | Patient | Controls | p_val_adj |
|---|---|---|---|---|---|
| APOBEC3G | 4.45E-75 | 0.706885736 | 0.475 | 0.305 | 2.22E-72 |
| JAK1 | 9.00E-74 | 0.578587297 | 0.88 | 0.721 | 4.48E-71 |
| CD40LG | 9.15E-70 | -0.788893354 | 0.215 | 0.468 | 4.56E-67 |
| IL2RB | 4.81E-66 | 0.606452877 | 0.799 | 0.615 | 2.39E-63 |
| S100A10 | 3.50E-59 | 0.757674688 | 0.618 | 0.504 | 1.74E-56 |
| FYN | 1.30E-52 | -0.48384544 | 0.801 | 0.886 | 6.49E-50 |
| GZMB | 2.97E-49 | 0.705481856 | 0.931 | 0.929 | 1.48E-46 |
| CXCR3 | 2.86E-46 | 0.736431832 | 0.39 | 0.188 | 1.43E-43 |
| IFNG | 7.40E-44 | 0.732725114 | 0.987 | 0.978 | 3.69E-41 |
| LDHA | 1.06E-40 | 0.318521542 | 1 | 0.998 | 5.30E-38 |
| BTG1 | 2.27E-38 | -0.494967065 | 0.979 | 0.992 | 1.13E-35 |
| GBP1 | 1.53E-32 | 0.498009249 | 0.732 | 0.614 | 7.64E-30 |
| TIGIT | 3.61E-31 | 0.234724906 | 0.145 | 0.045 | 1.80E-28 |
| YBX3 | 7.97E-30 | 0.301614173 | 0.306 | 0.146 | 3.97E-27 |
| LEF1 | 4.26E-29 | 0.491669469 | 0.606 | 0.454 | 2.12E-26 |
| DUSP6 | 5.28E-29 | -0.523274052 | 0.342 | 0.483 | 2.63E-26 |
| ITGA4 | 2.36E-28 | 0.18220038 | 0.188 | 0.076 | 1.17E-25 |
| PDIA6 | 2.57E-27 | 0.375299886 | 0.878 | 0.764 | 1.28E-24 |
| CCL5 | 2.11E-25 | 0.357077578 | 0.2 | 0.107 | 1.05E-22 |
| STAT3 | 8.97E-25 | 0.304329476 | 0.802 | 0.706 | 4.47E-22 |
| CD226 | 3.90E-24 | 0.226632832 | 0.191 | 0.083 | 1.94E-21 |
| DUSP2 | 4.54E-24 | -0.42073881 | 0.979 | 0.981 | 2.26E-21 |
| CD69 | 5.75E-23 | -0.294098167 | 1 | 1 | 2.86E-20 |
| TGFBR2 | 6.52E-23 | 0.378402352 | 0.728 | 0.573 | 3.25E-20 |
| IL7R | 8.15E-23 | -0.34067239 | 0.369 | 0.525 | 4.06E-20 |
| RGS1 | 1.30E-22 | 0.609791034 | 0.537 | 0.366 | 6.48E-20 |
| CCL3 | 4.62E-22 | 0.333512564 | 0.998 | 0.997 | 2.30E-19 |
| BCAT1 | 1.46E-20 | 0.170249199 | 0.147 | 0.064 | 7.28E-18 |
| FASLG | 1.18E-18 | -0.344436445 | 0.147 | 0.241 | 5.87E-16 |
| CD3E | 4.15E-18 | -0.306669407 | 0.773 | 0.797 | 2.07E-15 |
| IL17F | 4.26E-18 | 0.534662771 | 0.034 | 0.037 | 2.12E-15 |
| SLA | 5.25E-18 | -0.394358269 | 0.664 | 0.735 | 2.61E-15 |
| HLA.A | 8.69E-18 | 0.226852914 | 1 | 1 | 4.33E-15 |
| CSF1 | 2.52E-17 | 0.357361532 | 0.403 | 0.276 | 1.25E-14 |
| GBP2 | 5.81E-17 | 0.257426754 | 0.951 | 0.913 | 2.89E-14 |
| LAG3 | 7.33E-16 | 0.150904862 | 0.094 | 0.041 | 3.65E-13 |
| PSME2 | 1.06E-15 | 0.219864191 | 0.931 | 0.857 | 5.28E-13 |
| NFATC1 | 2.79E-15 | 0.188924812 | 0.297 | 0.193 | 1.39E-12 |
| EIF5A | 3.90E-15 | 0.175988926 | 0.998 | 0.998 | 1.94E-12 |
| PFN1 | 7.39E-15 | 0.183425514 | 1 | 0.999 | 3.68E-12 |
| CD83 | 1.07E-14 | -0.356493764 | 0.663 | 0.723 | 5.32E-12 |
| RPN2 | 2.30E-14 | 0.207284902 | 0.638 | 0.546 | 1.14E-11 |
| GIMAP5 | 2.83E-14 | 0.195447561 | 0.181 | 0.104 | 1.41E-11 |

| | | | | | |
|---|---|---|---|---|---|
| NAMPT | 3.39E-14 | 0.314451698 | 0.986 | 0.973 | 1.69E-11 |
| EVI2B | 3.54E-14 | -0.227916485 | 0.205 | 0.296 | 1.76E-11 |
| TBX21 | 4.77E-14 | 0.328989181 | 0.867 | 0.793 | 2.37E-11 |
| BCL11B | 8.70E-14 | 0.268945254 | 0.439 | 0.325 | 4.33E-11 |
| KLRK1 | 1.82E-13 | 0.367126526 | 0.363 | 0.214 | 9.04E-11 |
| CSF2 | 2.82E-13 | 0.327831154 | 0.885 | 0.868 | 1.40E-10 |
| PASK | 3.69E-13 | 0.215256042 | 0.142 | 0.087 | 1.84E-10 |
| CD8A | 6.52E-13 | 0.382802348 | 0.25 | 0.122 | 3.25E-10 |
| TGIF1 | 8.61E-13 | -0.252850118 | 0.455 | 0.543 | 4.29E-10 |
| STAT5A | 9.38E-13 | -0.178083019 | 0.993 | 0.992 | 4.67E-10 |
| IVNS1ABP | 1.02E-12 | 0.208405895 | 0.716 | 0.611 | 5.08E-10 |
| FURIN | 1.18E-12 | 0.273795102 | 0.659 | 0.611 | 5.88E-10 |
| IRF4 | 4.31E-12 | 0.198670625 | 0.999 | 0.988 | 2.15E-09 |
| IRF1 | 6.15E-12 | 0.225922266 | 0.874 | 0.824 | 3.06E-09 |
| CD109 | 2.13E-11 | -0.185889543 | 0.11 | 0.191 | 1.06E-08 |
| SRGN | 2.28E-11 | 0.194608163 | 0.881 | 0.804 | 1.13E-08 |
| CST7 | 2.49E-11 | 0.169673663 | 0.23 | 0.176 | 1.24E-08 |
| ORAI1 | 3.58E-11 | -0.188382213 | 0.445 | 0.534 | 1.78E-08 |
| CCL4 | 4.42E-11 | 0.235641962 | 1 | 1 | 2.20E-08 |
| EIF5 | 4.75E-11 | -0.172852142 | 0.866 | 0.899 | 2.37E-08 |
| IL6ST | 6.56E-11 | -0.220469754 | 0.391 | 0.454 | 3.27E-08 |
| TNF | 6.87E-11 | -0.335554866 | 0.895 | 0.906 | 3.42E-08 |
| IL2RG | 1.91E-10 | 0.174996246 | 0.885 | 0.837 | 9.50E-08 |
| EVI2A | 2.16E-10 | -0.194612258 | 0.298 | 0.364 | 1.08E-07 |
| CD74 | 3.64E-10 | 0.200634877 | 0.655 | 0.608 | 1.81E-07 |
| UGCG | 6.81E-10 | 0.151473608 | 0.295 | 0.224 | 3.39E-07 |
| HNRNPC | 1.68E-09 | 0.158601565 | 0.929 | 0.905 | 8.38E-07 |
| EIF1 | 2.81E-09 | 0.12273895 | 1 | 1 | 1.40E-06 |
| SLC7A1 | 8.83E-09 | 0.160466434 | 0.782 | 0.74 | 4.40E-06 |
| JUN | 1.14E-08 | -0.124816751 | 0.099 | 0.171 | 5.68E-06 |
| STIM1 | 1.41E-08 | -0.099390657 | 0.072 | 0.13 | 7.02E-06 |
| LAT | 1.47E-08 | -0.159406778 | 0.197 | 0.273 | 7.34E-06 |
| TRAC | 2.01E-08 | 0.20505855 | 0.979 | 0.942 | 1.00E-05 |
| FAS | 3.01E-08 | 0.110286201 | 0.172 | 0.111 | 1.50E-05 |
| TNFRSF4 | 3.77E-08 | 0.238404711 | 0.416 | 0.34 | 1.88E-05 |
| CREM | 4.08E-08 | -0.187950606 | 0.727 | 0.773 | 2.03E-05 |
| RUNX3 | 4.52E-08 | 0.155066464 | 0.391 | 0.34 | 2.25E-05 |
| LTA | 5.35E-08 | -0.226528698 | 0.468 | 0.525 | 2.66E-05 |
| UFM1 | 6.81E-08 | 0.147786281 | 0.963 | 0.942 | 3.39E-05 |
| ICOS | 9.77E-08 | 0.286579215 | 0.846 | 0.786 | 4.87E-05 |
| IL32 | 2.29E-07 | 0.175197765 | 0.595 | 0.522 | 0.000114051 |
| ARPC1B | 2.51E-07 | 0.12575058 | 0.349 | 0.28 | 0.00012522 |
| IL1R1 | 3.69E-07 | 0.158693948 | 0.198 | 0.13 | 0.000183627 |
| RAPH1 | 4.76E-07 | -0.07198986 | 0.04 | 0.083 | 0.000237148 |
| TNFAIP8 | 4.77E-07 | -0.142635983 | 0.994 | 0.991 | 0.000237572 |
| NR4A2 | 2.05E-06 | -0.236546012 | 0.319 | 0.349 | 0.001020722 |
| IKZF1 | 2.23E-06 | 0.122854183 | 0.33 | 0.268 | 0.001110522 |

| | | | | | |
|---|---|---|---|---|---|
| PRDM1 | 2.33E-06 | -0.087931752 | 0.058 | 0.098 | 0.001161725 |
| CXCR5 | 3.35E-06 | 0.247595692 | 0.29 | 0.232 | 0.001669666 |
| CXCL10 | 3.63E-06 | 0.131793456 | 0.136 | 0.129 | 0.001805391 |
| IGLC3 | 4.29E-06 | -0.172496267 | 0.256 | 0.269 | 0.002134664 |
| BCL10 | 4.33E-06 | 0.072445285 | 0.135 | 0.089 | 0.00215586 |
| MALT1 | 4.62E-06 | 0.091543978 | 0.181 | 0.119 | 0.002302988 |
| TNFSF14 | 5.93E-06 | 0.123588972 | 0.255 | 0.196 | 0.002953008 |
| TNFRSF8 | 6.33E-06 | 0.048909808 | 0.038 | 0.018 | 0.003154818 |
| RBCK1 | 8.09E-06 | 0.096095975 | 0.24 | 0.188 | 0.004027302 |
| PMCH | 1.59E-05 | 0.066504616 | 0.007 | 0.003 | 0.0079059 |
| BTLA | 2.27E-05 | 0.130380195 | 0.292 | 0.228 | 0.011304402 |
| TRDC | 2.29E-05 | 0.127560591 | 0.033 | 0.022 | 0.011399654 |
| LIF | 3.00E-05 | 0.280514705 | 0.344 | 0.284 | 0.014922551 |
| NFKBIZ | 4.83E-05 | -0.174342938 | 0.659 | 0.649 | 0.024078086 |
| PDIA4 | 5.83E-05 | 0.135516208 | 0.566 | 0.495 | 0.02902158 |
| CD63 | 9.44E-05 | 0.108972374 | 0.683 | 0.637 | 0.04699247 |
| FOXP1 | 0.000109679 | -0.112759541 | 0.523 | 0.583 | 0.054620363 |
| CCR4 | 0.000119783 | -0.202847222 | 0.378 | 0.457 | 0.059652006 |
| IL2 | 0.000128204 | -0.271903546 | 1 | 1 | 0.063845425 |
| FYB1 | 0.000154746 | -0.122211089 | 0.239 | 0.29 | 0.07706367 |
| CXCL8 | 0.000159764 | 0.267599343 | 0.884 | 0.875 | 0.079562517 |
| FOSL1 | 0.000165726 | -0.106535405 | 0.786 | 0.829 | 0.082531755 |
| GIMAP2 | 0.000171688 | 0.042548667 | 0.068 | 0.041 | 0.08550084 |
| IL22 | 0.000174689 | -0.171035702 | 0.058 | 0.06 | 0.086995368 |
| CD3D | 0.000181557 | 0.130671298 | 0.945 | 0.91 | 0.090415193 |
| BATF | 0.000184524 | -0.122061647 | 0.617 | 0.668 | 0.091892836 |
| CCND2 | 0.000193797 | 0.144463792 | 0.702 | 0.673 | 0.096510908 |
| OSM | 0.000216194 | 0.047038536 | 0.056 | 0.034 | 0.107664576 |
| IFITM2 | 0.000238294 | 0.095915893 | 0.251 | 0.219 | 0.118670483 |
| NKG7 | 0.000248867 | 0.097329935 | 0.326 | 0.275 | 0.12393569 |
| ILF2 | 0.000258138 | 0.080189859 | 0.984 | 0.981 | 0.128552512 |
| TNFSF8 | 0.000303793 | 0.120499151 | 0.214 | 0.157 | 0.151289082 |
| STAT4 | 0.000318332 | -0.137192864 | 0.769 | 0.813 | 0.158529475 |
| MAP2K3 | 0.000362791 | 0.129510983 | 0.811 | 0.76 | 0.180670069 |
| LRRC8B | 0.000372433 | -0.103797037 | 0.556 | 0.613 | 0.185471715 |
| CSRNP1 | 0.000385896 | -0.098997582 | 0.929 | 0.938 | 0.192176334 |
| CD8B | 0.000403846 | 0.023621775 | 0.024 | 0.009 | 0.201115438 |
| SNX9 | 0.000452127 | 0.114111492 | 0.761 | 0.719 | 0.225159453 |
| HIF1A | 0.000623821 | 0.114626955 | 0.718 | 0.682 | 0.31066284 |
| JAK3 | 0.000663131 | 0.089649704 | 0.509 | 0.44 | 0.330239359 |
| NFKB1 | 0.000736821 | -0.0785007 | 1 | 1 | 0.366936609 |
| XBP1 | 0.000810458 | 0.096400957 | 0.63 | 0.592 | 0.403608159 |
| PIK3AP1 | 0.000837628 | 0.041741255 | 0.068 | 0.052 | 0.41713892 |
| EGR3 | 0.00091236 | -0.073935991 | 0.112 | 0.137 | 0.454355051 |
| ITGB2 | 0.000983229 | 0.073419632 | 0.197 | 0.162 | 0.489647948 |
| NFKBID | 0.001035758 | -0.043406795 | 0.045 | 0.067 | 0.515807455 |
| NINJ2 | 0.001174614 | 0.01134933 | 0.01 | 0.002 | 0.584957665 |

| | | | | | |
|---|---|---|---|---|---|
| IL2RA | 0.001385692 | 0.09740703 | 0.651 | 0.619 | 0.690074424 |
| TRAF1 | 0.001739023 | -0.098643034 | 0.735 | 0.794 | 0.866033367 |
| SOCS3 | 0.001776826 | 0.092271795 | 0.428 | 0.378 | 0.884859501 |
| PSMB8 | 0.001788398 | 0.090370681 | 0.771 | 0.756 | 0.890622028 |
| CXCL1 | 0.002000664 | -0.069779528 | 0.036 | 0.042 | 0.996330613 |
| NT5E | 0.002024104 | 0.12137067 | 0.319 | 0.291 | 1 |
| CTSW | 0.002187804 | 0.027547004 | 0.04 | 0.023 | 1 |
| IRF9 | 0.002251426 | -0.092325585 | 0.382 | 0.409 | 1 |
| PSMB9 | 0.002829002 | 0.085125845 | 0.488 | 0.451 | 1 |
| VPS28 | 0.002905258 | 0.070717827 | 0.39 | 0.345 | 1 |
| FOSB | 0.002960352 | -0.092491216 | 0.203 | 0.254 | 1 |
| ZBED2 | 0.00306959 | -0.051312725 | 0.024 | 0.03 | 1 |
| CD200 | 0.003243719 | -0.17414426 | 0.601 | 0.585 | 1 |
| TNFRSF13C | 0.003407023 | 0.088154801 | 0.305 | 0.262 | 1 |
| PRF1 | 0.00341515 | 0.083991631 | 0.447 | 0.414 | 1 |
| GZMA | 0.003535925 | 0.027469435 | 0.037 | 0.022 | 1 |
| DOCK8 | 0.003844663 | 0.06201862 | 0.188 | 0.157 | 1 |
| CCR7 | 0.004651344 | -0.105937471 | 0.383 | 0.414 | 1 |
| GATA3 | 0.004664911 | -0.028859481 | 0.019 | 0.033 | 1 |
| NFKBIA | 0.004889568 | 0.097686496 | 0.997 | 0.998 | 1 |
| EIF3J | 0.005711589 | 0.072171292 | 0.553 | 0.517 | 1 |
| NANS | 0.00627858 | 0.077913601 | 0.55 | 0.515 | 1 |
| EOMES | 0.007020626 | 0.013295487 | 0.009 | 0.004 | 1 |
| BAX | 0.007429879 | -0.071525946 | 0.397 | 0.439 | 1 |
| CD27 | 0.007582254 | -0.03960489 | 0.038 | 0.056 | 1 |
| TNIK | 0.007955403 | -0.044069114 | 0.072 | 0.092 | 1 |
| PSME3 | 0.008357205 | 0.069476345 | 0.615 | 0.574 | 1 |
| ATF4 | 0.008394349 | 0.063076813 | 0.997 | 0.997 | 1 |
| LCK | 0.008551088 | 0.04750732 | 0.127 | 0.105 | 1 |
| SPAG9 | 0.008846793 | -0.066984222 | 0.295 | 0.334 | 1 |
| LIG4 | 0.009202384 | 0.04311662 | 0.164 | 0.128 | 1 |
| IFNGR1 | 0.009913644 | 0.05532367 | 0.193 | 0.157 | 1 |
| B4GALT5 | 0.010020679 | 0.034211067 | 0.085 | 0.062 | 1 |
| SLC1A5 | 0.010457284 | 0.077598585 | 0.93 | 0.914 | 1 |
| BIRC3 | 0.010999621 | 0.122568013 | 0.717 | 0.693 | 1 |
| CD274 | 0.011062686 | 0.10863873 | 0.626 | 0.565 | 1 |
| TNFRSF25 | 0.01157005 | 0.072139164 | 0.567 | 0.56 | 1 |
| F5 | 0.012724525 | -0.127367064 | 0.499 | 0.485 | 1 |
| AIM2 | 0.013383221 | 0.015230564 | 0.014 | 0.005 | 1 |
| IL31 | 0.013968441 | -0.040039117 | 0.01 | 0.011 | 1 |
| CD7 | 0.01535088 | 0.075679118 | 0.616 | 0.587 | 1 |
| IL6 | 0.016055458 | -0.069042109 | 0.27 | 0.287 | 1 |
| CXCL16 | 0.0164252 | -0.033568955 | 0.05 | 0.068 | 1 |
| IL4R | 0.01650037 | -0.07251605 | 0.852 | 0.855 | 1 |
| ANXA5 | 0.01828413 | 0.069962038 | 0.459 | 0.435 | 1 |
| GZMK | 0.021458536 | 0.026918135 | 0.018 | 0.009 | 1 |
| AHI1 | 0.021844385 | -0.024225454 | 0.028 | 0.04 | 1 |

| | | | | | |
|---|---|---|---|---|---|
| CCL20 | 0.024741081 | 0.125822724 | 0.435 | 0.451 | 1 |
| CHI3L2 | 0.025039335 | 0.008591207 | 0.009 | 0.004 | 1 |
| IL23A | 0.028693726 | -0.095317082 | 0.643 | 0.663 | 1 |
| LRRC32 | 0.030761394 | 0.046202465 | 0.033 | 0.025 | 1 |
| DPP4 | 0.03189827 | 0.027322748 | 0.038 | 0.028 | 1 |
| ICAM1 | 0.034461843 | 0.059641943 | 0.413 | 0.387 | 1 |
| ERBIN | 0.035274479 | -0.048652758 | 0.204 | 0.224 | 1 |
| CD48 | 0.035863354 | 0.076099587 | 0.958 | 0.966 | 1 |
| HLA.DQB1 | 0.040895426 | -0.061089623 | 0.243 | 0.255 | 1 |
| KLRC3 | 0.042197243 | 0.02400612 | 0.059 | 0.042 | 1 |
| REL | 0.045476537 | -0.070782292 | 0.836 | 0.859 | 1 |
| PTPN7 | 0.046425041 | 0.052296234 | 0.384 | 0.348 | 1 |
| PTPRC | 0.047057945 | 0.067740217 | 0.867 | 0.868 | 1 |
| ARID5A | 0.052355248 | -0.057688507 | 0.483 | 0.489 | 1 |
| CD80 | 0.056715161 | 0.008973096 | 0.013 | 0.007 | 1 |
| FOXO1 | 0.057671146 | 0.035486609 | 0.162 | 0.136 | 1 |
| NDFIP2 | 0.0587237 | -0.047526056 | 0.341 | 0.375 | 1 |
| POU2F2 | 0.06328808 | -0.050802252 | 0.473 | 0.487 | 1 |
| ARHGEF3 | 0.067351096 | -0.052573129 | 0.208 | 0.223 | 1 |
| ITK | 0.071481478 | -0.05610111 | 0.965 | 0.968 | 1 |
| IRF8 | 0.071665356 | -0.082772173 | 0.961 | 0.951 | 1 |
| FTH1 | 0.072493275 | 0.069204688 | 0.928 | 0.935 | 1 |
| TIMM9 | 0.074667865 | 0.047092358 | 0.731 | 0.716 | 1 |
| TRIB1 | 0.076430463 | 0.055203188 | 0.227 | 0.194 | 1 |
| LTB | 0.081568477 | -0.010680882 | 0.009 | 0.016 | 1 |
| IGHD | 0.08326832 | -0.007833501 | 0.003 | 0.009 | 1 |
| EGR1 | 0.085269854 | -0.063928798 | 0.567 | 0.575 | 1 |
| CXCL2 | 0.08973613 | -0.056181463 | 0.3 | 0.328 | 1 |
| IL18RAP | 0.091104457 | 0.018195438 | 0.038 | 0.03 | 1 |
| TNFSF10 | 0.096731145 | -0.012993803 | 0.015 | 0.021 | 1 |
| SMAD2 | 0.105736598 | 0.030095665 | 0.169 | 0.153 | 1 |
| PRNP | 0.10959553 | -0.05624274 | 0.852 | 0.862 | 1 |
| GNLY | 0.109927768 | 0.041457049 | 0.248 | 0.229 | 1 |
| IL4 | 0.110046447 | -0.027363859 | 0.055 | 0.056 | 1 |
| ARL4C | 0.110920034 | 0.022860038 | 0.061 | 0.05 | 1 |
| IL15RA | 0.111334805 | 0.047298898 | 0.475 | 0.445 | 1 |
| IL1RN | 0.112256768 | 0.040577222 | 0.188 | 0.182 | 1 |
| EEF1E1 | 0.112258147 | 0.038640711 | 0.349 | 0.332 | 1 |
| TGFB3 | 0.113726491 | -0.005413274 | 0.001 | 0.004 | 1 |
| TGFB1 | 0.114623496 | 0.008284303 | 0.013 | 0.008 | 1 |
| HLA.DRA | 0.115719075 | 0.048745747 | 0.541 | 0.525 | 1 |
| ATF6B | 0.116226138 | 0.018525706 | 0.062 | 0.049 | 1 |
| ADGRE1 | 0.116815778 | -0.009735747 | 0.007 | 0.011 | 1 |
| LILRB4 | 0.118210482 | -0.01006994 | 0.011 | 0.017 | 1 |
| FOS | 0.118588959 | -0.069169756 | 0.49 | 0.467 | 1 |
| TRIB2 | 0.119960715 | 0.02972961 | 0.092 | 0.076 | 1 |
| BCL2L1 | 0.123483121 | -0.047033983 | 0.439 | 0.449 | 1 |

| | | | | | |
|---|---|---|---|---|---|
| DCLRE1C | 0.124833887 | -0.019315757 | 0.048 | 0.058 | 1 |
| MCM4 | 0.130641485 | 0.018647624 | 0.066 | 0.053 | 1 |
| LGALS9 | 0.133087268 | 0.013583814 | 0.036 | 0.028 | 1 |
| FUT4 | 0.133433399 | 0.010103361 | 0.023 | 0.017 | 1 |
| BCL2 | 0.141215173 | -0.032736342 | 0.204 | 0.227 | 1 |
| TNIP3 | 0.143376235 | 0.009446827 | 0.016 | 0.01 | 1 |
| IL3 | 0.145508476 | -0.030193512 | 0.094 | 0.104 | 1 |
| CD40 | 0.145818264 | -0.031304205 | 0.193 | 0.208 | 1 |
| JCHAIN | 0.146003504 | -0.023541986 | 0.045 | 0.051 | 1 |
| PDCD1 | 0.153619561 | 0.018529505 | 0.046 | 0.036 | 1 |
| CD86 | 0.153638254 | -0.01392197 | 0.03 | 0.04 | 1 |
| VEGFA | 0.154425324 | -0.006549393 | 0.004 | 0.008 | 1 |
| IGBP1 | 0.155447128 | 0.022864414 | 0.128 | 0.112 | 1 |
| CCL22 | 0.155847219 | 0.031188638 | 0.172 | 0.149 | 1 |
| CD9 | 0.156520935 | 0.020015311 | 0.081 | 0.074 | 1 |
| SNX8 | 0.156722412 | -0.035342766 | 0.317 | 0.336 | 1 |
| LIPA | 0.158563442 | 0.024315601 | 0.13 | 0.122 | 1 |
| IFI6 | 0.166074606 | -0.010291041 | 0.014 | 0.021 | 1 |
| SDCBP | 0.174482833 | 0.046536265 | 0.755 | 0.704 | 1 |
| ADA | 0.180445499 | -0.019940584 | 0.056 | 0.063 | 1 |
| IL12RB1 | 0.182832704 | -0.003839139 | 0.001 | 0.004 | 1 |
| NFATC3 | 0.185242138 | 0.024173576 | 0.173 | 0.156 | 1 |
| ZNF341 | 0.189053785 | 0.007398324 | 0.017 | 0.012 | 1 |
| THEMIS | 0.19066913 | -0.013563575 | 0.025 | 0.027 | 1 |
| VPREB3 | 0.191797684 | -0.004232834 | 0.001 | 0.003 | 1 |
| CXCL5 | 0.192271966 | -0.037294655 | 0.234 | 0.242 | 1 |
| NFKB2 | 0.193084127 | -0.032878406 | 0.268 | 0.289 | 1 |
| HLA.DPA1 | 0.194438517 | 0.027946185 | 0.222 | 0.2 | 1 |
| CASP5 | 0.202469003 | 0.012207199 | 0.042 | 0.034 | 1 |
| GRB10 | 0.208924136 | -0.014736539 | 0.038 | 0.048 | 1 |
| CYCS | 0.209996442 | -0.029174309 | 0.979 | 0.987 | 1 |
| STAT1 | 0.215069632 | -0.040861773 | 0.772 | 0.77 | 1 |
| STAT6 | 0.218380992 | 0.033425071 | 0.441 | 0.407 | 1 |
| CD14 | 0.219123389 | -0.011390369 | 0.014 | 0.021 | 1 |
| SOCS2 | 0.22044689 | -0.038094659 | 0.172 | 0.17 | 1 |
| LAT2 | 0.22277814 | -0.008188118 | 0.013 | 0.018 | 1 |
| IL12RB2 | 0.223430908 | 0.043021709 | 0.223 | 0.214 | 1 |
| TCF7 | 0.226311704 | 0.016219242 | 0.073 | 0.063 | 1 |
| ENTPD1 | 0.228651606 | 0.002778934 | 0.004 | 0.002 | 1 |
| CD72 | 0.229361011 | 0.003868956 | 0.006 | 0.004 | 1 |
| LYN | 0.237623093 | 0.013510158 | 0.059 | 0.048 | 1 |
| KLRC1 | 0.2470444 | 0.011354057 | 0.041 | 0.033 | 1 |
| MAST4 | 0.248923727 | -0.031751053 | 0.229 | 0.251 | 1 |
| IL5 | 0.253022733 | -0.011570487 | 0.007 | 0.006 | 1 |
| CR2 | 0.254998289 | 0.009420254 | 0.021 | 0.015 | 1 |
| CTLA4 | 0.255353805 | -0.113677477 | 0.443 | 0.42 | 1 |
| CD160 | 0.256063438 | 0.003374843 | 0.005 | 0.003 | 1 |

| | | | | | |
|---|---|---|---|---|---|
| PAXBP1 | 0.258974546 | -0.027931194 | 0.335 | 0.354 | 1 |
| CCL13 | 0.260587192 | -0.003051426 | 0.001 | 0.003 | 1 |
| CNOT2 | 0.26460992 | 0.026714785 | 0.416 | 0.402 | 1 |
| PLCG2 | 0.265353589 | 0.005249824 | 0.011 | 0.008 | 1 |
| FCGR3A | 0.27221475 | 0.008915298 | 0.028 | 0.024 | 1 |
| PSEN1 | 0.279246561 | -0.011149794 | 0.038 | 0.045 | 1 |
| PCNA | 0.281816686 | -0.027080061 | 0.348 | 0.36 | 1 |
| TUBA1C | 0.28529441 | 0.030140785 | 0.969 | 0.962 | 1 |
| CXCR4 | 0.28588605 | 0.030785818 | 0.151 | 0.138 | 1 |
| AQP9 | 0.294263462 | -0.00590049 | 0.01 | 0.014 | 1 |
| PTGDR2 | 0.299097232 | -0.003625755 | 0.003 | 0.005 | 1 |
| GAB2 | 0.305103348 | 0.01185192 | 0.051 | 0.042 | 1 |
| TIAF1 | 0.305947515 | -0.005618106 | 0.009 | 0.012 | 1 |
| PIK3IP1 | 0.310731097 | 0.017828552 | 0.099 | 0.095 | 1 |
| BIN2 | 0.3110808 | -0.004482993 | 0.005 | 0.008 | 1 |
| CXCL9 | 0.314792781 | 0.01471841 | 0.052 | 0.053 | 1 |
| CXCL11 | 0.317550085 | 0.006520624 | 0.011 | 0.011 | 1 |
| CD247 | 0.31776166 | 0.010828248 | 0.05 | 0.042 | 1 |
| LAMP1 | 0.317843611 | 0.0265872 | 0.513 | 0.48 | 1 |
| IL11RA | 0.319942075 | 0.011653152 | 0.053 | 0.044 | 1 |
| RORC | 0.323063823 | -0.006679693 | 0.004 | 0.013 | 1 |
| CD300A | 0.327214401 | -0.006808606 | 0.014 | 0.019 | 1 |
| KLRC4 | 0.327674815 | 0.002980933 | 0.005 | 0.003 | 1 |
| FBXO11 | 0.33037663 | -0.022503514 | 0.225 | 0.235 | 1 |
| CD6 | 0.333087812 | -0.032058143 | 0.657 | 0.642 | 1 |
| SKAP1 | 0.342323338 | -0.005967217 | 0.013 | 0.016 | 1 |
| TGFBI | 0.342785741 | 0.013071647 | 0.071 | 0.067 | 1 |
| DUSP4 | 0.34290761 | 0.042420808 | 0.409 | 0.378 | 1 |
| IL23R | 0.348003251 | -0.003625755 | 0.003 | 0.004 | 1 |
| LAMP3 | 0.349447532 | -0.033209015 | 0.396 | 0.394 | 1 |
| GEM | 0.356078183 | 0.056117007 | 0.743 | 0.675 | 1 |
| B3GAT1 | 0.373296693 | 0.004564874 | 0.013 | 0.009 | 1 |
| TRAT1 | 0.37551761 | -0.013208699 | 0.056 | 0.068 | 1 |
| CD5 | 0.376278775 | 0.02568504 | 0.268 | 0.261 | 1 |
| CHD1 | 0.379376047 | -0.007552825 | 0.024 | 0.028 | 1 |
| IL6R | 0.381162554 | 0.012950827 | 0.088 | 0.084 | 1 |
| CYBB | 0.382146442 | 0.009590642 | 0.053 | 0.047 | 1 |
| IGHG3 | 0.383269498 | -0.003126499 | 0.004 | 0.006 | 1 |
| LGALS1 | 0.389743037 | -0.022933072 | 0.253 | 0.248 | 1 |
| WAS | 0.392345707 | 0.010385943 | 0.064 | 0.055 | 1 |
| SNCA | 0.394634338 | 0.001391277 | 0.002 | 0.001 | 1 |
| VSIR | 0.401531943 | -0.037354596 | 0.707 | 0.705 | 1 |
| CD52 | 0.405703023 | 0.026303512 | 0.48 | 0.442 | 1 |
| CCR5 | 0.40825535 | 0.007362025 | 0.038 | 0.032 | 1 |
| TMEM97 | 0.408871898 | 0.013224391 | 0.114 | 0.107 | 1 |
| ZAP70 | 0.408896982 | -0.012781265 | 0.088 | 0.093 | 1 |
| SLC7A7 | 0.412519829 | -0.003699132 | 0.005 | 0.007 | 1 |

| | | | | | |
|---|---|---|---|---|---|
| VMO1 | 0.415320471 | 0.007468658 | 0.03 | 0.023 | 1 |
| DUSP5 | 0.425104049 | -0.031196877 | 0.767 | 0.733 | 1 |
| DUSP1 | 0.427469614 | -0.022082278 | 0.34 | 0.358 | 1 |
| IFRD1 | 0.431913371 | 0.018059027 | 0.256 | 0.252 | 1 |
| RNASE6 | 0.434771059 | -0.002445958 | 0.003 | 0.004 | 1 |
| RPL6 | 0.435242206 | -0.013133354 | 1 | 1 | 1 |
| ITGAM | 0.435654358 | 0.003582812 | 0.011 | 0.008 | 1 |
| LGALS3 | 0.437445176 | 0.008928986 | 0.053 | 0.046 | 1 |
| LAP3 | 0.438733015 | -0.019285154 | 0.331 | 0.342 | 1 |
| CD28 | 0.443785915 | 0.013665629 | 0.116 | 0.109 | 1 |
| ZBTB16 | 0.447119106 | -0.00215779 | 0.002 | 0.003 | 1 |
| IL18R1 | 0.447977946 | -0.009645685 | 0.046 | 0.049 | 1 |
| POU2AF1 | 0.449292376 | -0.016145102 | 0.103 | 0.114 | 1 |
| SPP1 | 0.467360333 | 0.004079711 | 0.015 | 0.012 | 1 |
| IL3RA | 0.469415156 | 0.007623315 | 0.049 | 0.044 | 1 |
| PGM3 | 0.469545109 | -0.007690016 | 0.032 | 0.037 | 1 |
| NCAM1 | 0.475207108 | -0.00147471 | 0.001 | 0.002 | 1 |
| CD163 | 0.483626913 | -0.003201167 | 0.006 | 0.008 | 1 |
| JAK2 | 0.487858497 | -0.007599119 | 0.04 | 0.045 | 1 |
| CD44 | 0.48850806 | -0.017119476 | 1 | 1 | 1 |
| CHD7 | 0.492750072 | 0.011940584 | 0.134 | 0.125 | 1 |
| TCL1A | 0.499719085 | -0.002052477 | 0.003 | 0.004 | 1 |
| NFATC2 | 0.49977216 | -0.009287417 | 0.064 | 0.068 | 1 |
| CCL2 | 0.509165516 | 0.026736823 | 0.553 | 0.555 | 1 |
| MS4A1 | 0.51418086 | 0.005545834 | 0.031 | 0.027 | 1 |
| GZMH | 0.518752351 | -0.006783333 | 0.036 | 0.044 | 1 |
| FOXP3 | 0.519209725 | 0.011100689 | 0.018 | 0.014 | 1 |
| CBLB | 0.523591343 | 0.008504497 | 0.065 | 0.061 | 1 |
| HLA.DMA | 0.523962622 | -0.006510366 | 0.038 | 0.043 | 1 |
| CD3G | 0.525871681 | 0.005352187 | 0.024 | 0.02 | 1 |
| GNAI2 | 0.52990326 | -0.019415619 | 0.447 | 0.441 | 1 |
| IFITM3 | 0.53562826 | 0.009902892 | 0.109 | 0.097 | 1 |
| CD36 | 0.543165439 | -0.004797034 | 0.022 | 0.024 | 1 |
| MCM2 | 0.550902474 | -0.01046442 | 0.145 | 0.149 | 1 |
| CCR8 | 0.55135795 | -0.001869051 | 0.001 | 0.001 | 1 |
| SELL | 0.556676869 | -0.031874236 | 0.651 | 0.673 | 1 |
| KDELR1 | 0.559366602 | 0.010032635 | 0.137 | 0.13 | 1 |
| TGFBR1 | 0.56151832 | 0.008012819 | 0.08 | 0.076 | 1 |
| CD22 | 0.562943304 | 0.006490753 | 0.049 | 0.042 | 1 |
| MMP9 | 0.563748119 | 0.007583693 | 0.07 | 0.059 | 1 |
| CD33 | 0.572757904 | -0.001080262 | 0.001 | 0.002 | 1 |
| S100A9 | 0.57424972 | -0.006067642 | 0.035 | 0.036 | 1 |
| HAVCR2 | 0.577514037 | 0.006487877 | 0.046 | 0.047 | 1 |
| TREM1 | 0.590041924 | -0.001947344 | 0.004 | 0.004 | 1 |
| S100A12 | 0.592976135 | 0.001901555 | 0.006 | 0.005 | 1 |
| PAK1IP1 | 0.597174147 | -0.01465617 | 0.5 | 0.479 | 1 |
| CD4 | 0.606766708 | 0.003008991 | 0.016 | 0.014 | 1 |

| | | | | | |
|---|---|---|---|---|---|
| THBS1 | 0.60942097 | 0.007885608 | 0.038 | 0.041 | 1 |
| IL12A | 0.615632877 | 0.003998038 | 0.022 | 0.02 | 1 |
| KLRG1 | 0.621035821 | -0.002312249 | 0.007 | 0.008 | 1 |
| SP140 | 0.623916723 | -0.004861571 | 0.034 | 0.034 | 1 |
| BCL2A1 | 0.629443321 | -0.011865462 | 1 | 0.999 | 1 |
| IGHM | 0.638009441 | 0.003998038 | 0.023 | 0.018 | 1 |
| FCER2 | 0.638600861 | 0.003905935 | 0.027 | 0.022 | 1 |
| CCR1 | 0.644549617 | 0.007069972 | 0.103 | 0.096 | 1 |
| MITF | 0.647499121 | -0.001554079 | 0.004 | 0.004 | 1 |
| CARD11 | 0.647499122 | -0.001554079 | 0.003 | 0.005 | 1 |
| CXCL3 | 0.670603845 | 0.040575704 | 0.177 | 0.15 | 1 |
| IL13 | 0.671802541 | -0.019244046 | 0.086 | 0.072 | 1 |
| CD38 | 0.676728681 | -0.003888485 | 0.024 | 0.025 | 1 |
| CD70 | 0.677346333 | 0.002918746 | 0.015 | 0.016 | 1 |
| IGHG2 | 0.680873748 | -0.000975738 | 0.002 | 0.002 | 1 |
| CD79B | 0.684578013 | 0.001304165 | 0.004 | 0.003 | 1 |
| TSPAN32 | 0.685457646 | -0.001920479 | 0.007 | 0.008 | 1 |
| MYB | 0.691511701 | 0.011634183 | 0.167 | 0.166 | 1 |
| IL9 | 0.69358446 | 0.00583163 | 0.019 | 0.03 | 1 |
| IKZF2 | 0.696779065 | -0.000685706 | 0.001 | 0.001 | 1 |
| JUNB | 0.697322249 | 0.01081007 | 0.437 | 0.424 | 1 |
| CD2 | 0.732691877 | -0.009376846 | 0.991 | 0.986 | 1 |
| IGHG1 | 0.744420662 | 0.010315155 | 0.093 | 0.099 | 1 |
| RORA | 0.74777157 | 0.012742971 | 0.182 | 0.178 | 1 |
| TOP2A | 0.750294624 | 0.00060184 | 0.002 | 0.001 | 1 |
| CCR6 | 0.750294624 | 0.00060184 | 0.002 | 0.001 | 1 |
| IL1R2 | 0.751108244 | -0.000975738 | 0.002 | 0.002 | 1 |
| IL15 | 0.75229967 | -0.000871393 | 0.003 | 0.003 | 1 |
| CTSD | 0.760069861 | -0.004108886 | 0.058 | 0.057 | 1 |
| CXCL13 | 0.765720941 | 0.000704893 | 0.003 | 0.002 | 1 |
| IRF3 | 0.776652162 | -0.001503305 | 0.011 | 0.012 | 1 |
| TNFSF13B | 0.78275877 | 0.001725802 | 0.01 | 0.008 | 1 |
| MGST1 | 0.786573727 | -0.001320298 | 0.008 | 0.01 | 1 |
| FCER1G | 0.788331388 | -0.005362678 | 0.134 | 0.141 | 1 |
| ITGAX | 0.791237374 | -0.004508546 | 0.092 | 0.095 | 1 |
| QPCT | 0.792055612 | -0.001136618 | 0.007 | 0.008 | 1 |
| BCL6 | 0.796126315 | 0.001334774 | 0.012 | 0.01 | 1 |
| IGHA1 | 0.796643639 | -0.000581505 | 0.002 | 0.002 | 1 |
| CLEC4E | 0.796643639 | -0.000581505 | 0.002 | 0.002 | 1 |
| LY86 | 0.804367624 | 0.001233145 | 0.011 | 0.01 | 1 |
| FN1 | 0.818360374 | 0.004343008 | 0.121 | 0.114 | 1 |
| IL21 | 0.81881843 | -0.012470824 | 0.09 | 0.094 | 1 |
| MYC | 0.847676708 | -0.006417462 | 0.985 | 0.983 | 1 |
| SLC25A37 | 0.851255083 | -0.004875267 | 0.276 | 0.286 | 1 |
| CLEC10A | 0.855412701 | -0.000291041 | 0.001 | 0.001 | 1 |
| ZNF683 | 0.855412701 | -0.000291041 | 0.001 | 0.001 | 1 |
| TCF4 | 0.855412701 | -0.000291041 | 0.001 | 0.001 | 1 |

| | | | | | |
|---|---|---|---|---|---|
| IL1B | 0.858794861 | 0.006821206 | 0.392 | 0.4 | 1 |
| NGDN | 0.859087506 | 0.001956868 | 0.041 | 0.043 | 1 |
| IRF7 | 0.862714834 | 0.00383454 | 0.172 | 0.152 | 1 |
| CX3CR1 | 0.864296754 | -0.001403876 | 0.023 | 0.024 | 1 |
| EBF1 | 0.865818247 | 0.001178667 | 0.02 | 0.019 | 1 |
| TYMS | 0.870310246 | -0.000663237 | 0.004 | 0.004 | 1 |
| TNFRSF9 | 0.872883723 | 0.001523346 | 0.031 | 0.029 | 1 |
| TLR2 | 0.890176956 | 0.001481564 | 0.046 | 0.042 | 1 |
| FCN1 | 0.897831794 | 0.000757048 | 0.013 | 0.012 | 1 |
| CD79A | 0.8984427 | 0.000892217 | 0.021 | 0.021 | 1 |
| SOCS1 | 0.899789712 | 0.00031066 | 0.003 | 0.002 | 1 |
| TRBC2 | 0.903258927 | -0.004917864 | 0.954 | 0.966 | 1 |
| BACH2 | 0.915784975 | 0.002819819 | 0.332 | 0.334 | 1 |
| CD37 | 0.921731738 | 0.002467387 | 0.418 | 0.402 | 1 |
| IER3 | 0.93013009 | -0.0049952 | 0.945 | 0.941 | 1 |
| BLNK | 0.930234793 | -0.000187164 | 0.002 | 0.002 | 1 |
| GAPDH | 0.934154344 | -0.002756187 | 0.947 | 0.949 | 1 |
| CD24 | 0.941006357 | -0.000269971 | 0.003 | 0.005 | 1 |
| VSIG4 | 0.942041678 | 0.000103731 | 0.001 | 0.001 | 1 |
| NRP1 | 0.943788976 | 0.000675661 | 0.034 | 0.034 | 1 |
| ITGAE | 0.945273053 | 0.000709619 | 0.038 | 0.041 | 1 |
| ABCE1 | 0.951173781 | 0.001485268 | 0.904 | 0.911 | 1 |
| SELPLG | 0.955287481 | 0.000726424 | 0.04 | 0.035 | 1 |
| IGKC | 0.960363955 | 0.001952344 | 0.164 | 0.171 | 1 |
| MZB1 | 0.961646192 | -0.00024905 | 0.008 | 0.007 | 1 |
| IL1A | 0.979735954 | -0.000306184 | 0.039 | 0.041 | 1 |
| CD1C | 0.987417735 | -6.32E-05 | 0.005 | 0.006 | 1 |
| VNN2 | 0.993202779 | -0.00552153 | 0 | 0.004 | 1 |
| THBD | 0.993468375 | -0.002763406 | 0 | 0.002 | 1 |
| KLRF1 | 0.994542978 | 2.01E-05 | 0.004 | 0.004 | 1 |
| CD209 | 0.995841756 | -0.002368958 | 0 | 0.001 | 1 |
| TSLP | 0.995935991 | -0.001579737 | 0 | 0.001 | 1 |
| CXCR6 | 0.995935991 | -0.001579737 | 0 | 0.001 | 1 |

CD4+ Memory

| | p_val | avg_log2FC | Patient | Controls | p_val_adj |
|---|---|---|---|---|---|
| CD40LG | 0 | -1.384938948 | 0.183 | 0.598 | 0 |
| PFN1 | 1.44E-307 | 0.915837322 | 1 | 0.993 | 7.18E-305 |
| BTG1 | 2.00E-287 | -1.099372985 | 0.991 | 0.997 | 9.97E-285 |
| NFKBIZ | 3.76E-276 | -1.768792992 | 0.664 | 0.772 | 1.87E-273 |
| NR4A2 | 2.53E-263 | -2.311336207 | 0.416 | 0.702 | 1.26E-260 |
| DUSP2 | 6.10E-236 | -1.29004099 | 0.949 | 0.983 | 3.04E-233 |
| CREM | 1.40E-235 | -1.356566242 | 0.57 | 0.779 | 6.99E-233 |
| FOS | 6.20E-233 | -2.381676963 | 0.52 | 0.67 | 3.09E-230 |
| SDCBP | 1.57E-211 | -1.118249008 | 0.752 | 0.862 | 7.81E-209 |
| EGR1 | 4.46E-198 | -1.521070415 | 0.585 | 0.687 | 2.22E-195 |

| | | | | | |
|---|---|---|---|---|---|
| REL | 4.38E-192 | -0.924573005 | 0.829 | 0.933 | 2.18E-189 |
| TNFRSF4 | 7.16E-184 | 1.205484641 | 0.778 | 0.442 | 3.57E-181 |
| CSRNP1 | 2.34E-167 | -0.819229313 | 0.865 | 0.935 | 1.17E-164 |
| TRAC | 6.28E-166 | 0.841756226 | 0.975 | 0.9 | 3.13E-163 |
| TNF | 9.09E-162 | -1.627743025 | 0.947 | 0.964 | 4.53E-159 |
| FOSB | 9.78E-159 | -1.690505032 | 0.212 | 0.554 | 4.87E-156 |
| CD3D | 5.48E-154 | 0.788637379 | 0.965 | 0.85 | 2.73E-151 |
| TGIF1 | 5.26E-153 | -0.802460889 | 0.471 | 0.711 | 2.62E-150 |
| EGR3 | 5.11E-151 | -0.997201799 | 0.125 | 0.395 | 2.54E-148 |
| SLA | 3.40E-150 | -1.071830838 | 0.49 | 0.717 | 1.69E-147 |
| MYC | 2.78E-147 | -0.911636552 | 0.882 | 0.949 | 1.38E-144 |
| IL2RG | 5.56E-145 | 0.60178634 | 0.956 | 0.87 | 2.77E-142 |
| FYN | 2.49E-140 | -0.675907917 | 0.808 | 0.904 | 1.24E-137 |
| IL2RB | 2.71E-133 | 0.773482872 | 0.858 | 0.619 | 1.35E-130 |
| NFKBIA | 3.32E-133 | -0.77276789 | 0.995 | 1 | 1.65E-130 |
| IL6ST | 3.21E-127 | -0.688916718 | 0.314 | 0.565 | 1.60E-124 |
| S100A10 | 3.65E-121 | 0.939175854 | 0.763 | 0.638 | 1.82E-118 |
| FOSL1 | 8.61E-119 | -0.62067587 | 0.597 | 0.817 | 4.29E-116 |
| IFNG | 3.79E-117 | -1.52468296 | 0.991 | 0.991 | 1.89E-114 |
| TIGIT | 5.84E-112 | 0.597413058 | 0.345 | 0.088 | 2.91E-109 |
| JUN | 8.71E-112 | -0.665679405 | 0.117 | 0.356 | 4.34E-109 |
| BCL2A1 | 8.22E-109 | -0.596856216 | 0.99 | 0.992 | 4.09E-106 |
| FASLG | 6.29E-108 | -1.015077251 | 0.23 | 0.463 | 3.13E-105 |
| CSF2 | 2.32E-106 | -1.577727745 | 0.898 | 0.906 | 1.15E-103 |
| PRNP | 1.95E-105 | -0.630555657 | 0.732 | 0.832 | 9.71E-103 |
| BCAT1 | 5.09E-103 | 0.499596293 | 0.302 | 0.081 | 2.53E-100 |
| NFKB1 | 4.97E-102 | -0.518301461 | 0.996 | 0.998 | 2.47E-99 |
| CD52 | 3.36E-100 | 0.682615593 | 0.679 | 0.469 | 1.67E-97 |
| EIF5 | 2.08E-99 | -0.56561558 | 0.821 | 0.899 | 1.03E-96 |
| MAP2K3 | 2.25E-96 | 0.582055942 | 0.949 | 0.835 | 1.12E-93 |
| EVI2A | 1.09E-90 | -0.654855799 | 0.358 | 0.533 | 5.42E-88 |
| GBP1 | 1.48E-89 | 0.863511786 | 0.607 | 0.478 | 7.36E-87 |
| SPAG9 | 2.87E-84 | -0.558489528 | 0.342 | 0.524 | 1.43E-81 |
| DUSP1 | 1.17E-81 | -0.768439966 | 0.351 | 0.5 | 5.85E-79 |
| LTA | 1.44E-78 | -1.251381788 | 0.517 | 0.634 | 7.17E-76 |
| CST7 | 1.05E-76 | 0.609447859 | 0.513 | 0.305 | 5.21E-74 |
| EVI2B | 1.14E-76 | -0.58969606 | 0.275 | 0.451 | 5.69E-74 |
| FBXO11 | 7.23E-73 | -0.432430303 | 0.219 | 0.404 | 3.60E-70 |
| APOBEC3G | 1.07E-71 | 0.777296055 | 0.66 | 0.467 | 5.34E-69 |
| ITGA4 | 1.31E-70 | 0.356676459 | 0.245 | 0.085 | 6.53E-68 |
| CD83 | 4.05E-70 | -0.707579124 | 0.667 | 0.806 | 2.01E-67 |
| IL7R | 2.37E-68 | -0.548189832 | 0.399 | 0.58 | 1.18E-65 |
| CD226 | 6.86E-65 | 0.381884522 | 0.329 | 0.139 | 3.42E-62 |
| TRIB2 | 4.19E-64 | 0.387942246 | 0.193 | 0.058 | 2.09E-61 |
| NANS | 7.20E-63 | -0.480292055 | 0.429 | 0.561 | 3.58E-60 |
| PDIA6 | 5.81E-61 | 0.522489673 | 0.877 | 0.656 | 2.89E-58 |
| IL23A | 2.15E-57 | -0.630607827 | 0.737 | 0.8 | 1.07E-54 |

| | | | | | |
|---|---|---|---|---|---|
| JUNB | 4.01E-57 | -0.51995379 | 0.453 | 0.542 | 1.99E-54 |
| IFRD1 | 2.04E-53 | -0.406381587 | 0.288 | 0.423 | 1.01E-50 |
| GIMAP5 | 2.17E-52 | 0.421635103 | 0.303 | 0.147 | 1.08E-49 |
| NFKBID | 1.49E-51 | -0.250593347 | 0.062 | 0.208 | 7.41E-49 |
| JAK1 | 1.93E-51 | 0.407237596 | 0.81 | 0.698 | 9.63E-49 |
| TRBC2 | 2.23E-51 | 0.496570909 | 0.979 | 0.956 | 1.11E-48 |
| IL3 | 2.68E-51 | -2.019475861 | 0.108 | 0.121 | 1.33E-48 |
| TNFRSF25 | 3.48E-50 | 0.355225553 | 0.573 | 0.391 | 1.73E-47 |
| MAST4 | 5.07E-50 | -0.406549467 | 0.194 | 0.375 | 2.52E-47 |
| ARPC1B | 9.09E-49 | 0.329726712 | 0.393 | 0.227 | 4.53E-46 |
| CD5 | 3.66E-48 | 0.50019587 | 0.605 | 0.412 | 1.82E-45 |
| PTPN7 | 6.10E-47 | 0.33037195 | 0.481 | 0.303 | 3.04E-44 |
| CD2 | 1.98E-46 | 0.409525879 | 0.978 | 0.907 | 9.86E-44 |
| CD74 | 2.73E-44 | 0.478252852 | 0.711 | 0.63 | 1.36E-41 |
| ARHGEF3 | 4.92E-40 | -0.356204138 | 0.255 | 0.395 | 2.45E-37 |
| CCL4 | 2.32E-38 | -0.567134961 | 1 | 1 | 1.16E-35 |
| DUSP4 | 5.00E-38 | 0.547778671 | 0.786 | 0.575 | 2.49E-35 |
| CXCR3 | 1.66E-37 | 0.588582835 | 0.521 | 0.312 | 8.29E-35 |
| STAT5A | 1.30E-35 | -0.325576289 | 0.957 | 0.935 | 6.50E-33 |
| IL13 | 7.38E-35 | -1.730940614 | 0.102 | 0.134 | 3.68E-32 |
| CD69 | 1.08E-34 | -0.384530564 | 1 | 0.999 | 5.39E-32 |
| IL4 | 1.40E-34 | -1.450077424 | 0.069 | 0.108 | 6.98E-32 |
| DUSP5 | 3.77E-34 | -0.393669138 | 0.748 | 0.799 | 1.88E-31 |
| FTH1 | 1.37E-32 | -0.425755095 | 0.931 | 0.942 | 6.81E-30 |
| CCR7 | 1.42E-32 | -0.452984219 | 0.455 | 0.53 | 7.08E-30 |
| IRF7 | 7.98E-32 | -0.290941272 | 0.172 | 0.293 | 3.97E-29 |
| ZBED2 | 1.64E-31 | -0.670736909 | 0.05 | 0.111 | 8.16E-29 |
| IVNS1ABP | 1.85E-31 | -0.317342757 | 0.618 | 0.709 | 9.23E-29 |
| HNRNPC | 5.84E-31 | -0.277778686 | 0.841 | 0.888 | 2.91E-28 |
| F5 | 3.80E-30 | 0.470548521 | 0.483 | 0.285 | 1.89E-27 |
| EIF5A | 6.67E-30 | 0.24384537 | 0.994 | 0.965 | 3.32E-27 |
| CSF1 | 2.78E-29 | 0.584802908 | 0.432 | 0.323 | 1.38E-26 |
| IL12RB2 | 6.06E-29 | 0.339225833 | 0.279 | 0.156 | 3.02E-26 |
| JAK3 | 1.02E-28 | 0.247604062 | 0.459 | 0.325 | 5.08E-26 |
| VSIR | 2.34E-28 | 0.381197168 | 0.836 | 0.761 | 1.16E-25 |
| CD3E | 5.71E-28 | 0.318347849 | 0.851 | 0.709 | 2.84E-25 |
| IRF1 | 1.54E-27 | -0.394450036 | 0.777 | 0.827 | 7.69E-25 |
| CXCR4 | 1.70E-27 | -0.537221575 | 0.21 | 0.31 | 8.45E-25 |
| PDIA4 | 4.22E-27 | 0.279990674 | 0.532 | 0.389 | 2.10E-24 |
| GEM | 9.45E-27 | -0.612334026 | 0.766 | 0.714 | 4.70E-24 |
| CD6 | 1.03E-26 | 0.303738528 | 0.811 | 0.717 | 5.12E-24 |
| DUSP6 | 5.16E-26 | -0.399127638 | 0.307 | 0.383 | 2.57E-23 |
| NFATC1 | 5.80E-26 | -0.253875117 | 0.198 | 0.295 | 2.89E-23 |
| HIF1A | 1.32E-25 | -0.304777417 | 0.561 | 0.66 | 6.59E-23 |
| CXCL8 | 1.94E-25 | -0.491111354 | 0.872 | 0.889 | 9.68E-23 |
| BACH2 | 6.74E-25 | -0.222303666 | 0.197 | 0.311 | 3.36E-22 |
| LCK | 1.76E-24 | 0.201379455 | 0.22 | 0.131 | 8.79E-22 |

| | | | | | |
|---|---|---|---|---|---|
| BIRC3 | 3.69E-24 | -0.392716261 | 0.742 | 0.821 | 1.84E-21 |
| ARID5A | 3.74E-24 | -0.257403004 | 0.303 | 0.4 | 1.86E-21 |
| RPL6 | 4.96E-24 | 0.187980402 | 1 | 0.999 | 2.47E-21 |
| ATF4 | 7.58E-24 | -0.21561872 | 0.994 | 0.993 | 3.78E-21 |
| CD38 | 1.20E-23 | 0.104977314 | 0.063 | 0.013 | 5.98E-21 |
| GBP2 | 5.53E-23 | 0.286692029 | 0.842 | 0.756 | 2.76E-20 |
| TNFAIP8 | 6.36E-22 | 0.248674965 | 0.993 | 0.958 | 3.17E-19 |
| IL32 | 2.14E-21 | 0.269425814 | 0.516 | 0.374 | 1.07E-18 |
| TNFRSF8 | 3.04E-21 | 0.107454296 | 0.084 | 0.02 | 1.51E-18 |
| IL22 | 6.72E-21 | -1.977085191 | 0.061 | 0.092 | 3.35E-18 |
| LRRC8B | 9.60E-21 | 0.235694991 | 0.649 | 0.544 | 4.78E-18 |
| IL31 | 1.25E-20 | -0.542393944 | 0.011 | 0.023 | 6.21E-18 |
| TRAF1 | 1.59E-20 | -0.247846394 | 0.697 | 0.762 | 7.92E-18 |
| RPN2 | 1.81E-20 | 0.217396203 | 0.638 | 0.514 | 9.03E-18 |
| VPS28 | 2.84E-20 | 0.168745794 | 0.344 | 0.23 | 1.42E-17 |
| STAT4 | 5.15E-20 | -0.296729036 | 0.534 | 0.678 | 2.57E-17 |
| IER3 | 2.48E-19 | 0.437444643 | 0.956 | 0.924 | 1.24E-16 |
| CD7 | 3.81E-19 | 0.225851068 | 0.448 | 0.331 | 1.90E-16 |
| NINJ2 | 6.27E-19 | 0.074954781 | 0.047 | 0.006 | 3.12E-16 |
| SOCS3 | 4.31E-18 | -0.203917996 | 0.255 | 0.357 | 2.14E-15 |
| LEF1 | 9.23E-18 | 0.297639079 | 0.411 | 0.35 | 4.60E-15 |
| IRF4 | 7.73E-17 | 0.252339525 | 0.989 | 0.949 | 3.85E-14 |
| TRIB1 | 1.16E-16 | 0.310915333 | 0.58 | 0.387 | 5.80E-14 |
| NFKB2 | 6.56E-16 | -0.179785459 | 0.235 | 0.317 | 3.27E-13 |
| NT5E | 3.54E-15 | -0.263674688 | 0.148 | 0.224 | 1.76E-12 |
| SLC7A1 | 4.29E-15 | 0.189365061 | 0.734 | 0.637 | 2.13E-12 |
| PSME2 | 1.63E-14 | 0.185090787 | 0.807 | 0.752 | 8.11E-12 |
| IGLC3 | 2.25E-14 | 0.304279927 | 0.282 | 0.278 | 1.12E-11 |
| LIF | 6.06E-14 | -0.533148969 | 0.343 | 0.376 | 3.02E-11 |
| EIF1 | 6.25E-14 | 0.13529495 | 0.999 | 1 | 3.11E-11 |
| PIK3IP1 | 2.75E-13 | -0.188719969 | 0.182 | 0.208 | 1.37E-10 |
| LGALS1 | 1.33E-12 | -0.233265603 | 0.294 | 0.363 | 6.62E-10 |
| CXCL10 | 1.83E-12 | 0.26097653 | 0.151 | 0.147 | 9.10E-10 |
| CYCS | 2.22E-12 | -0.149826108 | 0.952 | 0.96 | 1.11E-09 |
| CCL20 | 3.60E-12 | -0.735629663 | 0.523 | 0.57 | 1.79E-09 |
| ITGB2 | 3.88E-12 | 0.129637041 | 0.182 | 0.12 | 1.93E-09 |
| MYB | 4.21E-12 | 0.111074755 | 0.092 | 0.047 | 2.09E-09 |
| ICOS | 4.75E-12 | 0.316090464 | 0.862 | 0.787 | 2.36E-09 |
| STAT6 | 1.43E-11 | 0.137964755 | 0.363 | 0.277 | 7.13E-09 |
| RAPH1 | 2.02E-11 | -0.100682774 | 0.072 | 0.132 | 1.00E-08 |
| SOCS2 | 3.03E-11 | -0.152102602 | 0.103 | 0.165 | 1.51E-08 |
| GATA3 | 4.22E-11 | -0.065913507 | 0.021 | 0.056 | 2.10E-08 |
| UGCG | 1.32E-10 | 0.135770282 | 0.281 | 0.235 | 6.57E-08 |
| LDHA | 2.99E-10 | 0.156478651 | 0.995 | 0.985 | 1.49E-07 |
| PRF1 | 3.06E-10 | -0.196930935 | 0.445 | 0.46 | 1.52E-07 |
| BCL11B | 3.82E-10 | 0.166101355 | 0.291 | 0.224 | 1.90E-07 |
| BTLA | 3.86E-10 | 0.177665714 | 0.318 | 0.238 | 1.92E-07 |

| | | | | | |
|---|---|---|---|---|---|
| CD40 | 4.43E-10 | -0.139664315 | 0.186 | 0.228 | 2.21E-07 |
| POU2F2 | 4.46E-10 | 0.146854708 | 0.541 | 0.442 | 2.22E-07 |
| LGALS9 | 5.89E-10 | 0.065798829 | 0.066 | 0.031 | 2.93E-07 |
| IKZF1 | 1.03E-09 | 0.092071993 | 0.137 | 0.092 | 5.15E-07 |
| POU2AF1 | 1.68E-09 | 0.213230646 | 0.257 | 0.178 | 8.38E-07 |
| PCNA | 2.28E-09 | -0.121216675 | 0.262 | 0.305 | 1.13E-06 |
| GZMB | 3.62E-09 | 0.256164495 | 0.934 | 0.935 | 1.80E-06 |
| SELPLG | 4.07E-09 | -0.076285421 | 0.027 | 0.051 | 2.03E-06 |
| TRAT1 | 5.43E-09 | -0.096474555 | 0.069 | 0.113 | 2.71E-06 |
| PASK | 5.58E-09 | 0.155319399 | 0.185 | 0.132 | 2.78E-06 |
| TBX21 | 7.94E-09 | 0.248294193 | 0.723 | 0.685 | 3.95E-06 |
| PSME3 | 9.46E-09 | 0.123806663 | 0.541 | 0.465 | 4.71E-06 |
| TNFSF14 | 2.41E-08 | 0.15276194 | 0.414 | 0.318 | 1.20E-05 |
| ERBIN | 6.22E-08 | -0.111463566 | 0.24 | 0.297 | 3.10E-05 |
| TNIK | 9.99E-08 | -0.091098195 | 0.105 | 0.159 | 4.98E-05 |
| CHD1 | 1.61E-07 | -0.046423635 | 0.025 | 0.057 | 8.02E-05 |
| GIMAP2 | 1.84E-07 | 0.055627599 | 0.087 | 0.05 | 9.15E-05 |
| IL6 | 2.28E-07 | -0.160173842 | 0.294 | 0.295 | 0.000113449 |
| TNFSF10 | 3.82E-07 | -0.09637791 | 0.038 | 0.08 | 0.000190386 |
| CD4 | 5.12E-07 | 0.048160014 | 0.054 | 0.029 | 0.000254796 |
| IGKC | 7.50E-07 | -0.163589553 | 0.167 | 0.181 | 0.000373591 |
| CD28 | 9.49E-07 | 0.097877922 | 0.205 | 0.165 | 0.000472651 |
| RORA | 9.76E-07 | 0.145320952 | 0.271 | 0.232 | 0.000486291 |
| NAMPT | 1.06E-06 | 0.151911249 | 0.986 | 0.978 | 0.000527871 |
| RBCK1 | 1.92E-06 | 0.078674205 | 0.212 | 0.153 | 0.000954036 |
| SNX8 | 2.24E-06 | 0.093178521 | 0.293 | 0.233 | 0.001115932 |
| TNFRSF13C | 2.87E-06 | 0.097103688 | 0.205 | 0.164 | 0.001429816 |
| RORC | 2.87E-06 | -0.121298432 | 0.056 | 0.092 | 0.001429992 |
| IL15RA | 3.80E-06 | 0.109099202 | 0.375 | 0.32 | 0.001893092 |
| TIMM9 | 4.46E-06 | 0.103676347 | 0.69 | 0.622 | 0.002220058 |
| CD70 | 5.02E-06 | 0.054951525 | 0.041 | 0.026 | 0.002497907 |
| PSMB8 | 5.45E-06 | -0.120586133 | 0.614 | 0.681 | 0.002712561 |
| IL2 | 5.86E-06 | -0.256725292 | 0.999 | 1 | 0.002918223 |
| SELL | 6.67E-06 | 0.215546455 | 0.504 | 0.463 | 0.003319996 |
| RUNX3 | 7.80E-06 | -0.131289238 | 0.527 | 0.582 | 0.003882001 |
| LAG3 | 9.04E-06 | 0.077899633 | 0.084 | 0.056 | 0.004502207 |
| CD63 | 1.02E-05 | 0.107876147 | 0.652 | 0.589 | 0.005072363 |
| CD200 | 1.29E-05 | 0.232257098 | 0.697 | 0.592 | 0.006412312 |
| CD48 | 1.42E-05 | -0.148989534 | 0.915 | 0.929 | 0.007059623 |
| IL1RN | 1.43E-05 | -0.10532339 | 0.19 | 0.202 | 0.00713157 |
| GZMK | 1.61E-05 | 0.041390388 | 0.024 | 0.013 | 0.008025322 |
| CXCL2 | 1.96E-05 | -0.117399542 | 0.324 | 0.338 | 0.009763098 |
| ATF6B | 2.20E-05 | 0.042153353 | 0.073 | 0.048 | 0.010959203 |
| PTPRC | 3.19E-05 | -0.13292934 | 0.692 | 0.771 | 0.015899103 |
| CD3G | 3.72E-05 | 0.028186408 | 0.033 | 0.015 | 0.01852206 |
| UFM1 | 4.60E-05 | 0.097655575 | 0.957 | 0.906 | 0.022916406 |
| BCL2 | 5.12E-05 | 0.071336462 | 0.223 | 0.176 | 0.02550341 |

| | | | | | |
|---|---|---|---|---|---|
| TSPAN32 | 6.53E-05 | 0.021452386 | 0.025 | 0.01 | 0.032497635 |
| WAS | 7.82E-05 | 0.042904146 | 0.076 | 0.056 | 0.038965236 |
| PSMB9 | 7.95E-05 | 0.095777605 | 0.374 | 0.317 | 0.039580861 |
| STAT1 | 9.12E-05 | 0.127044983 | 0.625 | 0.598 | 0.045439821 |
| CCR6 | 0.000127077 | 0.051529885 | 0.027 | 0.013 | 0.063284196 |
| CBLB | 0.000204184 | -0.042144683 | 0.05 | 0.067 | 0.101683813 |
| HLA.A | 0.000219291 | 0.080969027 | 1 | 1 | 0.109207099 |
| LAT | 0.000265467 | 0.111264225 | 0.369 | 0.302 | 0.132202483 |
| TGFBR1 | 0.000282605 | -0.043972637 | 0.075 | 0.105 | 0.140737397 |
| NFATC3 | 0.000318781 | -0.049664914 | 0.102 | 0.129 | 0.158752765 |
| GRB10 | 0.000355311 | -0.02799816 | 0.018 | 0.033 | 0.176944787 |
| TNFSF8 | 0.000355455 | 0.165816005 | 0.425 | 0.342 | 0.177016823 |
| LAMP1 | 0.000547453 | 0.073237067 | 0.442 | 0.394 | 0.272631552 |
| HAVCR2 | 0.000554728 | 0.0427509 | 0.06 | 0.049 | 0.276254694 |
| CD247 | 0.000635833 | -0.040384991 | 0.058 | 0.079 | 0.316644942 |
| CXCL9 | 0.000956071 | 0.066146313 | 0.063 | 0.058 | 0.476123406 |
| CD109 | 0.00129644 | -0.090162412 | 0.184 | 0.182 | 0.645627111 |
| CCR8 | 0.001395685 | -0.017693186 | 0.003 | 0.012 | 0.695051277 |
| NGDN | 0.001442027 | -0.026067238 | 0.029 | 0.047 | 0.718129222 |
| THBS1 | 0.001516649 | 0.085932011 | 0.062 | 0.058 | 0.755291054 |
| LIG4 | 0.001601662 | 0.041073313 | 0.131 | 0.097 | 0.797627788 |
| CCL3 | 0.001814357 | -0.127726214 | 0.998 | 0.998 | 0.903549849 |
| HLA.DPA1 | 0.001880552 | 0.066985383 | 0.276 | 0.229 | 0.936514777 |
| IL5 | 0.001988316 | -0.312714066 | 0.016 | 0.023 | 0.990181128 |
| SP140 | 0.001998002 | -0.040148394 | 0.072 | 0.087 | 0.995004809 |
| FOXP1 | 0.002007532 | 0.071921028 | 0.569 | 0.534 | 0.999750694 |
| CNOT2 | 0.002708092 | -0.057949411 | 0.333 | 0.361 | 1 |
| IFNGR1 | 0.00297739 | -0.050878357 | 0.154 | 0.177 | 1 |
| ABCE1 | 0.003055131 | -0.063966862 | 0.833 | 0.838 | 1 |
| IL1B | 0.003105873 | -0.086109934 | 0.41 | 0.438 | 1 |
| ADA | 0.003180054 | -0.041498191 | 0.059 | 0.08 | 1 |
| TUBA1C | 0.003327361 | -0.079130226 | 0.94 | 0.895 | 1 |
| CD24 | 0.004049511 | -0.013619022 | 0.003 | 0.009 | 1 |
| BAX | 0.0042997 | -0.057817524 | 0.286 | 0.312 | 1 |
| KLRC3 | 0.004847483 | -0.034242204 | 0.048 | 0.059 | 1 |
| FYB1 | 0.005739497 | -0.056283467 | 0.139 | 0.173 | 1 |
| CXCL3 | 0.006811516 | -0.073698313 | 0.148 | 0.16 | 1 |
| TGFB1 | 0.007497271 | -0.015472915 | 0.014 | 0.025 | 1 |
| NKG7 | 0.007620563 | -0.069204662 | 0.297 | 0.322 | 1 |
| ZBTB16 | 0.008376688 | -0.011846125 | 0.003 | 0.009 | 1 |
| IL18RAP | 0.013409746 | -0.033858623 | 0.044 | 0.059 | 1 |
| CD300A | 0.014618224 | -0.01575455 | 0.016 | 0.025 | 1 |
| IL9 | 0.017060957 | -0.54244164 | 0.031 | 0.035 | 1 |
| BCL6 | 0.018584124 | -0.015956851 | 0.019 | 0.03 | 1 |
| IL17RB | 0.02009491 | -0.018205456 | 0.001 | 0.003 | 1 |
| CTLA4 | 0.022119734 | -0.176903971 | 0.601 | 0.46 | 1 |
| DOCK8 | 0.023725972 | -0.043448001 | 0.21 | 0.238 | 1 |

| | | | | | |
|---|---|---|---|---|---|
| VEGFA | 0.026586517 | -0.008020468 | 0.003 | 0.007 | 1 |
| THEMIS | 0.030391305 | -0.023580619 | 0.036 | 0.051 | 1 |
| STIM1 | 0.03531248 | -0.031127649 | 0.12 | 0.137 | 1 |
| CXCL16 | 0.041954404 | -0.022077862 | 0.055 | 0.067 | 1 |
| IL3RA | 0.043074751 | -0.020889706 | 0.049 | 0.055 | 1 |
| QPCT | 0.048813992 | -0.011357596 | 0.014 | 0.021 | 1 |
| CXCL11 | 0.048960406 | -0.010027586 | 0.006 | 0.011 | 1 |
| IL26 | 0.048975986 | -0.014597227 | 0.01 | 0.018 | 1 |
| ITGAM | 0.050545963 | -0.011063315 | 0.011 | 0.017 | 1 |
| IFI6 | 0.051559501 | -0.011528129 | 0.014 | 0.02 | 1 |
| IL12RB1 | 0.055148145 | -0.004416683 | 0.001 | 0.004 | 1 |
| LILRB4 | 0.055865247 | -0.010111789 | 0.014 | 0.02 | 1 |
| IGHG1 | 0.063464735 | -0.036862299 | 0.105 | 0.111 | 1 |
| VMO1 | 0.063803526 | -0.016801633 | 0.025 | 0.033 | 1 |
| CXCL5 | 0.066244612 | -0.038764838 | 0.242 | 0.241 | 1 |
| ICAM1 | 0.071262655 | -0.053035136 | 0.444 | 0.448 | 1 |
| PLCG2 | 0.083472919 | -0.00845884 | 0.007 | 0.013 | 1 |
| PSEN1 | 0.083646194 | -0.014384449 | 0.038 | 0.049 | 1 |
| MCM4 | 0.09168417 | -0.015138156 | 0.047 | 0.057 | 1 |
| LYN | 0.094238298 | -0.018403343 | 0.055 | 0.06 | 1 |
| TGFBI | 0.095270335 | -0.021619463 | 0.075 | 0.086 | 1 |
| ZNF683 | 0.095752635 | -0.003969682 | 0.001 | 0.003 | 1 |
| EIF3J | 0.100848346 | -0.034000533 | 0.417 | 0.434 | 1 |
| CD160 | 0.107772836 | -0.006823688 | 0.006 | 0.009 | 1 |
| GZMA | 0.112742227 | -0.012637325 | 0.025 | 0.031 | 1 |
| ITK | 0.113061904 | -0.050683405 | 0.904 | 0.862 | 1 |
| MS4A1 | 0.114367221 | -0.013133418 | 0.033 | 0.038 | 1 |
| CCR1 | 0.127386807 | -0.020449991 | 0.095 | 0.104 | 1 |
| PTGDR2 | 0.133334015 | -0.003352222 | 0.002 | 0.004 | 1 |
| ILF2 | 0.135054823 | -0.028524311 | 0.956 | 0.956 | 1 |
| GNLY | 0.138417098 | -0.035655083 | 0.25 | 0.256 | 1 |
| IL4R | 0.140609896 | -0.037488222 | 0.797 | 0.787 | 1 |
| IL15 | 0.140762316 | -0.003905267 | 0.002 | 0.005 | 1 |
| HLA.DQB1 | 0.144910951 | -0.049905844 | 0.32 | 0.307 | 1 |
| ANXA5 | 0.166817697 | -0.038816202 | 0.56 | 0.555 | 1 |
| IL17F | 0.173561418 | -0.429323554 | 0.036 | 0.037 | 1 |
| CD27 | 0.176018943 | -0.016601222 | 0.042 | 0.044 | 1 |
| CXCR6 | 0.183431462 | -0.004627527 | 0.003 | 0.004 | 1 |
| S100A9 | 0.186285874 | -0.012485695 | 0.032 | 0.038 | 1 |
| CTSD | 0.187327391 | -0.014236236 | 0.054 | 0.063 | 1 |
| IGHM | 0.187487184 | -0.009757943 | 0.018 | 0.02 | 1 |
| MMP9 | 0.190397366 | -0.015348372 | 0.07 | 0.074 | 1 |
| CD79A | 0.195380508 | -0.007537873 | 0.018 | 0.022 | 1 |
| FCER1G | 0.195482734 | -0.022721529 | 0.147 | 0.156 | 1 |
| FN1 | 0.203219837 | -0.022633315 | 0.115 | 0.132 | 1 |
| GNAI2 | 0.206935623 | -0.035197572 | 0.468 | 0.505 | 1 |
| ITGAX | 0.218161845 | -0.019943101 | 0.1 | 0.102 | 1 |

| | | | | | |
|---|---|---|---|---|---|
| CXCR5 | 0.221047839 | -0.069119913 | 0.352 | 0.297 | 1 |
| KLRC1 | 0.225289497 | -0.013185418 | 0.041 | 0.043 | 1 |
| CXCL1 | 0.226666623 | -0.012804099 | 0.045 | 0.05 | 1 |
| CD163 | 0.233048743 | -0.004811874 | 0.008 | 0.011 | 1 |
| DPP4 | 0.236428364 | -0.010542811 | 0.02 | 0.025 | 1 |
| CD33 | 0.239299058 | -0.001624394 | 0 | 0.002 | 1 |
| CD274 | 0.255764503 | -0.039175233 | 0.454 | 0.437 | 1 |
| IL23R | 0.273303744 | -0.007165751 | 0.008 | 0.011 | 1 |
| ADGRE1 | 0.304414727 | -0.003988143 | 0.007 | 0.008 | 1 |
| IFITM2 | 0.329762555 | -0.021590823 | 0.224 | 0.246 | 1 |
| TGFBR2 | 0.334347217 | -0.029773257 | 0.644 | 0.644 | 1 |
| CLEC10A | 0.340396432 | -0.001176143 | 0 | 0.001 | 1 |
| SNX9 | 0.363691177 | -0.025311703 | 0.773 | 0.765 | 1 |
| CCL22 | 0.375088189 | -0.016179813 | 0.155 | 0.163 | 1 |
| EBF1 | 0.387227494 | -0.005315892 | 0.021 | 0.023 | 1 |
| IGBP1 | 0.388781803 | -0.011725864 | 0.13 | 0.13 | 1 |
| IL18R1 | 0.400924413 | -0.011083052 | 0.063 | 0.065 | 1 |
| KCNE3 | 0.403531548 | -0.001176143 | 0 | 0.001 | 1 |
| CD36 | 0.411536916 | -0.004671406 | 0.019 | 0.023 | 1 |
| IRF3 | 0.415191945 | -0.00465694 | 0.012 | 0.015 | 1 |
| TIAF1 | 0.431341912 | -0.003141776 | 0.009 | 0.011 | 1 |
| ITGAE | 0.437020509 | -0.00736733 | 0.04 | 0.044 | 1 |
| IGHG2 | 0.437711902 | -0.001840609 | 0.002 | 0.003 | 1 |
| IKZF2 | 0.443207664 | -0.002225609 | 0.003 | 0.004 | 1 |
| CCL2 | 0.44331025 | -0.025193078 | 0.585 | 0.56 | 1 |
| B4GALT5 | 0.460165664 | -0.007876784 | 0.07 | 0.074 | 1 |
| SLC25A37 | 0.462930317 | -0.012063566 | 0.14 | 0.145 | 1 |
| CR2 | 0.465241218 | -0.004047308 | 0.015 | 0.014 | 1 |
| TREM1 | 0.472103318 | -0.002163487 | 0.004 | 0.006 | 1 |
| VNN2 | 0.476305501 | -0.001006405 | 0.001 | 0.002 | 1 |
| CD79B | 0.481388473 | -0.001778885 | 0.003 | 0.005 | 1 |
| KLRC4 | 0.511071716 | -0.001886622 | 0.004 | 0.005 | 1 |
| TCF4 | 0.572550028 | -0.001115138 | 0.002 | 0.002 | 1 |
| FOXP3 | 0.572707593 | -0.014236567 | 0.031 | 0.033 | 1 |
| TNFRSF9 | 0.586476198 | -0.004344876 | 0.031 | 0.035 | 1 |
| ORAI1 | 0.593161587 | -0.012044999 | 0.449 | 0.422 | 1 |
| NFATC2 | 0.602049641 | -0.007869352 | 0.129 | 0.125 | 1 |
| CCL13 | 0.603518493 | -0.001393192 | 0.001 | 0.003 | 1 |
| CD86 | 0.611760863 | -0.004578264 | 0.043 | 0.047 | 1 |
| CCND2 | 0.638322992 | -0.015042255 | 0.577 | 0.572 | 1 |
| TMEM97 | 0.638472688 | -0.004010391 | 0.041 | 0.045 | 1 |
| LY86 | 0.646974474 | -0.001702138 | 0.008 | 0.009 | 1 |
| XBP1 | 0.656212677 | -0.010842694 | 0.573 | 0.54 | 1 |
| CYBB | 0.658431853 | -0.004384178 | 0.054 | 0.059 | 1 |
| TCL1A | 0.661646596 | -0.001054287 | 0.003 | 0.003 | 1 |
| IL21 | 0.677375125 | -0.041768782 | 0.312 | 0.158 | 1 |
| LAT2 | 0.697816935 | -0.002353139 | 0.021 | 0.023 | 1 |

| | | | | | |
|---|---|---|---|---|---|
| TNIP3 | 0.720608827 | -0.001318133 | 0.005 | 0.008 | 1 |
| CCR5 | 0.72266645 | -0.002760868 | 0.035 | 0.039 | 1 |
| GZMH | 0.732706363 | -0.003701126 | 0.048 | 0.054 | 1 |
| CLEC4E | 0.737915411 | -0.000885022 | 0.003 | 0.004 | 1 |
| BCL2L1 | 0.752150848 | -0.007942546 | 0.37 | 0.365 | 1 |
| MGST1 | 0.768145471 | -0.001088877 | 0.009 | 0.009 | 1 |
| IRF8 | 0.780616015 | -0.013097773 | 0.856 | 0.816 | 1 |
| THBD | 0.792948265 | -0.000497942 | 0.002 | 0.002 | 1 |
| BATF | 0.811570961 | -0.006699682 | 0.515 | 0.477 | 1 |
| IGHG3 | 0.815428554 | -0.000764249 | 0.006 | 0.006 | 1 |
| RNASE6 | 0.819372373 | -0.000655654 | 0.005 | 0.006 | 1 |
| BIN2 | 0.820603156 | -0.000812409 | 0.008 | 0.009 | 1 |
| CX3CR1 | 0.832649231 | -0.001507765 | 0.026 | 0.025 | 1 |
| ENTPD1 | 0.837145588 | -0.000497942 | 0.002 | 0.002 | 1 |
| CD72 | 0.846556463 | -0.000752318 | 0.009 | 0.01 | 1 |
| NDFIP2 | 0.849554594 | -0.002972228 | 0.141 | 0.151 | 1 |
| LTB | 0.925701196 | -0.000811675 | 0.033 | 0.035 | 1 |
| VPREB3 | 0.92572527 | -0.000109764 | 0.001 | 0.001 | 1 |
| KLRK1 | 0.928203962 | -0.0033171 | 0.254 | 0.238 | 1 |
| CD1C | 0.946785351 | -0.000209206 | 0.005 | 0.004 | 1 |
| FCN1 | 0.952956892 | -0.000335864 | 0.016 | 0.018 | 1 |
| PIK3AP1 | 0.962245956 | -0.000564539 | 0.073 | 0.069 | 1 |
| IL11RA | 0.968847888 | -0.000322439 | 0.035 | 0.039 | 1 |
| STAT3 | 0.970531736 | -0.000991517 | 0.753 | 0.706 | 1 |
| CHI3L2 | 0.971447128 | -0.000100003 | 0.004 | 0.004 | 1 |
| BLNK | 0.978049723 | -5.01E-05 | 0.002 | 0.002 | 1 |
| KIT | 0.994014271 | -0.002242648 | 0 | 0.002 | 1 |
| PRDM1 | 0.996151674 | -0.000129921 | 0.195 | 0.189 | 1 |
| PAX5 | 0.996211328 | -0.001794397 | 0 | 0.001 | 1 |
| TLR8 | 0.996269883 | -0.001346007 | 0 | 0.001 | 1 |
| CMKLR1 | 0.996269883 | -0.001346007 | 0 | 0.001 | 1 |
| PAXBP1 | 0.002238653 | 0.062752097 | 0.339 | 0.301 | 1 |
| CCR9 | 0.005076876 | 0.009576972 | 0.004 | 0.001 | 1 |
| FOXO1 | 0.005246073 | 0.045999154 | 0.166 | 0.143 | 1 |
| AIM2 | 0.005768242 | 0.022108619 | 0.044 | 0.026 | 1 |
| CCR4 | 0.005944858 | 0.120463705 | 0.575 | 0.498 | 1 |
| GAPDH | 0.006182534 | 0.089792964 | 0.97 | 0.902 | 1 |
| ZAP70 | 0.007834669 | 0.03142177 | 0.077 | 0.063 | 1 |
| PDCD1 | 0.008618603 | 0.048423861 | 0.154 | 0.12 | 1 |
| CD37 | 0.011668187 | 0.051551302 | 0.369 | 0.337 | 1 |
| TNFSF13B | 0.013867105 | 0.020728378 | 0.019 | 0.014 | 1 |
| AQP9 | 0.015605859 | 0.011076868 | 0.014 | 0.008 | 1 |
| IL6R | 0.019078776 | 0.03168998 | 0.116 | 0.092 | 1 |
| LRRC32 | 0.020984082 | 0.096452108 | 0.059 | 0.05 | 1 |
| LIPA | 0.021381393 | 0.03179599 | 0.107 | 0.091 | 1 |
| SLC1A5 | 0.021547496 | 0.058859076 | 0.949 | 0.927 | 1 |
| CD8A | 0.024043833 | 0.067283038 | 0.131 | 0.099 | 1 |

| | | | | | |
|---|---|---|---|---|---|
| S100A12 | 0.024062544 | 0.00644019 | 0.007 | 0.003 | 1 |
| MALT1 | 0.027698704 | 0.036768359 | 0.152 | 0.141 | 1 |
| PMCH | 0.029657685 | 0.015008149 | 0.01 | 0.005 | 1 |
| EOMES | 0.030608899 | 0.005158968 | 0.005 | 0.002 | 1 |
| CD209 | 0.030871925 | 0.00404115 | 0.003 | 0.001 | 1 |
| FAS | 0.031087707 | 0.034312674 | 0.123 | 0.108 | 1 |
| CD44 | 0.036707312 | 0.048918252 | 0.999 | 0.995 | 1 |
| LAP3 | 0.038688336 | 0.040269267 | 0.291 | 0.272 | 1 |
| TRDC | 0.042009953 | 0.02549505 | 0.024 | 0.019 | 1 |
| EEF1E1 | 0.042227985 | 0.036563397 | 0.273 | 0.245 | 1 |
| FURIN | 0.044113591 | 0.070279756 | 0.667 | 0.592 | 1 |
| BCL10 | 0.067850175 | 0.028307199 | 0.168 | 0.148 | 1 |
| MZB1 | 0.096763669 | 0.007535221 | 0.01 | 0.006 | 1 |
| TCF7 | 0.103343197 | 0.022994358 | 0.104 | 0.092 | 1 |
| FCER2 | 0.11786281 | 0.011913723 | 0.039 | 0.028 | 1 |
| RGS1 | 0.123912087 | 0.095599214 | 0.538 | 0.54 | 1 |
| YBX3 | 0.133873527 | 0.022507339 | 0.099 | 0.08 | 1 |
| HLA.DRA | 0.142862579 | 0.043508957 | 0.577 | 0.553 | 1 |
| TGFB3 | 0.151777137 | 0.002359547 | 0.003 | 0.001 | 1 |
| CD244 | 0.181352886 | 0.002191145 | 0.002 | 0.001 | 1 |
| IGHD | 0.181597548 | 0.004870744 | 0.009 | 0.005 | 1 |
| CTSW | 0.181629667 | 0.009302771 | 0.029 | 0.024 | 1 |
| SNCA | 0.199778222 | 0.005440255 | 0.001 | 0.001 | 1 |
| GAB2 | 0.203317873 | 0.013648265 | 0.06 | 0.054 | 1 |
| CASP5 | 0.203619471 | 0.009005803 | 0.036 | 0.027 | 1 |
| FCGR3A | 0.21007326 | 0.009047569 | 0.024 | 0.02 | 1 |
| TYMS | 0.226674978 | 0.005308496 | 0.012 | 0.007 | 1 |
| KLRG1 | 0.228466058 | 0.005089974 | 0.008 | 0.007 | 1 |
| KLRF1 | 0.246376943 | 0.002752003 | 0.005 | 0.003 | 1 |
| PAK1IP1 | 0.247897744 | 0.024766791 | 0.453 | 0.416 | 1 |
| AHI1 | 0.268993962 | 0.012380704 | 0.057 | 0.054 | 1 |
| CD9 | 0.297111972 | 0.012995425 | 0.077 | 0.075 | 1 |
| HLA.DMA | 0.318372939 | 0.010537294 | 0.063 | 0.055 | 1 |
| IL1R2 | 0.327207742 | 0.006073381 | 0.013 | 0.008 | 1 |
| CCL5 | 0.334491252 | 0.043851043 | 0.225 | 0.218 | 1 |
| CARD11 | 0.354170384 | 0.002136387 | 0.004 | 0.003 | 1 |
| CD14 | 0.379597235 | 0.007828365 | 0.021 | 0.02 | 1 |
| FUT4 | 0.403097646 | 0.004842308 | 0.022 | 0.018 | 1 |
| KDELR1 | 0.418313314 | 0.0105172 | 0.11 | 0.104 | 1 |
| LAMP3 | 0.424006021 | 0.022110237 | 0.292 | 0.26 | 1 |
| CHD7 | 0.425155718 | 0.008392842 | 0.072 | 0.066 | 1 |
| PGM3 | 0.446252951 | 0.007270646 | 0.054 | 0.048 | 1 |
| IL18 | 0.447959135 | 0.001742616 | 0.001 | 0.001 | 1 |
| CD8B | 0.450947141 | 0.002025106 | 0.005 | 0.003 | 1 |
| CD80 | 0.464967767 | 0.003085202 | 0.012 | 0.01 | 1 |
| TNFRSF17 | 0.467852727 | 0.001294225 | 0.002 | 0.001 | 1 |
| ZNF341 | 0.470245805 | 0.00269636 | 0.01 | 0.008 | 1 |

| | | | | | |
|---|---|---|---|---|---|
| DCLRE1C | 0.502509444 | 0.005604829 | 0.04 | 0.039 | 1 |
| CXCL13 | 0.52098456 | 0.042572925 | 0.001 | 0.002 | 1 |
| NCAM1 | 0.52171309 | 0.001352062 | 0.003 | 0.002 | 1 |
| VSIG4 | 0.548955193 | 0.001183491 | 0.003 | 0.002 | 1 |
| IL2RA | 0.555724736 | 0.015172856 | 0.514 | 0.492 | 1 |
| ARL4C | 0.57080743 | 0.011560419 | 0.108 | 0.119 | 1 |
| TOP2A | 0.577097657 | 0.001183491 | 0.003 | 0.002 | 1 |
| SLC7A7 | 0.587780499 | 0.001970871 | 0.006 | 0.005 | 1 |
| SOCS1 | 0.612489325 | 0.000845975 | 0.002 | 0.002 | 1 |
| JCHAIN | 0.628663645 | 0.008824351 | 0.045 | 0.056 | 1 |
| IL1R1 | 0.667751049 | 0.016127671 | 0.24 | 0.211 | 1 |
| TLR2 | 0.675479166 | 0.003609474 | 0.047 | 0.042 | 1 |
| MCM2 | 0.685792307 | 0.004368235 | 0.071 | 0.069 | 1 |
| NRP1 | 0.70757116 | 0.003290061 | 0.036 | 0.04 | 1 |
| SKAP1 | 0.720389035 | 0.001642835 | 0.012 | 0.012 | 1 |
| SMAD2 | 0.769553414 | 0.003896387 | 0.114 | 0.113 | 1 |
| JAK2 | 0.770345939 | 0.002698537 | 0.045 | 0.044 | 1 |
| IFITM3 | 0.770404715 | 0.002911115 | 0.052 | 0.051 | 1 |
| SRGN | 0.808117543 | 0.005858993 | 0.924 | 0.897 | 1 |
| IGHA1 | 0.819677611 | 0.000397863 | 0.002 | 0.002 | 1 |
| MITF | 0.823974262 | 0.000742523 | 0.006 | 0.006 | 1 |
| IL12A | 0.840748334 | 0.000980738 | 0.015 | 0.013 | 1 |
| OSM | 0.866636335 | 0.001737318 | 0.044 | 0.039 | 1 |
| IL1A | 0.934023346 | 0.001787584 | 0.048 | 0.056 | 1 |
| LGALS3 | 0.935055435 | 0.000942332 | 0.077 | 0.075 | 1 |
| IRF9 | 0.943682882 | 0.001957349 | 0.433 | 0.394 | 1 |
| SPP1 | 0.953174615 | 0.000387334 | 0.019 | 0.017 | 1 |
| B3GAT1 | 0.959476529 | 0.000262833 | 0.014 | 0.014 | 1 |
| CD22 | 0.980390624 | 0.00024737 | 0.046 | 0.044 | 1 |

CD8+ Naïve

| | p_val | avg_log2FC | Patient | Controls | p_val_adj |
|---|---|---|---|---|---|
| BTG1 | 5.69E-48 | -1.274781142 | 0.997 | 0.997 | 2.83E-45 |
| GZMB | 3.19E-32 | 1.387259827 | 0.931 | 0.915 | 1.59E-29 |
| FYN | 5.05E-29 | -0.707536404 | 0.794 | 0.932 | 2.52E-26 |
| NFKBIZ | 1.06E-25 | -1.352200384 | 0.68 | 0.767 | 5.27E-23 |
| GBP1 | 9.22E-23 | 0.799662034 | 0.68 | 0.448 | 4.59E-20 |
| CD69 | 1.20E-21 | -0.645363905 | 1 | 1 | 5.99E-19 |
| IL7R | 1.90E-17 | -0.641580535 | 0.392 | 0.636 | 9.45E-15 |
| LDHA | 4.92E-17 | 0.438109953 | 0.997 | 0.997 | 2.45E-14 |
| CREM | 2.98E-16 | -0.748129104 | 0.749 | 0.841 | 1.48E-13 |
| NR4A2 | 4.86E-16 | -1.784840649 | 0.278 | 0.518 | 2.42E-13 |
| GBP2 | 9.61E-16 | 0.536544928 | 0.969 | 0.873 | 4.79E-13 |
| PDIA6 | 1.28E-15 | 0.601673007 | 0.893 | 0.671 | 6.38E-13 |
| EVI2B | 1.37E-15 | -0.645233906 | 0.258 | 0.444 | 6.84E-13 |
| EGR3 | 7.14E-15 | -0.721429307 | 0.12 | 0.296 | 3.56E-12 |

| | | | | | |
|---|---|---|---|---|---|
| EGR1 | 3.13E-14 | -0.900256269 | 0.485 | 0.623 | 1.56E-11 |
| FOS | 6.11E-14 | -1.639330518 | 0.491 | 0.549 | 3.04E-11 |
| FOSB | 7.18E-13 | -1.432481217 | 0.247 | 0.465 | 3.58E-10 |
| DUSP2 | 1.77E-12 | -0.618264336 | 0.99 | 0.99 | 8.81E-10 |
| PFN1 | 2.40E-12 | 0.364107435 | 0.997 | 0.997 | 1.20E-09 |
| REL | 3.49E-12 | -0.55797996 | 0.935 | 0.955 | 1.74E-09 |
| SLA | 6.61E-12 | -0.633079706 | 0.794 | 0.836 | 3.29E-09 |
| EVI2A | 1.49E-11 | -0.508067127 | 0.357 | 0.487 | 7.41E-09 |
| TGIF1 | 3.35E-11 | -0.481770383 | 0.57 | 0.681 | 1.67E-08 |
| KLRK1 | 6.06E-11 | -0.469058495 | 0.89 | 0.93 | 3.02E-08 |
| SDCBP | 7.54E-11 | -0.604832122 | 0.835 | 0.81 | 3.75E-08 |
| IL2RA | 1.36E-10 | 0.406901149 | 0.749 | 0.575 | 6.79E-08 |
| GEM | 1.64E-10 | -0.845779272 | 0.814 | 0.797 | 8.15E-08 |
| IL2RB | 1.75E-10 | 0.449754388 | 0.835 | 0.703 | 8.70E-08 |
| CSRNP1 | 4.60E-10 | -0.433515214 | 0.935 | 0.961 | 2.29E-07 |
| FURIN | 3.14E-09 | 0.47444466 | 0.746 | 0.593 | 1.56E-06 |
| MYC | 3.75E-09 | -0.379191129 | 0.99 | 0.997 | 1.87E-06 |
| CD274 | 9.44E-09 | 0.505964336 | 0.701 | 0.473 | 4.70E-06 |
| SPAG9 | 1.06E-08 | -0.394104005 | 0.326 | 0.472 | 5.29E-06 |
| JAK1 | 1.84E-08 | 0.34187723 | 0.911 | 0.814 | 9.14E-06 |
| IFRD1 | 2.04E-08 | -0.350047835 | 0.275 | 0.398 | 1.01E-05 |
| JUN | 3.29E-08 | -0.391876884 | 0.12 | 0.271 | 1.64E-05 |
| APOBEC3G | 4.86E-08 | 0.378610248 | 0.443 | 0.314 | 2.42E-05 |
| TGFBR2 | 8.86E-08 | 0.364700211 | 0.918 | 0.744 | 4.41E-05 |
| CCR7 | 9.26E-08 | -0.527952184 | 0.478 | 0.532 | 4.61E-05 |
| PDIA4 | 1.24E-07 | 0.334579901 | 0.615 | 0.461 | 6.16E-05 |
| EIF5 | 2.30E-07 | -0.29453354 | 0.921 | 0.938 | 0.000114337 |
| FOSL1 | 4.06E-07 | -0.31055387 | 0.863 | 0.884 | 0.000202073 |
| STAT3 | 4.37E-07 | 0.299864489 | 0.852 | 0.755 | 0.000217679 |
| EIF5A | 7.84E-07 | 0.241180649 | 1 | 0.988 | 0.000390341 |
| CTLA4 | 1.90E-06 | -0.593153522 | 0.206 | 0.252 | 0.000945093 |
| CD3E | 1.97E-06 | -0.342338758 | 0.869 | 0.862 | 0.000982994 |
| DUSP1 | 2.96E-06 | -0.479230912 | 0.371 | 0.447 | 0.001475846 |
| RUNX3 | 3.03E-06 | -0.357459919 | 0.388 | 0.477 | 0.001508706 |
| CXCR4 | 4.30E-06 | -0.552159711 | 0.182 | 0.266 | 0.002142176 |
| NFKB1 | 4.68E-06 | -0.203820705 | 1 | 1 | 0.002329927 |
| IRF7 | 5.13E-06 | -0.273273062 | 0.189 | 0.291 | 0.00255251 |
| PSME2 | 7.12E-06 | 0.229568381 | 0.945 | 0.907 | 0.003544207 |
| CCND2 | 7.50E-06 | 0.34752782 | 0.784 | 0.676 | 0.003736441 |
| CD27 | 8.47E-06 | -0.245367379 | 0.096 | 0.196 | 0.004216791 |
| STAT6 | 9.55E-06 | 0.224901289 | 0.454 | 0.35 | 0.004758003 |
| ITGA4 | 1.12E-05 | 0.185069666 | 0.213 | 0.106 | 0.005561175 |
| CCL20 | 1.34E-05 | -0.425420944 | 0.412 | 0.438 | 0.00665357 |
| ARHGEF3 | 1.51E-05 | -0.308390065 | 0.241 | 0.385 | 0.007510193 |
| DUSP5 | 2.10E-05 | -0.344470846 | 0.773 | 0.762 | 0.010475888 |
| UGCG | 3.20E-05 | 0.192506329 | 0.265 | 0.176 | 0.015956134 |
| RPN2 | 3.22E-05 | 0.219221775 | 0.677 | 0.581 | 0.016029259 |

| | | | | | |
|---|---|---|---|---|---|
| CD40LG | 3.76E-05 | -0.232541777 | 0.065 | 0.119 | 0.01872655 |
| FBXO11 | 3.93E-05 | -0.256871564 | 0.323 | 0.426 | 0.019555154 |
| NFKBID | 4.46E-05 | -0.16594548 | 0.069 | 0.159 | 0.022206183 |
| PSMB9 | 6.41E-05 | 0.235292895 | 0.536 | 0.388 | 0.031909184 |
| SLC1A5 | 7.03E-05 | -0.227467978 | 0.955 | 0.947 | 0.035013461 |
| TBX21 | 7.11E-05 | 0.361476644 | 0.89 | 0.787 | 0.035416934 |
| TRAC | 9.90E-05 | 0.297182366 | 0.979 | 0.938 | 0.049314995 |
| MYB | 0.000117615 | 0.233127971 | 0.296 | 0.204 | 0.058572389 |
| TRIB1 | 0.000141221 | 0.209516786 | 0.158 | 0.101 | 0.070327887 |
| IKZF1 | 0.00015747 | 0.184480649 | 0.316 | 0.195 | 0.078420267 |
| S100A10 | 0.000233668 | 0.360135689 | 0.763 | 0.668 | 0.11636642 |
| LGALS1 | 0.000245824 | -0.19762695 | 0.182 | 0.279 | 0.122420381 |
| PASK | 0.000290539 | 0.34192835 | 0.309 | 0.242 | 0.144688582 |
| BCL2A1 | 0.000307613 | -0.199079303 | 1 | 0.995 | 0.153191474 |
| IL1RN | 0.000355167 | 0.214167943 | 0.237 | 0.18 | 0.176873174 |
| F5 | 0.000375713 | 0.341049773 | 0.364 | 0.203 | 0.187105051 |
| SNX9 | 0.000497401 | -0.228858735 | 0.825 | 0.863 | 0.247705636 |
| CXCL8 | 0.000558477 | -0.515691108 | 0.914 | 0.875 | 0.278121479 |
| JUNB | 0.00058299 | -0.280832677 | 0.447 | 0.466 | 0.290329032 |
| CXCL2 | 0.000664048 | -0.312051702 | 0.302 | 0.344 | 0.330695751 |
| IRF4 | 0.000738572 | 0.20582057 | 0.997 | 0.979 | 0.367808745 |
| CBLB | 0.001310424 | -0.114884085 | 0.062 | 0.119 | 0.652590963 |
| ARPC1B | 0.001434767 | 0.149711963 | 0.323 | 0.244 | 0.71451402 |
| IL23A | 0.001585472 | -0.285600879 | 0.804 | 0.824 | 0.789564879 |
| CD83 | 0.001599859 | -0.315207144 | 0.649 | 0.696 | 0.79672996 |
| STIM1 | 0.001655711 | -0.114627574 | 0.079 | 0.151 | 0.824543947 |
| EEF1E1 | 0.001777922 | 0.148280007 | 0.519 | 0.403 | 0.885405069 |
| SELPLG | 0.00206274 | -0.136051301 | 0.062 | 0.105 | 1 |
| CD5 | 0.002068315 | -0.22935752 | 0.199 | 0.252 | 1 |
| CXCR3 | 0.002123449 | 0.379713599 | 0.581 | 0.43 | 1 |
| RPL6 | 0.002743997 | 0.114115317 | 1 | 1 | 1 |
| LAT | 0.002894664 | -0.174883801 | 0.244 | 0.322 | 1 |
| IVNS1ABP | 0.002999105 | -0.189847394 | 0.746 | 0.76 | 1 |
| HIF1A | 0.003089583 | -0.197750952 | 0.77 | 0.775 | 1 |
| IFNG | 0.003216579 | -0.288882447 | 0.962 | 0.974 | 1 |
| GIMAP5 | 0.00334891 | 0.1633869 | 0.186 | 0.12 | 1 |
| SELL | 0.003353222 | 0.298887604 | 0.67 | 0.59 | 1 |
| FASLG | 0.003499252 | -0.199742284 | 0.141 | 0.172 | 1 |
| JAK3 | 0.00360727 | 0.153465324 | 0.502 | 0.401 | 1 |
| IFITM3 | 0.00393275 | 0.10518089 | 0.155 | 0.102 | 1 |
| TRAF1 | 0.004972569 | -0.168099254 | 0.866 | 0.886 | 1 |
| CD38 | 0.005630024 | 0.033545299 | 0.024 | 0.004 | 1 |
| TRDC | 0.006172317 | 0.263701874 | 0.031 | 0.017 | 1 |
| ORAI1 | 0.006609422 | -0.154923684 | 0.509 | 0.568 | 1 |
| VPS28 | 0.007086971 | 0.127808362 | 0.402 | 0.331 | 1 |
| TIGIT | 0.007416851 | 0.070397266 | 0.069 | 0.032 | 1 |
| PIK3IP1 | 0.008532563 | -0.112647015 | 0.062 | 0.11 | 1 |

| | | | | | |
|---|---|---|---|---|---|
| RGS1 | 0.009068542 | 0.269958564 | 0.44 | 0.322 | 1 |
| IL6ST | 0.009556416 | -0.170805338 | 0.447 | 0.51 | 1 |
| ATF4 | 0.009593677 | -0.124695064 | 1 | 0.997 | 1 |
| TGFBR1 | 0.010217084 | 0.086864858 | 0.151 | 0.098 | 1 |
| TRIB2 | 0.010489662 | 0.06956134 | 0.079 | 0.032 | 1 |
| NDFIP2 | 0.01052613 | 0.131782612 | 0.416 | 0.27 | 1 |
| PTPN7 | 0.011075408 | 0.121494064 | 0.347 | 0.249 | 1 |
| CSF1 | 0.012177958 | 0.246158656 | 0.426 | 0.305 | 1 |
| CD2 | 0.012399838 | 0.144194864 | 0.99 | 0.981 | 1 |
| IL4R | 0.012954931 | -0.144519849 | 0.863 | 0.893 | 1 |
| IGHG1 | 0.013152536 | -0.159001312 | 0.124 | 0.114 | 1 |
| TNIK | 0.015525564 | -0.086569299 | 0.065 | 0.112 | 1 |
| ABCE1 | 0.015569683 | -0.114788975 | 0.942 | 0.944 | 1 |
| IL18R1 | 0.015967891 | -0.052999427 | 0.014 | 0.044 | 1 |
| CD6 | 0.017660745 | -0.169478446 | 0.615 | 0.66 | 1 |
| KLRC1 | 0.01947603 | -0.064745358 | 0.024 | 0.05 | 1 |
| MAST4 | 0.019789045 | -0.150024483 | 0.306 | 0.372 | 1 |
| NAMPT | 0.019951366 | 0.18845366 | 0.986 | 0.983 | 1 |
| IFITM2 | 0.020098429 | 0.126453645 | 0.271 | 0.212 | 1 |
| RBCK1 | 0.020269828 | 0.092426773 | 0.244 | 0.203 | 1 |
| STAT5A | 0.020633635 | -0.125531311 | 0.993 | 0.983 | 1 |
| CCR5 | 0.020991577 | 0.038271003 | 0.048 | 0.021 | 1 |
| CD200 | 0.023550045 | -0.287425684 | 0.467 | 0.484 | 1 |
| BTLA | 0.023753204 | 0.144352778 | 0.268 | 0.205 | 1 |
| CHD1 | 0.025023651 | -0.050323499 | 0.021 | 0.05 | 1 |
| CD109 | 0.026189895 | -0.102351119 | 0.079 | 0.105 | 1 |
| NT5E | 0.027902726 | -0.154644146 | 0.801 | 0.819 | 1 |
| FYB1 | 0.029283938 | -0.148947647 | 0.292 | 0.353 | 1 |
| BACH2 | 0.030251928 | -0.129166593 | 0.498 | 0.534 | 1 |
| IGKC | 0.030659334 | -0.195694194 | 0.213 | 0.142 | 1 |
| NANS | 0.031926947 | -0.138936064 | 0.646 | 0.689 | 1 |
| XBP1 | 0.032732161 | 0.132957338 | 0.66 | 0.58 | 1 |
| IL22 | 0.03340632 | 0.069805949 | 0.065 | 0.045 | 1 |
| BCL2L1 | 0.033459423 | 0.135173298 | 0.515 | 0.424 | 1 |
| TNF | 0.035700735 | -0.158720495 | 0.869 | 0.851 | 1 |
| IL1R1 | 0.036043942 | 0.11138863 | 0.175 | 0.116 | 1 |
| FAS | 0.03726063 | 0.089386318 | 0.192 | 0.107 | 1 |
| IL31 | 0.037706931 | -0.066354074 | 0.007 | 0.018 | 1 |
| IL17F | 0.03994425 | -0.081725598 | 0.017 | 0.03 | 1 |
| POU2AF1 | 0.042117608 | -0.091185927 | 0.113 | 0.146 | 1 |
| CD226 | 0.045411658 | 0.090199333 | 0.155 | 0.109 | 1 |
| THEMIS | 0.04570732 | -0.058434496 | 0.045 | 0.068 | 1 |
| FOXP1 | 0.04833223 | -0.115463514 | 0.567 | 0.609 | 1 |
| CD52 | 0.049118481 | -0.145472791 | 0.512 | 0.552 | 1 |
| CD7 | 0.050648902 | 0.128486138 | 0.656 | 0.641 | 1 |
| CD3D | 0.050667177 | -0.157125632 | 0.962 | 0.953 | 1 |
| IRF9 | 0.051247254 | -0.120430118 | 0.392 | 0.402 | 1 |

| | | | | | |
|---|---|---|---|---|---|
| IL32 | 0.053474333 | 0.139984427 | 0.636 | 0.563 | 1 |
| KLRC3 | 0.053879638 | -0.066138031 | 0.062 | 0.092 | 1 |
| MAP2K3 | 0.054621775 | 0.130932764 | 0.869 | 0.833 | 1 |
| BCL11B | 0.057556631 | 0.138206927 | 0.478 | 0.385 | 1 |
| EOMES | 0.058843314 | 0.026137349 | 0.021 | 0.009 | 1 |
| TNFRSF25 | 0.061281186 | 0.104188943 | 0.488 | 0.463 | 1 |
| NFATC1 | 0.061473953 | -0.125337737 | 0.337 | 0.291 | 1 |
| DPP4 | 0.061934034 | 0.047193882 | 0.055 | 0.022 | 1 |
| PTPRC | 0.064002644 | 0.110132744 | 0.948 | 0.941 | 1 |
| IL13 | 0.064107512 | -0.078455369 | 0.058 | 0.059 | 1 |
| SPP1 | 0.065103552 | -0.028217648 | 0.003 | 0.022 | 1 |
| DUSP6 | 0.065452248 | -0.170670388 | 0.32 | 0.339 | 1 |
| CD3G | 0.068790571 | -0.053770577 | 0.027 | 0.052 | 1 |
| LEF1 | 0.070510157 | 0.155675635 | 0.581 | 0.532 | 1 |
| LIG4 | 0.071419839 | 0.069541507 | 0.206 | 0.159 | 1 |
| SMAD2 | 0.076998821 | 0.061781857 | 0.189 | 0.151 | 1 |
| IL2 | 0.079881787 | 0.221373869 | 1 | 0.999 | 1 |
| PSME3 | 0.081341954 | 0.090758789 | 0.667 | 0.637 | 1 |
| GNAI2 | 0.083221199 | -0.115321384 | 0.46 | 0.499 | 1 |
| IGBP1 | 0.084999822 | -0.065857063 | 0.144 | 0.19 | 1 |
| CXCL3 | 0.085091312 | 0.349602209 | 0.182 | 0.169 | 1 |
| TRBC2 | 0.087991907 | -0.135381512 | 0.969 | 0.986 | 1 |
| UFM1 | 0.09010761 | 0.091190815 | 0.983 | 0.963 | 1 |
| TCF7 | 0.091584556 | -0.065261496 | 0.107 | 0.141 | 1 |
| ITGB2 | 0.091799056 | 0.067965652 | 0.203 | 0.152 | 1 |
| STAT1 | 0.092915533 | 0.11232313 | 0.777 | 0.745 | 1 |
| LAP3 | 0.093431896 | -0.08101711 | 0.306 | 0.351 | 1 |
| FN1 | 0.096719186 | -0.081028812 | 0.089 | 0.119 | 1 |
| TNFRSF4 | 0.097646501 | 0.090128255 | 0.216 | 0.18 | 1 |
| IL4 | 0.10085842 | -0.051564507 | 0.045 | 0.056 | 1 |
| LRRC8B | 0.108437364 | -0.093009815 | 0.543 | 0.588 | 1 |
| PRF1 | 0.108661785 | 0.086390066 | 0.405 | 0.403 | 1 |
| BCAT1 | 0.116150789 | 0.047003 | 0.086 | 0.063 | 1 |
| ARID5A | 0.117054613 | -0.097268322 | 0.543 | 0.541 | 1 |
| MALT1 | 0.118475848 | 0.060174397 | 0.189 | 0.132 | 1 |
| CNOT2 | 0.127999923 | -0.079490594 | 0.43 | 0.465 | 1 |
| KLRG1 | 0.129587159 | 0.011073916 | 0.01 | 0.003 | 1 |
| NGDN | 0.129754438 | 0.038505149 | 0.065 | 0.035 | 1 |
| LIF | 0.134658665 | 0.286073025 | 0.471 | 0.349 | 1 |
| CD79A | 0.138809542 | -0.027211439 | 0.021 | 0.039 | 1 |
| ICAM1 | 0.140880058 | 0.077644465 | 0.412 | 0.372 | 1 |
| TUBA1C | 0.144210651 | -0.085838902 | 0.973 | 0.963 | 1 |
| IL3RA | 0.156627435 | -0.033275431 | 0.038 | 0.056 | 1 |
| CSF2 | 0.163560012 | -0.132913886 | 0.845 | 0.863 | 1 |
| TNFSF8 | 0.166054977 | 0.075565824 | 0.192 | 0.143 | 1 |
| IRF8 | 0.166320438 | 0.122729505 | 0.986 | 0.948 | 1 |
| CD22 | 0.168012015 | -0.030523145 | 0.031 | 0.047 | 1 |

| | | | | | |
|---|---|---|---|---|---|
| ITGAX | 0.179015861 | -0.039685299 | 0.055 | 0.096 | 1 |
| IL21 | 0.180395371 | 0.033158528 | 0.058 | 0.044 | 1 |
| RAPH1 | 0.18040629 | -0.038656876 | 0.045 | 0.065 | 1 |
| ITGAE | 0.181719495 | -0.029258178 | 0.034 | 0.052 | 1 |
| HAVCR2 | 0.184473576 | -0.028751881 | 0.031 | 0.047 | 1 |
| IL15RA | 0.186942178 | 0.085544252 | 0.577 | 0.497 | 1 |
| TNFSF10 | 0.18871506 | 0.013437465 | 0.017 | 0.008 | 1 |
| CD74 | 0.191970799 | 0.091012807 | 0.667 | 0.599 | 1 |
| IL18RAP | 0.193142565 | -0.028488399 | 0.017 | 0.025 | 1 |
| CCL4 | 0.195614393 | -0.078196007 | 1 | 1 | 1 |
| CCL3 | 0.196526161 | -0.078634973 | 1 | 0.995 | 1 |
| EIF3J | 0.197436829 | 0.07034387 | 0.574 | 0.544 | 1 |
| LILRB4 | 0.201022982 | 0.014608737 | 0.021 | 0.01 | 1 |
| ERBIN | 0.203937431 | -0.060306614 | 0.237 | 0.264 | 1 |
| IL6R | 0.206379371 | 0.044532083 | 0.155 | 0.123 | 1 |
| ITK | 0.209767376 | 0.07653449 | 0.986 | 0.968 | 1 |
| CD44 | 0.215286743 | 0.057790966 | 1 | 0.999 | 1 |
| POU2F2 | 0.21893876 | -0.072202824 | 0.536 | 0.553 | 1 |
| CXCL11 | 0.222021297 | -0.013570872 | 0.003 | 0.012 | 1 |
| NKG7 | 0.227418319 | -0.056788316 | 0.244 | 0.274 | 1 |
| CR2 | 0.228030309 | 0.024846066 | 0.041 | 0.032 | 1 |
| KDELR1 | 0.229553483 | 0.044616448 | 0.179 | 0.141 | 1 |
| CD72 | 0.230891081 | 0.00921597 | 0.01 | 0.004 | 1 |
| CHI3L2 | 0.230891081 | 0.00921597 | 0.01 | 0.004 | 1 |
| PIK3AP1 | 0.231532177 | 0.030346541 | 0.079 | 0.059 | 1 |
| IL6 | 0.233207312 | -0.058693946 | 0.251 | 0.284 | 1 |
| ADA | 0.234613981 | -0.045979562 | 0.052 | 0.047 | 1 |
| CD86 | 0.24001693 | 0.020806013 | 0.045 | 0.03 | 1 |
| IGLC3 | 0.245477348 | 0.165246123 | 0.261 | 0.27 | 1 |
| CTSD | 0.246721114 | 0.028594634 | 0.079 | 0.063 | 1 |
| PRDM1 | 0.254223805 | -0.026750015 | 0.038 | 0.056 | 1 |
| ANXA5 | 0.266228685 | -0.06758929 | 0.436 | 0.43 | 1 |
| MMP9 | 0.267411571 | -0.026018126 | 0.048 | 0.066 | 1 |
| B3GAT1 | 0.273074662 | 0.011589041 | 0.017 | 0.009 | 1 |
| CD37 | 0.274332193 | -0.059941514 | 0.45 | 0.456 | 1 |
| FTH1 | 0.278956604 | -0.08890747 | 0.952 | 0.953 | 1 |
| VMO1 | 0.27927736 | 0.021928153 | 0.041 | 0.03 | 1 |
| IGHD | 0.284735342 | 0.013437465 | 0.017 | 0.005 | 1 |
| IFNGR1 | 0.284807503 | 0.054787657 | 0.292 | 0.27 | 1 |
| CD8B | 0.2860437 | -0.030722348 | 0.055 | 0.084 | 1 |
| SOCS2 | 0.295998816 | -0.063525273 | 0.216 | 0.217 | 1 |
| PAK1IP1 | 0.306209188 | -0.05970518 | 0.485 | 0.463 | 1 |
| LCK | 0.312629389 | 0.044943353 | 0.189 | 0.143 | 1 |
| CD80 | 0.315893411 | 0.011589041 | 0.014 | 0.009 | 1 |
| MGST1 | 0.317256296 | 0.00921597 | 0.007 | 0.004 | 1 |
| FCER2 | 0.319260941 | -0.019862789 | 0.031 | 0.043 | 1 |
| KLRC4 | 0.323228439 | -0.01710964 | 0.01 | 0.021 | 1 |

| | | | | | |
|---|---|---|---|---|---|
| ARL4C | 0.324349008 | -0.04023474 | 0.076 | 0.075 | 1 |
| MZB1 | 0.324451613 | -0.009885825 | 0.003 | 0.009 | 1 |
| TNFSF14 | 0.324604727 | 0.070690225 | 0.306 | 0.288 | 1 |
| GAPDH | 0.327337124 | -0.072455868 | 0.966 | 0.951 | 1 |
| HLA.A | 0.327755906 | -0.054241066 | 1 | 1 | 1 |
| RORC | 0.329083692 | -0.012314235 | 0.007 | 0.013 | 1 |
| CTSW | 0.338431233 | -0.026306371 | 0.058 | 0.071 | 1 |
| CST7 | 0.339052743 | 0.040367737 | 0.168 | 0.145 | 1 |
| PRNP | 0.339165536 | -0.067144421 | 0.914 | 0.928 | 1 |
| HNRNPC | 0.340884582 | 0.048744334 | 0.942 | 0.942 | 1 |
| B4GALT5 | 0.343316371 | -0.025088182 | 0.062 | 0.079 | 1 |
| TSPAN32 | 0.348969833 | 0.014424261 | 0.031 | 0.019 | 1 |
| BCL10 | 0.360679919 | 0.031466342 | 0.134 | 0.094 | 1 |
| CD9 | 0.365110278 | -0.035552567 | 0.103 | 0.107 | 1 |
| VEGFA | 0.365385202 | -0.008039766 | 0.003 | 0.009 | 1 |
| LRRC32 | 0.365393645 | 0.01092369 | 0.021 | 0.013 | 1 |
| CD24 | 0.365922952 | 0.007360413 | 0.01 | 0.005 | 1 |
| TIMM9 | 0.372545453 | -0.048969711 | 0.801 | 0.784 | 1 |
| PAXBP1 | 0.384190704 | 0.046889428 | 0.44 | 0.393 | 1 |
| TNFRSF13C | 0.391170086 | 0.057270205 | 0.344 | 0.304 | 1 |
| ZBED2 | 0.39252962 | 0.017190234 | 0.041 | 0.028 | 1 |
| THBS1 | 0.393489371 | -0.021108776 | 0.027 | 0.027 | 1 |
| GZMA | 0.395349358 | 0.01092369 | 0.021 | 0.012 | 1 |
| FUT4 | 0.398585862 | 0.014908102 | 0.034 | 0.026 | 1 |
| CD70 | 0.399036563 | -0.011648327 | 0.014 | 0.022 | 1 |
| SNX8 | 0.400113891 | -0.040498777 | 0.299 | 0.305 | 1 |
| GIMAP2 | 0.401401554 | 0.018553834 | 0.065 | 0.05 | 1 |
| CD247 | 0.408557494 | 0.0252169 | 0.093 | 0.071 | 1 |
| CHD7 | 0.410391173 | 0.029195641 | 0.144 | 0.12 | 1 |
| LAT2 | 0.412716963 | -0.011648327 | 0.014 | 0.021 | 1 |
| AQP9 | 0.415159346 | -0.008638576 | 0.007 | 0.013 | 1 |
| SP140 | 0.415187268 | -0.016844008 | 0.034 | 0.043 | 1 |
| CD28 | 0.416573463 | -0.026160475 | 0.086 | 0.101 | 1 |
| TIAF1 | 0.423820084 | -0.010477576 | 0.007 | 0.01 | 1 |
| TLR2 | 0.430375588 | -0.017936945 | 0.038 | 0.049 | 1 |
| BATF | 0.439852594 | -0.05517085 | 0.509 | 0.488 | 1 |
| IRF3 | 0.440386966 | -0.008638576 | 0.003 | 0.013 | 1 |
| MITF | 0.442170818 | 0.007360413 | 0.007 | 0.005 | 1 |
| IL1A | 0.442705279 | -0.021468636 | 0.041 | 0.037 | 1 |
| LAMP1 | 0.442944291 | -0.03969883 | 0.505 | 0.514 | 1 |
| LYN | 0.445093987 | 0.015857283 | 0.048 | 0.037 | 1 |
| OSM | 0.446367255 | 0.02375765 | 0.072 | 0.062 | 1 |
| ILF2 | 0.451045209 | 0.033033649 | 0.99 | 0.988 | 1 |
| TCL1A | 0.451358025 | -0.006191342 | 0.003 | 0.008 | 1 |
| EIF1 | 0.455182222 | 0.030114154 | 0.997 | 1 | 1 |
| ATF6B | 0.460209531 | 0.022024683 | 0.082 | 0.063 | 1 |
| AHI1 | 0.463362827 | -0.019014738 | 0.045 | 0.05 | 1 |

| | | | | | |
|---|---|---|---|---|---|
| CCR1 | 0.467325195 | 0.022594999 | 0.1 | 0.09 | 1 |
| PDCD1 | 0.479152673 | -0.020959791 | 0.065 | 0.074 | 1 |
| CD36 | 0.490053351 | 0.010263108 | 0.021 | 0.016 | 1 |
| LIPA | 0.492219279 | 0.023912481 | 0.137 | 0.119 | 1 |
| TGFBI | 0.49449489 | 0.02052446 | 0.069 | 0.053 | 1 |
| GATA3 | 0.500791812 | -0.015061802 | 0.024 | 0.04 | 1 |
| LAG3 | 0.502140755 | 0.017441524 | 0.052 | 0.04 | 1 |
| ALAS2 | 0.511317994 | -0.009885825 | 0.003 | 0.004 | 1 |
| PSEN1 | 0.513492267 | -0.013277393 | 0.034 | 0.04 | 1 |
| GAB2 | 0.520996892 | 0.015665899 | 0.058 | 0.043 | 1 |
| S100A12 | 0.522154857 | 0.00550724 | 0.01 | 0.006 | 1 |
| QPCT | 0.522154857 | 0.00550724 | 0.01 | 0.006 | 1 |
| GZMK | 0.530374387 | 0.004300479 | 0.007 | 0.004 | 1 |
| IL12A | 0.533136733 | -0.011568759 | 0.024 | 0.027 | 1 |
| BAX | 0.53410693 | -0.032953797 | 0.515 | 0.523 | 1 |
| GRB10 | 0.535132562 | 0.018105062 | 0.052 | 0.028 | 1 |
| CYCS | 0.538770816 | -0.027661662 | 0.99 | 0.988 | 1 |
| DUSP4 | 0.544306731 | -0.045199722 | 0.323 | 0.306 | 1 |
| TMEM97 | 0.549453542 | -0.022129201 | 0.113 | 0.132 | 1 |
| MCM4 | 0.55761748 | 0.016838236 | 0.076 | 0.074 | 1 |
| BIN2 | 0.564490488 | -0.004340546 | 0.003 | 0.006 | 1 |
| SLC7A7 | 0.564490488 | -0.004340546 | 0.003 | 0.006 | 1 |
| LAMP3 | 0.569051098 | 0.040291769 | 0.419 | 0.397 | 1 |
| BCL6 | 0.570310527 | -0.007398744 | 0.01 | 0.013 | 1 |
| NFKBIA | 0.571100054 | -0.041461161 | 1 | 0.999 | 1 |
| IL12RB2 | 0.571741636 | 0.02647517 | 0.144 | 0.129 | 1 |
| DCLRE1C | 0.580472356 | 0.014832733 | 0.062 | 0.058 | 1 |
| GZMH | 0.588401305 | 0.011134873 | 0.045 | 0.037 | 1 |
| JCHAIN | 0.596029437 | 0.035165927 | 0.062 | 0.04 | 1 |
| FCN1 | 0.605762419 | -0.00495353 | 0.007 | 0.01 | 1 |
| PGM3 | 0.607621407 | -0.011977187 | 0.048 | 0.05 | 1 |
| MS4A1 | 0.611825807 | 0.007782203 | 0.027 | 0.021 | 1 |
| YBX3 | 0.619195943 | 0.030710411 | 0.478 | 0.463 | 1 |
| RORA | 0.623063685 | 0.034173522 | 0.199 | 0.204 | 1 |
| BCL2 | 0.630683603 | -0.02235726 | 0.247 | 0.236 | 1 |
| JAK2 | 0.635247647 | -0.009123074 | 0.027 | 0.035 | 1 |
| ICOS | 0.638114659 | 0.050276671 | 0.766 | 0.599 | 1 |
| IGHM | 0.641533005 | 0.00486076 | 0.014 | 0.01 | 1 |
| ZNF341 | 0.641533005 | 0.00486076 | 0.014 | 0.01 | 1 |
| NFATC3 | 0.649350375 | 0.01899699 | 0.196 | 0.186 | 1 |
| CYBB | 0.65040776 | -0.011339362 | 0.052 | 0.056 | 1 |
| TNFRSF9 | 0.654038339 | -0.010212422 | 0.045 | 0.052 | 1 |
| CXCL10 | 0.659076254 | -0.016275458 | 0.107 | 0.12 | 1 |
| S100A9 | 0.667501148 | 0.00766746 | 0.038 | 0.031 | 1 |
| PSMB8 | 0.668231653 | 0.023510241 | 0.797 | 0.806 | 1 |
| PLCG2 | 0.686813084 | 0.003656444 | 0.01 | 0.008 | 1 |
| FOXP3 | 0.696666406 | -0.00495353 | 0.003 | 0.008 | 1 |

| | | | | | |
|---|---|---|---|---|---|
| ITGAM | 0.699322537 | 0.00486076 | 0.01 | 0.009 | 1 |
| IL2RG | 0.702111577 | 0.021797435 | 0.897 | 0.875 | 1 |
| NFATC2 | 0.703682715 | 0.011335289 | 0.052 | 0.057 | 1 |
| FCER1G | 0.704230904 | -0.015627005 | 0.165 | 0.168 | 1 |
| CD8A | 0.708354916 | -0.031684916 | 0.838 | 0.76 | 1 |
| CCL2 | 0.719382436 | -0.025096737 | 0.553 | 0.57 | 1 |
| TGFB1 | 0.724933105 | 0.004218935 | 0.017 | 0.014 | 1 |
| BIRC3 | 0.73829443 | 0.033522929 | 0.663 | 0.674 | 1 |
| VNN2 | 0.742005959 | 0.002444923 | 0.007 | 0.005 | 1 |
| DOCK8 | 0.74303947 | 0.017035474 | 0.23 | 0.222 | 1 |
| HLA.DRA | 0.743199935 | 0.019125889 | 0.515 | 0.504 | 1 |
| CCR4 | 0.744455526 | -0.022077003 | 0.172 | 0.149 | 1 |
| MCM2 | 0.751651608 | 0.011161151 | 0.162 | 0.145 | 1 |
| LGALS9 | 0.752456538 | 0.005862963 | 0.038 | 0.031 | 1 |
| TYMS | 0.756108435 | -0.00310747 | 0.007 | 0.008 | 1 |
| IL11RA | 0.76672353 | -0.006676403 | 0.045 | 0.049 | 1 |
| NRP1 | 0.778372031 | -0.004928849 | 0.024 | 0.03 | 1 |
| SOCS1 | 0.785344557 | 0.002444923 | 0.003 | 0.005 | 1 |
| LGALS3 | 0.785695202 | 0.006297717 | 0.045 | 0.048 | 1 |
| CD40 | 0.793683682 | 0.010530898 | 0.192 | 0.182 | 1 |
| FOXO1 | 0.797253936 | 0.010959671 | 0.189 | 0.178 | 1 |
| FCGR3A | 0.804185169 | -0.003723086 | 0.021 | 0.022 | 1 |
| IER3 | 0.804968803 | 0.02900089 | 0.931 | 0.911 | 1 |
| SLC25A37 | 0.806356921 | -0.013183415 | 0.354 | 0.366 | 1 |
| CASP5 | 0.813613425 | -0.004322088 | 0.034 | 0.037 | 1 |
| CD48 | 0.821626571 | 0.015183599 | 0.99 | 0.988 | 1 |
| NFKB2 | 0.830641507 | 0.012324855 | 0.357 | 0.359 | 1 |
| CXCL16 | 0.832514495 | -0.006004027 | 0.082 | 0.081 | 1 |
| ZAP70 | 0.835677715 | 0.00721476 | 0.124 | 0.112 | 1 |
| STAT4 | 0.837073739 | 0.013575885 | 0.897 | 0.908 | 1 |
| CCL22 | 0.838291829 | 0.008314708 | 0.151 | 0.151 | 1 |
| CD300A | 0.843340534 | -0.002499945 | 0.014 | 0.014 | 1 |
| CXCL5 | 0.852412872 | 0.008701658 | 0.258 | 0.233 | 1 |
| TRAT1 | 0.857706433 | 0.004524274 | 0.048 | 0.043 | 1 |
| SRGN | 0.859623018 | -0.010867277 | 0.89 | 0.899 | 1 |
| IL9 | 0.875544343 | 0.00406072 | 0.031 | 0.022 | 1 |
| IL3 | 0.880077555 | 0.005740894 | 0.089 | 0.085 | 1 |
| CXCL1 | 0.880554216 | -0.003723086 | 0.048 | 0.045 | 1 |
| CXCL9 | 0.891774193 | 0.003297342 | 0.058 | 0.054 | 1 |
| HLA.DQB1 | 0.895181341 | 0.008349746 | 0.234 | 0.244 | 1 |
| VSIR | 0.899028296 | 0.009869996 | 0.784 | 0.758 | 1 |
| SLC7A1 | 0.900323361 | 0.007080366 | 0.814 | 0.76 | 1 |
| EBF1 | 0.901933646 | -0.00190035 | 0.021 | 0.019 | 1 |
| LTB | 0.906805797 | 0.001752044 | 0.017 | 0.018 | 1 |
| CD14 | 0.90703339 | 0.001752044 | 0.021 | 0.017 | 1 |
| PTGDR2 | 0.9169823 | -0.000631816 | 0.003 | 0.004 | 1 |
| TREM1 | 0.9169823 | -0.000631816 | 0.003 | 0.004 | 1 |

| | | | | | |
|---|---|---|---|---|---|
| NINJ2 | 0.9169823 | -0.000631816 | 0.003 | 0.004 | 1 |
| WAS | 0.918711121 | -0.002559153 | 0.055 | 0.065 | 1 |
| IRF1 | 0.922688749 | -0.007277049 | 0.9 | 0.836 | 1 |
| IL1B | 0.923026685 | 0.006057935 | 0.416 | 0.398 | 1 |
| TNFAIP8 | 0.930630557 | 0.006321963 | 0.99 | 0.961 | 1 |
| GNLY | 0.936699938 | -0.003216433 | 0.21 | 0.189 | 1 |
| LTA | 0.936825532 | -0.005747922 | 0.433 | 0.477 | 1 |
| IFI6 | 0.938978164 | -0.001308533 | 0.027 | 0.026 | 1 |
| HLA.DMA | 0.940346607 | 0.001590714 | 0.048 | 0.05 | 1 |
| CD4 | 0.94096897 | 0.00059175 | 0.007 | 0.006 | 1 |
| CARD11 | 0.94096897 | 0.00059175 | 0.007 | 0.006 | 1 |
| CD163 | 0.94096897 | 0.00059175 | 0.007 | 0.006 | 1 |
| CCL5 | 0.943017542 | -0.003169799 | 0.093 | 0.076 | 1 |
| LY86 | 0.949220933 | 0.00059175 | 0.003 | 0.006 | 1 |
| TGFB3 | 0.949220934 | 0.00059175 | 0.007 | 0.005 | 1 |
| SOCS3 | 0.951285342 | 0.003702021 | 0.443 | 0.397 | 1 |
| SKAP1 | 0.954448719 | -0.000663286 | 0.014 | 0.014 | 1 |
| CXCR5 | 0.954674422 | 0.002455489 | 0.168 | 0.164 | 1 |
| IL5 | 0.957280401 | 0.00059175 | 0.007 | 0.004 | 1 |
| CX3CR1 | 0.965537127 | 0.000547953 | 0.017 | 0.017 | 1 |
| CD63 | 0.967666952 | 0.002358745 | 0.615 | 0.612 | 1 |
| HLA.DPA1 | 0.967864792 | -0.001777442 | 0.213 | 0.229 | 1 |
| TNFRSF8 | 0.969082801 | 0.000547953 | 0.014 | 0.016 | 1 |
| PCNA | 0.97421298 | -0.001722399 | 0.385 | 0.385 | 1 |
| PMCH | 0.994249832 | -0.016678741 | 0 | 0.009 | 1 |
| CD1C | 0.994287777 | -0.014835041 | 0 | 0.01 | 1 |
| IL12RB1 | 0.994380458 | -0.011140558 | 0 | 0.008 | 1 |
| CLEC4E | 0.996186734 | 0.014797002 | 0.01 | 0 | 1 |
| SNCA | 0.996204493 | -0.029519087 | 0 | 0.005 | 1 |
| CD160 | 0.996475382 | -0.007436589 | 0 | 0.005 | 1 |
| RNASE6 | 0.996475382 | -0.007436589 | 0 | 0.005 | 1 |
| TCF4 | 0.996531597 | -0.005581032 | 0 | 0.004 | 1 |

CD8+ Memory

| | p_val | avg_log2FC | Patient | Controls | p_val_adj |
|---|---|---|---|---|---|
| TNF | 2.24E-165 | -1.750690731 | 0.947 | 0.973 | 1.12E-162 |
| GZMH | 6.57E-153 | 1.390854627 | 0.721 | 0.437 | 3.27E-150 |
| S100A10 | 8.11E-114 | 1.027642144 | 0.926 | 0.884 | 4.04E-111 |
| TGIF1 | 2.40E-85 | -0.728161648 | 0.54 | 0.778 | 1.20E-82 |
| GNLY | 3.32E-83 | 1.391519664 | 0.788 | 0.704 | 1.65E-80 |
| SLA | 1.61E-81 | -0.737424093 | 0.749 | 0.883 | 8.00E-79 |
| NR4A2 | 8.11E-79 | -0.972369321 | 0.498 | 0.722 | 4.04E-76 |
| IFNG | 2.50E-77 | -1.241846178 | 0.998 | 0.998 | 1.25E-74 |
| PFN1 | 1.20E-76 | 0.616782676 | 1 | 1 | 5.97E-74 |
| PRNP | 1.55E-75 | -0.677242239 | 0.767 | 0.882 | 7.70E-73 |
| DUSP2 | 7.84E-75 | -0.803301889 | 0.953 | 0.984 | 3.90E-72 |

| | | | | | |
|---|---|---|---|---|---|
| EVI2A | 6.93E-71 | -0.698432457 | 0.609 | 0.744 | 3.45E-68 |
| TRAC | 3.91E-70 | 0.859481761 | 0.909 | 0.792 | 1.95E-67 |
| FYN | 1.96E-68 | -0.546266772 | 0.835 | 0.924 | 9.78E-66 |
| IL2RG | 1.06E-65 | 0.513129428 | 0.978 | 0.937 | 5.29E-63 |
| FCGR3A | 6.70E-64 | 0.614134622 | 0.237 | 0.089 | 3.34E-61 |
| FASLG | 1.77E-63 | -0.685160555 | 0.663 | 0.832 | 8.82E-61 |
| IVNS1ABP | 4.29E-61 | 0.593699739 | 0.764 | 0.622 | 2.14E-58 |
| CD74 | 1.23E-56 | 0.812653892 | 0.851 | 0.733 | 6.12E-54 |
| ITGA4 | 1.91E-56 | 0.475403459 | 0.42 | 0.198 | 9.50E-54 |
| CD83 | 2.90E-55 | -0.827505636 | 0.695 | 0.809 | 1.44E-52 |
| CD72 | 1.17E-51 | -0.457199 | 0.027 | 0.219 | 5.82E-49 |
| CD3D | 5.84E-51 | 0.64088596 | 0.89 | 0.84 | 2.91E-48 |
| CSF2 | 8.71E-48 | -1.030658694 | 0.953 | 0.97 | 4.34E-45 |
| ICAM1 | 1.00E-46 | -0.719764839 | 0.547 | 0.668 | 4.98E-44 |
| TRIB2 | 3.85E-45 | 0.503323978 | 0.356 | 0.173 | 1.92E-42 |
| CST7 | 6.89E-45 | 0.57678709 | 0.896 | 0.814 | 3.43E-42 |
| CD52 | 8.88E-45 | 0.621311093 | 0.759 | 0.632 | 4.42E-42 |
| GNAI2 | 1.63E-44 | 0.535681066 | 0.79 | 0.631 | 8.13E-42 |
| ARPC1B | 1.97E-43 | 0.439863798 | 0.549 | 0.366 | 9.79E-41 |
| CCL4 | 3.51E-43 | -0.586477152 | 1 | 1 | 1.75E-40 |
| CD48 | 7.86E-42 | -0.55302077 | 0.879 | 0.954 | 3.91E-39 |
| KLRG1 | 9.79E-42 | 0.316378943 | 0.229 | 0.071 | 4.88E-39 |
| IL2RB | 5.60E-41 | 0.543271062 | 0.936 | 0.811 | 2.79E-38 |
| REL | 1.85E-39 | -0.444530757 | 0.904 | 0.964 | 9.19E-37 |
| IL32 | 3.18E-38 | 0.447528285 | 0.517 | 0.347 | 1.59E-35 |
| CD160 | 3.22E-38 | -0.345873304 | 0.02 | 0.168 | 1.60E-35 |
| IL18RAP | 2.28E-37 | -0.385279377 | 0.102 | 0.269 | 1.13E-34 |
| ARHGEF3 | 6.75E-37 | -0.433746084 | 0.331 | 0.51 | 3.36E-34 |
| PTPN7 | 1.41E-36 | 0.443003957 | 0.67 | 0.513 | 7.02E-34 |
| CD8A | 6.73E-36 | 0.700680154 | 0.563 | 0.36 | 3.35E-33 |
| CD226 | 2.50E-35 | 0.39496446 | 0.583 | 0.367 | 1.25E-32 |
| BTG1 | 1.05E-34 | -0.39680992 | 0.998 | 0.998 | 5.22E-32 |
| LTA | 4.69E-34 | -0.808718357 | 0.514 | 0.608 | 2.34E-31 |
| RGS1 | 5.74E-34 | 0.753009982 | 0.76 | 0.562 | 2.86E-31 |
| CD5 | 7.68E-34 | 0.419388201 | 0.27 | 0.148 | 3.82E-31 |
| IRF4 | 1.11E-31 | 0.463866856 | 0.977 | 0.957 | 5.51E-29 |
| NFKBIA | 3.46E-31 | -0.483109539 | 0.998 | 0.998 | 1.72E-28 |
| B3GAT1 | 4.64E-30 | 0.33912745 | 0.398 | 0.226 | 2.31E-27 |
| BCAT1 | 4.82E-28 | 0.359273521 | 0.278 | 0.151 | 2.40E-25 |
| KLRC1 | 1.90E-27 | -0.586891069 | 0.303 | 0.457 | 9.44E-25 |
| CX3CR1 | 1.22E-26 | 0.573687614 | 0.426 | 0.266 | 6.08E-24 |
| CD40LG | 1.93E-26 | -0.459838073 | 0.09 | 0.181 | 9.61E-24 |
| FOSB | 4.98E-26 | -0.497276148 | 0.233 | 0.408 | 2.48E-23 |
| JAK3 | 5.52E-26 | 0.285828596 | 0.505 | 0.349 | 2.75E-23 |
| NFKBID | 1.80E-24 | -0.209591605 | 0.063 | 0.182 | 8.98E-22 |
| TRBC2 | 7.51E-24 | 0.4420246 | 0.972 | 0.953 | 3.74E-21 |
| IRF8 | 7.78E-24 | -0.501779664 | 0.872 | 0.924 | 3.87E-21 |

| | | | | | |
|---|---|---|---|---|---|
| TNFRSF4 | 1.42E-23 | 0.476087806 | 0.31 | 0.237 | 7.06E-21 |
| BCL2 | 6.33E-22 | 0.255211135 | 0.411 | 0.278 | 3.15E-19 |
| LCK | 5.39E-21 | 0.233852689 | 0.233 | 0.125 | 2.68E-18 |
| GBP1 | 1.83E-19 | 0.432087878 | 0.416 | 0.341 | 9.09E-17 |
| IL6ST | 1.83E-19 | -0.279667959 | 0.23 | 0.368 | 9.10E-17 |
| SDCBP | 2.26E-19 | -0.32830655 | 0.864 | 0.879 | 1.12E-16 |
| HLA.A | 5.46E-19 | 0.232441541 | 1 | 1 | 2.72E-16 |
| IL1RN | 1.07E-18 | -0.449035509 | 0.214 | 0.259 | 5.34E-16 |
| ITK | 1.15E-18 | 0.330660885 | 0.925 | 0.87 | 5.73E-16 |
| JAK1 | 4.13E-18 | 0.311073217 | 0.916 | 0.835 | 2.06E-15 |
| SOCS2 | 1.19E-17 | 0.345873305 | 0.273 | 0.163 | 5.90E-15 |
| CSF1 | 1.65E-17 | 0.572712541 | 0.483 | 0.41 | 8.23E-15 |
| TBX21 | 4.58E-17 | 0.373412194 | 0.772 | 0.68 | 2.28E-14 |
| CD70 | 1.25E-16 | 0.219418826 | 0.119 | 0.051 | 6.20E-14 |
| CXCR3 | 2.71E-16 | 0.36626221 | 0.653 | 0.583 | 1.35E-13 |
| BIRC3 | 4.22E-16 | -0.366968183 | 0.89 | 0.906 | 2.10E-13 |
| TIGIT | 7.03E-16 | 0.289841202 | 0.367 | 0.246 | 3.50E-13 |
| GZMB | 8.02E-16 | 0.439129063 | 0.994 | 0.995 | 4.00E-13 |
| IL18R1 | 1.69E-15 | -0.194561036 | 0.107 | 0.192 | 8.41E-13 |
| APOBEC3G | 3.74E-15 | 0.26578881 | 0.974 | 0.965 | 1.86E-12 |
| RUNX3 | 4.12E-15 | 0.280074769 | 0.743 | 0.656 | 2.05E-12 |
| DUSP6 | 4.49E-15 | -0.348323018 | 0.269 | 0.395 | 2.24E-12 |
| IL2RA | 6.71E-15 | 0.290320907 | 0.629 | 0.458 | 3.34E-12 |
| IL7R | 7.19E-15 | -0.2777565 | 0.187 | 0.337 | 3.58E-12 |
| VPS28 | 8.82E-15 | 0.193801904 | 0.345 | 0.232 | 4.39E-12 |
| ITGB2 | 1.57E-14 | 0.26339256 | 0.338 | 0.231 | 7.79E-12 |
| EVI2B | 2.19E-14 | -0.28694993 | 0.621 | 0.671 | 1.09E-11 |
| CD44 | 9.25E-14 | 0.216954699 | 1 | 0.998 | 4.60E-11 |
| STAT5A | 1.06E-13 | -0.225166065 | 0.97 | 0.977 | 5.29E-11 |
| STAT3 | 2.59E-13 | 0.240562863 | 0.837 | 0.757 | 1.29E-10 |
| NFATC2 | 7.46E-13 | 0.194303913 | 0.33 | 0.207 | 3.71E-10 |
| IL3 | 1.55E-12 | -1.170795882 | 0.122 | 0.152 | 7.71E-10 |
| LGALS1 | 1.57E-12 | 0.390321739 | 0.655 | 0.555 | 7.80E-10 |
| RORA | 2.01E-12 | 0.306333526 | 0.446 | 0.345 | 9.99E-10 |
| CSRNP1 | 3.99E-12 | -0.219580478 | 0.899 | 0.93 | 1.99E-09 |
| DUSP4 | 5.79E-12 | 0.298443197 | 0.822 | 0.727 | 2.88E-09 |
| CD63 | 7.66E-12 | 0.228065153 | 0.812 | 0.739 | 3.81E-09 |
| IL31 | 2.10E-11 | -0.295823084 | 0.007 | 0.024 | 1.05E-08 |
| LEF1 | 2.85E-11 | 0.232291643 | 0.185 | 0.142 | 1.42E-08 |
| PASK | 6.68E-11 | 0.178684462 | 0.064 | 0.043 | 3.33E-08 |
| PDIA6 | 1.08E-10 | 0.228966121 | 0.854 | 0.831 | 5.39E-08 |
| NFATC1 | 1.47E-10 | -0.162594539 | 0.188 | 0.275 | 7.34E-08 |
| IGKC | 3.48E-10 | -0.71972128 | 0.182 | 0.196 | 1.74E-07 |
| IFITM2 | 1.05E-09 | 0.221465177 | 0.381 | 0.283 | 5.21E-07 |
| EGR3 | 1.10E-09 | -0.141211612 | 0.104 | 0.17 | 5.46E-07 |
| ZNF683 | 1.64E-09 | 0.086890364 | 0.086 | 0.031 | 8.17E-07 |
| CD38 | 1.73E-09 | 0.07532662 | 0.047 | 0.015 | 8.61E-07 |

| | | | | | |
|---|---|---|---|---|---|
| IRF1 | 2.87E-09 | 0.261944476 | 0.636 | 0.559 | 1.43E-06 |
| TGFBR2 | 3.21E-09 | 0.20290618 | 0.734 | 0.664 | 1.60E-06 |
| NFKB1 | 8.49E-09 | -0.170001402 | 0.996 | 0.997 | 4.23E-06 |
| BACH2 | 8.51E-09 | -0.164796833 | 0.245 | 0.336 | 4.24E-06 |
| CD2 | 1.42E-08 | 0.296056657 | 0.782 | 0.769 | 7.09E-06 |
| CCL20 | 2.33E-08 | -0.685594833 | 0.483 | 0.534 | 1.16E-05 |
| RORC | 2.39E-08 | -0.187062885 | 0.032 | 0.081 | 1.19E-05 |
| ARID5A | 2.59E-08 | 0.151268309 | 0.409 | 0.32 | 1.29E-05 |
| DOCK8 | 3.07E-08 | 0.143976052 | 0.35 | 0.25 | 1.53E-05 |
| STAT1 | 3.28E-08 | 0.217674223 | 0.441 | 0.384 | 1.63E-05 |
| ITGAX | 3.58E-08 | 0.217549221 | 0.277 | 0.196 | 1.79E-05 |
| PSMB9 | 5.25E-08 | 0.162911423 | 0.456 | 0.346 | 2.62E-05 |
| FAS | 1.02E-07 | 0.120475276 | 0.229 | 0.151 | 5.09E-05 |
| LAG3 | 1.03E-07 | 0.275861364 | 0.482 | 0.436 | 5.12E-05 |
| KLRF1 | 1.12E-07 | -0.072048309 | 0.022 | 0.057 | 5.60E-05 |
| KLRC3 | 1.77E-07 | -0.277796524 | 0.498 | 0.545 | 8.83E-05 |
| IL4 | 1.79E-07 | -0.505113424 | 0.061 | 0.084 | 8.90E-05 |
| TRDC | 1.82E-07 | 0.270897108 | 0.208 | 0.195 | 9.07E-05 |
| FCER1G | 1.87E-07 | -0.159461146 | 0.189 | 0.237 | 9.32E-05 |
| FOSL1 | 1.90E-07 | -0.187739701 | 0.456 | 0.552 | 9.47E-05 |
| CREM | 2.20E-07 | -0.200993591 | 0.651 | 0.718 | 0.000109437 |
| ORAI1 | 2.26E-07 | -0.163896589 | 0.541 | 0.594 | 0.000112542 |
| ITGAM | 2.42E-07 | 0.096441561 | 0.123 | 0.071 | 0.000120565 |
| CXCR4 | 2.89E-07 | -0.263756363 | 0.236 | 0.315 | 0.000143989 |
| CD37 | 3.36E-07 | 0.140158138 | 0.498 | 0.429 | 0.000167468 |
| ICOS | 3.45E-07 | 0.339060331 | 0.494 | 0.465 | 0.000171616 |
| TNFRSF8 | 3.92E-07 | 0.048715635 | 0.037 | 0.012 | 0.000195254 |
| IL13 | 4.03E-07 | -0.568525695 | 0.107 | 0.126 | 0.000200864 |
| LDHA | 5.13E-07 | -0.16088438 | 0.998 | 0.996 | 0.000255241 |
| TNIK | 6.79E-07 | -0.108837907 | 0.129 | 0.202 | 0.000338095 |
| DUSP5 | 7.65E-07 | -0.177298293 | 0.933 | 0.934 | 0.000380793 |
| ZBTB16 | 8.25E-07 | -0.047386055 | 0.006 | 0.035 | 0.000410703 |
| CD3E | 1.03E-06 | 0.188450757 | 0.807 | 0.735 | 0.000515284 |
| HLA.DPA1 | 1.36E-06 | 0.165334133 | 0.459 | 0.369 | 0.000678974 |
| STAT6 | 1.66E-06 | 0.122809938 | 0.383 | 0.322 | 0.000828832 |
| ZBED2 | 1.93E-06 | -0.224358625 | 0.047 | 0.086 | 0.000962811 |
| IRF7 | 2.04E-06 | -0.160012539 | 0.31 | 0.335 | 0.001014436 |
| CCL22 | 3.12E-06 | -0.149047286 | 0.173 | 0.197 | 0.001554758 |
| CCL2 | 3.58E-06 | -0.217509707 | 0.587 | 0.582 | 0.001782133 |
| SELPLG | 3.76E-06 | 0.100816967 | 0.137 | 0.075 | 0.001870348 |
| TUBA1C | 3.83E-06 | -0.173355182 | 0.857 | 0.865 | 0.001908008 |
| VSIR | 3.99E-06 | -0.156647006 | 0.874 | 0.927 | 0.001989068 |
| MYB | 4.15E-06 | 0.074009975 | 0.057 | 0.028 | 0.002064342 |
| GZMK | 4.45E-06 | 0.121578089 | 0.067 | 0.062 | 0.002213811 |
| CTSW | 6.37E-06 | 0.130644762 | 0.328 | 0.242 | 0.003170394 |
| PTPRC | 6.58E-06 | 0.152244702 | 0.902 | 0.882 | 0.003275689 |
| TNFRSF13C | 7.67E-06 | 0.143781892 | 0.288 | 0.247 | 0.003818518 |

| | | | | | |
|---|---|---|---|---|---|
| GZMA | 8.12E-06 | -0.26417939 | 0.163 | 0.175 | 0.004045208 |
| HAVCR2 | 8.44E-06 | 0.258192971 | 0.314 | 0.25 | 0.004202934 |
| PRF1 | 9.51E-06 | -0.21656296 | 0.918 | 0.935 | 0.004738207 |
| IL23A | 1.07E-05 | -0.296592073 | 0.4 | 0.451 | 0.005320498 |
| FOS | 1.32E-05 | -0.242966642 | 0.468 | 0.485 | 0.006581631 |
| CCL5 | 1.34E-05 | -0.178062749 | 0.858 | 0.855 | 0.006695567 |
| PIK3AP1 | 1.39E-05 | 0.148237196 | 0.435 | 0.342 | 0.006906094 |
| MYC | 1.42E-05 | -0.247409074 | 0.717 | 0.778 | 0.007088333 |
| NINJ2 | 1.63E-05 | 0.033691657 | 0.02 | 0.007 | 0.008134335 |
| RBCK1 | 1.65E-05 | 0.100342627 | 0.3 | 0.233 | 0.008221831 |
| ADA | 2.35E-05 | -0.089886081 | 0.066 | 0.113 | 0.011710767 |
| LAT2 | 2.68E-05 | 0.105317575 | 0.196 | 0.137 | 0.013327223 |
| CD28 | 3.68E-05 | 0.073116441 | 0.072 | 0.052 | 0.018317839 |
| PSMB8 | 3.69E-05 | 0.135972644 | 0.694 | 0.627 | 0.018367669 |
| SPAG9 | 3.83E-05 | -0.11844704 | 0.36 | 0.425 | 0.019056967 |
| UGCG | 4.34E-05 | 0.091920293 | 0.224 | 0.174 | 0.021617857 |
| IL17F | 4.68E-05 | 0.263146421 | 0.043 | 0.038 | 0.023329333 |
| CYCS | 5.77E-05 | -0.147686259 | 0.857 | 0.873 | 0.028723518 |
| ABCE1 | 6.03E-05 | -0.131615376 | 0.7 | 0.766 | 0.030045013 |
| UFM1 | 6.62E-05 | -0.130579821 | 0.914 | 0.918 | 0.032974863 |
| CXCL10 | 6.71E-05 | 0.207702594 | 0.146 | 0.155 | 0.033420663 |
| LAT | 7.33E-05 | 0.177575143 | 0.498 | 0.413 | 0.036499768 |
| IFI6 | 9.76E-05 | -0.05005182 | 0.019 | 0.036 | 0.048612601 |
| CTSD | 0.000104569 | -0.059547569 | 0.066 | 0.099 | 0.052075435 |
| CD3G | 0.000111981 | 0.032598131 | 0.043 | 0.02 | 0.055766783 |
| LIG4 | 0.000133795 | 0.064238644 | 0.15 | 0.103 | 0.066629912 |
| LRRC8B | 0.000133884 | 0.1232333 | 0.477 | 0.442 | 0.066674477 |
| ANXA5 | 0.000149853 | 0.15380351 | 0.783 | 0.729 | 0.074626895 |
| JAK2 | 0.000163724 | -0.040524397 | 0.022 | 0.051 | 0.081534573 |
| IL15RA | 0.000171212 | 0.121616025 | 0.425 | 0.373 | 0.085263591 |
| IL6 | 0.000185388 | -0.154067451 | 0.278 | 0.312 | 0.092323387 |
| TGFBR1 | 0.000201993 | -0.076941119 | 0.158 | 0.205 | 0.100592586 |
| PCNA | 0.000210657 | -0.086289509 | 0.209 | 0.255 | 0.104907423 |
| NT5E | 0.000223319 | -0.138696921 | 0.151 | 0.212 | 0.111212663 |
| GAPDH | 0.000235487 | -0.128585295 | 0.986 | 0.987 | 0.117272687 |
| THBS1 | 0.000240815 | 0.17117698 | 0.156 | 0.12 | 0.11992598 |
| WAS | 0.000241468 | 0.071813373 | 0.169 | 0.133 | 0.120251169 |
| HLA.DRA | 0.000267795 | 0.156192471 | 0.685 | 0.638 | 0.133361921 |
| EIF5 | 0.000327819 | -0.115380552 | 0.745 | 0.793 | 0.163254016 |
| PDIA4 | 0.000336202 | 0.122598993 | 0.511 | 0.453 | 0.167428364 |
| IFNGR1 | 0.000360786 | -0.097827566 | 0.299 | 0.351 | 0.179671424 |
| HNRNPC | 0.00039562 | -0.116200363 | 0.812 | 0.845 | 0.197018599 |
| PRDM1 | 0.000435536 | 0.12579895 | 0.456 | 0.394 | 0.216896915 |
| IL1B | 0.000439317 | -0.138371651 | 0.432 | 0.438 | 0.218779792 |
| CD40 | 0.000466442 | -0.123708053 | 0.223 | 0.218 | 0.232288182 |
| IRF9 | 0.000522125 | 0.115974585 | 0.48 | 0.408 | 0.260018204 |
| EIF1 | 0.000530257 | 0.101042666 | 0.997 | 0.998 | 0.264068117 |

| | | | | | |
|---|---|---|---|---|---|
| CXCL11 | 0.000589742 | 0.15882486 | 0.011 | 0.013 | 0.293691653 |
| IL12RB2 | 0.000590529 | -0.132091475 | 0.162 | 0.246 | 0.294083479 |
| CXCL5 | 0.000672081 | -0.08636904 | 0.224 | 0.26 | 0.334696561 |
| IL2 | 0.000756073 | -0.240403413 | 1 | 1 | 0.376524598 |
| RAPH1 | 0.000786749 | -0.0384299 | 0.025 | 0.049 | 0.391801236 |
| SP140 | 0.000801223 | -0.069468583 | 0.125 | 0.161 | 0.399009248 |
| FOXP1 | 0.000832556 | 0.100576146 | 0.472 | 0.429 | 0.414612992 |
| LAMP1 | 0.000952549 | -0.098481515 | 0.484 | 0.521 | 0.474369242 |
| TNFSF14 | 0.000979048 | 0.105489876 | 0.367 | 0.345 | 0.487565894 |
| SKAP1 | 0.00099349 | 0.0208185 | 0.023 | 0.01 | 0.494758164 |
| MAP2K3 | 0.001001246 | 0.098517091 | 0.978 | 0.969 | 0.498620752 |
| LYN | 0.00107513 | -0.065659382 | 0.106 | 0.136 | 0.53541479 |
| B4GALT5 | 0.001102451 | 0.057750613 | 0.156 | 0.123 | 0.549020402 |
| NKG7 | 0.001302556 | -0.141683005 | 0.909 | 0.922 | 0.648672672 |
| LIF | 0.001511714 | -0.356388072 | 0.23 | 0.286 | 0.752833523 |
| ERBIN | 0.001618545 | 0.096044218 | 0.536 | 0.488 | 0.806035522 |
| NFKBIZ | 0.001691731 | -0.137965574 | 0.643 | 0.651 | 0.842482273 |
| TRAF1 | 0.00228821 | -0.103882226 | 0.72 | 0.746 | 1 |
| CCR7 | 0.00345019 | -0.125683674 | 0.436 | 0.483 | 1 |
| QPCT | 0.004492202 | -0.023744321 | 0.016 | 0.031 | 1 |
| CD244 | 0.00524347 | -0.029818318 | 0.02 | 0.035 | 1 |
| PLCG2 | 0.005310043 | -0.028818236 | 0.025 | 0.042 | 1 |
| CBLB | 0.005383312 | -0.041884225 | 0.062 | 0.09 | 1 |
| STAT4 | 0.006473702 | -0.094363726 | 0.786 | 0.776 | 1 |
| GATA3 | 0.007244146 | -0.025471976 | 0.02 | 0.034 | 1 |
| DUSP1 | 0.009183912 | -0.088797149 | 0.331 | 0.342 | 1 |
| IL12RB1 | 0.011297609 | -0.018702989 | 0.011 | 0.021 | 1 |
| CD109 | 0.011541746 | -0.062355931 | 0.116 | 0.138 | 1 |
| CD24 | 0.012243175 | -0.015169148 | 0.003 | 0.011 | 1 |
| TLR2 | 0.012469262 | -0.02736908 | 0.033 | 0.048 | 1 |
| IL22 | 0.012654719 | -0.309874707 | 0.056 | 0.072 | 1 |
| CD80 | 0.014527785 | -0.015269605 | 0.006 | 0.016 | 1 |
| IL1A | 0.015036716 | -0.080774579 | 0.059 | 0.066 | 1 |
| CXCL8 | 0.015348327 | -0.123031668 | 0.895 | 0.893 | 1 |
| VEGFA | 0.017759213 | -0.015219453 | 0.005 | 0.012 | 1 |
| MCM4 | 0.019306779 | -0.022945215 | 0.025 | 0.041 | 1 |
| AHI1 | 0.022496673 | -0.033075331 | 0.051 | 0.071 | 1 |
| NCAM1 | 0.026972535 | -0.031170947 | 0.029 | 0.045 | 1 |
| FBXO11 | 0.02996802 | -0.056333821 | 0.333 | 0.351 | 1 |
| XBP1 | 0.032382706 | -0.066993873 | 0.718 | 0.708 | 1 |
| ILF2 | 0.033662973 | -0.068450097 | 0.891 | 0.907 | 1 |
| SLC25A37 | 0.034628393 | -0.045195888 | 0.127 | 0.15 | 1 |
| MMP9 | 0.037257614 | -0.028986091 | 0.059 | 0.073 | 1 |
| CXCL2 | 0.043056147 | -0.06343348 | 0.351 | 0.385 | 1 |
| FURIN | 0.043770823 | -0.080401608 | 0.662 | 0.68 | 1 |
| KLRB1 | 0.043858238 | -0.009023178 | 0.002 | 0.007 | 1 |
| IFRD1 | 0.045008636 | -0.05013321 | 0.215 | 0.25 | 1 |

| | | | | | |
|---|---|---|---|---|---|
| CD200 | 0.047998082 | -0.149561738 | 0.303 | 0.33 | 1 |
| MCM2 | 0.049177297 | -0.022824529 | 0.038 | 0.056 | 1 |
| IL5 | 0.065363161 | -0.021919702 | 0.009 | 0.008 | 1 |
| BCL6 | 0.074785498 | -0.011550799 | 0.009 | 0.017 | 1 |
| RPL6 | 0.075194849 | -0.056425377 | 1 | 1 | 1 |
| TGFB1 | 0.075377573 | -0.017617589 | 0.027 | 0.037 | 1 |
| TMEM97 | 0.084067219 | -0.018931612 | 0.032 | 0.046 | 1 |
| IL3RA | 0.091572967 | -0.03383008 | 0.056 | 0.064 | 1 |
| SLC7A1 | 0.106108018 | -0.051468447 | 0.676 | 0.696 | 1 |
| JCHAIN | 0.107803867 | -0.101463501 | 0.065 | 0.054 | 1 |
| CXCL1 | 0.126082407 | -0.02254101 | 0.041 | 0.054 | 1 |
| THBD | 0.138239394 | -0.00454044 | 0.002 | 0.005 | 1 |
| IGHA1 | 0.157765141 | -0.010191472 | 0.004 | 0.004 | 1 |
| RPN2 | 0.179741432 | -0.040930354 | 0.61 | 0.641 | 1 |
| FCN1 | 0.181402284 | -0.008828992 | 0.011 | 0.015 | 1 |
| IGHG2 | 0.193599073 | -0.004607199 | 0.003 | 0.006 | 1 |
| NRP1 | 0.201330056 | -0.015001573 | 0.028 | 0.03 | 1 |
| CNOT2 | 0.205650965 | -0.032466511 | 0.351 | 0.355 | 1 |
| PAK1IP1 | 0.211669144 | -0.034670353 | 0.288 | 0.305 | 1 |
| KLRC4 | 0.217590838 | -0.016411768 | 0.054 | 0.059 | 1 |
| GIMAP2 | 0.228525957 | -0.019093903 | 0.084 | 0.1 | 1 |
| PTGDR2 | 0.228877063 | -0.002832352 | 0.001 | 0.003 | 1 |
| HIF1A | 0.234723078 | -0.039232237 | 0.673 | 0.692 | 1 |
| CCL3 | 0.238202824 | -0.05470564 | 0.999 | 1 | 1 |
| IL12A | 0.242605769 | -0.007850015 | 0.014 | 0.018 | 1 |
| SLC7A7 | 0.248582685 | -0.004640503 | 0.004 | 0.006 | 1 |
| MS4A1 | 0.251572491 | -0.012095743 | 0.038 | 0.045 | 1 |
| BCL2A1 | 0.25178026 | -0.037695331 | 0.996 | 0.992 | 1 |
| CXCR5 | 0.260126523 | -0.051380448 | 0.223 | 0.223 | 1 |
| CLEC4E | 0.260234231 | -0.003459364 | 0.002 | 0.005 | 1 |
| CCR8 | 0.261462953 | -0.003390779 | 0.001 | 0.002 | 1 |
| TOP2A | 0.265811033 | -0.004640503 | 0.003 | 0.006 | 1 |
| PSME3 | 0.266254474 | -0.032034669 | 0.385 | 0.429 | 1 |
| IL21 | 0.272968112 | -0.069644379 | 0.09 | 0.085 | 1 |
| IGHD | 0.274270168 | -0.007083248 | 0.008 | 0.01 | 1 |
| ADGRG3 | 0.279859671 | -0.0058817 | 0.006 | 0.009 | 1 |
| TNFSF10 | 0.314906053 | -0.02925338 | 0.08 | 0.107 | 1 |
| CXCL13 | 0.318371622 | -0.04306823 | 0.002 | 0.002 | 1 |
| IL15 | 0.323953985 | -0.004806266 | 0.007 | 0.011 | 1 |
| IGHG3 | 0.327173844 | -0.004151542 | 0.005 | 0.008 | 1 |
| IL18 | 0.327230198 | -0.004573845 | 0.002 | 0.003 | 1 |
| CHD1 | 0.343892318 | -0.007728909 | 0.025 | 0.029 | 1 |
| ARL4C | 0.348733073 | -0.028038771 | 0.189 | 0.205 | 1 |
| CD1C | 0.366939883 | -0.003561853 | 0.005 | 0.007 | 1 |
| LRRC32 | 0.378181686 | -0.01137217 | 0.025 | 0.03 | 1 |
| TREM1 | 0.384456851 | -0.003561853 | 0.005 | 0.006 | 1 |
| CLEC10A | 0.385884425 | -0.00171485 | 0.001 | 0.002 | 1 |

| | | | | | |
|---|---|---|---|---|---|
| LAMP3 | 0.398925583 | -0.025674957 | 0.182 | 0.188 | 1 |
| LAP3 | 0.40034699 | -0.021387088 | 0.244 | 0.255 | 1 |
| LGALS3 | 0.401289758 | -0.013696071 | 0.086 | 0.098 | 1 |
| TRIB1 | 0.469509264 | -0.031606446 | 0.43 | 0.431 | 1 |
| CXCR6 | 0.471516981 | -0.004905123 | 0.008 | 0.01 | 1 |
| TNFRSF17 | 0.472507356 | -0.002867103 | 0.002 | 0.002 | 1 |
| STIM1 | 0.49956292 | -0.014189472 | 0.196 | 0.195 | 1 |
| PSME2 | 0.508670293 | -0.024554466 | 0.649 | 0.677 | 1 |
| CD209 | 0.515690498 | -0.001155774 | 0.001 | 0.002 | 1 |
| S100A9 | 0.518215191 | -0.008114745 | 0.041 | 0.043 | 1 |
| ZAP70 | 0.518327885 | -0.008749413 | 0.053 | 0.061 | 1 |
| IGHG1 | 0.52084983 | -0.045497554 | 0.113 | 0.134 | 1 |
| ZNF341 | 0.523798381 | -0.003177496 | 0.007 | 0.01 | 1 |
| RNASE6 | 0.536080943 | -0.002484008 | 0.005 | 0.007 | 1 |
| IL9 | 0.536949852 | -0.133690479 | 0.029 | 0.026 | 1 |
| NANS | 0.557235179 | -0.017948181 | 0.385 | 0.413 | 1 |
| KDELR1 | 0.558886814 | -0.011617072 | 0.198 | 0.194 | 1 |
| FOXP3 | 0.569355838 | -0.00696936 | 0.01 | 0.014 | 1 |
| TGFBI | 0.572368524 | -0.008988591 | 0.074 | 0.072 | 1 |
| CR2 | 0.576329686 | -0.004200257 | 0.016 | 0.02 | 1 |
| CCL13 | 0.576783295 | -0.001155774 | 0.001 | 0.001 | 1 |
| TCF4 | 0.590769771 | -0.001191823 | 0.002 | 0.002 | 1 |
| IL26 | 0.593277341 | -0.004584145 | 0.011 | 0.013 | 1 |
| CXCL16 | 0.598109065 | -0.007675874 | 0.071 | 0.068 | 1 |
| ENTPD1 | 0.604581651 | -0.002414121 | 0.002 | 0.004 | 1 |
| JUNB | 0.613733241 | -0.016171951 | 0.498 | 0.483 | 1 |
| IL23R | 0.620040875 | -0.003348117 | 0.009 | 0.012 | 1 |
| CHD7 | 0.632174358 | -0.005361383 | 0.039 | 0.045 | 1 |
| CD69 | 0.641978944 | -0.019985204 | 0.998 | 0.998 | 1 |
| LILRB4 | 0.652688125 | -0.003035554 | 0.017 | 0.019 | 1 |
| IRF3 | 0.654717264 | -0.003069586 | 0.017 | 0.02 | 1 |
| SRGN | 0.67230793 | -0.012428622 | 0.952 | 0.946 | 1 |
| JUN | 0.687594902 | -0.01020597 | 0.138 | 0.141 | 1 |
| MGST1 | 0.689253755 | -0.002208582 | 0.011 | 0.012 | 1 |
| TCF7 | 0.696581809 | -0.007197694 | 0.119 | 0.125 | 1 |
| TIMM9 | 0.724626648 | -0.01120264 | 0.534 | 0.566 | 1 |
| IGHM | 0.749652314 | -0.00279427 | 0.022 | 0.021 | 1 |
| VMO1 | 0.767185765 | -0.003031146 | 0.023 | 0.025 | 1 |
| EBF1 | 0.785030333 | -0.002419818 | 0.025 | 0.026 | 1 |
| CD79A | 0.791377197 | -0.002657799 | 0.029 | 0.029 | 1 |
| AQP9 | 0.837583345 | -0.001280262 | 0.014 | 0.016 | 1 |
| OSM | 0.842235533 | -0.002757879 | 0.037 | 0.034 | 1 |
| S100A12 | 0.845168304 | -0.000958818 | 0.008 | 0.008 | 1 |
| TCL1A | 0.859046709 | -0.000958818 | 0.008 | 0.007 | 1 |
| CD6 | 0.900977597 | -0.004837479 | 0.682 | 0.725 | 1 |
| CYBB | 0.925334218 | -0.001189474 | 0.069 | 0.066 | 1 |
| EIF5A | 0.927991643 | -0.003359503 | 0.956 | 0.946 | 1 |

| | | | | | |
|---|---|---|---|---|---|
| SOCS3 | 0.952590978 | -0.001567679 | 0.215 | 0.222 | 1 |
| SOCS1 | 0.956255964 | -0.000147551 | 0.003 | 0.003 | 1 |
| IFITM3 | 0.980175136 | -0.000443033 | 0.066 | 0.064 | 1 |
| KIT | 0.995335709 | -0.00391505 | 0 | 0.002 | 1 |
| IKZF2 | 0.995500444 | -3.16E-05 | 0.012 | 0.011 | 1 |
| BCL10 | 0.002393603 | 0.053395497 | 0.139 | 0.109 | 1 |
| IER3 | 0.002635308 | 0.170864888 | 0.922 | 0.918 | 1 |
| F5 | 0.003435214 | 0.138484842 | 0.153 | 0.127 | 1 |
| BATF | 0.004092679 | 0.105020011 | 0.596 | 0.534 | 1 |
| CASP5 | 0.006118528 | 0.032584007 | 0.072 | 0.052 | 1 |
| YBX3 | 0.006691109 | 0.055653271 | 0.107 | 0.088 | 1 |
| FTH1 | 0.00702232 | 0.105276424 | 0.926 | 0.909 | 1 |
| FOXO1 | 0.007122258 | 0.044840159 | 0.096 | 0.08 | 1 |
| VNN2 | 0.007726737 | 0.010610851 | 0.011 | 0.003 | 1 |
| FUT4 | 0.010329882 | 0.025863622 | 0.05 | 0.033 | 1 |
| IKZF1 | 0.011256688 | 0.047297494 | 0.113 | 0.083 | 1 |
| CCR5 | 0.012279216 | 0.034531171 | 0.062 | 0.049 | 1 |
| BAX | 0.01247242 | 0.065793724 | 0.364 | 0.317 | 1 |
| POU2AF1 | 0.012656886 | 0.082557615 | 0.122 | 0.102 | 1 |
| FYB1 | 0.014103985 | 0.049914928 | 0.116 | 0.101 | 1 |
| DPP4 | 0.016566401 | 0.022949717 | 0.023 | 0.014 | 1 |
| CD247 | 0.017464878 | 0.050447879 | 0.185 | 0.153 | 1 |
| TSPAN32 | 0.019210246 | 0.024241705 | 0.05 | 0.033 | 1 |
| HLA.DQB1 | 0.019850364 | 0.098838236 | 0.425 | 0.366 | 1 |
| TYMS | 0.019855632 | 0.028514559 | 0.037 | 0.029 | 1 |
| GEM | 0.02310688 | 0.159277062 | 0.546 | 0.547 | 1 |
| NAMPT | 0.029549241 | 0.077220118 | 0.989 | 0.986 | 1 |
| IL1R1 | 0.030777684 | 0.076463028 | 0.134 | 0.124 | 1 |
| IL1R2 | 0.035635111 | 0.007993041 | 0.005 | 0.002 | 1 |
| ATF4 | 0.040632143 | 0.064880808 | 0.991 | 0.984 | 1 |
| HLA.DMA | 0.046600162 | 0.033717629 | 0.103 | 0.081 | 1 |
| TNIP3 | 0.047491454 | 0.021825451 | 0.049 | 0.04 | 1 |
| PIK3IP1 | 0.0482497 | 0.043709882 | 0.17 | 0.151 | 1 |
| CD7 | 0.055677237 | 0.062271132 | 0.351 | 0.307 | 1 |
| CXCL3 | 0.063815364 | 0.069044158 | 0.16 | 0.165 | 1 |
| TNFRSF9 | 0.073733567 | 0.029408594 | 0.112 | 0.101 | 1 |
| EGR1 | 0.073822317 | 0.075667599 | 0.505 | 0.451 | 1 |
| CD8B | 0.08063572 | 0.011095356 | 0.02 | 0.013 | 1 |
| TNFRSF25 | 0.080691884 | 0.053184159 | 0.279 | 0.264 | 1 |
| SNX8 | 0.091764224 | 0.042325039 | 0.236 | 0.22 | 1 |
| PDCD1 | 0.11203883 | 0.029353394 | 0.091 | 0.075 | 1 |
| PMCH | 0.117462282 | 0.065923191 | 0.012 | 0.005 | 1 |
| LGALS9 | 0.120076172 | 0.018310227 | 0.043 | 0.035 | 1 |
| CD33 | 0.127864512 | 0.002685716 | 0.002 | 0 | 1 |
| LY86 | 0.131427465 | 0.006666156 | 0.011 | 0.007 | 1 |
| CCR4 | 0.141107257 | 0.068175296 | 0.155 | 0.137 | 1 |
| CD300A | 0.145752851 | 0.053338876 | 0.164 | 0.142 | 1 |

| | | | | | |
|---|---|---|---|---|---|
| VSIG4 | 0.147012959 | 0.00476433 | 0.004 | 0.002 | 1 |
| CD9 | 0.161377144 | 0.029646626 | 0.095 | 0.082 | 1 |
| CD27 | 0.164383766 | 0.036649721 | 0.034 | 0.047 | 1 |
| SELL | 0.165675234 | 0.073270434 | 0.275 | 0.264 | 1 |
| NDFIP2 | 0.166016414 | 0.025879196 | 0.141 | 0.129 | 1 |
| SLC1A5 | 0.173657697 | 0.042800715 | 0.962 | 0.949 | 1 |
| AIM2 | 0.179422236 | 0.009308013 | 0.023 | 0.016 | 1 |
| GBP2 | 0.18523878 | 0.051470838 | 0.752 | 0.768 | 1 |
| CD4 | 0.18933391 | 0.013212994 | 0.021 | 0.018 | 1 |
| BCL11B | 0.190347769 | 0.032458506 | 0.127 | 0.114 | 1 |
| KLRK1 | 0.191612319 | 0.050631252 | 0.873 | 0.838 | 1 |
| BTLA | 0.205079865 | 0.029408594 | 0.077 | 0.073 | 1 |
| EIF3J | 0.211439987 | 0.033958969 | 0.356 | 0.346 | 1 |
| TRAT1 | 0.217159901 | 0.015971615 | 0.038 | 0.039 | 1 |
| IL6R | 0.217907844 | 0.015231427 | 0.065 | 0.05 | 1 |
| IL4R | 0.23536027 | 0.037626664 | 0.876 | 0.868 | 1 |
| IGLC3 | 0.243721659 | 0.100850405 | 0.287 | 0.3 | 1 |
| MITF | 0.246489725 | 0.006109882 | 0.008 | 0.007 | 1 |
| EOMES | 0.26597159 | 0.008543942 | 0.025 | 0.02 | 1 |
| CD274 | 0.267542807 | 0.042434264 | 0.357 | 0.342 | 1 |
| TNFAIP8 | 0.288194793 | 0.033289156 | 0.993 | 0.989 | 1 |
| ATF6B | 0.293016004 | 0.017346876 | 0.129 | 0.118 | 1 |
| FN1 | 0.327506955 | 0.020184068 | 0.131 | 0.135 | 1 |
| PAXBP1 | 0.370447276 | 0.023283119 | 0.312 | 0.299 | 1 |
| PSEN1 | 0.38878755 | 0.010390046 | 0.066 | 0.059 | 1 |
| CD163 | 0.403860254 | 0.005228905 | 0.017 | 0.012 | 1 |
| GAB2 | 0.407491138 | 0.013024867 | 0.081 | 0.072 | 1 |
| ITGAE | 0.446408521 | 0.012939056 | 0.089 | 0.077 | 1 |
| CD86 | 0.4601699 | 0.012416435 | 0.062 | 0.049 | 1 |
| CD22 | 0.480871957 | 0.011329366 | 0.059 | 0.057 | 1 |
| THEMIS | 0.490018332 | 0.010138385 | 0.059 | 0.054 | 1 |
| TIAF1 | 0.497639236 | 0.005336506 | 0.023 | 0.02 | 1 |
| DCLRE1C | 0.511221013 | 0.008851164 | 0.062 | 0.052 | 1 |
| SMAD2 | 0.516029489 | 0.012424677 | 0.159 | 0.154 | 1 |
| SNX9 | 0.539581898 | 0.022833884 | 0.673 | 0.71 | 1 |
| LIPA | 0.544896141 | 0.012212684 | 0.135 | 0.122 | 1 |
| TNFSF8 | 0.552126307 | 0.023999279 | 0.118 | 0.126 | 1 |
| CCND2 | 0.555813062 | 0.024973456 | 0.604 | 0.611 | 1 |
| MZB1 | 0.558373751 | 0.008459059 | 0.014 | 0.011 | 1 |
| POU2F2 | 0.578826438 | 0.016679116 | 0.287 | 0.284 | 1 |
| CMKLR1 | 0.583991257 | 0.002414167 | 0.008 | 0.005 | 1 |
| PGM3 | 0.611128191 | 0.007887315 | 0.091 | 0.09 | 1 |
| BCL2L1 | 0.624538614 | 0.01613425 | 0.576 | 0.563 | 1 |
| SNCA | 0.62633987 | 0.001007187 | 0.002 | 0.002 | 1 |
| ADGRE1 | 0.629740669 | 0.001452346 | 0.005 | 0.003 | 1 |
| IL11RA | 0.632509433 | 0.004394306 | 0.028 | 0.028 | 1 |
| CHI3L2 | 0.641658831 | 0.00189565 | 0.006 | 0.005 | 1 |

| | | | | | |
|---|---|---|---|---|---|
| BIN2 | 0.64597114 | 0.004160594 | 0.036 | 0.033 | 1 |
| CD36 | 0.668158334 | 0.00349936 | 0.028 | 0.026 | 1 |
| IGBP1 | 0.675710217 | 0.007489662 | 0.143 | 0.138 | 1 |
| NFKB2 | 0.679151481 | 0.010693298 | 0.279 | 0.267 | 1 |
| NGDN | 0.682738615 | 0.003184117 | 0.024 | 0.02 | 1 |
| TNFSF13B | 0.710356942 | 0.003097893 | 0.013 | 0.008 | 1 |
| FCER2 | 0.71533968 | 0.004235934 | 0.037 | 0.037 | 1 |
| NFATC3 | 0.715860862 | 0.008925993 | 0.253 | 0.256 | 1 |
| CD14 | 0.749278291 | 0.002712825 | 0.016 | 0.019 | 1 |
| CCR1 | 0.767870504 | 0.005786971 | 0.128 | 0.109 | 1 |
| BLNK | 0.773340192 | 0.000894352 | 0.005 | 0.004 | 1 |
| CD79B | 0.787059038 | 0.000894352 | 0.005 | 0.003 | 1 |
| TGFB3 | 0.814585188 | 0.000894352 | 0.004 | 0.003 | 1 |
| CXCL9 | 0.817339336 | 0.011726526 | 0.058 | 0.059 | 1 |
| MALT1 | 0.817478967 | 0.004179991 | 0.131 | 0.126 | 1 |
| SPP1 | 0.839208511 | 0.001328878 | 0.017 | 0.016 | 1 |
| CARD11 | 0.86940467 | 0.000707432 | 0.008 | 0.008 | 1 |
| GIMAP5 | 0.88670825 | 0.00414767 | 0.251 | 0.275 | 1 |
| CCR6 | 0.925833551 | 0.000336573 | 0.003 | 0.004 | 1 |
| GRB10 | 0.930287649 | 0.000521924 | 0.009 | 0.011 | 1 |
| LTB | 0.938865812 | 0.000448112 | 0.013 | 0.012 | 1 |
| EEF1E1 | 0.950571814 | 0.001431268 | 0.224 | 0.227 | 1 |
| CTLA4 | 0.958365156 | 0.004198951 | 0.386 | 0.323 | 1 |
| MAST4 | 0.961098844 | 0.00112054 | 0.116 | 0.113 | 1 |

Naïve B-cells

| | p_val | avg_log2FC | Patient | Controls | p_val_adj |
|---|---|---|---|---|---|
| GBP2 | 1.56E-85 | 1.140188487 | 0.959 | 0.745 | 7.78E-83 |
| LTA | 1.09E-71 | -1.578247542 | 0.783 | 0.894 | 5.41E-69 |
| GBP1 | 1.14E-53 | 0.97036791 | 0.849 | 0.656 | 5.70E-51 |
| CR2 | 3.78E-51 | 0.856243349 | 0.565 | 0.269 | 1.88E-48 |
| JAK1 | 5.03E-51 | 0.682908498 | 0.921 | 0.783 | 2.50E-48 |
| TNF | 5.98E-49 | -1.218839884 | 0.972 | 0.974 | 2.98E-46 |
| IGHD | 4.34E-27 | 0.552958027 | 0.493 | 0.23 | 2.16E-24 |
| HLA.A | 1.13E-22 | 0.419614243 | 1 | 1 | 5.64E-20 |
| HLA.DQB1 | 2.35E-22 | 0.534120873 | 0.981 | 0.958 | 1.17E-19 |
| CSF2 | 2.71E-21 | -0.842190115 | 0.876 | 0.872 | 1.35E-18 |
| HLA.DRA | 1.01E-18 | 0.490177046 | 0.996 | 0.993 | 5.04E-16 |
| PIK3AP1 | 1.08E-18 | 0.419324162 | 0.721 | 0.506 | 5.40E-16 |
| SNX8 | 3.40E-18 | -0.419890269 | 0.625 | 0.784 | 1.69E-15 |
| DUSP4 | 3.04E-17 | -0.799108134 | 0.499 | 0.634 | 1.51E-14 |
| CD37 | 3.03E-15 | 0.3664796 | 0.849 | 0.765 | 1.51E-12 |
| CXCL8 | 7.41E-15 | 0.675629219 | 0.898 | 0.868 | 3.69E-12 |
| FURIN | 3.05E-14 | 0.418271207 | 0.64 | 0.494 | 1.52E-11 |
| CD74 | 2.30E-13 | 0.384229813 | 0.998 | 0.994 | 1.15E-10 |
| IFNGR1 | 2.65E-13 | 0.363858302 | 0.77 | 0.624 | 1.32E-10 |

| | | | | | |
|---|---|---|---|---|---|
| IVNS1ABP | 9.59E-13 | 0.319077323 | 0.787 | 0.665 | 4.77E-10 |
| ADA | 1.35E-12 | 0.394121351 | 0.273 | 0.169 | 6.74E-10 |
| POU2F2 | 2.07E-12 | 0.286298971 | 0.418 | 0.264 | 1.03E-09 |
| ANXA5 | 2.13E-12 | -0.468733129 | 0.776 | 0.86 | 1.06E-09 |
| CXCR5 | 2.28E-12 | 0.39854626 | 0.966 | 0.943 | 1.13E-09 |
| TNFAIP8 | 1.10E-11 | 0.276385439 | 0.998 | 0.985 | 5.46E-09 |
| CCL22 | 1.28E-11 | -1.23845171 | 0.226 | 0.28 | 6.38E-09 |
| STAT3 | 1.68E-10 | 0.284891347 | 0.951 | 0.883 | 8.35E-08 |
| LGALS1 | 1.69E-10 | -0.389756461 | 0.292 | 0.385 | 8.42E-08 |
| IL1A | 3.89E-10 | -0.403069116 | 0.085 | 0.155 | 1.94E-07 |
| CCL4 | 4.20E-10 | -0.500126328 | 1 | 1 | 2.09E-07 |
| IL15RA | 4.20E-10 | 0.333652593 | 0.642 | 0.495 | 2.09E-07 |
| LEF1 | 2.22E-09 | 0.191302457 | 0.115 | 0.056 | 1.11E-06 |
| TGIF1 | 4.00E-09 | 0.296470584 | 0.678 | 0.538 | 1.99E-06 |
| CCL3 | 4.06E-09 | -0.457690908 | 1 | 0.999 | 2.02E-06 |
| CD22 | 1.71E-08 | 0.40880293 | 0.674 | 0.555 | 8.53E-06 |
| LDHA | 4.71E-08 | 0.24612419 | 0.998 | 0.996 | 2.35E-05 |
| NFATC2 | 7.48E-08 | -0.171401669 | 0.062 | 0.162 | 3.73E-05 |
| CD86 | 9.30E-08 | 0.401249611 | 0.505 | 0.362 | 4.63E-05 |
| LAP3 | 1.36E-07 | 0.240290662 | 0.478 | 0.387 | 6.76E-05 |
| HIF1A | 1.53E-07 | 0.22056434 | 0.676 | 0.603 | 7.60E-05 |
| CD44 | 2.85E-07 | 0.210337691 | 1 | 1 | 0.000141826 |
| NFATC1 | 4.98E-07 | 0.203403322 | 0.448 | 0.34 | 0.00024811 |
| IGHG3 | 5.38E-07 | -0.170271733 | 0.011 | 0.064 | 0.000267756 |
| EGR1 | 5.94E-07 | -0.247172819 | 0.299 | 0.412 | 0.000295851 |
| FCER2 | 1.14E-06 | -0.304494676 | 0.418 | 0.521 | 0.000567758 |
| DUSP2 | 2.00E-06 | 0.286618058 | 0.989 | 0.985 | 0.000994057 |
| IGHG2 | 2.17E-06 | -0.141565307 | 0.011 | 0.06 | 0.001081266 |
| IL12A | 2.66E-06 | -0.192626836 | 0.066 | 0.11 | 0.001326228 |
| IGHM | 3.95E-06 | 0.414155244 | 0.367 | 0.245 | 0.001965146 |
| CD274 | 3.96E-06 | 0.299672782 | 0.744 | 0.605 | 0.001974066 |
| TRBC2 | 4.18E-06 | 0.230498415 | 0.825 | 0.752 | 0.002082622 |
| MYC | 4.24E-06 | 0.260109102 | 0.979 | 0.964 | 0.002113883 |
| ITGB2 | 5.62E-06 | 0.225536095 | 0.235 | 0.152 | 0.00279956 |
| EBF1 | 1.14E-05 | 0.210977872 | 0.569 | 0.458 | 0.005696546 |
| IL22 | 1.45E-05 | -0.36126767 | 0.049 | 0.066 | 0.007224629 |
| CD70 | 1.86E-05 | -0.213820762 | 0.038 | 0.114 | 0.009286952 |
| PCNA | 1.92E-05 | 0.173513684 | 0.452 | 0.375 | 0.009562769 |
| IGLC3 | 3.25E-05 | 0.520905343 | 0.51 | 0.458 | 0.016199593 |
| IL2RA | 4.73E-05 | 0.165894208 | 0.429 | 0.307 | 0.023540429 |
| CXCL10 | 4.89E-05 | 0.706286352 | 0.232 | 0.161 | 0.024337132 |
| JAK2 | 8.02E-05 | 0.046713761 | 0.047 | 0.015 | 0.039950609 |
| JUNB | 0.000112816 | 0.18222806 | 0.463 | 0.391 | 0.056182409 |
| TBX21 | 0.000127838 | 0.216418976 | 0.91 | 0.858 | 0.063663298 |
| NFKB1 | 0.000131156 | 0.134779506 | 1 | 1 | 0.065315522 |
| IL2 | 0.000143348 | 0.221217964 | 0.998 | 0.998 | 0.071387205 |
| SNX9 | 0.000164832 | -0.160420146 | 0.866 | 0.917 | 0.08208611 |

| | | | | | |
|---|---|---|---|---|---|
| ITGAX | 0.000192975 | 0.169896889 | 0.119 | 0.111 | 0.096101324 |
| MAP2K3 | 0.000236247 | 0.184619852 | 0.859 | 0.826 | 0.117650856 |
| IRF1 | 0.000262144 | 0.164776861 | 0.974 | 0.96 | 0.130547789 |
| LGALS9 | 0.000351433 | 0.119698502 | 0.222 | 0.161 | 0.175013772 |
| CD72 | 0.000375194 | 0.084188467 | 0.102 | 0.062 | 0.186846771 |
| CCND2 | 0.000419499 | 0.227128942 | 0.606 | 0.484 | 0.208910494 |
| IRF4 | 0.00050838 | 0.177161935 | 0.966 | 0.938 | 0.253173263 |
| TUBA1C | 0.000521066 | -0.169849314 | 0.985 | 0.976 | 0.259491006 |
| IKZF1 | 0.000576327 | 0.089201027 | 0.175 | 0.111 | 0.287010916 |
| PTPRC | 0.000751742 | 0.160713017 | 0.414 | 0.346 | 0.37436768 |
| FAS | 0.000890149 | 0.129486691 | 0.226 | 0.171 | 0.44329407 |
| BIRC3 | 0.000931195 | 0.268885608 | 0.921 | 0.852 | 0.463735302 |
| SDCBP | 0.000965914 | 0.170154104 | 0.712 | 0.656 | 0.481024955 |
| BTG1 | 0.000975267 | 0.133867753 | 1 | 0.997 | 0.485683098 |
| IL2RG | 0.001029218 | 0.142606859 | 0.834 | 0.815 | 0.512550597 |
| GAB2 | 0.001259608 | 0.110720048 | 0.237 | 0.183 | 0.627284896 |
| IFNG | 0.001585196 | 0.199939446 | 0.977 | 0.984 | 0.789427389 |
| PSMB9 | 0.001640966 | 0.161974582 | 0.618 | 0.534 | 0.817201189 |
| CXCL3 | 0.001907332 | -0.138792907 | 0.124 | 0.131 | 0.949851453 |
| CD1C | 0.001956972 | -0.180947904 | 0.021 | 0.049 | 0.974572162 |
| REL | 0.00196165 | -0.132612203 | 0.977 | 0.977 | 0.976901624 |
| STAT6 | 0.001980344 | 0.124798217 | 0.452 | 0.378 | 0.986211177 |
| GAPDH | 0.002997497 | -0.222630306 | 0.872 | 0.865 | 1 |
| PDIA6 | 0.003485138 | -0.19224327 | 0.757 | 0.769 | 1 |
| CD83 | 0.004897098 | -0.140938869 | 0.996 | 0.995 | 1 |
| IGHG1 | 0.006154943 | -0.232999527 | 0.177 | 0.237 | 1 |
| DCLRE1C | 0.007056845 | -0.043736614 | 0.017 | 0.042 | 1 |
| CD80 | 0.009225415 | -0.09298403 | 0.102 | 0.131 | 1 |
| CYBB | 0.010517026 | -0.098741375 | 0.107 | 0.155 | 1 |
| RGS1 | 0.011440252 | -0.254586667 | 0.544 | 0.622 | 1 |
| IL4R | 0.012459275 | -0.10579001 | 0.885 | 0.907 | 1 |
| TGFBI | 0.012498437 | -0.052003826 | 0.038 | 0.073 | 1 |
| IL3 | 0.013542947 | -0.082496556 | 0.068 | 0.103 | 1 |
| UGCG | 0.014370048 | -0.123084588 | 0.271 | 0.3 | 1 |
| GEM | 0.018816957 | -0.192672371 | 0.518 | 0.522 | 1 |
| IGHA1 | 0.020324738 | -0.031644922 | 0.004 | 0.022 | 1 |
| CD48 | 0.02426822 | -0.117043682 | 0.951 | 0.971 | 1 |
| RPL6 | 0.024717236 | -0.084507075 | 1 | 1 | 1 |
| CXCR4 | 0.027169873 | -0.187782932 | 0.358 | 0.361 | 1 |
| XBP1 | 0.027369217 | -0.112613803 | 0.704 | 0.745 | 1 |
| CD27 | 0.029238611 | -0.024997449 | 0.002 | 0.019 | 1 |
| IRF7 | 0.032572326 | -0.099604207 | 0.456 | 0.509 | 1 |
| CCR7 | 0.034390635 | -0.104990289 | 0.996 | 0.992 | 1 |
| HAVCR2 | 0.039760676 | -0.035144035 | 0.028 | 0.053 | 1 |
| IER3 | 0.042389728 | -0.139522826 | 0.733 | 0.717 | 1 |
| IRF8 | 0.042522985 | -0.114977679 | 0.996 | 0.992 | 1 |
| NFKBID | 0.046903357 | -0.050282902 | 0.085 | 0.121 | 1 |

| | | | | | |
|---|---|---|---|---|---|
| SMAD2 | 0.057752144 | -0.054054458 | 0.128 | 0.151 | 1 |
| FUT4 | 0.065246001 | -0.018871346 | 0.006 | 0.019 | 1 |
| FYN | 0.066534843 | -0.078122379 | 0.397 | 0.45 | 1 |
| CD9 | 0.071440617 | -0.055285952 | 0.113 | 0.141 | 1 |
| NAMPT | 0.082107677 | -0.093759622 | 0.981 | 0.964 | 1 |
| EIF1 | 0.085593483 | -0.061376113 | 1 | 1 | 1 |
| IL1RN | 0.08943078 | -0.062555954 | 0.186 | 0.22 | 1 |
| KLRC3 | 0.093366224 | -0.032177602 | 0.032 | 0.055 | 1 |
| RAPH1 | 0.099364456 | -0.014785539 | 0.004 | 0.014 | 1 |
| QPCT | 0.1019685 | -0.015302966 | 0.006 | 0.017 | 1 |
| PRDM1 | 0.112521487 | -0.038208995 | 0.055 | 0.074 | 1 |
| SP140 | 0.112905526 | -0.031494346 | 0.038 | 0.05 | 1 |
| LAT | 0.116127807 | -0.034219974 | 0.06 | 0.081 | 1 |
| IL4 | 0.116913391 | -0.040472038 | 0.047 | 0.059 | 1 |
| CYCS | 0.122028347 | -0.073937855 | 0.928 | 0.917 | 1 |
| SLC1A5 | 0.122626455 | -0.074001278 | 0.964 | 0.952 | 1 |
| PMCH | 0.12982328 | -0.049031141 | 0.002 | 0.004 | 1 |
| STIM1 | 0.137295414 | -0.031337465 | 0.058 | 0.085 | 1 |
| SPAG9 | 0.151003064 | -0.059900896 | 0.443 | 0.489 | 1 |
| CSRNP1 | 0.152498447 | -0.064879697 | 0.802 | 0.816 | 1 |
| FYB1 | 0.163785833 | -0.016641393 | 0.015 | 0.025 | 1 |
| IL9 | 0.19576927 | -0.018859441 | 0.021 | 0.029 | 1 |
| SLA | 0.196593131 | -0.073456648 | 0.503 | 0.516 | 1 |
| LILRB4 | 0.197132277 | -0.020965053 | 0.019 | 0.026 | 1 |
| SLC7A7 | 0.201473519 | -0.018456354 | 0.026 | 0.035 | 1 |
| TSPAN32 | 0.203767786 | -0.009762472 | 0.004 | 0.011 | 1 |
| IL1R1 | 0.216062279 | -0.020231759 | 0.036 | 0.046 | 1 |
| CD38 | 0.220542079 | -0.007788159 | 0.002 | 0.008 | 1 |
| FOSL1 | 0.221317826 | -0.053658147 | 0.868 | 0.853 | 1 |
| FBXO11 | 0.224880993 | -0.042095506 | 0.252 | 0.28 | 1 |
| TLR2 | 0.225664267 | -0.019047696 | 0.034 | 0.044 | 1 |
| CD200 | 0.241556624 | -0.092536256 | 0.482 | 0.494 | 1 |
| VNN2 | 0.242755083 | -0.007788159 | 0.002 | 0.007 | 1 |
| TCF7 | 0.246899071 | -0.011534906 | 0.006 | 0.015 | 1 |
| MS4A1 | 0.249204799 | -0.064026833 | 0.569 | 0.585 | 1 |
| IL6 | 0.249221747 | -0.129621431 | 0.81 | 0.782 | 1 |
| IL1B | 0.254363981 | -0.057110847 | 0.335 | 0.356 | 1 |
| FASLG | 0.267363039 | -0.030166176 | 0.096 | 0.113 | 1 |
| TRDC | 0.268940444 | -0.009575137 | 0.006 | 0.012 | 1 |
| BAX | 0.276069679 | -0.045468749 | 0.529 | 0.535 | 1 |
| ARPC1B | 0.277350276 | -0.055239299 | 0.339 | 0.38 | 1 |
| PAK1IP1 | 0.284318558 | -0.045448059 | 0.405 | 0.43 | 1 |
| CCL5 | 0.318635969 | -0.02831191 | 0.102 | 0.12 | 1 |
| CBLB | 0.320031738 | -0.00813962 | 0.006 | 0.011 | 1 |
| ABCE1 | 0.333727611 | -0.041260106 | 0.874 | 0.887 | 1 |
| BCL2L1 | 0.336834121 | -0.0502265 | 0.407 | 0.429 | 1 |
| PRF1 | 0.347297962 | -0.041357593 | 0.386 | 0.406 | 1 |

| | | | | | |
|---|---|---|---|---|---|
| CXCL11 | 0.351301041 | -0.043556866 | 0.017 | 0.015 | 1 |
| POU2AF1 | 0.351409891 | -0.073799046 | 0.546 | 0.56 | 1 |
| CCR5 | 0.364822311 | -0.011630225 | 0.023 | 0.03 | 1 |
| GZMA | 0.379721068 | -0.009544994 | 0.015 | 0.02 | 1 |
| LRRC32 | 0.381862295 | -0.064129615 | 0.292 | 0.324 | 1 |
| CD40LG | 0.382868123 | -0.017379013 | 0.058 | 0.063 | 1 |
| CD63 | 0.390945576 | -0.043854096 | 0.746 | 0.753 | 1 |
| IGBP1 | 0.400402316 | -0.024208721 | 0.141 | 0.156 | 1 |
| MALT1 | 0.405946616 | -0.024603247 | 0.16 | 0.179 | 1 |
| IL11RA | 0.420440297 | -0.011256033 | 0.028 | 0.033 | 1 |
| IL3RA | 0.42461951 | -0.016023097 | 0.051 | 0.063 | 1 |
| GNAI2 | 0.425296849 | -0.046979205 | 0.614 | 0.639 | 1 |
| BCL11B | 0.431296256 | -0.011957563 | 0.026 | 0.03 | 1 |
| TCF4 | 0.440711031 | -0.012791722 | 0.032 | 0.036 | 1 |
| CD160 | 0.447242578 | -0.004721856 | 0.004 | 0.008 | 1 |
| IFI6 | 0.451053183 | -0.010739513 | 0.021 | 0.029 | 1 |
| FCN1 | 0.457941317 | -0.006521813 | 0.006 | 0.012 | 1 |
| CD109 | 0.461478371 | -0.02292288 | 0.066 | 0.072 | 1 |
| AIM2 | 0.463054586 | -0.005264292 | 0.006 | 0.01 | 1 |
| CXCR3 | 0.497716761 | -0.015787213 | 0.07 | 0.084 | 1 |
| GZMB | 0.505625588 | -0.037426226 | 0.91 | 0.92 | 1 |
| RNASE6 | 0.505680421 | -0.004000328 | 0.004 | 0.007 | 1 |
| PAX5 | 0.505946307 | -0.007939377 | 0.015 | 0.02 | 1 |
| ZBED2 | 0.511865639 | -0.012658751 | 0.021 | 0.023 | 1 |
| TNFSF10 | 0.515254394 | -0.00562554 | 0.011 | 0.015 | 1 |
| CD40 | 0.530075125 | -0.033827426 | 0.97 | 0.967 | 1 |
| CTSD | 0.546545219 | -0.010946969 | 0.055 | 0.058 | 1 |
| TNFRSF9 | 0.590544702 | -0.003278439 | 0.004 | 0.006 | 1 |
| TNFSF14 | 0.602529364 | -0.006512865 | 0.026 | 0.029 | 1 |
| THBS1 | 0.608697437 | -0.010661834 | 0.032 | 0.036 | 1 |
| IL18R1 | 0.609951693 | -0.006338699 | 0.019 | 0.02 | 1 |
| S100A10 | 0.61062885 | -0.022673122 | 0.365 | 0.375 | 1 |
| TNFRSF4 | 0.613162482 | -0.018901411 | 0.139 | 0.164 | 1 |
| LIF | 0.614741951 | -0.03686527 | 0.183 | 0.212 | 1 |
| IL17F | 0.615428429 | -0.009304422 | 0.034 | 0.028 | 1 |
| NKG7 | 0.619983519 | -0.018908075 | 0.264 | 0.251 | 1 |
| CHI3L2 | 0.63790127 | -0.002005806 | 0.002 | 0.004 | 1 |
| IL5 | 0.665662759 | -0.002556188 | 0.004 | 0.006 | 1 |
| CD7 | 0.670416934 | -0.007022444 | 0.043 | 0.046 | 1 |
| DUSP5 | 0.671275768 | -0.020855256 | 0.962 | 0.967 | 1 |
| CARD11 | 0.676196055 | -0.004017041 | 0.013 | 0.014 | 1 |
| PDCD1 | 0.692894144 | -0.002930338 | 0.009 | 0.011 | 1 |
| ATF4 | 0.695366564 | -0.014262388 | 0.998 | 1 | 1 |
| PSMB8 | 0.70030729 | -0.021164838 | 0.691 | 0.651 | 1 |
| VEGFA | 0.709394199 | -0.003301241 | 0.013 | 0.015 | 1 |
| CD79B | 0.713275305 | -0.004556664 | 0.011 | 0.014 | 1 |
| CX3CR1 | 0.716591631 | -0.004207162 | 0.015 | 0.021 | 1 |

| | | | | | |
|---|---|---|---|---|---|
| LYN | 0.724296839 | -0.021970954 | 0.648 | 0.637 | 1 |
| CD69 | 0.728342678 | -0.017876312 | 0.998 | 0.998 | 1 |
| PLCG2 | 0.735743403 | -0.010345084 | 0.134 | 0.13 | 1 |
| TGFBR2 | 0.752633019 | -0.013868697 | 0.254 | 0.249 | 1 |
| PTPN7 | 0.752876797 | -0.00877868 | 0.136 | 0.149 | 1 |
| EEF1E1 | 0.776380835 | -0.010967069 | 0.33 | 0.34 | 1 |
| TYMS | 0.789150856 | -0.002383223 | 0.004 | 0.006 | 1 |
| BCL2 | 0.797650605 | -0.011006395 | 0.414 | 0.41 | 1 |
| CD300A | 0.797961502 | -0.00203942 | 0.011 | 0.012 | 1 |
| FCER1G | 0.814290951 | -0.006303869 | 0.102 | 0.115 | 1 |
| SLC7A1 | 0.817179425 | -0.009343568 | 0.891 | 0.887 | 1 |
| ITGAM | 0.828121393 | -0.001868574 | 0.013 | 0.014 | 1 |
| S100A12 | 0.841833559 | -0.001110602 | 0.002 | 0.005 | 1 |
| LIPA | 0.848649518 | -0.005829073 | 0.134 | 0.14 | 1 |
| CCR6 | 0.852858361 | -0.001662056 | 0.006 | 0.005 | 1 |
| S100A9 | 0.872882347 | -0.002304433 | 0.03 | 0.033 | 1 |
| CD209 | 0.879075108 | -0.000556589 | 0.002 | 0.003 | 1 |
| IL26 | 0.879075108 | -0.000556589 | 0.002 | 0.003 | 1 |
| DPP4 | 0.879075108 | -0.000556589 | 0.002 | 0.003 | 1 |
| CD244 | 0.879075108 | -0.000556589 | 0.002 | 0.003 | 1 |
| PASK | 0.884301941 | -0.001936829 | 0.028 | 0.029 | 1 |
| ARL4C | 0.90356724 | -0.00176936 | 0.023 | 0.03 | 1 |
| B4GALT5 | 0.91646261 | -0.002610813 | 0.117 | 0.115 | 1 |
| GIMAP2 | 0.938030811 | -0.001270964 | 0.032 | 0.033 | 1 |
| NFKBIZ | 0.940929349 | -0.004500552 | 0.934 | 0.886 | 1 |
| VMO1 | 0.943318472 | -0.000858002 | 0.023 | 0.025 | 1 |
| JCHAIN | 0.945249773 | -0.001905715 | 0.06 | 0.092 | 1 |
| IFITM3 | 0.945983697 | -0.001145877 | 0.045 | 0.045 | 1 |
| CST7 | 0.957524412 | -0.001526389 | 0.147 | 0.157 | 1 |
| SPP1 | 0.959519044 | -0.000434484 | 0.011 | 0.013 | 1 |
| CD226 | 0.961197344 | -0.000735584 | 0.036 | 0.032 | 1 |
| LGALS3 | 0.96123906 | -0.002065679 | 0.23 | 0.228 | 1 |
| SOCS2 | 0.968322004 | -0.00110201 | 0.049 | 0.051 | 1 |
| TRAT1 | 0.972356337 | -0.000218639 | 0.006 | 0.006 | 1 |
| LTB | 0.972356337 | -0.000218639 | 0.006 | 0.006 | 1 |
| CHD1 | 0.989883314 | -0.000149583 | 0.023 | 0.025 | 1 |
| SOCS1 | 0.992967606 | -0.008695325 | 0 | 0.006 | 1 |
| IGHG4 | 0.992967606 | -0.008695325 | 0 | 0.006 | 1 |
| CLEC10A | 0.995658482 | -0.004354214 | 0 | 0.003 | 1 |
| MITF | 0.995658482 | -0.004354214 | 0 | 0.003 | 1 |
| KLRF1 | 0.995658482 | -0.004354214 | 0 | 0.003 | 1 |
| IL17RB | 0.995658482 | -0.004354214 | 0 | 0.003 | 1 |
| THBD | 0.995658482 | -0.004354214 | 0 | 0.003 | 1 |
| ZNF683 | 0.99570371 | -0.003629423 | 0 | 0.003 | 1 |
| EOMES | 0.99570371 | -0.003629423 | 0 | 0.003 | 1 |
| IL12RB1 | 0.995759066 | -0.002904269 | 0 | 0.002 | 1 |
| PCLAF | 0.995830431 | -0.00217875 | 0 | 0.002 | 1 |

| | | | | | |
|---|---|---|---|---|---|
| TNFSF8 | 0.002341298 | 0.042107732 | 0.055 | 0.029 | 1 |
| YBX3 | 0.002863057 | 0.125545961 | 0.343 | 0.285 | 1 |
| VPS28 | 0.003091166 | 0.130459447 | 0.322 | 0.245 | 1 |
| SRGN | 0.004550801 | 0.134625994 | 0.81 | 0.809 | 1 |
| BLNK | 0.005144498 | 0.074790734 | 0.124 | 0.069 | 1 |
| NT5E | 0.005940335 | 0.093404941 | 0.16 | 0.114 | 1 |
| SELL | 0.006400769 | 0.087574225 | 0.198 | 0.156 | 1 |
| IGKC | 0.00659039 | 0.353497939 | 0.469 | 0.435 | 1 |
| TRAC | 0.007890897 | 0.116657012 | 0.38 | 0.359 | 1 |
| TRAF1 | 0.00923168 | 0.112308325 | 0.917 | 0.897 | 1 |
| TOP2A | 0.012459777 | 0.013985377 | 0.013 | 0.003 | 1 |
| LRRC8B | 0.012787348 | 0.075460418 | 0.222 | 0.181 | 1 |
| UFM1 | 0.0136941 | 0.099486416 | 0.906 | 0.896 | 1 |
| STAT1 | 0.014784542 | 0.121637525 | 0.678 | 0.633 | 1 |
| IL32 | 0.014891271 | 0.048744651 | 0.068 | 0.051 | 1 |
| IL15 | 0.015323875 | 0.040937386 | 0.062 | 0.031 | 1 |
| CXCL9 | 0.015375628 | 0.263870518 | 0.075 | 0.074 | 1 |
| FOSB | 0.016161616 | 0.146500389 | 0.397 | 0.37 | 1 |
| AQP9 | 0.017960585 | 0.02521096 | 0.03 | 0.013 | 1 |
| RPN2 | 0.020375319 | 0.101816753 | 0.638 | 0.581 | 1 |
| FOXP1 | 0.022191157 | 0.094788269 | 0.708 | 0.638 | 1 |
| ORAI1 | 0.022243832 | 0.104373316 | 0.544 | 0.481 | 1 |
| FOXO1 | 0.023824145 | 0.051842945 | 0.115 | 0.08 | 1 |
| ERBIN | 0.024000351 | 0.106094479 | 0.529 | 0.483 | 1 |
| STAT4 | 0.027276214 | 0.10624342 | 0.446 | 0.409 | 1 |
| ZBTB16 | 0.027749391 | 0.008472321 | 0.006 | 0.001 | 1 |
| TIAF1 | 0.028896781 | 0.028722619 | 0.041 | 0.023 | 1 |
| BCL2A1 | 0.029070174 | 0.079288752 | 1 | 0.999 | 1 |
| LAMP1 | 0.029864505 | 0.092550078 | 0.542 | 0.479 | 1 |
| JUN | 0.030594813 | 0.073045865 | 0.205 | 0.164 | 1 |
| CD2 | 0.030614581 | 0.092506202 | 0.42 | 0.402 | 1 |
| FOS | 0.031783003 | 0.153124756 | 0.467 | 0.519 | 1 |
| HLA.DPA1 | 0.032730889 | 0.137220578 | 0.928 | 0.862 | 1 |
| IL6ST | 0.032849455 | 0.071396583 | 0.222 | 0.184 | 1 |
| CD52 | 0.033387739 | 0.123866489 | 0.452 | 0.386 | 1 |
| CD36 | 0.036730368 | 0.024724401 | 0.032 | 0.018 | 1 |
| CSF1 | 0.038501842 | 0.07735298 | 0.23 | 0.17 | 1 |
| VPREB3 | 0.040939517 | 0.028616685 | 0.036 | 0.019 | 1 |
| CASP5 | 0.043972144 | 0.037768316 | 0.085 | 0.06 | 1 |
| PDIA4 | 0.044129981 | 0.104495151 | 0.369 | 0.309 | 1 |
| FCGR3A | 0.04497714 | 0.024241993 | 0.041 | 0.022 | 1 |
| GIMAP5 | 0.04497714 | 0.024241993 | 0.038 | 0.023 | 1 |
| TRIB2 | 0.04885101 | 0.031255981 | 0.047 | 0.026 | 1 |
| NRP1 | 0.049364367 | 0.025997927 | 0.047 | 0.027 | 1 |
| PSME3 | 0.049520481 | 0.08813676 | 0.648 | 0.63 | 1 |
| NFATC3 | 0.05056466 | 0.052161721 | 0.141 | 0.109 | 1 |
| NFKBIA | 0.051436438 | 0.111554015 | 1 | 0.999 | 1 |

| | | | | | |
|---|---|---|---|---|---|
| CD8A | 0.054432404 | 0.03283252 | 0.064 | 0.046 | 1 |
| CTLA4 | 0.055585191 | 0.079271536 | 0.271 | 0.229 | 1 |
| MZB1 | 0.060466565 | 0.045716742 | 0.064 | 0.033 | 1 |
| ICOS | 0.063013467 | 0.079401864 | 0.292 | 0.286 | 1 |
| NIBAN3 | 0.067056059 | 0.022822017 | 0.036 | 0.024 | 1 |
| MAST4 | 0.068973538 | 0.027947375 | 0.055 | 0.034 | 1 |
| ZAP70 | 0.071131333 | 0.011089852 | 0.013 | 0.005 | 1 |
| VSIG4 | 0.074882406 | 0.009347992 | 0.006 | 0.002 | 1 |
| CD163 | 0.076432961 | 0.016107926 | 0.023 | 0.012 | 1 |
| TNFRSF13C | 0.083997469 | 0.069017661 | 0.23 | 0.206 | 1 |
| CD3D | 0.084035261 | 0.070129032 | 0.365 | 0.351 | 1 |
| TCL1A | 0.091366077 | 0.086117246 | 0.288 | 0.237 | 1 |
| PFN1 | 0.095591202 | 0.08695431 | 0.998 | 0.998 | 1 |
| SELPLG | 0.097951056 | 0.013679194 | 0.019 | 0.009 | 1 |
| NGDN | 0.124532213 | 0.023635105 | 0.051 | 0.036 | 1 |
| CXCL5 | 0.125049415 | 0.059685789 | 0.294 | 0.257 | 1 |
| HLA.DMA | 0.141069921 | 0.070074416 | 0.397 | 0.326 | 1 |
| STAT5A | 0.148599537 | 0.07448701 | 0.923 | 0.923 | 1 |
| SLC25A37 | 0.149011362 | 0.06060045 | 0.299 | 0.277 | 1 |
| TRIB1 | 0.151302939 | 0.08670204 | 0.339 | 0.255 | 1 |
| PGM3 | 0.153491517 | 0.024895651 | 0.066 | 0.042 | 1 |
| TMEM97 | 0.155490299 | 0.029112006 | 0.083 | 0.068 | 1 |
| TREM1 | 0.155728561 | 0.010366877 | 0.013 | 0.005 | 1 |
| CD4 | 0.158199852 | 0.010512658 | 0.015 | 0.007 | 1 |
| NINJ2 | 0.16073581 | 0.009644265 | 0.011 | 0.006 | 1 |
| RORC | 0.160735816 | 0.009644265 | 0.013 | 0.006 | 1 |
| DOCK8 | 0.161229918 | 0.039437409 | 0.164 | 0.133 | 1 |
| IFITM2 | 0.164447475 | 0.084503699 | 0.473 | 0.428 | 1 |
| IL12RB2 | 0.16463792 | 0.039737074 | 0.104 | 0.067 | 1 |
| CXCL2 | 0.170159119 | 0.067770554 | 0.286 | 0.276 | 1 |
| F5 | 0.170258975 | 0.034038185 | 0.081 | 0.057 | 1 |
| MGST1 | 0.171132853 | 0.008049397 | 0.011 | 0.005 | 1 |
| APOBEC3G | 0.178796477 | 0.06109768 | 0.495 | 0.466 | 1 |
| ARID5A | 0.181370893 | 0.058061216 | 0.606 | 0.583 | 1 |
| VSIR | 0.183034241 | 0.052052399 | 0.264 | 0.236 | 1 |
| NDFIP2 | 0.183590541 | 0.014364984 | 0.03 | 0.019 | 1 |
| RUNX3 | 0.184916511 | 0.064586722 | 0.652 | 0.628 | 1 |
| BCL10 | 0.18715182 | 0.028202311 | 0.113 | 0.093 | 1 |
| CCR1 | 0.19170579 | 0.0295271 | 0.104 | 0.088 | 1 |
| GZMK | 0.192649756 | 0.012536888 | 0.006 | 0.003 | 1 |
| DUSP1 | 0.194544938 | 0.060740513 | 0.401 | 0.371 | 1 |
| IL18 | 0.201930166 | 0.005569513 | 0.006 | 0.003 | 1 |
| LAMP3 | 0.235711342 | 0.052624317 | 0.145 | 0.138 | 1 |
| ITK | 0.236800414 | 0.052222632 | 0.352 | 0.329 | 1 |
| CHD7 | 0.239647541 | 0.02534663 | 0.107 | 0.09 | 1 |
| TGFBR1 | 0.241872193 | 0.03903559 | 0.277 | 0.246 | 1 |
| IL21 | 0.255959673 | 0.025445145 | 0.049 | 0.047 | 1 |

| | | | | | |
|---|---|---|---|---|---|
| DUSP6 | 0.262796236 | 0.053829904 | 0.222 | 0.199 | 1 |
| CXCL1 | 0.282810571 | 0.017221872 | 0.043 | 0.039 | 1 |
| SNCA | 0.289102811 | 0.004844723 | 0.006 | 0.003 | 1 |
| TNIK | 0.320189015 | 0.014903705 | 0.03 | 0.024 | 1 |
| EIF5 | 0.32944525 | 0.041550835 | 0.855 | 0.854 | 1 |
| B3GAT1 | 0.334576638 | 0.006757431 | 0.013 | 0.008 | 1 |
| FN1 | 0.337991314 | 0.028846322 | 0.115 | 0.111 | 1 |
| IFRD1 | 0.340464073 | 0.035741501 | 0.339 | 0.317 | 1 |
| NR4A2 | 0.341011312 | 0.049716617 | 0.316 | 0.291 | 1 |
| HNRNPC | 0.341855035 | 0.04253816 | 0.936 | 0.906 | 1 |
| ITGA4 | 0.343831327 | 0.034727601 | 0.247 | 0.238 | 1 |
| LCK | 0.345165453 | 0.011374941 | 0.026 | 0.017 | 1 |
| BTLA | 0.352902882 | 0.034723857 | 0.173 | 0.168 | 1 |
| ATF6B | 0.356098718 | 0.022695835 | 0.107 | 0.08 | 1 |
| IL6R | 0.359121255 | 0.0197539 | 0.072 | 0.06 | 1 |
| CXCL16 | 0.359264992 | 0.018396708 | 0.072 | 0.066 | 1 |
| IL18RAP | 0.359505177 | 0.009513057 | 0.019 | 0.013 | 1 |
| TLR7 | 0.361919076 | 0.007478598 | 0.013 | 0.006 | 1 |
| RORA | 0.368832629 | 0.015128481 | 0.049 | 0.037 | 1 |
| IL31 | 0.37456298 | 0.008499734 | 0.011 | 0.011 | 1 |
| JAK3 | 0.378126861 | 0.035308998 | 0.399 | 0.375 | 1 |
| CD3G | 0.386730758 | 0.003234868 | 0.004 | 0.002 | 1 |
| GATA3 | 0.388195132 | 0.004120297 | 0.006 | 0.004 | 1 |
| ILF2 | 0.39423203 | 0.033554354 | 0.985 | 0.983 | 1 |
| ENTPD1 | 0.395633605 | 0.006603811 | 0.009 | 0.005 | 1 |
| IRF9 | 0.398387359 | 0.039885731 | 0.599 | 0.564 | 1 |
| BCAT1 | 0.401709001 | 0.04565592 | 0.275 | 0.246 | 1 |
| IL13 | 0.410765495 | 0.019702982 | 0.094 | 0.079 | 1 |
| CD24 | 0.411156052 | 0.02453201 | 0.09 | 0.093 | 1 |
| BACH2 | 0.42116987 | 0.026450717 | 0.217 | 0.206 | 1 |
| CREM | 0.494477595 | 0.035810826 | 0.783 | 0.804 | 1 |
| ITGAE | 0.503524436 | 0.009112202 | 0.034 | 0.027 | 1 |
| CD14 | 0.504013567 | 0.008796902 | 0.015 | 0.011 | 1 |
| SOCS3 | 0.507625794 | 0.031398096 | 0.52 | 0.506 | 1 |
| LY86 | 0.519491566 | 0.014301194 | 0.087 | 0.074 | 1 |
| WAS | 0.521631204 | 0.023410123 | 0.13 | 0.119 | 1 |
| PSEN1 | 0.528393421 | 0.009132328 | 0.047 | 0.039 | 1 |
| TNFRSF25 | 0.532390322 | 0.01256947 | 0.081 | 0.076 | 1 |
| CCL20 | 0.533661977 | 0.038189231 | 0.452 | 0.436 | 1 |
| LAT2 | 0.541142796 | 0.025007527 | 0.19 | 0.211 | 1 |
| CLEC4E | 0.541481427 | 0.003396234 | 0.006 | 0.004 | 1 |
| CCL2 | 0.553217513 | 0.033836268 | 0.539 | 0.563 | 1 |
| EVI2B | 0.554174783 | 0.021757387 | 0.252 | 0.239 | 1 |
| EIF5A | 0.557707535 | 0.025443368 | 0.996 | 0.996 | 1 |
| GZMH | 0.57827342 | 0.008866472 | 0.051 | 0.044 | 1 |
| KLRG1 | 0.578848783 | 0.003556935 | 0.009 | 0.006 | 1 |
| LIG4 | 0.587736401 | 0.017244995 | 0.241 | 0.23 | 1 |

| | | | | | |
|---|---|---|---|---|---|
| TNFSF13B | 0.605698004 | 0.002672534 | 0.006 | 0.005 | 1 |
| MCM4 | 0.608846218 | 0.013536298 | 0.109 | 0.11 | 1 |
| IRF3 | 0.622095607 | 0.006696357 | 0.028 | 0.024 | 1 |
| THEMIS | 0.639674561 | 0.002672534 | 0.004 | 0.005 | 1 |
| CD247 | 0.644143259 | 0.002834685 | 0.009 | 0.007 | 1 |
| SKAP1 | 0.646461725 | 0.004035094 | 0.015 | 0.011 | 1 |
| CTSW | 0.65362157 | 0.004193179 | 0.015 | 0.014 | 1 |
| NFKB2 | 0.662882786 | 0.019408247 | 0.414 | 0.388 | 1 |
| EVI2A | 0.67133121 | 0.013086144 | 0.147 | 0.127 | 1 |
| AHI1 | 0.681240707 | 0.005128293 | 0.028 | 0.025 | 1 |
| CD28 | 0.681472586 | 0.003156992 | 0.013 | 0.011 | 1 |
| ZNF341 | 0.685468607 | 0.004350619 | 0.017 | 0.014 | 1 |
| CD6 | 0.687194872 | 0.009936772 | 0.124 | 0.125 | 1 |
| IL23A | 0.690099047 | 0.016229172 | 0.226 | 0.21 | 1 |
| PAXBP1 | 0.708733421 | 0.01414641 | 0.322 | 0.316 | 1 |
| BCL6 | 0.710796277 | 0.003951316 | 0.023 | 0.02 | 1 |
| PIK3IP1 | 0.71108035 | 0.005943077 | 0.047 | 0.044 | 1 |
| BIN2 | 0.739271261 | 0.001949198 | 0.006 | 0.005 | 1 |
| GNLY | 0.757067895 | 0.011314378 | 0.232 | 0.225 | 1 |
| TIGIT | 0.765130555 | 0.004179907 | 0.034 | 0.031 | 1 |
| KLRC1 | 0.765173305 | 0.005098064 | 0.049 | 0.038 | 1 |
| TNIP3 | 0.765671097 | 0.000894085 | 0.002 | 0.002 | 1 |
| BATF | 0.780437347 | 0.014878037 | 0.418 | 0.361 | 1 |
| ARHGEF3 | 0.780628711 | 0.006239093 | 0.072 | 0.068 | 1 |
| CCR4 | 0.808253198 | 0.005711672 | 0.068 | 0.069 | 1 |
| EIF3J | 0.810303483 | 0.009577064 | 0.48 | 0.495 | 1 |
| LAG3 | 0.811072206 | 0.003942579 | 0.038 | 0.034 | 1 |
| PSME2 | 0.823015807 | 0.011062741 | 0.957 | 0.948 | 1 |
| KLRK1 | 0.829145592 | 0.00641057 | 0.158 | 0.154 | 1 |
| TIMM9 | 0.834185244 | 0.009142237 | 0.719 | 0.698 | 1 |
| GRB10 | 0.858169003 | 0.001555635 | 0.011 | 0.008 | 1 |
| NANS | 0.872604409 | 0.006995438 | 0.478 | 0.463 | 1 |
| CD5 | 0.876819555 | 0.002631526 | 0.047 | 0.048 | 1 |
| MMP9 | 0.883093893 | 0.002716033 | 0.06 | 0.056 | 1 |
| MYB | 0.89996536 | 0.001751987 | 0.032 | 0.027 | 1 |
| FTH1 | 0.907064559 | 0.00685482 | 0.974 | 0.977 | 1 |
| PRNP | 0.91262483 | 0.005962543 | 0.849 | 0.852 | 1 |
| CD3E | 0.919517295 | 0.003072834 | 0.149 | 0.171 | 1 |
| TGFB1 | 0.91952918 | 0.000670101 | 0.009 | 0.008 | 1 |
| CNOT2 | 0.925250274 | 0.003420755 | 0.341 | 0.34 | 1 |
| EGR3 | 0.934389508 | 0.002883576 | 0.173 | 0.168 | 1 |
| IL2RB | 0.937741719 | 0.00284193 | 0.209 | 0.209 | 1 |
| OSM | 0.941443006 | 0.001149364 | 0.043 | 0.037 | 1 |
| TNFRSF8 | 0.94818891 | 0.001149364 | 0.036 | 0.033 | 1 |
| FOXP3 | 0.948708659 | 0.000336434 | 0.004 | 0.004 | 1 |
| ICAM1 | 0.951791405 | 0.003452624 | 0.915 | 0.911 | 1 |
| RBCK1 | 0.955740804 | 0.001804574 | 0.209 | 0.209 | 1 |

| | | | | | |
|---|---|---|---|---|---|
| CD79A | 0.955947537 | 0.003849843 | 0.352 | 0.299 | 1 |
| CCL13 | 0.959713274 | 0.000168565 | 0.002 | 0.002 | 1 |
| PTGDR2 | 0.959713274 | 0.000168565 | 0.002 | 0.002 | 1 |
| KDELR1 | 0.9606193 | 0.001501352 | 0.156 | 0.152 | 1 |
| MCM2 | 0.985821537 | 0.000607581 | 0.096 | 0.111 | 1 |
| IL7R | 0.989966452 | 0.000304043 | 0.083 | 0.079 | 1 |

Memory B-cells

| | p_val | avg_log2FC | Patient | Controls | p_val_adj |
|---|---|---|---|---|---|
| LTA | 1.50E-120 | -2.504890183 | 0.83 | 0.957 | 7.45E-118 |
| TNF | 5.75E-87 | -1.964334109 | 0.986 | 0.996 | 2.87E-84 |
| DUSP4 | 4.64E-46 | -1.139369448 | 0.663 | 0.925 | 2.31E-43 |
| GBP2 | 1.49E-39 | 1.078497054 | 0.948 | 0.753 | 7.40E-37 |
| CSF2 | 5.77E-38 | -1.790186735 | 0.906 | 0.925 | 2.87E-35 |
| LGALS1 | 3.08E-33 | -1.267535475 | 0.358 | 0.597 | 1.53E-30 |
| CR2 | 2.63E-28 | 0.801602414 | 0.479 | 0.206 | 1.31E-25 |
| IGHG1 | 8.32E-27 | -1.723224923 | 0.222 | 0.472 | 4.15E-24 |
| IL1A | 2.00E-26 | -1.468894894 | 0.135 | 0.334 | 9.95E-24 |
| IGHD | 8.94E-25 | 0.78016697 | 0.573 | 0.21 | 4.45E-22 |
| CCL22 | 9.03E-25 | -2.334511733 | 0.344 | 0.551 | 4.50E-22 |
| JAK1 | 4.62E-20 | 0.520793561 | 0.951 | 0.89 | 2.30E-17 |
| IRF8 | 7.33E-18 | -0.63144634 | 0.997 | 0.998 | 3.65E-15 |
| GBP1 | 1.39E-17 | 0.705043222 | 0.889 | 0.754 | 6.94E-15 |
| CCR7 | 2.38E-15 | -0.445632629 | 1 | 0.996 | 1.18E-12 |
| CD70 | 5.80E-15 | -0.762710268 | 0.087 | 0.364 | 2.89E-12 |
| CD40 | 1.16E-13 | -0.543906994 | 0.993 | 0.986 | 5.78E-11 |
| SNX8 | 2.17E-13 | -0.466468438 | 0.684 | 0.823 | 1.08E-10 |
| NFATC2 | 6.52E-13 | -0.398631929 | 0.128 | 0.363 | 3.24E-10 |
| CD37 | 8.43E-13 | 0.441094126 | 0.872 | 0.787 | 4.20E-10 |
| ADA | 2.28E-12 | 0.616577591 | 0.358 | 0.198 | 1.14E-09 |
| RPL6 | 7.82E-12 | -0.328776005 | 1 | 1 | 3.89E-09 |
| IVNS1ABP | 1.10E-11 | 0.384315785 | 0.878 | 0.736 | 5.49E-09 |
| JCHAIN | 1.71E-11 | -1.583966295 | 0.059 | 0.117 | 8.50E-09 |
| PIK3AP1 | 9.30E-11 | 0.430417741 | 0.75 | 0.582 | 4.63E-08 |
| LEF1 | 1.28E-10 | 0.325620842 | 0.163 | 0.087 | 6.37E-08 |
| IL12A | 2.55E-10 | -0.538132361 | 0.08 | 0.199 | 1.27E-07 |
| CD83 | 2.56E-10 | -0.362672898 | 0.997 | 0.997 | 1.28E-07 |
| GAPDH | 3.58E-10 | -0.652620043 | 0.958 | 0.972 | 1.78E-07 |
| NAMPT | 8.18E-10 | -0.422907032 | 0.99 | 0.995 | 4.07E-07 |
| IGHG2 | 1.19E-09 | -0.389306965 | 0.035 | 0.159 | 5.91E-07 |
| IGHG3 | 2.35E-09 | -0.406801394 | 0.035 | 0.171 | 1.17E-06 |
| PDIA6 | 5.88E-09 | -0.477193441 | 0.882 | 0.923 | 2.93E-06 |
| ANXA5 | 5.30E-08 | -0.43662679 | 0.878 | 0.936 | 2.64E-05 |
| IER3 | 7.30E-08 | -0.534909148 | 0.799 | 0.812 | 3.64E-05 |
| LYN | 8.91E-08 | -0.353605929 | 0.75 | 0.891 | 4.43E-05 |
| FURIN | 1.27E-07 | 0.370046806 | 0.677 | 0.577 | 6.30E-05 |

| | | | | | |
|---|---|---|---|---|---|
| CCL20 | 1.29E-07 | -0.580573868 | 0.497 | 0.486 | 6.42E-05 |
| CD80 | 2.47E-07 | -0.334367448 | 0.226 | 0.434 | 0.000122905 |
| CYCS | 2.64E-07 | -0.297956109 | 0.979 | 0.983 | 0.000131318 |
| FAS | 3.57E-07 | 0.282434861 | 0.306 | 0.206 | 0.000177948 |
| HLA.DPA1 | 3.61E-07 | -0.440635657 | 0.976 | 0.968 | 0.000179608 |
| IGLC3 | 3.61E-07 | -1.152173647 | 0.587 | 0.543 | 0.000179628 |
| SLC1A5 | 7.75E-07 | -0.2957297 | 0.983 | 0.987 | 0.000386059 |
| CCL3 | 8.71E-07 | -0.563417724 | 1 | 1 | 0.000433817 |
| CCL4 | 1.08E-06 | -0.563632272 | 1 | 1 | 0.000539198 |
| IGHM | 1.11E-06 | 0.60696389 | 0.472 | 0.272 | 0.000553344 |
| CD22 | 1.64E-06 | 0.518841006 | 0.75 | 0.548 | 0.000817531 |
| IL2 | 1.77E-06 | -0.493539924 | 1 | 1 | 0.000880062 |
| REL | 1.91E-06 | -0.260677028 | 0.979 | 0.991 | 0.00095241 |
| CD48 | 2.32E-06 | -0.293392583 | 0.99 | 0.995 | 0.001154578 |
| IGHA1 | 3.11E-06 | -0.286192959 | 0.007 | 0.073 | 0.001550709 |
| IFNGR1 | 6.88E-06 | 0.298585911 | 0.826 | 0.676 | 0.003426741 |
| YBX3 | 7.14E-06 | 0.255638343 | 0.358 | 0.234 | 0.003553245 |
| EBF1 | 1.90E-05 | 0.253100092 | 0.694 | 0.558 | 0.009462474 |
| GZMB | 2.19E-05 | -0.471771286 | 0.931 | 0.954 | 0.010921101 |
| EIF1 | 2.20E-05 | -0.200309987 | 1 | 1 | 0.010954918 |
| TCL1A | 2.58E-05 | 0.34517936 | 0.306 | 0.122 | 0.012847761 |
| IFNG | 2.94E-05 | -0.511845008 | 0.997 | 0.986 | 0.014661538 |
| BATF | 3.02E-05 | -0.295106725 | 0.531 | 0.611 | 0.015036188 |
| NFATC1 | 4.25E-05 | 0.212938244 | 0.497 | 0.386 | 0.021161018 |
| STAT4 | 7.20E-05 | 0.268573342 | 0.51 | 0.383 | 0.035852005 |
| BCL2L1 | 7.57E-05 | -0.299126377 | 0.5 | 0.607 | 0.037720954 |
| PCNA | 8.30E-05 | 0.242274564 | 0.528 | 0.436 | 0.041344936 |
| ERBIN | 0.000131555 | 0.221567018 | 0.667 | 0.565 | 0.06551444 |
| SOCS2 | 0.000151534 | -0.297638662 | 0.104 | 0.179 | 0.075463712 |
| TUBA1C | 0.000194982 | -0.25460439 | 0.993 | 0.982 | 0.097101005 |
| SP140 | 0.000201282 | -0.163164333 | 0.073 | 0.146 | 0.100238555 |
| CXCL11 | 0.000260844 | 0.165006051 | 0.045 | 0.016 | 0.129900391 |
| EIF5A | 0.000351574 | -0.191511049 | 0.997 | 0.999 | 0.175083799 |
| CD1C | 0.00037368 | -0.442266107 | 0.045 | 0.126 | 0.186092762 |
| NFKBIZ | 0.000418974 | -0.269631137 | 0.948 | 0.933 | 0.208648841 |
| IL6 | 0.000455857 | -0.519988525 | 0.788 | 0.782 | 0.227016599 |
| NFKBID | 0.000536202 | -0.142603876 | 0.115 | 0.206 | 0.267028562 |
| CD63 | 0.000615409 | -0.23703859 | 0.858 | 0.862 | 0.306473642 |
| IL23A | 0.000622986 | -0.313550672 | 0.257 | 0.279 | 0.310247132 |
| BTLA | 0.000636847 | 0.166105376 | 0.181 | 0.103 | 0.317150008 |
| IL22 | 0.000727037 | -0.394388817 | 0.056 | 0.063 | 0.362064301 |
| PSMB8 | 0.000825723 | -0.241962271 | 0.764 | 0.825 | 0.41121027 |
| LIF | 0.000917283 | -0.413822579 | 0.236 | 0.285 | 0.456806872 |
| MZB1 | 0.001038357 | -0.331894294 | 0.101 | 0.092 | 0.517101735 |
| HLA.DMA | 0.001147659 | -0.262468621 | 0.545 | 0.585 | 0.571533952 |
| CD27 | 0.001281941 | -0.105888299 | 0.014 | 0.078 | 0.638406815 |
| POU2F2 | 0.00130703 | 0.187648468 | 0.444 | 0.335 | 0.650900994 |

| | | | | | |
|---|---|---|---|---|---|
| CREM | 0.001481464 | 0.232767141 | 0.806 | 0.777 | 0.737769086 |
| JAK2 | 0.00182518 | 0.058458847 | 0.066 | 0.024 | 0.908939394 |
| RBCK1 | 0.002264657 | -0.144870663 | 0.243 | 0.334 | 1 |
| IL3 | 0.002454559 | -0.230922625 | 0.115 | 0.122 | 1 |
| ATF4 | 0.002458652 | -0.131451451 | 1 | 1 | 1 |
| STAT3 | 0.002475998 | 0.159420025 | 0.976 | 0.945 | 1 |
| MYC | 0.002717208 | 0.20386946 | 0.99 | 0.98 | 1 |
| EGR1 | 0.002734868 | -0.202110322 | 0.337 | 0.42 | 1 |
| FOS | 0.002737131 | 0.343167114 | 0.476 | 0.499 | 1 |
| NIBAN3 | 0.002995584 | 0.059283711 | 0.073 | 0.034 | 1 |
| STAT6 | 0.003131215 | 0.159117171 | 0.608 | 0.486 | 1 |
| FOSB | 0.003547926 | 0.30270202 | 0.392 | 0.333 | 1 |
| IKZF1 | 0.004541185 | 0.106274829 | 0.201 | 0.132 | 1 |
| UGCG | 0.004922772 | -0.190165271 | 0.323 | 0.419 | 1 |
| HIF1A | 0.005042608 | 0.161082387 | 0.729 | 0.693 | 1 |
| CD3G | 0.005601951 | 0.018994763 | 0.014 | 0.001 | 1 |
| CXCL10 | 0.00604527 | 0.643146346 | 0.302 | 0.207 | 1 |
| ARID5A | 0.006309144 | 0.152053157 | 0.694 | 0.604 | 1 |
| GRB10 | 0.006567185 | 0.042358373 | 0.031 | 0.006 | 1 |
| PRF1 | 0.007107434 | -0.280566652 | 0.431 | 0.478 | 1 |
| SNX9 | 0.007960548 | 0.136854561 | 0.938 | 0.927 | 1 |
| BCAT1 | 0.008259275 | -0.219764026 | 0.438 | 0.48 | 1 |
| LAP3 | 0.008421481 | 0.165149352 | 0.559 | 0.485 | 1 |
| TGIF1 | 0.008578004 | 0.177530132 | 0.753 | 0.648 | 1 |
| GEM | 0.008606162 | -0.343794584 | 0.573 | 0.574 | 1 |
| ICAM1 | 0.009517923 | -0.194080713 | 0.948 | 0.957 | 1 |
| LRRC8B | 0.010678734 | 0.110603521 | 0.285 | 0.219 | 1 |
| CD200 | 0.010855053 | 0.291326806 | 0.476 | 0.388 | 1 |
| NKG7 | 0.011204988 | -0.242983994 | 0.295 | 0.323 | 1 |
| TRIB2 | 0.011457343 | 0.064932529 | 0.069 | 0.043 | 1 |
| GZMK | 0.012108521 | 0.048073426 | 0.017 | 0.006 | 1 |
| PFN1 | 0.012870844 | -0.18258134 | 1 | 1 | 1 |
| TOP2A | 0.0132476 | 0.021624547 | 0.021 | 0.006 | 1 |
| CD109 | 0.01357802 | -0.148448153 | 0.076 | 0.122 | 1 |
| PLCG2 | 0.014139539 | -0.133892036 | 0.198 | 0.26 | 1 |
| IL15RA | 0.014225071 | 0.161833107 | 0.74 | 0.71 | 1 |
| PRDM1 | 0.014743286 | -0.137428722 | 0.09 | 0.138 | 1 |
| EVI2A | 0.014756708 | -0.149484433 | 0.188 | 0.229 | 1 |
| IL1RN | 0.0148165 | -0.124400341 | 0.205 | 0.256 | 1 |
| TRIB1 | 0.014904096 | -0.201467702 | 0.34 | 0.386 | 1 |
| CXCR5 | 0.015249587 | 0.167330197 | 0.976 | 0.978 | 1 |
| JUNB | 0.018607585 | 0.137609565 | 0.503 | 0.479 | 1 |
| LAMP3 | 0.018675603 | 0.167290006 | 0.181 | 0.137 | 1 |
| CCL5 | 0.019205037 | -0.16593515 | 0.149 | 0.193 | 1 |
| CD3D | 0.020031722 | 0.176288633 | 0.392 | 0.408 | 1 |
| IL4R | 0.021137197 | -0.125214695 | 0.934 | 0.932 | 1 |
| SELL | 0.021906053 | 0.124556511 | 0.24 | 0.192 | 1 |

| | | | | | |
|---|---|---|---|---|---|
| POU2AF1 | 0.022123402 | -0.22971562 | 0.597 | 0.694 | 1 |
| TBX21 | 0.022538447 | 0.145576052 | 0.965 | 0.941 | 1 |
| TIGIT | 0.023346562 | 0.06872224 | 0.062 | 0.054 | 1 |
| DUSP2 | 0.023805439 | 0.167216369 | 0.993 | 0.992 | 1 |
| IL32 | 0.023977414 | 0.08557359 | 0.09 | 0.086 | 1 |
| PRNP | 0.024094809 | 0.150349358 | 0.878 | 0.866 | 1 |
| ARHGEF3 | 0.025270172 | -0.074608525 | 0.062 | 0.102 | 1 |
| CD163 | 0.02557358 | 0.024223499 | 0.028 | 0.011 | 1 |
| STAT5A | 0.02669585 | -0.130435223 | 0.951 | 0.975 | 1 |
| IRF4 | 0.027681089 | 0.139542814 | 0.979 | 0.977 | 1 |
| IGBP1 | 0.028687404 | -0.09667432 | 0.191 | 0.246 | 1 |
| FCGR3A | 0.029011558 | 0.042681952 | 0.062 | 0.036 | 1 |
| TCF4 | 0.029778833 | -0.06819642 | 0.049 | 0.081 | 1 |
| LAT | 0.030168489 | -0.086938308 | 0.083 | 0.136 | 1 |
| VPREB3 | 0.032914752 | 0.049975969 | 0.059 | 0.029 | 1 |
| LILRB4 | 0.034023156 | -0.074984971 | 0.035 | 0.055 | 1 |
| TRBC2 | 0.034929426 | 0.158412827 | 0.823 | 0.832 | 1 |
| IGHG4 | 0.036040196 | -0.043939088 | 0.003 | 0.021 | 1 |
| CD40LG | 0.036805047 | -0.08429451 | 0.059 | 0.088 | 1 |
| XBP1 | 0.037396797 | -0.137411207 | 0.806 | 0.831 | 1 |
| PTPRC | 0.042612459 | 0.156156749 | 0.51 | 0.449 | 1 |
| CD226 | 0.043584999 | -0.072203365 | 0.059 | 0.09 | 1 |
| TNFRSF25 | 0.043859463 | 0.071993501 | 0.135 | 0.112 | 1 |
| CXCL8 | 0.045173335 | 0.228059792 | 0.92 | 0.903 | 1 |
| BTG1 | 0.045642661 | 0.119910441 | 1 | 0.997 | 1 |
| GZMH | 0.045715009 | 0.089497976 | 0.059 | 0.056 | 1 |
| CCR1 | 0.04743503 | 0.067454168 | 0.139 | 0.097 | 1 |
| TRDC | 0.049129677 | -0.043837214 | 0.014 | 0.03 | 1 |
| MCM4 | 0.049826966 | 0.084949813 | 0.163 | 0.143 | 1 |
| RAPH1 | 0.049888232 | -0.033402011 | 0.003 | 0.022 | 1 |
| QPCT | 0.050293193 | -0.035151486 | 0.014 | 0.036 | 1 |
| GNAI2 | 0.050960409 | -0.173921643 | 0.74 | 0.747 | 1 |
| KLRC3 | 0.051212083 | -0.10233048 | 0.049 | 0.061 | 1 |
| F5 | 0.053371623 | 0.072060874 | 0.104 | 0.072 | 1 |
| SLC25A37 | 0.056433807 | 0.102326866 | 0.344 | 0.29 | 1 |
| BCL2 | 0.059273783 | -0.110507978 | 0.476 | 0.538 | 1 |
| ARPC1B | 0.059353254 | -0.155681363 | 0.486 | 0.564 | 1 |
| SLA | 0.061684778 | -0.155604983 | 0.51 | 0.514 | 1 |
| LGALS3 | 0.062692535 | -0.119795031 | 0.323 | 0.342 | 1 |
| ITGB2 | 0.063275042 | 0.15987964 | 0.372 | 0.3 | 1 |
| PDCD1 | 0.063302001 | -0.028988747 | 0.003 | 0.021 | 1 |
| IL6ST | 0.064257883 | 0.087689129 | 0.222 | 0.188 | 1 |
| IL2RA | 0.066178916 | 0.110004766 | 0.538 | 0.444 | 1 |
| LGALS9 | 0.068185059 | 0.087893677 | 0.257 | 0.211 | 1 |
| ICOS | 0.068784131 | 0.125409579 | 0.323 | 0.316 | 1 |
| TCF7 | 0.070322478 | -0.038584917 | 0.021 | 0.045 | 1 |
| VMO1 | 0.070998769 | -0.031960525 | 0.017 | 0.04 | 1 |

| | | | | | |
|---|---|---|---|---|---|
| CD79A | 0.072525447 | -0.206773863 | 0.483 | 0.482 | 1 |
| FYN | 0.075745665 | -0.115209492 | 0.566 | 0.581 | 1 |
| TNFAIP8 | 0.076495538 | 0.093438009 | 1 | 0.993 | 1 |
| JAK3 | 0.077918515 | 0.090211904 | 0.521 | 0.462 | 1 |
| CCL2 | 0.078367485 | 0.129389065 | 0.545 | 0.614 | 1 |
| AQP9 | 0.08057514 | 0.019341278 | 0.021 | 0.011 | 1 |
| BACH2 | 0.084464271 | -0.081034399 | 0.243 | 0.291 | 1 |
| HNRNPC | 0.084528002 | -0.103014589 | 0.972 | 0.961 | 1 |
| IL15 | 0.084588765 | 0.04342575 | 0.08 | 0.045 | 1 |
| DCLRE1C | 0.084826672 | -0.037686825 | 0.035 | 0.058 | 1 |
| GNLY | 0.087829926 | 0.157267414 | 0.285 | 0.272 | 1 |
| PSME2 | 0.091627293 | -0.105910797 | 0.972 | 0.98 | 1 |
| ZBED2 | 0.092287442 | -0.099059985 | 0.035 | 0.046 | 1 |
| CD69 | 0.093661263 | 0.125175782 | 0.997 | 0.997 | 1 |
| DUSP6 | 0.094725286 | 0.097435168 | 0.229 | 0.2 | 1 |
| CTSD | 0.096548762 | -0.044765138 | 0.062 | 0.087 | 1 |
| TLR2 | 0.100845152 | 0.035205701 | 0.062 | 0.044 | 1 |
| SPP1 | 0.101617117 | 0.019752795 | 0.024 | 0.014 | 1 |
| CX3CR1 | 0.101769697 | -0.042879933 | 0.017 | 0.035 | 1 |
| TREM1 | 0.102263307 | 0.020237084 | 0.021 | 0.009 | 1 |
| DUSP5 | 0.104773795 | 0.105498001 | 0.962 | 0.967 | 1 |
| ILF2 | 0.107669106 | -0.079380716 | 0.993 | 0.995 | 1 |
| ENTPD1 | 0.109404763 | -0.034568591 | 0.014 | 0.029 | 1 |
| FOXO1 | 0.112325135 | 0.057897664 | 0.167 | 0.131 | 1 |
| PASK | 0.115362378 | -0.03765002 | 0.045 | 0.066 | 1 |
| LAMP1 | 0.118779359 | 0.087671179 | 0.622 | 0.594 | 1 |
| BAX | 0.119593904 | -0.087433738 | 0.635 | 0.644 | 1 |
| FCN1 | 0.120395133 | 0.018446028 | 0.017 | 0.011 | 1 |
| CTLA4 | 0.127318149 | 0.096693942 | 0.299 | 0.247 | 1 |
| CD72 | 0.128257861 | 0.057577812 | 0.101 | 0.073 | 1 |
| ORAI1 | 0.128367446 | -0.091843813 | 0.608 | 0.606 | 1 |
| CD9 | 0.132523054 | -0.066033681 | 0.122 | 0.136 | 1 |
| ABCE1 | 0.135767159 | -0.080300986 | 0.941 | 0.944 | 1 |
| PIK3IP1 | 0.139081654 | 0.036754682 | 0.059 | 0.046 | 1 |
| CYBB | 0.140110628 | -0.081882188 | 0.149 | 0.194 | 1 |
| FASLG | 0.140779504 | -0.091287665 | 0.135 | 0.144 | 1 |
| BIRC3 | 0.145956212 | -0.157045347 | 0.951 | 0.873 | 1 |
| SNCA | 0.150020358 | 0.009530066 | 0.01 | 0.004 | 1 |
| CD4 | 0.151094958 | 0.013113553 | 0.017 | 0.008 | 1 |
| AIM2 | 0.158665243 | -0.0253037 | 0.017 | 0.036 | 1 |
| IL18R1 | 0.159538223 | -0.027914914 | 0.021 | 0.033 | 1 |
| B3GAT1 | 0.161686196 | 0.027938321 | 0.028 | 0.021 | 1 |
| RUNX3 | 0.164803952 | -0.091668487 | 0.694 | 0.745 | 1 |
| HAVCR2 | 0.168398971 | -0.04228557 | 0.045 | 0.073 | 1 |
| KDELR1 | 0.171390627 | -0.066350717 | 0.236 | 0.263 | 1 |
| MAST4 | 0.17169888 | -0.032291141 | 0.038 | 0.057 | 1 |
| CXCR4 | 0.17517413 | -0.145085546 | 0.378 | 0.367 | 1 |

| | | | | | |
|---|---|---|---|---|---|
| IFI6 | 0.178129291 | -0.02999699 | 0.031 | 0.049 | 1 |
| TRAT1 | 0.190880914 | 0.017551334 | 0.021 | 0.01 | 1 |
| FOXP1 | 0.19111401 | 0.070685201 | 0.747 | 0.679 | 1 |
| MS4A1 | 0.1921872 | -0.099762692 | 0.684 | 0.683 | 1 |
| B4GALT5 | 0.193591586 | -0.047686003 | 0.135 | 0.164 | 1 |
| ZAP70 | 0.19390488 | 0.013547229 | 0.021 | 0.011 | 1 |
| NFKBIA | 0.197097655 | -0.098294985 | 1 | 1 | 1 |
| TNFRSF13C | 0.204595825 | 0.069665447 | 0.267 | 0.26 | 1 |
| CD38 | 0.20756473 | -0.023121064 | 0.007 | 0.012 | 1 |
| NFKB2 | 0.212832796 | -0.074419501 | 0.514 | 0.534 | 1 |
| IL2RG | 0.213526123 | 0.073567458 | 0.917 | 0.906 | 1 |
| CD74 | 0.218697596 | -0.088467827 | 0.997 | 0.996 | 1 |
| CXCL2 | 0.221027713 | 0.080596988 | 0.382 | 0.369 | 1 |
| EGR3 | 0.221941642 | -0.06355664 | 0.215 | 0.24 | 1 |
| FCER1G | 0.223426991 | -0.047564285 | 0.139 | 0.168 | 1 |
| HLA.A | 0.227329285 | 0.064278623 | 1 | 1 | 1 |
| TRAC | 0.22839893 | 0.089051084 | 0.451 | 0.465 | 1 |
| STIM1 | 0.228967771 | -0.037732523 | 0.08 | 0.115 | 1 |
| PAX5 | 0.229838141 | 0.018382749 | 0.024 | 0.016 | 1 |
| IL3RA | 0.231027435 | -0.038405563 | 0.059 | 0.068 | 1 |
| TGFBR2 | 0.231309952 | -0.074956527 | 0.319 | 0.336 | 1 |
| SELPLG | 0.234615223 | 0.026673354 | 0.031 | 0.015 | 1 |
| CD79B | 0.235282246 | -0.030257452 | 0.021 | 0.032 | 1 |
| NCAM1 | 0.239869213 | 0.00636831 | 0.007 | 0.003 | 1 |
| CD3E | 0.242843316 | 0.069899815 | 0.233 | 0.205 | 1 |
| CXCL1 | 0.249028663 | 0.02744694 | 0.049 | 0.048 | 1 |
| ITGA4 | 0.249768244 | 0.060176092 | 0.302 | 0.27 | 1 |
| CD28 | 0.255205293 | 0.012216076 | 0.014 | 0.008 | 1 |
| RNASE6 | 0.259934721 | -0.016559297 | 0.003 | 0.008 | 1 |
| NGDN | 0.260126521 | 0.024592293 | 0.059 | 0.038 | 1 |
| ARL4C | 0.263576707 | 0.032968344 | 0.052 | 0.038 | 1 |
| S100A10 | 0.264031488 | -0.099053385 | 0.455 | 0.598 | 1 |
| TIMM9 | 0.265592326 | -0.063306631 | 0.802 | 0.832 | 1 |
| CD8A | 0.268816964 | 0.057504416 | 0.076 | 0.07 | 1 |
| IL2RB | 0.268994969 | -0.083076641 | 0.25 | 0.282 | 1 |
| IL9 | 0.269949741 | -0.032273976 | 0.031 | 0.029 | 1 |
| CD6 | 0.270303067 | 0.050238949 | 0.181 | 0.157 | 1 |
| CXCL5 | 0.270498291 | 0.057183733 | 0.372 | 0.322 | 1 |
| LAT2 | 0.270557054 | -0.073382866 | 0.285 | 0.332 | 1 |
| ITK | 0.275125053 | -0.090488644 | 0.375 | 0.433 | 1 |
| VNN2 | 0.280409509 | -0.013358095 | 0.007 | 0.014 | 1 |
| LTB | 0.281850065 | -0.011199433 | 0.003 | 0.009 | 1 |
| IL12RB1 | 0.282023647 | -0.011199433 | 0.003 | 0.009 | 1 |
| SMAD2 | 0.283070091 | -0.045863528 | 0.212 | 0.24 | 1 |
| CHD7 | 0.289225632 | -0.032131824 | 0.101 | 0.126 | 1 |
| CHD1 | 0.299559897 | -0.020208356 | 0.031 | 0.042 | 1 |
| FUT4 | 0.299615399 | -0.014973604 | 0.014 | 0.023 | 1 |

| | | | | | |
|---|---|---|---|---|---|
| IL1B | 0.299906372 | -0.086241495 | 0.417 | 0.467 | 1 |
| LIG4 | 0.304670087 | 0.042163318 | 0.326 | 0.306 | 1 |
| DOCK8 | 0.307744246 | 0.048868574 | 0.264 | 0.226 | 1 |
| CD86 | 0.310938201 | 0.11297547 | 0.542 | 0.473 | 1 |
| NR4A2 | 0.311631644 | 0.100267625 | 0.358 | 0.315 | 1 |
| KLRC1 | 0.313887017 | -0.042779676 | 0.049 | 0.052 | 1 |
| TNFRSF17 | 0.32197146 | -0.030755673 | 0.003 | 0.004 | 1 |
| TGFB1 | 0.322191938 | 0.010864808 | 0.021 | 0.013 | 1 |
| TNIK | 0.322570242 | 0.019136006 | 0.038 | 0.027 | 1 |
| TIAF1 | 0.331501491 | 0.018602068 | 0.052 | 0.04 | 1 |
| FBXO11 | 0.337442472 | -0.045356613 | 0.312 | 0.347 | 1 |
| SLC7A1 | 0.34784535 | -0.044638566 | 0.944 | 0.948 | 1 |
| SDCBP | 0.34833357 | 0.069424234 | 0.743 | 0.725 | 1 |
| CD274 | 0.348804346 | 0.066255253 | 0.858 | 0.846 | 1 |
| EIF5 | 0.349292105 | 0.048764603 | 0.931 | 0.92 | 1 |
| PMCH | 0.35016975 | -0.025448389 | 0.003 | 0.004 | 1 |
| ATF6B | 0.355692041 | -0.037706758 | 0.16 | 0.169 | 1 |
| S100A9 | 0.356515601 | -0.020542297 | 0.028 | 0.041 | 1 |
| IL18 | 0.358298547 | 0.006827545 | 0.01 | 0.006 | 1 |
| MCM2 | 0.361352169 | -0.061149886 | 0.156 | 0.227 | 1 |
| CD160 | 0.361788086 | -0.013358095 | 0.007 | 0.01 | 1 |
| BCL6 | 0.365289749 | 0.013059145 | 0.031 | 0.021 | 1 |
| THBS1 | 0.373157324 | -0.024938969 | 0.035 | 0.045 | 1 |
| TNFSF14 | 0.374391007 | 0.024292388 | 0.066 | 0.054 | 1 |
| NT5E | 0.380592737 | 0.041671112 | 0.149 | 0.119 | 1 |
| TSPAN32 | 0.388137549 | -0.00979148 | 0.007 | 0.012 | 1 |
| TNFRSF8 | 0.395611394 | -0.032646011 | 0.083 | 0.08 | 1 |
| CBLB | 0.397599844 | -0.013933508 | 0.021 | 0.028 | 1 |
| CXCL3 | 0.399927582 | -0.038602172 | 0.163 | 0.176 | 1 |
| S100A12 | 0.402804643 | 0.007727823 | 0.007 | 0.004 | 1 |
| TNFSF13B | 0.402804643 | 0.007727823 | 0.007 | 0.004 | 1 |
| IGKC | 0.413659125 | -0.150905645 | 0.583 | 0.543 | 1 |
| EVI2B | 0.419195301 | -0.047244313 | 0.33 | 0.332 | 1 |
| JUN | 0.419323321 | -0.0391095 | 0.181 | 0.211 | 1 |
| DPP4 | 0.426925181 | -0.006717619 | 0.003 | 0.008 | 1 |
| CARD11 | 0.428086897 | -0.011435417 | 0.014 | 0.019 | 1 |
| CXCR3 | 0.428530657 | -0.033217727 | 0.104 | 0.122 | 1 |
| VSIR | 0.436695265 | -0.055616031 | 0.326 | 0.303 | 1 |
| BIN2 | 0.44033134 | -0.008898447 | 0.007 | 0.011 | 1 |
| CHI3L2 | 0.441141229 | 0.005927829 | 0.01 | 0.006 | 1 |
| RPN2 | 0.449548676 | -0.047586814 | 0.712 | 0.693 | 1 |
| SPAG9 | 0.451231593 | -0.042495645 | 0.521 | 0.572 | 1 |
| PSEN1 | 0.453893303 | -0.017424294 | 0.052 | 0.063 | 1 |
| GZMA | 0.457535918 | -0.032307965 | 0.024 | 0.033 | 1 |
| TGFBI | 0.458978537 | -0.019590049 | 0.069 | 0.073 | 1 |
| FN1 | 0.45926089 | -0.027804231 | 0.122 | 0.134 | 1 |
| CXCL9 | 0.478140629 | 0.136687352 | 0.094 | 0.1 | 1 |

| | | | | | |
|---|---|---|---|---|---|
| CD44 | 0.480801697 | -0.036223581 | 1 | 1 | 1 |
| EIF3J | 0.485027877 | -0.036213664 | 0.625 | 0.651 | 1 |
| IL21 | 0.486992589 | 0.03620527 | 0.066 | 0.056 | 1 |
| CD5 | 0.509214046 | 0.016266361 | 0.066 | 0.062 | 1 |
| SKAP1 | 0.516031194 | 0.006842904 | 0.017 | 0.013 | 1 |
| IRF1 | 0.517812695 | 0.03636262 | 0.986 | 0.976 | 1 |
| IFITM3 | 0.520500825 | 0.014231782 | 0.049 | 0.048 | 1 |
| PTPN7 | 0.523956562 | -0.030020526 | 0.198 | 0.21 | 1 |
| IL23R | 0.526289814 | 0.004563813 | 0.007 | 0.003 | 1 |
| MAP2K3 | 0.527853211 | -0.042130586 | 0.91 | 0.917 | 1 |
| IL31 | 0.536237103 | 0.009079294 | 0.014 | 0.012 | 1 |
| TGFBR1 | 0.536481798 | 0.02757155 | 0.344 | 0.316 | 1 |
| NANS | 0.542757035 | -0.036683272 | 0.594 | 0.577 | 1 |
| NINJ2 | 0.544003611 | 0.007295987 | 0.017 | 0.016 | 1 |
| CD52 | 0.551120891 | 0.06314807 | 0.615 | 0.609 | 1 |
| PDIA4 | 0.554670601 | -0.049493166 | 0.451 | 0.447 | 1 |
| NFATC3 | 0.560056564 | 0.022555526 | 0.177 | 0.169 | 1 |
| SRGN | 0.573565944 | -0.036275947 | 0.885 | 0.885 | 1 |
| GIMAP2 | 0.578265096 | -0.01608463 | 0.059 | 0.073 | 1 |
| LRRC32 | 0.580966051 | 0.058595137 | 0.323 | 0.306 | 1 |
| DUSP1 | 0.592144614 | -0.032779041 | 0.42 | 0.428 | 1 |
| APOBEC3G | 0.594839157 | -0.045694179 | 0.542 | 0.552 | 1 |
| THEMIS | 0.596950492 | 0.006386861 | 0.01 | 0.008 | 1 |
| ZNF341 | 0.599417116 | -0.009411405 | 0.024 | 0.031 | 1 |
| TYMS | 0.610440178 | 0.012604513 | 0.017 | 0.016 | 1 |
| MMP9 | 0.612080584 | -0.012193504 | 0.059 | 0.065 | 1 |
| PAK1IP1 | 0.615286947 | -0.029762632 | 0.549 | 0.534 | 1 |
| ITGAM | 0.621093678 | -0.008156355 | 0.021 | 0.028 | 1 |
| LIPA | 0.624278502 | -0.02386198 | 0.201 | 0.217 | 1 |
| BLNK | 0.624840379 | -0.020388866 | 0.115 | 0.097 | 1 |
| IL1R1 | 0.634143191 | 0.014536898 | 0.059 | 0.06 | 1 |
| CASP5 | 0.634916882 | -0.012615777 | 0.076 | 0.086 | 1 |
| IL12RB2 | 0.636728701 | -0.02540619 | 0.184 | 0.155 | 1 |
| FTH1 | 0.643604114 | 0.034705102 | 0.986 | 0.98 | 1 |
| KLRF1 | 0.644636281 | -0.003122115 | 0.003 | 0.006 | 1 |
| MYB | 0.647980627 | -0.011482029 | 0.052 | 0.052 | 1 |
| CD247 | 0.65196688 | 0.00640516 | 0.014 | 0.014 | 1 |
| TLR7 | 0.653466468 | -0.007000446 | 0.014 | 0.013 | 1 |
| CNOT2 | 0.656053557 | 0.022766951 | 0.444 | 0.451 | 1 |
| IL17F | 0.659657271 | -0.008418738 | 0.035 | 0.036 | 1 |
| HLA.DQB1 | 0.667403391 | 0.031114761 | 0.99 | 0.989 | 1 |
| TMEM97 | 0.677632185 | 0.013915148 | 0.108 | 0.099 | 1 |
| FCER2 | 0.678309844 | -0.037681763 | 0.33 | 0.306 | 1 |
| SLC7A7 | 0.680701905 | -0.009046338 | 0.049 | 0.049 | 1 |
| TRAF1 | 0.683556369 | 0.023555891 | 0.948 | 0.923 | 1 |
| RORA | 0.684547633 | 0.015221045 | 0.066 | 0.074 | 1 |
| TNFRSF9 | 0.685565353 | -0.005617217 | 0.017 | 0.019 | 1 |

| | | | | | |
|---|---|---|---|---|---|
| FYB1 | 0.6915039 | -0.006784342 | 0.028 | 0.029 | 1 |
| MALT1 | 0.69212724 | 0.016603611 | 0.215 | 0.209 | 1 |
| NRP1 | 0.692968752 | 0.007343199 | 0.049 | 0.042 | 1 |
| IRF9 | 0.69820804 | 0.024816955 | 0.688 | 0.643 | 1 |
| IL7R | 0.70161824 | -0.013692014 | 0.083 | 0.092 | 1 |
| IL13 | 0.701964655 | -0.018333332 | 0.111 | 0.085 | 1 |
| CCR4 | 0.709374737 | -0.015329664 | 0.094 | 0.095 | 1 |
| MITF | 0.715751504 | 0.00276157 | 0.003 | 0.005 | 1 |
| FOXP3 | 0.71575151 | 0.00276157 | 0.007 | 0.004 | 1 |
| CCND2 | 0.716704215 | 0.02973617 | 0.698 | 0.664 | 1 |
| WAS | 0.721992833 | 0.021639951 | 0.253 | 0.234 | 1 |
| AHI1 | 0.726983626 | 0.006902298 | 0.042 | 0.036 | 1 |
| TNFSF10 | 0.730620741 | -0.004730769 | 0.014 | 0.019 | 1 |
| TNFSF8 | 0.73101349 | -0.010991081 | 0.052 | 0.041 | 1 |
| PAXBP1 | 0.73509078 | 0.017023184 | 0.392 | 0.367 | 1 |
| BCL10 | 0.736486174 | 0.011790036 | 0.16 | 0.142 | 1 |
| UFM1 | 0.742693476 | 0.016482266 | 0.948 | 0.961 | 1 |
| SOCS3 | 0.745708093 | -0.019468605 | 0.573 | 0.575 | 1 |
| LY86 | 0.756701535 | 0.01256547 | 0.135 | 0.124 | 1 |
| CCR5 | 0.764404177 | -0.005708786 | 0.042 | 0.044 | 1 |
| CD36 | 0.765482055 | 0.005136903 | 0.035 | 0.033 | 1 |
| TNFRSF4 | 0.768538375 | -0.016915915 | 0.205 | 0.248 | 1 |
| CCL13 | 0.771468846 | 0.001384902 | 0.003 | 0.003 | 1 |
| CD209 | 0.771468846 | 0.001384902 | 0.003 | 0.003 | 1 |
| ITGAX | 0.786059185 | 0.027397986 | 0.229 | 0.342 | 1 |
| IL5 | 0.793407762 | -0.002631689 | 0.007 | 0.007 | 1 |
| IL6R | 0.79451046 | 0.008678267 | 0.115 | 0.107 | 1 |
| CSRNP1 | 0.800051729 | 0.016042793 | 0.812 | 0.777 | 1 |
| OSM | 0.800968563 | -0.006540381 | 0.066 | 0.068 | 1 |
| CD2 | 0.809058766 | 0.018291786 | 0.455 | 0.442 | 1 |
| PGM3 | 0.816918901 | 0.00745097 | 0.111 | 0.09 | 1 |
| CTSW | 0.816945827 | 0.004684849 | 0.028 | 0.024 | 1 |
| LDHA | 0.819556475 | -0.013442564 | 1 | 0.999 | 1 |
| LAG3 | 0.819877284 | 0.009495716 | 0.049 | 0.051 | 1 |
| CD244 | 0.821252653 | 0.001384902 | 0.003 | 0.002 | 1 |
| BLK | 0.826108488 | -0.001320997 | 0.003 | 0.004 | 1 |
| KLRG1 | 0.836993245 | -0.00375655 | 0.024 | 0.02 | 1 |
| CSF1 | 0.837495158 | 0.013672405 | 0.292 | 0.264 | 1 |
| IFRD1 | 0.838684349 | 0.010508696 | 0.417 | 0.396 | 1 |
| CD300A | 0.845491602 | -0.004152487 | 0.024 | 0.021 | 1 |
| MGST1 | 0.847231268 | 0.002804748 | 0.014 | 0.008 | 1 |
| LCK | 0.851663678 | 0.004325764 | 0.049 | 0.04 | 1 |
| IL11RA | 0.858255435 | 0.003911426 | 0.049 | 0.048 | 1 |
| GAB2 | 0.86094664 | -0.008572724 | 0.253 | 0.241 | 1 |
| IL4 | 0.861176091 | 0.005288822 | 0.052 | 0.06 | 1 |
| IFITM2 | 0.86731276 | 0.014533724 | 0.597 | 0.567 | 1 |
| STAT1 | 0.869688311 | -0.010902749 | 0.736 | 0.752 | 1 |

| | | | | | |
|---|---|---|---|---|---|
| IL18RAP | 0.873180728 | 0.002889358 | 0.024 | 0.02 | 1 |
| BCL11B | 0.876224812 | 0.003945142 | 0.049 | 0.05 | 1 |
| EEF1E1 | 0.877104491 | 0.008567064 | 0.469 | 0.473 | 1 |
| NDFIP2 | 0.882080219 | 0.003012078 | 0.049 | 0.042 | 1 |
| CCR6 | 0.882877845 | 0.00284734 | 0.021 | 0.011 | 1 |
| RGS1 | 0.892503126 | -0.018357365 | 0.625 | 0.614 | 1 |
| CD24 | 0.894032956 | -0.005590999 | 0.132 | 0.129 | 1 |
| HLA.DRA | 0.89530498 | -0.009342316 | 0.993 | 0.996 | 1 |
| VEGFA | 0.897377257 | -0.001992864 | 0.021 | 0.024 | 1 |
| CST7 | 0.903750831 | 0.008420379 | 0.167 | 0.203 | 1 |
| GIMAP5 | 0.912862887 | -0.00224285 | 0.042 | 0.037 | 1 |
| KLRK1 | 0.91550098 | -0.007386247 | 0.212 | 0.199 | 1 |
| RORC | 0.923523144 | -0.001660421 | 0.014 | 0.008 | 1 |
| CD7 | 0.935182166 | 0.002411714 | 0.059 | 0.075 | 1 |
| CXCL16 | 0.941957341 | 0.002062883 | 0.094 | 0.088 | 1 |
| IRF7 | 0.951970978 | -0.003807425 | 0.5 | 0.458 | 1 |
| BCL2A1 | 0.957703936 | -0.002622296 | 1 | 0.999 | 1 |
| PSMB9 | 0.964128791 | -0.002929027 | 0.767 | 0.746 | 1 |
| FOSL1 | 0.96485486 | 0.002518596 | 0.882 | 0.861 | 1 |
| PSME3 | 0.965485446 | 0.002551954 | 0.764 | 0.762 | 1 |
| NFKB1 | 0.967233353 | -0.001645485 | 1 | 1 | 1 |
| IRF3 | 0.980402533 | -0.000506755 | 0.038 | 0.036 | 1 |
| CD14 | 0.983477118 | -0.000290337 | 0.017 | 0.014 | 1 |
| SOCS1 | 0.99117481 | -0.014409059 | 0 | 0.009 | 1 |
| VPS28 | 0.99327812 | -0.00056765 | 0.483 | 0.442 | 1 |
| IL17RB | 0.994239528 | -0.018882536 | 0 | 0.008 | 1 |
| GATA3 | 0.994446772 | -0.009921668 | 0 | 0.006 | 1 |
| PCLAF | 0.994446772 | -0.009921668 | 0 | 0.006 | 1 |
| TGFB3 | 0.994641039 | -0.005420276 | 0 | 0.004 | 1 |
| TNIP3 | 0.994641039 | -0.005420276 | 0 | 0.003 | 1 |
| EOMES | 0.994699473 | -0.004518309 | 0 | 0.003 | 1 |
| THBD | 0.994699473 | -0.004518309 | 0 | 0.003 | 1 |
| CLEC4E | 0.996634035 | -0.003615779 | 0 | 0.003 | 1 |
| NFATC4 | 0.996634035 | -0.003615779 | 0 | 0.002 | 1 |
| KCNE3 | 0.996689959 | -0.002712683 | 0 | 0.002 | 1 |
| ZNF683 | 0.996689959 | -0.002712683 | 0 | 0.002 | 1 |
| CLEC10A | 0.996689959 | -0.002712683 | 0 | 0.002 | 1 |
| ITGAE | 0.997140854 | 7.72E-05 | 0.056 | 0.056 | 1 |

Monocytes

| | p_val | avg_log2FC | Patient | Controls | p_val_adj |
|---|---|---|---|---|---|
| CD9 | 8.72E-25 | -1.363133152 | 0.662 | 0.812 | 4.34E-22 |
| FCGR3A | 3.87E-23 | 1.268009281 | 0.802 | 0.603 | 1.93E-20 |
| HLA.A | 2.48E-18 | 0.550019701 | 1 | 1 | 1.23E-15 |
| NAMPT | 1.91E-14 | 0.708491559 | 1 | 0.998 | 9.53E-12 |
| FN1 | 2.05E-14 | -2.369728418 | 0.198 | 0.481 | 1.02E-11 |

| | | | | | |
|---|---|---|---|---|---|
| TRAC | 9.96E-14 | 0.905375155 | 0.633 | 0.49 | 4.96E-11 |
| GZMH | 3.47E-13 | 1.152579867 | 0.251 | 0.145 | 1.73E-10 |
| FCER2 | 2.26E-12 | 1.204024436 | 0.454 | 0.238 | 1.13E-09 |
| HIF1A | 3.51E-11 | 0.54371891 | 0.894 | 0.843 | 1.75E-08 |
| SRGN | 4.27E-11 | 0.445044877 | 0.995 | 0.986 | 2.13E-08 |
| MGST1 | 1.55E-10 | -0.716371678 | 0.222 | 0.49 | 7.71E-08 |
| ITGA4 | 4.19E-10 | 0.412317254 | 0.401 | 0.215 | 2.09E-07 |
| THBS1 | 1.33E-09 | 1.529008929 | 0.522 | 0.367 | 6.64E-07 |
| LAT | 1.35E-09 | -0.737320309 | 0.464 | 0.582 | 6.72E-07 |
| ATF4 | 3.33E-09 | 0.401203906 | 1 | 0.998 | 1.66E-06 |
| IL6 | 5.66E-09 | -0.896017131 | 0.309 | 0.353 | 2.82E-06 |
| IFNG | 9.46E-09 | -1.044889291 | 0.99 | 0.991 | 4.71E-06 |
| TREM1 | 1.54E-08 | -0.51945908 | 0.169 | 0.342 | 7.65E-06 |
| LTA | 2.17E-08 | -0.878993845 | 0.444 | 0.487 | 1.08E-05 |
| IVNS1ABP | 4.79E-08 | 0.46705986 | 0.952 | 0.95 | 2.39E-05 |
| IRF4 | 4.87E-08 | 0.674688314 | 0.758 | 0.733 | 2.42E-05 |
| S100A12 | 9.27E-08 | -0.570672583 | 0.203 | 0.361 | 4.61E-05 |
| ITGAM | 9.42E-08 | -0.442904882 | 0.169 | 0.361 | 4.69E-05 |
| MMP9 | 1.60E-07 | -0.899055012 | 0.589 | 0.761 | 7.97E-05 |
| TGFBR2 | 2.09E-07 | 0.448957308 | 0.546 | 0.362 | 0.000104314 |
| SPAG9 | 2.64E-07 | 0.341136007 | 0.957 | 0.963 | 0.000131549 |
| CD3D | 2.70E-07 | 0.72889267 | 0.57 | 0.495 | 0.000134316 |
| CXCL16 | 4.06E-07 | 0.477811564 | 0.913 | 0.93 | 0.00020221 |
| VMO1 | 5.44E-07 | 1.14470679 | 0.594 | 0.402 | 0.000270918 |
| GNLY | 7.49E-07 | 1.218040224 | 0.459 | 0.377 | 0.000372809 |
| B4GALT5 | 9.47E-07 | 0.29444021 | 0.585 | 0.415 | 0.000471559 |
| IL3 | 1.30E-06 | -0.786596362 | 0.097 | 0.146 | 0.000646587 |
| IRF7 | 1.72E-06 | 0.344748639 | 0.976 | 0.948 | 0.000855676 |
| FOSB | 2.09E-06 | 0.581675803 | 0.643 | 0.645 | 0.001039863 |
| ICOS | 2.74E-06 | 0.455797577 | 0.473 | 0.409 | 0.001365343 |
| CD2 | 2.79E-06 | 0.491543485 | 0.623 | 0.546 | 0.001391167 |
| KLRG1 | 3.11E-06 | 0.146203947 | 0.097 | 0.026 | 0.001549047 |
| CXCL5 | 3.76E-06 | -1.317763719 | 0.594 | 0.597 | 0.001872169 |
| CXCL1 | 4.19E-06 | 1.64378778 | 0.333 | 0.318 | 0.002085933 |
| TRBC2 | 5.27E-06 | 0.597618214 | 0.754 | 0.715 | 0.002623149 |
| CSF2 | 5.33E-06 | -0.827861289 | 0.937 | 0.936 | 0.002653705 |
| PTPRC | 6.87E-06 | 0.395012596 | 0.937 | 0.848 | 0.003420729 |
| S100A9 | 7.20E-06 | -0.64104673 | 0.589 | 0.752 | 0.003584543 |
| FOSL1 | 9.54E-06 | 0.399964621 | 0.836 | 0.795 | 0.004749306 |
| GZMB | 1.14E-05 | 0.787786479 | 0.966 | 0.976 | 0.00569362 |
| DUSP2 | 1.41E-05 | 0.572561686 | 0.957 | 0.926 | 0.007030725 |
| POU2F2 | 2.01E-05 | 0.243896984 | 0.304 | 0.201 | 0.01002717 |
| NFKB1 | 2.13E-05 | 0.416730005 | 0.986 | 0.956 | 0.010599588 |
| NR4A2 | 2.53E-05 | 0.602416516 | 0.546 | 0.473 | 0.012620627 |
| HAVCR2 | 2.54E-05 | 0.529320319 | 0.744 | 0.66 | 0.012634671 |
| EIF5 | 2.65E-05 | 0.30350298 | 0.952 | 0.94 | 0.013195799 |
| EIF1 | 2.98E-05 | 0.246408598 | 1 | 0.999 | 0.01485244 |

| | | | | | |
|---|---|---|---|---|---|
| CD69 | 3.18E-05 | 0.445453107 | 0.995 | 0.981 | 0.015859152 |
| IL1RN | 3.57E-05 | -0.844384794 | 0.691 | 0.777 | 0.01778431 |
| LYN | 3.57E-05 | 0.403169362 | 0.71 | 0.594 | 0.017797937 |
| PFN1 | 3.68E-05 | 0.307625147 | 1 | 1 | 0.018326714 |
| CD5 | 3.76E-05 | 0.281458507 | 0.164 | 0.088 | 0.018736295 |
| CLEC4E | 6.48E-05 | -0.315920689 | 0.121 | 0.228 | 0.032269516 |
| IL9 | 7.27E-05 | 0.187900464 | 0.058 | 0.029 | 0.036218004 |
| CD3E | 8.33E-05 | 0.405556446 | 0.406 | 0.316 | 0.041482806 |
| SNX8 | 9.13E-05 | 0.252343144 | 0.966 | 0.952 | 0.045476134 |
| CD44 | 9.52E-05 | -0.264839384 | 1 | 0.998 | 0.047426465 |
| STAT6 | 9.74E-05 | 0.244073519 | 0.444 | 0.321 | 0.048516198 |
| FOS | 0.000121848 | 0.880580267 | 0.686 | 0.582 | 0.060680511 |
| IL4 | 0.000144613 | -0.435918947 | 0.058 | 0.08 | 0.072017292 |
| CD8A | 0.000156679 | 0.568537171 | 0.188 | 0.124 | 0.078026355 |
| ZNF683 | 0.000167864 | 0.053495347 | 0.039 | 0.005 | 0.083596136 |
| IFNGR1 | 0.000170164 | 0.284937508 | 0.705 | 0.596 | 0.084741909 |
| SDCBP | 0.000172397 | 0.210542196 | 0.995 | 1 | 0.085853784 |
| IFRD1 | 0.0001789 | 0.276121953 | 0.435 | 0.373 | 0.089092174 |
| S100A10 | 0.000188717 | -0.388033894 | 0.932 | 0.972 | 0.093981188 |
| FYB1 | 0.000219869 | 0.370701031 | 0.522 | 0.439 | 0.109494709 |
| HLA.DRA | 0.000220439 | 0.494579556 | 0.986 | 0.942 | 0.109778452 |
| CX3CR1 | 0.000228835 | 0.365743616 | 0.164 | 0.1 | 0.113960067 |
| RORA | 0.000249093 | 0.317690444 | 0.208 | 0.129 | 0.124048522 |
| EGR3 | 0.000264173 | 0.313343175 | 0.382 | 0.276 | 0.131558084 |
| CD14 | 0.000266461 | 0.728432838 | 0.411 | 0.414 | 0.13269777 |
| NRP1 | 0.000274997 | -0.4044931 | 0.705 | 0.819 | 0.136948571 |
| CD7 | 0.000296049 | 0.207598474 | 0.208 | 0.117 | 0.14743261 |
| PRNP | 0.000300479 | 0.371201901 | 0.855 | 0.774 | 0.149638501 |
| ITK | 0.000337728 | 0.44163788 | 0.512 | 0.505 | 0.168188304 |
| NFKBIZ | 0.000379737 | 0.403519656 | 0.812 | 0.745 | 0.189108998 |
| B3GAT1 | 0.000417064 | 0.191161988 | 0.155 | 0.072 | 0.207697926 |
| CREM | 0.000541504 | 0.33328624 | 0.928 | 0.905 | 0.269669231 |
| LAP3 | 0.00067295 | 0.304244697 | 0.976 | 0.917 | 0.335128906 |
| ICAM1 | 0.000675967 | 0.319309483 | 0.976 | 0.984 | 0.336631688 |
| CCR7 | 0.000710141 | -0.582870031 | 0.464 | 0.442 | 0.353650418 |
| NT5E | 0.000747352 | -0.372183787 | 0.159 | 0.245 | 0.372181153 |
| BCAT1 | 0.000807845 | -0.268647739 | 0.729 | 0.816 | 0.402306683 |
| PSEN1 | 0.000850676 | 0.174440777 | 0.435 | 0.345 | 0.423636662 |
| IL32 | 0.000999599 | 0.312567522 | 0.28 | 0.147 | 0.497800353 |
| CXCR4 | 0.001145435 | -0.494870475 | 0.271 | 0.362 | 0.570426415 |
| CCL3 | 0.001194555 | 0.370516354 | 1 | 1 | 0.594888296 |
| NFATC1 | 0.001292243 | 0.229818119 | 0.396 | 0.242 | 0.643537116 |
| IL7R | 0.001349216 | -0.27694138 | 0.232 | 0.322 | 0.671909572 |
| EEF1E1 | 0.001429728 | -0.186661545 | 0.159 | 0.249 | 0.712004486 |
| LEF1 | 0.001531233 | 0.164250154 | 0.13 | 0.095 | 0.762553956 |
| ERBIN | 0.001594111 | 0.216195758 | 0.942 | 0.926 | 0.79386749 |
| CCL2 | 0.001629374 | 0.57146127 | 0.928 | 0.928 | 0.811428348 |

| | | | | | |
|---|---|---|---|---|---|
| TIGIT | 0.001911248 | 0.198102761 | 0.159 | 0.083 | 0.951801426 |
| SELL | 0.002017524 | 0.213454616 | 0.28 | 0.197 | 1 |
| TNFRSF4 | 0.002185416 | 0.265713844 | 0.314 | 0.266 | 1 |
| APOBEC3G | 0.002214947 | 0.536825566 | 0.604 | 0.559 | 1 |
| IL2RB | 0.002359689 | 0.470619069 | 0.425 | 0.353 | 1 |
| CSRNP1 | 0.002451105 | 0.212713257 | 0.971 | 0.973 | 1 |
| HLA.DMA | 0.002497895 | 0.384592997 | 0.778 | 0.633 | 1 |
| CXCL3 | 0.002587608 | 0.670249251 | 0.72 | 0.695 | 1 |
| TRIB2 | 0.002601375 | 0.173710483 | 0.121 | 0.08 | 1 |
| CD72 | 0.002966648 | -0.126371226 | 0.019 | 0.061 | 1 |
| HLA.DQB1 | 0.003276295 | 0.500188706 | 0.826 | 0.639 | 1 |
| HLA.DPA1 | 0.003326835 | 0.507871583 | 0.889 | 0.711 | 1 |
| IL1R1 | 0.003652305 | 0.303961872 | 0.734 | 0.762 | 1 |
| KLRK1 | 0.00378475 | 0.479078724 | 0.401 | 0.325 | 1 |
| AQP9 | 0.003854636 | -0.378003399 | 0.367 | 0.545 | 1 |
| CASP5 | 0.003911455 | 0.1811779 | 0.696 | 0.625 | 1 |
| PASK | 0.0039588 | 0.114979746 | 0.039 | 0.035 | 1 |
| IFITM2 | 0.003960184 | 0.287934677 | 0.469 | 0.403 | 1 |
| GBP2 | 0.004359535 | 0.233885807 | 0.932 | 0.868 | 1 |
| WAS | 0.005354328 | 0.17834765 | 0.671 | 0.599 | 1 |
| IGLC3 | 0.005366571 | -0.284256152 | 0.406 | 0.369 | 1 |
| LIPA | 0.005565926 | 0.438951307 | 0.831 | 0.768 | 1 |
| STAT1 | 0.005831099 | 0.203453844 | 0.981 | 0.981 | 1 |
| PTPN7 | 0.006619506 | 0.235202911 | 0.512 | 0.519 | 1 |
| CXCL10 | 0.007275036 | 0.978478625 | 0.512 | 0.366 | 1 |
| CD63 | 0.007822621 | 0.165358848 | 0.995 | 0.999 | 1 |
| IFI6 | 0.007864994 | -0.29772672 | 0.391 | 0.362 | 1 |
| CD38 | 0.008013685 | 0.052410858 | 0.043 | 0.014 | 1 |
| CD74 | 0.008132036 | 0.391877549 | 0.981 | 0.934 | 1 |
| CCL20 | 0.008154703 | -0.493600906 | 0.589 | 0.55 | 1 |
| F5 | 0.008224579 | 0.142799887 | 0.13 | 0.091 | 1 |
| IL22 | 0.008390087 | -0.22642257 | 0.092 | 0.087 | 1 |
| PIK3AP1 | 0.008406985 | 0.264276746 | 0.667 | 0.603 | 1 |
| ARID5A | 0.008687771 | 0.184788378 | 0.739 | 0.708 | 1 |
| TGFBR1 | 0.008712273 | 0.16652579 | 0.473 | 0.377 | 1 |
| LCK | 0.009042787 | 0.099424587 | 0.097 | 0.056 | 1 |
| MCM2 | 0.010340308 | 0.08469654 | 0.121 | 0.066 | 1 |
| CD160 | 0.010757017 | -0.087382904 | 0.014 | 0.049 | 1 |
| IER3 | 0.011069342 | 0.30657423 | 0.995 | 0.981 | 1 |
| ZNF341 | 0.01225536 | 0.033520094 | 0.034 | 0.01 | 1 |
| IRF3 | 0.012629454 | 0.067297174 | 0.097 | 0.053 | 1 |
| RAPH1 | 0.013134561 | 0.137723421 | 0.242 | 0.18 | 1 |
| TLR2 | 0.013542902 | 0.24756954 | 0.865 | 0.874 | 1 |
| ENTPD1 | 0.01367045 | 0.074779574 | 0.063 | 0.025 | 1 |
| GNAI2 | 0.013899804 | 0.17325649 | 0.986 | 0.985 | 1 |
| IL12A | 0.013921577 | -0.297483143 | 0.019 | 0.023 | 1 |
| GAB2 | 0.014625219 | 0.177551446 | 0.56 | 0.515 | 1 |

| | | | | | |
|---|---|---|---|---|---|
| PSME2 | 0.015165205 | 0.191416498 | 0.99 | 0.952 | 1 |
| BCL11B | 0.015768161 | 0.077342089 | 0.097 | 0.06 | 1 |
| IL2RG | 0.016164796 | 0.294863089 | 0.749 | 0.747 | 1 |
| JUN | 0.016308454 | 0.266755812 | 0.768 | 0.742 | 1 |
| MAP2K3 | 0.017593858 | 0.171253026 | 0.995 | 0.99 | 1 |
| CCL22 | 0.01945929 | 0.741921757 | 0.217 | 0.258 | 1 |
| ITGAE | 0.020096212 | -0.211169223 | 0.222 | 0.27 | 1 |
| IGHG1 | 0.022896797 | -0.171886518 | 0.15 | 0.159 | 1 |
| MS4A1 | 0.023187091 | -0.082557403 | 0.039 | 0.072 | 1 |
| TNFRSF13C | 0.02406634 | 0.107423428 | 0.159 | 0.097 | 1 |
| PSMB9 | 0.025397478 | 0.181191747 | 0.855 | 0.808 | 1 |
| JAK3 | 0.025468252 | 0.175847633 | 0.449 | 0.383 | 1 |
| NINJ2 | 0.025500945 | 0.055608567 | 0.068 | 0.039 | 1 |
| CXCR3 | 0.026454585 | 0.224993157 | 0.242 | 0.168 | 1 |
| IL6R | 0.028163424 | 0.156918584 | 0.826 | 0.813 | 1 |
| LY86 | 0.028344855 | 0.203464765 | 0.357 | 0.321 | 1 |
| ANXA5 | 0.029419106 | -0.118838879 | 0.995 | 0.997 | 1 |
| CXCL2 | 0.029726315 | 0.470791991 | 0.802 | 0.822 | 1 |
| TNIK | 0.03122507 | -0.118860809 | 0.126 | 0.192 | 1 |
| EVI2A | 0.031727151 | -0.195398457 | 0.725 | 0.773 | 1 |
| IL11RA | 0.03358989 | 0.035023526 | 0.034 | 0.013 | 1 |
| TRDC | 0.034374292 | 0.182332192 | 0.082 | 0.064 | 1 |
| CD52 | 0.034653181 | -0.294572417 | 0.778 | 0.773 | 1 |
| PAXBP1 | 0.036562721 | 0.111508692 | 0.251 | 0.203 | 1 |
| JAK1 | 0.037846331 | 0.137543907 | 0.976 | 0.964 | 1 |
| GAPDH | 0.038302519 | -0.134709864 | 1 | 0.998 | 1 |
| LGALS1 | 0.038538973 | -0.219750883 | 0.942 | 0.96 | 1 |
| LAG3 | 0.044644129 | 0.246586734 | 0.159 | 0.111 | 1 |
| THEMIS | 0.045985262 | 0.044626513 | 0.039 | 0.018 | 1 |
| CD36 | 0.046588104 | 0.295291764 | 0.623 | 0.58 | 1 |
| HNRNPC | 0.049415193 | 0.149455067 | 0.937 | 0.877 | 1 |
| RNASE2 | 0.051140462 | 0.026528299 | 0.019 | 0.005 | 1 |
| TCF4 | 0.055172257 | 0.030691995 | 0.019 | 0.006 | 1 |
| IL18RAP | 0.056226167 | -0.0905597 | 0.043 | 0.09 | 1 |
| SPP1 | 0.057151991 | -0.600411727 | 0.072 | 0.234 | 1 |
| NKG7 | 0.05795879 | 0.424378989 | 0.464 | 0.453 | 1 |
| TRAT1 | 0.060404203 | 0.037993421 | 0.039 | 0.019 | 1 |
| GBP1 | 0.063014448 | 0.214763977 | 0.942 | 0.929 | 1 |
| TGFB1 | 0.063581001 | -0.070125243 | 0.068 | 0.111 | 1 |
| FOXP1 | 0.063923676 | 0.114361278 | 0.271 | 0.239 | 1 |
| MYB | 0.06399965 | 0.05541425 | 0.048 | 0.025 | 1 |
| CD6 | 0.064845631 | 0.187676003 | 0.304 | 0.245 | 1 |
| PSMB8 | 0.065353031 | 0.132239631 | 0.942 | 0.918 | 1 |
| CTSD | 0.066938012 | 0.259471376 | 0.816 | 0.843 | 1 |
| CD37 | 0.068658753 | 0.129901637 | 0.575 | 0.55 | 1 |
| SOCS3 | 0.06926046 | 0.14339749 | 0.85 | 0.827 | 1 |
| TIMM9 | 0.069782452 | -0.136101075 | 0.449 | 0.493 | 1 |

| | | | | | |
|---|---|---|---|---|---|
| SOCS2 | 0.070938309 | 0.160567937 | 0.179 | 0.123 | 1 |
| EBF1 | 0.074582777 | -0.052134436 | 0.029 | 0.053 | 1 |
| CD22 | 0.078528608 | -0.123134004 | 0.179 | 0.253 | 1 |
| CD209 | 0.079255012 | 0.197840656 | 0.193 | 0.113 | 1 |
| CD274 | 0.079507419 | 0.179885444 | 0.894 | 0.906 | 1 |
| IRF1 | 0.082674597 | 0.114278696 | 0.995 | 0.991 | 1 |
| YBX3 | 0.083171108 | 0.115273415 | 0.3 | 0.27 | 1 |
| ZBED2 | 0.084860256 | -0.071151125 | 0.034 | 0.044 | 1 |
| RPN2 | 0.08558569 | -0.132351343 | 0.841 | 0.863 | 1 |
| BATF | 0.086506483 | 0.156563076 | 0.372 | 0.376 | 1 |
| FTH1 | 0.087648465 | -0.105386887 | 1 | 1 | 1 |
| CD226 | 0.102001458 | 0.126251689 | 0.188 | 0.214 | 1 |
| TNFRSF9 | 0.102599342 | 0.050181631 | 0.039 | 0.024 | 1 |
| IKZF1 | 0.102968977 | 0.047021637 | 0.063 | 0.054 | 1 |
| ILF2 | 0.103257217 | -0.116587669 | 0.966 | 0.965 | 1 |
| BIRC3 | 0.104729783 | -0.257336172 | 0.725 | 0.669 | 1 |
| FCER1G | 0.104759449 | -0.134317955 | 0.957 | 0.966 | 1 |
| RBCK1 | 0.107687073 | 0.106181771 | 0.459 | 0.453 | 1 |
| CD79A | 0.109147493 | -0.0502289 | 0.024 | 0.038 | 1 |
| BCL10 | 0.112998234 | 0.095828557 | 0.498 | 0.462 | 1 |
| TCF7 | 0.114489163 | -0.04148148 | 0.024 | 0.044 | 1 |
| FURIN | 0.114658457 | -0.135410153 | 0.942 | 0.922 | 1 |
| BTG1 | 0.115554228 | -0.157026361 | 0.971 | 0.992 | 1 |
| FOXO1 | 0.115795825 | 0.041401207 | 0.058 | 0.038 | 1 |
| CD83 | 0.115950956 | 0.204917204 | 0.923 | 0.878 | 1 |
| KLRC3 | 0.117729491 | 0.257514682 | 0.179 | 0.155 | 1 |
| RORC | 0.119135208 | -0.044387223 | 0.01 | 0.02 | 1 |
| CCND2 | 0.119414103 | 0.141988293 | 0.338 | 0.276 | 1 |
| IGKC | 0.120050953 | 0.178888487 | 0.251 | 0.228 | 1 |
| NFKB2 | 0.122207461 | 0.101895323 | 0.353 | 0.288 | 1 |
| REL | 0.12287894 | 0.163170343 | 0.758 | 0.696 | 1 |
| NANS | 0.126312706 | -0.101095302 | 0.986 | 0.959 | 1 |
| CXCL11 | 0.128744711 | 0.547927249 | 0.188 | 0.104 | 1 |
| CD40LG | 0.12997072 | -0.106836897 | 0.111 | 0.099 | 1 |
| PDIA6 | 0.130123027 | -0.120459911 | 0.961 | 0.962 | 1 |
| LRRC8B | 0.130977559 | 0.087430906 | 0.271 | 0.219 | 1 |
| LDHA | 0.131013406 | -0.155253018 | 0.986 | 0.981 | 1 |
| TNFRSF25 | 0.131420087 | 0.084842191 | 0.193 | 0.168 | 1 |
| JCHAIN | 0.131448587 | 0.059270516 | 0.111 | 0.087 | 1 |
| KLRC1 | 0.13314192 | -0.223524391 | 0.14 | 0.121 | 1 |
| STIM1 | 0.141336885 | 0.062355909 | 0.179 | 0.125 | 1 |
| IL18 | 0.145597008 | 0.069019968 | 0.092 | 0.087 | 1 |
| TGIF1 | 0.146490532 | 0.122339824 | 0.821 | 0.845 | 1 |
| CCR1 | 0.148743574 | -0.119586843 | 0.952 | 0.962 | 1 |
| IL1B | 0.15507248 | -0.308204301 | 0.797 | 0.834 | 1 |
| CD163 | 0.156116849 | 0.179283257 | 0.478 | 0.505 | 1 |
| IL13 | 0.157304464 | 0.209445625 | 0.135 | 0.104 | 1 |

| | | | | | |
|---|---|---|---|---|---|
| CYCS | 0.158141867 | -0.100569301 | 0.942 | 0.941 | 1 |
| JAK2 | 0.159684758 | 0.055683207 | 0.121 | 0.086 | 1 |
| CXCL8 | 0.161794782 | 0.189947326 | 1 | 0.995 | 1 |
| SLC7A1 | 0.167732931 | 0.109261422 | 0.633 | 0.498 | 1 |
| BCL6 | 0.17035854 | 0.055137352 | 0.14 | 0.122 | 1 |
| IGHG3 | 0.174342343 | -0.026994571 | 0.005 | 0.015 | 1 |
| PMCH | 0.181306046 | -0.093815459 | 0.005 | 0.006 | 1 |
| LAMP3 | 0.181522758 | -0.378009771 | 0.14 | 0.127 | 1 |
| RGS1 | 0.182322436 | 0.263460369 | 0.43 | 0.473 | 1 |
| POU2AF1 | 0.191052447 | -0.074661005 | 0.087 | 0.114 | 1 |
| CCR5 | 0.1928943 | 0.165038463 | 0.739 | 0.762 | 1 |
| BCL2A1 | 0.193849257 | 0.145940412 | 0.981 | 0.979 | 1 |
| IFITM3 | 0.194382302 | 0.17821669 | 0.802 | 0.684 | 1 |
| VSIG4 | 0.196501 | 0.123635741 | 0.121 | 0.124 | 1 |
| CSF1 | 0.199259763 | 0.248371066 | 0.599 | 0.531 | 1 |
| FUT4 | 0.202046903 | 0.044703272 | 0.106 | 0.097 | 1 |
| TRAF1 | 0.206266451 | 0.139446433 | 0.652 | 0.603 | 1 |
| IL3RA | 0.210905722 | 0.129248604 | 0.899 | 0.91 | 1 |
| PDIA4 | 0.211019128 | -0.125301645 | 0.836 | 0.866 | 1 |
| CD70 | 0.213249174 | 0.100562586 | 0.048 | 0.028 | 1 |
| IL6ST | 0.213511582 | -0.08546487 | 0.614 | 0.636 | 1 |
| PRF1 | 0.21627593 | 0.253290878 | 0.676 | 0.621 | 1 |
| CXCR5 | 0.219507297 | -0.097541407 | 0.285 | 0.24 | 1 |
| CCL4 | 0.224742295 | 0.19235402 | 1 | 1 | 1 |
| NFATC2 | 0.225142448 | 0.055950831 | 0.116 | 0.088 | 1 |
| ADA | 0.225393203 | 0.137175338 | 0.44 | 0.441 | 1 |
| CD244 | 0.228319186 | 0.031538399 | 0.029 | 0.02 | 1 |
| EVI2B | 0.228785844 | -0.110517927 | 0.681 | 0.682 | 1 |
| CD28 | 0.229191999 | 0.030250853 | 0.039 | 0.02 | 1 |
| ARPC1B | 0.229418372 | 0.088324531 | 0.952 | 0.947 | 1 |
| LGALS9 | 0.238191443 | 0.108441292 | 0.836 | 0.789 | 1 |
| IL15RA | 0.239043246 | 0.127480561 | 0.498 | 0.42 | 1 |
| CBLB | 0.244182265 | 0.041869207 | 0.097 | 0.081 | 1 |
| SNX9 | 0.245075732 | 0.088447643 | 0.937 | 0.933 | 1 |
| GRB10 | 0.245171124 | -0.066929368 | 0.082 | 0.098 | 1 |
| NDFIP2 | 0.248416304 | 0.034386524 | 0.063 | 0.056 | 1 |
| TYMS | 0.249864217 | -0.01663406 | 0.005 | 0.015 | 1 |
| MAST4 | 0.254647991 | 0.030451956 | 0.072 | 0.057 | 1 |
| UFM1 | 0.255501695 | 0.073977286 | 0.986 | 0.977 | 1 |
| PGM3 | 0.25575276 | -0.087245881 | 0.309 | 0.377 | 1 |
| CLEC4D | 0.261060522 | -0.0179332 | 0.005 | 0.013 | 1 |
| MITF | 0.261116621 | 0.091957626 | 0.401 | 0.375 | 1 |
| CD4 | 0.26972239 | 0.105631133 | 0.449 | 0.36 | 1 |
| VNN2 | 0.274419673 | -0.03741231 | 0.029 | 0.043 | 1 |
| CCR4 | 0.274885151 | 0.059925035 | 0.126 | 0.095 | 1 |
| IL2RA | 0.276396049 | 0.120800044 | 0.251 | 0.197 | 1 |
| CD200 | 0.277278041 | 0.141929254 | 0.3 | 0.266 | 1 |

| | | | | | |
|---|---|---|---|---|---|
| CCR6 | 0.278417528 | 0.021282129 | 0.01 | 0.007 | 1 |
| FBXO11 | 0.281651251 | 0.067125692 | 0.551 | 0.511 | 1 |
| CST7 | 0.285696226 | 0.270195152 | 0.377 | 0.317 | 1 |
| LILRB4 | 0.286089903 | -0.105995979 | 0.696 | 0.734 | 1 |
| FASLG | 0.288037261 | -0.141789495 | 0.3 | 0.276 | 1 |
| IL18R1 | 0.301588088 | -0.047868159 | 0.063 | 0.079 | 1 |
| IGHM | 0.313262739 | -0.020460399 | 0.019 | 0.03 | 1 |
| ORAI1 | 0.315792597 | -0.080820906 | 0.507 | 0.525 | 1 |
| TGFBI | 0.320653525 | -0.120051796 | 0.879 | 0.892 | 1 |
| IL21 | 0.320990462 | -0.095494508 | 0.092 | 0.082 | 1 |
| DPP4 | 0.323414823 | -0.012729605 | 0.005 | 0.013 | 1 |
| GZMA | 0.324461188 | -0.126486728 | 0.116 | 0.086 | 1 |
| TNFSF13B | 0.325093216 | 0.082503794 | 0.208 | 0.133 | 1 |
| CNOT2 | 0.327634135 | 0.058768092 | 0.469 | 0.438 | 1 |
| CD109 | 0.327764474 | 0.083382324 | 0.783 | 0.851 | 1 |
| IL2 | 0.329460852 | -0.134553468 | 1 | 1 | 1 |
| NGDN | 0.331097153 | 0.019731365 | 0.043 | 0.027 | 1 |
| SLC1A5 | 0.331529563 | -0.098129285 | 0.725 | 0.805 | 1 |
| IL1A | 0.331555181 | -0.276615636 | 0.203 | 0.294 | 1 |
| SELPLG | 0.343269528 | 0.062291482 | 0.203 | 0.141 | 1 |
| IL31 | 0.344967421 | -0.02731407 | 0.014 | 0.016 | 1 |
| EGR1 | 0.344994487 | 0.141572876 | 0.657 | 0.564 | 1 |
| LAMP1 | 0.351464709 | -0.09547684 | 0.894 | 0.847 | 1 |
| FCN1 | 0.352937414 | -0.162099907 | 0.382 | 0.453 | 1 |
| IRF9 | 0.369155193 | -0.056181584 | 0.937 | 0.94 | 1 |
| PIK3IP1 | 0.370126821 | -0.062561527 | 0.198 | 0.224 | 1 |
| ARL4C | 0.371658924 | 0.094985049 | 0.271 | 0.269 | 1 |
| CTSW | 0.373052165 | 0.051558063 | 0.126 | 0.08 | 1 |
| AHI1 | 0.376213609 | -0.034626343 | 0.077 | 0.106 | 1 |
| ATF6B | 0.38425166 | -0.051337585 | 0.386 | 0.44 | 1 |
| CXCL9 | 0.384284777 | 0.415941004 | 0.28 | 0.208 | 1 |
| BCL2 | 0.386980744 | 0.053246413 | 0.222 | 0.229 | 1 |
| CHD1 | 0.395474159 | 0.021015473 | 0.034 | 0.022 | 1 |
| IL23A | 0.402711881 | -0.12130173 | 0.314 | 0.271 | 1 |
| IL12RB1 | 0.410256949 | -0.01447862 | 0.014 | 0.022 | 1 |
| CD1C | 0.41117981 | -0.025562774 | 0.005 | 0.02 | 1 |
| RNASE6 | 0.413306298 | -0.06470536 | 0.285 | 0.321 | 1 |
| SP140 | 0.41357638 | -0.035538537 | 0.097 | 0.108 | 1 |
| PDCD1 | 0.41439662 | 0.023282962 | 0.058 | 0.036 | 1 |
| KCNE3 | 0.416519563 | -0.029573115 | 0.039 | 0.071 | 1 |
| IL17F | 0.41703054 | -0.041813066 | 0.068 | 0.055 | 1 |
| IGHD | 0.419413305 | -0.010120753 | 0.005 | 0.01 | 1 |
| TSPAN32 | 0.422822354 | 0.01689247 | 0.039 | 0.022 | 1 |
| BTLA | 0.42789137 | 0.025013932 | 0.043 | 0.045 | 1 |
| CD247 | 0.429072672 | 0.029831373 | 0.082 | 0.053 | 1 |
| DUSP4 | 0.44937775 | 0.095023908 | 0.585 | 0.559 | 1 |
| TIAF1 | 0.449863698 | -0.029934648 | 0.082 | 0.1 | 1 |

| | | | | | |
|---|---|---|---|---|---|
| LRRC32 | 0.45316253 | 0.025811788 | 0.029 | 0.032 | 1 |
| EIF3J | 0.45592887 | 0.044713788 | 0.406 | 0.412 | 1 |
| TNIP3 | 0.459424768 | 0.015606072 | 0.034 | 0.024 | 1 |
| IGBP1 | 0.464461924 | -0.036426786 | 0.251 | 0.292 | 1 |
| AIM2 | 0.469279546 | 0.009233702 | 0.019 | 0.013 | 1 |
| TNFSF8 | 0.47527836 | 0.037879258 | 0.087 | 0.077 | 1 |
| STAT5A | 0.484765251 | 0.09094539 | 0.681 | 0.688 | 1 |
| PSME3 | 0.488056491 | 0.044281283 | 0.498 | 0.47 | 1 |
| TBX21 | 0.496073948 | -0.089178107 | 0.638 | 0.682 | 1 |
| GATA3 | 0.496268094 | 0.009233702 | 0.014 | 0.013 | 1 |
| ITGAX | 0.503787195 | 0.042795888 | 0.971 | 0.975 | 1 |
| CD33 | 0.50895721 | 0.024830251 | 0.101 | 0.083 | 1 |
| TNFSF10 | 0.516381115 | -0.106039193 | 0.213 | 0.161 | 1 |
| DUSP1 | 0.526709543 | -0.080067083 | 0.942 | 0.948 | 1 |
| QPCT | 0.531081504 | -0.027141075 | 0.077 | 0.093 | 1 |
| PLCG2 | 0.546821704 | 0.033691081 | 0.353 | 0.328 | 1 |
| SLC25A37 | 0.547796243 | -0.067783866 | 0.758 | 0.813 | 1 |
| GZMK | 0.548473786 | -0.01277284 | 0.019 | 0.021 | 1 |
| STAT4 | 0.549831567 | -0.065559739 | 0.372 | 0.35 | 1 |
| MZB1 | 0.556396589 | 0.009233702 | 0.019 | 0.01 | 1 |
| DOCK8 | 0.560659654 | 0.04192159 | 0.401 | 0.375 | 1 |
| BLNK | 0.571648352 | 0.011753747 | 0.034 | 0.027 | 1 |
| CHD7 | 0.576642294 | -0.011952342 | 0.019 | 0.036 | 1 |
| STAT3 | 0.578168178 | 0.029545385 | 0.99 | 0.991 | 1 |
| ZAP70 | 0.579927468 | 0.008249565 | 0.024 | 0.018 | 1 |
| RUNX3 | 0.580580407 | -0.056493254 | 0.628 | 0.672 | 1 |
| LIF | 0.589316932 | 0.102894917 | 0.28 | 0.324 | 1 |
| TNFRSF8 | 0.59147672 | 0.007929865 | 0.014 | 0.013 | 1 |
| LTB | 0.59510561 | 0.008916948 | 0.014 | 0.005 | 1 |
| TUBA1C | 0.599220815 | -0.04175145 | 0.995 | 0.988 | 1 |
| CCL5 | 0.599413622 | 0.09887525 | 0.295 | 0.26 | 1 |
| CD3G | 0.607060673 | 0.006298625 | 0.014 | 0.009 | 1 |
| IGHG2 | 0.610315307 | -0.011013786 | 0.01 | 0.008 | 1 |
| GIMAP2 | 0.611423363 | -0.036115961 | 0.425 | 0.417 | 1 |
| CD27 | 0.612871644 | 0.009547537 | 0.024 | 0.013 | 1 |
| BIN2 | 0.616304666 | -0.019928757 | 0.145 | 0.144 | 1 |
| CMKLR1 | 0.620733256 | 0.01292536 | 0.053 | 0.042 | 1 |
| LGALS3 | 0.620742956 | -0.046270228 | 0.908 | 0.915 | 1 |
| TMEM97 | 0.621906037 | 0.010166607 | 0.034 | 0.022 | 1 |
| IRF8 | 0.641302875 | -0.035671969 | 0.99 | 0.993 | 1 |
| CD300A | 0.641509546 | 0.059041272 | 0.343 | 0.313 | 1 |
| CD40 | 0.641572899 | -0.07428623 | 0.667 | 0.56 | 1 |
| TPSAB1 | 0.64949452 | -0.012729605 | 0.005 | 0.003 | 1 |
| LAT2 | 0.664627871 | 0.033182276 | 0.836 | 0.837 | 1 |
| DUSP5 | 0.668415645 | -0.028790247 | 0.995 | 0.997 | 1 |
| GEM | 0.672545889 | 0.061909756 | 0.652 | 0.69 | 1 |
| NFATC3 | 0.676393706 | 0.020975328 | 0.184 | 0.167 | 1 |

| | | | | | |
|---|---|---|---|---|---|
| IL12RB2 | 0.677580556 | 0.024661772 | 0.087 | 0.091 | 1 |
| CTLA4 | 0.684197161 | 0.05134688 | 0.382 | 0.332 | 1 |
| ITGB2 | 0.6843209 | 0.056731809 | 0.812 | 0.714 | 1 |
| NCAM1 | 0.690149727 | 0.010774472 | 0.024 | 0.028 | 1 |
| TNF | 0.690295168 | -0.056932843 | 0.986 | 0.991 | 1 |
| MYC | 0.720140991 | 0.047532387 | 0.507 | 0.551 | 1 |
| PCNA | 0.723755091 | -0.024169757 | 0.362 | 0.357 | 1 |
| CD80 | 0.72874079 | -0.024043555 | 0.087 | 0.083 | 1 |
| BACH2 | 0.741092069 | -0.015002974 | 0.111 | 0.114 | 1 |
| TRIB1 | 0.741913038 | -0.028372245 | 0.918 | 0.94 | 1 |
| DCLRE1C | 0.743256015 | -0.016122266 | 0.184 | 0.19 | 1 |
| VEGFA | 0.752209225 | -0.010429318 | 0.043 | 0.056 | 1 |
| KLRC4 | 0.75672909 | -0.005425372 | 0.014 | 0.014 | 1 |
| TLR8 | 0.75790466 | -0.005044098 | 0.019 | 0.022 | 1 |
| TNFSF14 | 0.761756367 | -0.02289175 | 0.28 | 0.274 | 1 |
| TNFAIP8 | 0.762114444 | 0.032149196 | 0.947 | 0.951 | 1 |
| VSIR | 0.763933343 | 0.037783615 | 0.507 | 0.425 | 1 |
| NFKBID | 0.77822996 | 0.009942803 | 0.097 | 0.09 | 1 |
| CARD11 | 0.804499265 | 0.001677819 | 0.005 | 0.004 | 1 |
| IGHA1 | 0.804499265 | 0.001677819 | 0.005 | 0.004 | 1 |
| CD86 | 0.809415206 | -0.038927601 | 0.546 | 0.542 | 1 |
| FYN | 0.809593442 | -0.024344993 | 0.58 | 0.571 | 1 |
| CLEC10A | 0.810729198 | -0.022794583 | 0.082 | 0.103 | 1 |
| OSM | 0.813687933 | -0.03037849 | 0.169 | 0.161 | 1 |
| ADGRE1 | 0.816530775 | -0.007850702 | 0.029 | 0.037 | 1 |
| DUSP6 | 0.819462451 | 0.018190394 | 0.961 | 0.957 | 1 |
| LIG4 | 0.82329516 | -0.011662918 | 0.3 | 0.31 | 1 |
| BAX | 0.824701503 | 0.015465208 | 0.57 | 0.556 | 1 |
| SLC7A7 | 0.82569376 | -0.019231171 | 0.343 | 0.382 | 1 |
| PRDM1 | 0.830914949 | 0.020023108 | 0.512 | 0.561 | 1 |
| FAS | 0.831685917 | -0.012504529 | 0.15 | 0.134 | 1 |
| SKAP1 | 0.833681461 | -0.002265769 | 0.005 | 0.005 | 1 |
| CD48 | 0.842702655 | -0.017695307 | 0.937 | 0.963 | 1 |
| EIF5A | 0.845891639 | -0.015072312 | 0.971 | 0.962 | 1 |
| GIMAP5 | 0.862099043 | 0.019731365 | 0.551 | 0.53 | 1 |
| CR2 | 0.863007015 | -0.004291997 | 0.029 | 0.022 | 1 |
| JUNB | 0.874928266 | -0.013925969 | 0.754 | 0.733 | 1 |
| IL15 | 0.878735327 | -0.011965746 | 0.246 | 0.201 | 1 |
| IL5 | 0.898749966 | -0.00282826 | 0.014 | 0.008 | 1 |
| CCL13 | 0.904457042 | 0.025905533 | 0.063 | 0.038 | 1 |
| IL4R | 0.904513133 | -0.008926262 | 0.85 | 0.914 | 1 |
| SOCS1 | 0.905147242 | -0.000952437 | 0.005 | 0.005 | 1 |
| ZBTB16 | 0.917210148 | -0.000952437 | 0.005 | 0.005 | 1 |
| UGCG | 0.920293377 | -0.010580869 | 0.609 | 0.593 | 1 |
| KDELR1 | 0.921248018 | -0.006836544 | 0.652 | 0.634 | 1 |
| IL1R2 | 0.924573515 | 0.004132242 | 0.024 | 0.03 | 1 |
| MALT1 | 0.926656123 | 0.006411919 | 0.473 | 0.484 | 1 |

| | | | | | |
|---|---|---|---|---|---|
| VPS28 | 0.930026261 | 0.00604512 | 0.618 | 0.609 | 1 |
| NFKBIA | 0.931411735 | -0.00941764 | 0.995 | 0.998 | 1 |
| PAK1IP1 | 0.934236495 | -0.004893482 | 0.275 | 0.256 | 1 |
| KLRF1 | 0.940784148 | -0.001729482 | 0.029 | 0.022 | 1 |
| FOXP3 | 0.94926881 | 0.000720807 | 0.005 | 0.009 | 1 |
| ABCE1 | 0.949687945 | 0.004530098 | 0.86 | 0.781 | 1 |
| CYBB | 0.94997826 | 0.005829668 | 0.894 | 0.914 | 1 |
| SNCA | 0.952608219 | -0.002492085 | 0.087 | 0.096 | 1 |
| RPL6 | 0.952719145 | 0.004193476 | 1 | 1 | 1 |
| CHI3L2 | 0.958298874 | 0.000720807 | 0.01 | 0.006 | 1 |
| XBP1 | 0.964332654 | -0.003381585 | 0.855 | 0.827 | 1 |
| IL23R | 0.967407151 | 0.000362091 | 0.005 | 0.004 | 1 |
| SMAD2 | 0.96803313 | -0.002356984 | 0.435 | 0.414 | 1 |
| SLA | 0.968254169 | 0.004263116 | 0.652 | 0.633 | 1 |
| THBD | 0.972985999 | -0.003253562 | 0.159 | 0.136 | 1 |
| BCL2L1 | 0.977057545 | 0.00242163 | 0.507 | 0.507 | 1 |
| ARHGEF3 | 0.98068893 | 0.001863884 | 0.193 | 0.201 | 1 |
| MCM4 | 0.990481208 | 0.000258108 | 0.039 | 0.036 | 1 |
| MME | 0.992271613 | -0.02488596 | 0 | 0.016 | 1 |
| TCL1A | 0.992271613 | -0.02488596 | 0 | 0.012 | 1 |
| APOE | 0.992348598 | -0.020985025 | 0 | 0.011 | 1 |
| CD24 | 0.992408417 | -0.018378529 | 0 | 0.012 | 1 |
| AURKB | 0.995215135 | -0.013151368 | 0 | 0.009 | 1 |
| CCR3 | 0.995403476 | -0.006590669 | 0 | 0.005 | 1 |
| PCLAF | 0.995464109 | -0.005274942 | 0 | 0.004 | 1 |
| CD8B | 0.995464109 | -0.005274942 | 0 | 0.004 | 1 |
| IKZF2 | 0.995464109 | -0.005274942 | 0 | 0.004 | 1 |
| TOP2A | 0.995464109 | -0.005274942 | 0 | 0.004 | 1 |
| EOMES | 0.995464109 | -0.005274942 | 0 | 0.004 | 1 |
| IL26 | 0.995464109 | -0.005274942 | 0 | 0.004 | 1 |
| CXCL13 | 0.996640489 | -0.075914977 | 0 | 0.006 | 1 |
| CD79B | 0.997017202 | -0.007905198 | 0 | 0.003 | 1 |
| F13A1 | 0.997017202 | -0.007905198 | 0 | 0.003 | 1 |
| PTGDR2 | 0.997131438 | -0.003958014 | 0 | 0.003 | 1 |
| ADGRG3 | 0.997131438 | -0.003958014 | 0 | 0.003 | 1 |

NK cells

| | p_val | avg_log2FC | Patient | Controls | p_val_adj |
|---|---|---|---|---|---|
| FCGR3A | 2.47E-22 | 0.694878833 | 0.37 | 0.077 | 1.23E-19 |
| HLA.A | 5.14E-19 | 0.438285293 | 1 | 1 | 2.56E-16 |
| BTG1 | 2.19E-15 | -0.556990603 | 0.994 | 0.999 | 1.09E-12 |
| GZMH | 2.13E-11 | 0.696579044 | 0.58 | 0.369 | 1.06E-08 |
| IL2 | 6.81E-11 | 0.722922646 | 1 | 1 | 3.39E-08 |
| CD83 | 8.04E-11 | -0.981458739 | 0.679 | 0.784 | 4.00E-08 |
| PTPRC | 5.17E-10 | 0.536422354 | 0.815 | 0.747 | 2.57E-07 |
| RUNX3 | 6.39E-10 | 0.559423914 | 0.667 | 0.498 | 3.18E-07 |

| | | | | | |
|---|---|---|---|---|---|
| SNX9 | 6.51E-10 | -0.513992699 | 0.58 | 0.778 | 3.24E-07 |
| VSIR | 1.67E-09 | -0.522936616 | 0.642 | 0.875 | 8.33E-07 |
| LDHA | 5.20E-09 | -0.398168327 | 1 | 0.999 | 2.59E-06 |
| IGLC3 | 9.52E-09 | 0.872938282 | 0.29 | 0.274 | 4.74E-06 |
| CXCR4 | 1.55E-08 | -0.861732408 | 0.377 | 0.566 | 7.72E-06 |
| GNAI2 | 2.15E-08 | 0.450941045 | 0.877 | 0.808 | 1.07E-05 |
| ITGA4 | 3.08E-08 | 0.369923105 | 0.352 | 0.201 | 1.53E-05 |
| SELPLG | 7.98E-08 | 0.304805754 | 0.198 | 0.06 | 3.97E-05 |
| GAPDH | 1.06E-07 | -0.41440459 | 0.988 | 0.997 | 5.30E-05 |
| S100A10 | 1.37E-07 | 0.52874286 | 0.852 | 0.757 | 6.83E-05 |
| IVNS1ABP | 1.53E-07 | 0.414353169 | 0.679 | 0.557 | 7.64E-05 |
| RPL6 | 1.88E-07 | -0.311608655 | 1 | 1 | 9.39E-05 |
| DUSP5 | 2.63E-07 | -0.398882597 | 0.957 | 0.976 | 0.000130826 |
| NFKBIA | 2.86E-07 | -0.563841603 | 1 | 0.993 | 0.000142294 |
| IRF8 | 3.88E-07 | -0.46297022 | 0.951 | 0.955 | 0.000193142 |
| CYCS | 4.38E-07 | -0.396089826 | 0.833 | 0.881 | 0.000217977 |
| IL2RG | 4.74E-07 | 0.317774787 | 0.963 | 0.957 | 0.00023617 |
| ICAM1 | 6.62E-07 | -0.667220944 | 0.685 | 0.799 | 0.000329745 |
| TUBA1C | 1.17E-06 | -0.441916042 | 0.772 | 0.87 | 0.00058315 |
| IL32 | 1.20E-06 | 0.353231329 | 0.358 | 0.201 | 0.000598224 |
| RGS1 | 1.25E-06 | 0.730672407 | 0.846 | 0.733 | 0.000620197 |
| TNF | 1.63E-06 | -0.720515279 | 0.889 | 0.906 | 0.000813364 |
| PFN1 | 2.73E-06 | 0.289723521 | 1 | 1 | 0.001358214 |
| SOCS2 | 3.78E-06 | 0.411606522 | 0.302 | 0.171 | 0.00188106 |
| CD74 | 5.50E-06 | 0.466372718 | 0.802 | 0.771 | 0.002737898 |
| IFITM2 | 7.93E-06 | 0.464004348 | 0.568 | 0.433 | 0.003951254 |
| NFKB1 | 2.42E-05 | -0.261055289 | 0.988 | 0.997 | 0.012043806 |
| ABCE1 | 2.64E-05 | -0.322422931 | 0.519 | 0.697 | 0.013142018 |
| ARPC1B | 4.11E-05 | 0.287878008 | 0.568 | 0.418 | 0.020488025 |
| GNLY | 4.62E-05 | 0.671049647 | 0.815 | 0.753 | 0.022999038 |
| PSMB9 | 4.85E-05 | 0.277746194 | 0.352 | 0.264 | 0.024134501 |
| ITGB2 | 4.98E-05 | 0.404202786 | 0.488 | 0.379 | 0.024811388 |
| CST7 | 5.32E-05 | 0.348941606 | 0.951 | 0.865 | 0.026476439 |
| IL18RAP | 5.57E-05 | -0.315475645 | 0.272 | 0.457 | 0.027762248 |
| CD160 | 5.77E-05 | -0.246366351 | 0.025 | 0.146 | 0.028741623 |
| PRNP | 8.97E-05 | -0.31360323 | 0.741 | 0.83 | 0.044657217 |
| REL | 0.000105379 | -0.333266472 | 0.889 | 0.923 | 0.052478883 |
| ITGAM | 0.000175625 | 0.242883584 | 0.253 | 0.133 | 0.087461014 |
| KLRC1 | 0.000194201 | -0.524991059 | 0.488 | 0.588 | 0.096711958 |
| DUSP2 | 0.000211132 | -0.418219922 | 0.926 | 0.97 | 0.105143642 |
| CD3E | 0.0002236 | 0.282828212 | 0.414 | 0.278 | 0.111352565 |
| HLA.DRA | 0.00022533 | 0.337501627 | 0.63 | 0.544 | 0.112214242 |
| IL18R1 | 0.000238767 | -0.257290182 | 0.136 | 0.27 | 0.118906129 |
| STAT4 | 0.000270993 | -0.274662801 | 0.821 | 0.895 | 0.134954566 |
| STAT5A | 0.000297502 | -0.302443522 | 0.895 | 0.929 | 0.148156114 |
| TRAF1 | 0.000301151 | -0.274088344 | 0.716 | 0.834 | 0.14997299 |
| HNRNPC | 0.000301957 | -0.262954657 | 0.741 | 0.847 | 0.150374547 |

| | | | | | |
|---|---|---|---|---|---|
| FASLG | 0.000349333 | -0.356484951 | 0.759 | 0.845 | 0.173967809 |
| NKG7 | 0.000398447 | -0.274908214 | 0.957 | 0.983 | 0.198426609 |
| SLC7A1 | 0.000399724 | -0.264622356 | 0.611 | 0.745 | 0.199062507 |
| LAG3 | 0.000480015 | 0.251160022 | 0.136 | 0.061 | 0.239047453 |
| PRF1 | 0.000539245 | -0.273623745 | 0.963 | 0.993 | 0.268543968 |
| CCL4 | 0.000540591 | -0.439574555 | 1 | 1 | 0.269214178 |
| ILF2 | 0.00055161 | -0.252365601 | 0.883 | 0.899 | 0.274701993 |
| FYN | 0.000580251 | -0.281848351 | 0.735 | 0.781 | 0.288965201 |
| TRDC | 0.000594237 | 0.309105185 | 0.37 | 0.205 | 0.295930006 |
| FOSL1 | 0.000771267 | -0.28070475 | 0.42 | 0.539 | 0.384091069 |
| HAVCR2 | 0.00081801 | -0.392925683 | 0.537 | 0.591 | 0.407369106 |
| HLA.DPA1 | 0.000911571 | 0.218380626 | 0.389 | 0.293 | 0.453962513 |
| JAK1 | 0.001061544 | -0.268309843 | 0.932 | 0.956 | 0.52864895 |
| DUSP6 | 0.001086003 | -0.374322486 | 0.352 | 0.446 | 0.540829728 |
| MYC | 0.001130987 | -0.44617235 | 0.636 | 0.733 | 0.563231619 |
| CCR7 | 0.001457897 | -0.335861239 | 0.333 | 0.495 | 0.726032508 |
| EIF3J | 0.001689163 | -0.205785064 | 0.253 | 0.351 | 0.841203302 |
| IFNG | 0.001773726 | -0.49591549 | 1 | 0.997 | 0.883315673 |
| XBP1 | 0.00185671 | -0.229702975 | 0.753 | 0.818 | 0.924641333 |
| IGKC | 0.002016281 | -0.426427824 | 0.198 | 0.17 | 1 |
| STAT1 | 0.002086835 | 0.203779476 | 0.389 | 0.276 | 1 |
| IL1RN | 0.002303512 | -0.45024364 | 0.222 | 0.244 | 1 |
| IL21 | 0.002724716 | 0.274765872 | 0.062 | 0.052 | 1 |
| IL12RB2 | 0.002746378 | -0.2992731 | 0.265 | 0.432 | 1 |
| MAP2K3 | 0.002792669 | -0.241929952 | 0.926 | 0.936 | 1 |
| TIGIT | 0.003013574 | 0.174725386 | 0.198 | 0.132 | 1 |
| TGIF1 | 0.003092962 | -0.240182161 | 0.383 | 0.493 | 1 |
| UFM1 | 0.003361396 | -0.234825509 | 0.852 | 0.865 | 1 |
| CSF2 | 0.003522158 | -0.567501242 | 0.895 | 0.94 | 1 |
| ARL4C | 0.003615116 | -0.267049435 | 0.21 | 0.27 | 1 |
| ITGAX | 0.004982735 | 0.364598361 | 0.519 | 0.405 | 1 |
| BACH2 | 0.006177017 | -0.21959186 | 0.333 | 0.476 | 1 |
| ATF6B | 0.006356806 | 0.109087842 | 0.13 | 0.072 | 1 |
| JAK3 | 0.008343085 | 0.181986839 | 0.543 | 0.42 | 1 |
| IL4R | 0.008555055 | -0.208521184 | 0.852 | 0.887 | 1 |
| CCL5 | 0.009341574 | -0.292607412 | 0.704 | 0.778 | 1 |
| CASP5 | 0.009398388 | 0.076011672 | 0.105 | 0.053 | 1 |
| CD2 | 0.009407416 | 0.225015417 | 0.543 | 0.498 | 1 |
| RORA | 0.009411139 | 0.234887714 | 0.265 | 0.174 | 1 |
| LCK | 0.009462858 | 0.138837003 | 0.21 | 0.151 | 1 |
| SLC25A37 | 0.009649416 | -0.120234816 | 0.056 | 0.114 | 1 |
| IRF1 | 0.009752259 | 0.232678802 | 0.704 | 0.566 | 1 |
| IRF9 | 0.010830475 | 0.194198159 | 0.549 | 0.474 | 1 |
| CD8A | 0.011008264 | 0.257743 | 0.235 | 0.186 | 1 |
| VPS28 | 0.011740034 | 0.137789684 | 0.265 | 0.175 | 1 |
| BCL2 | 0.012659493 | 0.172896632 | 0.407 | 0.329 | 1 |
| ORAI1 | 0.014044639 | -0.168625448 | 0.346 | 0.436 | 1 |

| | | | | | |
|---|---|---|---|---|---|
| IGHG1 | 0.014368345 | -0.237480102 | 0.086 | 0.1 | 1 |
| B3GAT1 | 0.014568568 | 0.213344011 | 0.327 | 0.274 | 1 |
| CD44 | 0.015770451 | -0.181228944 | 1 | 0.999 | 1 |
| SNX8 | 0.016482134 | -0.145227167 | 0.154 | 0.229 | 1 |
| ANXA5 | 0.017168829 | -0.26782621 | 0.778 | 0.811 | 1 |
| FOSB | 0.019297468 | -0.235492404 | 0.16 | 0.288 | 1 |
| CD70 | 0.020760955 | 0.056836824 | 0.056 | 0.017 | 1 |
| CSRNP1 | 0.022415588 | -0.16801009 | 0.914 | 0.94 | 1 |
| CD52 | 0.022461303 | 0.268394749 | 0.63 | 0.502 | 1 |
| GBP1 | 0.022633356 | 0.164715275 | 0.321 | 0.247 | 1 |
| TRIB1 | 0.024336464 | -0.22789779 | 0.253 | 0.335 | 1 |
| CX3CR1 | 0.025074732 | -0.22973861 | 0.191 | 0.273 | 1 |
| PRDM1 | 0.02935875 | 0.169943047 | 0.401 | 0.321 | 1 |
| CD40LG | 0.03004782 | -0.085651284 | 0.043 | 0.089 | 1 |
| CD300A | 0.030483578 | -0.264203072 | 0.463 | 0.503 | 1 |
| CXCL10 | 0.032445886 | -0.110732822 | 0.08 | 0.144 | 1 |
| WAS | 0.034272056 | 0.097069076 | 0.167 | 0.127 | 1 |
| POU2F2 | 0.03681536 | -0.140292944 | 0.235 | 0.331 | 1 |
| CD200 | 0.039371401 | -0.134488291 | 0.142 | 0.18 | 1 |
| CD63 | 0.040517972 | 0.167257911 | 0.877 | 0.858 | 1 |
| CD3D | 0.04054114 | 0.195750012 | 0.414 | 0.422 | 1 |
| IFRD1 | 0.04106814 | 0.094309835 | 0.216 | 0.148 | 1 |
| PSEN1 | 0.042531597 | 0.068873407 | 0.13 | 0.081 | 1 |
| TNFAIP8 | 0.043358576 | -0.178337292 | 0.963 | 0.971 | 1 |
| BAX | 0.044042768 | -0.131508072 | 0.265 | 0.348 | 1 |
| IL4 | 0.048135766 | -0.080652617 | 0.037 | 0.069 | 1 |
| BCAT1 | 0.049346472 | 0.107275148 | 0.123 | 0.107 | 1 |
| BCL2L1 | 0.050787052 | -0.165227735 | 0.537 | 0.61 | 1 |
| TNFSF14 | 0.053813067 | -0.139116123 | 0.167 | 0.191 | 1 |
| ERBIN | 0.055042853 | 0.134038934 | 0.667 | 0.578 | 1 |
| HLA.DMA | 0.059237942 | 0.063878264 | 0.117 | 0.069 | 1 |
| NFATC3 | 0.060947314 | 0.111549613 | 0.296 | 0.243 | 1 |
| PTPN7 | 0.061947004 | 0.149853824 | 0.673 | 0.613 | 1 |
| PDIA4 | 0.066511445 | -0.146604089 | 0.401 | 0.491 | 1 |
| CXCL8 | 0.06735332 | 0.206541345 | 0.895 | 0.854 | 1 |
| IL1B | 0.06919655 | 0.177310917 | 0.432 | 0.406 | 1 |
| CD72 | 0.072959303 | -0.065434174 | 0.031 | 0.069 | 1 |
| FOXP1 | 0.074832795 | -0.125420866 | 0.352 | 0.45 | 1 |
| DOCK8 | 0.074933313 | 0.106527637 | 0.315 | 0.229 | 1 |
| LILRB4 | 0.077751337 | 0.039963006 | 0.049 | 0.017 | 1 |
| TRBC2 | 0.078971615 | -0.144666527 | 0.889 | 0.929 | 1 |
| IL17F | 0.082898001 | -0.182661752 | 0.037 | 0.037 | 1 |
| TNIK | 0.083387517 | -0.092169729 | 0.136 | 0.192 | 1 |
| IL15RA | 0.084106862 | -0.137642512 | 0.34 | 0.422 | 1 |
| PSME3 | 0.086821636 | -0.106772246 | 0.284 | 0.344 | 1 |
| EIF5A | 0.087464041 | -0.133762486 | 0.951 | 0.945 | 1 |
| IL7R | 0.0874697 | -0.11450313 | 0.136 | 0.182 | 1 |

| | | | | | |
|---|---|---|---|---|---|
| LAMP1 | 0.08871051 | -0.117878759 | 0.444 | 0.509 | 1 |
| IRF3 | 0.090679543 | 0.040314979 | 0.049 | 0.024 | 1 |
| PCNA | 0.09092215 | -0.098013454 | 0.191 | 0.265 | 1 |
| CD5 | 0.093560697 | 0.051486739 | 0.074 | 0.044 | 1 |
| CXCL2 | 0.095759481 | 0.13687255 | 0.333 | 0.342 | 1 |
| SOCS3 | 0.096085758 | 0.086034176 | 0.228 | 0.154 | 1 |
| IFITM3 | 0.099275127 | 0.07392442 | 0.13 | 0.076 | 1 |
| EIF1 | 0.104345843 | -0.093416882 | 1 | 0.998 | 1 |
| BATF | 0.105215209 | 0.137864065 | 0.599 | 0.568 | 1 |
| JCHAIN | 0.106612388 | 0.087313161 | 0.037 | 0.047 | 1 |
| TBX21 | 0.10826777 | 0.152702939 | 0.617 | 0.556 | 1 |
| CCND2 | 0.11055532 | 0.170850945 | 0.58 | 0.54 | 1 |
| TNFRSF25 | 0.111033114 | -0.102610111 | 0.191 | 0.276 | 1 |
| ZNF683 | 0.111582526 | 0.023614736 | 0.025 | 0.007 | 1 |
| VNN2 | 0.111582526 | 0.023614736 | 0.025 | 0.007 | 1 |
| IL3RA | 0.116292599 | -0.059301561 | 0.037 | 0.059 | 1 |
| CXCL9 | 0.116503785 | -0.057764326 | 0.037 | 0.06 | 1 |
| LGALS1 | 0.120412317 | 0.194144643 | 0.562 | 0.524 | 1 |
| KLRG1 | 0.120976978 | 0.045813286 | 0.056 | 0.029 | 1 |
| CCL2 | 0.1228361 | -0.162298948 | 0.568 | 0.56 | 1 |
| FCER1G | 0.12406619 | -0.146927072 | 0.352 | 0.443 | 1 |
| OSM | 0.124646766 | 0.080857908 | 0.043 | 0.039 | 1 |
| IKZF1 | 0.125207555 | -0.060265965 | 0.062 | 0.09 | 1 |
| VEGFA | 0.127404149 | -0.053347246 | 0.025 | 0.043 | 1 |
| MALT1 | 0.128110384 | 0.068913127 | 0.154 | 0.119 | 1 |
| CCL20 | 0.128982408 | 0.305970026 | 0.432 | 0.461 | 1 |
| RPN2 | 0.131469571 | -0.111958761 | 0.58 | 0.651 | 1 |
| RORC | 0.132433641 | -0.052868222 | 0.006 | 0.017 | 1 |
| TGFB1 | 0.139126916 | -0.049202312 | 0.037 | 0.072 | 1 |
| HIF1A | 0.145598525 | 0.112185656 | 0.679 | 0.677 | 1 |
| STAT6 | 0.146343397 | 0.085146614 | 0.309 | 0.266 | 1 |
| CBLB | 0.147824586 | -0.043120181 | 0.031 | 0.056 | 1 |
| TRIB2 | 0.15109127 | 0.1407234 | 0.296 | 0.261 | 1 |
| CTSW | 0.152370088 | 0.11519617 | 0.506 | 0.453 | 1 |
| HLA.DQB1 | 0.152914601 | 0.11306897 | 0.296 | 0.275 | 1 |
| CCR1 | 0.153798288 | -0.059720523 | 0.086 | 0.115 | 1 |
| DUSP1 | 0.167236091 | -0.101339612 | 0.265 | 0.295 | 1 |
| IFNGR1 | 0.168552847 | -0.090580673 | 0.29 | 0.364 | 1 |
| LTA | 0.168660384 | -0.210709543 | 0.475 | 0.425 | 1 |
| CXCL5 | 0.168670438 | 0.102673166 | 0.228 | 0.237 | 1 |
| TSPAN32 | 0.168792934 | 0.039757372 | 0.062 | 0.041 | 1 |
| LIF | 0.169633694 | 0.467209448 | 0.253 | 0.335 | 1 |
| STIM1 | 0.1717031 | 0.059361749 | 0.191 | 0.148 | 1 |
| GATA3 | 0.174231716 | -0.043120181 | 0.031 | 0.051 | 1 |
| KLRF1 | 0.175515941 | 0.060579136 | 0.117 | 0.096 | 1 |
| TIMM9 | 0.176509392 | -0.091480951 | 0.414 | 0.501 | 1 |
| IL22 | 0.177274468 | -0.091620607 | 0.074 | 0.072 | 1 |

| | | | | | |
|---|---|---|---|---|---|
| NRP1 | 0.184381315 | -0.033682662 | 0.019 | 0.038 | 1 |
| THEMIS | 0.186062796 | 0.021500253 | 0.006 | 0.003 | 1 |
| IL12RB1 | 0.191077935 | -0.036069255 | 0.025 | 0.045 | 1 |
| NFATC2 | 0.19217449 | -0.102628781 | 0.383 | 0.418 | 1 |
| CD36 | 0.193623275 | -0.028049806 | 0.012 | 0.031 | 1 |
| CXCL16 | 0.196634713 | -0.042301208 | 0.037 | 0.056 | 1 |
| CD244 | 0.198501621 | -0.061472102 | 0.056 | 0.084 | 1 |
| TCF7 | 0.199643936 | -0.073985489 | 0.167 | 0.224 | 1 |
| DPP4 | 0.200360033 | 0.021968763 | 0.019 | 0.008 | 1 |
| QPCT | 0.201034395 | -0.032089404 | 0.019 | 0.037 | 1 |
| NDFIP2 | 0.202230526 | -0.045891254 | 0.068 | 0.096 | 1 |
| CD79A | 0.204508354 | -0.023949091 | 0.006 | 0.022 | 1 |
| NANS | 0.2110916 | -0.091065369 | 0.364 | 0.402 | 1 |
| TGFBR2 | 0.21710566 | 0.092769408 | 0.574 | 0.529 | 1 |
| EIF5 | 0.223049094 | -0.085479503 | 0.784 | 0.81 | 1 |
| CD7 | 0.236736089 | -0.07279087 | 0.198 | 0.22 | 1 |
| IL2RB | 0.245118399 | 0.085417412 | 0.969 | 0.967 | 1 |
| LIPA | 0.246132986 | 0.042413755 | 0.117 | 0.083 | 1 |
| CD226 | 0.248249044 | 0.077084408 | 0.333 | 0.262 | 1 |
| IL6R | 0.25027088 | -0.033711267 | 0.031 | 0.047 | 1 |
| TMEM97 | 0.254154679 | -0.032943154 | 0.037 | 0.058 | 1 |
| IL23R | 0.256142264 | -0.032040503 | 0.006 | 0.013 | 1 |
| CD28 | 0.256239483 | -0.020699776 | 0.006 | 0.018 | 1 |
| GEM | 0.271478665 | -0.182339649 | 0.481 | 0.545 | 1 |
| GIMAP5 | 0.284656256 | -0.068173273 | 0.198 | 0.215 | 1 |
| CR2 | 0.291333536 | 0.017042082 | 0.019 | 0.013 | 1 |
| LAT2 | 0.292648659 | 0.083677798 | 0.568 | 0.476 | 1 |
| CHD7 | 0.292950379 | 0.023789125 | 0.049 | 0.031 | 1 |
| GZMK | 0.294874469 | -0.03132094 | 0.019 | 0.036 | 1 |
| APOBEC3G | 0.297775533 | -0.072411731 | 0.981 | 0.988 | 1 |
| FAS | 0.299120525 | 0.067425377 | 0.235 | 0.205 | 1 |
| TLR2 | 0.308858238 | 0.025831583 | 0.049 | 0.035 | 1 |
| CARD11 | 0.319604295 | -0.019985531 | 0.012 | 0.024 | 1 |
| IFI6 | 0.319892382 | 0.027009428 | 0.037 | 0.025 | 1 |
| TNFSF10 | 0.321145659 | 0.049078864 | 0.099 | 0.07 | 1 |
| GZMA | 0.327739228 | -0.195301352 | 0.352 | 0.344 | 1 |
| YBX3 | 0.328146051 | 0.023789125 | 0.037 | 0.031 | 1 |
| FURIN | 0.33129026 | -0.080383778 | 0.852 | 0.918 | 1 |
| FCN1 | 0.337087398 | 0.017533011 | 0.031 | 0.017 | 1 |
| GIMAP2 | 0.337713761 | 0.033734029 | 0.099 | 0.071 | 1 |
| ENTPD1 | 0.339624219 | -0.023216653 | 0.012 | 0.02 | 1 |
| LGALS9 | 0.340320441 | -0.025823358 | 0.031 | 0.045 | 1 |
| DUSP4 | 0.340848575 | 0.117808033 | 0.753 | 0.635 | 1 |
| ARID5A | 0.342539182 | 0.067511882 | 0.457 | 0.381 | 1 |
| MCM2 | 0.346235496 | -0.023372187 | 0.025 | 0.038 | 1 |
| SMAD2 | 0.34687351 | 0.042599168 | 0.179 | 0.166 | 1 |
| SPAG9 | 0.348989993 | -0.055495937 | 0.302 | 0.367 | 1 |

| | | | | | |
|---|---|---|---|---|---|
| JAK2 | 0.34963625 | 0.015403586 | 0.025 | 0.013 | 1 |
| CD48 | 0.359065222 | -0.079137977 | 0.778 | 0.795 | 1 |
| SOCS1 | 0.361520956 | 0.009424899 | 0.012 | 0.006 | 1 |
| SLC7A7 | 0.361520956 | 0.009424899 | 0.012 | 0.006 | 1 |
| SDCBP | 0.362867323 | -0.080273382 | 0.802 | 0.788 | 1 |
| CD9 | 0.363504471 | -0.058117279 | 0.111 | 0.1 | 1 |
| AQP9 | 0.367425471 | -0.014179109 | 0.006 | 0.015 | 1 |
| BIRC3 | 0.370584227 | -0.111086483 | 0.864 | 0.857 | 1 |
| NT5E | 0.37347829 | -0.050410278 | 0.08 | 0.11 | 1 |
| B4GALT5 | 0.373861185 | 0.043428666 | 0.191 | 0.136 | 1 |
| STAT3 | 0.384974203 | 0.062910002 | 0.858 | 0.833 | 1 |
| S100A9 | 0.386652368 | 0.024668378 | 0.062 | 0.038 | 1 |
| F5 | 0.402155317 | -0.029149841 | 0.056 | 0.064 | 1 |
| MCM4 | 0.403500005 | 0.021035916 | 0.056 | 0.038 | 1 |
| ZAP70 | 0.404948018 | 0.027925599 | 0.062 | 0.061 | 1 |
| CD14 | 0.411177715 | -0.01512524 | 0.012 | 0.022 | 1 |
| FCER2 | 0.419187318 | -0.018581828 | 0.025 | 0.037 | 1 |
| ITGAE | 0.420225405 | -0.063262218 | 0.216 | 0.23 | 1 |
| FOS | 0.424448905 | -0.103385523 | 0.377 | 0.357 | 1 |
| PLCG2 | 0.427046961 | -0.022815546 | 0.043 | 0.056 | 1 |
| KLRK1 | 0.429095994 | 0.069962309 | 0.846 | 0.819 | 1 |
| CREM | 0.430168314 | -0.071921638 | 0.685 | 0.704 | 1 |
| SLC1A5 | 0.437106087 | -0.067712569 | 0.883 | 0.892 | 1 |
| TYMS | 0.43735159 | -0.025098137 | 0.031 | 0.043 | 1 |
| MAST4 | 0.445246404 | 0.038531699 | 0.099 | 0.075 | 1 |
| RBCK1 | 0.445253805 | 0.044341764 | 0.247 | 0.245 | 1 |
| CD38 | 0.448527648 | 0.015903768 | 0.031 | 0.016 | 1 |
| IL11RA | 0.448851059 | 0.014276362 | 0.025 | 0.021 | 1 |
| CYBB | 0.449778366 | -0.021248251 | 0.043 | 0.056 | 1 |
| IL6 | 0.455605376 | -0.053332987 | 0.278 | 0.272 | 1 |
| EVI2B | 0.461145243 | 0.062857237 | 0.568 | 0.563 | 1 |
| CD40 | 0.461527607 | -0.045042065 | 0.167 | 0.236 | 1 |
| PDIA6 | 0.463224549 | -0.057404201 | 0.802 | 0.85 | 1 |
| TNFRSF13C | 0.464295804 | 0.039341498 | 0.123 | 0.11 | 1 |
| CCL3 | 0.470704499 | -0.097391561 | 1 | 1 | 1 |
| JUNB | 0.474449796 | -0.053304904 | 0.481 | 0.528 | 1 |
| IGHD | 0.476639682 | 0.007775162 | 0.012 | 0.007 | 1 |
| FBXO11 | 0.476763426 | 0.042187824 | 0.327 | 0.29 | 1 |
| EVI2A | 0.485836033 | 0.054430784 | 0.568 | 0.467 | 1 |
| LRRC8B | 0.489619897 | -0.045944798 | 0.315 | 0.306 | 1 |
| PASK | 0.492411916 | -0.015379396 | 0.025 | 0.035 | 1 |
| GZMB | 0.492675384 | -0.077206718 | 1 | 0.999 | 1 |
| IL13 | 0.499353111 | 0.047699103 | 0.099 | 0.102 | 1 |
| NCAM1 | 0.509759298 | 0.032572411 | 0.056 | 0.062 | 1 |
| KLRB1 | 0.51140521 | -0.009269195 | 0.006 | 0.012 | 1 |
| KDELR1 | 0.517791406 | 0.032392625 | 0.228 | 0.2 | 1 |
| IL1R1 | 0.519483501 | 0.058525678 | 0.167 | 0.128 | 1 |

| | | | | | |
|---|---|---|---|---|---|
| PSMB8 | 0.52209396 | 0.043590194 | 0.654 | 0.633 | 1 |
| NR4A2 | 0.526553734 | -0.074971947 | 0.414 | 0.391 | 1 |
| EGR1 | 0.535527822 | 0.052057475 | 0.358 | 0.273 | 1 |
| IL31 | 0.5491599 | -0.011875926 | 0.012 | 0.016 | 1 |
| JUN | 0.553025544 | 0.025769999 | 0.093 | 0.085 | 1 |
| ADA | 0.554517991 | -0.024096613 | 0.08 | 0.085 | 1 |
| NGDN | 0.55548561 | -0.014707326 | 0.031 | 0.038 | 1 |
| BCL2A1 | 0.557158294 | 0.053704768 | 0.981 | 0.955 | 1 |
| SP140 | 0.559308877 | -0.02327198 | 0.074 | 0.079 | 1 |
| CNOT2 | 0.561046769 | -0.036641866 | 0.327 | 0.333 | 1 |
| NFKBID | 0.563015206 | -0.01377551 | 0.019 | 0.031 | 1 |
| NFKBIZ | 0.564909173 | 0.051965593 | 0.531 | 0.526 | 1 |
| SLA | 0.564946886 | -0.051176147 | 0.765 | 0.808 | 1 |
| LEF1 | 0.566022728 | -0.020060321 | 0.056 | 0.075 | 1 |
| BCL11B | 0.566197292 | 0.014171123 | 0.049 | 0.037 | 1 |
| PIK3AP1 | 0.575959233 | -0.049442379 | 0.667 | 0.648 | 1 |
| LIG4 | 0.583237964 | -0.023181571 | 0.13 | 0.142 | 1 |
| MZB1 | 0.584359944 | 0.011607449 | 0.006 | 0.008 | 1 |
| ADGRG3 | 0.586354754 | 0.020815809 | 0.037 | 0.047 | 1 |
| ATF4 | 0.589460566 | 0.037391391 | 0.988 | 0.984 | 1 |
| SPP1 | 0.590304498 | 0.008324865 | 0.012 | 0.013 | 1 |
| TNFSF13B | 0.594361719 | 0.006127309 | 0.012 | 0.008 | 1 |
| CD163 | 0.594361719 | 0.006127309 | 0.012 | 0.008 | 1 |
| GRB10 | 0.599614872 | -0.007628837 | 0.006 | 0.009 | 1 |
| CMKLR1 | 0.599614872 | -0.007628837 | 0.006 | 0.009 | 1 |
| CXCL11 | 0.59992637 | -0.007628837 | 0.006 | 0.009 | 1 |
| IL18 | 0.59992637 | -0.007628837 | 0.006 | 0.009 | 1 |
| CSF1 | 0.60255085 | 0.102822143 | 0.284 | 0.315 | 1 |
| TRAC | 0.604964898 | 0.044484528 | 0.438 | 0.455 | 1 |
| LGALS3 | 0.605006642 | 0.020628325 | 0.123 | 0.108 | 1 |
| MYB | 0.607090867 | -0.008619276 | 0.012 | 0.017 | 1 |
| CXCL1 | 0.60861617 | 0.023259196 | 0.068 | 0.045 | 1 |
| IRF7 | 0.610983692 | -0.04162878 | 0.364 | 0.374 | 1 |
| CD24 | 0.614781292 | 0.003906193 | 0.006 | 0.003 | 1 |
| LY86 | 0.614781292 | 0.003906193 | 0.006 | 0.003 | 1 |
| IL5 | 0.623486858 | -0.005986611 | 0.006 | 0.01 | 1 |
| ZBTB16 | 0.638068922 | -0.011515521 | 0.031 | 0.036 | 1 |
| IL9 | 0.643463823 | 0.011548766 | 0.031 | 0.024 | 1 |
| CCL22 | 0.660743302 | -0.020802498 | 0.154 | 0.166 | 1 |
| CXCR5 | 0.662907769 | 0.026383564 | 0.167 | 0.196 | 1 |
| FOXO1 | 0.666726713 | -0.012736751 | 0.049 | 0.053 | 1 |
| THBS1 | 0.682085913 | 0.043028334 | 0.16 | 0.116 | 1 |
| UGCG | 0.682662463 | -0.017210227 | 0.117 | 0.121 | 1 |
| SELL | 0.683534913 | 0.033639717 | 0.173 | 0.16 | 1 |
| NFATC1 | 0.685396447 | -0.022893866 | 0.191 | 0.196 | 1 |
| PSME2 | 0.686256065 | 0.031044941 | 0.735 | 0.664 | 1 |
| IL3 | 0.692671863 | -0.22815133 | 0.148 | 0.133 | 1 |

| | | | | | |
|---|---|---|---|---|---|
| CD86 | 0.69602672 | 0.011999185 | 0.062 | 0.045 | 1 |
| TGFBR1 | 0.696256519 | -0.018009283 | 0.148 | 0.169 | 1 |
| CXCR3 | 0.707203188 | -0.034080702 | 0.259 | 0.366 | 1 |
| ZNF341 | 0.710502922 | 0.004481336 | 0.012 | 0.009 | 1 |
| ICOS | 0.716266195 | -0.027950522 | 0.383 | 0.331 | 1 |
| NAMPT | 0.716603796 | 0.037234877 | 0.901 | 0.896 | 1 |
| EEF1E1 | 0.723569951 | -0.019088492 | 0.191 | 0.22 | 1 |
| TNFRSF9 | 0.728451222 | -0.013793541 | 0.086 | 0.096 | 1 |
| CD80 | 0.729054953 | -0.004342514 | 0.006 | 0.008 | 1 |
| PDCD1 | 0.736824262 | -0.005355259 | 0.012 | 0.015 | 1 |
| SRGN | 0.740562287 | -0.024132341 | 0.877 | 0.9 | 1 |
| CTLA4 | 0.742239773 | -0.039811656 | 0.259 | 0.226 | 1 |
| CXCL3 | 0.742393153 | 0.021458892 | 0.136 | 0.153 | 1 |
| MGST1 | 0.75438744 | -0.004342514 | 0.006 | 0.007 | 1 |
| FOXP3 | 0.75438744 | -0.004342514 | 0.006 | 0.007 | 1 |
| KLRC4 | 0.761287027 | 0.006166756 | 0.031 | 0.025 | 1 |
| TNFSF8 | 0.76306016 | -0.008648214 | 0.043 | 0.043 | 1 |
| IL23A | 0.767762944 | -0.027225119 | 0.29 | 0.265 | 1 |
| NFKB2 | 0.768155803 | -0.018755512 | 0.284 | 0.281 | 1 |
| SKAP1 | 0.772082613 | 0.005611501 | 0.025 | 0.018 | 1 |
| AHI1 | 0.774021184 | -0.008971959 | 0.043 | 0.056 | 1 |
| IGBP1 | 0.775861085 | 0.011704108 | 0.142 | 0.136 | 1 |
| CD69 | 0.775995775 | -0.026645072 | 0.988 | 0.999 | 1 |
| TNFRSF4 | 0.781113329 | -0.01813143 | 0.222 | 0.197 | 1 |
| TGFBI | 0.782552654 | -0.010211087 | 0.074 | 0.067 | 1 |
| TRAT1 | 0.78383265 | 0.005611501 | 0.025 | 0.017 | 1 |
| BIN2 | 0.788348303 | 0.008324865 | 0.074 | 0.063 | 1 |
| CCR5 | 0.79189532 | 0.008849195 | 0.049 | 0.045 | 1 |
| DCLRE1C | 0.792484134 | -0.008353876 | 0.062 | 0.061 | 1 |
| TNIP3 | 0.79296344 | 0.002252675 | 0.006 | 0.005 | 1 |
| POU2AF1 | 0.795381154 | -0.0114072 | 0.074 | 0.077 | 1 |
| LAP3 | 0.809588456 | -0.013048786 | 0.198 | 0.207 | 1 |
| CD274 | 0.815531989 | 0.01858277 | 0.29 | 0.314 | 1 |
| MS4A1 | 0.818592795 | -0.005110649 | 0.031 | 0.033 | 1 |
| ARHGEF3 | 0.818979758 | 0.016920146 | 0.346 | 0.304 | 1 |
| ITK | 0.822288069 | 0.019470248 | 0.895 | 0.904 | 1 |
| IGHM | 0.822408447 | -0.003720477 | 0.012 | 0.013 | 1 |
| EOMES | 0.82289805 | -0.002696541 | 0.006 | 0.007 | 1 |
| CD109 | 0.82728096 | -0.014271748 | 0.148 | 0.132 | 1 |
| CCR4 | 0.834340107 | 0.010318915 | 0.086 | 0.078 | 1 |
| PAXBP1 | 0.835980112 | -0.013646549 | 0.278 | 0.283 | 1 |
| ZBED2 | 0.838526369 | -0.005110649 | 0.031 | 0.029 | 1 |
| NINJ2 | 0.838612866 | 0.002837239 | 0.006 | 0.01 | 1 |
| PIK3IP1 | 0.840285866 | -0.014323874 | 0.315 | 0.316 | 1 |
| IER3 | 0.855785746 | -0.020344151 | 0.889 | 0.894 | 1 |
| EGR3 | 0.859720798 | -0.007470294 | 0.08 | 0.069 | 1 |
| LAMP3 | 0.870077557 | -0.007890721 | 0.13 | 0.138 | 1 |

| | | | | | |
|---|---|---|---|---|---|
| CD6 | 0.884466125 | 0.012329825 | 0.34 | 0.291 | 1 |
| GBP2 | 0.887247629 | 0.010979318 | 0.568 | 0.581 | 1 |
| BTLA | 0.888145727 | 0.002935635 | 0.031 | 0.028 | 1 |
| KLRC3 | 0.890835674 | -0.015131364 | 0.667 | 0.665 | 1 |
| BCL10 | 0.8994511 | -0.004426633 | 0.08 | 0.083 | 1 |
| FN1 | 0.89988203 | 0.005876714 | 0.08 | 0.112 | 1 |
| IL1A | 0.90335521 | 0.013004386 | 0.074 | 0.066 | 1 |
| CD22 | 0.913907812 | -0.003297055 | 0.037 | 0.045 | 1 |
| GAB2 | 0.914282814 | 0.004278032 | 0.086 | 0.079 | 1 |
| LRRC32 | 0.915812066 | 0.001782019 | 0.019 | 0.016 | 1 |
| CD4 | 0.917406354 | -0.001048689 | 0.006 | 0.007 | 1 |
| CD79B | 0.93347658 | 0.001195013 | 0.012 | 0.01 | 1 |
| IL12A | 0.93347658 | 0.001195013 | 0.012 | 0.01 | 1 |
| PMCH | 0.938711245 | -0.001048689 | 0.006 | 0.005 | 1 |
| FYB1 | 0.941796939 | -0.002516695 | 0.056 | 0.069 | 1 |
| LYN | 0.943060134 | 0.003889893 | 0.191 | 0.208 | 1 |
| IRF4 | 0.947406977 | -0.006599012 | 0.877 | 0.918 | 1 |
| TOP2A | 0.94882582 | 0.000601049 | 0.006 | 0.006 | 1 |
| IL6ST | 0.949741668 | -0.003736753 | 0.191 | 0.181 | 1 |
| CD37 | 0.960791547 | -0.003179646 | 0.327 | 0.367 | 1 |
| MMP9 | 0.965534151 | -0.001771013 | 0.068 | 0.085 | 1 |
| CD247 | 0.968218663 | 0.00248162 | 0.333 | 0.315 | 1 |
| VMO1 | 0.976165099 | 0.00087456 | 0.031 | 0.036 | 1 |
| PGM3 | 0.979300753 | -0.00128673 | 0.117 | 0.123 | 1 |
| IL2RA | 0.981909164 | 0.002021123 | 0.71 | 0.707 | 1 |
| LAT | 0.982366355 | -0.002161326 | 0.494 | 0.432 | 1 |
| PAK1IP1 | 0.986606399 | -0.001002024 | 0.253 | 0.236 | 1 |
| FUT4 | 0.988336574 | 0.000436955 | 0.056 | 0.048 | 1 |
| RAPH1 | 0.988891528 | -0.052170929 | 0 | 0.03 | 1 |
| FTH1 | 0.992196605 | 0.000840362 | 0.864 | 0.904 | 1 |
| TNFRSF8 | 0.992963759 | -0.040918654 | 0 | 0.021 | 1 |
| CHD1 | 0.99301442 | -0.036069255 | 0 | 0.024 | 1 |
| TIAF1 | 0.993052192 | -0.032827242 | 0 | 0.022 | 1 |
| S100A12 | 0.993289119 | -0.018147347 | 0 | 0.013 | 1 |
| EBF1 | 0.993769402 | 0.000150933 | 0.019 | 0.015 | 1 |
| CXCR6 | 0.995712148 | -0.016506988 | 0 | 0.007 | 1 |
| BCL6 | 0.995737474 | -0.014864763 | 0 | 0.01 | 1 |
| SNCA | 0.995797884 | -0.011574693 | 0 | 0.008 | 1 |
| IL15 | 0.995834938 | -0.00992684 | 0 | 0.007 | 1 |
| TCL1A | 0.995878764 | -0.008277103 | 0 | 0.006 | 1 |
| CD27 | 0.995932403 | -0.006625477 | 0 | 0.005 | 1 |
| BLNK | 0.995932403 | -0.006625477 | 0 | 0.005 | 1 |
| IGHG2 | 0.995932403 | -0.006625477 | 0 | 0.005 | 1 |
| IL26 | 0.995932403 | -0.006625477 | 0 | 0.005 | 1 |
| CCL13 | 0.995932403 | -0.006625477 | 0 | 0.003 | 1 |
| VSIG4 | 0.995932403 | -0.006625477 | 0 | 0.003 | 1 |
| CD209 | 0.996001556 | -0.004971958 | 0 | 0.003 | 1 |

| | | | | | |
|---|---|---|---|---|---|
| CLEC4E | 0.996001556 | -0.004971958 | 0 | 0.003 | 1 |
| IGHG3 | 0.996001556 | -0.004971958 | 0 | 0.003 | 1 |
| IKZF2 | 0.996001556 | -0.004971958 | 0 | 0.003 | 1 |
| PTGDR2 | 0.996001556 | -0.004971958 | 0 | 0.003 | 1 |
| CTSD | 0.999672759 | 1.80E-05 | 0.123 | 0.115 | 1 |

Undefined

| | p_val | avg_log2FC | Patient | Controls | p_val_adj |
|---|---|---|---|---|---|
| IGLC3 | 3.27E-31 | 3.19246586 | 0.561 | 0.517 | 1.63E-28 |
| TNF | 1.88E-21 | -1.439728779 | 0.994 | 0.996 | 9.37E-19 |
| CCL22 | 2.34E-19 | -2.428197236 | 0.271 | 0.45 | 1.16E-16 |
| IGKC | 2.65E-18 | -2.875753419 | 0.497 | 0.503 | 1.32E-15 |
| LTA | 1.41E-15 | -1.325751888 | 0.845 | 0.929 | 7.01E-13 |
| CSF2 | 1.85E-13 | -1.446684696 | 0.955 | 0.971 | 9.23E-11 |
| TNFRSF4 | 1.96E-10 | 0.799497402 | 0.535 | 0.38 | 9.77E-08 |
| FYN | 4.33E-10 | -0.513794532 | 0.845 | 0.932 | 2.15E-07 |
| GZMH | 1.90E-09 | 0.991193026 | 0.432 | 0.258 | 9.46E-07 |
| S100A10 | 2.44E-08 | 0.746492053 | 0.845 | 0.804 | 1.21E-05 |
| DUSP4 | 3.25E-08 | -0.686227412 | 0.845 | 0.848 | 1.62E-05 |
| ITGA4 | 1.51E-07 | 0.404922295 | 0.484 | 0.336 | 7.53E-05 |
| CD40LG | 2.46E-07 | -0.621805947 | 0.135 | 0.26 | 0.000122312 |
| TRAC | 2.55E-07 | 0.647368873 | 0.884 | 0.83 | 0.000126813 |
| IL3 | 6.86E-07 | -1.472975445 | 0.103 | 0.15 | 0.000341471 |
| CD5 | 1.69E-06 | 0.450940696 | 0.329 | 0.17 | 0.000840735 |
| GBP2 | 2.55E-06 | 0.429215676 | 0.955 | 0.929 | 0.001267888 |
| CCL20 | 4.05E-06 | -1.402911311 | 0.542 | 0.554 | 0.002016183 |
| FCGR3A | 5.96E-06 | 0.358325118 | 0.174 | 0.073 | 0.002967723 |
| REL | 6.80E-06 | -0.313508563 | 0.987 | 0.993 | 0.003387863 |
| CR2 | 8.09E-06 | 0.42976382 | 0.355 | 0.205 | 0.004027456 |
| SLA | 1.01E-05 | -0.43817767 | 0.774 | 0.852 | 0.005049008 |
| IL2RG | 1.72E-05 | 0.296786495 | 0.974 | 0.985 | 0.008572561 |
| GAPDH | 2.16E-05 | -0.375776609 | 0.994 | 0.991 | 0.010781063 |
| EVI2A | 2.39E-05 | -0.415582158 | 0.497 | 0.603 | 0.011906527 |
| IFNG | 2.89E-05 | -0.881893222 | 1 | 0.998 | 0.014410806 |
| IL1RN | 3.00E-05 | -0.424150246 | 0.174 | 0.307 | 0.014937815 |
| FASLG | 3.69E-05 | -0.528045293 | 0.426 | 0.578 | 0.018365341 |
| IL22 | 4.52E-05 | -1.416018675 | 0.084 | 0.079 | 0.022505221 |
| ARHGEF3 | 4.65E-05 | -0.363414591 | 0.239 | 0.371 | 0.023146945 |
| IL13 | 4.75E-05 | -0.859058817 | 0.077 | 0.126 | 0.023660464 |
| IL23A | 5.15E-05 | -0.630211214 | 0.535 | 0.565 | 0.025656745 |
| IGHD | 6.43E-05 | 0.320684384 | 0.355 | 0.185 | 0.032009176 |
| IL4 | 7.50E-05 | -0.78988055 | 0.077 | 0.102 | 0.037352245 |
| CCR7 | 0.000105778 | -0.358623736 | 0.948 | 0.996 | 0.05267727 |
| JAK1 | 0.000109072 | 0.299315321 | 0.955 | 0.951 | 0.054317727 |
| IRF8 | 0.000127841 | -0.402319402 | 0.987 | 0.996 | 0.063664909 |
| IL1A | 0.000140428 | -0.640555805 | 0.148 | 0.234 | 0.069933077 |

| | | | | | |
|---|---|---|---|---|---|
| LYN | 0.000160329 | -0.388581539 | 0.6 | 0.698 | 0.079843848 |
| CXCR3 | 0.000201901 | 0.459163677 | 0.535 | 0.415 | 0.100546876 |
| IL32 | 0.000262627 | 0.333728565 | 0.51 | 0.406 | 0.130788307 |
| TRIB2 | 0.000300374 | 0.339585186 | 0.271 | 0.128 | 0.149586123 |
| STAT5A | 0.000334282 | -0.247924163 | 0.994 | 0.991 | 0.166472209 |
| CCL4 | 0.000356155 | -0.508902072 | 1 | 1 | 0.17736501 |
| CD22 | 0.000356195 | 0.496348843 | 0.555 | 0.534 | 0.177385261 |
| CD83 | 0.000489907 | -0.298135124 | 0.994 | 0.996 | 0.243973808 |
| BACH2 | 0.00055288 | -0.265825738 | 0.31 | 0.439 | 0.275334296 |
| CD48 | 0.000666614 | -0.288382147 | 0.987 | 0.991 | 0.331973991 |
| CD226 | 0.000796702 | 0.282415147 | 0.413 | 0.272 | 0.396757354 |
| TUBA1C | 0.000951059 | -0.237292874 | 0.994 | 0.998 | 0.473627409 |
| CD70 | 0.001805873 | -0.385103342 | 0.11 | 0.21 | 0.899324555 |
| CXCL11 | 0.001857924 | 0.340313531 | 0.032 | 0.02 | 0.925246371 |
| IL2RB | 0.001889688 | 0.352631365 | 0.865 | 0.742 | 0.941064672 |
| BTG1 | 0.001982464 | -0.227090753 | 1 | 1 | 0.987266955 |
| HLA.A | 0.00216407 | 0.179312745 | 1 | 1 | 1 |
| STAT3 | 0.002223701 | 0.20656078 | 1 | 0.954 | 1 |
| EVI2B | 0.002332547 | -0.310412973 | 0.497 | 0.6 | 1 |
| CD40 | 0.002431935 | -0.331480231 | 0.942 | 0.958 | 1 |
| CD27 | 0.002501086 | -0.146155898 | 0.013 | 0.099 | 1 |
| TIGIT | 0.002528931 | 0.28175695 | 0.303 | 0.177 | 1 |
| HAVCR2 | 0.002536035 | -0.34994636 | 0.148 | 0.192 | 1 |
| TRAF1 | 0.002565496 | -0.21682799 | 0.929 | 0.969 | 1 |
| LEF1 | 0.0028037 | 0.377317833 | 0.31 | 0.256 | 1 |
| EGR3 | 0.003564465 | -0.231289252 | 0.194 | 0.302 | 1 |
| RGS1 | 0.003622074 | 0.475274111 | 0.865 | 0.766 | 1 |
| IL1B | 0.004026617 | 0.349494151 | 0.516 | 0.453 | 1 |
| TGFBR2 | 0.004213687 | 0.26328259 | 0.748 | 0.658 | 1 |
| PTPN7 | 0.005046816 | 0.243576946 | 0.535 | 0.426 | 1 |
| CD80 | 0.005049229 | -0.23425866 | 0.135 | 0.221 | 1 |
| PRNP | 0.005135793 | -0.237084544 | 0.916 | 0.962 | 1 |
| NFKBIZ | 0.005568573 | -0.273930284 | 0.961 | 0.945 | 1 |
| FTH1 | 0.005760611 | 0.26024853 | 0.994 | 0.989 | 1 |
| NFKBIA | 0.006092744 | -0.239485397 | 1 | 1 | 1 |
| CYCS | 0.006395305 | -0.179112887 | 0.981 | 0.993 | 1 |
| CD3D | 0.007368606 | 0.305136376 | 0.897 | 0.839 | 1 |
| GNLY | 0.007809798 | 0.668825153 | 0.568 | 0.532 | 1 |
| CD38 | 0.008175833 | 0.068074998 | 0.065 | 0.011 | 1 |
| GBP1 | 0.0088509 | 0.307736964 | 0.858 | 0.786 | 1 |
| IGHG1 | 0.009401042 | 1.59953084 | 0.161 | 0.249 | 1 |
| PASK | 0.009847282 | 0.15632813 | 0.142 | 0.119 | 1 |
| TGIF1 | 0.009862261 | -0.254775608 | 0.716 | 0.819 | 1 |
| ORAI1 | 0.009958907 | -0.209850961 | 0.697 | 0.722 | 1 |
| IL6ST | 0.010095602 | -0.213876621 | 0.355 | 0.424 | 1 |
| CXCL2 | 0.01022401 | 0.24634844 | 0.413 | 0.408 | 1 |
| LIF | 0.010541004 | -0.581282663 | 0.368 | 0.391 | 1 |

| | | | | | |
|---|---|---|---|---|---|
| PFN1 | 0.010746598 | 0.147526605 | 1 | 1 | 1 |
| ANXA5 | 0.010810469 | -0.256433618 | 0.961 | 0.938 | 1 |
| STAT4 | 0.011288481 | -0.226769746 | 0.781 | 0.854 | 1 |
| IL18R1 | 0.011316036 | -0.127566283 | 0.065 | 0.137 | 1 |
| NFKBID | 0.011397398 | -0.146948736 | 0.129 | 0.223 | 1 |
| ARID5A | 0.011838359 | 0.190580908 | 0.787 | 0.722 | 1 |
| ABCE1 | 0.012020521 | -0.159554007 | 0.948 | 0.967 | 1 |
| CTLA4 | 0.012427789 | 0.693023461 | 0.445 | 0.369 | 1 |
| EEF1E1 | 0.012520069 | -0.183870033 | 0.426 | 0.525 | 1 |
| HLA.DPA1 | 0.012670399 | -0.26831277 | 0.916 | 0.927 | 1 |
| LAG3 | 0.012952316 | 0.376809036 | 0.29 | 0.227 | 1 |
| LCK | 0.013469419 | 0.137764861 | 0.206 | 0.115 | 1 |
| KLRG1 | 0.01600193 | 0.11185669 | 0.097 | 0.038 | 1 |
| IRF4 | 0.016129919 | 0.187582374 | 0.994 | 0.993 | 1 |
| ARL4C | 0.016232842 | -0.157786238 | 0.071 | 0.13 | 1 |
| RPL6 | 0.017000264 | -0.117272489 | 1 | 1 | 1 |
| CD8A | 0.017309535 | 0.408748562 | 0.394 | 0.285 | 1 |
| JAK3 | 0.01748777 | 0.180280986 | 0.639 | 0.525 | 1 |
| TNFSF8 | 0.018674961 | 0.277012928 | 0.213 | 0.174 | 1 |
| LIPA | 0.021244759 | -0.129809858 | 0.123 | 0.21 | 1 |
| SLC7A7 | 0.022907701 | 0.053045798 | 0.058 | 0.02 | 1 |
| IGHA1 | 0.022950671 | -0.161496207 | 0.032 | 0.053 | 1 |
| NFKB1 | 0.023141972 | -0.114313726 | 1 | 1 | 1 |
| KLRC1 | 0.024129806 | -0.37518274 | 0.226 | 0.234 | 1 |
| FAS | 0.024626794 | 0.163817243 | 0.413 | 0.305 | 1 |
| CD63 | 0.025184132 | -0.180040206 | 0.884 | 0.896 | 1 |
| OSM | 0.026868055 | -0.082388715 | 0.026 | 0.075 | 1 |
| DUSP2 | 0.027105003 | -0.203128532 | 0.994 | 0.998 | 1 |
| TNFSF14 | 0.028204162 | 0.186014797 | 0.297 | 0.236 | 1 |
| CD52 | 0.028507182 | 0.226939702 | 0.761 | 0.682 | 1 |
| ATF4 | 0.028763617 | -0.111909804 | 1 | 1 | 1 |
| ADA | 0.030055741 | 0.205380813 | 0.206 | 0.183 | 1 |
| YBX3 | 0.032314539 | 0.164773198 | 0.439 | 0.291 | 1 |
| JUNB | 0.036191537 | 0.186917878 | 0.645 | 0.638 | 1 |
| GIMAP2 | 0.037091401 | 0.07661301 | 0.11 | 0.071 | 1 |
| BCL2L1 | 0.037567257 | -0.185438873 | 0.652 | 0.722 | 1 |
| EBF1 | 0.037706071 | 0.179906828 | 0.516 | 0.475 | 1 |
| SNX8 | 0.03998721 | -0.159740916 | 0.768 | 0.826 | 1 |
| MAP2K3 | 0.041147082 | 0.150931475 | 0.968 | 0.974 | 1 |
| ICAM1 | 0.041609399 | -0.206744839 | 0.942 | 0.954 | 1 |
| CXCL10 | 0.042008746 | 0.661083681 | 0.219 | 0.181 | 1 |
| FOS | 0.046085631 | -0.300754072 | 0.665 | 0.642 | 1 |
| CD160 | 0.046853138 | -0.082667072 | 0.013 | 0.051 | 1 |
| IL18RAP | 0.046861842 | -0.135566989 | 0.077 | 0.121 | 1 |
| TSPAN32 | 0.047889431 | 0.06119536 | 0.058 | 0.022 | 1 |
| HNRNPC | 0.050261706 | -0.122261407 | 0.994 | 0.993 | 1 |
| FOSB | 0.050851252 | -0.224737757 | 0.445 | 0.475 | 1 |

| | | | | | |
|---|---|---|---|---|---|
| IL1R1 | 0.051286116 | 0.178009025 | 0.194 | 0.137 | 1 |
| JAK2 | 0.058302527 | 0.060582408 | 0.065 | 0.029 | 1 |
| DUSP6 | 0.06317227 | -0.221937384 | 0.394 | 0.433 | 1 |
| ICOS | 0.063342854 | 0.327741323 | 0.619 | 0.603 | 1 |
| IL15RA | 0.064395657 | 0.158592102 | 0.761 | 0.726 | 1 |
| JUN | 0.065829167 | -0.142562032 | 0.206 | 0.269 | 1 |
| LGALS1 | 0.068661246 | -0.246246364 | 0.594 | 0.658 | 1 |
| CHI3L2 | 0.070088254 | 0.030403055 | 0.019 | 0.004 | 1 |
| MALT1 | 0.071266781 | -0.116119669 | 0.194 | 0.254 | 1 |
| SELL | 0.071706159 | -0.281872606 | 0.368 | 0.446 | 1 |
| SDCBP | 0.072296587 | -0.156875394 | 0.916 | 0.918 | 1 |
| UFM1 | 0.073066354 | 0.114666054 | 0.994 | 0.989 | 1 |
| SLC1A5 | 0.074113424 | -0.120852847 | 0.994 | 0.996 | 1 |
| SPP1 | 0.074946565 | 0.027235776 | 0.026 | 0.007 | 1 |
| FOSL1 | 0.078584268 | -0.130691612 | 0.897 | 0.921 | 1 |
| LY86 | 0.078982613 | -0.083342283 | 0.065 | 0.106 | 1 |
| EIF1 | 0.08023212 | -0.092660126 | 1 | 1 | 1 |
| LDHA | 0.080869969 | -0.096102576 | 1 | 1 | 1 |
| BCAT1 | 0.084358023 | 0.170358158 | 0.394 | 0.313 | 1 |
| BCL2 | 0.085474146 | 0.131068012 | 0.574 | 0.541 | 1 |
| LRRC8B | 0.085742031 | 0.139091721 | 0.503 | 0.525 | 1 |
| NFATC1 | 0.086434082 | -0.136996769 | 0.439 | 0.455 | 1 |
| GAB2 | 0.086628833 | 0.10117844 | 0.258 | 0.205 | 1 |
| NIBAN3 | 0.087058675 | -0.043871893 | 0.006 | 0.038 | 1 |
| RORA | 0.087444844 | 0.180460504 | 0.335 | 0.267 | 1 |
| PSMB9 | 0.089673649 | 0.146613448 | 0.761 | 0.72 | 1 |
| IL2RA | 0.09169449 | 0.140736969 | 0.703 | 0.68 | 1 |
| IRF3 | 0.092094228 | 0.049378904 | 0.045 | 0.029 | 1 |
| SPAG9 | 0.092150326 | -0.12741171 | 0.606 | 0.691 | 1 |
| TCF4 | 0.094966138 | -0.043871893 | 0.006 | 0.033 | 1 |
| CD72 | 0.096181111 | -0.098299732 | 0.077 | 0.126 | 1 |
| EIF5 | 0.099385314 | -0.114056969 | 0.948 | 0.974 | 1 |
| CD28 | 0.102788952 | 0.073641443 | 0.097 | 0.06 | 1 |
| IGHM | 0.106017961 | 0.212258587 | 0.277 | 0.199 | 1 |
| RORC | 0.106421997 | -0.117572974 | 0.026 | 0.064 | 1 |
| GATA3 | 0.107502949 | -0.049843225 | 0.019 | 0.046 | 1 |
| IVNS1ABP | 0.109125944 | 0.133363662 | 0.877 | 0.859 | 1 |
| FN1 | 0.112405214 | 0.096957814 | 0.123 | 0.099 | 1 |
| MYC | 0.116488553 | -0.143633263 | 0.987 | 0.985 | 1 |
| VSIR | 0.12090767 | -0.143131906 | 0.8 | 0.885 | 1 |
| BIRC3 | 0.125846239 | -0.189938393 | 0.968 | 0.943 | 1 |
| IL31 | 0.128260557 | -0.178793199 | 0.026 | 0.018 | 1 |
| EOMES | 0.129355972 | 0.024075435 | 0.026 | 0.009 | 1 |
| GZMA | 0.130114735 | -0.192789062 | 0.077 | 0.132 | 1 |
| IL21 | 0.132610692 | -0.331136028 | 0.116 | 0.093 | 1 |
| BTLA | 0.135683365 | 0.136047402 | 0.297 | 0.223 | 1 |
| VPREB3 | 0.136750969 | -0.037719666 | 0.013 | 0.038 | 1 |

| | | | | | |
|---|---|---|---|---|---|
| RBCK1 | 0.140703082 | 0.09916214 | 0.368 | 0.298 | 1 |
| CCL2 | 0.141084226 | 0.146109857 | 0.665 | 0.62 | 1 |
| IL7R | 0.141856749 | -0.138070191 | 0.31 | 0.364 | 1 |
| IKZF1 | 0.143146825 | 0.08455236 | 0.245 | 0.181 | 1 |
| ITK | 0.14453702 | 0.146726527 | 0.903 | 0.909 | 1 |
| EIF5A | 0.147964651 | -0.082596317 | 1 | 1 | 1 |
| PAXBP1 | 0.148488973 | -0.110008527 | 0.432 | 0.481 | 1 |
| TCL1A | 0.151079522 | 0.122607692 | 0.206 | 0.124 | 1 |
| CARD11 | 0.161664086 | 0.032669662 | 0.032 | 0.013 | 1 |
| ZBED2 | 0.161723938 | -0.19054224 | 0.058 | 0.073 | 1 |
| CD86 | 0.166519936 | -0.205260328 | 0.368 | 0.4 | 1 |
| CST7 | 0.166527344 | 0.209727797 | 0.677 | 0.561 | 1 |
| TGFB1 | 0.172090152 | -0.046764806 | 0.019 | 0.046 | 1 |
| LILRB4 | 0.172135356 | -0.040779466 | 0.013 | 0.029 | 1 |
| IL6 | 0.179197995 | -0.274589272 | 0.774 | 0.799 | 1 |
| PSME2 | 0.180678502 | 0.088616067 | 0.994 | 0.985 | 1 |
| CD8B | 0.181668293 | 0.024075435 | 0.026 | 0.007 | 1 |
| IL4R | 0.181871059 | -0.089407105 | 0.968 | 0.978 | 1 |
| BCL2A1 | 0.182602261 | -0.078458523 | 1 | 1 | 1 |
| FCER2 | 0.183420575 | 0.16670258 | 0.465 | 0.349 | 1 |
| CSF1 | 0.183787252 | 0.189873161 | 0.432 | 0.444 | 1 |
| TBX21 | 0.185686056 | 0.121186487 | 0.955 | 0.962 | 1 |
| CXCL3 | 0.186085277 | -0.129462235 | 0.174 | 0.201 | 1 |
| BIN2 | 0.187935919 | -0.031580529 | 0.013 | 0.033 | 1 |
| PSEN1 | 0.188723787 | 0.051133725 | 0.103 | 0.071 | 1 |
| NKG7 | 0.189473935 | -0.244203358 | 0.626 | 0.675 | 1 |
| NR4A2 | 0.190599172 | -0.174146953 | 0.594 | 0.638 | 1 |
| TMEM97 | 0.194570375 | -0.057502278 | 0.077 | 0.121 | 1 |
| LGALS9 | 0.198131086 | 0.072424107 | 0.2 | 0.166 | 1 |
| ZNF341 | 0.198626209 | -0.028441852 | 0.006 | 0.022 | 1 |
| PIK3AP1 | 0.205832672 | 0.118191894 | 0.652 | 0.611 | 1 |
| TRBC2 | 0.2076254 | 0.128796051 | 1 | 0.976 | 1 |
| ARPC1B | 0.210338557 | 0.101765388 | 0.677 | 0.558 | 1 |
| EIF3J | 0.210748108 | 0.094872551 | 0.735 | 0.68 | 1 |
| IL15 | 0.211744903 | -0.037719666 | 0.019 | 0.035 | 1 |
| PTPRC | 0.214932879 | 0.113694753 | 0.826 | 0.857 | 1 |
| BATF | 0.215716405 | -0.113440682 | 0.69 | 0.744 | 1 |
| CCL5 | 0.21585862 | -0.190566194 | 0.523 | 0.587 | 1 |
| MZB1 | 0.231899937 | 0.26024853 | 0.019 | 0.038 | 1 |
| NFATC2 | 0.23655433 | -0.09283313 | 0.226 | 0.318 | 1 |
| PRF1 | 0.236751209 | -0.24292 | 0.716 | 0.742 | 1 |
| HLA.DRA | 0.239985169 | 0.123384325 | 0.994 | 0.993 | 1 |
| CD7 | 0.240583817 | 0.104724599 | 0.419 | 0.377 | 1 |
| PRDM1 | 0.24916168 | 0.124212545 | 0.361 | 0.296 | 1 |
| CHD1 | 0.250987814 | -0.034710916 | 0.026 | 0.055 | 1 |
| PDIA4 | 0.256127412 | 0.098969006 | 0.626 | 0.592 | 1 |
| S100A9 | 0.257535508 | -0.028559667 | 0.019 | 0.038 | 1 |

| | | | | | |
|---|---|---|---|---|---|
| PCNA | 0.25787636 | 0.082862 | 0.548 | 0.503 | 1 |
| CD3G | 0.259259741 | 0.029536772 | 0.039 | 0.011 | 1 |
| IFITM2 | 0.260948506 | 0.113048508 | 0.548 | 0.508 | 1 |
| IL17F | 0.263405904 | -0.037719666 | 0.026 | 0.038 | 1 |
| SELPLG | 0.264226734 | 0.089222165 | 0.103 | 0.071 | 1 |
| HLA.DMA | 0.264294247 | -0.104567264 | 0.329 | 0.395 | 1 |
| ADGRG3 | 0.272176408 | 0.018133569 | 0.013 | 0.007 | 1 |
| TLR2 | 0.272599806 | 0.033539017 | 0.065 | 0.044 | 1 |
| WAS | 0.274797832 | 0.064460729 | 0.2 | 0.163 | 1 |
| BCL6 | 0.276779614 | 0.02329133 | 0.039 | 0.022 | 1 |
| FYB1 | 0.278010699 | -0.078505281 | 0.168 | 0.163 | 1 |
| MCM4 | 0.27866062 | -0.053929893 | 0.135 | 0.161 | 1 |
| SLC7A1 | 0.287705431 | 0.067106822 | 0.974 | 0.96 | 1 |
| TRIB1 | 0.291221872 | 0.133142455 | 0.484 | 0.492 | 1 |
| FCER1G | 0.296617326 | -0.071666998 | 0.219 | 0.234 | 1 |
| PIK3IP1 | 0.297489223 | 0.070690946 | 0.181 | 0.161 | 1 |
| NT5E | 0.304161454 | -0.101393411 | 0.31 | 0.294 | 1 |
| IFITM3 | 0.304280691 | 0.058956353 | 0.097 | 0.088 | 1 |
| ZAP70 | 0.308189754 | 0.041714802 | 0.058 | 0.042 | 1 |
| LAT2 | 0.317451902 | 0.07517439 | 0.245 | 0.234 | 1 |
| IRF9 | 0.320186976 | 0.080560685 | 0.71 | 0.715 | 1 |
| NINJ2 | 0.322625855 | 0.029111843 | 0.039 | 0.018 | 1 |
| KDELR1 | 0.322953056 | 0.059891131 | 0.329 | 0.258 | 1 |
| TNFRSF9 | 0.323170994 | 0.035232935 | 0.077 | 0.06 | 1 |
| TCF7 | 0.327521213 | -0.045987282 | 0.11 | 0.143 | 1 |
| IFNGR1 | 0.332531234 | 0.086898599 | 0.703 | 0.7 | 1 |
| IGBP1 | 0.334010076 | 0.057857994 | 0.252 | 0.245 | 1 |
| CX3CR1 | 0.33858198 | 0.102190356 | 0.161 | 0.172 | 1 |
| IL5 | 0.340310723 | -0.02222333 | 0.006 | 0.013 | 1 |
| AHI1 | 0.342435691 | -0.034784375 | 0.058 | 0.075 | 1 |
| PSME3 | 0.346681334 | 0.065620084 | 0.787 | 0.781 | 1 |
| KLRC4 | 0.346873536 | 0.026005932 | 0.032 | 0.024 | 1 |
| CCL3 | 0.353786883 | -0.123279123 | 1 | 1 | 1 |
| MCM2 | 0.353977685 | -0.048426172 | 0.135 | 0.166 | 1 |
| MS4A1 | 0.357818574 | -0.083289982 | 0.542 | 0.587 | 1 |
| IGHG3 | 0.359753992 | -0.046489876 | 0.039 | 0.057 | 1 |
| DUSP1 | 0.363773319 | -0.088884427 | 0.419 | 0.481 | 1 |
| TNIK | 0.365416818 | -0.051165197 | 0.123 | 0.148 | 1 |
| LAP3 | 0.365756664 | 0.071457282 | 0.632 | 0.539 | 1 |
| CD244 | 0.368334798 | -0.015977887 | 0.006 | 0.015 | 1 |
| PSMB8 | 0.369670105 | -0.071136216 | 0.91 | 0.876 | 1 |
| BCL11B | 0.381157653 | -0.07254503 | 0.161 | 0.163 | 1 |
| TNFRSF8 | 0.389805252 | 0.037317388 | 0.103 | 0.053 | 1 |
| IL2 | 0.392523479 | -0.169566867 | 1 | 1 | 1 |
| GZMB | 0.396603257 | 0.161142608 | 0.994 | 0.989 | 1 |
| KLRC3 | 0.399426156 | -0.14046884 | 0.245 | 0.302 | 1 |
| ITGAE | 0.399809397 | -0.031878787 | 0.058 | 0.079 | 1 |

| | | | | | |
|---|---|---|---|---|---|
| MAST4 | 0.401747793 | -0.050871034 | 0.129 | 0.163 | 1 |
| MMP9 | 0.409511529 | -0.034837157 | 0.065 | 0.091 | 1 |
| CD4 | 0.413240774 | 0.014635739 | 0.026 | 0.015 | 1 |
| SLC25A37 | 0.417192318 | -0.06104074 | 0.323 | 0.349 | 1 |
| CD1C | 0.418389841 | 0.044742494 | 0.052 | 0.026 | 1 |
| UGCG | 0.423465768 | -0.067698823 | 0.419 | 0.422 | 1 |
| B4GALT5 | 0.427178668 | 0.045582981 | 0.252 | 0.221 | 1 |
| CD9 | 0.434439617 | -0.045847547 | 0.11 | 0.132 | 1 |
| ATF6B | 0.437555888 | 0.042450683 | 0.148 | 0.13 | 1 |
| PAX5 | 0.446141927 | -0.012844997 | 0.006 | 0.013 | 1 |
| POU2F2 | 0.447255696 | 0.062544458 | 0.503 | 0.442 | 1 |
| NAMPT | 0.450157928 | -0.061470263 | 1 | 0.998 | 1 |
| CD6 | 0.453437277 | 0.071237318 | 0.645 | 0.651 | 1 |
| CTSD | 0.457614595 | 0.035557941 | 0.11 | 0.079 | 1 |
| CD36 | 0.457701528 | -0.022800246 | 0.045 | 0.062 | 1 |
| SRGN | 0.45916563 | -0.052944844 | 0.961 | 0.978 | 1 |
| CXCL16 | 0.459848747 | -0.034921033 | 0.103 | 0.113 | 1 |
| FURIN | 0.460344376 | 0.069921057 | 0.819 | 0.781 | 1 |
| FOXP1 | 0.463691789 | -0.05470702 | 0.755 | 0.768 | 1 |
| PMCH | 0.464240243 | -0.046879665 | 0.013 | 0.007 | 1 |
| THBS1 | 0.465517235 | -0.079388845 | 0.116 | 0.079 | 1 |
| CCR6 | 0.467884349 | 0.023680879 | 0.019 | 0.009 | 1 |
| MGST1 | 0.47021056 | 0.014973228 | 0.019 | 0.004 | 1 |
| IL6R | 0.478042582 | 0.036280915 | 0.135 | 0.106 | 1 |
| CXCL8 | 0.478324302 | -0.127542968 | 0.923 | 0.94 | 1 |
| DUSP5 | 0.481133592 | -0.054309361 | 0.987 | 0.993 | 1 |
| CD69 | 0.490572516 | 0.063853523 | 1 | 1 | 1 |
| LAMP1 | 0.491384973 | 0.049984934 | 0.71 | 0.698 | 1 |
| TRDC | 0.491637273 | 0.07558764 | 0.135 | 0.108 | 1 |
| APOBEC3G | 0.493532841 | 0.097122876 | 0.832 | 0.81 | 1 |
| CBLB | 0.497357352 | -0.026224027 | 0.077 | 0.099 | 1 |
| CD14 | 0.497794797 | -0.022517259 | 0.026 | 0.026 | 1 |
| LRRC32 | 0.498153065 | 0.084810809 | 0.277 | 0.256 | 1 |
| GNAI2 | 0.502541567 | 0.059813191 | 0.806 | 0.834 | 1 |
| CXCL1 | 0.502849882 | 0.028855809 | 0.077 | 0.06 | 1 |
| GIMAP5 | 0.503877946 | 0.047169232 | 0.219 | 0.234 | 1 |
| KLRF1 | 0.50617495 | -0.025570583 | 0.019 | 0.024 | 1 |
| GZMK | 0.508983964 | -0.022612774 | 0.019 | 0.038 | 1 |
| NCAM1 | 0.511621288 | -0.0130048 | 0.013 | 0.02 | 1 |
| CXCR4 | 0.512914217 | -0.107103797 | 0.439 | 0.499 | 1 |
| TIMM9 | 0.515216924 | -0.044344056 | 0.865 | 0.885 | 1 |
| BLNK | 0.517333574 | 0.030184399 | 0.103 | 0.079 | 1 |
| RAPH1 | 0.520214317 | -0.019685743 | 0.039 | 0.046 | 1 |
| NANS | 0.521052888 | -0.049226766 | 0.626 | 0.653 | 1 |
| NRP1 | 0.527766234 | 0.023309767 | 0.09 | 0.064 | 1 |
| EGR1 | 0.530054175 | -0.068144561 | 0.626 | 0.623 | 1 |
| PGM3 | 0.530593209 | 0.025516135 | 0.097 | 0.071 | 1 |

| | | | | | |
|---|---|---|---|---|---|
| CASP5 | 0.532430435 | -0.023249183 | 0.077 | 0.093 | 1 |
| KLRK1 | 0.538777835 | 0.088232995 | 0.587 | 0.629 | 1 |
| FUT4 | 0.545961531 | -0.016391157 | 0.026 | 0.031 | 1 |
| JCHAIN | 0.549175138 | 0.380696512 | 0.11 | 0.128 | 1 |
| BAX | 0.55085851 | -0.041944252 | 0.652 | 0.693 | 1 |
| HIF1A | 0.551822276 | -0.044852907 | 0.903 | 0.885 | 1 |
| IER3 | 0.553290479 | 0.088502186 | 0.955 | 0.954 | 1 |
| CD3E | 0.558832498 | 0.052855787 | 0.787 | 0.733 | 1 |
| STAT1 | 0.561194787 | 0.048968455 | 0.832 | 0.81 | 1 |
| BCL10 | 0.566304999 | -0.029682347 | 0.142 | 0.157 | 1 |
| CCR4 | 0.572385718 | 0.084702437 | 0.297 | 0.274 | 1 |
| DOCK8 | 0.574627335 | 0.038583701 | 0.29 | 0.294 | 1 |
| LIG4 | 0.583613289 | -0.035394613 | 0.265 | 0.285 | 1 |
| LAMP3 | 0.588620731 | -0.051678124 | 0.342 | 0.32 | 1 |
| CD79A | 0.591080586 | -0.063190353 | 0.329 | 0.349 | 1 |
| IL11RA | 0.59771164 | -0.016918347 | 0.052 | 0.06 | 1 |
| F5 | 0.604595933 | 0.074372842 | 0.316 | 0.23 | 1 |
| PAK1IP1 | 0.607817791 | 0.037989994 | 0.645 | 0.596 | 1 |
| CCR5 | 0.607947151 | 0.020344309 | 0.084 | 0.062 | 1 |
| CSRNP1 | 0.613760224 | -0.036217641 | 0.994 | 0.969 | 1 |
| IFRD1 | 0.622155549 | -0.035903813 | 0.484 | 0.494 | 1 |
| HLA.DQB1 | 0.623259936 | 0.051084736 | 0.974 | 0.965 | 1 |
| CD109 | 0.626736592 | -0.040171145 | 0.168 | 0.177 | 1 |
| ITGAM | 0.629361647 | -0.020125606 | 0.045 | 0.064 | 1 |
| CD24 | 0.633288532 | 0.021174023 | 0.129 | 0.093 | 1 |
| IFI6 | 0.640748548 | -0.016789008 | 0.045 | 0.042 | 1 |
| FCN1 | 0.642749798 | -0.010063323 | 0.019 | 0.024 | 1 |
| CD200 | 0.648447493 | -0.073343576 | 0.632 | 0.592 | 1 |
| IL12A | 0.65008246 | -0.042746483 | 0.097 | 0.146 | 1 |
| LGALS3 | 0.661593013 | 0.031686341 | 0.284 | 0.283 | 1 |
| DPP4 | 0.662794378 | -0.010063323 | 0.013 | 0.024 | 1 |
| IL12RB1 | 0.664639746 | -0.009885458 | 0.013 | 0.013 | 1 |
| SOCS2 | 0.670034089 | 0.042922518 | 0.245 | 0.245 | 1 |
| POU2AF1 | 0.677253361 | -0.055136719 | 0.606 | 0.631 | 1 |
| NFATC3 | 0.679424387 | 0.026729042 | 0.284 | 0.227 | 1 |
| ERBIN | 0.68589698 | 0.034291579 | 0.658 | 0.625 | 1 |
| SNX9 | 0.688479993 | -0.028981329 | 0.948 | 0.954 | 1 |
| SP140 | 0.689315762 | -0.021325228 | 0.135 | 0.119 | 1 |
| AQP9 | 0.691400005 | -0.006759357 | 0.013 | 0.018 | 1 |
| CD2 | 0.697526654 | 0.049422897 | 0.845 | 0.863 | 1 |
| CD274 | 0.700193318 | -0.040769762 | 0.806 | 0.788 | 1 |
| SOCS3 | 0.704415079 | -0.03328742 | 0.542 | 0.583 | 1 |
| DCLRE1C | 0.710401151 | 0.014431616 | 0.077 | 0.071 | 1 |
| TYMS | 0.712638369 | 0.008376749 | 0.019 | 0.018 | 1 |
| TNIP3 | 0.712638378 | 0.008376749 | 0.026 | 0.015 | 1 |
| TNFRSF13C | 0.729543135 | 0.029919051 | 0.439 | 0.375 | 1 |
| FBXO11 | 0.747487708 | 0.023444197 | 0.523 | 0.453 | 1 |

| | | | | | |
|---|---|---|---|---|---|
| CNOT2 | 0.753279706 | -0.022506572 | 0.561 | 0.558 | 1 |
| CTSW | 0.764777493 | 0.021383854 | 0.168 | 0.137 | 1 |
| VPS28 | 0.770293042 | -0.022059322 | 0.413 | 0.408 | 1 |
| CD37 | 0.77144668 | -0.022279304 | 0.8 | 0.828 | 1 |
| NFKB2 | 0.774749089 | 0.022844776 | 0.523 | 0.554 | 1 |
| CCND2 | 0.774758734 | -0.029814468 | 0.735 | 0.768 | 1 |
| IRF7 | 0.775195448 | -0.024215402 | 0.594 | 0.578 | 1 |
| CD300A | 0.775795787 | -0.021417854 | 0.103 | 0.086 | 1 |
| MITF | 0.778948066 | -0.003405301 | 0.006 | 0.009 | 1 |
| IKZF2 | 0.778948066 | -0.003405301 | 0.006 | 0.009 | 1 |
| IGHG2 | 0.780509588 | -0.014212913 | 0.026 | 0.051 | 1 |
| TIAF1 | 0.791447552 | -0.007346017 | 0.026 | 0.033 | 1 |
| TNFRSF25 | 0.794020917 | 0.021415458 | 0.387 | 0.364 | 1 |
| IL12RB2 | 0.80269534 | -0.021600001 | 0.206 | 0.225 | 1 |
| RUNX3 | 0.803966591 | 0.02186913 | 0.781 | 0.81 | 1 |
| THEMIS | 0.81858443 | -0.008275888 | 0.058 | 0.062 | 1 |
| RNASE6 | 0.826754672 | 0.010880973 | 0.019 | 0.013 | 1 |
| PLCG2 | 0.829005222 | -0.009974675 | 0.11 | 0.121 | 1 |
| SMAD2 | 0.830949957 | 0.012403813 | 0.277 | 0.26 | 1 |
| CHD7 | 0.832772901 | -0.010446736 | 0.129 | 0.135 | 1 |
| CCR1 | 0.833990153 | -0.009813669 | 0.103 | 0.113 | 1 |
| CD44 | 0.840008935 | -0.011474359 | 1 | 1 | 1 |
| IRF1 | 0.84264338 | 0.014872506 | 0.994 | 0.98 | 1 |
| ITGAX | 0.845568126 | 0.025642395 | 0.219 | 0.267 | 1 |
| ILF2 | 0.84690096 | -0.009898238 | 1 | 0.998 | 1 |
| MYB | 0.857089059 | -0.009150514 | 0.058 | 0.079 | 1 |
| CYBB | 0.857347561 | -0.011488124 | 0.135 | 0.148 | 1 |
| CD74 | 0.85787208 | 0.016603347 | 0.994 | 0.998 | 1 |
| SKAP1 | 0.85893477 | -0.004060355 | 0.026 | 0.026 | 1 |
| QPCT | 0.863615646 | -0.004273183 | 0.032 | 0.033 | 1 |
| VMO1 | 0.864001009 | -0.004483336 | 0.039 | 0.042 | 1 |
| AIM2 | 0.867102266 | -0.004060355 | 0.026 | 0.024 | 1 |
| PDIA6 | 0.867240843 | -0.013838598 | 0.961 | 0.954 | 1 |
| STIM1 | 0.869630625 | -0.00960603 | 0.2 | 0.183 | 1 |
| IL9 | 0.870018292 | -0.004273183 | 0.026 | 0.033 | 1 |
| TREM1 | 0.87284667 | 0.002659796 | 0.013 | 0.009 | 1 |
| PDCD1 | 0.87827066 | 0.006134624 | 0.058 | 0.062 | 1 |
| ZNF683 | 0.879922941 | 0.004984785 | 0.026 | 0.018 | 1 |
| CXCL9 | 0.881167041 | 0.03918832 | 0.11 | 0.091 | 1 |
| IL3RA | 0.88502577 | 0.007723305 | 0.084 | 0.073 | 1 |
| TGFBI | 0.893240949 | -0.006262234 | 0.097 | 0.075 | 1 |
| GEM | 0.901381707 | 0.02044336 | 0.8 | 0.715 | 1 |
| LTB | 0.901408767 | 0.002400643 | 0.019 | 0.015 | 1 |
| TNFSF10 | 0.901897622 | 0.007723305 | 0.065 | 0.062 | 1 |
| ITGB2 | 0.902255409 | -0.01039906 | 0.316 | 0.313 | 1 |
| FOXP3 | 0.907044546 | -0.0038448 | 0.006 | 0.013 | 1 |
| NDFIP2 | 0.907839935 | -0.00669277 | 0.181 | 0.172 | 1 |

| | | | | | |
|---|---|---|---|---|---|
| STAT6 | 0.912732784 | -0.00803675 | 0.652 | 0.589 | 1 |
| FOXO1 | 0.931963187 | 0.0047156 | 0.142 | 0.152 | 1 |
| CREM | 0.935038944 | 0.006942199 | 0.955 | 0.932 | 1 |
| B3GAT1 | 0.940474983 | 0.005500863 | 0.148 | 0.155 | 1 |
| RPN2 | 0.948805514 | 0.004998334 | 0.871 | 0.779 | 1 |
| GRB10 | 0.96495119 | -0.00119379 | 0.032 | 0.026 | 1 |
| VEGFA | 0.968717358 | -0.000725458 | 0.019 | 0.02 | 1 |
| CD79B | 0.970989222 | -0.000725458 | 0.019 | 0.018 | 1 |
| CD247 | 0.971216241 | -0.001754169 | 0.129 | 0.11 | 1 |
| LAT | 0.971403628 | -0.00323129 | 0.368 | 0.369 | 1 |
| TGFBR1 | 0.972598301 | -0.00231433 | 0.329 | 0.318 | 1 |
| TRAT1 | 0.980310977 | 0.000912994 | 0.045 | 0.046 | 1 |
| S100A12 | 0.981935877 | -0.000244961 | 0.006 | 0.007 | 1 |
| NGDN | 0.984356712 | 0.000675713 | 0.052 | 0.06 | 1 |
| CXCR5 | 0.987381848 | -0.0016638 | 0.929 | 0.945 | 1 |
| XBP1 | 0.991545673 | -0.000831637 | 0.903 | 0.916 | 1 |
| ZBTB16 | 0.99303792 | -0.050076872 | 0 | 0.029 | 1 |
| CD163 | 0.993220117 | -0.031501143 | 0 | 0.018 | 1 |
| ENTPD1 | 0.995609766 | -0.037719666 | 0 | 0.009 | 1 |
| IL23R | 0.995705102 | -0.0252557 | 0 | 0.015 | 1 |
| IL26 | 0.995772744 | -0.018983103 | 0 | 0.011 | 1 |
| VNN2 | 0.995935721 | -0.009522774 | 0 | 0.007 | 1 |
| TOP2A | 0.99731942 | -0.015836548 | 0 | 0.007 | 1 |
| CXCL5 | 0.997576501 | -0.000244961 | 0.31 | 0.347 | 1 |
| TNFAIP8 | 0.99882897 | -9.46E-05 | 1 | 1 | 1 |
