## Supplementary Table 5 for "Human germline biallelic complete NFAT1 deficiency causes the triad of progressive joint contractures, osteochondromas, and susceptibility to B cell malignancy"

Supplementary Table 5: List of 500 genes used for single cell targeted panel

| | | | | | | | | |
|---|---|---|---|---|---|---|---|---|
| *ABCE1* | *CCR7* | *CMTM2* | *FCER1A* | *IGHM* | *ITGAX* | *MZB1* | *RNASE6* | *TNFAIP8* |
| *ADA* | *CCR8* | *CNOT2* | *FCER1G* | *IGKC* | *ITGB2* | *NAMPT* | *RORA* | *TNFRSF13C* |
| *ADGRE1* | *CCR9* | *CR2* | *FCER2* | *IGLC3* | *ITK* | *NANS* | *RORC* | *TNFRSF17* |
| *ADGRG3* | *CD109* | *CREM* | *FCGR3A* | *IKZF1* | *IVNS1ABP* | *NCAM1* | *RPL6* | *TNFRSF25* |
| *AHI1* | *CD14* | *CSF1* | *FCN1* | *IKZF2* | *JAK1* | *NDFIP2* | *RPN2* | *TNFRSF4* |
| *AIM2* | *CD160* | *CSF2* | *FN1* | *IL11RA* | *JAK2* | *NFATC1* | *RUNX3* | *TNFRSF8* |
| *ALAS2* | *CD163* | *CSRNP1* | *FOS* | *IL12A* | *JAK3* | *NFATC2* | *S100A10* | *TNFRSF9* |
| *ANXA5* | *CD1C* | *CST7* | *FOSB* | *IL12RB1* | *JCHAIN* | *NFATC3* | *S100A12* | *TNFSF10* |
| *APOBEC3G* | *CD2* | *CTLA4* | *FOSL1* | *IL12RB2* | *JUN* | *NFATC4* | *S100A9* | *TNFSF13B* |
| *APOE* | *CD200* | *CTSD* | *FOXO1* | *IL13* | *JUNB* | *NFKB1* | *SDCBP* | *TNFSF14* |
| *AQP9* | *CD209* | *CTSG* | *FOXP1* | *IL15* | *KCNE3* | *NFKB2* | *SELL* | *TNFSF8* |
| *ARG1* | *CD22* | *CTSW* | *FOXP3* | *IL15RA* | *KDELR1* | *NFKBIA* | *SELPLG* | *TNIK* |
| *ARHGEF3* | *CD226* | *CX3CR1* | *FTH1* | *IL17A* | *KIAA0101* | *NFKBID* | *SKAP1* | *TNIP3* |
| *ARID5A* | *CD24* | *CXCL1* | *FURIN* | *IL17F* | *KIR2DL1* | *NFKBIZ* | *SLA* | *TOP2A* |
| *ARL4C* | *CD244* | *CXCL10* | *FUT4* | *IL17RB* | *KIT* | *NGDN* | *SLC1A5* | *TPSAB1* |
| *ARPC1B* | *CD247* | *CXCL11* | *FYB/FYB1* | *IL18* | *KLRB1* | *NINJ2* | *SLC25A37* | *TRAC* |
| *ATF4* | *CD27* | *CXCL13* | *FYN* | *IL18R1* | *KLRC1* | *NKG7* | *SLC7A1* | *TRAF1* |
| *ATF6B* | *CD274* | *CXCL16* | *GAB2* | *IL18RAP* | *KLRC3* | *NR4A2* | *SLC7A7* | *TRAT1* |
| *AURKB* | *CD28* | *CXCL2* | *GAPDH* | *IL1A* | *KLRC4* | *NRP1* | *SMAD2* | *TRBC2* |
| *B3GAT1* | *CD300A* | *CXCL3* | *GATA3* | *IL1B* | *KLRF1* | *NT5E* | *SMAD3* | *TRDC* |
| *B4GALT5* | *CD33* | *CXCL5* | *GBP1* | *IL1R1* | *KLRG1* | *ORAI1* | *SNCA* | *TREM1* |
| *BACH2* | *CD34* | *CXCL8* | *GBP2* | *IL1R2* | *KLRK1* | *OSM* | *SNX8* | *TRIB1* |
| *BATF* | *CD36* | *CXCL9* | *GEM* | *IL1RL1* | *LAG3* | *PAK1IP1* | *SNX9* | *TRIB2* |
| *BAX* | *CD37* | *CXCR2* | *GIMAP2* | *IL1RN* | *LAMP1* | *PASK* | *SOCS1* | *TSLP* |
| *BCAT1* | *CD38* | *CXCR3* | *GIMAP5* | *IL2* | *LAMP3* | *PAX5* | *SOCS2* | *TSPAN32* |
| *BCL10* | *CD3D* | *CXCR4* | *GNAI2* | *IL21* | *LAP3* | *PAXBP1* | *SOCS3* | *TUBA1C* |
| *BCL11B* | *CD3E* | *CXCR5* | *GNLY* | *IL22* | *LAT* | *PCNA* | *SP140* | *TXK* |
| *BCL2* | *CD3G* | *CXCR6* | *GRB10* | *IL23A* | *LAT2* | *PDCD1* | *SPAG9* | *TYMS* |
| *BCL2A1* | *CD4* | *CYBB* | *GZMA* | *IL23R* | *LCK* | *PDIA4* | *SPP1* | *UFM1* |
| *BCL2L1* | *CD40* | *CYCS* | *GZMB* | *IL26* | *LDHA* | *PDIA6* | *SRGN* | *UGCG* |
| *BCL6* | *CD40LG* | *DCLRE1C* | *GZMH* | *IL2RA* | *LEF1* | *PFN1* | *STAT1* | *VEGFA* |
| *BIN2* | *CD44* | *DOCK8* | *GZMK* | *IL2RB* | *LGALS1* | *PGM3* | *STAT3* | *VMO1* |
| *BIRC3* | *CD48* | *DPP4* | *HAVCR2* | *IL2RG* | *LGALS3* | *PIGF* | *STAT4* | *VNN2* |
| *BLK* | *CD5* | *DUSP1* | *HIF1A* | *IL3* | *LGALS9* | *PIK3AP1* | *STAT5A* | *VPREB3* |
| *BLNK* | *CD52* | *DUSP2* | *HLA-A* | *IL31* | *LIF* | *PIK3IP1* | *STAT6* | *VPS28* |
| *BTG1* | *CD6* | *DUSP4* | *HLA-DMA* | *IL32* | *LIG4* | *PLCG2* | *STIM1* | *VSIG4* |
| *BTLA* | *CD63* | *DUSP5* | *HLA-DPA1* | *IL33* | *LILRB4* | *PMCH* | *TBX21* | *WAS* |
| *C10orf54/VSI*[illegible] | *CD69* | *DUSP6* | *HLA-DQB1* | *IL3RA* | *LIPA* | *POU2AF1* | *TCF4* | *XBP1* |
| *CARD11* | *CD7* | *EBF1* | *HLA-DRA* | *IL4* | *LRRC32* | *POU2F2* | *TCF7* | *YBX3* |
| *CASP5* | *CD70* | *EEF1E1* | *HNRNPC* | *IL4R* | *LRRC8B* | *PRDM1* | *TCL1A* | *ZAP70* |
| *CBLB* | *CD72* | *EGR1* | *ICAM1* | *IL5* | *LTA* | *PRF1* | *TGFB1* | *ZBED2* |
| *CCL11* | *CD74* | *EGR3* | *ICOS* | *IL5RA* | *LTB* | *PRNP* | *TGFB3* | *ZBTB16* |
| *CCL13* | *CD79A* | *EIF1* | *IER3* | *IL6* | *LY86* | *PSEN1* | *TGFBI* | *ZNF341* |
| *CCL2* | *CD79B* | *EIF3J* | *IFI6* | *IL6R* | *LYN* | *PSMB8* | *TGFBR1* | *ZNF683* |

| | | | | | | | |
|---|---|---|---|---|---|---|---|
| *CCL20* | *CD80* | *EIF5* | *IFITM2* | *IL6ST* | *MALT1* | *PSMB9* | *TGFBR2* |
| *CCL22* | *CD83* | *EIF5A* | *IFITM3* | *IL7R* | *MAP2K3* | *PSME2* | *TGIF1* |
| *CCL3* | *CD86* | *ENTPD1* | *IFNG* | *IL9* | *MAST4* | *PSME3* | *THBD* |
| *CCL4* | *CD8A* | *EOMES* | *IFNGR1* | *ILF2* | *MCM2* | *PTGDR2* | *THBS1* |
| *CCL5* | *CD8B* | *ERBB2IP* | *IFRD1* | *IRF1* | *MCM4* | *PTPN7* | *THEMIS* |
| *CCND2* | *CD9* | *EVI2A* | *IGBP1* | *IRF3* | *MGST1* | *PTPRC* | *TIAF1* |
| *CCR1* | *CHD1* | *EVI2B* | *IGHA1* | *IRF4* | *MITF* | *QPCT* | *TIGIT* |
| *CCR10* | *CHD7* | *F13A1* | *IGHD* | *IRF7* | *MKI67* | *RAG1* | *TIMM9* |
| *CCR2* | *CHI3L2* | *F5* | *IGHE* | *IRF8* | *MME* | *RAPH1* | *TLR2* |
| *CCR3* | *CLEC10A* | *FAM129C* | *IGHG1* | *IRF9* | *MMP9* | *RBCK1* | *TLR7* |
| *CCR4* | *CLEC4D* | *FAS* | *IGHG2* | *ITGA4* | *MS4A1* | *REL* | *TLR8* |
| *CCR5* | *CLEC4E* | *FASLG* | *IGHG3* | *ITGAE* | *MYB* | *RGS1* | *TMEM97* |
| *CCR6* | *CMKLR1* | *FBXO11* | *IGHG4* | *ITGAM* | *MYC* | *RNASE2* | *TNF* |
