## Supplementary Table 6 for "Human germline biallelic complete NFAT1 deficiency causes the triad of progressive joint contractures, osteochondromas, and susceptibility to B cell malignancy"

Supplementary Table 6: List of antibodies used in six different flow cytometry panels

| *Marker* | *Fluorophore* | *Clone* | *Catalogue #:* |
|---|---|---|---|
| **T-cell panel** | | | |
| Cell surface markers | | | |
| Anti-CD3 | BV510 | UCHT1 | 563109 (BD) |
| Anti-CD8 | BV570 | HIT8a | 563550 (BD) |
| Anti-CD4 | BUV395 | SK3 | 624298 (BD) |
| Anti-CD27 | BV605 | L128 | 562655 (BD) |
| Anti-CD45RA | BV421 | HI100 | 562885 (BD) |
| Anti-PD-1 | BUV737 | EH12.1 | 612792 (BD) |
| Anti-CXCR5 | PeCy7 | MU5UBEE | 25-9185-42 (eBiosciences) |
| Anti-CD154 | PeCy5 | TRAP1 | 561722 (BD) |
| Anti-CD69 | PerCPCy5.5 | FN50 | 560738 (BD) |
| Intracellular markers | | | |
| Anti-TNF | R718 | Mab11 | 566957 (BD) |
| Anti-IL-2 | BV650 | MQ1-17H12 | 564166 (BD) |
| Anti-IFNγ | BV711 | 4S.B3 | 564793 (BD) |
| Anti-IL-4 | PeCF594 | MP4-25D2 | 565161 (BD) |
| Anti-IL-17A | AF488 | N49-653 | 560488 (BD) |
| Anti-IL-21 | AF647 | 3A3-N2.1 | 560493 (BD) |
| Fixable viability stain | AF780 | | 565388 (BD) |
| **B-cell panel** | | | |
| Cell surface markers | | | |
| Anti-CD19 | BUV737 | HIB19 | 741829 (BD) |

| | | | |
|---|---|---|---|
| Anti-IgD | BV605 | IA6-2 | 563313 (BD) |
| Anti-IgM | BB515 | G20-127 | 564622 (BD) |
| Anti-CD38 | BV786 | HIT2 | 563964 (BD) |
| Anti-CD27 | BB630 | M-T271 | Custom (BD) |
| Intracellular markers | | | |
| Anti-IL-6 | BV421 | MQ2-13A5 | 563279 (BD) |
| Anti-IL-10 | BB700 | JES3-19F1 | 566568 (BD) |
| Anti-TNF | BUV395 | Mab11 | 563996 (BD) |
| Anti-Ki67 | AF647 | Ki-67 | 350514 (BD) |
| Fixable viability stain | AF780 | | 565388 (BD) |
| **T-reg panel** | | | |
| Cell surface markers | | | |
| Anti-CD3 | BV510 | UCTH1 | 563109 (BD) |
| Anti-CD4 | BUV395 | SK3 | 563550 (BD) |
| Anti-CD8 | BV570 | HIT8a | 624298 (BD) |
| Anti-CD45RA | APC | 5H9 | 550855 (BD) |
| Anti-CD25 | BUV805 | 2A3 | 742070 (BD) |
| Anti-CD69 | PerCPCy5.5 | FN50 | 560738 (BD) |
| Anti-PD-1 | PeCy7 | EH12.1 | 561272 (BD) |
| Anti-CD152 | BV421 | BNI3 | 562743 (BD) |
| Anti-CD69 | PerCPCy5.5 | FN50 | 560738 (BD) |
| Anti-CD278 | BV786 | DX29 | 741017 (BD) |
| Anti-CD127 | PeCF594 | HIL-7R-M21 | 562397 (BD) |
| Intracellular markers | | | |
| Anti-FoxP3 | AF488 | 236A/E7 | 561181 (BD) |
| Anti-Helios | PE | 22F6 | 137206 (BD) |
| Anti-TNF | R718 | Mab11 | 566957 (BD) |
| Anti-IL-2 | BV650 | MQ1-17H12 | 564166 (BD) |
| Anti-IFNγ | BV711 | 4S_B3 | 564793 (BD) |
| Fixable viability stain | AF780 | | 565388 (BD) |
| **Plasmablasts differentiation panel** | | | |
| Cell surface markers | | | |
| Anti-CD19 | BUV737 | HIB19 | 741829 (BD) |
| Anti-IgD | BUV615 | IA6-2 | 613008 (BD) |
| Anti-IgM | BB515 | G20-127 | 564622 (BD) |

| | | | |
|---|---|---|---|
| Anti-CD38 | BV786 | HIT2 | 563964 (BD) |
| Anti-CD27 | BV605 | L128 | 562655 (BD) |
| Anti-CD21 | PeCF594 | B-ly4 | 563474 (BD) |
| Fixable viability stain | AF780 | | 565388 (BD) |
| **T-cell rescue panel** | | | |
| Cell surface markers | | | |
| Anti-CD3 | BV510 | UCHT1 | 563109 (BD) |
| Anti-CD8 | BV570 | HIT8a | 563550 (BD) |
| Anti-CD4 | BUV395 | SK3 | 624298 (BD) |
| Anti-CD27 | BV605 | L128 | 562655 (BD) |
| Anti-CD45RA | BV421 | HI100 | 562885 (BD) |
| Anti-CD154 | PeCy5 | TRAP1 | 561722 (BD) |
| Anti-CD69 | PerCPCy5.5 | FN50 | 560738 (BD) |
| GFP | GFP/AF488 | | |
| Intracellular markers | | | |
| Anti-TNF | R718 | Mab11 | 566957 (BD) |
| Anti-p65(pS529) | PeCy7 | K10-895.12.50 | 560335 (BD) |
| *Primary rabbit-anti-NFATC2<br><br>Secondary anti-rabbit | AF647 | JA11-08 | NBP2-66974 (Novus biologicals)<br><br>A32733 (Thermo Fisher) |
| Fixable viability stain | AF780 | | 565388 (BD) |
| **NFAT1 staining panel** | | | |
| Cell surface markers | | | |
| Anti-CD3 | APC-H7 | SK7 | 560176 (BD) |
| Anti-CD4 | BUV563 | L200 | 749214 (BD) |
| Anti-CD8 | BV570 | HIT8a | 624298 (BD) |
| Anti-CD27 | BB630 | M-T271 | Custom (BD) |
| Anti-CD45RA | BV421 | HI100 | 562885 (BD) |
| Anti-CD19 | BUV395 | SJ25C1 | 563594 (BD) |
| Anti-CXCR5 | PeCy7 | MU5UBEE | 25-9185-42 (eBiosciences) |
| Anti-IgD | BV605 | IA6-2 | 563313 (BD) |
| Anti-IgM | BB515 | G20-127 | 564622 (BD) |
| Anti-PD-1 | BUV737 | EH12.1 | 612792 (BD) |

| | | | |
|---|---|---|---|
| Anti-CD38 | BV786 | HIT2 | 563964 (BD) |
| Intracellular makers | | | |
| Anti-p65(pS529) | PeCy7 | K10-895.12.50 | 560335 (BD) |
| *Primary rabbit-anti-NFATC2<br><br>Secondary anti-rabbit | AF647 | JA11-08 | NBP2-66974 (Novus biologicals)<br><br>A32733 (Thermo Fisher) |
| Fixable viability stain | AF780 | | 565388 (BD) |

*A primary/secondary approach was used to stain for NFAT1, primary antibody for rabit-anti-human-NFATC2 (Novus Biologicals, Clone: JA11-08, Cat#: NBP2-66974) was stained for 1hour at room temperature followed by a wash and secondary stain anti-rabbit-AF647 (Thermo Fisher, Cat#: A32733) for 30 mins.
