## Supplementary Table 4 for "Human germline biallelic complete NFAT1 deficiency causes the triad of progressive joint contractures, osteochondromas, and susceptibility to B cell malignancy"

Supplementary Table 4 : Differential gene expression under different conditions
between patient cells transduced with EV and patient cells rescued with WT-NFATC2

**Unstimulated**

| | logFC | AveExpr | t | P.Value | adj.P.Val | B |
|---|---|---|---|---|---|---|
| NFATC2 | -7.147768522 | 5.463722861 | -29.44800906 | 1.29E-06 | 0.017787582 | 4.829772182 |
| PI16 | 1.311839336 | 2.049845425 | 20.83253335 | 6.71E-06 | 0.036348563 | 4.087490906 |
| ALPL | 1.451726786 | 1.620900964 | 19.36002852 | 9.52E-06 | 0.036348563 | 3.888878194 |
| ADGRD1 | 1.296688413 | 3.964517623 | 18.78771239 | 1.10E-05 | 0.036348563 | 3.803297492 |
| PENK | -1.111111635 | 9.564690436 | -17.39896379 | 1.58E-05 | 0.036348563 | 3.573052769 |
| RAC2 | 1.132293274 | 1.216732964 | 16.57587647 | 1.99E-05 | 0.036348563 | 3.419545568 |
| CPE | 1.084217334 | 3.210758922 | 16.3755655 | 2.11E-05 | 0.036348563 | 3.380054229 |
| IFNE | -1.058077172 | 1.506284467 | -16.37032853 | 2.11E-05 | 0.036348563 | 3.379010033 |
| TACSTD2 | -1.391405102 | 3.68162159 | -15.71140587 | 2.56E-05 | 0.036348563 | 3.242668526 |
| GJB2 | 1.021529271 | 2.202718991 | 15.63045615 | 2.63E-05 | 0.036348563 | 3.225216456 |
| SFRP2 | 1.479501219 | 4.979006505 | 15.09498408 | 3.10E-05 | 0.038963026 | 3.105668393 |
| TRNP1 | 1.063678594 | 2.305997464 | 14.81147127 | 3.39E-05 | 0.039061579 | 3.039360367 |
| PXDNL | 0.990918773 | 3.289484915 | 14.55812077 | 3.68E-05 | 0.039067983 | 2.978251512 |
| TGFA | 0.96358577 | 3.722195933 | 14.33484165 | 3.95E-05 | 0.039067983 | 2.922891077 |
| SERPING1 | 1.023923595 | 7.178674808 | 14.08789107 | 4.29E-05 | 0.039575676 | 2.859961547 |
| CTAG2 | 0.876902949 | 0.832941349 | 13.64291796 | 4.99E-05 | 0.041656112 | 2.741859296 |
| DSG2 | 1.674426436 | 3.546963477 | 13.33771971 | 5.55E-05 | 0.041656112 | 2.657177193 |
| PTGS2 | -0.847595848 | 7.952260681 | -12.85150802 | 6.61E-05 | 0.041656112 | 2.515699191 |
| WISP2 | 0.819023706 | 9.09580925 | 12.80053995 | 6.73E-05 | 0.041656112 | 2.500380955 |
| MALAT1 | -0.779711412 | 11.95791843 | -12.48172498 | 7.58E-05 | 0.041656112 | 2.402370857 |
| NRN1 | -0.943756423 | 4.002110436 | -12.32853969 | 8.03E-05 | 0.041656112 | 2.353898242 |
| PARM1 | 0.984244263 | 1.309596235 | 12.281165 | 8.18E-05 | 0.041656112 | 2.338721149 |
| FAM20A | 0.927616761 | 5.43867527 | 12.23899403 | 8.31E-05 | 0.041656112 | 2.32513618 |
| WSCD2 | -1.373766627 | 3.814912178 | -12.23750348 | 8.31E-05 | 0.041656112 | 2.324654715 |
| PLXDC1 | 1.025000718 | 4.428188006 | 12.22806178 | 8.34E-05 | 0.041656112 | 2.321602879 |
| FAM107A | -0.957390685 | 3.853674966 | -12.21753886 | 8.38E-05 | 0.041656112 | 2.318197352 |
| C2 | 0.80999298 | 3.521464943 | 12.20348106 | 8.42E-05 | 0.041656112 | 2.313640908 |
| CPXM1 | 0.909409674 | 3.647219236 | 12.20094692 | 8.43E-05 | 0.041656112 | 2.312818691 |
| GPRC5B | 0.793917157 | 2.116174616 | 12.00542582 | 9.09E-05 | 0.043377142 | 2.248594541 |
| CDH2 | 0.938922678 | 2.327522411 | 11.82120347 | 9.77E-05 | 0.044193519 | 2.186632797 |
| XPNPEP2 | 0.803948017 | 1.897968276 | 11.73095274 | 0.000101295 | 0.044193519 | 2.155751884 |
| MBP | 1.021926879 | 2.883987439 | 11.70835063 | 0.000102211 | 0.044193519 | 2.147963175 |
| ADORA1 | 1.343471927 | 3.284002892 | 11.57533805 | 0.00010781 | 0.045201982 | 2.101674892 |
| SEPP1 | 0.721536834 | 6.544012569 | 11.49560051 | 0.000111344 | 0.045310455 | 2.073551171 |
| COMP | -0.753994526 | 10.18608658 | -11.4205359 | 0.000114798 | 0.045381427 | 2.046814719 |
| CELF2 | 0.873911203 | 5.377059053 | 11.3186549 | 0.000119695 | 0.046002817 | 2.010116525 |
| SYT12 | -0.764830543 | 3.878803729 | -11.22644278 | 0.000124345 | 0.046340726 | 1.976487874 |
| CTSH | 0.956201373 | 3.253526568 | 11.10338438 | 0.000130893 | 0.046340726 | 1.930987806 |
| SORL1 | -1.019746433 | 2.38639005 | -11.10002242 | 0.000131078 | 0.046340726 | 1.929734645 |
| NLGN4X | 0.727355047 | 0.433601406 | 11.04805831 | 0.000133971 | 0.046340726 | 1.910296163 |
| PLA2G4A | -0.729976203 | 5.618134251 | -10.90964121 | 0.000142063 | 0.047304947 | 1.857878847 |
| LINC00342 | -0.744109618 | 1.255359526 | -10.74783455 | 0.000152277 | 0.047304947 | 1.795403537 |

| | | | | | | |
|---|---|---|---|---|---|---|
| CD70 | -0.859463562 | 3.759634018 | -10.66209997 | 0.000158047 | 0.047304947 | 1.761764818 |
| ADAM23 | 0.677927274 | 4.194987006 | 10.63783386 | 0.000159727 | 0.047304947 | 1.752175414 |
| C3 | 0.666602476 | 6.186116963 | 10.6130967 | 0.000161463 | 0.047304947 | 1.742368576 |
| SERPINB2 | 1.288414779 | 3.480108568 | 10.60056046 | 0.00016235 | 0.047304947 | 1.737386589 |
| STC1 | -0.712983626 | 4.813246794 | -10.57729764 | 0.000164014 | 0.047304947 | 1.728120148 |
| CXCL12 | 1.160124892 | 6.328412738 | 10.57594938 | 0.000164111 | 0.047304947 | 1.727582226 |
| OXTR | 0.782141969 | 2.466124736 | 10.49402065 | 0.000170138 | 0.048041527 | 1.694715801 |
| RANBP17 | 0.801287524 | 1.012601172 | 10.4442259 | 0.000173932 | 0.048130395 | 1.674567152 |
| LGR4 | 0.720777546 | 5.509086841 | 10.36153688 | 0.000180458 | 0.048722945 | 1.640815871 |
| SNX10 | 0.692326353 | 0.613606967 | 10.32887843 | 0.000183116 | 0.048722945 | 1.627384126 |

**PMA/Iono (50ng/mL PMA + 1uM Ionomycin) stimulation**

| | logFC | AveExpr | t | P.Value | adj.P.Val | B |
|---|---|---|---|---|---|---|
| NFATC2 | -9.036233532 | 6.168695986 | -108.1963557 | 4.77E-09 | 6.60E-05 | 6.289218494 |
| ZNF462 | -1.910790323 | 3.996195306 | -22.92552857 | 6.12E-06 | 0.024150944 | 4.448779892 |
| EPB41L3 | 1.695413206 | 3.064610593 | 22.47897315 | 6.70E-06 | 0.024150944 | 4.393089696 |
| DSG2 | 1.868932185 | 3.599451862 | 22.10081409 | 7.24E-06 | 0.024150944 | 4.34421581 |
| CA2 | -1.290131508 | 1.160670352 | -19.40952087 | 1.31E-05 | 0.024150944 | 3.944667803 |
| FAM107A | -1.976076275 | 4.285047957 | -19.36774428 | 1.33E-05 | 0.024150944 | 3.937663636 |
| S1PR1 | -1.246402989 | 3.654275909 | -18.80910903 | 1.52E-05 | 0.024150944 | 3.841341726 |
| CFI | 1.260350059 | 1.186076656 | 17.60852093 | 2.05E-05 | 0.024150944 | 3.616335739 |
| SLC7A2 | 1.281829094 | 2.699731515 | 17.56444648 | 2.07E-05 | 0.024150944 | 3.607573987 |
| F3 | 1.252668551 | 6.731918179 | 16.76318666 | 2.56E-05 | 0.024150944 | 3.441567778 |
| HOXD1 | -1.628558251 | 0.944716891 | -16.57509742 | 2.70E-05 | 0.024150944 | 3.400673494 |
| SERPINB2 | 1.653858213 | 5.189966948 | 16.4742594 | 2.78E-05 | 0.024150944 | 3.378434153 |
| TACSTD2 | -2.422200398 | 3.387711643 | -16.30145849 | 2.91E-05 | 0.024150944 | 3.339802336 |
| SERPING1 | 1.084038555 | 7.167263275 | 16.26641176 | 2.94E-05 | 0.024150944 | 3.33188584 |
| PIM1 | -1.136411732 | 4.378463978 | -15.94053283 | 3.23E-05 | 0.024150944 | 3.256931637 |
| HAGLR | -1.229293156 | 2.315965754 | -15.74641909 | 3.41E-05 | 0.024150944 | 3.211104175 |
| UCP2 | -1.089044524 | 5.631806133 | -15.72566452 | 3.43E-05 | 0.024150944 | 3.206150984 |
| SFRP2 | 1.35433507 | 4.9172706 | 15.17058259 | 4.04E-05 | 0.024150944 | 3.069726802 |
| TNF | 1.011465156 | 1.597944938 | 15.04670063 | 4.19E-05 | 0.024150944 | 3.038208579 |
| PENK | -0.974416657 | 10.14329792 | -14.99629234 | 4.26E-05 | 0.024150944 | 3.025268159 |
| FAM20A | 1.043399982 | 5.225530627 | 14.95363727 | 4.31E-05 | 0.024150944 | 3.014265419 |
| CXCL12 | 1.155547387 | 6.197956514 | 14.88962916 | 4.40E-05 | 0.024150944 | 2.997663526 |
| ADGRD1 | 1.296544873 | 4.170345809 | 14.8658113 | 4.43E-05 | 0.024150944 | 2.991457746 |
| MMP9 | 1.752068893 | 5.241566355 | 14.62107444 | 4.78E-05 | 0.024150944 | 2.92679516 |
| GJB2 | 1.196490163 | 3.855109751 | 14.58191582 | 4.84E-05 | 0.024150944 | 2.91629534 |
| PPARGC1A | 1.062100104 | 1.50469468 | 14.47380787 | 5.00E-05 | 0.024150944 | 2.887083921 |
| ARL4D | -1.446771607 | 4.63895439 | -14.46812623 | 5.01E-05 | 0.024150944 | 2.885539557 |
| TANC1 | -1.01195971 | 4.172467092 | -14.40478194 | 5.11E-05 | 0.024150944 | 2.868259149 |
| TXNIP | 1.096334724 | 4.690449895 | 14.30571494 | 5.27E-05 | 0.024150944 | 2.841002273 |
| CMKLR1 | -1.371797179 | 6.449047332 | -14.23999032 | 5.39E-05 | 0.024150944 | 2.822761812 |
| RGS4 | 1.32930976 | 5.42764791 | 14.22547484 | 5.41E-05 | 0.024150944 | 2.818716304 |
| RASGRP3 | -1.018344419 | 1.211454762 | -14.00616984 | 5.81E-05 | 0.024542466 | 2.756834595 |
| E2F7 | 1.046268857 | 3.278753944 | 13.98097078 | 5.85E-05 | 0.024542466 | 2.74963165 |
| MCF2L | 0.892089096 | 2.86167652 | 13.84677986 | 6.12E-05 | 0.024841697 | 2.710947705 |

| | | | | | | |
|---|---|---|---|---|---|---|
| FOXP2 | 0.917822291 | 1.656009478 | 13.72959522 | 6.36E-05 | 0.024841697 | 2.676710796 |
| SYNPO | -1.071997368 | 5.790556148 | -13.56251893 | 6.72E-05 | 0.024841697 | 2.627148952 |
| REM1 | 0.893949489 | 2.432780396 | 13.54506774 | 6.76E-05 | 0.024841697 | 2.621920741 |
| MEGF10 | -1.069514024 | 1.123452751 | -13.51597408 | 6.82E-05 | 0.024841697 | 2.613182704 |
| MARCKSL1 | 1.011427336 | 6.548128302 | 13.3955409 | 7.10E-05 | 0.025205185 | 2.576718773 |
| LMO4 | 0.916305941 | 6.502006246 | 13.22427081 | 7.53E-05 | 0.025356512 | 2.524038185 |
| AMPH | 1.048881358 | 2.774515392 | 13.17012299 | 7.67E-05 | 0.025356512 | 2.507178252 |
| HS3ST3B1 | 0.963534424 | 4.722756741 | 13.16039621 | 7.70E-05 | 0.025356512 | 2.504139102 |
| SYNGAP1 | -0.910519806 | 4.15569567 | -12.98675045 | 8.17E-05 | 0.025686745 | 2.449337155 |
| KCNK3 | 1.148761887 | 1.563069504 | 12.92635982 | 8.35E-05 | 0.025686745 | 2.430032945 |
| DNAJB4 | 1.08111858 | 6.68772012 | 12.92393902 | 8.35E-05 | 0.025686745 | 2.429256457 |
| COL6A6 | 1.746053201 | 2.071843533 | 12.8217009 | 8.66E-05 | 0.02604604 | 2.396273669 |
| IL1RN | 1.081091066 | 3.136920457 | 12.63721637 | 9.24E-05 | 0.026219288 | 2.335809535 |
| CSF3 | 1.344646143 | 2.194471936 | 12.55330854 | 9.53E-05 | 0.026219288 | 2.307898219 |
| HOXD10 | -0.817076313 | 2.010903835 | -12.51970146 | 9.64E-05 | 0.026219288 | 2.29664598 |
| PPARG | 0.841711968 | 2.847529355 | 12.47611986 | 9.80E-05 | 0.026219288 | 2.281991349 |
| BTG2 | -0.87114033 | 5.362037704 | -12.44699104 | 9.90E-05 | 0.026219288 | 2.272156862 |
| WISP2 | 0.834794255 | 9.032051807 | 12.43847368 | 9.93E-05 | 0.026219288 | 2.26927519 |
| CILP | 0.890994664 | 2.9205418 | 12.31233245 | 0.00010397 | 0.026219288 | 2.226275678 |
| STARD9 | -1.324790566 | 3.682875161 | -12.27789627 | 0.000105291 | 0.026219288 | 2.214431161 |
| IGSF3 | -1.032839132 | 2.574844216 | -12.26215775 | 0.000105901 | 0.026219288 | 2.209002575 |
| PLEKHH3 | -1.337547969 | 4.67842254 | -12.25653221 | 0.00010612 | 0.026219288 | 2.207059871 |
| DLX5 | 1.002868356 | 4.286344876 | 12.20430908 | 0.000108181 | 0.026259623 | 2.18896664 |
| PMEL | 0.894872263 | 2.267349642 | 12.13840425 | 0.000110852 | 0.026443995 | 2.165981174 |
| DEPDC1 | 0.940546313 | 1.944945713 | 11.66740035 | 0.000132454 | 0.030318989 | 1.996624146 |
| ORAI1 | -0.764560962 | 3.67780411 | -11.575873 | 0.000137225 | 0.030318989 | 1.962645612 |
| KCNJ6 | 1.043287423 | 1.613993903 | 11.54778609 | 0.000138731 | 0.030318989 | 1.952146855 |
| FAM84A | 1.089429688 | 1.424170397 | 11.52736645 | 0.000139838 | 0.030318989 | 1.944492769 |
| C2orf88 | 0.923205532 | 1.315885373 | 11.50487702 | 0.000141069 | 0.030318989 | 1.936042002 |
| CHI3L1 | 1.137569975 | 12.11822063 | 11.47339448 | 0.000142816 | 0.030318989 | 1.924175085 |
| CTSH | 0.884694236 | 3.344396743 | 11.45287289 | 0.000143968 | 0.030318989 | 1.916416525 |
| RUSC1 | -1.438190908 | 4.521256223 | -11.40442329 | 0.000146734 | 0.030318989 | 1.898026173 |
| KCND2 | 1.006370575 | 4.048269903 | 11.34868654 | 0.000149996 | 0.030318989 | 1.876741943 |
| LGR5 | -1.094943408 | 4.554504935 | -11.28837488 | 0.000153624 | 0.030318989 | 1.853555306 |
| FAM49A | -0.824057489 | 4.302016845 | -11.28516579 | 0.00015382 | 0.030318989 | 1.852317028 |
| CTAG2 | 1.175090661 | 0.865567272 | 11.2757791 | 0.000154396 | 0.030318989 | 1.848692374 |
| HSPB8 | -1.71574487 | 7.96187276 | -11.2515544 | 0.000155892 | 0.030318989 | 1.839319762 |
| PTGES | 0.77859271 | 5.021875231 | 11.19974128 | 0.000159152 | 0.030318989 | 1.819184213 |
| ADORA1 | 0.899234652 | 2.555475089 | 11.16465894 | 0.000161406 | 0.030318989 | 1.805481387 |
| SLC37A1 | 1.01858422 | 1.821907814 | 11.13318854 | 0.000163461 | 0.030318989 | 1.793141532 |
| CNTN3 | 1.101988167 | 3.177000301 | 11.09457966 | 0.000166026 | 0.030318989 | 1.777940508 |
| PCDH9 | 0.730137864 | 2.659489355 | 11.04765399 | 0.000169208 | 0.030318989 | 1.759372314 |
| SYNDIG1 | 0.881504957 | 1.883406746 | 10.92679029 | 0.000177752 | 0.030318989 | 1.711073914 |
| WSCD2 | -1.413285183 | 3.801873942 | -10.90241596 | 0.000179538 | 0.030318989 | 1.701250211 |
| RASGEF1B | 0.738184419 | 1.857606377 | 10.89870616 | 0.000179812 | 0.030318989 | 1.699752557 |
| ZNF385A | -0.785584215 | 3.811212555 | -10.88632762 | 0.000180729 | 0.030318989 | 1.694750569 |
| CCL5 | 0.89626474 | 2.571880653 | 10.86103908 | 0.000182619 | 0.030318989 | 1.684509112 |

| | | | | | | |
|---|---|---|---|---|---|---|
| FAM19A5 | -0.722258964 | 4.891630739 | -10.84760573 | 0.000183633 | 0.030318989 | 1.67905637 |
| LONRF3 | -0.906439111 | 3.14612447 | -10.84313863 | 0.000183972 | 0.030318989 | 1.677241209 |
| TNRC18 | -1.004997537 | 5.945481023 | -10.82625651 | 0.000185259 | 0.030318989 | 1.670372668 |
| C3 | 0.803370592 | 6.262022089 | 10.78126505 | 0.000188743 | 0.030318989 | 1.65200064 |
| ADAM19 | 0.839833512 | 0.840929575 | 10.7404862 | 0.000191969 | 0.030318989 | 1.635264022 |
| SLC22A4 | -0.978834814 | 4.28938809 | -10.7323206 | 0.000192622 | 0.030318989 | 1.631902931 |
| CPE | 0.82986969 | 3.22970343 | 10.72966868 | 0.000192835 | 0.030318989 | 1.630810654 |
| TMEM255A | 0.781948273 | 1.231111258 | 10.65596365 | 0.00019887 | 0.030820392 | 1.600314888 |
| EML6 | -1.151734196 | 3.875220476 | -10.63675695 | 0.00020048 | 0.030820392 | 1.592324024 |
| CYP7B1 | 1.175084381 | 1.805626754 | 10.60346187 | 0.000203308 | 0.030911706 | 1.57842835 |
| IL1A | 0.90497705 | 3.192060377 | 10.54971012 | 0.000207976 | 0.031277771 | 1.555878271 |
| HOXC10 | -3.444615605 | 6.451150284 | -10.46294698 | 0.000215788 | 0.031784437 | 1.519171913 |
| PXDNL | 0.989237703 | 3.368317777 | 10.42774919 | 0.000219058 | 0.031784437 | 1.504171789 |
| FCRLB | 0.875305253 | 2.699671658 | 10.3941946 | 0.000222231 | 0.031784437 | 1.489812756 |
| MEG3 | -0.678557347 | 6.566729905 | -10.3752032 | 0.000224051 | 0.031784437 | 1.481660038 |
| GLDN | -1.346121156 | 2.355161473 | -10.33589065 | 0.000227878 | 0.031784437 | 1.464724383 |
| DAPK1 | 0.775596571 | 4.116112218 | 10.28861994 | 0.000232583 | 0.031784437 | 1.444253768 |
| C17orf96 | -0.723392658 | 3.843666407 | -10.27228878 | 0.000234236 | 0.031784437 | 1.437154336 |
| KCND3 | 0.861471793 | 2.147057732 | 10.26649684 | 0.000234826 | 0.031784437 | 1.434633114 |
| PIK3CD | 0.68015684 | 3.524933024 | 10.26454034 | 0.000235026 | 0.031784437 | 1.433781054 |
| PI16 | 1.144088703 | 1.988797996 | 10.24934985 | 0.000236582 | 0.031784437 | 1.427158685 |
| DOCK10 | 0.660256866 | 5.395519268 | 10.24903836 | 0.000236614 | 0.031784437 | 1.427022762 |
| LINC00607 | 0.69903617 | 1.771460087 | 10.18102842 | 0.00024374 | 0.031917298 | 1.397222518 |
| THBS1 | 0.674001416 | 11.29678294 | 10.15438705 | 0.000246601 | 0.031917298 | 1.385481744 |
| PNLIPRP3 | 0.766512175 | 0.613413967 | 10.15383058 | 0.000246661 | 0.031917298 | 1.385236108 |
| BCAR1 | 0.704386393 | 6.212819086 | 10.10854623 | 0.000251621 | 0.031917298 | 1.365190621 |
| IFI44L | -1.841847209 | 2.552985631 | -10.09036936 | 0.000253645 | 0.031917298 | 1.35711335 |
| JAG1 | -0.803640999 | 7.640617208 | -10.08004436 | 0.000254804 | 0.031917298 | 1.352517251 |
| TGFA | 0.650405038 | 2.745941665 | 10.00694465 | 0.000263193 | 0.031917298 | 1.319811275 |
| IRS2 | 0.660484633 | 5.549151157 | 9.9993583 | 0.000264083 | 0.031917298 | 1.316400258 |
| IL1RL1 | 0.872672151 | 4.19427918 | 9.995912282 | 0.000264488 | 0.031917298 | 1.314849795 |
| COL27A1 | -0.659092379 | 5.303614508 | -9.978186769 | 0.000266585 | 0.031917298 | 1.306864233 |
| CILP2 | -0.692879376 | 4.65504607 | -9.977271998 | 0.000266693 | 0.031917298 | 1.306451647 |
| PLXDC1 | 0.881920453 | 4.460749587 | 9.957646845 | 0.000269039 | 0.031917298 | 1.297589039 |
| TDRP | 0.992585249 | 1.755482655 | 9.955575937 | 0.000269288 | 0.031917298 | 1.296652584 |
| IL1B | 1.475555288 | 4.547017525 | 9.949143171 | 0.000270064 | 0.031917298 | 1.293742203 |
| GUCY1A2 | 0.848587559 | 2.838095837 | 9.883354401 | 0.000278149 | 0.031917298 | 1.263845075 |
| KIAA1467 | 0.731313617 | 2.432904323 | 9.870544529 | 0.000279757 | 0.031917298 | 1.25799558 |
| NPTX1 | -0.702967099 | 9.61265331 | -9.86093316 | 0.000280971 | 0.031917298 | 1.253600591 |
| PALM | 0.688338782 | 5.493872766 | 9.834548756 | 0.000284337 | 0.031917298 | 1.241509032 |
| COL1A1 | -0.726682167 | 10.14318049 | -9.811435483 | 0.000287325 | 0.031917298 | 1.230884233 |
| SEPP1 | 0.665678118 | 6.513522484 | 9.805412276 | 0.00028811 | 0.031917298 | 1.228110484 |
| RARRES2 | 1.066627035 | 0.677152242 | 9.796152205 | 0.000289322 | 0.031917298 | 1.223842103 |
| SLC37A2 | 0.666557493 | 5.552752903 | 9.794459913 | 0.000289544 | 0.031917298 | 1.223061522 |
| SFRP4 | 0.971531819 | 8.892622146 | 9.78597655 | 0.000290661 | 0.031917298 | 1.219146058 |
| SOCS1 | 0.833176372 | 3.629504873 | 9.764078545 | 0.000293567 | 0.031931402 | 1.209020134 |
| MYOZ3 | -0.779169211 | 0.549247488 | -9.75036781 | 0.000295405 | 0.031931402 | 1.202666145 |

| | | | | | | |
|---|---|---|---|---|---|---|
| NTRK2 | -0.807092379 | 7.501061046 | -9.707267318 | 0.000301273 | 0.032163888 | 1.182621663 |
| PIK3C2B | -2.953481886 | 1.896428101 | -9.647413723 | 0.000309657 | 0.032163888 | 1.154607712 |
| CDH2 | 1.218628113 | 1.807054591 | 9.638770619 | 0.000310891 | 0.032163888 | 1.150545154 |
| HES7 | -1.007503774 | 0.634509485 | -9.61488306 | 0.000314332 | 0.032163888 | 1.139294443 |
| AGTR1 | 0.960341541 | 2.879680434 | 9.595159179 | 0.000317208 | 0.032163888 | 1.12997953 |
| DCLK3 | 1.273521637 | 1.999432561 | 9.591725301 | 0.000317712 | 0.032163888 | 1.128355488 |
| CXorf57 | 0.695904945 | 0.784965202 | 9.57467133 | 0.000320229 | 0.032163888 | 1.120279562 |
| RASL12 | -1.539588784 | 1.52857923 | -9.560465191 | 0.000322344 | 0.032163888 | 1.11353911 |
| CDH4 | 0.769401056 | 1.029960012 | 9.53184259 | 0.000326657 | 0.032163888 | 1.099922077 |
| ALPL | 1.326897146 | 1.165484037 | 9.529108837 | 0.000327073 | 0.032163888 | 1.098618962 |
| GALNT12 | 0.639599909 | 5.289977256 | 9.527537927 | 0.000327312 | 0.032163888 | 1.097869946 |
| LOC153684 | -1.053365256 | 0.822857602 | -9.511506398 | 0.000329764 | 0.032163888 | 1.090217635 |
| KLF6 | 0.668993742 | 8.095507546 | 9.484077949 | 0.000334011 | 0.032163888 | 1.077089619 |
| IL11 | -0.836016801 | 8.999952211 | -9.434777491 | 0.000341812 | 0.032163888 | 1.053379406 |
| GPRC5B | 0.712882964 | 1.895135967 | 9.413968518 | 0.000345171 | 0.032163888 | 1.04332761 |
| ADAMTS9 | -1.867954603 | 3.956979542 | -9.411433442 | 0.000345583 | 0.032163888 | 1.042101244 |
| FLVCR2 | -1.165404995 | 2.39236706 | -9.372869192 | 0.000351922 | 0.032163888 | 1.023397181 |
| ENPP4 | 1.016277915 | 1.825073143 | 9.37019876 | 0.000352366 | 0.032163888 | 1.022098633 |
| BIRC5 | 0.667144632 | 2.679249059 | 9.362497569 | 0.000353651 | 0.032163888 | 1.018351338 |
| KLHL3 | 0.890355047 | 0.992255658 | 9.347336035 | 0.000356197 | 0.032163888 | 1.010963318 |
| MME | 0.612323433 | 9.821355825 | 9.340323525 | 0.000357382 | 0.032163888 | 1.007541438 |
| ACE | 0.64519403 | 5.831421184 | 9.324232577 | 0.000360119 | 0.032163888 | 0.999678132 |
| PACSIN3 | -0.871699175 | 4.102638047 | -9.323186236 | 0.000360298 | 0.032163888 | 0.999166255 |
| CDA | 0.621815851 | 2.737630743 | 9.303841227 | 0.000363624 | 0.032163888 | 0.989690362 |
| MAFB | 0.713947401 | 7.465714582 | 9.293171318 | 0.000365474 | 0.032163888 | 0.984453944 |
| PAPL | 0.911920895 | 3.217926205 | 9.285940255 | 0.000366734 | 0.032163888 | 0.980901178 |
| FER1L6 | 0.628042481 | 4.044953705 | 9.282184458 | 0.000367391 | 0.032163888 | 0.979054599 |
| QPRT | 0.750411706 | 2.490059834 | 9.260519038 | 0.000371207 | 0.032163888 | 0.968385431 |
| PBK | 0.698619039 | 2.988695953 | 9.251129397 | 0.000372876 | 0.032163888 | 0.963752402 |
| PDGFD | 0.705108536 | 5.50122224 | 9.24491997 | 0.000373985 | 0.032163888 | 0.960685527 |
| ENPP5 | 0.889847964 | 0.918143511 | 9.233520803 | 0.00037603 | 0.032163888 | 0.95504913 |
| NFKBID | -0.934505605 | 4.819382821 | -9.233328052 | 0.000376065 | 0.032163888 | 0.954953753 |
| TRAF1 | 0.706645445 | 6.285509841 | 9.224030845 | 0.000377743 | 0.032163888 | 0.950350545 |
| PPP1R1B | 0.678128096 | 0.516801078 | 9.222986988 | 0.000377932 | 0.032163888 | 0.949833374 |
| TNFRSF21 | 0.618826571 | 4.293570112 | 9.208354344 | 0.000380594 | 0.032163888 | 0.942576548 |
| TSPAN2 | -0.739860215 | 1.541278092 | -9.204806024 | 0.000381243 | 0.032163888 | 0.940814786 |
| AMOTL2 | -0.748001561 | 6.462193162 | -9.154777494 | 0.000390535 | 0.032598547 | 0.915890716 |
| SMURF2 | 0.636564765 | 6.262973357 | 9.134479892 | 0.000394382 | 0.032598547 | 0.905733253 |
| AMOT | 0.90496342 | 3.309883555 | 9.129537887 | 0.000395326 | 0.032598547 | 0.903256169 |
| PTGER4 | -0.624189701 | 3.441786001 | -9.126001934 | 0.000396003 | 0.032598547 | 0.901482885 |
| BCR | -0.604281975 | 6.203339091 | -9.109621009 | 0.000399157 | 0.032598547 | 0.893257415 |
| ANKRD52 | -1.102718426 | 6.273581892 | -9.098097359 | 0.000401394 | 0.032598547 | 0.887460673 |
| TRABD2A | 0.824689284 | 0.829461074 | 9.072198657 | 0.000406477 | 0.032598547 | 0.874401734 |
| MID2 | -1.231270062 | 4.884067544 | -9.067815816 | 0.000407345 | 0.032598547 | 0.872187498 |
| ADAMTS8 | -1.090898522 | 2.101749166 | -9.066531722 | 0.0004076 | 0.032598547 | 0.871538531 |
| CEMIP | 0.853451161 | 10.45036592 | 9.006057638 | 0.000419815 | 0.032969445 | 0.84085475 |
| DMKN | 0.643462959 | 6.724283549 | 8.995906339 | 0.000421908 | 0.032969445 | 0.835680809 |

| | | | | | | |
|---|---|---|---|---|---|---|
| MGC12916 | 1.043772103 | 1.952531893 | 8.988489842 | 0.000423446 | 0.032969445 | 0.831896495 |
| PTCH1 | -0.870705593 | 2.071298658 | -8.986375969 | 0.000423885 | 0.032969445 | 0.83081722 |
| NRN1 | -0.837475638 | 4.027355394 | -8.985096186 | 0.000424152 | 0.032969445 | 0.830163662 |
| STXBP6 | 0.603760524 | 3.838662375 | 8.929829036 | 0.000435848 | 0.033680147 | 0.801837335 |
| TACC2 | -0.587365712 | 6.233348642 | -8.919098599 | 0.000438163 | 0.033680147 | 0.79631428 |
| WISP1 | -3.797700015 | 4.454768465 | -8.898598142 | 0.000442629 | 0.033730471 | 0.785741301 |
| ABCB5 | 0.940317397 | 1.421362137 | 8.880697691 | 0.000446573 | 0.033730471 | 0.776486427 |
| TENM4 | 0.705576415 | 1.85489363 | 8.880526184 | 0.000446611 | 0.033730471 | 0.776397652 |
| APBB1IP | 0.578020378 | 5.704689406 | 8.843680598 | 0.000454864 | 0.033730471 | 0.757280063 |
| ADAM23 | 0.667494189 | 4.188356808 | 8.836758501 | 0.000456435 | 0.033730471 | 0.75367834 |
| CCL3 | 0.910489497 | 3.330367944 | 8.832272609 | 0.000457457 | 0.033730471 | 0.751342511 |
| IRF4 | 0.823434394 | 2.852090937 | 8.829864796 | 0.000458007 | 0.033730471 | 0.750088192 |
| ABCC3 | -0.694186849 | 4.032020811 | -8.828488702 | 0.000458321 | 0.033730471 | 0.749371158 |
| GLCCI1 | -0.645248694 | 0.948656834 | -8.817225287 | 0.000460904 | 0.033732943 | 0.743497401 |
| SPON1 | 0.899876041 | 4.168987919 | 8.80587482 | 0.000463525 | 0.033732943 | 0.737569596 |
| TLR4 | 0.676558203 | 4.867980619 | 8.796648148 | 0.000465669 | 0.033732943 | 0.732744537 |
| COL15A1 | 0.63872808 | 5.092564843 | 8.784085176 | 0.000468607 | 0.033768981 | 0.726165509 |
| GPSM2 | -0.625013803 | 4.975061226 | -8.758958564 | 0.000474551 | 0.033865247 | 0.712974974 |
| SIPA1L2 | -0.846300883 | 4.145960969 | -8.743529708 | 0.000478245 | 0.033865247 | 0.704854104 |
| EVI2A | -0.780463825 | 4.296093364 | -8.736090916 | 0.000480039 | 0.033865247 | 0.700932942 |
| RNF144B | 0.583328217 | 4.487248091 | 8.727788205 | 0.000482051 | 0.033865247 | 0.696551918 |
| FAM124A | 0.646013311 | 3.88712458 | 8.727253429 | 0.000482181 | 0.033865247 | 0.696269576 |
| EGR3 | -0.716705898 | 6.726849064 | -8.681604186 | 0.000493428 | 0.034480169 | 0.672096011 |
| LSAMP | 0.62637421 | 3.742798667 | 8.601680189 | 0.000513892 | 0.035562114 | 0.629424994 |
| KLF2 | 0.644191843 | 3.835457976 | 8.601071103 | 0.000514052 | 0.035562114 | 0.629098096 |
| PRDM1 | 0.702215244 | 4.564764941 | 8.581096623 | 0.000519327 | 0.035748301 | 0.618363325 |
| ZNF92 | 0.699528907 | 3.359091897 | 8.562620751 | 0.000524264 | 0.035909521 | 0.608408968 |
| AQP1 | -0.641130806 | 6.707949304 | -8.552396968 | 0.000527021 | 0.035920503 | 0.602890294 |
| PCDHGA3 | -0.774067236 | 1.735492812 | -8.498618275 | 0.000541811 | 0.036421475 | 0.573739249 |
| FOXP4 | -0.852642798 | 5.409554054 | -8.498507285 | 0.000541843 | 0.036421475 | 0.573678873 |
| ADCY8 | 1.063890217 | 4.378173878 | 8.49698554 | 0.000542268 | 0.036421475 | 0.572850997 |
| NAV3 | 0.59430085 | 6.195864597 | 8.482755866 | 0.000546269 | 0.03651295 | 0.565101609 |
| CNIH3 | 0.699513433 | 5.118306585 | 8.469546446 | 0.000550015 | 0.036541984 | 0.557894874 |
| SYNM | -0.658635346 | 6.61158863 | -8.462640896 | 0.000551986 | 0.036541984 | 0.554122395 |
| PRR16 | 0.785749605 | 3.375611936 | 8.453318677 | 0.000554659 | 0.036544135 | 0.549024262 |
| ROR2 | 0.841600814 | 0.82531931 | 8.426450126 | 0.000562453 | 0.036716196 | 0.534295372 |
| CELF2 | 0.677571494 | 5.643195364 | 8.40796057 | 0.000567894 | 0.036716196 | 0.524129401 |
| HPD | -0.781565863 | 1.816156654 | -8.402914924 | 0.000569389 | 0.036716196 | 0.521350887 |
| ARMCX5 | -0.72212636 | 3.334631735 | -8.396361056 | 0.000571339 | 0.036716196 | 0.517739072 |
| RFX2 | -1.15936246 | 4.073454244 | -8.375731882 | 0.000577528 | 0.036716196 | 0.506349998 |
| SLC39A10 | -0.687843336 | 5.347788279 | -8.374812316 | 0.000577806 | 0.036716196 | 0.505841596 |
| GCOM1 | 0.825127475 | 1.289246856 | 8.357406869 | 0.000583095 | 0.036716196 | 0.496206973 |
| PSD4 | 0.721008142 | 3.02506644 | 8.356413608 | 0.000583398 | 0.036716196 | 0.495656494 |
| LZTS1 | 0.60914841 | 0.620115925 | 8.345951978 | 0.000586607 | 0.036716196 | 0.489854125 |
| ARHGAP42 | -0.615965787 | 3.130849349 | -8.345671207 | 0.000586693 | 0.036716196 | 0.489698289 |
| FOXL1 | 0.545419614 | 4.732841625 | 8.330064441 | 0.00059152 | 0.036716196 | 0.481027001 |
| PDK4 | -0.681264098 | 4.19392401 | -8.327884306 | 0.000592199 | 0.036716196 | 0.479814269 |

| | | | | | | |
|---|---|---|---|---|---|---|
| NGFR | -2.271893369 | 1.897817794 | -8.322312237 | 0.000593936 | 0.036716196 | 0.476713132 |
| LINC00856 | 0.566994522 | 3.829991723 | 8.320755758 | 0.000594422 | 0.036716196 | 0.475846465 |
| RAB36 | 0.699336476 | 3.045200902 | 8.310082063 | 0.000597771 | 0.03675893 | 0.4698984 |
| RBP4 | -0.534489425 | 8.404274372 | -8.30139751 | 0.000600512 | 0.036764108 | 0.465052603 |
| TTC9 | -0.670313946 | 4.812539024 | -8.280001267 | 0.000607331 | 0.036897826 | 0.453090128 |
| PLXNC1 | 0.719473331 | 4.93884454 | 8.277824185 | 0.00060803 | 0.036897826 | 0.451871034 |
| JHDM1D-AS1 | 0.538679189 | 6.304204514 | 8.199542335 | 0.000633821 | 0.038294986 | 0.407800706 |
| PDGFA | -0.817089716 | 4.18454607 | -8.138035175 | 0.000655014 | 0.039403363 | 0.372850288 |
| CNTNAP3 | -0.801707576 | 2.518287027 | -8.090318481 | 0.000672045 | 0.040252852 | 0.345537559 |
| TNC | -0.529134072 | 11.19594146 | -8.049697537 | 0.000686964 | 0.040622293 | 0.322148445 |
| FAM65C | 0.747860588 | 4.387713668 | 8.048889257 | 0.000687265 | 0.040622293 | 0.321681751 |
| ZFHX2 | -1.117258366 | 1.467480099 | -8.04831242 | 0.00068748 | 0.040622293 | 0.32134866 |
| LGI2 | 0.966430167 | 4.175970098 | 8.041677798 | 0.000689957 | 0.040622293 | 0.317515677 |
| PTGS1 | 0.703025544 | 5.936739867 | 8.026072382 | 0.000695825 | 0.040794194 | 0.308486572 |
| DCHS1 | -0.543502642 | 8.274401661 | -7.997851987 | 0.00070659 | 0.041105851 | 0.292110417 |
| HDAC9 | 0.589171955 | 2.189486546 | 7.991114037 | 0.000709189 | 0.041105851 | 0.288191215 |
| LBH | 0.720456678 | 7.483186874 | 7.98888149 | 0.000710053 | 0.041105851 | 0.286891845 |
| KCNA1 | -0.643922882 | 5.872316645 | -7.957562642 | 0.000722308 | 0.04148031 | 0.26862263 |
| ZBTB21 | -0.601083756 | 6.398168815 | -7.936903032 | 0.000730531 | 0.04148031 | 0.25652896 |
| SH3BP2 | 0.579667012 | 2.57163587 | 7.9363769 | 0.000730742 | 0.04148031 | 0.256220534 |
| PARM1 | 1.214766288 | 2.422626654 | 7.93549935 | 0.000731094 | 0.04148031 | 0.255706052 |
| TRPM3 | -0.602464323 | 1.277543627 | -7.934457719 | 0.000731512 | 0.04148031 | 0.255095295 |
| CCDC8 | 0.767693122 | 1.551832575 | 7.900844766 | 0.000745155 | 0.041823657 | 0.235340248 |
| ABAT | 0.872856596 | 2.854467849 | 7.890156057 | 0.000749557 | 0.041823657 | 0.229039426 |
| DKK2 | -0.70725476 | 1.142280337 | -7.883827119 | 0.000752179 | 0.041823657 | 0.22530432 |
| SDC1 | 0.590764562 | 6.734127589 | 7.880125341 | 0.000753717 | 0.041823657 | 0.223118184 |
| CDC42EP3 | 0.598521669 | 6.686076074 | 7.877452315 | 0.00075483 | 0.041823657 | 0.221538909 |
| CDH13 | -0.677294588 | 7.048671453 | -7.853185233 | 0.000765028 | 0.041823657 | 0.20717526 |
| SDK1 | 0.550907752 | 0.862989223 | 7.843115589 | 0.000769308 | 0.041823657 | 0.201201165 |
| TMEM132B | -1.072657317 | 3.368254144 | -7.832428091 | 0.000773882 | 0.041823657 | 0.194851576 |
| CSF2 | -1.031201459 | 4.767688823 | -7.826727811 | 0.000776335 | 0.041823657 | 0.191461192 |
| FAM225B | 0.574785349 | 1.165259997 | 7.825788046 | 0.00077674 | 0.041823657 | 0.190901991 |
| CCKAR | -0.973501192 | 2.405697132 | -7.818888376 | 0.000779724 | 0.041823657 | 0.186794203 |
| JAK2 | -1.01042731 | 4.05611588 | -7.818055053 | 0.000780085 | 0.041823657 | 0.186297816 |
| TSLP | 0.654639603 | 3.9651435 | 7.81731553 | 0.000780406 | 0.041823657 | 0.185857255 |
| RHOB | 0.597012494 | 7.206829325 | 7.808612665 | 0.000784193 | 0.041823657 | 0.180669313 |
| RAB27B | 0.584989821 | 4.22489528 | 7.805226374 | 0.000785673 | 0.041823657 | 0.178649022 |
| TAGLN | 0.580094383 | 7.646638763 | 7.796126742 | 0.000789665 | 0.041823657 | 0.173215491 |
| NR4A3 | -0.693647255 | 10.37641574 | -7.790992791 | 0.000791928 | 0.041823657 | 0.170146961 |
| GRIN2A | 0.618321565 | 3.631368462 | 7.787257963 | 0.00079358 | 0.041823657 | 0.167913331 |
| FBLIM1 | 0.944403497 | 2.54429454 | 7.783825852 | 0.000795101 | 0.041823657 | 0.165859741 |
| FRK | -0.86540835 | 0.993779048 | -7.777254861 | 0.000798023 | 0.041823657 | 0.161925335 |
| IRAK3 | -1.212401624 | 4.959034972 | -7.751231541 | 0.000809723 | 0.042017825 | 0.146309145 |
| PI3 | 0.749280363 | 3.610877624 | 7.739665197 | 0.00081499 | 0.042017825 | 0.13935056 |
| AMMECR1 | -0.825411693 | 3.65441149 | -7.739263716 | 0.000815173 | 0.042017825 | 0.139108822 |
| BCAR3 | 0.6415115 | 6.089514471 | 7.73378259 | 0.000817684 | 0.042017825 | 0.13580723 |
| MAP7D3 | -0.528855612 | 4.990621767 | -7.724770196 | 0.000821833 | 0.042017825 | 0.130373184 |

| | | | | | | |
|---|---|---|---|---|---|---|
| NPAT | -0.542413947 | 3.890799557 | -7.72182941 | 0.000823192 | 0.042017825 | 0.128598582 |
| SEMA6D | 0.869820351 | 1.957394998 | 7.713353832 | 0.000827125 | 0.042017825 | 0.123480051 |
| ID2 | 0.510371914 | 8.260845723 | 7.705812968 | 0.000830642 | 0.042017825 | 0.118921022 |
| PCOLCE2 | -0.505997748 | 9.011698792 | -7.703580571 | 0.000831687 | 0.042017825 | 0.117570466 |
| EN1 | 0.727704701 | 4.908540909 | 7.695250883 | 0.0008356 | 0.042017825 | 0.11252753 |
| TRPV4 | -0.704766588 | 3.709793649 | -7.682076909 | 0.000841833 | 0.042017825 | 0.104540042 |
| STON2 | 0.969992747 | 3.260961994 | 7.681908008 | 0.000841913 | 0.042017825 | 0.104437543 |
| C5orf30 | 0.496157634 | 1.873700557 | 7.678117481 | 0.000843717 | 0.042017825 | 0.102136594 |
| PGM2L1 | 0.543052851 | 6.516537916 | 7.669029564 | 0.000848062 | 0.042017825 | 0.096615126 |
| SMAD7 | 0.510900171 | 7.087704928 | 7.667419365 | 0.000848834 | 0.042017825 | 0.095636115 |
| LINC01426 | 0.581720797 | 3.114658104 | 7.664332848 | 0.000850317 | 0.042017825 | 0.093758888 |
| PDE4B | -0.587542661 | 6.304242806 | -7.648136476 | 0.000858152 | 0.042116295 | 0.083895198 |
| AXIN2 | 0.528103311 | 6.224296226 | 7.646299977 | 0.000859045 | 0.042116295 | 0.082775375 |
| MEIS2 | 0.934724706 | 4.055718934 | 7.641281052 | 0.000861494 | 0.042116295 | 0.079713602 |
| LTBP4 | -0.891616774 | 6.496952067 | -7.635170054 | 0.000864486 | 0.042116295 | 0.075982767 |
| CRYBG3 | -0.630521425 | 5.383322918 | -7.623707103 | 0.000870133 | 0.042242657 | 0.068976061 |
| COMP | -0.645740156 | 10.02919853 | -7.605649728 | 0.000879118 | 0.042529654 | 0.057916136 |
| IL33 | 0.915385751 | 5.932505824 | 7.596289828 | 0.00088382 | 0.042608121 | 0.052172504 |
| NRBF2 | 0.546224171 | 6.051391473 | 7.563125659 | 0.000900723 | 0.043177538 | 0.031761953 |
| TRNP1 | 0.803571787 | 2.240740395 | 7.555971548 | 0.00090442 | 0.043177538 | 0.027346796 |
| LOC728743 | -0.673482814 | 0.971341541 | -7.554866157 | 0.000904993 | 0.043177538 | 0.026664217 |
| ASPHD2 | -0.52569491 | 2.76902074 | -7.521035391 | 0.000922739 | 0.043778488 | 0.005723313 |
| HBP1 | -0.810417763 | 5.046302209 | -7.518817147 | 0.000923917 | 0.043778488 | 0.004346825 |
| YPEL2 | 0.627653803 | 4.634399389 | 7.49163798 | 0.0009385 | 0.044065351 | -0.012552951 |
| CLCF1 | -0.504793417 | 8.570625392 | -7.490420148 | 0.00093916 | 0.044065351 | -0.013311675 |
| SLC47A2 | -1.019373076 | 1.429046836 | -7.486621777 | 0.000941222 | 0.044065351 | -0.015678927 |
| SLC8A1 | -0.524217719 | 3.321020647 | -7.483884515 | 0.000942711 | 0.044065351 | -0.017385637 |
| COL21A1 | -0.562625572 | 2.300330146 | -7.448938234 | 0.000961973 | 0.044652033 | -0.039231896 |
| SYT14 | 0.571374592 | 2.659153829 | 7.443679189 | 0.000964912 | 0.044652033 | -0.042528688 |
| SCAMP1-AS1 | -0.682375435 | 1.036586969 | -7.432201809 | 0.000971365 | 0.044652033 | -0.049731983 |
| RGS10 | -0.621001498 | 3.966604877 | -7.427523828 | 0.00097401 | 0.044652033 | -0.052671211 |
| RGMB | 0.501603769 | 7.264579342 | 7.426535061 | 0.00097457 | 0.044652033 | -0.053292709 |
| PROB1 | -0.496138408 | 1.49861935 | -7.422827262 | 0.000976674 | 0.044652033 | -0.055624036 |
| FAM167A | -0.817517913 | 4.152384723 | -7.420755253 | 0.000977852 | 0.044652033 | -0.056927361 |
| ZNF367 | 0.527291951 | 3.452515955 | 7.407475007 | 0.000985444 | 0.044726308 | -0.065289742 |
| RGS2 | 0.514554785 | 7.78366618 | 7.406604668 | 0.000985944 | 0.044726308 | -0.065838321 |
| CACNA2D4 | -0.554649076 | 0.711810495 | -7.398317515 | 0.00099072 | 0.044796103 | -0.071065073 |
| RGL1 | -0.87047615 | 4.807930437 | -7.37902517 | 0.001001947 | 0.044967904 | -0.083256201 |
| AFF2 | -0.498468163 | 5.001357431 | -7.379008166 | 0.001001957 | 0.044967904 | -0.08326696 |
| C5orf38 | -0.502788691 | 0.921672249 | -7.375066225 | 0.00100427 | 0.044967904 | -0.085761965 |
| TNFAIP8 | 0.527585611 | 7.608046542 | 7.364127435 | 0.001010722 | 0.044979732 | -0.092692712 |
| COL14A1 | -0.523635002 | 10.39095518 | -7.356284891 | 0.001015378 | 0.044979732 | -0.097668196 |
| PLTP | -0.47231575 | 5.962121487 | -7.353065994 | 0.001017296 | 0.044979732 | -0.099711909 |
| SNTB1 | -0.50694421 | 5.399259197 | -7.352661038 | 0.001017538 | 0.044979732 | -0.099969085 |
| RTKN | -0.661821098 | 3.162999096 | -7.335507328 | 0.001027839 | 0.045290363 | -0.110876245 |
| DKK3 | -0.500245909 | 8.51080502 | -7.324451257 | 0.001034544 | 0.045389507 | -0.117920064 |
| HTATSF1P2 | 0.525747691 | 2.792280049 | 7.320996983 | 0.00103665 | 0.045389507 | -0.120123008 |

| | | | | | | |
|---|---|---|---|---|---|---|
| SLC16A9 | 0.819461671 | 2.99529336 | 7.312068305 | 0.001042116 | 0.045484932 | -0.125822145 |
| ADGRG2 | -1.384266902 | 2.366212331 | -7.272545885 | 0.001066735 | 0.046390441 | -0.151134654 |
| TMCC2 | -0.559332974 | 2.164833501 | -7.26806725 | 0.001069569 | 0.046390441 | -0.15401187 |
| RHOU | -1.133892367 | 1.670444713 | -7.249568957 | 0.001081368 | 0.046680125 | -0.165914876 |
| EPB41L4A | 0.770176247 | 2.835101334 | 7.241935478 | 0.001086283 | 0.046680125 | -0.170835746 |
| NUAK2 | -1.289928148 | 1.662517604 | -7.241802696 | 0.001086369 | 0.046680125 | -0.17092139 |
| LAMB1 | 0.610639519 | 7.681648257 | 7.218746904 | 0.001101378 | 0.047178553 | -0.185816427 |
| OC10028815 | -0.612213731 | 2.772783956 | -7.205547377 | 0.001110084 | 0.047390008 | -0.194365596 |
| ZNF189 | -0.475361142 | 4.942193671 | -7.200904573 | 0.001113165 | 0.047390008 | -0.197376449 |
| LIN7A | -0.482614291 | 1.935758331 | -7.189825921 | 0.00112056 | 0.047558487 | -0.204568879 |
| FXYD1 | -0.459438975 | 3.261817936 | -7.162017496 | 0.001139383 | 0.048059792 | -0.22267194 |
| TSHZ2 | 0.509507272 | 4.54078191 | 7.136947161 | 0.001156678 | 0.048059792 | -0.239053347 |
| SYT12 | -0.671620347 | 2.48630045 | -7.133358294 | 0.00115918 | 0.048059792 | -0.241403114 |
| SLC29A1 | -0.646836512 | 6.222446577 | -7.128109905 | 0.00116285 | 0.048059792 | -0.244841572 |
| ANK3 | 0.978264244 | 1.588537945 | 7.128023778 | 0.001162911 | 0.048059792 | -0.244898019 |
| RAB11FIP5 | -0.502927445 | 6.72779304 | -7.127049822 | 0.001163593 | 0.048059792 | -0.245536389 |
| SLC2A1 | 0.495136189 | 7.434177235 | 7.125372772 | 0.00116477 | 0.048059792 | -0.246635802 |
| RSPO3 | 0.584256296 | 3.757152306 | 7.12514644 | 0.001164929 | 0.048059792 | -0.246784197 |
| RGS3 | 0.466324342 | 8.528155688 | 7.122512733 | 0.00116678 | 0.048059792 | -0.248511338 |
| LCE1F | 1.054453254 | 2.022617952 | 7.120338904 | 0.00116831 | 0.048059792 | -0.249937383 |
| GABPB1 | -0.588591042 | 5.114378961 | -7.117120121 | 0.00117058 | 0.048059792 | -0.252049726 |
| FADS2 | 0.594393732 | 6.029954988 | 7.111294854 | 0.001174703 | 0.048086356 | -0.255875027 |
| USP54 | 0.764503165 | 4.532387281 | 7.100730181 | 0.001182224 | 0.048251474 | -0.262820599 |
| GDF15 | 0.612856004 | 5.466154761 | 7.088575092 | 0.001190949 | 0.04846461 | -0.270824591 |
| SLIT3 | 0.486224459 | 10.18519078 | 7.079937565 | 0.001197196 | 0.04854026 | -0.276520664 |
| SEMA3E | -0.541819206 | 1.655065867 | -7.076319327 | 0.001199824 | 0.04854026 | -0.278908802 |
| FRY | 0.746151825 | 4.281996428 | 7.063190786 | 0.00120942 | 0.048691416 | -0.287584269 |
| RILPL2 | -0.871926231 | 4.321994979 | -7.061586477 | 0.001210599 | 0.048691416 | -0.288645517 |
| NUAK1 | 0.873026015 | 5.657019081 | 7.044074521 | 0.001223557 | 0.049068655 | -0.300245328 |
| ALDH1A3 | 0.460963554 | 5.192113711 | 7.039364669 | 0.001227071 | 0.049068655 | -0.303370016 |
| H19 | 0.687302673 | 4.161057153 | 7.02623611 | 0.001236929 | 0.049287847 | -0.312090992 |
| CNKSR3 | -0.452004192 | 3.844501176 | -7.016792803 | 0.001244079 | 0.049287847 | -0.318373988 |
| CPXM2 | 0.471326254 | 6.196999712 | 7.010919073 | 0.001248551 | 0.049287847 | -0.322286252 |
| BASP1 | 0.461565391 | 8.631156332 | 7.010566983 | 0.001248819 | 0.049287847 | -0.32252087 |
| ZNF503 | 0.503001451 | 7.70691627 | 7.00392102 | 0.001253904 | 0.049287847 | -0.326951643 |
| ST6GALNAC4 | 0.521427333 | 7.501832837 | 7.003892103 | 0.001253926 | 0.049287847 | -0.326970931 |
| RNF19B | 0.499048035 | 6.153964667 | 6.995606819 | 0.0012603 | 0.049398043 | -0.332500495 |
| TMEM151A | -0.665426966 | 1.98230925 | -6.985633004 | 0.001268024 | 0.04948504 | -0.339165611 |
| KCTD12 | 0.491511803 | 6.992862069 | 6.983514083 | 0.001269673 | 0.04948504 | -0.34058282 |
| TPPP3 | 0.675352919 | 1.912908638 | 6.961539387 | 0.001286919 | 0.049745072 | -0.355305466 |
| ALPK2 | 0.672091059 | 2.682690961 | 6.960664273 | 0.001287612 | 0.049745072 | -0.355892729 |
| NCAPG | 0.79781661 | 2.562097653 | 6.956014818 | 0.001291299 | 0.049745072 | -0.359014068 |
| SPX | -0.585387966 | 0.718507706 | -6.953955719 | 0.001292936 | 0.049745072 | -0.360397072 |
| SHOX2 | -0.771897484 | 4.508996276 | -6.949278474 | 0.001296664 | 0.049745072 | -0.363540072 |
| PTPN22 | 0.48712858 | 1.854313663 | 6.940700217 | 0.001303535 | 0.049745072 | -0.369309902 |
| PVRL3 | -0.53075214 | 5.683066881 | -6.938867001 | 0.001305009 | 0.049745072 | -0.370543857 |
| MITF | 0.468133922 | 4.760929569 | 6.934578935 | 0.001308464 | 0.049745072 | -0.373431454 |

| | | | | | | |
|---|---|---|---|---|---|---|
| KCNK15 | 0.466150124 | 5.212030008 | 6.92917082 | 0.001312838 | 0.049745072 | -0.377075811 |
| SAMD4A | -0.484871525 | 7.196836385 | -6.925402497 | 0.001315896 | 0.049745072 | -0.379616825 |
| SYTL4 | -0.735945842 | 2.069022403 | -6.924981885 | 0.001316238 | 0.049745072 | -0.379900532 |
| PHLDA3 | 0.457290471 | 7.467224057 | 6.920989314 | 0.001319488 | 0.049745072 | -0.382594409 |
| ZNF785 | -0.444586001 | 2.11390264 | -6.912534912 | 0.001326403 | 0.049869863 | -0.388303846 |

**IL1B (20ng/mL) stimulation**

| | logFC | AveExpr | t | P.Value | adj.P.Val | B |
|---|---|---|---|---|---|---|
| NFATC2 | -7.889369833 | 5.868501206 | -29.84378071 | 2.18E-06 | 0.017175883 | 4.090289736 |
| EPB41L3 | 1.86892663 | 3.153695179 | 28.98474657 | 2.48E-06 | 0.017175883 | 4.050278576 |
| SFRP2 | 1.414418015 | 5.160573527 | 21.99250845 | 8.64E-06 | 0.039836356 | 3.578969179 |
